# Syndemic Geographic Patterns of Cancer Types in a Health Deprived Area of England: a new Paradigm for Public Health Cancer Interventions?

**DOI:** 10.1101/2024.02.24.24303312

**Authors:** Catherine Jones, Tom Keegan, Andy Knox, Alison Birtle, Jessica A. Mendes, Kelly Heys, Peter Atkinson, Luigi Sedda

**Affiliations:** University Hospitals of Morecambe Bay NHS Foundation Trust, Kendal, LA9 7RG, UK; Lancaster Medical School, Lancaster University, Lancaster, LA1 4YG, UK; Lancaster Ecology and Epidemiology Group (LEEG), Lancaster University, Lancaster, LA1 4YG, UK; NHS Lancashire and South Cumbria Integrated Care Board, Preston, PR1 8XB; Rosemere Cancer Centre, Lancashire Teaching Hospitals, Preston, PR 29HT; Centre for Tropical Medicine and Global Health, University of Oxford, Oxford, OX3 7LG, UK; Faculty of Science and Technology, Lancaster University, Lancaster, LA1 4YG, UK

**Keywords:** Synchronic diseases, geospatial analyses, joint modelling, left-censoring, variable selection, common cancers, North West of England, Morecambe Bay

## Abstract

Cancer poses a significant public health challenge, and accurate tools are crucial for effective intervention, especially in high-risk areas. The North West of England, historically identified as a region with high cancer incidence, has become a focus for public health initiatives. This study aims to analyse cancer risk factors, demographic trends and spatial patterns in this region by employing a novel spatial joint modelling framework designed to account for large frequencies of left-censored data.

Cancer diagnoses were collected at the postcode sector level. The dataset was left-censored due to confidentiality issues, and categorised as interval censored. Demographic and behavioural factors, alongside socio-economic variables, both at individual and geographic unit levels, were obtained from the linkage of primary and secondary health data and various open source datasets. An ecological investigation was conducted using joint spatial modelling on nine cancer types (breast, colorectal, gynaecology, haematology, head and neck, lung, skin, upper GI, urology), for which explanatory factors were selected by employing an accelerated failure model with lognormal distribution. Post-processing included principal components analysis and hierarchical clustering to delineate geographic areas with similar spatial patterns of different cancer types.

The study included 15,506 cancer diagnoses from 2017 to 2022, with the highest incidence in skin, breast and urology cancers. Preliminary censoring adjustments reduced censored records from 86% to 60%. Factors such as age, ethnicity, frailty and comorbidities were associated with cancer risk. The analysis identified 22 relevant variables, with comorbidities and ethnicity being prominent. The spatial distribution of the risk and cumulative risk of the cancer types revealed regional variations, with five clusters identified. Rural areas were the least affected by cancer and Barrow-in-Furness was the area with the highest cancer risk.

This study emphasizes the need for targeted interventions addressing health inequalities in different geographical regions. The findings suggest the need for tailored public health interventions, considering specific risk factors and socio-economic disparities. Policymakers can utilize the spatial patterns identified to allocate resources effectively and implement targeted cancer prevention programmes.

## Introduction

Geographic mapping is an essential public health tool that can identify spatial patterns of disease and differences in disease diagnosis, burden and mortality across one or more regions (Downing et al., 2008). This includes geographical distribution incidence, prevalence, survival and relative risks. Maps can be used for prevention and control programmes, and prioritisation of intervention, and services management, for example, by targeting high risk communities, but also to investigate the aetiology of a disease (DeChello et al., 2006).

The advantages of disease mapping can be enhanced by employing syndemic frameworks where multiple diseases that share common risk factors are modelled and mapped jointly (Downing et al., 2008). A geographic syndemic framework is composed of two or more geographically co-occurring diseases that interact with each other and the environment (in the ecological sense) (Shrestha et al., 2020). This framework requires shared risk factors or common components such as those defining the spatial variation of the diseases, but allows for other risk factors or components to exist at the individual disease level.

Cancers are an ideal candidate disease complex to be analysed by syndemic frameworks due to the shared risk factors of different cancer types (socio-demographic, anthropometric and lifestyle factors), and their common empirical geographical patterns (Jahan et al., 2020, Gomez-Rubio et al., 2019). Methodologically, syndemic frameworks can be analysed by employing joint models whose joint component can include the risk factors affecting the average risk (named fixed effects) of the cancers and/or variables influencing the variability of the cancers (named random effects, as for example, spatial, temporal and spatial–temporal autocorrelations) caused by the dependency and association between measurements in space and/or time (Król et al., 2016). It has been shown in other studies that joint modelling results in better performance than individual models with an independent set of random effects (Wah et al., 2020, Kouame et al., 2023). Shared random effects can identify geographic areas with unmeasured or unknown risk factors that are common for different cancers among multiple population groups for gender, ethnicity and age distributions (Wah et al., 2020).

Providing accurate and advanced public health tools and information to tackle cancer morbidity and mortality are particularly needed in the areas at most risk for cancer occurrence. The North West of England is one of these areas. Historically, in the scientific literature, the North West has been identified as one of the regions most affected by cancer in England. In fact, it had the highest oral cancer incidence rates in the period 1990-1999 (for both males and females driven by the over 45 years old) (Conway et al., 2006); the highest uveal melanoma admission rates between 1999-2010 (Keenan et al., 2012) with Lancashire also experiencing the highest cutaneous melanoma; the second highest glioblastoma (a fast growing type of brain tumour) incidence and the lowest survival in 2007-2011 (Brodbelt et al., 2015); higher incidence (2008-2010) and mortality (2009-2011) rates, compared to England, of head and neck cancer in the Merseyside and Cheshire region (Taib et al., 2018); and finally the lowest incidence of basal cell carcinoma (the most common skin cancer) between 2004 and 2010 (Musah et al., 2013). Compared with London, the North West of England had a higher incidence rate for liver cancer subtypes hepatocellular carcinoma and intrahepatic cholangiocarcinoma between 2008 and 2018 (Liao et al., 2023); while compared to the whole UK, the North of England had the highest rates of lung cancer in non-smokers for men and women during the period 1998 to 2007 (Rait and Horsfall, 2020).

Morecambe Bay and South Cumbria, the focus of the present study, have been recognized as priority areas for public health initiatives, due to poor health outcomes for some portions of their populations. According to a recent report on coastal towns (Asthana and Gibson, 2021), 20% of people smoke in Morecambe (16.6% nationally), a town of almost 33,000 inhabitants. Further, residents have high rates of hospital admission for alcohol-related harm, and are more likely to have hypertension or depression than the national average, with a quarter having a limiting, long-term illness or disability, significantly more than the national average. In addition, Morecambe has worse values for all emergency hospital admission indicators, and higher standardised mortality ratios for all ages and under 75s. According to the same study, people in Morecambe are more likely to have lung cancer, peripheral artery disease, COPD, dementia, stroke, coronary heart disease, kidney disease, epilepsy and diabetes than the national averages. Deprivation rates are significantly worse than the England average.

Morecambe Bay and South Cumbria area also notable for the presence of nuclear power stations at Heysham in Lancashire and Sellafield in Cumbria. The latter raised concerns in 1957 after an uncontrolled release of iodine-131 into the atmosphere; the health consequences of this event, such as an possible increase in cancers, have been debated with most studies not finding evidence for increased cancer risk (McNally et al., 2016, Bunch et al., 2014b). Similar results were found for other point exposures (where the exposure is fixed at a location), including powerlines (Bunch et al., 2014a, Elliott et al., 2013, Bithell et al., 2013, Stark et al., 2007, Sehmer et al., 2014).

Socioeconomic differences in incidence and survival due to environment, lifestyle, biological effects, access to health care and health seeking behaviour, exist for different cancers even between neighbouring geographical areas (Phillips et al., 2019) and in particular for breast, cervix, lung and malignant melanoma (Shack et al., 2008). However, this has not been established for childhood leukaemia prevalence in 1993–1994 in England, Wales, and Scotland (Smith et al., 2006), or colorectal cancer survival between 1991 and 1997 in Bolton (North West of England) (Lyratzopoulos et al., 2004), although other studies have shown an association with survival and uptake of screening (Mansouri et al., 2013, McCaffery et al., 2002). An inverse socio-economic gradient was found for non-melanoma skin cancer rates in different studies covering Ireland (1994-2003) (Carsin et al., 2011), England (2006-2008) (Wheeler et al., 2013), and the whole of the UK (2004-2010) (Musah et al., 2013), and for breast cancer incidence in Wales (1985-2012)(Abdulrahman, 2014).

Given the complexity of relationships between demographic, behavioural and socio-economic conditions, and cancer morbidity measures, evaluation of geographical risk must consider the local scale factors influencing cancer dynamics. Variation within the study region, apart from revealing important local differences in risk can, in turn, provide aetiological clues. In the present study, we developed a Bayesian hierarchical joint spatial analysis for left-censored counts of newly diagnosed cancer types (censored to remove patients’ identifiability), provided for each postcode sector, to investigate the variation in each cancer risk, joint cancer risk and cancers’ correlations, across the Morecambe Bay region in the North West of England.

## Data

### Study area

The study area is Morecambe Bay in the North West of England. The Morecambe Bay study area was defined using the limits of the Morecambe Bay ex Clinical Commissioning Group (Figure 1, red border) to simplify data acquisition and homogenisation. CCGs formed the lowest level of the health geography hierarchy in England which was introduced by the Health and Social Care Act 2012. Since patients were recruited from a larger region (Figure 1, black border), the study area was extended to include these patients.

**Figure 1.**
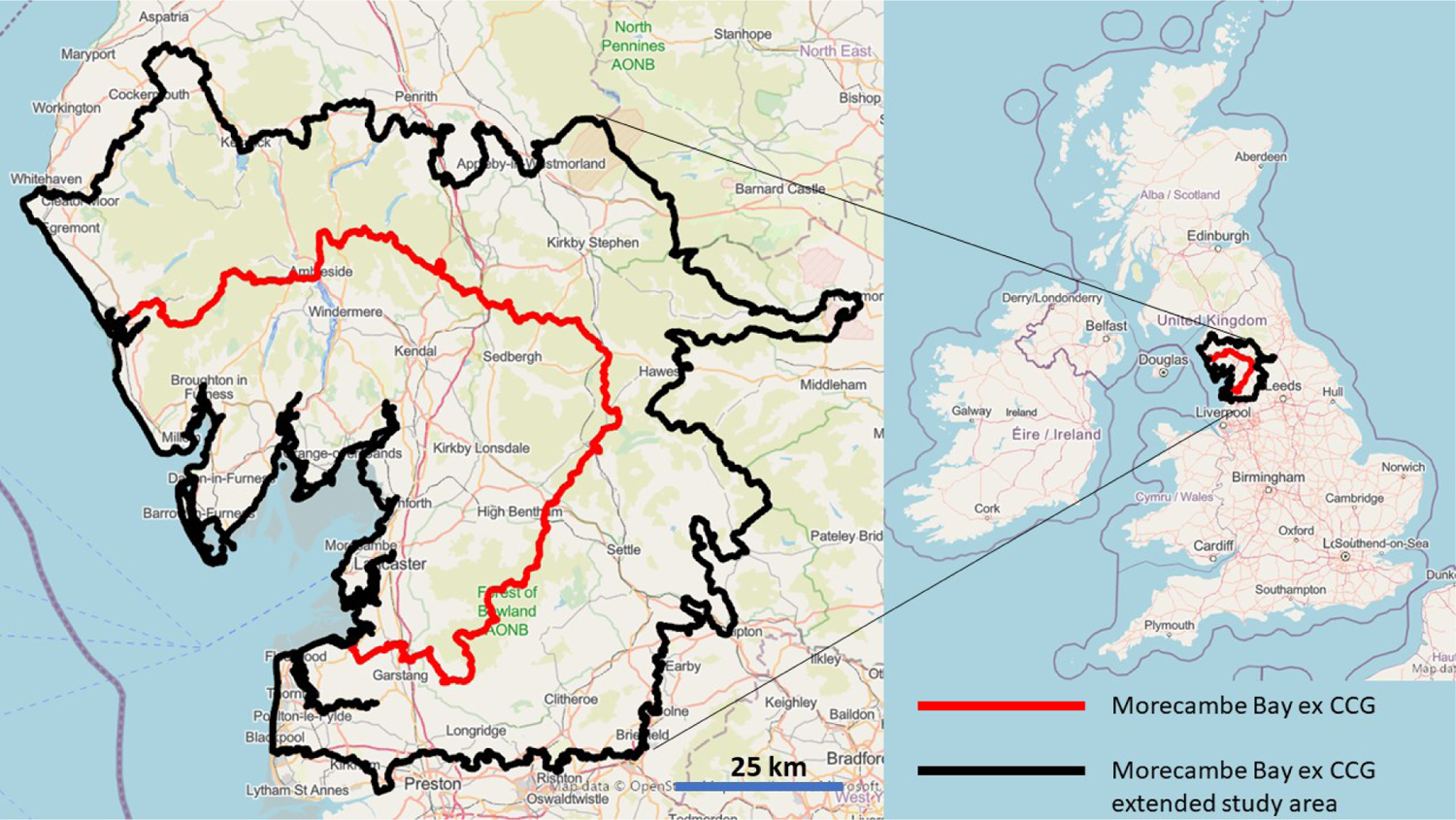
Morecambe Bay ex CCG (red border) and study area (black border). Basemap from openstreetmap under the Open Database licence (https://www.openstreetmap.org/copyright), Microsoft, Facebook Inc and its affiliates, and Esri Community Maps contributors. Map layer by Esri.

In 2000, Morecambe Bay ex CCG had a population of 334,287. The Index of Multiple Deprivation of 2019 ranked Morecambe Bay CCG 99^th^ out of 191, but 53^rd^ for its proportion of Lower Super Output Areas (LSOAs) in the 10% most deprived LSOAs in England^1^. A lower super output area typically has between 400 and 1,200 households and a resident population of between 1,000 and 3,000 residents^2^.

The age distribution of Morecambe Bay ex CCG is reported in Supplementary File 1.

### Study design and data collection

An ecological study design was used for this research. The primary outcome is the 6-months new cancer diagnosis count by cancer type from January 2017 to December 2022 (for a total of 12 temporal measurements). The data were extracted in August 2023. The geographic unit of analysis is the postcode sector which includes the first part of the postcode (postcode district), the single space, and the first character of the second part of the postcode (inward code)^3^, for a total of 75 postcode sectors. The same data were also provided at Local Authority district level, for a total of 13 districts. The data were provided by University Hospitals of Morecambe Bay for the Morecambe Bay ex CCG extended study area (Figure 1).

Cancers were classified into different types by their anatomic location (site)(Rachet et al., 2008). Nine cancer types were considered in this study: lung, skin, breast, colorectal, haematology, upper gastrointestinal (upper GI), urology, head and neck and gynaecology. Sarcoma, brain and central nervous system cancers were excluded because they were fully censored. The ICD10 Codes used to identify each cancer by its tumour site are reported in Supplementary File 2. Lung cancer contributes to the largest number of cancer deaths in the world (Rait and Horsfall, 2020) while skin cancer is the most common in the general population (Saleh et al., 2017), although prostate cancer is the most common in men, and breast cancer in female (Nogueira et al., 2019).

After lung cancer, colorectal cancer is the second highest cause of cancer deaths in the United Kingdom (Lal et al., 2020). Haematological cancers are the third largest cancer killers in the UK^4^ and increasingly important worldwide, with 1.3 million new cases in 2020 and 700,000 deaths (Chen et al., 2022). Within the upper GI cancers, oesophageal cancer in the UK has one of the highest age-standardised rates within the oesophageal cancers world wide (Arnold et al., 2020). Head and neck cancer, a diverse group of more than 30 different subsites, is the 6th most common cancer worldwide and one of the most debilitating, although it constitutes only 3% of all cancers in the UK (Taib et al., 2018). Finally, every year in the UK, more than 21 000 women (5% of all cancers) are diagnosed with a gynaecological cancer and around 8000 die^5^ (Knapp et al., 2021).

### Censored cancers

Public health data, such as new cancer diagnoses, are often left-censored due to confidentiality issues. The postcode sector and Local Authority district count data used here, of cancer new diagnosis by type, were censored, with values less than or equal to 5 replaced by the words ‘less than or equal to 5’. However, because zero was not included in the censoring, it was more appropriate to describe the censoring as interval censoring, with the censored values being larger than 0 and smaller than 6 (Wong and Yu, 1999). This means that the dataset contains nonignorable or ‘missing not random’ values (Leacy et al., 2017). In other words, the missing data isn’t just randomly scattered throughout the dataset, but instead, there’s a pattern. To handle this, it was necessary to make statistical assumptions on the distribution of the missing data in order to estimate the missing values.

### Exposure and predictor variables

The following demographic and behavioural factors aggregated by postcode sector and provided as counts of patients by cancer type were obtained from the University Hospital of Morecambe Bay. These factors acquired by linking primary and secondary data are:

- age group;
- gender;
- ethnicity (African, any other Asian background, any other black background, any other ethnic group, any other mixed background, any other white background, Asian and Chinese, British, British Asian, Caribbean, Chinese, Chinese and white, English, Filipino, Indian, Irish, Italian, Northern-Irish, not known, other mixed white, other white or white unspecified, other white European, Pakistani, Polish, Scottish, Welsh, white and Asian, white and black African, white and black Caribbean);
- frailty score (fit, mild, moderate and severe);
- smoking information (never smoked, ex smoker, current smoker, unknown);
- co-morbidities (chronic kidney disease and stage, chronic obstructive pulmonary disease, congestive heart failure, coronary heart disease, depression, anxiety, hypertension, diabetes type and COVID19).

In addition, and for each local authority other factors were extracted from the following datasets: police crime reports (from data.police.uk), household data (Office of National Statistics, Census of 2011), the index of multiple deprivation (Ministry for Housing, Communities, and Local Government), and place-based longitudinal data which includes health, social and behavioural variables (pldr.org). The full list of the 787 variables included in the analyses is available in Supplementary File 3.

The Index of Multiple Deprivation (IMD) is the official measure of relative deprivation for LSOAs in England. Every LSOA in England is ranked from 1 (most deprived or poor) to 5 (quintile) or 10 (decile) (least deprived or most affluent), but there is no definitive cut-off below which an area is considered ‘deprived’ (Rogers et al., 2019).

While linked primary and secondary data were available at postcode sector level, the remainder of the data were available at a coarser resolution (LA or LSOA), as common in geographic health analyses (McNally et al., 2003, Manda et al., 2009, Wheeler et al., 2013). Population estimates were required for calculating cancer risk and rates. These were obtained at LSOA level from the 2011 Census for England and Wales.

## Statistical analyses

The current analysis incorporated an ecological investigation to assess cancer risk at individual and population level exposures (Chidumwa et al., 2021). The first analysis reduced the number of censored cancers by combining local authority with postcode sector data. The next analysis identified important factors associated to cancers counts, followed by joint spatial modelling. Finally, we calculated summary statistics and produced geographic cancer risk maps for individual cancer type, cumulative risk for all cancers, and correlated cancer risks.

### Censoring reduction

Before any statistical analyses started, the number of censored data was reduced by combining the Local Authority (LA) with the postcode sector (PS) data. The following rules were used (for each cancer type and time period):

a. if within an LA (containing multiple PS) only one PS was censored, then the number of patients in the censored PS was obtained by the difference of the number of diagnosis in the LA minus the sum of patients in the non-censored PS belonging to that LA;
b. if the number of diagnosis in the LA (or the residual number of diagnosis after deducting diagnosis from non-censored PS) was equal to the number of censored PS, then each censored PS was associated with one diagnosis;
c. if the number of diagnosis in the LA (or the residual number of diagnosis after deducting diagnosis from non-censored PS) is equal to five times the number of censored PS, then each censored PS was associated with five diagnoses.

In all the other cases, the number of censored patients in each PS and time period, was converted to 1 to 2, 1 to 3, 1 to 4 and 1 to 5 censoring level depending on the total number or residual number of diagnoses at the LA unit level. For example, if an LA had nine censored diagnosis and three censored PS, each censored PS will have a censoring level of 1 to 5 (since it is still possible for one PS to have 5 diagnoses). However, if the LA had 5 diagnoses, then the three censored PS will be censored 1 to 3 since none of them can have all five or four diagnoses.

### Descriptive statistics

Due to the high proportion of censoring in the cancer data, it was not possible to provide statistics to describe the demographic (age, sex, ethnicity), socio-economic and clinical characteristics (comorbidities and frailty) of the real study population. Post-analyses statistics were provided as crude cancer rates by cancer type, and age-adjusted rates averaged by postcode sector and 6 months period (age-specific incidence rate). We considered two age groups, 0-50 and 50+ because the level of censoring did not allow to create multiple age thresholds. A single age threshold was used in other cancer research (Downing et al., 2010, Shack et al., 2008). The standard errors of the incidence and age-adjusted incidence rates were calculated using the Poisson approximation method (Boyle and Parkin, 1991).

### Variable selection

Using a Bayesian joint model for variables selection would have required exceptionally lengthy computations. For this reason, we employed a deterministic method that can account for censored and clustered data within the entire cancer dataset.

A stepwise selection method was applied to an accelerated failure model with lognormal distribution. The accelerated failure model allows the inclusion of censored data as the outcome (Kalbfleisch and Prentice, 2011), while cancer type and postcode sector were used as clusters for robust variance computation (to account for correlation within each cancer type). For the cancer counts *C*, the accelerated failure model is:

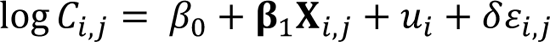

where *β*_0_ is the intercept, **β**_1_ is a vector of regression coefficients, *u* is the random effect, *δ* is scale parameter and, *ε* is the error distribution assumed to have a normal distribution. The subscript *i* refers to post-code sector at a given time period, and *j* to the cancer type.

The accelerated failure model was estimated using the maximum likelihood estimation method. The model’s performance was assessed using the Bayesian Information Criterion (BIC) and the best model was chosen as the one with the lowest BIC. The described model is mostly used in parametric mixed survival problems with left truncated data or Tobit regression, where the time to occurrence of an event was substituted here with the number of patients with a cancer type. The use of the proposed method for non-survival data has been described elsewhere (Kong and Nan, 2016).

### Inference and prediction

The joint modelling of two or more cancers allows the identification of shared and divergent trends among the cancers in terms of geographic patterns and risk factors. Rather than treating a cancer as a proxy for unmeasured risk factors affecting another cancer, the proposed model treats the different cancer types symmetrically and assumes that the area-specific relative risks of each depend on a shared latent component plus additional latent components specific to one or other cancer types (Best and Hansell, 2009). The joint model employed in this study is the shared component model (Cai et al., 2020) for Poisson distributed data.

The log of the number of cases observed in each postcode sector was the dependent variable, and the individual and ecological variables obtained from the variable selection step were the independent variables. The joint model is spatially explicit (by modelling the spatial autocorrelation common to all the cancer types) and Bayesian, and for these reasons adapted to stabilise risk estimates based on small numbers at postcode sector level (Jarup et al., 2002). This means that the resulting relative risk estimate for each postcode sector is a form of smoothed average of the observed risk and the mean relative risk in the neighbouring postcode sectors conditioned to a set of prior assumptions.

The common shared component is the spatial dependence, which results from common factors interacting with the cancers, but not represented by independent variables. If spatial dependence is disregarded, inferences are likely to be biased (Nogueira et al., 2019).

Since the cancer counts are censored, the joint model’s maximum likelihood estimation method integrates the censored data into the likelihood function. Therefore, the observed cancers’ counts are simultaneously modelled with the censored data. Within this approach the method first models the complete response values, including both the observed and the unobserved values, and then models the censored values conditioned on the complete response values (Qu et al., 2023). To facilitate the computation of the spatial dependence structure the data were modelled using a log Gaussian process (Qu et al., 2023). The main advantage of the likelihood-based approach is that it utilizes all the information available (Leung et al., 1997).

Formally, let *Y_j_*_,*s*_ represent the observed (including censored) number of cases of cancer type *j* at postcode sector *s*. The joint modelling follows the multivariate spatial Bayesian log-Gaussian model for left-censored data proposed by Sahoo and colleagues (Sahoo and Hazra, 2021) adapted to Poisson (Po) processes (Gomez-Rubio et al., 2019) and to account for censoring levels varying among locations. *Y* follows a Poisson distribution with mean *E_j_*_,*s*_*θ_j_*_,*s*_:

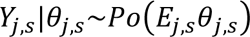

where *E* is the expected number of cases and *θ* is the relative risk modelled using a Poisson log-linear model within a multivariate spatial Bayesian framework:

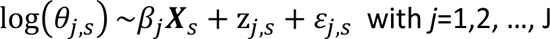

where **X** is the (full rank) design matrix containing the covariate values (obtained from the variable selection step described above and common to all cancers), *β* are unknown regression coefficients, *z* is a multivariate spatial zero mean Gaussian process, at every location *s* and for each joint process *j* with non-singular covariance matrix **∑**:

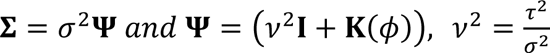

where **Κ** is the exponential spatial correlation matrix, *ν* is the ratio between the spatial variance *τ*^2^ and the total variance *σ*^2^, and **Ι** is the identity matrix. Finally, *ε* is the multivariate nugget effect (pure error term), zero-mean normally distributed with covariance (1-ν^2^)**∑**.

The data of size *n* consist of *m* exact observations and *n*-*m* (censored) interval observations at locations *s*\*. Therefore, *Y*^*c*^_j_ is left-censored with censoring level *u_j_*_,*s*\*_, with *u_j_*_,*s*\*_ being one of the following integer intervals: ⟦1,2⟧, ⟦1,3⟧, ⟦1,4⟧ and ⟦1,5⟧, depending on the cancer type and location.

The likelihood for censored spatial data of cancer *j*, in its generic form, is:

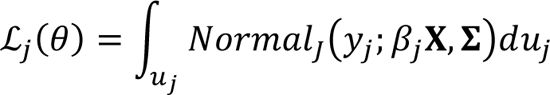

The set of parameters and hyperparameters are:

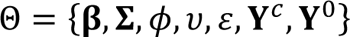

where **Y**^c^ is the set of censored cancer observations and **Y**^0^ the set of cancers to be predicted. Inference on these parameters is based on Markov chain Monte Carlo (MCMC) sampling. The regression coefficients **β** are assumed to have normal priors with zero mean and ten standard deviations; the covariance **Σ** follows an Inverse-Wishart distribution with scale 0.01 and degrees of freedom 0.01**I**; the spatial range parameter ø is uniformely distributed between 0 and 50 km (approximately half of the maximum distance between two postcode-sectors); the ratio of spatial to total variance ν is uniformely distributed between its natural limits of 0 and 1; finally the pure error term ε is normally distributed with mean zero and variance νΣ_s_⊕Σ, with **Σ**_s_ been the spatial covariance. Censored values are inputted at every iteration from a Truncated-Normal distribution for each cancer type. In practice, by using conventional equations the truncated-normal distribution probability at each censored value is used to estimate the inputted binomial value (Horgan, 2019).

Θ relative posteriors and full MCMC algorithm are described elsewhere (Sahoo and Hazra, 2021).

We ran the MCMC for 100,000 iterations and discarded the first 20,000 iterations. The remaining rest of 80,000 iterations were thinned by keeping one every fifty. Convergence was assessed by a visual examination of trace plots. Results were based on posterior sample sizes sufficient to give Monte Carlo standard errors less than 5% of the posterior standard deviation for the parameters of interest (Best and Hansell, 2009)

Predictions were made at the LSOA level. The following outcomes were mapped:

1. cancer risk prevalence by type for the general population, over 50 years old and 50 years old or under;
2. cancer cumulative risk prevalence by number of cancers for the general population;
3. geographical correlation (co-regionalisation) between pairs of cancer types;
4. cancers’ geographic clusters.

All the outcomes are given in terms of model posterior means (Gomez-Rubio et al., 2019). Uncertainty is represented by the posterior standard deviation of the outcome or parameter of interest, with smaller values indicating more certainty in the inference of the parameter or the prediction of the outcome. Uncertainty enables a better understanding of the information produced and more informed decision making based on this information. For example, large uncertainties may be dependent on the vaguely defined or non-informative assumptions made for the model, the absence of model components, and/or the lack of predictive capacity from the employed risk factors. As stressed by Roberts and colleagues (Roberts et al., 2016), ignoring and/or not understanding uncertainty information can result in misinterpretation of model outputs, substandard decision making, or disregard of important information due to its large uncertainty. To facilitate comparisons, all maps are shown on the same scale (Carsin et al., 2011). Finally, we carried out a mediational analysis (Tingley et al., 2014) to examine the effects of including each of the potential mediating variables in the individual-level cancer type generalised linear models on the odds ratios associated to each selected variable.

### Model performance and validation

Model performance was assessed for each individual cancer type through the mean error and mean squared error difference between the observed and the predicted outcomes. To account for the complex parameterisation of the joint model and to evaluate the global and individual (each individual model for cancer type of the joint model) predictive accuracy we used two information criteria: Deviance Information Criterion (DIC) and Watanabe-Akaike Information Criterion (WAIC).

Both measures are designed for Bayesian analyses. However, the WAIC averages over the posterior distribution rather than conditioning on a point estimate as in the DIC (Gelman et al., 2013). WAIC often produces values with small differences between models with similar structure. Therefore, we decided to report both WAIC and DIC estimates to provide evidence of consensus between the two statistics.

Finally, robustness of the model was assessed using cross-validation by leaving out 10% of the data for each cancer type. Validation assessment was carried out by measuring the Root Mean Square Error and the Mean Squared Deviation Ratio (Liao et al., 2022).

### Clustering

Principal components analysis (PCA) was used to identify co-regionalisation between cancers and geographic cancer clusters. PCA was employed on the posterior predictions of the latent variable for each cancer type. Based on the first two principal components (explaining the greatest variability) a centroid hierarchical cluster analysis (Vichi et al., 2022) was employed to cluster the LSOAs based on similarity of the cancer patterns.

### Software

All the analyses were performed in the R-cran software (R Core Team, 2023) using various packages and the authors’ written codes for model inference, predictions, and mapping.

## Results

### Descriptive statistics and censoring reduction

During 2017–2022 the University Hospitals of Morecambe Bay recorded 15,506 individuals diagnosed with one of the nine cancer types in the Morecambe Bay ex CCG extended area. These cancer types were 4,599 skin, 2,450 urology, 2,076 breast, 1,606 colorectal, 1,535 lung, 1,039 upper GI, 992 haematology, 670 gynaecology, and 539 head and neck cancers. Our model predictions were accurate with 15,243 estimated cancers (an error of 1.7%). However, the accuracy was reduced for upper GI (9.2% underestimation) and, to a lesser extent, relatively high for lung cancer (2.3% overestimation) (Table 1).

**Table 1.** Predicted and real total number of diagnosed cancers, and relative rates, in the study area between 2017 and 2022. 95%CRL is the lower limit of the 95% credible interval for the model-predicated cancer counts; 95%CRU is the upper limit of the 95% credible interval for the model-predicated cancer counts. * data from the Office of National Statistics for the year 2017. ** data from Cancer Research UK for the period 2016-2018.

| Cancer type | Cancer counts |  |  |  |  | Rates x 100,000 per year |  |  |  |
| --- | --- | --- | --- | --- | --- | --- | --- | --- | --- |
|  | Model predicted | Real | Error (%) | 95%CRL | 95%CRU | Model | Real | North West* | England** |
| Breast | 2011 | 2076 | 3.13 | 1389 | 2181 | 100.26 | 103.5 | 95 | 83 |
| Colorectal | 1680 | 1606 | -4.61 | 822 | 2200 | 83.76 | 80.07 | 72 | 69 |
| Gynaecology | 708 | 670 | -5.67 | 490 | 994 | 35.3 | 33.4 | 36 | 32 |
| Haematology | 947 | 992 | 4.54 | 590 | 1131 | 47.21 | 49.46 | 48 | 47 |
| Head and Neck | 519 | 539 | 3.71 | 391 | 931 | 25.88 | 26.87 | 28 | 18 |
| Lung | 1570 | 1535 | -2.28 | 1135 | 2009 | 78.28 | 76.53 | 92 | 76 |
| Skin | 4276 | 4599 | 7.02 | 3276 | 4922 | 213.19 | 229.29 | 254 | 257 |
| Upper GI | 943 | 1039 | 9.23 | 628 | 1145 | 47.02 | 51.8 | 61 | 50 |
| Urology | 2589 | 2450 | -5.67 | 1999 | 2714 | 129.08 | 122.15 | 108 | 98 |

A direct comparison with cancer incidence at national and regional level was not possible because open-source data was available for different length of periods than the one considered in our study (Table 1): only available for one year from the Office of National Statistics, and slightly older data from Cancer Research UK (2016-2018). Compared to these official rates we found that the rates for breast, colorectal, haematology and urology cancers were larger in Morecambe Bay ex CCG extended area than in the North West and England. For urology, this difference was 24 new diagnoses every 100,000 more per year than in England and 14 more than in the North West. Skin cancers were lower compared to the North West and England, with 25 new diagnoses per year less than in the North West, and 28 less than in England.

The capacity of the model to return accurate results was also attributable to the adopted preliminary censoring reduction step. In fact, this preliminary adjustment reduced the number of censored records (one record containing the number of diagnoses for a cancer type in a postcode sector during a 6 months period) to up 86% (for cancer type ‘others’ not considered in this study). Head and neck cancer censored records reduced by 84% (from 366 censored records to only 57), but breast and colorectal of less than 25% (Supplementary File 4).

As expected, all cancer types had a larger value of incidence in the over 50 years old group compared to the 50 years old and under group, with around a 20-fold difference for lung and upper GI; around 15-fold for skin and urology; and 11-fold for colorectal cancers. The largest incidence was in skin cancer, followed by urology, breast, colorectal and lung (all above 1 new case per 1000 people per post-code sector every 6-months) (Table 2). Geographically, four postcode sectors experienced all 12 cancer types between 2017 and 2022; 72% of postcode sectors had at least 10 cancer types during the same period (Supplementary File 5).

**Table 2.** Descriptive statistics for model-based cancer counts by cancer type per postcode sector and 6 months period. Incidence refers to 1000 people. SD = standard deviation, SEI = standard error incidence, SEI50 standard error for incidence in 50 years old and under, SEIO50 = standard error for incidence in over 50 years old. Standard errors were estimated using a Poisson approximation method (see methods).

| Cancer type | Mean | SD | Incidence | Incidence for 50 and under | Incidence for over 50 | SEI | SEI50 | SEIO50 |
| --- | --- | --- | --- | --- | --- | --- | --- | --- |
| Breast | 2.79 | 1.57 | 1.65 | 0.57 | 2.84 | 1.51 | 0.93 | 4.49 |
| Colorectal | 2.33 | 0.8 | 1.38 | 0.24 | 2.65 | 1.21 | 0.52 | 6.13 |
| Gynaecology | 0.98 | 0.47 | 0.58 | 0.16 | 0.82 | 0.46 | 0.35 | 1.34 |
| Haematology | 1.31 | 0.51 | 0.78 | 0.15 | 1.33 | 0.55 | 0.41 | 1.84 |
| Head and Neck | 0.95 | 0.45 | 0.56 | 0.14 | 0.69 | 0.42 | 0.35 | 1.28 |
| Lung | 2.18 | 1.02 | 1.29 | 0.11 | 2.37 | 1.03 | 0.41 | 3.82 |
| Skin | 9.55 | 6.06 | 5.68 | 0.81 | 12.41 | 5.34 | 1.28 | 17.77 |
| Upper GI | 1.31 | 0.46 | 0.78 | 0.07 | 1.43 | 0.49 | 0.21 | 1.94 |
| Urology | 3.59 | 2.07 | 2.13 | 0.32 | 4.53 | 1.87 | 0.71 | 6.17 |

### Variable selection

Variable selection reduced the initial 787 variables to 22. These variables, obtained from the best fitting model where all the cancer types are modelled simultaneously, can be grouped into three domains: demographic (age, frailty, ethnicity and comorbidities), behavioural (smoking status) and socio-economic (employment). Despite the absence of sex, all these domains are commonly associated with incident diagnosis of cancer (Liao, Coupland et al. 2023). Six out of 22 selected variables are area-based variables (or ecological variables) while the rest (16) are defined at the individual level (variables associated to cancer patients instead of the area where they live).

Summary statistics for these 22 variables are provided in Supplementary File 6. In addition to these 22 variables, time was also included to account for the presence of temporal trends (taking the overall number of variables to 23). When Time is a risk factor, it meant that from 2017 to 2022 there was an increase in cancer risk for those cancers that had time as important factor (breast, colorectal and urology).

### Inference and prediction: incidence and prevalence risk rates

Seventeen out of 23 factors were risk factors for one or more cancer types, with comorbidities such as ‘chronic kidney disease’ and ‘COVID19’ being statistically significant factors for six out of nine cancer types – associated to higher risk of diagnoses for five and four cancer types, respectively (Table 3). ‘Age above 80’, ‘depression’ and ‘congestive heart failure’ were risk factors and protective factors for the same number of cancer types; while being ‘fit’ or ‘inemployed, self employed with one parent working’ or ‘northern Irish’ were mostly protective factors.

**Table 3.** Summary of the number of associations between selected variables and cancer types. It is important to emphasise that the term ‘Protective factor’ does not have a biological or medical meaning, but it is an epidemiological term to represent negative association between the factor and the likelihood to be diagnosed with cancer. *The name of this variable is the same as the one reported in the 2011 Census for England and Wales.

| **Factor** | **Number of cancer types** | **Risk factor** | **Protective factor** |
| --- | --- | --- | --- |
| Chronic Kidney disease | 6 | 5 | 1 |
| COVID19 | 6 | 4 | 2 |
| Smoker | 5 | 3 | 2 |
| Age above 80 | 4 | 2 | 2 |
| Depression | 4 | 2 | 2 |
| Diabetes | 4 | 3 | 1 |
| White and Black Caribbean | 4 | 4 | 0 |
| Ex Smoker | 3 | 2 | 1 |
| Northern Irish (area) | 3 | 1 | 2 |
| Any other Black background | 3 | 3 | 0 |
| Other White background (area) | 3 | 1 | 2 |
| Time | 3 | 3 | 0 |
| Age 60 to 80 | 2 | 2 | 0 |
| British | 2 | 2 | 0 |
| Congestive Heart Failure | 2 | 1 | 1 |
| Coronary Heart Disease | 2 | 2 | 0 |
| Frailty (number of fit) | 2 | 0 | 2 |
| Inemployed, self employed (including fulltime students) one parent working (area)* | 2 | 0 | 2 |
| Scottish (area) | 2 | 2 | 0 |
| Hypertension | 1 | 1 | 0 |
| Pakistani | 1 | 1 | 0 |
| Male Pakistani (area) | 1 | 1 | 0 |
| Female age 25-49 mixed white and Asian (area) | 1 | 1 | 0 |

The odds ratios and relative credible intervals of the statistically significant variables for each cancer type are provided in Supplementary file 7. Increasing cancer incidence over time was found for the breast, colorectal and urology cancer types, with breast cancer types increasing faster than the other two cancer types. Being a current smoker or ex-smoker is usually associated to an increased risk of cancer diagnosis. However, ‘smoking’ was found protective for upper GI and gynaecology cancer types. Apart from breast cancer, all the other cancer types were associated to one or more variables belonging to the ethnicity categorisation. Fifteen times they appeared as a risk factor and only four times as protective. Large risks were found for ‘female age 25-49 mixed white and Asian’ for lung cancer type and for ‘any other black background’ for skin cancer type. ‘White and black Caribbean’ and ‘any other black background’ were the most common risk factors appearing statistically significant for four and three cancer types, respectively (Table 3).

Ethnicity and comorbidities may act as proxies for other socio-economic conditions. By employing a Poisson generalised linear model for any selected important factor (listed in Table 3) as the outcome and each one of the remaining 787 variables considered in this study as a predictor, we found that the selected variables were mostly associated to higher values of ecological variables such as Small Area Mental Health Index (meaning worst mental health conditions), unemployment, criminality (shoplifting, criminal damage) and lone parenting (Supplementary File 8). The Small Area Mental Health Index takes into consideration depression rates, incapacity benefit and employment support allowance for mental illness, prescription of antidepressant, and mental health related hospital attendances. In the mediation analysis, the risk factors of only two cancer types mediated other factors. For breast cancer the effect of diabetes was weakly influenced by ‘age over 60’ (coefficient of the average causal mediation effects for the variable 0.23, *p-value* = 0.04) and also by the ‘Scottish ethnicity’ (coefficient −0.15, *p-value* = 0.04); for urology cancer, the ‘COVID19’ variable was weakly influenced by ‘never smoker’ (coefficient −0.09, *p-value* = 0.02) and ‘Scottish ethnicity’ (coefficient 0.32, *p-value* = 0.02).

In Supplementary File 9, the maps for standardised risk prevalence of a cancer type in the Morecambe Bay ex CCG extended study area and more specifically for three regions (Morecambe and Lancaster, Barrow in Furness, and Kendal) are presented alongside their uncertainties. In these maps, areas indicated with low uncertainty risk are those for which the prediction is more accurate. LSOAs without colour are those where cancers were not recorded during 2017-2022. For all the cancer types, the North Cumbria and Forest of Bowland areas were generally at low risk, apart from Bentham, although associated with large uncertainty. Locally, Morecambe and Barrow-in-Furness suffer the highest risk prevalence for cancers, especially in areas such as Vickerstown (Barrow) and Torrisholme (Morecambe), which had all cancer types in the top ten of the areas ranked risks.

When adjusted by age (Supplementary File 10), one LSOA (located in Westgate/White Lund in Morecambe) was in the top ten LSOAs for the risk of six of the cancer types in under 50 years old. For the over 50 years old, two LSOA areas were consistently in the top ten LSOAs for the risk of nine cancer types: Bowerham south and Freehold West in Lancaster.

Figure 2 presents the cumulative risk for the number of cancer types, with the maximum found in urban areas. Areas such as the Forest of Bowland, Yorkshire borders and Windermere West exibit the presence of most cancer types, but with low cumulative risk (less or equal to 10%, or in lay terms one in ten people was likely to get one of the nine cancer types during the 2017-2022 period). With the same number of cancer types, some areas in Morecambe, Lancaster and Barrow-in-Furness had up to six times more cumulative risk than Windemere West, Burton in Lonsdale or Quernmore.

**Figure 2.**
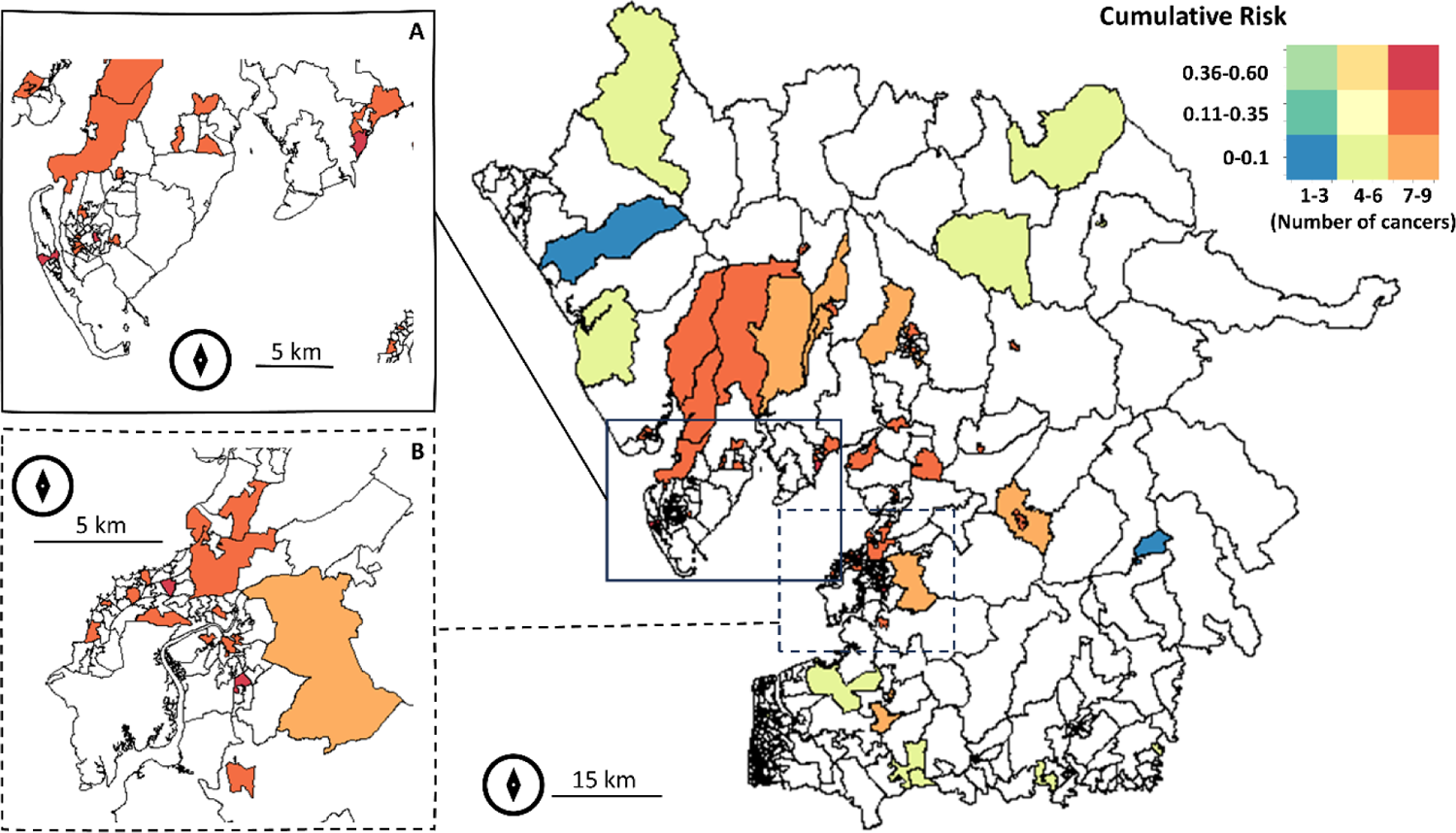
Cumulative risk for number of cancer types for the Morecambe Bay ex CCG extended area (main) zoomed to (A) Barrow-in-Furness and (B) Morecambe and Lancaster. White polygons show no cases of cancer types during 2017-2022. Map created in R (sf and sp packages).

### Inference and prediction: co-regionalisation

The correlation structure between cancer types was quantified implicitly. Moreover the correspondence between the correlation and co-regionalisation of any pair of cancer types’ spatial patterns and residuals were identified.

Lung and skin cancer types were negatively correlated (spatial patterns not coincident) between themselves and between each of them and the rest of the cancer types (Figure 3). Apart from the negative associations with the lung and skin cancer types, breast cancer had no significant associations with any other cancer types. A positive association was found between the colorectal, haematology, upper GI and urology cancer types; and between colorectal, haematology, upper GI and head and neck cancer types.

**Figure 3.**
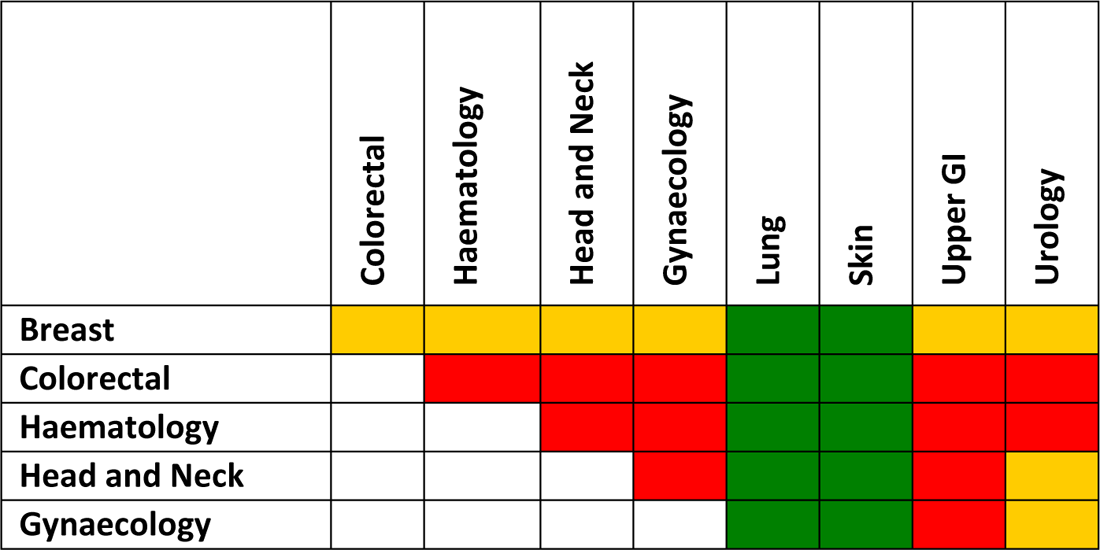
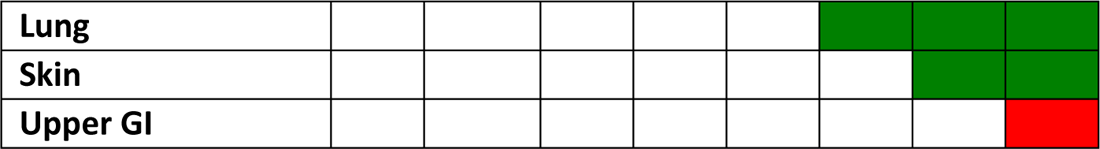
Median correlation between posterior samples of cancer types in the Morecambe Bay ex CCG extended area. Orange = no correlation; green = negative correlation (areas with high incidence in one cancer type have low incidence in the other cancer type); and red = positive correlation (both cancer types have high incidence in the same area). The correlation threshold is arbitrary: negative correlation is lower than −0.3; positive correlation is above 0.3 and no correlation (or weak correlation) is between −0.3 and 0.3.

Spatially, most of the associations were homogeneous over the region, but not all of them. For example, between the colorectal, haematology, head and neck, gynaecology, upper GI and urology cancer types: some areas exhibited no association (e.g., Windemere), while being contiguous with areas with positive association (e.g., Windemere West). The positive associations between these cancer types were also found in both rural and urban areas (see co-regionalisation maps in Supplementary File 11). Correlations between upper GI and urology, upper GI and head and neck, upper GI and gynaecology, were found to be generally positive and homogeneous over the region.

The common spatial range for all cancer types was estimated by the joint model to be 15.2 km (95% CI, 14.6–15.8). This shows a relatively moderate range of influence of the shared spatial component. For example, it allows for statistical dependence (or influence) between cancer types in Lancaster and Morecambe with Arnside and Silverdale, but not between Lancaster and Morecambe and Barrow-in-Furness.

### Clusters

As described in the methods section, the first two principal components from the PCA of the posteriors for the nine cancer types were employed in conjunction with a centroid hierarchical clustering to cluster the LSOAs in the Morecambe Bay ex CCG extended area. The first two principal components explained 91% of the variance (64% PC1 and 27% PC2). The centroid hierarchical clustering identified five clusters (Figure 4):

- Cluster 1 (rural). Large rural LSOAs with large, but generally younger population. Low level of comorbidities and smoking. General cancer risk low.
- Cluster 2 (Windemere East to Coniston). Similar to Cluster 1, but with an older population and larger proportion of British ethnic group individuals compared to other clusters. High number of cancers with low-to-moderate risk.
- Cluster 3 (Morecambe and Lancaster). This urban cluster is characterised by a high level of unemployment, and a high proportion of chronic diseases and mental health conditions. High number of cancers with moderate-to-high risk.
- Cluster 4 (Dalton-in-Furness). This isolated cluster presents a high level of comorbidities in a generally younger population than the study area as a whole. High number of cancers with moderate risk.
- Cluster 5 (Barrow-in-Furness). The worst cluster in terms of cancer risk prevalence. The local population is affected by high level of comorbidities, lower population density and a high level of smoking. High number of cancers with moderate-to-high risk.

**Figure 4.**
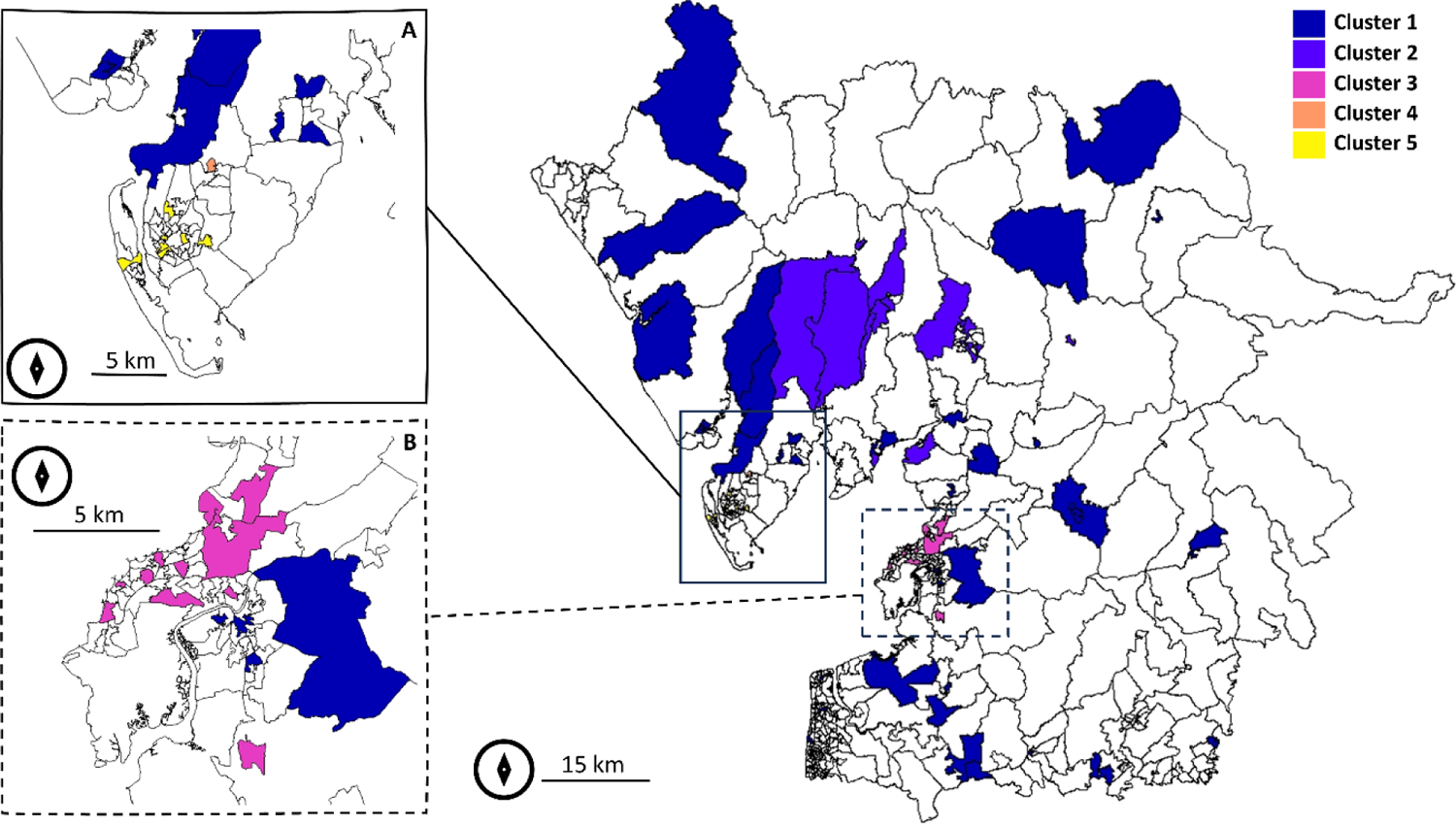
Cancer type clustering by LSOA for the Morecambe Bay ex CCG extended area (main) zoomed to (A) Barrow-in-Furness and (B) Morecambe and Lancaster. White polygons were not clustered due to the absence of cases of cancer types during 2017-2022. Map created in R (sf and sp packages).

### Validation

Model validation results indicate a precise joint model with an error of maximum of 2.5 cases on average per time period (Upper GI, mean squared error) (Table 4). Similar results were obtained when leaving out 10% of the data (root mean squared error and mean squared deviation ratio). The best fitting models based on the DIC and WAIC statistics were for the cancer types highly correlated to each other: gynaecology, haematology, head and neck and upper GI as shown elsewhere (Held et al., 2005). The DIC and WAIC results were consistent, although for the latter upper GI was the best fitting model (instead of haematology for DIC).

**Table 4.** Joint model validation statistics for all data and cross-validated data (indicated with *)

| Cancer type | Mean error | Mean squared error | WAIC | DIC | Root Mean squared error* | Mean Squared Deviation Ratio* |
| --- | --- | --- | --- | --- | --- | --- |
| Breast | -1 | 1 | -116.19 | 224.96 | 1 | 0.22 |
| Colorectal | -0.8 | 0.93 | -98.17 | 79.36 | 0.96 | 0.55 |
| Gynaecology | -0.8 | 1.03 | -93.71 | 69.92 | 1.01 | 2.02 |
| Haematology | -0.86 | 0.86 | -97.34 | 68.17 | 0.93 | 1.34 |
| Head and Neck | -0.58 | 0.89 | -91.14 | 68.21 | 0.94 | 0.79 |
| Lung | -0.66 | 0.8 | -98.45 | 128.61 | 0.89 | 0.31 |
| Skin | -1.4 | 2.33 | -103.44 | 156.94 | 1.52 | 0.32 |
| Upper GI | -0.46 | 2.46 | -88.62 | 69.77 | 1.57 | 4.92 |
| Urology | -1.05 | 1.48 | -99.56 | 77.64 | 1.21 | 3.3 |

## Discussion

This is the first population-based study to investigate spatial patterns in multiple cancers in the Morecambe Bay area. It is possible to state with certainty that the nine cancer types considered in this research are co-regionalised in three quarters of the Morecambe Bay ex CCG extended area for the LSOAs where cancers were found between 2017 and 2022. 19% of LSOAs had only one cancer type during the study period. In addition, the rates for the breast, colorectal and urology cancer types rates were above England and North West rates for the 2017 and 2016-2018 periods respectively. The major difference was in urology cancer type with 14 new cases per 100,000 people per year more than in the North West and 24 new cases more than in England. In contrast, the rates for the lung, skin and upper GI cancer types were generally below the North West and England rates. Between these, skin cancer had 25 new cases per 100,000 people per year less than the rest of the North West.

Estimated counts in censored areas can be considered accurate due to the good agreement between the real data and model predictions (error at individual cancer type up to 10%). The incidence rates were driven by the over 50 years old population since the incidence rates for the general population and those for the over 50s had the same ranking of cancer types. In the uder 50 year old population, skin cancer type remains the most common cancer followed by breast cancer type instead of urology, the second cancer for incidence in the over 50s. Also, in the under 50 group the least common cancer type was upper GI instead of head and neck cancer whose incidence was the lowest in the over 50 group.

Apart from co-occurring, cancers tend to associate geographically with positive associations (co-occurring with high risk prevalence) or negative associations (where one cancer has high risk prevalence, the other has low risk prevalence) (Chidumwa et al., 2021). The joint modelling implemented using a Bayesian hierarchical model enabled the analyses and calculation of the geographical correlations between any two cancer types since estimates were obtained from a common variance/covariance matrix updated during the MCMC computation, while adjusting for individual and ecological factors associated with cancer risk (comorbidities, ethnicity, frailty, age, socio-economic and population density). The proposed model allowed for shared local space dependence, in addition to cancer specific components. Similar frameworks have been proposed elsewhere for cancers and other diseases (Manda et al., 2009), although none of them are censored cancer-specific. This allowed to investigate spatial inequalities in cancer, especially between urban and rural areas.

A positive spatial correlation can support the theory of shared environmental aetiology (Manda et al., 2009), although differences in risk factors selected for each individual cancer type suggest that this is not the only relevant component (DeChello et al., 2006). Upper GI and gynaecology, upper GI and head and neck and upper GI and urology correlations were the most widespread positive associations. This is likely attributable to the fact that these cancers share risk factors such as alcohol consumption, ethnicity, obesity and smoking, but also infectious diseases^6^. This reinforces the ‘integrating intervention’ paradigm, where interventions are designed to reduce and eliminate multiple cancers instead of tackling individual cancers especially when risk factors are identified (Subramanian et al., 2022, Villalobos and Chambers, 2023). The latter was a designed-in goal of this research, which aimed not only to select the risk factors for each cancer type, but also to find areas with correlated residuals (or in other words residual patterns of co-occurrence)(Pollock et al., 2014) which are indicative of the presence of factors or biological processes associated to both cancers, but not considered in the model (Jahan et al., 2020).

The use of a large number of candidate factors (787), and the selection of 22 of them, indicates that individual-level characteristics and area-level socio-economic characteristics were far from providing a comprehensive explanation for the observed spatial heterogeneities in cancer types (Ribeiro et al., 2015).

Chronic Kidney Disease and COVID19 were the most important factors (speculatively, COVID19 may have increased the cancer diagnosis rate once people were hospitalised), both being associated to six of the nine cancer types, followed by ‘current smoker’ associated five of the nine cancer types included in the joint modelling. Overall, comorbidities (seven risk factors) and ethnicities (eight risk factors) were the most important factors associated to cancer type counts.

In the literature, age is a common risk factor for cancers (Liao et al., 2023). For cancer types where age was not found to be a statistically significant factor, frailty or comorbidities may have proxied the age effect. Apart from breast cancer type, at least one ethnic group was always associated to the rest of the cancer types. In terms of ethnicity, the present findings are in agreement with other research for the head and neck (Moles et al., 2008), and urology cancer types (DeChello et al., 2006), although for other cancer types the present findings are novel. Smoking has been confirmed one of the major risk factors (Edwards et al., 2006), and consistent with other research, for lung cancer (Tomintz et al., 2016) and head and neck cancer (Taib et al., 2018). Interestingly, smoking was found to be protective for the upper GI and gynaecology cancer types, both explained by frailty (that appeared only in these two cancers) which may have masked the effect of smoking (Taib et al., 2018). The presence of comorbidities as risk factor for most of the cancer types (but not for the skin cancer), has been described for different cancers elsewhere as potential diagnostic driver or because of the interlink between comorbidities, socio-economic status and behaviour (Grose et al., 2014).

For example, diabetes may represent the effects of obesity (a variable not available in this study), which itself is linked to several comorbidities and cancers (Ellaway et al., 2016).

Only one socio-economic variable was selected among the many considered in this study (inemployed, self employed with one parent working), but it was shown that most of the selected variables were correlated with other socio-economic and health factors (Chaturvedi, 2001).

Deprivation was found to be not significant, in contrast with the large amount of literature linking deprivation or socio-economic status with cancer diagnosis and survival rates (Liao et al., 2023, Bithell et al., 2013, Chambers et al., 2020, Shack et al., 2008, Edwards et al., 2006, Rogers et al., 2019, Rafiq et al., 2019, Phillips et al., 2019, Taib et al., 2018).

Spatial heterogeneity has been observed in other studies where similar risk factors were considered. For example, age, socioeconomic deprivation, ethnicity, and geographical region were all significantly associated with an incident diagnosis of liver cancer (from primary care data) at the population level (Liao et al., 2023, Burton et al., 2022); for certain leukaemias and lymphomas in the North West (McNally et al., 2003); head and neck cancer in UK (Taib et al., 2018); and for joint modelling of lung, bowel and melanoma cancer (Jahan et al., 2020).

Spatial heterogeneity was confirmed by the presence of geographic clusters for cancers risk. Although the clustering analysis was affected by a level of uncertainty due to the use of only two principal components, a clear pattern emerged: Barrow-in-Furness (identified cluster 5) cancer risk was three times more than the surrounding rural areas (cluster 1) and higher than Morecambe and Lancaster urban centres (cluster 3).

Policymakers may potentially use our spatial results for the purposes of resource allocation and education in holistic public health interventions and programs targeted to reduce the burden from geographically correlated and co-regionalised cancer types (Chidumwa et al., 2021) by taking action on common risk factors, deploying research for hidden common risk factors or prioritising cancers that are on the rise (as we found for the breast, colorectal and urology). However, some cancer types are not associated to others or negatively associated (breast, skin, lung) meaning that cancer-type specific interventions are still likely to have greater impact for certain cancer types.

Most of the factors associated to cancer risk by type were grouped by ethnicity and comorbidities, which may help health commissioners and policymakers to consider health equality in different geographical regions and reduce health inequities in ethnic minority groups (Liao et al., 2023), but also to potentially include comorbidities in screening programmes (Vrinzen et al., 2023). This complies with the goals set by the NHS Long Term Plan^7^ which aims by 2028 to reach a target of 55,000 people each year that will survive for five years or more following their cancer diagnosis. The plan set up the actions to improve and extend screening (including lung screening pilots) and reach ethnic minority backgrounds. However, focusing only on individual factors may not be beneficial if the healthcare system is not improved as well (Exarchakou et al., 2018) and, in this sense, our maps showing the cumulative risk versus the number of cancer types may inform on the necessity for expertise and facilities in response to the complexity of local cancer type dynamics.

Identifying regional and subregional inequalities is essential for the distribution of resources (Tomintz et al., 2016). Some cancers were localised. Therefore, while the rates may be generally low, some communities may experience larger than expected risks. This requires further investigation into the causes, and possible interventions. In the long-term, reducing the socioeconomic variation in incidence should have a substantial impact on the burden of cancer (Shack et al., 2008) and fulfil the goal of universal health coverage by targeting the most vulnerable members of society first (Burki, 2018). Mobile services such as ‘stop smoking services’, ‘drink aware’ and ‘low dose CT scan’ (the latter used in US) could be successful in reaching hard-to-reach populations and improving their health (Vohra et al., 2016). These interventions will benefit non-cancer diseases too (such as diabetes and heart disease) due to the common risk factors shared between cancers and comorbidities.

Finally, it is also essential to enhance health literacy and reduce patients’ misconceptions about cancer screening in the high-risk areas (Lal et al., 2020, Wardle et al., 2004).

### Limitations

The following limitations affected the present study:

- Each cancer type is effectively a collection of different cancers (Supplementary File 2) which may present different detection and under-detection rates (e.g., non-melanoma skin cancer versus melanoma skin cancer). For example, prostate cancer has the greatest incidence of urological cancers but urothelial cancer may present the largest mortality threat.
- Border effects: some patients within the extended portion of the Morecambe Bay ex CCG may have chosen to go to a different CCG if they lived closer to it. This could have created a potential bias in the most peripheral areas of the study region.
- Identification of important factors was carried out for all cancers together. While this promotes shared risk factors, it may reduce the significance of risk factors for less common cancers.
- Some factors may be affected by reverse causation (e.g., depression and cancer) although this analysis focused on new diagnosis instead of cancer prevalence, which should have reduced this risk.
- Some maps show high levels of uncertainty (standard errors), often based on very small numbers and, therefore, interpretation should be done cautiously. As described by (Goovaerts, 2010), mapping and interpreting cancer incidence rates faces three major hurdles: (1) the presence of unreliable rates that occur for sparsely populated areas and/or rare cancers, (2) the visual bias caused by the aggregation of health data within administrative units of widely different sizes and shapes (Roberts et al., 2003), and (3) the mismatch of spatial supports for cancer rates and explanatory variables that prevent their direct use in correlation analysis (Carsin et al., 2011).
- The study did not consider rare cancers due to the total censoring for these cancers, or cancer stage at diagnosis, which was necessary to remove biases and support correct inference(Stromberg et al., 2020).
- Postcode sector of patient residence is related to the last known address and, therefore, local factors may or may not be involved in the development of the cancer since cancers have a complex aetiology and long latency (Wah et al., 2020, Downing et al., 2008). Therefore, the findings should be treated as hypotheses-generating (DeChello et al., 2006).

## Conclusions

Striking geographical, socioeconomic, behavioural and demographic variations were observed in Morecambe Bay ex CCG extended area in relation to nine cancer types during the 2017-2022 period. The joint model provided a richer perspective on spatial variation in disease risk than a standard disease mapping analysis by providing the different geographic levels of association between cancers, and between cancers and explanatory factors. The results illustrated how joint mapping can help to better understand the cancer burden for different cancers for an area of interest, and can help inform etiologic debate about specific causes of disease, generate new hypotheses, or aid policy formulation and evaluation or resource allocation (Best and Hansell, 2009), and future research. As such, this study calls for demographic and geographic-specific understanding to better control disease within at-risk communities.

## Supporting information

Supplementary Files_From_01_to_11

## Data Availability

To guarantee the confidentiality of personal and health information of patients, only the named authors have had full access to the data during the study, in accordance with the relevant licence agreements. Data is not publicly available but request under ethical approval can be made to Kelly Heys.

## Acknowledgements

The authors thank the many people who helped make this study possible, that provided useful discussions and support: Faye Bennett, Maria Angela Ferrario, Amy Gadoud, Beccy Harrison, Rebecca Henderson, Fiona Macdonald, Steve Milan, Alicia Rice, Alastair Richards, Helen Stansfield, Hannah Timpson, Angela Waind, Lizzie Wrench and Sam Winder. We also thank the Morecambe Bay population for their kindness and support during this work. We hope that this research is a catalyst to improve their health outcomes.

## Contributors

LS, AK, AB KH and PA secured the funding for this study. LS is the project lead and the guarantor of this study. LS, AK and AB conceived the study. CJ acquired and manipulated the cancer data with input from KH. JAM obtained the ecological open source data, and led the ethical approval. LS conducted the literature review, designed the statistical analysis plan, with methodological input from PA and TK. LS performed statistical analyses and prepared the maps, tables and figures. All authors contributed to the interpretation of the findings. LS drafted the whole manuscript. All authors read and commented on earlier drafts, contributed to revision of the manuscript, approved the final version of the manuscript, and had final responsibility for the decision to submit for publication.

## Software

The joint modelling analysis was performed using a modified version of the MCMC software written by Arnab Hazra available at https://github.com/arnabstatswithR/Arsenic-contamination-mapping.

## Patient and public involvement statement

No patients were involved in forming the research question or selecting the outcome measures, nor were they involved in developing the study design. No patients were asked to advise on the interpretation or writing up of results. Results will be shared through patient charities, regionally and nationally.

## Ethics approval

Project data management and analyses were approved by the Faculty of Health and Medicine Research Ethics Committee at Lancaster University (UK) with reference number FHM-2022-0899-IRAS-2, and by the HRA and Health and Care Research Wales (314994 SL45) under Lancaster University sponsorship.

## Consent for publication

All authors provided consent to publish this manuscript.

## Transparency

The lead author (LS) affirms that the manuscript is an honest, accurate, and transparent account of the study being reported; that no important aspects of the study have been omitted; and that any discrepancies from the study as planned have been explained.

## Footnotes

1 https://www.gov.uk/government/statistics/english-indices-of-deprivation-2019

2 https://www.ons.gov.uk/methodology/geography/ukgeographies/censusgeographies/census2021geographies

3 https://www.ons.gov.uk/methodology/geography/ukgeographies/postalgeography

4 https://bloodcancer.org.uk/news/blood-cancer-facts/#

5 https://www.macmillan.org.uk/about-us/what-we-do/research/cancer-statistics-fact-sheet#

6 https://www.cancerresearchuk.org/about-cancer/type

7 https://www.longtermplan.nhs.uk/areas-of-work/cancer/#:~:text=Our%20NHS%20Long%20Term%20Plan,more%20following%20their%20cancer%20diagnosis

## References

Abdulrahman, G. O. J. 2014. Breast cancer in Wales: time trends and geographical distribution. Gland Surg, 3, 237–42.

Arnold, M., Abnet, C. C., Neale, R. E., Vignat, J., Giovannucci, E. L., Mcglynn, K. A. & Bray, F. 2020. Global Burden of 5 Major Types of Gastrointestinal Cancer. Gastroenterology, 159, 335–349 e15.

Asthana, S. & Gibson, A. 2021. Analysis of Coastal health outcomes. Chief Medical Officer Annual Report, 2021: Health in Coastal Communities. Department of Health and Social Care.

Best, N. & Hansell, A. L. 2009. Geographic variations in risk: adjusting for unmeasured confounders through joint modeling of multiple diseases. Epidemiology, 20, 400–10.

Bithell, J. F., Murphy, M. F., Stiller, C. A., Toumpakari, E., Vincent, T. & Wakeford, R. 2013. Leukaemia in young children in the vicinity of British nuclear power plants: a case-control study. Br J Cancer, 109, 2880–5.

Boyle, P. & Parkin, D. M. 1991. Statistical methods for registries. Cancer registration: principles and methods, 95, 126–158.

Brodbelt, A., Greenberg, D., Winters, T., Williams, M., Vernon, S., Collins, V. P. & National Cancer Information Network Brain Tumour, G. 2015. Glioblastoma in England: 2007-2011. Eur J Cancer, 51, 533–542.

Bunch, K. J., Keegan, T. J., Swanson, J., Vincent, T. J. & Murphy, M. F. 2014a. Residential distance at birth from overhead high-voltage powerlines: childhood cancer risk in Britain 1962-2008. Br J Cancer, 110, 1402–8.

Bunch, K. J., Vincent, T. J., Black, R. J., Pearce, M. S., Mcnally, R. J., Mckinney, P. A., Parker, L., Craft, A. W. & Murphy, M. F. 2014b. Updated investigations of cancer excesses in individuals born or resident in the vicinity of Sellafield and Dounreay. Br J Cancer, 111, 1814–23.

Burki, T. K. 2018. No impact of English national cancer policies on survival. Lancet Oncology, 19, E230–E230.

Burton, A., Balachandrakumar, V. K., Driver, R. J., Tataru, D., Paley, L., Marshall, A., Alexander, G., Rowe, I. A., Partnership, H.-U. B. N., Palmer, D. H. & Cross, T. J. S. 2022. Regional variations in hepatocellular carcinoma incidence, routes to diagnosis, treatment and survival in England. Br J Cancer, 126, 804–814.

Cai, J., Xie, Y., Deng, M., Tang, X., Li, Y. & Shekhar, S. 2020. Significant spatial co-distribution pattern discovery. Computers, Environment and Urban Systems, 84.

Carsin, A. E., Sharp, L. & Comber, H. 2011. Geographical, urban/rural and socioeconomic variations in nonmelanoma skin cancer incidence: a population-based study in Ireland. Br J Dermatol, 164, 822–9.

Chambers, A. C., Dixon, S. W., White, P., Williams, A. C., Thomas, M. G. & Messenger, D. E. 2020. Demographic trends in the incidence of young-onset colorectal cancer: a population-based study. Br J Surg, 107, 595–605.

Chaturvedi, N. 2001. Ethnicity as an epidemiological determinant—crudely racist or crucially important? International Journal of Epidemiology, 30, 925–927 %@ 0300-5771.

Chen, L., Zheng, Y., Yu, K., Chen, S., Wang, W., Gale, R. P., Liu, Z. X. & Liang, Y. 2022. Changing causes of death in persons with haematological cancers 1975-2016. Leukemia, 36, 1850–1860.

Chidumwa, G., Maposa, I., Kowal, P., Micklesfield, L. K. & Ware, L. J. 2021. Bivariate Joint Spatial Modeling to Identify Shared Risk Patterns of Hypertension and Diabetes in South Africa: Evidence from WHO SAGE South Africa Wave 2. Int J Environ Res Public Health, 18.

Conway, D. I., Stockton, D. L., Warnakulasuriya, K. A., Ogden, G. & Macpherson, L. M. 2006. Incidence of oral and oropharyngeal cancer in United Kingdom (1990-1999) -- recent trends and regional variation. Oral Oncol, 42, 586–92.

Dechello, L. M., Gregorio, D. I. & Samociuk, H. 2006. Race-specific geography of prostate cancer incidence. Int J Health Geogr, 5, 59.

Downing, A., Forman, D., Gilthorpe, M. S., Edwards, K. L. & Manda, S. O. 2008. Joint disease mapping using six cancers in the Yorkshire region of England. Int J Health Geogr, 7, 41.

Downing, A., Harrison, W. J., West, R. M., Forman, D. & Gilthorpe, M. S. 2010. Latent class modelling of the association between socioeconomic background and breast cancer survival status at 5 years incorporating stage of disease. J Epidemiol Community Health, 64, 772–6.

Edwards, R., Pless-Mulloli, T., Howel, D., Chadwick, T., Bhopal, R., Harrison, R. & Gribbin, H. 2006. Does living near heavy industry cause lung cancer in women? A case-control study using life grid interviews. Thorax, 61, 1076–82.

Ellaway, A., Lamb, K. E., Ferguson, N. S. & Ogilvie, D. 2016. Associations between access to recreational physical activity facilities and body mass index in Scottish adults. BMC Public Health, 16, 756.

Elliott, P., Shaddick, G., Douglass, M., De Hoogh, K., Briggs, D. J. & Toledano, M. B. 2013. Adult cancers near high-voltage overhead power lines. Epidemiology, 24, 184–90.

Exarchakou, A., Rachet, B., Belot, A., Maringe, C. & Coleman, M. P. 2018. Impact of national cancer policies on cancer survival trends and socioeconomic inequalities in England, 1996-2013: population based study. Bmj-British Medical Journal, 360.

Gelman, A., Hwang, J. & Vehtari, A. 2013. Understanding predictive information criteria for Bayesian models. Statistics and Computing, 24, 997–1016.

Gomez-Rubio, V., Palmi-Perales, F., Lopez-Abente, G., Ramis-Prieto, R. & Fernandez-Navarro, P. 2019. Bayesian joint spatio-temporal analysis of multiple diseases. Sort-Statistics and Operations Research Transactions, 43, 51–74.

Goovaerts, P. 2010. Geostatistical Analysis of County-Level Lung Cancer Mortality Rates in the Southeastern United States. Geogr Anal, 42, 32–52.

Grose, D., Morrison, D. S., Devereux, G., Jones, R., Sharma, D., Selby, C., Docherty, K., Mcintosh, D., Louden, G., Nicolson, M., Mcmillan, D. C., Milroy, R. & Scottish Lung Cancer, F. 2014. Comorbidities in lung cancer: prevalence, severity and links with socioeconomic status and treatment. Postgrad Med J, 90, 305–10.

Held, L., Natario, I., Fenton, S. E., Rue, H. & Becker, N. 2005. Towards joint disease mapping. Statistical Methods in Medical Research, 14, 61–82.

Horgan, J. M. 2019. Probability with R: an introduction with computer science applications, John Wiley & Sons.

Jahan, F., Duncan, E. W., Cramb, S. M., Baade, P. D. & Mengersen, K. L. 2020. Multivariate Bayesian meta-analysis: joint modelling of multiple cancer types using summary statistics. Int J Health Geogr, 19, 42.

Jarup, L., Best, N., Toledano, M. B., Wakefield, J. & Elliott, P. 2002. Geographical epidemiology of prostate cancer in Great Britain. Int J Cancer, 97, 695–9.

Kalbfleisch, J. D. & Prentice, R. L. 2011. The statistical analysis of failure time data, John Wiley & Sons.

Keenan, T. D., Yeates, D. & Goldacre, M. J. 2012. Uveal melanoma in England: trends over time and geographical variation. Br J Ophthalmol, 96, 1415–9.

Knapp, S., Henderson, L., Duncan, T. & Gray, T. 2021. Gynaecological cancers. InnovAiT: Education and inspiration for general practice, 14, 449–457.

Kong, S. & Nan, B. 2016. Semiparametric approach to regression with a covariate subject to a detection limit. Biometrika, 103, 161–174.

Kouame, R. M. A., Edi, A. V. C., Cain, R. J., Weetman, D., Donnelly, M. J. & Sedda, L. 2023. Joint spatial modelling of malaria incidence and vector’s abundance shows heterogeneity in malaria-vector geographical relationships. Journal of Applied Ecology.

Król, A., Ferrer, L., Pignon, J.-P., Proust-Lima, C., Ducreux, M., Bouché, O., Michiels, S. & Rondeau, V. 2016. Joint model for left-censored longitudinal data, recurrent events and terminal event: Predictive abilities of tumor burden for cancer evolution with application to the FFCD 2000–05 trial. Biometrics, 72, 907–916.

Lal, N., Singh, H. K., Majeed, A. & Pawa, N. 2020. The impact of socioeconomic deprivation on the uptake of colorectal cancer screening in London. J Med Screen, 969141320916206.

Leacy, F. P., Floyd, S., Yates, T. A. & White, I. R. 2017. Analyses of Sensitivity to the Missing-at-Random Assumption Using Multiple Imputation With Delta Adjustment: Application to a Tuberculosis/HIV Prevalence Survey With Incomplete HIV-Status Data. Am J Epidemiol, 185, 304–315.

Leung, K.-M., Elashoff, R. M. & Afifi, A. A. 1997. Censoring issues in survival analysis. Annual review of public health, 18, 83–104.

Liao, W., Coupland, C. A. C., Innes, H., Jepsen, P., Matthews, P. C., Campbell, C., De, L. C., Barnes, E. & Hippisley-Cox, J. 2023. Disparities in care and outcomes for primary liver cancer in England during 2008-2018: a cohort study of 8.52 million primary care population using the QResearch database. EClinicalMedicine, 59, 101969.

Liao, W., Jepsen, P., Coupland, C., Innes, H., Matthews, P. C., Campbell, C., Barnes, E., Hippisley-Cox, J. & De, L. C. 2022. Development and validation of personalised risk prediction models for early detection and diagnosis of primary liver cancer among the English primary care population using the QResearch(R) database: research protocol and statistical analysis plan. Diagn Progn Res, 6, 21.

Lyratzopoulos, G., Sheridan, G. F., Michie, H. R., Mcelduff, P. & Hobbiss, J. H. 2004. Absence of socioeconomic variation in survival from colorectal cancer in patients receiving surgical treatment in one health district: cohort study. Colorectal Dis, 6, 512–7.

Manda, S. O., Feltbower, R. G. & Gilthorpe, M. S. 2009. Investigating spatio-temporal similarities in the epidemiology of childhood leukaemia and diabetes. Eur J Epidemiol, 24, 743–52.

Mansouri, D., Mcmillan, D. C., Grant, Y., Crighton, E. M. & Horgan, P. G. 2013. The impact of age, sex and socioeconomic deprivation on outcomes in a colorectal cancer screening programme. PLoS One, 8, e66063.

Mccaffery, K., Wardle, J., Nadel, M. & Atkin, W. 2002. Socioeconomic variation in participation in colorectal cancer screening. J Med Screen, 9, 104–8.

Mcnally, R. J., Alston, R. D., Cairns, D. P., Eden, O. B. & Birch, J. M. 2003. Geographical and ecological analyses of childhood acute leukaemias and lymphomas in north-west England. Br J Haematol, 123, 60–5.

Mcnally, R. J., Wakeford, R., James, P. W., Basta, N. O., Alston, R. D., Pearce, M. S. & Elliott, A. T. 2016. A geographical study of thyroid cancer incidence in north-west England following the Windscale nuclear reactor fire of 1957. J Radiol Prot, 36, 934–952.

Moles, D. R., Fedele, S., Speight, P. M., Porter, S. R. & Dos Santos Silva, I. 2008. Oral and pharyngeal cancer in South Asians and non-South Asians in relation to socioeconomic deprivation in South East England. Br J Cancer, 98, 633–5.

Musah, A., Gibson, J. E., Leonardi-Bee, J., Cave, M. R., Ander, E. L. & Bath-Hextall, F. 2013. Regional variations of basal cell carcinoma incidence in the U.K. using The Health Improvement Network database (2004-10). Br J Dermatol, 169, 1093–9.

Nogueira, M. C., Fayer, V. A., Correa, C. S. L., Guerra, M. R., Stavola, B., Dos-Santos-Silva, I., Bustamante-Teixeira, M. T. & Silva, G. A. E. 2019. Inequities in access to mammographic screening in Brazil. Cad Saude Publica, 35, e00099817.

Phillips, A., Kehoe, S., Singh, K., Elattar, A., Nevin, J., Balega, J., Pounds, R., Elmodir, A., Pascoe, J., Fernando, I. & Sundar, S. 2019. Socioeconomic differences impact overall survival in advanced ovarian cancer (AOC) prior to achievement of standard therapy. Arch Gynecol Obstet, 300, 1261–1270.

Pollock, L. J., Tingley, R., Morris, W. K., Golding, N., O’hara, R. B., Parris, K. M., Vesk, P. A., Mccarthy, M. A. & Mcpherson, J. 2014. Understanding co-occurrence by modelling species simultaneously with a Joint Species Distribution Model (JSDM). Methods in Ecology and Evolution, 5, 397–406.

Qu, T., Li, B., Sally Chan, M. P. & Albarracin, D. 2023. Bias correction for nonignorable missing counts of areal HIV new diagnosis. Stat, 12.

R Core Team 2023. R: A language and environment for statistical computing. Vienna, Austria: R Foundation for Statistical Computing,.

Rachet, B., Woods, L. M., Mitry, E., Riga, M., Cooper, N., Quinn, M. J., Steward, J., Brenner, H., Esteve, J., Sullivan, R. & Coleman, M. P. 2008. Cancer survival in England and Wales at the end of the 20th century. Br J Cancer, 99 Suppl 1, S2–10.

Rafiq, M., Hayward, A., Warren-Gash, C., Denaxas, S., Gonzalez-Izquierdo, A., Lyratzopoulos, G. & Thomas, S. 2019. Socioeconomic deprivation and regional variation in Hodgkin’s lymphoma incidence in the UK: a population-based cohort study of 10 million individuals. BMJ Open, 9, e029228.

Rait, G. & Horsfall, L. 2020. Twenty-year sociodemographic trends in lung cancer in non-smokers: A UK-based cohort study of 3.7 million people. Cancer Epidemiol, 67, 101771.

Ribeiro, A. I., De Pina Mde, F. & Mitchell, R. 2015. Development of a measure of multiple physical environmental deprivation. After United Kingdom and New Zealand, Portugal. Eur J Public Health, 25, 610–7.

Roberts, J., Cramb, S., Baade, P. & Mengersen, K. 2016. Communicating statistical outputs through health maps.

Roberts, R. J., Steward, J. & John, G. 2003. Cement, cancers and clusters: an investigation of a claim of a local excess cancer risk related to a cement works. J Public Health Med, 25, 351–7.

Rogers, S. N., Staunton, A., Girach, R., Langton, S. & Lowe, D. 2019. Audit of the two-week pathway for patients with suspected cancer of the head and neck and the influence of socioeconomic status. Br J Oral Maxillofac Surg, 57, 419–424.

Sahoo, I. & Hazra, A. 2021. Contamination mapping in Bangladesh using a multivariate spatial Bayesian model for left-censored data. arXiv preprint arXiv:2106.15730.

Saleh, G. M., Desai, P., Collin, J. R., Ives, A., Jones, T. & Hussain, B. 2017. Incidence of eyelid basal cell carcinoma in England: 2000-2010. Br J Ophthalmol, 101, 209–212.

Sehmer, E. A., Hall, G. J., Greenberg, D. C., O’hara, C., Wallingford, S. C., Wright, K. A. & Green, A. C. 2014. Incidence of glioma in a northwestern region of England, 2006-2010. Neuro Oncol, 16, 971–4.

Shack, L., Jordan, C., Thomson, C. S., Mak, V., Moller, H. & Registries, U. K. A. O. C. 2008. Variation in incidence of breast, lung and cervical cancer and malignant melanoma of skin by socioeconomic group in England. BMC Cancer, 8, 271.

Shrestha, S., Bauer, C. X. C., Hendricks, B. & Stopka, T. J. 2020. Spatial epidemiology: An empirical framework for syndemics research. Soc Sci Med, 113352.

Smith, A., Roman, E., Simpson, J., Ansell, P., Fear, N. T. & Eden, T. 2006. Childhood leukaemia and socioeconomic status: fact or artefact? A report from the United Kingdom childhood cancer study (UKCCS). Int J Epidemiol, 35, 1504–13.

Stark, J. M., Black, R. J. & Brewster, D. H. 2007. Risk of leukaemia among children living near the Solway coast of Dumfries and Galloway Health Board area, Scotland, 1975-2002. Occup Environ Med, 64, 66–8.

Stromberg, U., Parkes, B. L., Holmen, A., Peterson, S., Holmberg, E., Baigi, A. & Piel, F. B. 2020. Disease mapping of early- and late-stage cancer to monitor inequalities in early detection: a study of cutaneous malignant melanoma. Eur J Epidemiol, 35, 537–547.

Subramanian, S., Tangka, F. K. L., Hoover, S. & Degroff, A. 2022. Integrated interventions and supporting activities to increase uptake of multiple cancer screenings: conceptual framework, determinants of implementation success, measurement challenges, and research priorities. Implement Sci Commun, 3, 105.

Taib, B. G., Oakley, J., Dailey, Y., Hodge, I., Wright, P., Du Plessis, R., Rylands, J., Taylor-Robinson, D., Povall, S., Schache, A., Shaw, R., Dingle, A. & Jones, T. M. 2018. Socioeconomic deprivation and the burden of head and neck cancer-Regional variations of incidence and mortality in Merseyside and Cheshire, North West, England. Clin Otolaryngol, 43, 846–853.

Tingley, D., Yamamoto, T., Hirose, K., Keele, L. & Imai, K. 2014. Mediation: R package for causal mediation analysis. R package version 4.5.0.

Tomintz, M., Kosar, B. & Clarke, G. 2016. smokeSALUD: exploring the effect of demographic change on the smoking prevalence at municipality level in Austria. Int J Health Geogr, 15, 36.

Vichi, M., Cavicchia, C. & Groenen, P. J. F. 2022. Hierarchical Means Clustering. Journal of Classification, 39, 553–577.

Villalobos, A. & Chambers, D. A. 2023. Advancing the science of integrating multiple interventions by blending and bundling. JNCI Cancer Spectr, 7.

Vohra, J., Marmot, M. G., Bauld, L. & Hiatt, R. A. 2016. Socioeconomic position in childhood and cancer in adulthood: a rapid-review. J Epidemiol Community Health, 70, 629–34.

Vrinzen, C. E. J., Delfgou, L., Stadhouders, N., Hermens, R., Merkx, M. A. W., Bloemendal, H. J. & Jeurissen, P. P. T. 2023. A Systematic Review and Multilevel Regression Analysis Reveals the Comorbidity Prevalence in Cancer. Cancer Res, 83, 1147–1157.

Wah, W., Ahern, S. & Earnest, A. 2020. A systematic review of Bayesian spatial-temporal models on cancer incidence and mortality. Int J Public Health, 65, 673–682.

Wardle, J., Mccaffery, K., Nadel, M. & Atkin, W. 2004. Socioeconomic differences in cancer screening participation: comparing cognitive and psychosocial explanations. Soc Sci Med, 59, 249–61.

Wheeler, B. W., Kothencz, G. & Pollard, A. S. 2013. Geography of non-melanoma skin cancer and ecological associations with environmental risk factors in England. Br J Cancer, 109, 235–41.

Wong, G. Y. C. & Yu, Q. 1999. Generalized MLE of a Joint Distribution Function with Multivariate Interval-Censored Data. Journal of Multivariate Analysis, 69, 155–166.

