## Supplementary Files_From_01_to_11 for "Syndemic Geographic Patterns of Cancer Types in a Health Deprived Area of England: a new Paradigm for Public Health Cancer Interventions?"

Supplementary Table S1. Age distribution in Morecambe Bay ex CCG

| **Age group** | **Male and Female (%)** | **Male (%)** | **Female ( %)** |
|---|---|---|---|
| Under 6 | 5.62 | 2.85 | 2.77 |
| 6-10 | 5.25 | 2.71 | 2.54 |
| 11-15 | 5.36 | 2.81 | 2.55 |
| 16-20 | 6.32 | 3.31 | 3.01 |
| 21-25 | 6.79 | 3.68 | 3.11 |
| 26-30 | 6.06 | 3.18 | 2.88 |
| 31-35 | 5.07 | 2.45 | 2.62 |
| 36-40 | 5.16 | 2.50 | 2.66 |
| 41-45 | 4.97 | 2.40 | 2.58 |
| 46-50 | 6.23 | 3.00 | 3.23 |
| 51-55 | 7.26 | 3.53 | 3.73 |
| 56-60 | 7.28 | 3.53 | 3.74 |
| 61-65 | 6.50 | 3.16 | 3.34 |
| 66-70 | 5.99 | 2.90 | 3.09 |
| 71-75 | 6.25 | 3.09 | 3.16 |

| | | | |
|---|---|---|---|
| 76-80 | 4.30 | 2.06 | 2.25 |
| 81-85 | 3.02 | 1.34 | 1.68 |
| 86 and over | 2.57 | 0.92 | 1.65 |
| ***All Ages (total count)*** | ***334,287*** | ***165,161*** | ***169,126*** |

| ICD10 Code | ICD10 Group | ICD10 Code Description | TumourSite |
|---|---|---|---|
| C50 | C50 | Malignant neoplasm of breast | Breast |
| C500 | C50 | Malignant neoplasm of nipple and areola | Breast |
| C501 | C50 | Malignant neoplasm of central portion of breast | Breast |
| C502 | C50 | Malignant neoplasm of upper-inner quadrant of breast | Breast |
| C503 | C50 | Malignant neoplasm of lower-inner quadrant of breast | Breast |
| C504 | C50 | Malignant neoplasm of upper-outer quadrant of breast | Breast |
| C505 | C50 | Malignant neoplasm of lower-outer quadrant of breast | Breast |
| C506 | C50 | Malignant neoplasm of axillary tail of breast | Breast |
| C508 | C50 | Malignant neoplasm, overlapping lesion of breast | Breast |
| C509 | C50 | Malignant neoplasm of breast, unspecified | Breast |
| D05 | D05 | Carcinoma in situ of breast | Breast |
| D050 | D05 | Lobular carcinoma in situ | Breast |
| D051 | D05 | Intraductal carcinoma in situ | Breast |
| D057 | D05 | Other carcinoma in situ of breast | Breast |
| D059 | D05 | Carcinoma in situ of breast, unspecified | Breast |
| C17 | C17 | Malignant neoplasm of small intestine | Colorectal |
| C170 | C17 | Malignant neoplasm of small intestine, duodenum | Colorectal |
| C171 | C17 | Malignant neoplasm of small intestine, jejunum | Colorectal |
| C172 | C17 | Malignant neoplasm of small intestine, ileum | Colorectal |
| C173 | C17 | Malignant neoplasm of small intestine, Meckel's diverticulum | Colorectal |
| C178 | C17 | Malignant neoplasm overlapping lesion of small intestine | Colorectal |
| C179 | C17 | Malignant neoplasm of small intestine, unspecified | Colorectal |
| C18 | C18 | Malignant neoplasm of colon | Colorectal |
| C180 | C18 | Malignant neoplasm of caecum | Colorectal |
| C181 | C18 | Malignant neoplasm of appendix | Colorectal |
| C182 | C18 | Malignant neoplasm of ascending colon | Colorectal |
| C183 | C18 | Malignant neoplasm of hepatic flexure | Colorectal |
| C184 | C18 | Malignant neoplasm of transverse colon | Colorectal |
| C185 | C18 | Malignant neoplasm of splenic flexure | Colorectal |
| C186 | C18 | Malignant neoplasm of descending colon | Colorectal |
| C187 | C18 | Malignant neoplasm of sigmoid colon | Colorectal |
| C188 | C18 | Malignant neoplasm overlapping lesion of colon | Colorectal |

| | | | |
|---|---|---|---|
| C189 | C18 | Malignant neoplasm of colon, unspecified | Colorectal |
| C19X | C19 | Malignant neoplasm of rectosigmoid junction | Colorectal |
| C20X | C20 | Malignant neoplasm of rectum | Colorectal |
| C21 | C21 | Malignant neoplasm of anus and anal canal | Colorectal |
| C210 | C21 | Malignant neoplasm of anus, unspecified | Colorectal |
| C211 | C21 | Malignant neoplasm of anal canal | Colorectal |
| C212 | C21 | Malignant neoplasm of cloacogenic zone | Colorectal |
| C218 | C21 | Malig neo, overlapping lesion of rectum, anus and anal canal | Colorectal |
| C26 | C26 | Malignant neoplasm of other and ill-defined digestive organs | Colorectal |
| C260 | C26 | Malignant neoplasm of intestinal tract, part unsp | Colorectal |
| C261 | C26 | Malignant neoplasm of spleen | Colorectal |
| C268 | C26 | Malignant neoplasm, overlapping lesion of digestive system | Colorectal |
| C269 | C26 | Malignant neoplasm of ill-def sites within digestive system | Colorectal |
| C784 | C78 | Secondary malignant neoplasm of small intestine | Colorectal |
| C785 | C78 | Secondary malignant neoplasm of large intest & rectum | Colorectal |
| C788 | C78 | Secondary malignant neoplasm of other & unsp digestive orgs | Colorectal |
| C51 | C51 | Malignant neoplasm of vulva | Gynaecology |
| C510 | C51 | Malignant neoplasm of labium majus | Gynaecology |
| C511 | C51 | Malignant neoplasm of labium minus | Gynaecology |
| C512 | C51 | Malignant neoplasm of clitoris | Gynaecology |
| C518 | C51 | Malignant neoplasm of overlapping lesion of vulva | Gynaecology |
| C519 | C51 | Malignant neoplasm of vulva, unspecified | Gynaecology |
| C52X | C52 | Malignant neoplasm of vagina | Gynaecology |
| C53 | C53 | Malignant neoplasm of cervix uteri | Gynaecology |
| C530 | C53 | Malignant neoplasm of endocervix | Gynaecology |
| C531 | C53 | Malignant neoplasm of exocervix | Gynaecology |
| C538 | C53 | Malignant neoplasm, overlapping lesion of cervix uteri | Gynaecology |
| C539 | C53 | Malignant neoplasm of cervix uteri, unsp | Gynaecology |
| C54 | C54 | Malignant neoplasm of corpus uteri | Gynaecology |
| C540 | C54 | Malignant neoplasm of isthmus uteri | Gynaecology |
| C541 | C54 | Malignant neoplasm of endometrium | Gynaecology |
| C542 | C54 | Malignant neoplasm of myometrium | Gynaecology |
| C543 | C54 | Malignant neoplasm of fundus uteri | Gynaecology |

| | | | |
|---|---|---|---|
| C548 | C54 | Malignant neoplasm overlapping lesion of corpus uteri | Gynaecology |
| C549 | C54 | Malignant neoplasm of corpus uteri, unsp | Gynaecology |
| C55X | C55 | Malignant neoplasm of uterus, part unspecified | Gynaecology |
| C56X | C56 | Malignant neoplasm of ovary | Gynaecology |
| C57 | C57 | Malignant neoplasm of other and unspec female genital orgs | Gynaecology |
| C570 | C57 | Malignant neoplasm of fallopian tube | Gynaecology |
| C571 | C57 | Malignant neoplasm of broad ligament | Gynaecology |
| C572 | C57 | Malignant neoplasm of round ligament | Gynaecology |
| C573 | C57 | Malignant neoplasm of parametrium | Gynaecology |
| C574 | C57 | Malignant neoplasm of uterine adnexa, unsp | Gynaecology |
| C577 | C57 | Malignant neoplasm of other specified female genital organs | Gynaecology |
| C578 | C57 | Malignant neoplasm, overlapping lesion female genital organs | Gynaecology |
| C579 | C57 | Malignant neoplasm of female genital organ, unspecified | Gynaecology |
| C58X | C58 | Malignant neoplasm of placenta | Gynaecology |
| C796 | C79 | Secondary malignant neoplasm of ovary | Gynaecology |
| C81 | C81 | Hodgkin lymphoma | Haematology |
| C810 | C81 | Nodular lymphocyte predominant Hodgkin lymphoma | Haematology |
| C811 | C81 | Nodular sclerosis classical Hodgkin lymphoma | Haematology |
| C812 | C81 | Mixed cellularity classical Hodgkin lymphoma | Haematology |
| C813 | C81 | Lymphocyte depleted classical Hodgkin lymphoma | Haematology |
| C814 | C81 | Lymphocyte-rich classical Hodgkin lymphoma | Haematology |
| C817 | C81 | Other classical Hodgkin lymphoma | Haematology |
| C819 | C81 | Hodgkin lymphoma, unspecified | Haematology |
| C82 | C82 | Follicular lymphoma | Haematology |
| C820 | C82 | Follicular lymphoma grade I | Haematology |
| C821 | C82 | Follicular lymphoma grade II | Haematology |
| C822 | C82 | Follicular lymphoma grade III, unspecified | Haematology |
| C823 | C82 | Follicular lymphoma grade IIIa | Haematology |
| C824 | C82 | Follicular lymphoma grade IIIb | Haematology |
| C825 | C82 | Diffuse follicle centre lymphoma | Haematology |
| C826 | C82 | Cutaneous follicle centre lymphoma | Haematology |
| C827 | C82 | Other types of follicular lymphoma | Haematology |
| C829 | C82 | Follicular lymphoma, unspecified | Haematology |

| | | | |
|---|---|---|---|
| C83 | C83 | Non-follicular lymphoma | Haematology |
| C830 | C83 | Small cell B-cell lymphoma | Haematology |
| C831 | C83 | Mantle cell lymphoma | Haematology |
| C832 | C83 | Diffuse non-Hodgkin mixed sml & lge cell (diffuse) lymphoma | Haematology |
| C833 | C83 | Diffuse large B-cell lymphoma | Haematology |
| C834 | C83 | Diffuse non-Hodgkin's immunoblastic (diffuse) lymphoma | Haematology |
| C835 | C83 | Lymphoblastic (diffuse) lymphoma | Haematology |
| C836 | C83 | Diffuse non-Hodgkin's lymphoma undifferentiated (diffuse) | Haematology |
| C837 | C83 | Burkitt lymphoma | Haematology |
| C838 | C83 | Other non-follicular lymphoma | Haematology |
| C839 | C83 | Non-follicular (diffuse) lymphoma, unspecified | Haematology |
| C84 | C84 | Mature T/NK-cell lymphomas | Haematology |
| C840 | C84 | Peripheral and cutaneous T-cell lymphomas, mycosis fungoides | Haematology |
| C841 | C84 | SezÚry disease | Haematology |
| C842 | C84 | Peripheral and cutaneous T-cell lymphomas, T-zone lymphoma | Haematology |
| C843 | C84 | Periph & cutan T-cell lymphomas, lymphoepithelioid lymphoma | Haematology |
| C844 | C84 | Peripheral T-cell lymphoma, not elsewhere classified | Haematology |
| C845 | C84 | Other mature T/NK-cell lymphomas | Haematology |
| C846 | C84 | Anaplastic large cell lymphoma, ALK-positive | Haematology |
| C847 | C84 | Anaplastic large cell lymphoma, ALK-negative | Haematology |
| C848 | C84 | Cutaneous T-cell lymphoma, unspecified | Haematology |
| C849 | C84 | Mature T/NK-cell lymphoma, unspecified | Haematology |
| C85 | C85 | Other and unspecified types of non-Hodgkin lymphoma | Haematology |
| C850 | C85 | Oth & unspec types of non-Hodgkin lymphoma, lymphosarcoma | Haematology |
| C851 | C85 | Oth & unsp types non-Hodgkin B-cell lymphoma, unsp | Haematology |
| C852 | C85 | Mediastinal (thymic) large B-cell lymphoma | Haematology |
| C857 | C85 | Other specified types of non-Hodgkin lymphoma | Haematology |
| C859 | C85 | Non-Hodgkin lymphoma, unspecified | Haematology |
| C86 | C86 | Other specified types of T/NK-cell lymphoma | Haematology |
| C860 | C86 | Extranodal NK/T-cell lymphoma, nasal type | Haematology |
| C861 | C86 | Hepatosplenic T-cell lymphoma | Haematology |
| C862 | C86 | Enteropathy-type (intestinal) T-cell lymphoma | Haematology |
| C863 | C86 | Subcutaneous panniculitis-like T-cell lymphoma | Haematology |

| | | | |
|---|---|---|---|
| C864 | C86 | Blastic NK-cell lymphoma | Haematology |
| C865 | C86 | Angioimmunoblastic T-cell lymphoma | Haematology |
| C866 | C86 | Primary cutaneous CD30-positive T-cell proliferations | Haematology |
| C88 | C88 | Malignant immunoproliferative diseases | Haematology |
| C880 | C88 | Waldenstrom macroglobulinaemia | Haematology |
| C881 | C88 | Alpha heavy chain disease | Haematology |
| C882 | C88 | Other heavy chain disease | Haematology |
| C883 | C88 | Malignant immunoproliferative small intestinal disease | Haematology |
| C884 | C88 | Extranodal marginal zone B-cell lymphoma of mucosa-associated lymphoid tissue [MALT-lymphoma] | Haematology |
| C887 | C88 | Other malignant immunoproliferative diseases | Haematology |
| C889 | C88 | Malignant immunoproliferative disease, unspecified | Haematology |
| C90 | C90 | Multiple myeloma and malignant plasma cell neoplasms | Haematology |
| C900 | C90 | Multiple myeloma | Haematology |
| C901 | C90 | Plasma cell leukaemia | Haematology |
| C902 | C90 | Extramedullary plasmacytoma | Haematology |
| C903 | C90 | Solitary plasmacytoma | Haematology |
| C91 | C91 | Lymphoid leukaemia | Haematology |
| C910 | C91 | Acute lymphoblastic leukaemia [ALL] | Haematology |
| C911 | C91 | Chronic lymphocytic leukaemia of B-cell type | Haematology |
| C912 | C91 | Subacute lymphocytic leukaemia | Haematology |
| C913 | C91 | Prolymphocytic leukaemia of B-cell type | Haematology |
| C914 | C91 | Hairy-cell leukaemia | Haematology |
| C915 | C91 | Adult T-cell lymphoma/leukaemia (HTLV-1-associated) | Haematology |
| C916 | C91 | Prolymphocytic leukaemia of T-cell type | Haematology |
| C917 | C91 | Other lymphoid leukaemia | Haematology |
| C918 | C91 | Mature B-cell leukaemia Burkitt-type | Haematology |
| C919 | C91 | Lymphoid leukaemia, unspecified | Haematology |
| C92 | C92 | Myeloid leukaemia | Haematology |
| C920 | C92 | Acute myeloblastic leukaemia [AML] | Haematology |
| C921 | C92 | Chronic myeloid leukaemia [CML], BCR/ABL-positive | Haematology |
| C922 | C92 | Atypical chronic myeloid leukaemia, BCR/ABL-negative | Haematology |
| C923 | C92 | Myeloid sarcoma | Haematology |
| C924 | C92 | Acute promyelocytic leukaemia [PML] | Haematology |

| | | | |
|---|---|---|---|
| C925 | C92 | Acute myelomonocytic leukaemia | Haematology |
| C926 | C92 | Acute myeloid leukaemia with 11q23-abnormality | Haematology |
| C927 | C92 | Other myeloid leukaemia | Haematology |
| C928 | C92 | Acute myeloid leukaemia with multilineage dysplasia | Haematology |
| C929 | C92 | Myeloid leukaemia, unspecified | Haematology |
| C93 | C93 | Monocytic leukaemia | Haematology |
| C930 | C93 | Acute monoblastic/monocytic leukaemia | Haematology |
| C931 | C93 | Chronic myelomonocytic leukaemia | Haematology |
| C932 | C93 | Subacute monocytic leukaemia | Haematology |
| C933 | C93 | Juvenile myelomonocytic leukaemia | Haematology |
| C937 | C93 | Other monocytic leukaemia | Haematology |
| C939 | C93 | Monocytic leukaemia, unspecified | Haematology |
| C94 | C94 | Other leukaemias of specified cell type | Haematology |
| C940 | C94 | Acute erythroid leukaemia | Haematology |
| C941 | C94 | Chronic erythraemia | Haematology |
| C942 | C94 | Acute megakaryoblastic leukaemia | Haematology |
| C943 | C94 | Mast cell leukaemia | Haematology |
| C944 | C94 | Acute panmyelosis with myelofibrosis | Haematology |
| C945 | C94 | Acute myelofibrosis | Haematology |
| C946 | C94 | Myelodysplastic and myeloproliferative disease, not elsewhere classified | Haematology |
| C947 | C94 | Other specified leukaemias | Haematology |
| C95 | C95 | Leukaemia of unspecified cell type | Haematology |
| C950 | C95 | Acute leukaemia of unsp cell type | Haematology |
| C951 | C95 | Chronic leukaemia unsp cell type | Haematology |
| C952 | C95 | Subacute leukaemia unsp cell type | Haematology |
| C957 | C95 | Other leukaemia unspecified cell type | Haematology |
| C959 | C95 | Leukaemia, unspecified | Haematology |
| C96 | C96 | Oth & unspec malig neop lymphoid, haematapoietic & rel tiss | Haematology |
| C960 | C96 | Multifocal and multisystemic (disseminated) Langerhans-cell histiocytosis [Letterer-Siwe disease] | Haematology |
| C961 | C96 | Malignant histiocytosis | Haematology |
| C962 | C96 | Malignant mast cell tumour | Haematology |
| C963 | C96 | True histiocytic lymphoma | Haematology |
| C964 | C96 | Sarcoma of dendritic cells (accessory cells) | Haematology |

| | | | |
|---|---|---|---|
| C965 | C96 | Multifocal and unisystemic Langerhans-cell histiocytosis | Haematology |
| C966 | C96 | Unifocal Langerhans-cell histiocytosis | Haematology |
| C967 | C96 | Oth spec malig neop lymphoid h'poietic & related tissue | Haematology |
| C968 | C96 | Histiocytic sarcoma | Haematology |
| C969 | C96 | Malig neop lymphoid haematopoietic and related tissue unspec | Haematology |
| C00 | C00 | Malignant neoplasm of lip | Head and Neck |
| C000 | C00 | Malignant neoplasm of external upper lip | Head and Neck |
| C001 | C00 | Malignant neoplasm of external lower lip | Head and Neck |
| C002 | C00 | Malignant neoplasm of external lip, unspecified | Head and Neck |
| C003 | C00 | Malignant neoplasm of upper lip, inner aspect | Head and Neck |
| C004 | C00 | Malignant neoplasm of lower lip, inner aspect | Head and Neck |
| C005 | C00 | Malignant neoplasm of lip, unspecified, inner aspect | Head and Neck |
| C006 | C00 | Malignant neoplasm of commissure of lip | Head and Neck |
| C008 | C00 | Malignant neoplasm of overlapping lesion of lip | Head and Neck |
| C009 | C00 | Malignant neoplasm of lip, unspecified | Head and Neck |
| C01X | C01 | Malignant neoplasm of base of tongue | Head and Neck |
| C02 | C02 | Malignant neoplasm of other and unspecified parts of tongue | Head and Neck |
| C020 | C02 | Malignant neoplasm of dorsal surface tongue | Head and Neck |
| C021 | C02 | Malignant neoplasm of border of tongue | Head and Neck |
| C022 | C02 | Malignant neoplasm of ventral surface of tongue | Head and Neck |
| C023 | C02 | Malignant neo of anterior two-thirds of tongue, part uns | Head and Neck |
| C024 | C02 | Malignant neoplasm of lingual tonsil | Head and Neck |
| C028 | C02 | Malignant neoplasm of overlapping lesion of tongue | Head and Neck |
| C029 | C02 | Malignant neoplasm of tongue, unspecified | Head and Neck |
| C03 | C03 | Malignant neoplasm of gum | Head and Neck |
| C030 | C03 | Malignant neoplasm of upper gum | Head and Neck |
| C031 | C03 | Malignant neoplasm of lower gum | Head and Neck |
| C039 | C03 | Malignant neoplasm of gum unspecified | Head and Neck |
| C04 | C04 | Malignant neoplasm of floor of mouth | Head and Neck |
| C040 | C04 | Malignant neoplasm of floor of anterior floor of mouth | Head and Neck |
| C041 | C04 | Malignant neoplasm of lateral floor of mouth | Head and Neck |
| C048 | C04 | Malignant neoplasm, overlapping lesion of floor of mouth | Head and Neck |
| C049 | C04 | Malignant neoplasm of floor of mouth, floor of mouth, unspec | Head and Neck |

| | | | |
|---|---|---|---|
| C05 | C05 | Malignant neoplasm of palate | Head and Neck |
| C050 | C05 | Malignant neoplasm of hard palate | Head and Neck |
| C051 | C05 | Malignant neoplasm of soft palate | Head and Neck |
| C052 | C05 | Malignant neoplasm of uvula | Head and Neck |
| C058 | C05 | Malignant neoplasm, overlapping lesion of palate | Head and Neck |
| C059 | C05 | Malignant neoplasm of palate, unspecified | Head and Neck |
| C06 | C06 | Malignant neoplasm of other and unspecified parts of mouth | Head and Neck |
| C060 | C06 | Malignant neoplasm cheek mucosa | Head and Neck |
| C061 | C06 | Malignant neoplasm of vestibule of mouth | Head and Neck |
| C062 | C06 | Malignant neoplasm of retromolar area | Head and Neck |
| C068 | C06 | Malignant neoplasm, overlap les of oth & unsp part of mouth | Head and Neck |
| C069 | C06 | Malignant neoplasm of part of mouth, unspecified | Head and Neck |
| C07X | C07 | Malignant neoplasm of parotid gland | Head and Neck |
| C08 | C08 | Maligt neoplasm of oth and unspec major saliv glands | Head and Neck |
| C080 | C08 | Malignant neoplasm of submandibular gland | Head and Neck |
| C081 | C08 | Malignant neoplasm of sublingual gland | Head and Neck |
| C088 | C08 | Malignant neoplasm, overlapping lesion of major saliv gland | Head and Neck |
| C089 | C08 | Malignant neoplasm of major salivary gland, unspecified | Head and Neck |
| C09 | C09 | Malignant neoplasm of tonsil | Head and Neck |
| C090 | C09 | Malignant neoplasm tonsillar fossa | Head and Neck |
| C091 | C09 | Malig neo of tonsillar pillar (anterior)(posterior) | Head and Neck |
| C098 | C09 | Malignant neoplasm of overlapping lesion of tonsil | Head and Neck |
| C099 | C09 | Malignant neoplasm of tonsil unspecified | Head and Neck |
| C10 | C10 | Malignant neoplasm of oropharynx | Head and Neck |
| C100 | C10 | Malignant neoplasm of vallecula | Head and Neck |
| C101 | C10 | Malignant neoplasm of anterior surface of epiglottis | Head and Neck |
| C102 | C10 | Malignant neoplasm of lateral wall of oropharynx | Head and Neck |
| C103 | C10 | Malignant neoplasm of posterior wall of oropharynx | Head and Neck |
| C104 | C10 | Malignant neoplasm of branchial cleft | Head and Neck |
| C108 | C10 | Malignant neoplasm overlapping lesion of oropharynx | Head and Neck |
| C109 | C10 | Malignant neoplasm of oropharynx unspecified | Head and Neck |
| C11 | C11 | Malignant neoplasm of nasopharynx | Head and Neck |
| C110 | C11 | Malignant neoplasm of superior wall of nasopharynx | Head and Neck |

| | | | |
|---|---|---|---|
| C111 | C11 | Malignant neoplasm of posterior wall of nasopharynx | Head and Neck |
| C112 | C11 | Malignant neoplasm of lateral wall of nasopharynx | Head and Neck |
| C113 | C11 | Malignant neoplasm of anterior wall of nasopharynx | Head and Neck |
| C118 | C11 | Malignant neoplasm overlapping lesion of nasopharynx | Head and Neck |
| C119 | C11 | Malignant neoplasm of nasopharynx unspecified | Head and Neck |
| C12X | C12 | Malignant neoplasm of pyriform sinus | Head and Neck |
| C13 | C13 | Malignant neoplasm of hypopharynx | Head and Neck |
| C130 | C13 | Malignant neoplasm of hypopharynx, postcricoid region | Head and Neck |
| C131 | C13 | Malig neoplasm aryepiglottic fold, hypopharyngeal aspect | Head and Neck |
| C132 | C13 | Malignant neoplasm posterior wall of hypopharynx | Head and Neck |
| C138 | C13 | Malignant neoplasm overlapping lesion of hypopharynx | Head and Neck |
| C139 | C13 | Malignant neoplasm of hypopharynx unspecified | Head and Neck |
| C14 | C14 | Mal neo oth ill-def sites lip/oral cavity/pharynx | Head and Neck |
| C140 | C14 | Malignant neoplasm of pharynx, unsp | Head and Neck |
| C142 | C14 | Malignant neoplasm of Waldeyer's ring | Head and Neck |
| C148 | C14 | Malig neo, overlapping lesion of lip, oral cavity & pharynx | Head and Neck |
| C30 | C30 | Malignant neoplasm of nasal cavity and middle ear | Head and Neck |
| C300 | C30 | Malignant neoplasm of nasal cavity | Head and Neck |
| C301 | C30 | Malignant neoplasm of middle ear | Head and Neck |
| C31 | C31 | Malignant neoplasm of accessory sinuses | Head and Neck |
| C310 | C31 | Malignant neoplasm of maxillary sinus | Head and Neck |
| C311 | C31 | Malignant neoplasm of ethmoidal sinus | Head and Neck |
| C312 | C31 | Malignant neoplasm of frontal sinus | Head and Neck |
| C313 | C31 | Malignant neoplasm of sphenoidal sinus | Head and Neck |
| C318 | C31 | Malignant neoplasm, overlapping lesion accessory sinuses | Head and Neck |
| C319 | C31 | Malignant neoplasm of accessory sinus, unsp | Head and Neck |
| C32 | C32 | Malignant neoplasm of larynx | Head and Neck |
| C320 | C32 | Malignant neoplasm of glottis | Head and Neck |
| C320A | C32 | Vocal cords, true | Head and Neck |
| C320B | C32 | Anterior commissure | Head and Neck |
| C320C | C32 | Posterior commissure | Head and Neck |
| C321 | C32 | Malignant neoplasm of supraglottis | Head and Neck |
| C321A | C32 | Suprahyoid epiglottis (tip, laryngeal surface) | Head and Neck |

| | | | |
|---|---|---|---|
| C321B | C32 | Aryepiglottic fold, laryngeal aspect | Head and Neck |
| C321C | C32 | Arytenoid | Head and Neck |
| C321D | C32 | Infrahyoid epiglottis | Head and Neck |
| C321E | C32 | False cords | Head and Neck |
| C322 | C32 | Malignant neoplasm of subglottis | Head and Neck |
| C323 | C32 | Malignant neoplasm of laryngeal cartilage | Head and Neck |
| C323A | C32 | Arytenoid cartilage | Head and Neck |
| C323B | C32 | Cricoid cartilage | Head and Neck |
| C323C | C32 | Thyroid cartilage | Head and Neck |
| C328 | C32 | Malignant neoplasm, overlapping lesion of larynx | Head and Neck |
| C329 | C32 | Malignant neoplasm of larynx, unspecified | Head and Neck |
| C73X | C73 | Malignant neoplasm of thyroid gland | Head and Neck |
| C770 | C77 | Sec & uns malig neoplasm of lymph nodes of head, face & neck | Head and Neck |
| C339A | C33 | Tracheal stoma | Lung |
| C33X | C33 | Malignant neoplasm of trachea | Lung |
| C34 | C34 | Malignant neoplasm of bronchus and lung | Lung |
| C340 | C34 | Malignant neoplasm of main bronchus | Lung |
| C341 | C34 | Malignant neoplasm of upper lobe, bronchus or lung | Lung |
| C342 | C34 | Malignant neoplasm of middle lobe, bronchus or lung | Lung |
| C343 | C34 | Malignant neoplasm of lower lobe, bronchus or lung | Lung |
| C348 | C34 | Malignant neoplasm of overlap les of bronchus & lung | Lung |
| C349 | C34 | Malignant neoplasm of bronchus or lung, unspec | Lung |
| C37X | C37 | Malignant neoplasm of thymus | Lung |
| C38 | C38 | Malignant neoplasm of heart, mediastinum and pleura | Lung |
| C380 | C38 | Malignant neoplasm of heart, mediastinum & pleura, heart | Lung |
| C381 | C38 | Malignant neoplasm of anterior mediastinum | Lung |
| C382 | C38 | Malignant neoplasm of posterior mediastinum | Lung |
| C383 | C38 | Malig neo heart, mediastinum & pleura,mediastinum,part unsp | Lung |
| C384 | C38 | Malignant neoplasm of pleura | Lung |
| C388 | C38 | Malig neo, overlapping lesion of heart, mediastinum & pleura | Lung |
| C39 | C39 | Malig neo oth + ill-def sites resp sys + intrathorac orgs | Lung |
| C390 | C39 | Malignant neoplasm of upper respiratory tract, part unsp | Lung |
| C398 | C39 | Malignant neoplasm, overlap lesion of resp & intrathor orgs | Lung |

| | | | |
|---|---|---|---|
| C399 | C39 | Malignant neoplasm of ill-def sites within the resp sys | Lung |
| C45 | C45 | Mesothelioma | Lung |
| C450 | C45 | Mesothelioma of pleura | Lung |
| C451 | C45 | Mesothelioma of peritoneum | Lung |
| C452 | C45 | Mesothelioma of pericardium | Lung |
| C457 | C45 | Mesothelioma of other sites | Lung |
| C459 | C45 | Mesothelioma, unspecified | Lung |
| C78 | C78 | Sec malignant neoplasm of respiratory and digestive organs | Lung |
| C780 | C78 | Secondary malignant neoplasm of lung | Lung |
| C781 | C78 | Secondary malignant neoplasm of mediastinum | Lung |
| C782 | C78 | Secondary malignant neoplasm of pleura | Lung |
| C783 | C78 | Secondary malignant neoplasm of oth & unsp respiratory orgs | Lung |
| C43 | C43 | Malignant melanoma of skin | Skin |
| C430 | C43 | Malignant melanoma of lip | Skin |
| C431 | C43 | Malignant melanoma of eyelid, including canthus | Skin |
| C432 | C43 | Malignant melanoma of ear and ext auricular canal | Skin |
| C433 | C43 | Malignant melanoma of other and unspecified parts of face | Skin |
| C434 | C43 | Malignant melanoma of scalp and neck | Skin |
| C435 | C43 | Malignant melanoma of trunk | Skin |
| C436 | C43 | Malignant melanoma of upper limb, including shoulder | Skin |
| C437 | C43 | Malignant melanoma of lower limb, including hip | Skin |
| C438 | C43 | Malignant melanoma of skin | Skin |
| C439 | C43 | Malignant melanoma of skin, unsp | Skin |
| C44 | C44 | Other malignant neoplasms of skin | Skin |
| C440 | C44 | Other malignant neoplasms of skin of lip | Skin |
| C441 | C44 | Other malignant neoplasms of skin of eyelid, incl canthus | Skin |
| C442 | C44 | Oth malignant neoplasms of skin of ear & ext auricular canal | Skin |
| C443 | C44 | Oth malignant neoplasm of skin of oth & unsp parts of face | Skin |
| C444 | C44 | Other malignant neoplasms of skin of scalp and neck | Skin |
| C445 | C44 | Other malignant neoplasms of skin of trunk | Skin |
| C446 | C44 | Oth malignant neoplasms of skin of upper limb, incl shoulder | Skin |
| C447 | C44 | Other malignant neoplasms of skin of lower limb, incl hip | Skin |
| C448 | C44 | Other malignant neoplasms, overlapping lesion of skin | Skin |

| | | | |
|---|---|---|---|
| C449 | C44 | Other malignant neoplasms of skin, unspecified | Skin |
| C792 | C79 | Secondary malignant neoplasm of skin | Skin |
| C15 | C15 | Malignant neoplasm of oesophagus | Upper GI |
| C150 | C15 | Malignant neoplasm of cervical part of oesophagus | Upper GI |
| C151 | C15 | Malignant neoplasm of thoracic part of oesophagus | Upper GI |
| C152 | C15 | Malignant neo of abdominal part of oesophagus | Upper GI |
| C153 | C15 | Malignant neoplasm of upper third of oesophagus | Upper GI |
| C154 | C15 | Malignant neoplasm of middle third of oesophagus | Upper GI |
| C155 | C15 | Malignant neoplasm of lower third of oesophagus | Upper GI |
| C158 | C15 | Malignant neoplasm overlapping lesion of oesophagus | Upper GI |
| C159 | C15 | Malignant neoplasm of oesophagus unspecified | Upper GI |
| C16 | C16 | Malignant neoplasm of stomach | Upper GI |
| C160 | C16 | Malignant neoplasm of cardia of stomach | Upper GI |
| C161 | C16 | Malignant neoplasm of fundus of stomach | Upper GI |
| C162 | C16 | Malignant neoplasm of body of stomach | Upper GI |
| C163 | C16 | Malignant neoplasm of pyloric antrum | Upper GI |
| C164 | C16 | Malignant neoplasm of pylorus | Upper GI |
| C165 | C16 | Malignant neoplasm of lesser curvature of stomach, unsp | Upper GI |
| C166 | C16 | Malignant neoplasm of greater curvature of stomach, unsp | Upper GI |
| C168 | C16 | Malignant neoplasm overlapping lesion of stomach | Upper GI |
| C169 | C16 | Malignant neoplasm of stomach, unspecified | Upper GI |
| C22 | C22 | Malignant neoplasm of liver and intrahepatic bile ducts | Upper GI |
| C220 | C22 | Malignant neoplasm, liver cell carcinoma | Upper GI |
| C221 | C22 | Malignant neoplasm, intrahep bile duct carcinoma | Upper GI |
| C222 | C22 | Malignant neoplasm, hepatoblastoma | Upper GI |
| C223 | C22 | Malignant neoplasm, angiosarcoma of liver | Upper GI |
| C224 | C22 | Malignant neoplasm, other sarcomas of liver | Upper GI |
| C227 | C22 | Malignant neoplasm, oth spec carcinomas of liver | Upper GI |
| C229 | C22 | Malignant neoplasm, liver, unspecified | Upper GI |
| C23X | C23 | Malignant neoplasm of gallbladder | Upper GI |
| C24 | C24 | Maligt neoplasm of other and unspec parts biliary tract | Upper GI |
| C240 | C24 | Malignant neoplasm of extrahepatic bile duct | Upper GI |
| C241 | C24 | Malignant neoplasm of Ampulla of Vater | Upper GI |

| | | | |
|---|---|---|---|
| C248 | C24 | Malignant neoplasm overlapping lesion of biliary tract | Upper GI |
| C249 | C24 | Malignant neoplasm of biliary tract, unspecified | Upper GI |
| C25 | C25 | Malignant neoplasm of pancreas | Upper GI |
| C250 | C25 | Malignant neoplasm of head of pancreas | Upper GI |
| C251 | C25 | Malignant neoplasm of body of pancreas | Upper GI |
| C252 | C25 | Malignant neoplasm of tail of pancreas | Upper GI |
| C253 | C25 | Malignant neoplasm of pancreatic duct | Upper GI |
| C254 | C25 | Malignant neoplasm of endocrine pancreas | Upper GI |
| C257 | C25 | Malignant neoplasm of other parts of pancreas | Upper GI |
| C258 | C25 | Malignant neoplasm, overlapping lesion of pancreas | Upper GI |
| C259 | C25 | Malignant neoplasm of pancreas, unspecified | Upper GI |
| C787 | C78 | Secondary malignant neoplasm of liver and intrahepatic bile duct | Upper GI |
| C60 | C60 | Malignant neoplasm of penis | Urology |
| C600 | C60 | Malignant neoplasm of prepuce | Urology |
| C601 | C60 | Malignant neoplasm of glans penis | Urology |
| C602 | C60 | Malignant neoplasm of body of penis | Urology |
| C608 | C60 | Malignant neoplasm, overlapping lesion of penis | Urology |
| C609 | C60 | Malignant neoplasm of penis, unspecified | Urology |
| C61X | C61 | Malignant neoplasm of prostate | Urology |
| C62 | C62 | Malignant neoplasm of testis | Urology |
| C620 | C62 | Malignant neoplasm of undescended testis | Urology |
| C621 | C62 | Malignant neoplasm of descended testis | Urology |
| C629 | C62 | Malignant neoplasm of testis, unspecified | Urology |
| C63 | C63 | Malignant neoplasm of other and unspec male genital organs | Urology |
| C630 | C63 | Malignant neoplasm of epididymis | Urology |
| C631 | C63 | Malignant neoplasm of spermatic cord | Urology |
| C632 | C63 | Malignant neoplasm of scrotum | Urology |
| C637 | C63 | Malignant neoplasm of other specified male genital orgs | Urology |
| C638 | C63 | Malignant neoplasm, overlapping lesion male genital orgs | Urology |
| C639 | C63 | Malignant neoplasm of male genital organ, unspecified | Urology |
| C64X | C64 | Malignant neoplasm of kidney, except renal pelvis | Urology |
| C65X | C65 | Malignant neoplasm of renal pelvis | Urology |
| C66X | C66 | Malignant neoplasm of ureter | Urology |

| | | | |
|---|---|---|---|
| C67 | C67 | Malignant neoplasm of bladder | Urology |
| C670 | C67 | Malignant neoplasm of trigone of bladder | Urology |
| C671 | C67 | Malignant neoplasm of dome of bladder | Urology |
| C672 | C67 | Malignant neoplasm of lateral wall of bladder | Urology |
| C673 | C67 | Malignant neoplasm of anterior wall of bladder | Urology |
| C674 | C67 | Malignant neoplasm of posterior wall of bladder | Urology |
| C675 | C67 | Malignant neoplasm of bladder neck | Urology |
| C676 | C67 | Malignant neoplasm of ureteric orifice | Urology |
| C677 | C67 | Malignant neoplasm of urachus | Urology |
| C678 | C67 | Malignant neoplasm, overlapping lesion of bladder | Urology |
| C679 | C67 | Malignant neoplasm of bladder, unspecified | Urology |
| C68 | C68 | Malignant neoplasm of other and unspecified urinary organs | Urology |
| C680 | C68 | Malignant neoplasm of urethra | Urology |
| C681 | C68 | Malignant neoplasm of paraurethral gland | Urology |
| C688 | C68 | Malignant neoplasm of overlapping lesion urinary organs | Urology |
| C689 | C68 | Malignant neoplasm of urinary organ, unspecified | Urology |
| C790 | C79 | Secondary malignant neoplasm of kidney & renal pelvis | Urology |
| C791 | C79 | Secondary malignant neoplasm of oth & uns urinary organs | Urology |

| ID | ID relative to data source | *Aggregated data from Hospital Information* |
|---|---|---|
| 1 | 1 | Month |
| 2 | 2 | CancerCount |
| 3 | 3 | PostcodeSector |
| 4 | 4 | CancerSite |
| 5 | 5 | AgeUnder18 |
| 6 | 6 | Age18-40 |
| 7 | 7 | Age40-60 |
| 8 | 8 | Age60-80 |
| 9 | 9 | AgeMoreThan80 |
| 10 | 10 | Male |
| 11 | 11 | Female |
| 12 | 12 | CKDStage1 |
| 13 | 13 | CKDStage2 |
| 14 | 14 | CKDStage3 |
| 15 | 15 | CKDStage4 |
| 16 | 16 | CKDStage5 |
| 17 | 17 | DiabetesType1 |
| 18 | 18 | DiabetesType2 |
| 19 | 19 | Diabetes |
| 20 | 20 | CKD |
| 21 | 21 | CongestiveHeartFailure |
| 22 | 22 | COPD |
| 23 | 23 | CoronaryHeartDisease |
| 24 | 24 | Covid19 |
| 25 | 25 | Depression |
| 26 | 26 | Hypertensive |
| 27 | 27 | FrailtyFit |
| 28 | 28 | FrailtyMild |
| 29 | 29 | FrailtyModerate |
| 30 | 30 | FrailtySevere |
| 31 | 31 | UnknownSmoker |
| 32 | 32 | NeverSmoker |
| 33 | 33 | ExSmoker |
| 34 | 34 | CurrentSmoker |

| | | |
|---|---|---|
| 35 | 35 | Anxiety |
| 36 | 36 | African |
| 37 | 37 | British |
| 38 | 38 | AnyotherWhitebackground |
| 39 | 39 | AnyotherBlackbackground |
| 40 | 40 | AnyotherAsianbackground |
| 41 | 41 | Anyothermixedbackground |
| 42 | 42 | Anyotherethnicgroup |
| 43 | 43 | English-ethniccategory2001census |
| 44 | 44 | Irish |
| 45 | 45 | Scottish-ethniccategory2001census |
| 46 | 46 | Chinese |
| 47 | 47 | Indian |
| 48 | 48 | Italian-ethniccategory2001census |
| 49 | 49 | WhiteandBlackAfrican |
| 50 | 50 | WhiteandBlackCaribbean |
| 51 | 51 | WhiteandAsian |
| 52 | 52 | OthermixedWhite-ethniccategory2001census |
| 53 | 53 | OthWhiteEuropean/Europeanunsp/MixedEuropean2001census |
| 54 | 54 | OtherWhiteorWhiteunspecifiedethniccategory2001census |
| 55 | 55 | Polish-ethniccategory2001census |
| 56 | 56 | Pakistani |
| 57 | 57 | Filipino-ethniccategory2001census |
| 58 | 58 | Welsh-ethniccategory2001census |
| 59 | 59 | Bangladeshi |
| 60 | 60 | EastAfricanAsian-ethniccategory2001census |
| 61 | 61 | Caribbean |
| 62 | 62 | ChineseandWhite-ethniccategory2001census |
| 63 | 63 | NorthernIrish-ethniccategory2001census |
| 64 | 64 | Notknown |
| 65 | 65 | Notstated |
| | | ***Geography*** |
| 66 | 1 | Longitude |
| 67 | 2 | Latitude |
| 68 | 3 | LSOA |
| 69 | 4 | Local Authority |
| 70 | 5 | Total Area |
| | | ***Crime*** |

| | | |
|---|---|---|
| 71 | 1 | Criminaldamageandarson |
| 72 | 2 | Vehiclecrime |
| 73 | 3 | Violenceandsexualoffences |
| 74 | 4 | Anti-socialbehaviour |
| 75 | 5 | Publicorder |
| 76 | 6 | Burglary |
| 77 | 7 | Shoplifting |
| 78 | 8 | Drugs |
| 79 | 9 | Bicycletheft |
| 80 | 10 | Theftfromtheperson |
| 81 | 11 | Robbery |
| 82 | 12 | Possessionofweapons |
| 83 | 13 | Othercrime |
| 84 | 14 | Othertheft |
| | | ***Census 2011*** |
| 85 | 1 | Sex_Allpersons;Age_Allcategories_Age;EthnicGroup_White_Total;measures_Value |
| 86 | 2 | Sex_Allpersons;Age_Allcategories_Age;EthnicGroup_White_English/Welsh/Scottish/NorthernIrish/British;measures_Value |
| 87 | 3 | Sex_Allpersons;Age_Allcategories_Age;EthnicGroup_White_Irish;measures_Value |
| 88 | 4 | Sex_Allpersons;Age_Allcategories_Age;EthnicGroup_White_GypsyorIrishTraveller;measures_Value |
| 89 | 5 | Sex_Allpersons;Age_Allcategories_Age;EthnicGroup_White_OtherWhite;measures_Value |
| 90 | 6 | Sex_Allpersons;Age_Allcategories_Age;EthnicGroup_Mixed/multipleethnicgroup_Total;measures_Value |
| 91 | 7 | Sex_Allpersons;Age_Allcategories_Age;EthnicGroup_Mixed/multipleethnicgroup_WhiteandBlackCaribbean;measures_Value |
| 92 | 8 | Sex_Allpersons;Age_Allcategories_Age;EthnicGroup_Mixed/multipleethnicgroup_WhiteandBlackAfrican;measures_Value |
| 93 | 9 | Sex_Allpersons;Age_Allcategories_Age;EthnicGroup_Mixed/multipleethnicgroup_WhiteandAsian;measures_Value |
| 94 | 10 | Sex_Allpersons;Age_Allcategories_Age;EthnicGroup_Mixed/multipleethnicgroup_OtherMixed;measures_Value |
| 95 | 11 | Sex_Allpersons;Age_Allcategories_Age;EthnicGroup_Asian/AsianBritish_Total;measures_Value |
| 96 | 12 | Sex_Allpersons;Age_Allcategories_Age;EthnicGroup_Asian/AsianBritish_Indian;measures_Value |
| 97 | 13 | Sex_Allpersons;Age_Allcategories_Age;EthnicGroup_Asian/AsianBritish_Pakistani;measures_Value |
| 98 | 14 | Sex_Allpersons;Age_Allcategories_Age;EthnicGroup_Asian/AsianBritish_Bangladeshi;measures_Value |
| 99 | 15 | Sex_Allpersons;Age_Allcategories_Age;EthnicGroup_Asian/AsianBritish_Chinese;measures_Value |
| 100 | 16 | Sex_Allpersons;Age_Allcategories_Age;EthnicGroup_Asian/AsianBritish_OtherAsian;measures_Value |
| 101 | 17 | Sex_Allpersons;Age_Allcategories_Age;EthnicGroup_Black/African/Caribbean/BlackBritish_Total;measures_Value |
| 102 | 18 | Sex_Allpersons;Age_Allcategories_Age;EthnicGroup_Black/African/Caribbean/BlackBritish_African;measures_Value |
| 103 | 19 | Sex_Allpersons;Age_Allcategories_Age;EthnicGroup_Black/African/Caribbean/BlackBritish_Caribbean;measures_Value |
| 104 | 20 | Sex_Allpersons;Age_Allcategories_Age;EthnicGroup_Black/African/Caribbean/BlackBritish_OtherBlack;measures_Value |
| 105 | 21 | Sex_Allpersons;Age_Allcategories_Age;EthnicGroup_Otherethnicgroup_Total;measures_Value |
| 106 | 22 | Sex_Allpersons;Age_Allcategories_Age;EthnicGroup_Otherethnicgroup_Arab;measures_Value |
| 107 | 23 | Sex_Allpersons;Age_Allcategories_Age;EthnicGroup_Otherethnicgroup_Anyotherethnicgroup;measures_Value |

| | | |
|---|---|---|
| 108 | 24 | Sex_Allpersons;Age_Age0to24;EthnicGroup_Allcategories_Ethnicgroup;measures_Value |
| 109 | 25 | Sex_Allpersons;Age_Age0to24;EthnicGroup_White_Total;measures_Value |
| 110 | 26 | Sex_Allpersons;Age_Age0to24;EthnicGroup_White_English/Welsh/Scottish/NorthernIrish/British;measures_Value |
| 111 | 27 | Sex_Allpersons;Age_Age0to24;EthnicGroup_White_Irish;measures_Value |
| 112 | 28 | Sex_Allpersons;Age_Age0to24;EthnicGroup_White_GypsyorIrishTraveller;measures_Value |
| 113 | 29 | Sex_Allpersons;Age_Age0to24;EthnicGroup_White_OtherWhite;measures_Value |
| 114 | 30 | Sex_Allpersons;Age_Age0to24;EthnicGroup_Mixed/multipleethnicgroup_Total;measures_Value |
| 115 | 31 | Sex_Allpersons;Age_Age0to24;EthnicGroup_Mixed/multipleethnicgroup_WhiteandBlackCaribbean;measures_Value |
| 116 | 32 | Sex_Allpersons;Age_Age0to24;EthnicGroup_Mixed/multipleethnicgroup_WhiteandBlackAfrican;measures_Value |
| 117 | 33 | Sex_Allpersons;Age_Age0to24;EthnicGroup_Mixed/multipleethnicgroup_WhiteandAsian;measures_Value |
| 118 | 34 | Sex_Allpersons;Age_Age0to24;EthnicGroup_Mixed/multipleethnicgroup_OtherMixed;measures_Value |
| 119 | 35 | Sex_Allpersons;Age_Age0to24;EthnicGroup_Asian/AsianBritish_Total;measures_Value |
| 120 | 36 | Sex_Allpersons;Age_Age0to24;EthnicGroup_Asian/AsianBritish_Indian;measures_Value |
| 121 | 37 | Sex_Allpersons;Age_Age0to24;EthnicGroup_Asian/AsianBritish_Pakistani;measures_Value |
| 122 | 38 | Sex_Allpersons;Age_Age0to24;EthnicGroup_Asian/AsianBritish_Bangladeshi;measures_Value |
| 123 | 39 | Sex_Allpersons;Age_Age0to24;EthnicGroup_Asian/AsianBritish_Chinese;measures_Value |
| 124 | 40 | Sex_Allpersons;Age_Age0to24;EthnicGroup_Asian/AsianBritish_OtherAsian;measures_Value |
| 125 | 41 | Sex_Allpersons;Age_Age0to24;EthnicGroup_Black/African/Caribbean/BlackBritish_Total;measures_Value |
| 126 | 42 | Sex_Allpersons;Age_Age0to24;EthnicGroup_Black/African/Caribbean/BlackBritish_African;measures_Value |
| 127 | 43 | Sex_Allpersons;Age_Age0to24;EthnicGroup_Black/African/Caribbean/BlackBritish_Caribbean;measures_Value |
| 128 | 44 | Sex_Allpersons;Age_Age0to24;EthnicGroup_Black/African/Caribbean/BlackBritish_OtherBlack;measures_Value |
| 129 | 45 | Sex_Allpersons;Age_Age0to24;EthnicGroup_Otherethnicgroup_Total;measures_Value |
| 130 | 46 | Sex_Allpersons;Age_Age0to24;EthnicGroup_Otherethnicgroup_Arab;measures_Value |
| 131 | 47 | Sex_Allpersons;Age_Age0to24;EthnicGroup_Otherethnicgroup_Anyotherethnicgroup;measures_Value |
| 132 | 48 | Sex_Allpersons;Age_Age25to49;EthnicGroup_Allcategories_Ethnicgroup;measures_Value |
| 133 | 49 | Sex_Allpersons;Age_Age25to49;EthnicGroup_White_Total;measures_Value |
| 134 | 50 | Sex_Allpersons;Age_Age25to49;EthnicGroup_White_English/Welsh/Scottish/NorthernIrish/British;measures_Value |
| 135 | 51 | Sex_Allpersons;Age_Age25to49;EthnicGroup_White_Irish;measures_Value |
| 136 | 52 | Sex_Allpersons;Age_Age25to49;EthnicGroup_White_GypsyorIrishTraveller;measures_Value |
| 137 | 53 | Sex_Allpersons;Age_Age25to49;EthnicGroup_White_OtherWhite;measures_Value |
| 138 | 54 | Sex_Allpersons;Age_Age25to49;EthnicGroup_Mixed/multipleethnicgroup_Total;measures_Value |
| 139 | 55 | Sex_Allpersons;Age_Age25to49;EthnicGroup_Mixed/multipleethnicgroup_WhiteandBlackCaribbean;measures_Value |
| 140 | 56 | Sex_Allpersons;Age_Age25to49;EthnicGroup_Mixed/multipleethnicgroup_WhiteandBlackAfrican;measures_Value |
| 141 | 57 | Sex_Allpersons;Age_Age25to49;EthnicGroup_Mixed/multipleethnicgroup_WhiteandAsian;measures_Value |
| 142 | 58 | Sex_Allpersons;Age_Age25to49;EthnicGroup_Mixed/multipleethnicgroup_OtherMixed;measures_Value |
| 143 | 59 | Sex_Allpersons;Age_Age25to49;EthnicGroup_Asian/AsianBritish_Total;measures_Value |
| 144 | 60 | Sex_Allpersons;Age_Age25to49;EthnicGroup_Asian/AsianBritish_Indian;measures_Value |
| 145 | 61 | Sex_Allpersons;Age_Age25to49;EthnicGroup_Asian/AsianBritish_Pakistani;measures_Value |

| | | |
|---|---|---|
| 146 | 62 | Sex_Allpersons;Age_Age25to49;EthnicGroup_Asian/AsianBritish_Bangladeshi;measures_Value |
| 147 | 63 | Sex_Allpersons;Age_Age25to49;EthnicGroup_Asian/AsianBritish_Chinese;measures_Value |
| 148 | 64 | Sex_Allpersons;Age_Age25to49;EthnicGroup_Asian/AsianBritish_OtherAsian;measures_Value |
| 149 | 65 | Sex_Allpersons;Age_Age25to49;EthnicGroup_Black/African/Caribbean/BlackBritish_Total;measures_Value |
| 150 | 66 | Sex_Allpersons;Age_Age25to49;EthnicGroup_Black/African/Caribbean/BlackBritish_African;measures_Value |
| 151 | 67 | Sex_Allpersons;Age_Age25to49;EthnicGroup_Black/African/Caribbean/BlackBritish_Caribbean;measures_Value |
| 152 | 68 | Sex_Allpersons;Age_Age25to49;EthnicGroup_Black/African/Caribbean/BlackBritish_OtherBlack;measures_Value |
| 153 | 69 | Sex_Allpersons;Age_Age25to49;EthnicGroup_Otherethnicgroup_Total;measures_Value |
| 154 | 70 | Sex_Allpersons;Age_Age25to49;EthnicGroup_Otherethnicgroup_Arab;measures_Value |
| 155 | 71 | Sex_Allpersons;Age_Age25to49;EthnicGroup_Otherethnicgroup_Anyotherethnicgroup;measures_Value |
| 156 | 72 | Sex_Allpersons;Age_Age50to64;EthnicGroup_Allcategories_Ethnicgroup;measures_Value |
| 157 | 73 | Sex_Allpersons;Age_Age50to64;EthnicGroup_White_Total;measures_Value |
| 158 | 74 | Sex_Allpersons;Age_Age50to64;EthnicGroup_White_English/Welsh/Scottish/NorthernIrish/British;measures_Value |
| 159 | 75 | Sex_Allpersons;Age_Age50to64;EthnicGroup_White_Irish;measures_Value |
| 160 | 76 | Sex_Allpersons;Age_Age50to64;EthnicGroup_White_GypsyorIrishTraveller;measures_Value |
| 161 | 77 | Sex_Allpersons;Age_Age50to64;EthnicGroup_White_OtherWhite;measures_Value |
| 162 | 78 | Sex_Allpersons;Age_Age50to64;EthnicGroup_Mixed/multipleethnicgroup_Total;measures_Value |
| 163 | 79 | Sex_Allpersons;Age_Age50to64;EthnicGroup_Mixed/multipleethnicgroup_WhiteandBlackCaribbean;measures_Value |
| 164 | 80 | Sex_Allpersons;Age_Age50to64;EthnicGroup_Mixed/multipleethnicgroup_WhiteandBlackAfrican;measures_Value |
| 165 | 81 | Sex_Allpersons;Age_Age50to64;EthnicGroup_Mixed/multipleethnicgroup_WhiteandAsian;measures_Value |
| 166 | 82 | Sex_Allpersons;Age_Age50to64;EthnicGroup_Mixed/multipleethnicgroup_OtherMixed;measures_Value |
| 167 | 83 | Sex_Allpersons;Age_Age50to64;EthnicGroup_Asian/AsianBritish_Total;measures_Value |
| 168 | 84 | Sex_Allpersons;Age_Age50to64;EthnicGroup_Asian/AsianBritish_Indian;measures_Value |
| 169 | 85 | Sex_Allpersons;Age_Age50to64;EthnicGroup_Asian/AsianBritish_Pakistani;measures_Value |
| 170 | 86 | Sex_Allpersons;Age_Age50to64;EthnicGroup_Asian/AsianBritish_Bangladeshi;measures_Value |
| 171 | 87 | Sex_Allpersons;Age_Age50to64;EthnicGroup_Asian/AsianBritish_Chinese;measures_Value |
| 172 | 88 | Sex_Allpersons;Age_Age50to64;EthnicGroup_Asian/AsianBritish_OtherAsian;measures_Value |
| 173 | 89 | Sex_Allpersons;Age_Age50to64;EthnicGroup_Black/African/Caribbean/BlackBritish_Total;measures_Value |
| 174 | 90 | Sex_Allpersons;Age_Age50to64;EthnicGroup_Black/African/Caribbean/BlackBritish_African;measures_Value |
| 175 | 91 | Sex_Allpersons;Age_Age50to64;EthnicGroup_Black/African/Caribbean/BlackBritish_Caribbean;measures_Value |
| 176 | 92 | Sex_Allpersons;Age_Age50to64;EthnicGroup_Black/African/Caribbean/BlackBritish_OtherBlack;measures_Value |
| 177 | 93 | Sex_Allpersons;Age_Age50to64;EthnicGroup_Otherethnicgroup_Total;measures_Value |
| 178 | 94 | Sex_Allpersons;Age_Age50to64;EthnicGroup_Otherethnicgroup_Arab;measures_Value |
| 179 | 95 | Sex_Allpersons;Age_Age50to64;EthnicGroup_Otherethnicgroup_Anyotherethnicgroup;measures_Value |
| 180 | 96 | Sex_Allpersons;Age_Age65andover;EthnicGroup_Allcategories_Ethnicgroup;measures_Value |
| 181 | 97 | Sex_Allpersons;Age_Age65andover;EthnicGroup_White_Total;measures_Value |
| 182 | 98 | Sex_Allpersons;Age_Age65andover;EthnicGroup_White_English/Welsh/Scottish/NorthernIrish/British;measures_Value |
| 183 | 99 | Sex_Allpersons;Age_Age65andover;EthnicGroup_White_Irish;measures_Value |

| | | |
|---|---|---|
| 184 | 100 | Sex_Allpersons;Age_Age65andover;EthnicGroup_White_GypsyorIrishTraveller;measures_Value |
| 185 | 101 | Sex_Allpersons;Age_Age65andover;EthnicGroup_White_OtherWhite;measures_Value |
| 186 | 102 | Sex_Allpersons;Age_Age65andover;EthnicGroup_Mixed/multipleethnicgroup_Total;measures_Value |
| 187 | 103 | Sex_Allpersons;Age_Age65andover;EthnicGroup_Mixed/multipleethnicgroup_WhiteandBlackCaribbean;measures_Value |
| 188 | 104 | Sex_Allpersons;Age_Age65andover;EthnicGroup_Mixed/multipleethnicgroup_WhiteandBlackAfrican;measures_Value |
| 189 | 105 | Sex_Allpersons;Age_Age65andover;EthnicGroup_Mixed/multipleethnicgroup_WhiteandAsian;measures_Value |
| 190 | 106 | Sex_Allpersons;Age_Age65andover;EthnicGroup_Mixed/multipleethnicgroup_OtherMixed;measures_Value |
| 191 | 107 | Sex_Allpersons;Age_Age65andover;EthnicGroup_Asian/AsianBritish_Total;measures_Value |
| 192 | 108 | Sex_Allpersons;Age_Age65andover;EthnicGroup_Asian/AsianBritish_Indian;measures_Value |
| 193 | 109 | Sex_Allpersons;Age_Age65andover;EthnicGroup_Asian/AsianBritish_Pakistani;measures_Value |
| 194 | 110 | Sex_Allpersons;Age_Age65andover;EthnicGroup_Asian/AsianBritish_Bangladeshi;measures_Value |
| 195 | 111 | Sex_Allpersons;Age_Age65andover;EthnicGroup_Asian/AsianBritish_Chinese;measures_Value |
| 196 | 112 | Sex_Allpersons;Age_Age65andover;EthnicGroup_Asian/AsianBritish_OtherAsian;measures_Value |
| 197 | 113 | Sex_Allpersons;Age_Age65andover;EthnicGroup_Black/African/Caribbean/BlackBritish_Total;measures_Value |
| 198 | 114 | Sex_Allpersons;Age_Age65andover;EthnicGroup_Black/African/Caribbean/BlackBritish_African;measures_Value |
| 199 | 115 | Sex_Allpersons;Age_Age65andover;EthnicGroup_Black/African/Caribbean/BlackBritish_Caribbean;measures_Value |
| 200 | 116 | Sex_Allpersons;Age_Age65andover;EthnicGroup_Black/African/Caribbean/BlackBritish_OtherBlack;measures_Value |
| 201 | 117 | Sex_Allpersons;Age_Age65andover;EthnicGroup_Otherethnicgroup_Total;measures_Value |
| 202 | 118 | Sex_Allpersons;Age_Age65andover;EthnicGroup_Otherethnicgroup_Arab;measures_Value |
| 203 | 119 | Sex_Allpersons;Age_Age65andover;EthnicGroup_Otherethnicgroup_Anyotherethnicgroup;measures_Value |
| 204 | 120 | Sex_Males;Age_Allcategories_Age;EthnicGroup_Allcategories_Ethnicgroup;measures_Value |
| 205 | 121 | Sex_Males;Age_Allcategories_Age;EthnicGroup_White_Total;measures_Value |
| 206 | 122 | Sex_Males;Age_Allcategories_Age;EthnicGroup_White_English/Welsh/Scottish/NorthernIrish/British;measures_Value |
| 207 | 123 | Sex_Males;Age_Allcategories_Age;EthnicGroup_White_Irish;measures_Value |
| 208 | 124 | Sex_Males;Age_Allcategories_Age;EthnicGroup_White_GypsyorIrishTraveller;measures_Value |
| 209 | 125 | Sex_Males;Age_Allcategories_Age;EthnicGroup_White_OtherWhite;measures_Value |
| 210 | 126 | Sex_Males;Age_Allcategories_Age;EthnicGroup_Mixed/multipleethnicgroup_Total;measures_Value |
| 211 | 127 | Sex_Males;Age_Allcategories_Age;EthnicGroup_Mixed/multipleethnicgroup_WhiteandBlackCaribbean;measures_Value |
| 212 | 128 | Sex_Males;Age_Allcategories_Age;EthnicGroup_Mixed/multipleethnicgroup_WhiteandBlackAfrican;measures_Value |
| 213 | 129 | Sex_Males;Age_Allcategories_Age;EthnicGroup_Mixed/multipleethnicgroup_WhiteandAsian;measures_Value |
| 214 | 130 | Sex_Males;Age_Allcategories_Age;EthnicGroup_Mixed/multipleethnicgroup_OtherMixed;measures_Value |
| 215 | 131 | Sex_Males;Age_Allcategories_Age;EthnicGroup_Asian/AsianBritish_Total;measures_Value |
| 216 | 132 | Sex_Males;Age_Allcategories_Age;EthnicGroup_Asian/AsianBritish_Indian;measures_Value |
| 217 | 133 | Sex_Males;Age_Allcategories_Age;EthnicGroup_Asian/AsianBritish_Pakistani;measures_Value |
| 218 | 134 | Sex_Males;Age_Allcategories_Age;EthnicGroup_Asian/AsianBritish_Bangladeshi;measures_Value |
| 219 | 135 | Sex_Males;Age_Allcategories_Age;EthnicGroup_Asian/AsianBritish_Chinese;measures_Value |
| 220 | 136 | Sex_Males;Age_Allcategories_Age;EthnicGroup_Asian/AsianBritish_OtherAsian;measures_Value |
| 221 | 137 | Sex_Males;Age_Allcategories_Age;EthnicGroup_Black/African/Caribbean/BlackBritish_Total;measures_Value |

| | | |
|---|---|---|
| 222 | 138 | Sex_Males;Age_Allcategories_Age;EthnicGroup_Black/African/Caribbean/BlackBritish_African;measures_Value |
| 223 | 139 | Sex_Males;Age_Allcategories_Age;EthnicGroup_Black/African/Caribbean/BlackBritish_Caribbean;measures_Value |
| 224 | 140 | Sex_Males;Age_Allcategories_Age;EthnicGroup_Black/African/Caribbean/BlackBritish_OtherBlack;measures_Value |
| 225 | 141 | Sex_Males;Age_Allcategories_Age;EthnicGroup_Otherethnicgroup_Total;measures_Value |
| 226 | 142 | Sex_Males;Age_Allcategories_Age;EthnicGroup_Otherethnicgroup_Arab;measures_Value |
| 227 | 143 | Sex_Males;Age_Allcategories_Age;EthnicGroup_Otherethnicgroup_Anyotherethnicgroup;measures_Value |
| 228 | 144 | Sex_Males;Age_Age0to24;EthnicGroup_Allcategories_Ethnicgroup;measures_Value |
| 229 | 145 | Sex_Males;Age_Age0to24;EthnicGroup_White_Total;measures_Value |
| 230 | 146 | Sex_Males;Age_Age0to24;EthnicGroup_White_English/Welsh/Scottish/NorthernIrish/British;measures_Value |
| 231 | 147 | Sex_Males;Age_Age0to24;EthnicGroup_White_Irish;measures_Value |
| 232 | 148 | Sex_Males;Age_Age0to24;EthnicGroup_White_GypsyorIrishTraveller;measures_Value |
| 233 | 149 | Sex_Males;Age_Age0to24;EthnicGroup_White_OtherWhite;measures_Value |
| 234 | 150 | Sex_Males;Age_Age0to24;EthnicGroup_Mixed/multipleethnicgroup_Total;measures_Value |
| 235 | 151 | Sex_Males;Age_Age0to24;EthnicGroup_Mixed/multipleethnicgroup_WhiteandBlackCaribbean;measures_Value |
| 236 | 152 | Sex_Males;Age_Age0to24;EthnicGroup_Mixed/multipleethnicgroup_WhiteandBlackAfrican;measures_Value |
| 237 | 153 | Sex_Males;Age_Age0to24;EthnicGroup_Mixed/multipleethnicgroup_WhiteandAsian;measures_Value |
| 238 | 154 | Sex_Males;Age_Age0to24;EthnicGroup_Mixed/multipleethnicgroup_OtherMixed;measures_Value |
| 239 | 155 | Sex_Males;Age_Age0to24;EthnicGroup_Asian/AsianBritish_Total;measures_Value |
| 240 | 156 | Sex_Males;Age_Age0to24;EthnicGroup_Asian/AsianBritish_Indian;measures_Value |
| 241 | 157 | Sex_Males;Age_Age0to24;EthnicGroup_Asian/AsianBritish_Pakistani;measures_Value |
| 242 | 158 | Sex_Males;Age_Age0to24;EthnicGroup_Asian/AsianBritish_Bangladeshi;measures_Value |
| 243 | 159 | Sex_Males;Age_Age0to24;EthnicGroup_Asian/AsianBritish_Chinese;measures_Value |
| 244 | 160 | Sex_Males;Age_Age0to24;EthnicGroup_Asian/AsianBritish_OtherAsian;measures_Value |
| 245 | 161 | Sex_Males;Age_Age0to24;EthnicGroup_Black/African/Caribbean/BlackBritish_Total;measures_Value |
| 246 | 162 | Sex_Males;Age_Age0to24;EthnicGroup_Black/African/Caribbean/BlackBritish_African;measures_Value |
| 247 | 163 | Sex_Males;Age_Age0to24;EthnicGroup_Black/African/Caribbean/BlackBritish_Caribbean;measures_Value |
| 248 | 164 | Sex_Males;Age_Age0to24;EthnicGroup_Black/African/Caribbean/BlackBritish_OtherBlack;measures_Value |
| 249 | 165 | Sex_Males;Age_Age0to24;EthnicGroup_Otherethnicgroup_Total;measures_Value |
| 250 | 166 | Sex_Males;Age_Age0to24;EthnicGroup_Otherethnicgroup_Arab;measures_Value |
| 251 | 167 | Sex_Males;Age_Age0to24;EthnicGroup_Otherethnicgroup_Anyotherethnicgroup;measures_Value |
| 252 | 168 | Sex_Males;Age_Age25to49;EthnicGroup_Allcategories_Ethnicgroup;measures_Value |
| 253 | 169 | Sex_Males;Age_Age25to49;EthnicGroup_White_Total;measures_Value |
| 254 | 170 | Sex_Males;Age_Age25to49;EthnicGroup_White_English/Welsh/Scottish/NorthernIrish/British;measures_Value |
| 255 | 171 | Sex_Males;Age_Age25to49;EthnicGroup_White_Irish;measures_Value |
| 256 | 172 | Sex_Males;Age_Age25to49;EthnicGroup_White_GypsyorIrishTraveller;measures_Value |
| 257 | 173 | Sex_Males;Age_Age25to49;EthnicGroup_White_OtherWhite;measures_Value |
| 258 | 174 | Sex_Males;Age_Age25to49;EthnicGroup_Mixed/multipleethnicgroup_Total;measures_Value |
| 259 | 175 | Sex_Males;Age_Age25to49;EthnicGroup_Mixed/multipleethnicgroup_WhiteandBlackCaribbean;measures_Value |

| | | |
|---|---|---|
| 260 | 176 | Sex_Males;Age_Age25to49;EthnicGroup_Mixed/multipleethnicgroup_WhiteandBlackAfrican;measures_Value |
| 261 | 177 | Sex_Males;Age_Age25to49;EthnicGroup_Mixed/multipleethnicgroup_WhiteandAsian;measures_Value |
| 262 | 178 | Sex_Males;Age_Age25to49;EthnicGroup_Mixed/multipleethnicgroup_OtherMixed;measures_Value |
| 263 | 179 | Sex_Males;Age_Age25to49;EthnicGroup_Asian/AsianBritish_Total;measures_Value |
| 264 | 180 | Sex_Males;Age_Age25to49;EthnicGroup_Asian/AsianBritish_Indian;measures_Value |
| 265 | 181 | Sex_Males;Age_Age25to49;EthnicGroup_Asian/AsianBritish_Pakistani;measures_Value |
| 266 | 182 | Sex_Males;Age_Age25to49;EthnicGroup_Asian/AsianBritish_Bangladeshi;measures_Value |
| 267 | 183 | Sex_Males;Age_Age25to49;EthnicGroup_Asian/AsianBritish_Chinese;measures_Value |
| 268 | 184 | Sex_Males;Age_Age25to49;EthnicGroup_Asian/AsianBritish_OtherAsian;measures_Value |
| 269 | 185 | Sex_Males;Age_Age25to49;EthnicGroup_Black/African/Caribbean/BlackBritish_Total;measures_Value |
| 270 | 186 | Sex_Males;Age_Age25to49;EthnicGroup_Black/African/Caribbean/BlackBritish_African;measures_Value |
| 271 | 187 | Sex_Males;Age_Age25to49;EthnicGroup_Black/African/Caribbean/BlackBritish_Caribbean;measures_Value |
| 272 | 188 | Sex_Males;Age_Age25to49;EthnicGroup_Black/African/Caribbean/BlackBritish_OtherBlack;measures_Value |
| 273 | 189 | Sex_Males;Age_Age25to49;EthnicGroup_Otherethnicgroup_Total;measures_Value |
| 274 | 190 | Sex_Males;Age_Age25to49;EthnicGroup_Otherethnicgroup_Arab;measures_Value |
| 275 | 191 | Sex_Males;Age_Age25to49;EthnicGroup_Otherethnicgroup_Anyotherethnicgroup;measures_Value |
| 276 | 192 | Sex_Males;Age_Age50to64;EthnicGroup_Allcategories_Ethnicgroup;measures_Value |
| 277 | 193 | Sex_Males;Age_Age50to64;EthnicGroup_White_Total;measures_Value |
| 278 | 194 | Sex_Males;Age_Age50to64;EthnicGroup_White_English/Welsh/Scottish/NorthernIrish/British;measures_Value |
| 279 | 195 | Sex_Males;Age_Age50to64;EthnicGroup_White_Irish;measures_Value |
| 280 | 196 | Sex_Males;Age_Age50to64;EthnicGroup_White_GypsyorIrishTraveller;measures_Value |
| 281 | 197 | Sex_Males;Age_Age50to64;EthnicGroup_White_OtherWhite;measures_Value |
| 282 | 198 | Sex_Males;Age_Age50to64;EthnicGroup_Mixed/multipleethnicgroup_Total;measures_Value |
| 283 | 199 | Sex_Males;Age_Age50to64;EthnicGroup_Mixed/multipleethnicgroup_WhiteandBlackCaribbean;measures_Value |
| 284 | 200 | Sex_Males;Age_Age50to64;EthnicGroup_Mixed/multipleethnicgroup_WhiteandBlackAfrican;measures_Value |
| 285 | 201 | Sex_Males;Age_Age50to64;EthnicGroup_Mixed/multipleethnicgroup_WhiteandAsian;measures_Value |
| 286 | 202 | Sex_Males;Age_Age50to64;EthnicGroup_Mixed/multipleethnicgroup_OtherMixed;measures_Value |
| 287 | 203 | Sex_Males;Age_Age50to64;EthnicGroup_Asian/AsianBritish_Total;measures_Value |
| 288 | 204 | Sex_Males;Age_Age50to64;EthnicGroup_Asian/AsianBritish_Indian;measures_Value |
| 289 | 205 | Sex_Males;Age_Age50to64;EthnicGroup_Asian/AsianBritish_Pakistani;measures_Value |
| 290 | 206 | Sex_Males;Age_Age50to64;EthnicGroup_Asian/AsianBritish_Bangladeshi;measures_Value |
| 291 | 207 | Sex_Males;Age_Age50to64;EthnicGroup_Asian/AsianBritish_Chinese;measures_Value |
| 292 | 208 | Sex_Males;Age_Age50to64;EthnicGroup_Asian/AsianBritish_OtherAsian;measures_Value |
| 293 | 209 | Sex_Males;Age_Age50to64;EthnicGroup_Black/African/Caribbean/BlackBritish_Total;measures_Value |
| 294 | 210 | Sex_Males;Age_Age50to64;EthnicGroup_Black/African/Caribbean/BlackBritish_African;measures_Value |
| 295 | 211 | Sex_Males;Age_Age50to64;EthnicGroup_Black/African/Caribbean/BlackBritish_Caribbean;measures_Value |
| 296 | 212 | Sex_Males;Age_Age50to64;EthnicGroup_Black/African/Caribbean/BlackBritish_OtherBlack;measures_Value |
| 297 | 213 | Sex_Males;Age_Age50to64;EthnicGroup_Otherethnicgroup_Total;measures_Value |

| | | |
|---|---|---|
| 298 | 214 | Sex_Males;Age_Age50to64;EthnicGroup_Otherethnicgroup_Arab;measures_Value |
| 299 | 215 | Sex_Males;Age_Age50to64;EthnicGroup_Otherethnicgroup_Anyotherethnicgroup;measures_Value |
| 300 | 216 | Sex_Males;Age_Age65andover;EthnicGroup_Allcategories_Ethnicgroup;measures_Value |
| 301 | 217 | Sex_Males;Age_Age65andover;EthnicGroup_White_Total;measures_Value |
| 302 | 218 | Sex_Males;Age_Age65andover;EthnicGroup_White_English/Welsh/Scottish/NorthernIrish/British;measures_Value |
| 303 | 219 | Sex_Males;Age_Age65andover;EthnicGroup_White_Irish;measures_Value |
| 304 | 220 | Sex_Males;Age_Age65andover;EthnicGroup_White_GypsyorIrishTraveller;measures_Value |
| 305 | 221 | Sex_Males;Age_Age65andover;EthnicGroup_White_OtherWhite;measures_Value |
| 306 | 222 | Sex_Males;Age_Age65andover;EthnicGroup_Mixed/multipleethnicgroup_Total;measures_Value |
| 307 | 223 | Sex_Males;Age_Age65andover;EthnicGroup_Mixed/multipleethnicgroup_WhiteandBlackCaribbean;measures_Value |
| 308 | 224 | Sex_Males;Age_Age65andover;EthnicGroup_Mixed/multipleethnicgroup_WhiteandBlackAfrican;measures_Value |
| 309 | 225 | Sex_Males;Age_Age65andover;EthnicGroup_Mixed/multipleethnicgroup_WhiteandAsian;measures_Value |
| 310 | 226 | Sex_Males;Age_Age65andover;EthnicGroup_Mixed/multipleethnicgroup_OtherMixed;measures_Value |
| 311 | 227 | Sex_Males;Age_Age65andover;EthnicGroup_Asian/AsianBritish_Total;measures_Value |
| 312 | 228 | Sex_Males;Age_Age65andover;EthnicGroup_Asian/AsianBritish_Indian;measures_Value |
| 313 | 229 | Sex_Males;Age_Age65andover;EthnicGroup_Asian/AsianBritish_Pakistani;measures_Value |
| 314 | 230 | Sex_Males;Age_Age65andover;EthnicGroup_Asian/AsianBritish_Bangladeshi;measures_Value |
| 315 | 231 | Sex_Males;Age_Age65andover;EthnicGroup_Asian/AsianBritish_Chinese;measures_Value |
| 316 | 232 | Sex_Males;Age_Age65andover;EthnicGroup_Asian/AsianBritish_OtherAsian;measures_Value |
| 317 | 233 | Sex_Males;Age_Age65andover;EthnicGroup_Black/African/Caribbean/BlackBritish_Total;measures_Value |
| 318 | 234 | Sex_Males;Age_Age65andover;EthnicGroup_Black/African/Caribbean/BlackBritish_African;measures_Value |
| 319 | 235 | Sex_Males;Age_Age65andover;EthnicGroup_Black/African/Caribbean/BlackBritish_Caribbean;measures_Value |
| 320 | 236 | Sex_Males;Age_Age65andover;EthnicGroup_Black/African/Caribbean/BlackBritish_OtherBlack;measures_Value |
| 321 | 237 | Sex_Males;Age_Age65andover;EthnicGroup_Otherethnicgroup_Total;measures_Value |
| 322 | 238 | Sex_Males;Age_Age65andover;EthnicGroup_Otherethnicgroup_Arab;measures_Value |
| 323 | 239 | Sex_Males;Age_Age65andover;EthnicGroup_Otherethnicgroup_Anyotherethnicgroup;measures_Value |
| 324 | 240 | Sex_Females;Age_Allcategories_Age;EthnicGroup_Allcategories_Ethnicgroup;measures_Value |
| 325 | 241 | Sex_Females;Age_Allcategories_Age;EthnicGroup_White_Total;measures_Value |
| 326 | 242 | Sex_Females;Age_Allcategories_Age;EthnicGroup_White_English/Welsh/Scottish/NorthernIrish/British;measures_Value |
| 327 | 243 | Sex_Females;Age_Allcategories_Age;EthnicGroup_White_Irish;measures_Value |
| 328 | 244 | Sex_Females;Age_Allcategories_Age;EthnicGroup_White_GypsyorIrishTraveller;measures_Value |
| 329 | 245 | Sex_Females;Age_Allcategories_Age;EthnicGroup_White_OtherWhite;measures_Value |
| 330 | 246 | Sex_Females;Age_Allcategories_Age;EthnicGroup_Mixed/multipleethnicgroup_Total;measures_Value |
| 331 | 247 | Sex_Females;Age_Allcategories_Age;EthnicGroup_Mixed/multipleethnicgroup_WhiteandBlackCaribbean;measures_Value |
| 332 | 248 | Sex_Females;Age_Allcategories_Age;EthnicGroup_Mixed/multipleethnicgroup_WhiteandBlackAfrican;measures_Value |
| 333 | 249 | Sex_Females;Age_Allcategories_Age;EthnicGroup_Mixed/multipleethnicgroup_WhiteandAsian;measures_Value |
| 334 | 250 | Sex_Females;Age_Allcategories_Age;EthnicGroup_Mixed/multipleethnicgroup_OtherMixed;measures_Value |
| 335 | 251 | Sex_Females;Age_Allcategories_Age;EthnicGroup_Asian/AsianBritish_Total;measures_Value |

| | | |
|---|---|---|
| 336 | 252 | Sex_Females;Age_Allcategories_Age;EthnicGroup_Asian/AsianBritish_Indian;measures_Value |
| 337 | 253 | Sex_Females;Age_Allcategories_Age;EthnicGroup_Asian/AsianBritish_Pakistani;measures_Value |
| 338 | 254 | Sex_Females;Age_Allcategories_Age;EthnicGroup_Asian/AsianBritish_Bangladeshi;measures_Value |
| 339 | 255 | Sex_Females;Age_Allcategories_Age;EthnicGroup_Asian/AsianBritish_Chinese;measures_Value |
| 340 | 256 | Sex_Females;Age_Allcategories_Age;EthnicGroup_Asian/AsianBritish_OtherAsian;measures_Value |
| 341 | 257 | Sex_Females;Age_Allcategories_Age;EthnicGroup_Black/African/Caribbean/BlackBritish_Total;measures_Value |
| 342 | 258 | Sex_Females;Age_Allcategories_Age;EthnicGroup_Black/African/Caribbean/BlackBritish_African;measures_Value |
| 343 | 259 | Sex_Females;Age_Allcategories_Age;EthnicGroup_Black/African/Caribbean/BlackBritish_Caribbean;measures_Value |
| 344 | 260 | Sex_Females;Age_Allcategories_Age;EthnicGroup_Black/African/Caribbean/BlackBritish_OtherBlack;measures_Value |
| 345 | 261 | Sex_Females;Age_Allcategories_Age;EthnicGroup_Otherethnicgroup_Total;measures_Value |
| 346 | 262 | Sex_Females;Age_Allcategories_Age;EthnicGroup_Otherethnicgroup_Arab;measures_Value |
| 347 | 263 | Sex_Females;Age_Allcategories_Age;EthnicGroup_Otherethnicgroup_Anyotherethnicgroup;measures_Value |
| 348 | 264 | Sex_Females;Age_Age0to24;EthnicGroup_Allcategories_Ethnicgroup;measures_Value |
| 349 | 265 | Sex_Females;Age_Age0to24;EthnicGroup_White_Total;measures_Value |
| 350 | 266 | Sex_Females;Age_Age0to24;EthnicGroup_White_English/Welsh/Scottish/NorthernIrish/British;measures_Value |
| 351 | 267 | Sex_Females;Age_Age0to24;EthnicGroup_White_Irish;measures_Value |
| 352 | 268 | Sex_Females;Age_Age0to24;EthnicGroup_White_GypsyorIrishTraveller;measures_Value |
| 353 | 269 | Sex_Females;Age_Age0to24;EthnicGroup_White_OtherWhite;measures_Value |
| 354 | 270 | Sex_Females;Age_Age0to24;EthnicGroup_Mixed/multipleethnicgroup_Total;measures_Value |
| 355 | 271 | Sex_Females;Age_Age0to24;EthnicGroup_Mixed/multipleethnicgroup_WhiteandBlackCaribbean;measures_Value |
| 356 | 272 | Sex_Females;Age_Age0to24;EthnicGroup_Mixed/multipleethnicgroup_WhiteandBlackAfrican;measures_Value |
| 357 | 273 | Sex_Females;Age_Age0to24;EthnicGroup_Mixed/multipleethnicgroup_WhiteandAsian;measures_Value |
| 358 | 274 | Sex_Females;Age_Age0to24;EthnicGroup_Mixed/multipleethnicgroup_OtherMixed;measures_Value |
| 359 | 275 | Sex_Females;Age_Age0to24;EthnicGroup_Asian/AsianBritish_Total;measures_Value |
| 360 | 276 | Sex_Females;Age_Age0to24;EthnicGroup_Asian/AsianBritish_Indian;measures_Value |
| 361 | 277 | Sex_Females;Age_Age0to24;EthnicGroup_Asian/AsianBritish_Pakistani;measures_Value |
| 362 | 278 | Sex_Females;Age_Age0to24;EthnicGroup_Asian/AsianBritish_Bangladeshi;measures_Value |
| 363 | 279 | Sex_Females;Age_Age0to24;EthnicGroup_Asian/AsianBritish_Chinese;measures_Value |
| 364 | 280 | Sex_Females;Age_Age0to24;EthnicGroup_Asian/AsianBritish_OtherAsian;measures_Value |
| 365 | 281 | Sex_Females;Age_Age0to24;EthnicGroup_Black/African/Caribbean/BlackBritish_Total;measures_Value |
| 366 | 282 | Sex_Females;Age_Age0to24;EthnicGroup_Black/African/Caribbean/BlackBritish_African;measures_Value |
| 367 | 283 | Sex_Females;Age_Age0to24;EthnicGroup_Black/African/Caribbean/BlackBritish_Caribbean;measures_Value |
| 368 | 284 | Sex_Females;Age_Age0to24;EthnicGroup_Black/African/Caribbean/BlackBritish_OtherBlack;measures_Value |
| 369 | 285 | Sex_Females;Age_Age0to24;EthnicGroup_Otherethnicgroup_Total;measures_Value |
| 370 | 286 | Sex_Females;Age_Age0to24;EthnicGroup_Otherethnicgroup_Arab;measures_Value |
| 371 | 287 | Sex_Females;Age_Age0to24;EthnicGroup_Otherethnicgroup_Anyotherethnicgroup;measures_Value |
| 372 | 288 | Sex_Females;Age_Age25to49;EthnicGroup_Allcategories_Ethnicgroup;measures_Value |
| 373 | 289 | Sex_Females;Age_Age25to49;EthnicGroup_White_Total;measures_Value |

| | | |
|---|---|---|
| 374 | 290 | Sex_Females;Age_Age25to49;EthnicGroup_White_English/Welsh/Scottish/NorthernIrish/British;measures_Value |
| 375 | 291 | Sex_Females;Age_Age25to49;EthnicGroup_White_Irish;measures_Value |
| 376 | 292 | Sex_Females;Age_Age25to49;EthnicGroup_White_GypsyorIrishTraveller;measures_Value |
| 377 | 293 | Sex_Females;Age_Age25to49;EthnicGroup_White_OtherWhite;measures_Value |
| 378 | 294 | Sex_Females;Age_Age25to49;EthnicGroup_Mixed/multipleethnicgroup_Total;measures_Value |
| 379 | 295 | Sex_Females;Age_Age25to49;EthnicGroup_Mixed/multipleethnicgroup_WhiteandBlackCaribbean;measures_Value |
| 380 | 296 | Sex_Females;Age_Age25to49;EthnicGroup_Mixed/multipleethnicgroup_WhiteandBlackAfrican;measures_Value |
| 381 | 297 | Sex_Females;Age_Age25to49;EthnicGroup_Mixed/multipleethnicgroup_WhiteandAsian;measures_Value |
| 382 | 298 | Sex_Females;Age_Age25to49;EthnicGroup_Mixed/multipleethnicgroup_OtherMixed;measures_Value |
| 383 | 299 | Sex_Females;Age_Age25to49;EthnicGroup_Asian/AsianBritish_Total;measures_Value |
| 384 | 300 | Sex_Females;Age_Age25to49;EthnicGroup_Asian/AsianBritish_Indian;measures_Value |
| 385 | 301 | Sex_Females;Age_Age25to49;EthnicGroup_Asian/AsianBritish_Pakistani;measures_Value |
| 386 | 302 | Sex_Females;Age_Age25to49;EthnicGroup_Asian/AsianBritish_Bangladeshi;measures_Value |
| 387 | 303 | Sex_Females;Age_Age25to49;EthnicGroup_Asian/AsianBritish_Chinese;measures_Value |
| 388 | 304 | Sex_Females;Age_Age25to49;EthnicGroup_Asian/AsianBritish_OtherAsian;measures_Value |
| 389 | 305 | Sex_Females;Age_Age25to49;EthnicGroup_Black/African/Caribbean/BlackBritish_Total;measures_Value |
| 390 | 306 | Sex_Females;Age_Age25to49;EthnicGroup_Black/African/Caribbean/BlackBritish_African;measures_Value |
| 391 | 307 | Sex_Females;Age_Age25to49;EthnicGroup_Black/African/Caribbean/BlackBritish_Caribbean;measures_Value |
| 392 | 308 | Sex_Females;Age_Age25to49;EthnicGroup_Black/African/Caribbean/BlackBritish_OtherBlack;measures_Value |
| 393 | 309 | Sex_Females;Age_Age25to49;EthnicGroup_Otherethnicgroup_Total;measures_Value |
| 394 | 310 | Sex_Females;Age_Age25to49;EthnicGroup_Otherethnicgroup_Arab;measures_Value |
| 395 | 311 | Sex_Females;Age_Age25to49;EthnicGroup_Otherethnicgroup_Anyotherethnicgroup;measures_Value |
| 396 | 312 | Sex_Females;Age_Age50to64;EthnicGroup_Allcategories_Ethnicgroup;measures_Value |
| 397 | 313 | Sex_Females;Age_Age50to64;EthnicGroup_White_Total;measures_Value |
| 398 | 314 | Sex_Females;Age_Age50to64;EthnicGroup_White_English/Welsh/Scottish/NorthernIrish/British;measures_Value |
| 399 | 315 | Sex_Females;Age_Age50to64;EthnicGroup_White_Irish;measures_Value |
| 400 | 316 | Sex_Females;Age_Age50to64;EthnicGroup_White_GypsyorIrishTraveller;measures_Value |
| 401 | 317 | Sex_Females;Age_Age50to64;EthnicGroup_White_OtherWhite;measures_Value |
| 402 | 318 | Sex_Females;Age_Age50to64;EthnicGroup_Mixed/multipleethnicgroup_Total;measures_Value |
| 403 | 319 | Sex_Females;Age_Age50to64;EthnicGroup_Mixed/multipleethnicgroup_WhiteandBlackCaribbean;measures_Value |
| 404 | 320 | Sex_Females;Age_Age50to64;EthnicGroup_Mixed/multipleethnicgroup_WhiteandBlackAfrican;measures_Value |
| 405 | 321 | Sex_Females;Age_Age50to64;EthnicGroup_Mixed/multipleethnicgroup_WhiteandAsian;measures_Value |
| 406 | 322 | Sex_Females;Age_Age50to64;EthnicGroup_Mixed/multipleethnicgroup_OtherMixed;measures_Value |
| 407 | 323 | Sex_Females;Age_Age50to64;EthnicGroup_Asian/AsianBritish_Total;measures_Value |
| 408 | 324 | Sex_Females;Age_Age50to64;EthnicGroup_Asian/AsianBritish_Indian;measures_Value |
| 409 | 325 | Sex_Females;Age_Age50to64;EthnicGroup_Asian/AsianBritish_Pakistani;measures_Value |
| 410 | 326 | Sex_Females;Age_Age50to64;EthnicGroup_Asian/AsianBritish_Bangladeshi;measures_Value |
| 411 | 327 | Sex_Females;Age_Age50to64;EthnicGroup_Asian/AsianBritish_Chinese;measures_Value |

| | | |
|---|---|---|
| 412 | 328 | Sex_Females;Age_Age50to64;EthnicGroup_Asian/AsianBritish_OtherAsian;measures_Value |
| 413 | 329 | Sex_Females;Age_Age50to64;EthnicGroup_Black/African/Caribbean/BlackBritish_Total;measures_Value |
| 414 | 330 | Sex_Females;Age_Age50to64;EthnicGroup_Black/African/Caribbean/BlackBritish_African;measures_Value |
| 415 | 331 | Sex_Females;Age_Age50to64;EthnicGroup_Black/African/Caribbean/BlackBritish_Caribbean;measures_Value |
| 416 | 332 | Sex_Females;Age_Age50to64;EthnicGroup_Black/African/Caribbean/BlackBritish_OtherBlack;measures_Value |
| 417 | 333 | Sex_Females;Age_Age50to64;EthnicGroup_Otherethnicgroup_Total;measures_Value |
| 418 | 334 | Sex_Females;Age_Age50to64;EthnicGroup_Otherethnicgroup_Arab;measures_Value |
| 419 | 335 | Sex_Females;Age_Age50to64;EthnicGroup_Otherethnicgroup_Anyotherethnicgroup;measures_Value |
| 420 | 336 | Sex_Females;Age_Age65andover;EthnicGroup_Allcategories_Ethnicgroup;measures_Value |
| 421 | 337 | Sex_Females;Age_Age65andover;EthnicGroup_White_Total;measures_Value |
| 422 | 338 | Sex_Females;Age_Age65andover;EthnicGroup_White_English/Welsh/Scottish/NorthernIrish/British;measures_Value |
| 423 | 339 | Sex_Females;Age_Age65andover;EthnicGroup_White_Irish;measures_Value |
| 424 | 340 | Sex_Females;Age_Age65andover;EthnicGroup_White_GypsyorIrishTraveller;measures_Value |
| 425 | 341 | Sex_Females;Age_Age65andover;EthnicGroup_White_OtherWhite;measures_Value |
| 426 | 342 | Sex_Females;Age_Age65andover;EthnicGroup_Mixed/multipleethnicgroup_Total;measures_Value |
| 427 | 343 | Sex_Females;Age_Age65andover;EthnicGroup_Mixed/multipleethnicgroup_WhiteandBlackCaribbean;measures_Value |
| 428 | 344 | Sex_Females;Age_Age65andover;EthnicGroup_Mixed/multipleethnicgroup_WhiteandBlackAfrican;measures_Value |
| 429 | 345 | Sex_Females;Age_Age65andover;EthnicGroup_Mixed/multipleethnicgroup_WhiteandAsian;measures_Value |
| 430 | 346 | Sex_Females;Age_Age65andover;EthnicGroup_Mixed/multipleethnicgroup_OtherMixed;measures_Value |
| 431 | 347 | Sex_Females;Age_Age65andover;EthnicGroup_Asian/AsianBritish_Total;measures_Value |
| 432 | 348 | Sex_Females;Age_Age65andover;EthnicGroup_Asian/AsianBritish_Indian;measures_Value |
| 433 | 349 | Sex_Females;Age_Age65andover;EthnicGroup_Asian/AsianBritish_Pakistani;measures_Value |
| 434 | 350 | Sex_Females;Age_Age65andover;EthnicGroup_Asian/AsianBritish_Bangladeshi;measures_Value |
| 435 | 351 | Sex_Females;Age_Age65andover;EthnicGroup_Asian/AsianBritish_Chinese;measures_Value |
| 436 | 352 | Sex_Females;Age_Age65andover;EthnicGroup_Asian/AsianBritish_OtherAsian;measures_Value |
| 437 | 353 | Sex_Females;Age_Age65andover;EthnicGroup_Black/African/Caribbean/BlackBritish_Total;measures_Value |
| 438 | 354 | Sex_Females;Age_Age65andover;EthnicGroup_Black/African/Caribbean/BlackBritish_African;measures_Value |
| 439 | 355 | Sex_Females;Age_Age65andover;EthnicGroup_Black/African/Caribbean/BlackBritish_Caribbean;measures_Value |
| 440 | 356 | Sex_Females;Age_Age65andover;EthnicGroup_Black/African/Caribbean/BlackBritish_OtherBlack;measures_Value |
| 441 | 357 | Sex_Females;Age_Age65andover;EthnicGroup_Otherethnicgroup_Total;measures_Value |
| 442 | 358 | Sex_Females;Age_Age65andover;EthnicGroup_Otherethnicgroup_Arab;measures_Value |
| 443 | 359 | Sex_Females;Age_Age65andover;EthnicGroup_Otherethnicgroup_Anyotherethnicgroup;measures_Value |
| 444 | 360 | Familystatus_Allcategories_Familystatusbynumberofparentsworking;Economicactivity_Economicallyactive_Total;measures_Value |
| 445 | 361 | Familystatus_Allcategories_Familystatusbynumberofparentsworking;Economicactivity_Economicallyactive_Inemployment_Total;measures_Value |
| 446 | 362 | Familystatus_Allcategories_Familystatusbynumberofparentsworking;Economicactivity_Economicallyactive_Inemployment_Employee(includingfull-timestudents);measures_Value |
| 447 | 363 | Familystatus_Allcategories_Familystatusbynumberofparentsworking;Economicactivity_Economicallyactive_Inemployment_Self-employed(includingfull-timestudents);measures_Value |
| 448 | 364 | Familystatus_Allcategories_Familystatusbynumberofparentsworking;Economicactivity_Economicallyactive_Unemployed(includingfull-timestudents);measures_Value |
| 449 | 365 | Familystatus_Allcategories_Familystatusbynumberofparentsworking;Economicactivity_Economicallyinactive_Total;measures_Value |

| | | |
|---|---|---|
| 450 | 366 | Familystatus_Allcategories_Familystatusbynumberofparentsworking;Economicactivity_Economicallyinactive_Retired;measures_Value |
| 451 | 367 | Familystatus_Allcategories_Familystatusbynumberofparentsworking;Economicactivity_Economicallyinactive_Student(includingfull-timestudents);measures_Value |
| 452 | 368 | Familystatus_Allcategories_Familystatusbynumberofparentsworking;Economicactivity_Economicallyinactive_Lookingafterhomeorfamily;measures_Value |
| 453 | 369 | Familystatus_Allcategories_Familystatusbynumberofparentsworking;Economicactivity_Economicallyinactive_Long-termsickordisabled;measures_Value |
| 454 | 370 | Familystatus_Allcategories_Familystatusbynumberofparentsworking;Economicactivity_Economicallyinactive_Other;measures_Value |
| 455 | 371 | Familystatus_Couplefamily_Total;Economicactivity_Allcategories_Economicactivity;measures_Value |
| 456 | 372 | Familystatus_Couplefamily_Total;Economicactivity_Economicallyactive_Total;measures_Value |
| 457 | 373 | Familystatus_Couplefamily_Total;Economicactivity_Economicallyactive_Inemployment_Total;measures_Value |
| 458 | 374 | Familystatus_Couplefamily_Total;Economicactivity_Economicallyactive_Inemployment_Employee(includingfull-timestudents);measures_Value |
| 459 | 375 | Familystatus_Couplefamily_Total;Economicactivity_Economicallyactive_Inemployment_Self-employed(includingfull-timestudents);measures_Value |
| 460 | 376 | Familystatus_Couplefamily_Total;Economicactivity_Economicallyactive_Unemployed(includingfull-timestudents);measures_Value |
| 461 | 377 | Familystatus_Couplefamily_Total;Economicactivity_Economicallyinactive_Total;measures_Value |
| 462 | 378 | Familystatus_Couplefamily_Total;Economicactivity_Economicallyinactive_Retired;measures_Value |
| 463 | 379 | Familystatus_Couplefamily_Total;Economicactivity_Economicallyinactive_Student(includingfull-timestudents);measures_Value |
| 464 | 380 | Familystatus_Couplefamily_Total;Economicactivity_Economicallyinactive_Lookingafterhomeorfamily;measures_Value |
| 465 | 381 | Familystatus_Couplefamily_Total;Economicactivity_Economicallyinactive_Long-termsickordisabled;measures_Value |
| 466 | 382 | Familystatus_Couplefamily_Total;Economicactivity_Economicallyinactive_Other;measures_Value |
| 467 | 383 | Familystatus_Couplefamily_Bothparentsworking;Economicactivity_Allcategories_Economicactivity;measures_Value |
| 468 | 384 | Familystatus_Couplefamily_Bothparentsworking;Economicactivity_Economicallyactive_Total;measures_Value |
| 469 | 385 | Familystatus_Couplefamily_Bothparentsworking;Economicactivity_Economicallyactive_Inemployment_Total;measures_Value |
| 470 | 386 | Familystatus_Couplefamily_Bothparentsworking;Economicactivity_Economicallyactive_Inemployment_Employee(includingfull-timestudents);measures_Value |
| 471 | 387 | Familystatus_Couplefamily_Bothparentsworking;Economicactivity_Economicallyactive_Inemployment_Self-employed(includingfull-timestudents);measures_Value |
| 472 | 388 | Familystatus_Couplefamily_Bothparentsworking;Economicactivity_Economicallyactive_Unemployed(includingfull-timestudents);measures_Value |
| 473 | 389 | Familystatus_Couplefamily_Bothparentsworking;Economicactivity_Economicallyinactive_Total;measures_Value |
| 474 | 390 | Familystatus_Couplefamily_Bothparentsworking;Economicactivity_Economicallyinactive_Retired;measures_Value |
| 475 | 391 | Familystatus_Couplefamily_Bothparentsworking;Economicactivity_Economicallyinactive_Student(includingfull-timestudents);measures_Value |
| 476 | 392 | Familystatus_Couplefamily_Bothparentsworking;Economicactivity_Economicallyinactive_Lookingafterhomeorfamily;measures_Value |
| 477 | 393 | Familystatus_Couplefamily_Bothparentsworking;Economicactivity_Economicallyinactive_Long-termsickordisabled;measures_Value |
| 478 | 394 | Familystatus_Couplefamily_Bothparentsworking;Economicactivity_Economicallyinactive_Other;measures_Value |
| 479 | 395 | Familystatus_Couplefamily_Oneparentworking;Economicactivity_Allcategories_Economicactivity;measures_Value |
| 480 | 396 | Familystatus_Couplefamily_Oneparentworking;Economicactivity_Economicallyactive_Total;measures_Value |
| 481 | 397 | Familystatus_Couplefamily_Oneparentworking;Economicactivity_Economicallyactive_Inemployment_Total;measures_Value |
| 482 | 398 | Familystatus_Couplefamily_Oneparentworking;Economicactivity_Economicallyactive_Inemployment_Employee(includingfull-timestudents);measures_Value |
| 483 | 399 | Familystatus_Couplefamily_Oneparentworking;Economicactivity_Economicallyactive_Inemployment_Self-employed(includingfull-timestudents);measures_Value |
| 484 | 400 | Familystatus_Couplefamily_Oneparentworking;Economicactivity_Economicallyactive_Unemployed(includingfull-timestudents);measures_Value |
| 485 | 401 | Familystatus_Couplefamily_Oneparentworking;Economicactivity_Economicallyinactive_Total;measures_Value |
| 486 | 402 | Familystatus_Couplefamily_Oneparentworking;Economicactivity_Economicallyinactive_Retired;measures_Value |
| 487 | 403 | Familystatus_Couplefamily_Oneparentworking;Economicactivity_Economicallyinactive_Student(includingfull-timestudents);measures_Value |

| | | |
|---|---|---|
| 488 | 404 | Familystatus_Couplefamily_Oneparentworking;Economicactivity_Economicallyinactive_Lookingafterhomeorfamily;measures_Value |
| 489 | 405 | Familystatus_Couplefamily_Oneparentworking;Economicactivity_Economicallyinactive_Long-termsickordisabled;measures_Value |
| 490 | 406 | Familystatus_Couplefamily_Oneparentworking;Economicactivity_Economicallyinactive_Other;measures_Value |
| 491 | 407 | Familystatus_Couplefamily_Noparentsworking;Economicactivity_Allcategories_Economicactivity;measures_Value |
| 492 | 408 | Familystatus_Couplefamily_Noparentsworking;Economicactivity_Economicallyactive_Total;measures_Value |
| 493 | 409 | Familystatus_Couplefamily_Noparentsworking;Economicactivity_Economicallyactive_Inemployment_Total;measures_Value |
| 494 | 410 | Familystatus_Couplefamily_Noparentsworking;Economicactivity_Economicallyactive_Inemployment_Employee(includingfull-timestudents);measures_Value |
| 495 | 411 | Familystatus_Couplefamily_Noparentsworking;Economicactivity_Economicallyactive_Inemployment_Self-employed(includingfull-timestudents);measures_Value |
| 496 | 412 | Familystatus_Couplefamily_Noparentsworking;Economicactivity_Economicallyactive_Unemployed(includingfull-timestudents);measures_Value |
| 497 | 413 | Familystatus_Couplefamily_Noparentsworking;Economicactivity_Economicallyinactive_Total;measures_Value |
| 498 | 414 | Familystatus_Couplefamily_Noparentsworking;Economicactivity_Economicallyinactive_Retired;measures_Value |
| 499 | 415 | Familystatus_Couplefamily_Noparentsworking;Economicactivity_Economicallyinactive_Student(includingfull-timestudents);measures_Value |
| 500 | 416 | Familystatus_Couplefamily_Noparentsworking;Economicactivity_Economicallyinactive_Lookingafterhomeorfamily;measures_Value |
| 501 | 417 | Familystatus_Couplefamily_Noparentsworking;Economicactivity_Economicallyinactive_Long-termsickordisabled;measures_Value |
| 502 | 418 | Familystatus_Couplefamily_Noparentsworking;Economicactivity_Economicallyinactive_Other;measures_Value |
| 503 | 419 | Familystatus_Loneparentfamily_Total;Economicactivity_Allcategories_Economicactivity;measures_Value |
| 504 | 420 | Familystatus_Loneparentfamily_Total;Economicactivity_Economicallyactive_Total;measures_Value |
| 505 | 421 | Familystatus_Loneparentfamily_Total;Economicactivity_Economicallyactive_Inemployment_Total;measures_Value |
| 506 | 422 | Familystatus_Loneparentfamily_Total;Economicactivity_Economicallyactive_Inemployment_Employee(includingfull-timestudents);measures_Value |
| 507 | 423 | Familystatus_Loneparentfamily_Total;Economicactivity_Economicallyactive_Inemployment_Self-employed(includingfull-timestudents);measures_Value |
| 508 | 424 | Familystatus_Loneparentfamily_Total;Economicactivity_Economicallyactive_Unemployed(includingfull-timestudents);measures_Value |
| 509 | 425 | Familystatus_Loneparentfamily_Total;Economicactivity_Economicallyinactive_Total;measures_Value |
| 510 | 426 | Familystatus_Loneparentfamily_Total;Economicactivity_Economicallyinactive_Retired;measures_Value |
| 511 | 427 | Familystatus_Loneparentfamily_Total;Economicactivity_Economicallyinactive_Student(includingfull-timestudents);measures_Value |
| 512 | 428 | Familystatus_Loneparentfamily_Total;Economicactivity_Economicallyinactive_Lookingafterhomeorfamily;measures_Value |
| 513 | 429 | Familystatus_Loneparentfamily_Total;Economicactivity_Economicallyinactive_Long-termsickordisabled;measures_Value |
| 514 | 430 | Familystatus_Loneparentfamily_Total;Economicactivity_Economicallyinactive_Other;measures_Value |
| 515 | 431 | Familystatus_Loneparentfamily_Parentworking;Economicactivity_Allcategories_Economicactivity;measures_Value |
| 516 | 432 | Familystatus_Loneparentfamily_Parentworking;Economicactivity_Economicallyactive_Total;measures_Value |
| 517 | 433 | Familystatus_Loneparentfamily_Parentworking;Economicactivity_Economicallyactive_Inemployment_Total;measures_Value |
| 518 | 434 | Familystatus_Loneparentfamily_Parentworking;Economicactivity_Economicallyactive_Inemployment_Employee(includingfull-timestudents);measures_Value |
| 519 | 435 | Familystatus_Loneparentfamily_Parentworking;Economicactivity_Economicallyactive_Inemployment_Self-employed(includingfull-timestudents);measures_Value |
| 520 | 436 | Familystatus_Loneparentfamily_Parentworking;Economicactivity_Economicallyactive_Unemployed(includingfull-timestudents);measures_Value |
| 521 | 437 | Familystatus_Loneparentfamily_Parentworking;Economicactivity_Economicallyinactive_Total;measures_Value |
| 522 | 438 | Familystatus_Loneparentfamily_Parentworking;Economicactivity_Economicallyinactive_Retired;measures_Value |
| 523 | 439 | Familystatus_Loneparentfamily_Parentworking;Economicactivity_Economicallyinactive_Student(includingfull-timestudents);measures_Value |
| 524 | 440 | Familystatus_Loneparentfamily_Parentworking;Economicactivity_Economicallyinactive_Lookingafterhomeorfamily;measures_Value |
| 525 | 441 | Familystatus_Loneparentfamily_Parentworking;Economicactivity_Economicallyinactive_Long-termsickordisabled;measures_Value |

| | | |
|---|---|---|
| 526 | 442 | Familystatus_Loneparentfamily_Parentworking;Economicactivity_Economicallyinactive_Other;measures_Value |
| 527 | 443 | Familystatus_Loneparentfamily_Parentnotworking;Economicactivity_Allcategories_Economicactivity;measures_Value |
| 528 | 444 | Familystatus_Loneparentfamily_Parentnotworking;Economicactivity_Economicallyactive_Total;measures_Value |
| 529 | 445 | Familystatus_Loneparentfamily_Parentnotworking;Economicactivity_Economicallyactive_Inemployment_Total;measures_Value |
| 530 | 446 | Familystatus_Loneparentfamily_Parentnotworking;Economicactivity_Economicallyactive_Inemployment_Employee(includingfull-timestudents);measures_Value |
| 531 | 447 | Familystatus_Loneparentfamily_Parentnotworking;Economicactivity_Economicallyactive_Inemployment_Self-employed(includingfull-timestudents);measures_Value |
| 532 | 448 | Familystatus_Loneparentfamily_Parentnotworking;Economicactivity_Economicallyactive_Unemployed(includingfull-timestudents);measures_Value |
| 533 | 449 | Familystatus_Loneparentfamily_Parentnotworking;Economicactivity_Economicallyinactive_Total;measures_Value |
| 534 | 450 | Familystatus_Loneparentfamily_Parentnotworking;Economicactivity_Economicallyinactive_Retired;measures_Value |
| 535 | 451 | Familystatus_Loneparentfamily_Parentnotworking;Economicactivity_Economicallyinactive_Student(includingfull-timestudents);measures_Value |
| 536 | 452 | Familystatus_Loneparentfamily_Parentnotworking;Economicactivity_Economicallyinactive_Lookingafterhomeorfamily;measures_Value |
| 537 | 453 | Familystatus_Loneparentfamily_Parentnotworking;Economicactivity_Economicallyinactive_Long-termsickordisabled;measures_Value |
| 538 | 454 | Familystatus_Loneparentfamily_Parentnotworking;Economicactivity_Economicallyinactive_Other;measures_Value |
| 539 | 455 | disability/health/care_Allcategories_Long-termhealthproblemordisability;measures_Value |
| 540 | 456 | disability/health/care_Allcategories_Long-termhealthproblemordisability;measures_Percent |
| 541 | 457 | disability/health/care_Day-to-dayactivitieslimitedalot;measures_Value |
| 542 | 458 | disability/health/care_Day-to-dayactivitieslimitedalot;measures_Percent |
| 543 | 459 | disability/health/care_Day-to-dayactivitieslimitedalittle;measures_Value |
| 544 | 460 | disability/health/care_Day-to-dayactivitieslimitedalittle;measures_Percent |
| 545 | 461 | disability/health/care_Day-to-dayactivitiesnotlimited;measures_Value |
| 546 | 462 | disability/health/care_Day-to-dayactivitiesnotlimited;measures_Percent |
| 547 | 463 | disability/health/care_Day-to-dayactivitieslimitedalot_Age16to64;measures_Value |
| 548 | 464 | disability/health/care_Day-to-dayactivitieslimitedalot_Age16to64;measures_Percent |
| 549 | 465 | disability/health/care_Day-to-dayactivitieslimitedalittle_Age16to64;measures_Value |
| 550 | 466 | disability/health/care_Day-to-dayactivitieslimitedalittle_Age16to64;measures_Percent |
| 551 | 467 | disability/health/care_Day-to-dayactivitiesnotlimited_Age16to64;measures_Value |
| 552 | 468 | disability/health/care_Day-to-dayactivitiesnotlimited_Age16to64;measures_Percent |
| 553 | 469 | disability/health/care_Verygoodhealth;measures_Value |
| 554 | 470 | disability/health/care_Verygoodhealth;measures_Percent |
| 555 | 471 | disability/health/care_Goodhealth;measures_Value |
| 556 | 472 | disability/health/care_Goodhealth;measures_Percent |
| 557 | 473 | disability/health/care_Fairhealth;measures_Value |
| 558 | 474 | disability/health/care_Fairhealth;measures_Percent |
| 559 | 475 | disability/health/care_Badhealth;measures_Value |
| 560 | 476 | disability/health/care_Badhealth;measures_Percent |
| 561 | 477 | disability/health/care_Verybadhealth;measures_Value |
| 562 | 478 | disability/health/care_Verybadhealth;measures_Percent |
| 563 | 479 | disability/health/care_Providesnounpaidcare;measures_Value |

| | | |
|---|---|---|
| 564 | 480 | disability/health/care_Providesnounpaidcare;measures_Percent |
| 565 | 481 | disability/health/care_Provides1to19hoursunpaidcareaweek;measures_Value |
| 566 | 482 | disability/health/care_Provides1to19hoursunpaidcareaweek;measures_Percent |
| 567 | 483 | disability/health/care_Provides20to49hoursunpaidcareaweek;measures_Value |
| 568 | 484 | disability/health/care_Provides20to49hoursunpaidcareaweek;measures_Percent |
| 569 | 485 | disability/health/care_Provides50ormorehoursunpaidcareaweek;measures_Value |
| 570 | 486 | disability/health/care_Provides50ormorehoursunpaidcareaweek;measures_Percent |
| 571 | 487 | HouseholdComposition_Allcategories_Householdcomposition;EthnicGroupofHRP_White_Total;measures_Value |
| 572 | 488 | HouseholdComposition_Allcategories_Householdcomposition;EthnicGroupofHRP_White_English/Welsh/Scottish/NorthernIrish/British;measures_Value |
| 573 | 489 | HouseholdComposition_Allcategories_Householdcomposition;EthnicGroupofHRP_White_Irish;measures_Value |
| 574 | 490 | HouseholdComposition_Allcategories_Householdcomposition;EthnicGroupofHRP_White_OtherWhite;measures_Value |
| 575 | 491 | HouseholdComposition_Allcategories_Householdcomposition;EthnicGroupofHRP_Mixed/multipleethnicgroup;measures_Value |
| 576 | 492 | HouseholdComposition_Allcategories_Householdcomposition;EthnicGroupofHRP_Asian/AsianBritish;measures_Value |
| 577 | 493 | HouseholdComposition_Allcategories_Householdcomposition;EthnicGroupofHRP_Black/African/Caribbean/BlackBritish;measures_Value |
| 578 | 494 | HouseholdComposition_Allcategories_Householdcomposition;EthnicGroupofHRP_Otherethnicgroup;measures_Value |
| 579 | 495 | HouseholdComposition_Onepersonhousehold;EthnicGroupofHRP_Allcategories_EthnicgroupofHRP;measures_Value |
| 580 | 496 | HouseholdComposition_Onepersonhousehold;EthnicGroupofHRP_White_Total;measures_Value |
| 581 | 497 | HouseholdComposition_Onepersonhousehold;EthnicGroupofHRP_White_English/Welsh/Scottish/NorthernIrish/British;measures_Value |
| 582 | 498 | HouseholdComposition_Onepersonhousehold;EthnicGroupofHRP_White_Irish;measures_Value |
| 583 | 499 | HouseholdComposition_Onepersonhousehold;EthnicGroupofHRP_White_OtherWhite;measures_Value |
| 584 | 500 | HouseholdComposition_Onepersonhousehold;EthnicGroupofHRP_Mixed/multipleethnicgroup;measures_Value |
| 585 | 501 | HouseholdComposition_Onepersonhousehold;EthnicGroupofHRP_Asian/AsianBritish;measures_Value |
| 586 | 502 | HouseholdComposition_Onepersonhousehold;EthnicGroupofHRP_Black/African/Caribbean/BlackBritish;measures_Value |
| 587 | 503 | HouseholdComposition_Onepersonhousehold;EthnicGroupofHRP_Otherethnicgroup;measures_Value |
| 588 | 504 | HouseholdComposition_Onefamilyonly_Total;EthnicGroupofHRP_Allcategories_EthnicgroupofHRP;measures_Value |
| 589 | 505 | HouseholdComposition_Onefamilyonly_Total;EthnicGroupofHRP_White_Total;measures_Value |
| 590 | 506 | HouseholdComposition_Onefamilyonly_Total;EthnicGroupofHRP_White_English/Welsh/Scottish/NorthernIrish/British;measures_Value |
| 591 | 507 | HouseholdComposition_Onefamilyonly_Total;EthnicGroupofHRP_White_Irish;measures_Value |
| 592 | 508 | HouseholdComposition_Onefamilyonly_Total;EthnicGroupofHRP_White_OtherWhite;measures_Value |
| 593 | 509 | HouseholdComposition_Onefamilyonly_Total;EthnicGroupofHRP_Mixed/multipleethnicgroup;measures_Value |
| 594 | 510 | HouseholdComposition_Onefamilyonly_Total;EthnicGroupofHRP_Asian/AsianBritish;measures_Value |
| 595 | 511 | HouseholdComposition_Onefamilyonly_Total;EthnicGroupofHRP_Black/African/Caribbean/BlackBritish;measures_Value |
| 596 | 512 | HouseholdComposition_Onefamilyonly_Total;EthnicGroupofHRP_Otherethnicgroup;measures_Value |
| 597 | 513 | HouseholdComposition_Onefamilyonly_Allaged65andover;EthnicGroupofHRP_Allcategories_EthnicgroupofHRP;measures_Value |
| 598 | 514 | HouseholdComposition_Onefamilyonly_Allaged65andover;EthnicGroupofHRP_White_Total;measures_Value |
| 599 | 515 | HouseholdComposition_Onefamilyonly_Allaged65andover;EthnicGroupofHRP_White_English/Welsh/Scottish/NorthernIrish/British;measures_Value |
| 600 | 516 | HouseholdComposition_Onefamilyonly_Allaged65andover;EthnicGroupofHRP_White_Irish;measures_Value |
| 601 | 517 | HouseholdComposition_Onefamilyonly_Allaged65andover;EthnicGroupofHRP_White_OtherWhite;measures_Value |

| | | |
|---|---|---|
| 602 | 518 | HouseholdComposition_Onefamilyonly_Allaged65andover;EthnicGroupofHRP_Mixed/multipleethnicgroup;measures_Value |
| 603 | 519 | HouseholdComposition_Onefamilyonly_Allaged65andover;EthnicGroupofHRP_Asian/AsianBritish;measures_Value |
| 604 | 520 | HouseholdComposition_Onefamilyonly_Allaged65andover;EthnicGroupofHRP_Black/African/Caribbean/BlackBritish;measures_Value |
| 605 | 521 | HouseholdComposition_Onefamilyonly_Allaged65andover;EthnicGroupofHRP_Otherethnicgroup;measures_Value |
| 606 | 522 | HouseholdComposition_Onefamilyonly_Married,same-sexcivilpartnershiporcohabitingcouple;EthnicGroupofHRP_Allcategories_EthnicgroupofHRP;measures_Value |
| 607 | 523 | HouseholdComposition_Onefamilyonly_Married,same-sexcivilpartnershiporcohabitingcouple;EthnicGroupofHRP_White_Total;measures_Value |
| 608 | 524 | HouseholdComposition_Onefamilyonly_Married,same-sexcivilpartnershiporcohabitingcouple;EthnicGroupofHRP_White_English/Welsh/Scottish/NorthernIrish/British;measures_Value |
| 609 | 525 | HouseholdComposition_Onefamilyonly_Married,same-sexcivilpartnershiporcohabitingcouple;EthnicGroupofHRP_White_Irish;measures_Value |
| 610 | 526 | HouseholdComposition_Onefamilyonly_Married,same-sexcivilpartnershiporcohabitingcouple;EthnicGroupofHRP_White_OtherWhite;measures_Value |
| 611 | 527 | HouseholdComposition_Onefamilyonly_Married,same-sexcivilpartnershiporcohabitingcouple;EthnicGroupofHRP_Mixed/multipleethnicgroup;measures_Value |
| 612 | 528 | HouseholdComposition_Onefamilyonly_Married,same-sexcivilpartnershiporcohabitingcouple;EthnicGroupofHRP_Asian/AsianBritish;measures_Value |
| 613 | 529 | HouseholdComposition_Onefamilyonly_Married,same-sexcivilpartnershiporcohabitingcouple;EthnicGroupofHRP_Black/African/Caribbean/BlackBritish;measures_Value |
| 614 | 530 | HouseholdComposition_Onefamilyonly_Married,same-sexcivilpartnershiporcohabitingcouple;EthnicGroupofHRP_Otherethnicgroup;measures_Value |
| 615 | 531 | HouseholdComposition_Onefamilyonly_Loneparent;EthnicGroupofHRP_Allcategories_EthnicgroupofHRP;measures_Value |
| 616 | 532 | HouseholdComposition_Onefamilyonly_Loneparent;EthnicGroupofHRP_White_Total;measures_Value |
| 617 | 533 | HouseholdComposition_Onefamilyonly_Loneparent;EthnicGroupofHRP_White_English/Welsh/Scottish/NorthernIrish/British;measures_Value |
| 618 | 534 | HouseholdComposition_Onefamilyonly_Loneparent;EthnicGroupofHRP_White_Irish;measures_Value |
| 619 | 535 | HouseholdComposition_Onefamilyonly_Loneparent;EthnicGroupofHRP_White_OtherWhite;measures_Value |
| 620 | 536 | HouseholdComposition_Onefamilyonly_Loneparent;EthnicGroupofHRP_Mixed/multipleethnicgroup;measures_Value |
| 621 | 537 | HouseholdComposition_Onefamilyonly_Loneparent;EthnicGroupofHRP_Asian/AsianBritish;measures_Value |
| 622 | 538 | HouseholdComposition_Onefamilyonly_Loneparent;EthnicGroupofHRP_Black/African/Caribbean/BlackBritish;measures_Value |
| 623 | 539 | HouseholdComposition_Onefamilyonly_Loneparent;EthnicGroupofHRP_Otherethnicgroup;measures_Value |
| 624 | 540 | HouseholdComposition_Otherhouseholdtypes;EthnicGroupofHRP_Allcategories_EthnicgroupofHRP;measures_Value |
| 625 | 541 | HouseholdComposition_Otherhouseholdtypes;EthnicGroupofHRP_White_Total;measures_Value |
| 626 | 542 | HouseholdComposition_Otherhouseholdtypes;EthnicGroupofHRP_White_English/Welsh/Scottish/NorthernIrish/British;measures_Value |
| 627 | 543 | HouseholdComposition_Otherhouseholdtypes;EthnicGroupofHRP_White_Irish;measures_Value |
| 628 | 544 | HouseholdComposition_Otherhouseholdtypes;EthnicGroupofHRP_White_OtherWhite;measures_Value |
| 629 | 545 | HouseholdComposition_Otherhouseholdtypes;EthnicGroupofHRP_Mixed/multipleethnicgroup;measures_Value |
| 630 | 546 | HouseholdComposition_Otherhouseholdtypes;EthnicGroupofHRP_Asian/AsianBritish;measures_Value |
| 631 | 547 | HouseholdComposition_Otherhouseholdtypes;EthnicGroupofHRP_Black/African/Caribbean/BlackBritish;measures_Value |
| 632 | 548 | HouseholdComposition_Otherhouseholdtypes;EthnicGroupofHRP_Otherethnicgroup;measures_Value |
| 633 | 549 | Age_Allcategories_Age;livingarrangements_Livinginacouple_Total;measures_Value |
| 634 | 550 | Age_Allcategories_Age;livingarrangements_Livinginacouple_Marriedorinaregisteredsame-sexcivilpartnership;measures_Value |
| 635 | 551 | Age_Allcategories_Age;livingarrangements_Livinginacouple_Cohabiting;measures_Value |
| 636 | 552 | Age_Allcategories_Age;livingarrangements_Notlivinginacouple_Total;measures_Value |
| 637 | 553 | Age_Allcategories_Age;livingarrangements_Notlivinginacouple_Single(nevermarriedorneverregisteredasame-sexcivilpartnership);measures_Value |
| 638 | 554 | Age_Allcategories_Age;livingarrangements_Notlivinginacouple_Marriedorinaregisteredsame-sexcivilpartnership;measures_Value |
| 639 | 555 | Age_Allcategories_Age;livingarrangements_Notlivinginacouple_Separated(butstilllegallymarriedorstilllegallyinasame-sexcivilpartnership);measures_Value |

| | | |
|---|---|---|
| 640 | 556 | Age_Allcategories_Age;livingarrangements_Notlivinginacouple_Divorcedorformerlyinasame-sexcivilpartnershipwhichisnowlegallydissolved;measures_Value |
| 641 | 557 | Age_Allcategories_Age;livingarrangements_Notlivinginacouple_Widowedorsurvivingpartnerfromasame-sexcivilpartnership;measures_Value |
| 642 | 558 | Age_Age24andunder;livingarrangements_Allcategories_Livingarrangements;measures_Value |
| 643 | 559 | Age_Age24andunder;livingarrangements_Livinginacouple_Total;measures_Value |
| 644 | 560 | Age_Age24andunder;livingarrangements_Livinginacouple_Marriedorinaregisteredsame-sexcivilpartnership;measures_Value |
| 645 | 561 | Age_Age24andunder;livingarrangements_Livinginacouple_Cohabiting;measures_Value |
| 646 | 562 | Age_Age24andunder;livingarrangements_Notlivinginacouple_Total;measures_Value |
| 647 | 563 | Age_Age24andunder;livingarrangements_Notlivinginacouple_Single(nevermarriedorneverregisteredasame-sexcivilpartnership);measures_Value |
| 648 | 564 | Age_Age24andunder;livingarrangements_Notlivinginacouple_Marriedorinaregisteredsame-sexcivilpartnership;measures_Value |
| 649 | 565 | Age_Age24andunder;livingarrangements_Notlivinginacouple_Separated(butstilllegallymarriedorstilllegallyinasame-sexcivilpartnership);measures_Value |
| 650 | 566 | Age_Age24andunder;livingarrangements_Notlivinginacouple_Divorcedorformerlyinasame-sexcivilpartnershipwhichisnowlegallydissolved;measures_Value |
| 651 | 567 | Age_Age24andunder;livingarrangements_Notlivinginacouple_Widowedorsurvivingpartnerfromasame-sexcivilpartnership;measures_Value |
| 652 | 568 | Age_Age25to34;livingarrangements_Allcategories_Livingarrangements;measures_Value |
| 653 | 569 | Age_Age25to34;livingarrangements_Livinginacouple_Total;measures_Value |
| 654 | 570 | Age_Age25to34;livingarrangements_Livinginacouple_Marriedorinaregisteredsame-sexcivilpartnership;measures_Value |
| 655 | 571 | Age_Age25to34;livingarrangements_Livinginacouple_Cohabiting;measures_Value |
| 656 | 572 | Age_Age25to34;livingarrangements_Notlivinginacouple_Total;measures_Value |
| 657 | 573 | Age_Age25to34;livingarrangements_Notlivinginacouple_Single(nevermarriedorneverregisteredasame-sexcivilpartnership);measures_Value |
| 658 | 574 | Age_Age25to34;livingarrangements_Notlivinginacouple_Marriedorinaregisteredsame-sexcivilpartnership;measures_Value |
| 659 | 575 | Age_Age25to34;livingarrangements_Notlivinginacouple_Separated(butstilllegallymarriedorstilllegallyinasame-sexcivilpartnership);measures_Value |
| 660 | 576 | Age_Age25to34;livingarrangements_Notlivinginacouple_Divorcedorformerlyinasame-sexcivilpartnershipwhichisnowlegallydissolved;measures_Value |
| 661 | 577 | Age_Age25to34;livingarrangements_Notlivinginacouple_Widowedorsurvivingpartnerfromasame-sexcivilpartnership;measures_Value |
| 662 | 578 | Age_Age35to49;livingarrangements_Allcategories_Livingarrangements;measures_Value |
| 663 | 579 | Age_Age35to49;livingarrangements_Livinginacouple_Total;measures_Value |
| 664 | 580 | Age_Age35to49;livingarrangements_Livinginacouple_Marriedorinaregisteredsame-sexcivilpartnership;measures_Value |
| 665 | 581 | Age_Age35to49;livingarrangements_Livinginacouple_Cohabiting;measures_Value |
| 666 | 582 | Age_Age35to49;livingarrangements_Notlivinginacouple_Total;measures_Value |
| 667 | 583 | Age_Age35to49;livingarrangements_Notlivinginacouple_Single(nevermarriedorneverregisteredasame-sexcivilpartnership);measures_Value |
| 668 | 584 | Age_Age35to49;livingarrangements_Notlivinginacouple_Marriedorinaregisteredsame-sexcivilpartnership;measures_Value |
| 669 | 585 | Age_Age35to49;livingarrangements_Notlivinginacouple_Separated(butstilllegallymarriedorstilllegallyinasame-sexcivilpartnership);measures_Value |
| 670 | 586 | Age_Age35to49;livingarrangements_Notlivinginacouple_Divorcedorformerlyinasame-sexcivilpartnershipwhichisnowlegallydissolved;measures_Value |
| 671 | 587 | Age_Age35to49;livingarrangements_Notlivinginacouple_Widowedorsurvivingpartnerfromasame-sexcivilpartnership;measures_Value |
| 672 | 588 | Age_Age50to64;livingarrangements_Allcategories_Livingarrangements;measures_Value |
| 673 | 589 | Age_Age50to64;livingarrangements_Livinginacouple_Total;measures_Value |
| 674 | 590 | Age_Age50to64;livingarrangements_Livinginacouple_Marriedorinaregisteredsame-sexcivilpartnership;measures_Value |
| 675 | 591 | Age_Age50to64;livingarrangements_Livinginacouple_Cohabiting;measures_Value |
| 676 | 592 | Age_Age50to64;livingarrangements_Notlivinginacouple_Total;measures_Value |
| 677 | 593 | Age_Age50to64;livingarrangements_Notlivinginacouple_Single(nevermarriedorneverregisteredasame-sexcivilpartnership);measures_Value |

| | | |
|---|---|---|
| 678 | 594 | Age_Age50to64;livingarrangements_Notlivinginacouple_Marriedorinaregisteredsame-sexcivilpartnership;measures_Value |
| 679 | 595 | Age_Age50to64;livingarrangements_Notlivinginacouple_Separated(butstilllegallymarriedorstilllegallyinasame-sexcivilpartnership);measures_Value |
| 680 | 596 | Age_Age50to64;livingarrangements_Notlivinginacouple_Divorcedorformerlyinasame-sexcivilpartnershipwhichisnowlegallydissolved;measures_Value |
| 681 | 597 | Age_Age50to64;livingarrangements_Notlivinginacouple_Widowedorsurvivingpartnerfromasame-sexcivilpartnership;measures_Value |
| 682 | 598 | Age_Age65andover;livingarrangements_Allcategories_Livingarrangements;measures_Value |
| 683 | 599 | Age_Age65andover;livingarrangements_Livinginacouple_Total;measures_Value |
| 684 | 600 | Age_Age65andover;livingarrangements_Livinginacouple_Marriedorinaregisteredsame-sexcivilpartnership;measures_Value |
| 685 | 601 | Age_Age65andover;livingarrangements_Livinginacouple_Cohabiting;measures_Value |
| 686 | 602 | Age_Age65andover;livingarrangements_Notlivinginacouple_Total;measures_Value |
| 687 | 603 | Age_Age65andover;livingarrangements_Notlivinginacouple_Single(nevermarriedorneverregisteredasame-sexcivilpartnership);measures_Value |
| 688 | 604 | Age_Age65andover;livingarrangements_Notlivinginacouple_Marriedorinaregisteredsame-sexcivilpartnership;measures_Value |
| 689 | 605 | Age_Age65andover;livingarrangements_Notlivinginacouple_Separated(butstilllegallymarriedorstilllegallyinasame-sexcivilpartnership);measures_Value |
| 690 | 606 | Age_Age65andover;livingarrangements_Notlivinginacouple_Divorcedorformerlyinasame-sexcivilpartnershipwhichisnowlegallydissolved;measures_Value |
| 691 | 607 | Age_Age65andover;livingarrangements_Notlivinginacouple_Widowedorsurvivingpartnerfromasame-sexcivilpartnership;measures_Value |
| 692 | 608 | RuralUrban |
| 693 | 609 | Qualifications_Allcategories_Highestlevelofqualification;measures_Value |
| 694 | 610 | Qualifications_Allcategories_Highestlevelofqualification;measures_Percent |
| 695 | 611 | Qualifications_Noqualifications;measures_Value |
| 696 | 612 | Qualifications_Noqualifications;measures_Percent |
| 697 | 613 | Qualifications_Highestlevelofqualification_Level1qualifications;measures_Value |
| 698 | 614 | Qualifications_Highestlevelofqualification_Level1qualifications;measures_Percent |
| 699 | 615 | Qualifications_Highestlevelofqualification_Level2qualifications;measures_Value |
| 700 | 616 | Qualifications_Highestlevelofqualification_Level2qualifications;measures_Percent |
| 701 | 617 | Qualifications_Highestlevelofqualification_Apprenticeship;measures_Value |
| 702 | 618 | Qualifications_Highestlevelofqualification_Apprenticeship;measures_Percent |
| 703 | 619 | Qualifications_Highestlevelofqualification_Level3qualifications;measures_Value |
| 704 | 620 | Qualifications_Highestlevelofqualification_Level3qualifications;measures_Percent |
| 705 | 621 | Qualifications_Highestlevelofqualification_Level4qualificationsandabove;measures_Value |
| 706 | 622 | Qualifications_Highestlevelofqualification_Level4qualificationsandabove;measures_Percent |
| 707 | 623 | Qualifications_Highestlevelofqualification_Otherqualifications;measures_Value |
| 708 | 624 | Qualifications_Highestlevelofqualification_Otherqualifications;measures_Percent |
| 709 | 625 | Qualifications_Schoolchildrenandfull-timestudents_Age16to17;measures_Value |
| 710 | 626 | Qualifications_Schoolchildrenandfull-timestudents_Age16to17;measures_Percent |
| 711 | 627 | Qualifications_Schoolchildrenandfull-timestudents_Age18andover;measures_Value |
| 712 | 628 | Qualifications_Schoolchildrenandfull-timestudents_Age18andover;measures_Percent |
| 713 | 629 | Qualifications_Full-timestudents_Age18to74_Economicallyactive_Inemployment;measures_Value |
| 714 | 630 | Qualifications_Full-timestudents_Age18to74_Economicallyactive_Inemployment;measures_Percent |
| 715 | 631 | Qualifications_Full-timestudents_Age18to74_Economicallyactive_Unemployed;measures_Value |

| | | |
|---|---|---|
| 716 | 632 | Qualifications_Full-timestudents_Age18to74_Economicallyactive_Unemployed;measures_Percent |
| 717 | 633 | Qualifications_Full-timestudents_Age18to74_Economicallyinactive;measures_Value |
| 718 | 634 | Qualifications_Full-timestudents_Age18to74_Economicallyinactive;measures_Percent |
| 719 | 635 | Age_Allcategories_Age;ResidenceType_Livesinahousehold;measures_Value |
| 720 | 636 | Age_Allcategories_Age;ResidenceType_Livesinacommunalestablishment;measures_Value |
| 721 | 637 | Age_Age0to15;ResidenceType_Allcategories_Residencetype;measures_Value |
| 722 | 638 | Age_Age0to15;ResidenceType_Livesinahousehold;measures_Value |
| 723 | 639 | Age_Age0to15;ResidenceType_Livesinacommunalestablishment;measures_Value |
| 724 | 640 | Age_Age16to24;ResidenceType_Allcategories_Residencetype;measures_Value |
| 725 | 641 | Age_Age16to24;ResidenceType_Livesinahousehold;measures_Value |
| 726 | 642 | Age_Age16to24;ResidenceType_Livesinacommunalestablishment;measures_Value |
| 727 | 643 | Age_Age25to34;ResidenceType_Allcategories_Residencetype;measures_Value |
| 728 | 644 | Age_Age25to34;ResidenceType_Livesinahousehold;measures_Value |
| 729 | 645 | Age_Age25to34;ResidenceType_Livesinacommunalestablishment;measures_Value |
| 730 | 646 | Age_Age35to49;ResidenceType_Allcategories_Residencetype;measures_Value |
| 731 | 647 | Age_Age35to49;ResidenceType_Livesinahousehold;measures_Value |
| 732 | 648 | Age_Age35to49;ResidenceType_Livesinacommunalestablishment;measures_Value |
| 733 | 649 | Age_Age50to64;ResidenceType_Allcategories_Residencetype;measures_Value |
| 734 | 650 | Age_Age50to64;ResidenceType_Livesinahousehold;measures_Value |
| 735 | 651 | Age_Age50to64;ResidenceType_Livesinacommunalestablishment;measures_Value |
| 736 | 652 | Age_Age65andover;ResidenceType_Allcategories_Residencetype;measures_Value |
| 737 | 653 | Age_Age65andover;ResidenceType_Livesinahousehold;measures_Value |
| 738 | 654 | Age_Age65andover;ResidenceType_Livesinacommunalestablishment;measures_Value |
| | | ***Deprivation*** |
| 739 | 1 | IMDSCORE |
| 740 | 2 | RANKOFIMDSCORE(where1ismostdeprived) |
| 741 | 3 | IMDDecile_1isMostDeprived |
| | | ***Place based longitudinal data*** |
| 742 | 1 | Employment and Support Allowance claimants |
| 743 | 2 | Distance to nearest Gambling Outlet (minutes) |
| 744 | 3 | Distance to nearest Fast Food Outlet (minutes) |
| 745 | 4 | Distance to nearest Pubs/Bars/Nightclub (minutes) |
| 746 | 5 | Distance to nearest Off licence (minutes) |
| 747 | 6 | Distance to nearest Tobacconists/Vape Store (minutes) |
| 748 | 7 | Distance to nearest GP Practice (minutes) |
| 749 | 8 | Distance to nearest GP Practice (minutes) |
| 750 | 9 | Distance to nearest Hospital (minutes) |
| 751 | 10 | Distance to nearest Hospital (minutes) |

| | | |
|---|---|---|
| 752 | 11 | Distance to nearest Dentist (minutes) |
| 753 | 12 | Distance to nearest Dentist (minutes) |
| 754 | 13 | Distance to nearest Pharmacy (minutes) |
| 755 | 14 | Distance to nearest Pharmacy (minutes) |
| 756 | 15 | Distance to nearest Pharmacy (minutes) |
| 757 | 16 | Distance to nearest Leisure Centre (minutes) |
| 758 | 17 | Distance to nearest Blue space (minutes) |
| 759 | 18 | NVDI value indicating Passive Green Space |
| 760 | 19 | Annual mean Nitrogen Dioxide ($\mu gm^3$) |
| 761 | 20 | Annual mean Particulate Matter ($\mu gm^3$) |
| 762 | 21 | Annual mean Sulpur Dioxide ($\mu gm^3$) |
| 763 | 22 | Health Domain Score |
| 764 | 23 | Green/Bluespace Domain Score |
| 765 | 24 | Air quality Domain Score |
| 766 | 25 | Retail Domain Score |
| 767 | 26 | Access to Healthy Assets and Hazards index score |
| 768 | 27 | AHAH Index ranked |
| 769 | 28 | AHAH Index percentiles |
| 770 | 29 | SmallAreaMentalHealthIndex_index.2011 |
| 771 | 30 | SmallAreaMentalHealthIndex_decile2011 |
| 772 | 31 | SmallAreaMentalHealthIndex_index.2012 |
| 773 | 32 | SmallAreaMentalHealthIndex_decile2012 |
| 774 | 33 | SmallAreaMentalHealthIndex_index.2013 |
| 775 | 34 | SmallAreaMentalHealthIndex_decile2013 |
| 776 | 35 | SmallAreaMentalHealthIndex_index.2014 |
| 777 | 36 | SmallAreaMentalHealthIndex_decile2014 |
| 778 | 37 | SmallAreaMentalHealthIndex_index.2015 |
| 779 | 38 | SmallAreaMentalHealthIndex_decile2015 |
| 780 | 39 | SmallAreaMentalHealthIndex_index.2016 |
| 781 | 40 | SmallAreaMentalHealthIndex_decile2016 |
| 782 | 41 | SmallAreaMentalHealthIndex_index.2017 |
| 783 | 42 | SmallAreaMentalHealthIndex_decile2017 |
| 784 | 43 | SmallAreaMentalHealthIndex_index.2018 |
| 785 | 44 | SmallAreaMentalHealthIndex_decile2018 |
| 786 | 45 | SmallAreaMentalHealthIndex_index.2019 |
| 787 | 46 | SmallAreaMentalHealthIndex_decile2019 |

Supplementary Table S4. Censoring correction by cancer type.

| Cancer | N. of records | Censored records (%) | Censored records after correction (%) | Mean number of diagnoses for uncensored records | Mean number of diagnoses for uncensored records after correction | % records with censored values between 1 and 2 | % records with censored values between 1 and 3 | % records with censored values between 1 and 4 | % records with censored values between 1 and 5 | Sum of diagnoses from uncensored records before correction | Sum of diagnoses from uncensored records after correction | Number of censored records after correction |
|---|---|---|---|---|---|---|---|---|---|---|---|---|
| Brain/CNS | 7 | 100 | 100 | NA | NA | 0 | 0 | 0 | 100 | 0 | 0 | 7 |
| Breast | 606 | 81.35 | 77.72 | 7.77 | 6.67 | 45.54 | 21.29 | 2.97 | 7.92 | 878 | 900 | 471 |
| Colorectal | 588 | 92.18 | 85.71 | 6.93 | 4.25 | 59.52 | 19.22 | 0 | 6.97 | 319 | 357 | 504 |
| Gynaecology | 406 | 99.51 | 43.84 | 6 | 1.04 | 37.19 | 0 | 0 | 6.65 | 12 | 238 | 178 |
| Haematology | 479 | 97.49 | 62.84 | 6.58 | 1.38 | 55.53 | 0.42 | 0 | 6.89 | 79 | 245 | 301 |
| Head and Neck | 366 | 100 | 15.57 | NA | 1 | 8.2 | 0 | 0 | 7.38 | 0 | 309 | 57 |
| Lung | 531 | 88.14 | 66.1 | 7.25 | 3.19 | 35.4 | 21.66 | 0.38 | 8.66 | 457 | 574 | 351 |
| Other | 266 | 99.62 | 13.91 | 6 | 1.02 | 1.88 | 0 | 0 | 12.03 | 6 | 234 | 37 |
| Sarcoma | 54 | 100 | 100 | NA | NA | 0 | 0 | 0 | 100 | 0 | 0 | 54 |
| Skin | 669 | 50.82 | 45.59 | 11.66 | 10.7 | 17.19 | 16.14 | 4.93 | 7.32 | 3837 | 3893 | 305 |
| Upper GI | 498 | 98.19 | 64.66 | 6.33 | 1.27 | 46.79 | 8.03 | 0 | 9.84 | 57 | 224 | 322 |
| Urology | 640 | 75.62 | 74.69 | 7.73 | 7.49 | 42.97 | 22.34 | 3.91 | 5.47 | 1206 | 1214 | 478 |

Supplementary Table S5. Number of cancer types in postcode sectors

| **Number of cancer types in each postcode sector** | **Number of postcode sectors** | **Range in the proportion of the population in postcode sectors respect to the overall in the Morecambe Bay ex CCG extended area.** |
|---|---|---|
| 12 | 4 | 0.83% - 2.30% |
| 11 | 30 | 0.49% - 3.26% |
| 10 | 20 | 0.26% - 2.53% |
| 9 | 2 | 0.22% - 0.63% |
| 8 | 1 | 1.90% |
| 6 | 1 | 2.38% |
| 4 | 2 | 1.02% - 1.07% |
| 2 | 1 | 1.40% |
| 1 | 14 | 0.2% - 2.73% |

Supplementary Table S6. Variable selection method: summary statistics for selected candidate variables for joint modelling. SD = standard deviation; min = minimum, max = maximum, 25PER = 25th percentile or first quartile (the value below which 25% of the data falls in a dataset), 75PER = 75th percentile or third quartile (the value below which 75% of the data falls in a dataset).

| Risk Factor (number of people with) | Mean | SD | Min | 25PER | 75PER | Max |
|---|---|---|---|---|---|---|
| Age 60 to 80 | 7.9 | 5.8 | 0 | 4 | 11 | 42 |
| Age above 80 | 4.3 | 3.7 | 0 | 2 | 5 | 21 |
| Any other Black background | 0.0014 | 0.037 | 0 | 0 | 0 | 1 |
| British | 8.9 | 7.3 | 0 | 4 | 12 | 45 |
| Chronic Kidney Disease | 4.7 | 3.6 | 0 | 2 | 6 | 24 |
| Congestive Hearth Failure | 1.4 | 1.3 | 0 | 0 | 2 | 10 |
| Coronary Heart Disease | 14 | 11 | 0 | 6 | 20 | 54 |
| COVID19 | 0.73 | 1.4 | 0 | 0 | 1 | 8 |
| Depression | 6.3 | 4.4 | 0 | 3 | 8 | 26 |
| Diabetes | 5.6 | 4.1 | 0 | 3 | 7 | 25 |
| Ex Smoker | 0.84 | 0.96 | 0 | 0 | 1 | 6 |
| Female age 25-49 mixed white and Asian (area) | 3.9 | 5.1 | 0 | 0 | 8 | 20 |
| Frailty (number of fit) | 5.9 | 4.2 | 0 | 3 | 7 | 24 |
| Hypertension | 11 | 8.1 | 0 | 5 | 15 | 45 |
| Inemployed, self employed (including fulltime students) one parent working (area) | 30 | 20 | 0 | 15 | 45 | 110 |
| Male Pakistani (area) | 12 | 29 | 0 | 0 | 9 | 220 |
| Northern Irish (area) | 0.0014 | 0.037 | 0 | 0 | 0 | 1 |
| Other White background (area) | 0.0055 | 0.074 | 0 | 0 | 0 | 1 |
| Pakistani | 0.0028 | 0.053 | 0 | 0 | 0 | 1 |
| Scottish (area) | 0.022 | 0.15 | 0 | 0 | 0 | 1 |
| Smoker | 0.89 | 1 | 0 | 0 | 1 | 5 |
| White and Black Caribbean | 0.0014 | 0.037 | 0 | 0 | 0 | 1 |

### Joint modelling: statistically significant risk factors for Breast cancer

| Risk factor | OR | Low CI | Upp CI |
|---|---|---|---|
| Chronic Kidney Disease | 1.14 | 1.07 | 1.21 |
| Congestive Heart Failure | 1.08 | 1.04 | 1.13 |
| COVID19 | 1.11 | 1.02 | 1.20 |
| Depression | 1.00 | 1.00 | 1.01 |
| Diabetes | 1.07 | 1.02 | 1.13 |
| Age above 80 | 1.24 | 1.04 | 1.50 |
| Time | 1.34 | 1.09 | 1.61 |

### Joint modelling: statistically significant risk factors for Colorectal cancer

| Risk factor | OR | Low CI | Upp CI |
|---|---|---|---|
| Coronary Hearth Disease | 1.04 | 1.02 | 1.06 |
| COVID19 | 1.06 | 1.01 | 1.10 |
| Depression | 1.10 | 1.03 | 1.13 |
| Diabetes | 1.31 | 1.20 | 1.44 |
| Hypertension | 1.06 | 1.00 | 1.14 |
| Age 60 to 80 | 5.14 | 2.30 | 11.39 |
| Age above 80 | 0.73 | 0.59 | 0.95 |
| Ex Smoker | 1.23 | 1.10 | 1.37 |
| Inemployed, self employed (including fulltime students) one parent working (area) | 0.95 | 0.91 | 0.99 |
| British | 1.52 | 1.26 | 1.81 |
| Any other Black background | 1.09 | 1.03 | 1.16 |
| Other White background (area) | 0.89 | 0.83 | 0.95 |
| Pakistani | 1.11 | 1.03 | 1.19 |
| Scottish (area) | 1.19 | 1.11 | 1.28 |
| White and Black Caribbean (area) | 1.18 | 1.03 | 1.36 |
| Time | 1.08 | 1.02 | 1.15 |

### Joint modelling: statistically significant risk factors for Gynaecology cancer

| Risk factor | OR | Low CI | Upp CI |
|---|---|---|---|
| Chronic Kidney Disease | 1.00 | 1.00 | 1.00 |
| Depression | 0.91 | 0.84 | 1.00 |
| Diabetes | 1.04 | 1.01 | 1.07 |
| Frailty (number of fit) | 0.89 | 0.83 | 0.95 |
| Age above 80 | 1.26 | 1.17 | 1.34 |
| Smoker | 0.92 | 0.84 | 0.99 |
| Other White background (area) | 1.01 | 1.00 | 1.02 |

### Joint modelling: statistically significant risk factors for Haematology cancer

| **Risk factor** | **OR** | **Low CI** | **Upp CI** |
|---|---|---|---|
| Chronic Kidney Disease | 1.04 | 1.01 | 1.06 |
| Covid19 | 1.16 | 1.03 | 1.31 |
| Smoker | 1.12 | 1.01 | 1.23 |
| White and Black Caribbean | 1.11 | 1.01 | 1.22 |

### Joint modelling: statistically significant risk factors for Head and Neck cancer

| Risk factor | OR | Low CI | Upp CI |
|---|---|---|---|
| Chronic Kidney Disease | 0.98 | 0.96 | 1.00 |
| COVID19 | 1.16 | 1.04 | 1.30 |
| Smoker | 1.12 | 1.02 | 1.24 |
| White and Black Caribbean | 1.11 | 1.01 | 1.21 |
| Northern Irish (area) | 0.90 | 0.82 | 0.98 |

### Joint modelling: statistically significant risk factors for Lung cancer

| Risk factor | Odd ratios (OR) | Low 95% Confidence interval | Upper 95% Confidence interval |
|---|---|---|---|
| Congestive Heart Failure | 0.35 | 0.14 | 0.87 |
| Chronic Kidney Disease | 1.28 | 1.16 | 1.41 |
| Coronary Heart Disease | 1.18 | 1.11 | 1.26 |
| Smoker | 1.05 | 1.03 | 1.07 |
| Inemployed, self employed (including fulltime students) one parent working (area) | 0.92 | 0.86 | 0.98 |
| Female age 25-49 mixed white and Asian (area) | 11.53 | 2.81 | 55.79 |
| Male Pakistani (area) | 1.07 | 1.00 | 1.14 |
| Northern Irish (area) | 1.30 | 1.02 | 1.69 |

### Joint modelling: statistically significant risk factors for Skin cancer

| Risk factor | OR | Low CI | Upp CI |
|---|---|---|---|
| Ex Smoker | 0.69 | 0.55 | 0.87 |
| Any other Black background | 8.23 | 3.44 | 18.54 |

### Joint modelling: statistically significant risk factors for Upper GI cancer

| Risk factor | OR | Low CI | Upp CI |
|---|---|---|---|
| Chronic Kidney Disease | 1.00 | 1.00 | 1.00 |
| Covid19 | 0.98 | 0.97 | 0.99 |
| Depression | 0.90 | 0.83 | 0.99 |
| Frailty (number of fit) | 0.89 | 0.83 | 0.96 |
| Smoker | 0.91 | 0.84 | 0.99 |
| Northern Irish (area) | 0.97 | 0.95 | 1.00 |

### Joint modelling: statistically significant risk factors for Urology cancer

| Risk factor | OR | Low CI | Upp CI |
|---|---|---|---|
| COVID19 | 0.95 | 0.90 | 1.00 |
| Diabetes | 0.69 | 0.67 | 0.80 |
| Age 60 to 80 | 1.20 | 1.12 | 1.28 |
| Age above 80 | 0.73 | 0.58 | 0.93 |
| Ex Smoker | 1.45 | 1.24 | 1.70 |
| British | 1.49 | 1.24 | 1.79 |
| Any other Black background | 1.09 | 1.02 | 1.16 |
| Other White background (area) | 0.89 | 0.82 | 0.95 |
| White and Black Caribbean | 1.19 | 1.04 | 1.37 |
| Scottish (area) | 1.20 | 1.12 | 1.28 |
| Time | 1.15 | 1.06 | 1.26 |

Supplementary Table S8. Largest statistically significant associations between selected important variables (selected variable) and other variables in the dataset.

| Selected variable | Correlated variable | Regression coefficient | P-value |
|---|---|---|---|
| Age 60 to 80 | Small Area Mental Health Index (higher values: worst) | 0.066 | <0.001 |
| Age above 80 | Small Area Mental Health Index (higher values: worst) | 0.063 | <0.001 |
| Any other Black background | Not living in a couple | 0.649 | 0.006 |
| British | Small Area Mental Health Index (higher values: worst) | 0.095 | <0.001 |
| Chronic Kidney Disease | Distance to nearest Tobacconists/Vape Store (minutes) | -0.015 | <0.001 |
| Congestive Hearth Failure | Small Area Mental Health Index (higher values: worst) | 0.086 | <0.001 |
| Coronary Heart Disease | Small Area Mental Health Index (higher values: worst) | 0.076 | <0.001 |
| COVID19 | Shoplifting | 0.063 | 0.001 |
| Depression | Small Area Mental Health Index (higher values: worst) | 0.065 | <0.001 |
| Diabetes | Small Area Mental Health Index (higher values: worst) | 0.063 | <0.001 |
| Ex Smoker | Distance to nearest GP Practice (minutes) | -0.036 | <0.001 |
| Frailty (number of fit) | Small Area Mental Health Index (higher values: worst) | 0.052 | <0.001 |
| Hypertension | Small Area Mental Health Index (higher values: worst) | 0.073 | <0.001 |
| Female age 25-49 mixed white and Asian (area) | Cohabitating | -0.055 | <0.001 |
| Inemployed, self employed (including fulltime students) one parent working (area) | Shoplifting | 0.011 | <0.001 |
| Northern Irish (area) | Asian, Asian British | 0.298 | 0.006 |
| Other White background (area) | Criminal damage and arson | 0.778 | 0.009 |
| Pakistani | Shoplifting | 0.007 | 0.006 |
| Scottish (area) | Loneparenting | 0.057 | 0.006 |
| Smoker | Small Area Mental Health Index (higher values: worst) | 0.093 | <0.001 |
| White and Black Caribbean | Small Area Mental Health Index (higher values: worst) | 1.751 | 0.010 |

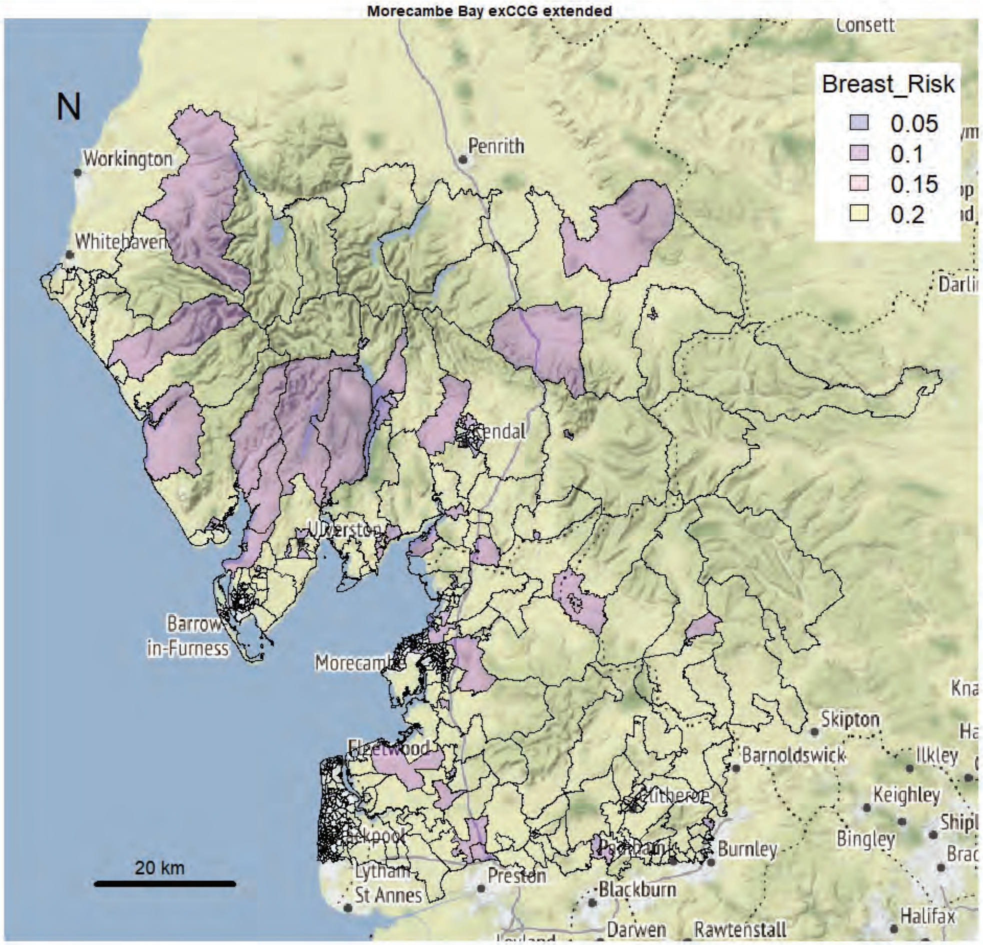

Morecambe Bay exCCG extended
N
Breast_Risk
0.05
0.1
0.15
0.2
Workington
Whitehaven
Penrith
Consett
Kendal
Barrow-in-Furness
Morecambe
Fleetwood
Lytham St Annes
Preston
Blackburn
Darwen
Rawtenstall
Burnley
Clitheroe
Skipton
Barnoldswick
Ilkley
Keighley
Bingley
Halifax
20 km


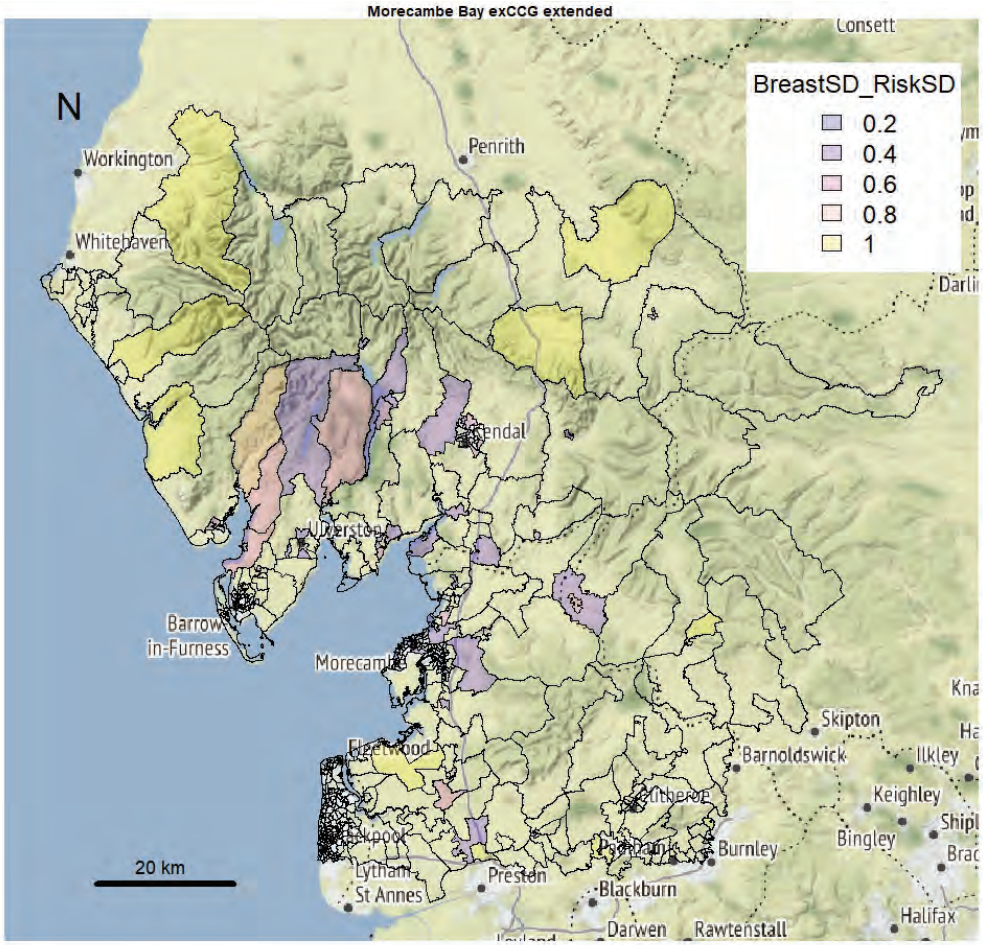

Morecambe Bay exCCG extended
N
BreastSD_RiskSD
0.2
0.4
0.6
0.8
1
Workington
Whitehaven
Penrith
Consett
Kendal
Barrow-in-Furness
Morecambe
Fleetwood
Lytham St Annes
Preston
Blackburn
Darwen
Rawtenstall
Burnley
Clitheroe
Skipton
Barnoldswick
Ilkley
Keighley
Bingley
Halifax
20 km

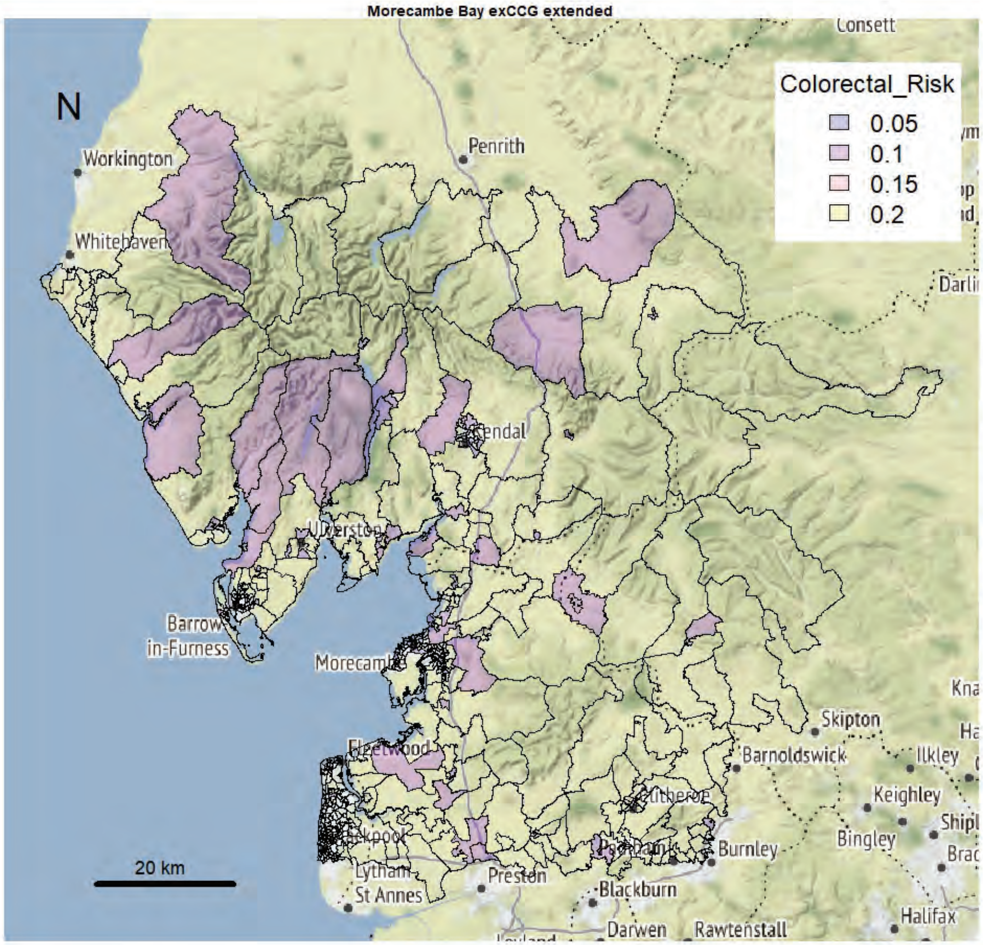
Morecambe Bay exCCG extended
Colorectal_Risk
0.05
0.1
0.15
0.2
N
20 km

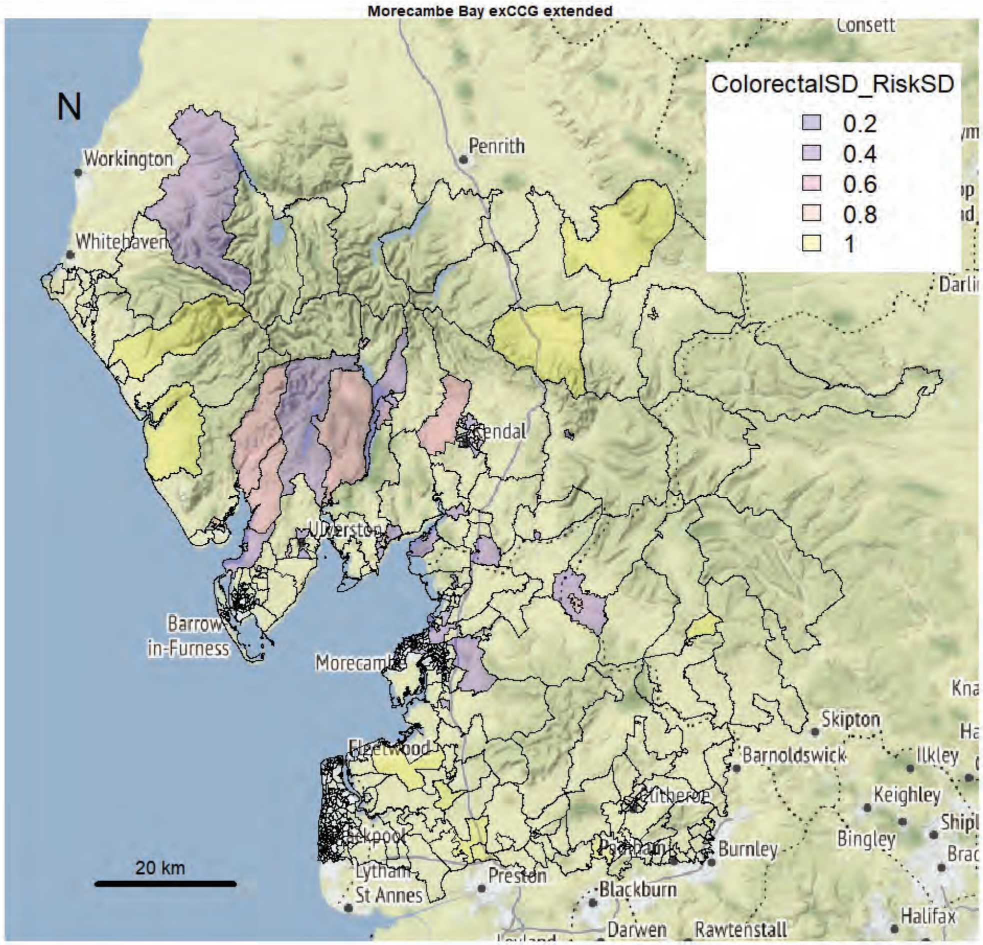
Morecambe Bay exCCG extended
ColorectalSD_RiskSD
0.2
0.4
0.6
0.8
1
N
20 km

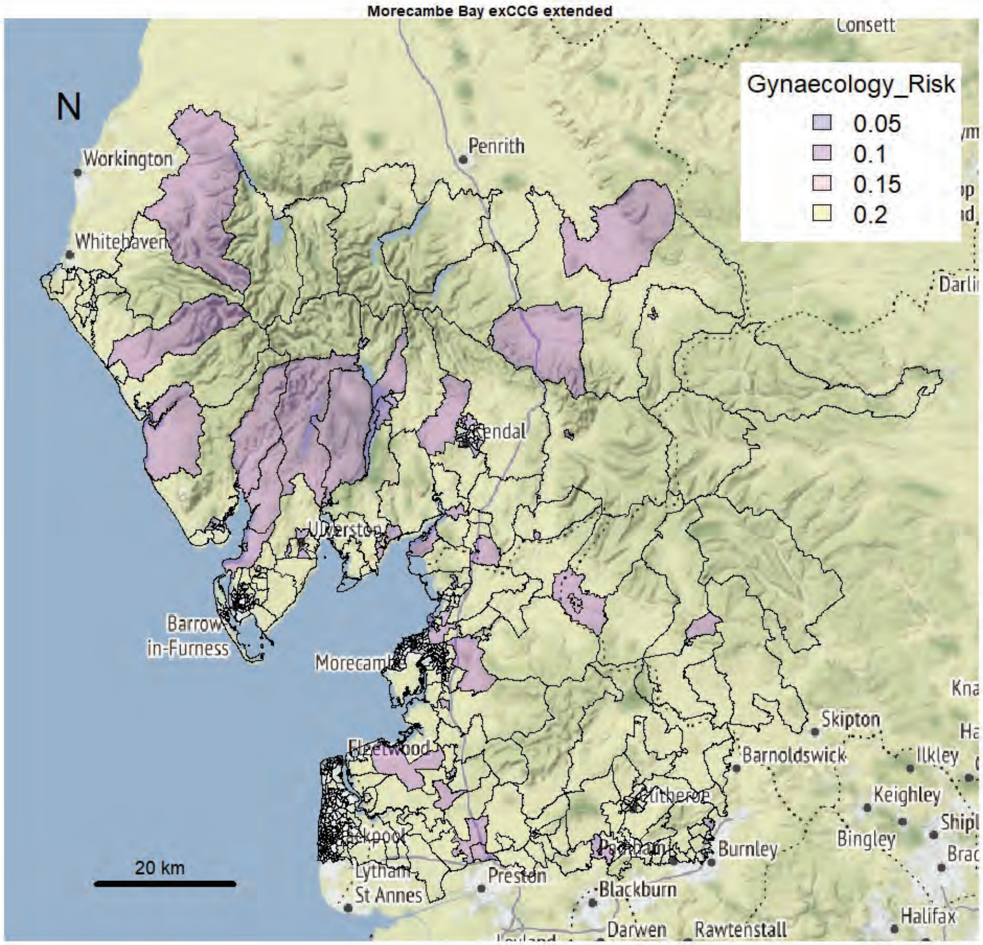

Morecambe Bay exCCG extended
Gynaecology_Risk
0.05
0.1
0.15
0.2
N
20 km
Workington
Whitehaven
Penrith
Consett
Kendal
Barrow in-Furness
Morecambe
Fleetwood
Lytham St Annes
Preston
Blackburn
Darwen
Rawtenstall
Burnley
Barnoldswick
Skipton
Ilkley
Keighley
Bingley
Halifax


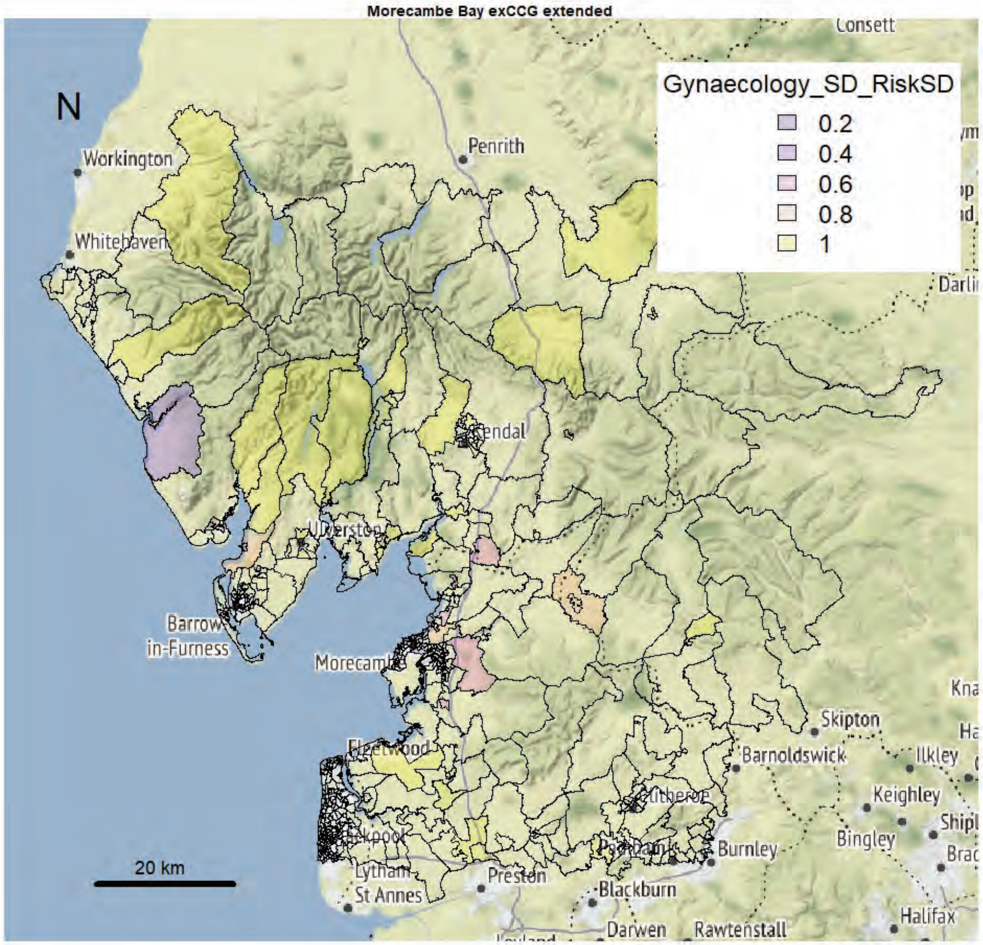

Morecambe Bay exCCG extended
Gynaecology_SD_RiskSD
0.2
0.4
0.6
0.8
1
N
20 km
Workington
Whitehaven
Penrith
Consett
Kendal
Barrow in-Furness
Morecambe
Fleetwood
Lytham St Annes
Preston
Blackburn
Darwen
Rawtenstall
Burnley
Barnoldswick
Skipton
Ilkley
Keighley
Bingley
Halifax

Morecambe Bay exCCG extended

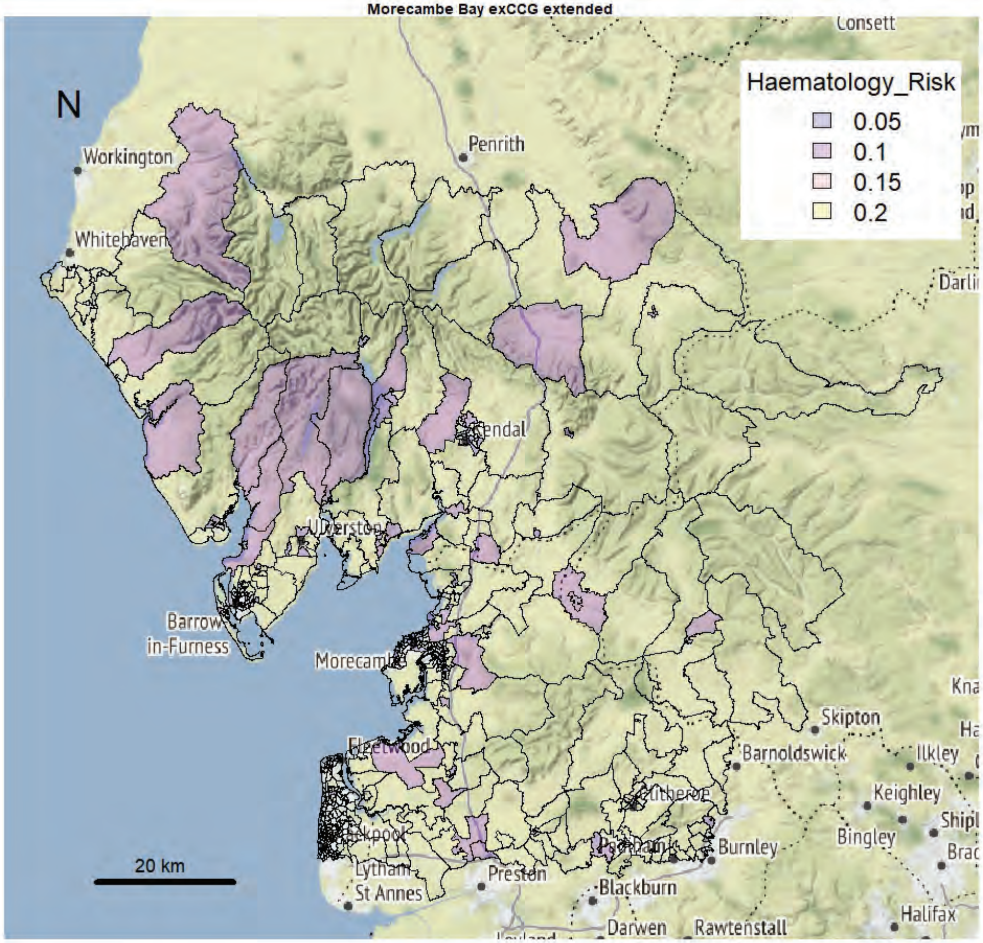


Morecambe Bay exCCG extended

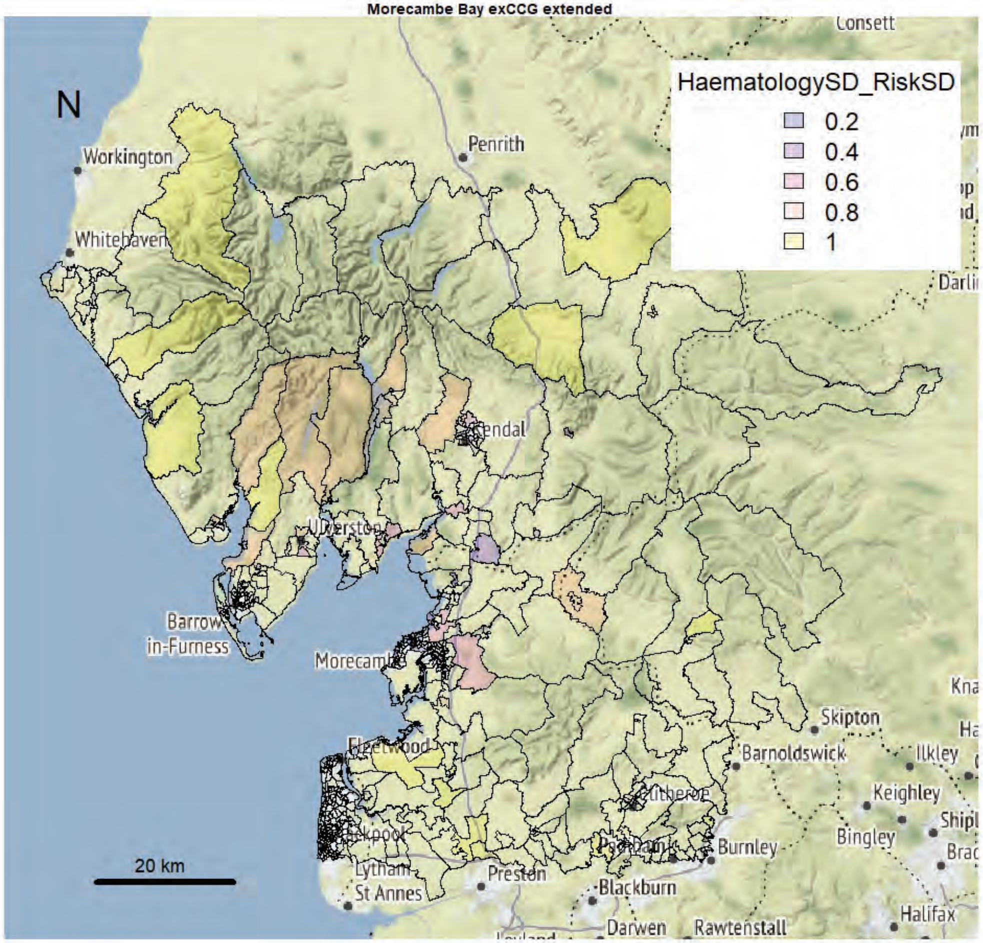

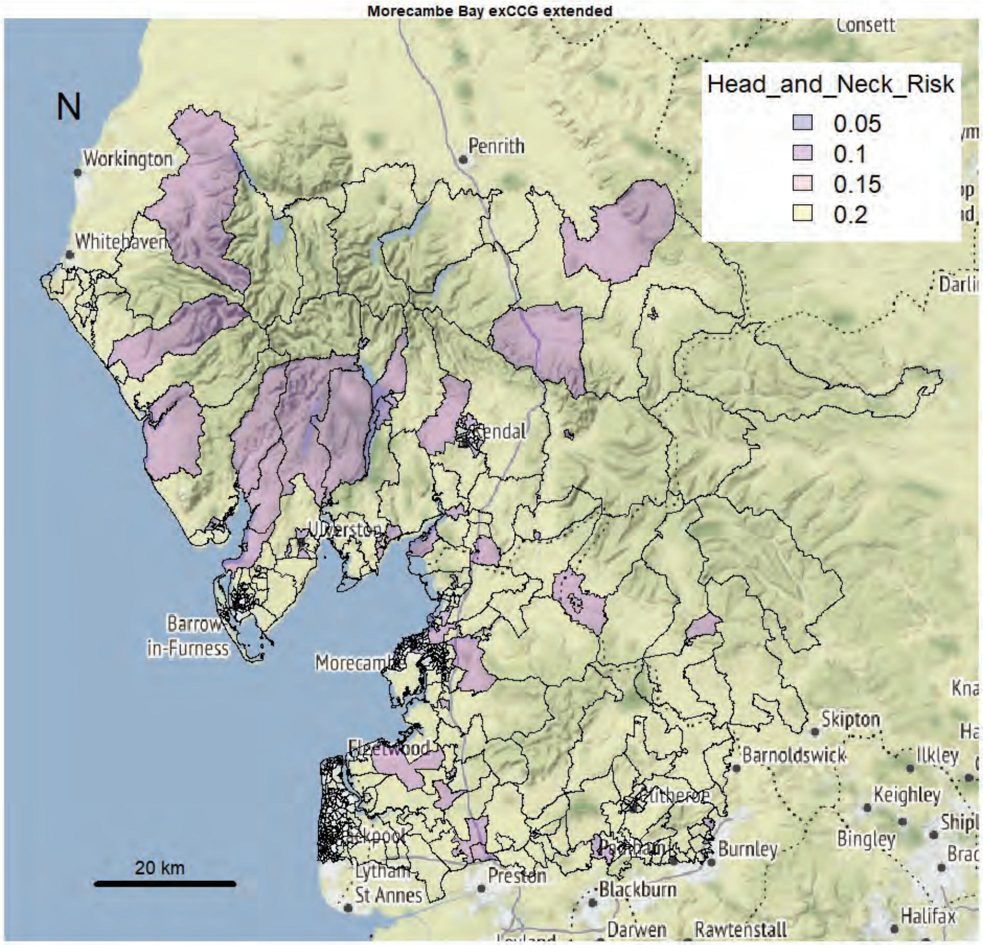
Morecambe Bay exCCG extended
N
Head_and_Neck_Risk
0.05
0.1
0.15
0.2
Workington
Whitehaven
Penrith
Consett
Barrow in-Furness
Morecambe
Fleetwood
Lytham St Annes
Preston
Blackburn
Darwen
Rawtenstall
Burnley
Barnoldswick
Skipton
Ilkley
Keighley
Bingley
Halifax
20 km

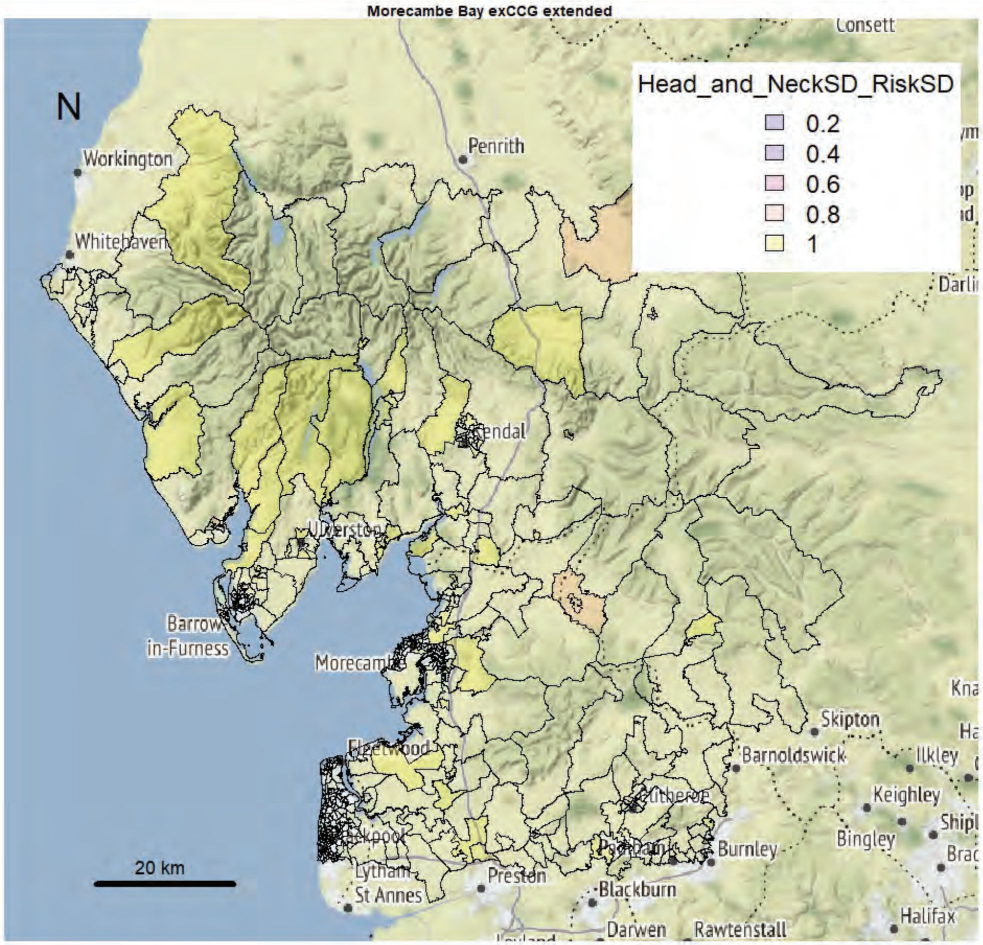
Morecambe Bay exCCG extended
N
Head_and_NeckSD_RiskSD
0.2
0.4
0.6
0.8
1
Workington
Whitehaven
Penrith
Consett
Barrow in-Furness
Morecambe
Fleetwood
Lytham St Annes
Preston
Blackburn
Darwen
Rawtenstall
Burnley
Barnoldswick
Skipton
Ilkley
Keighley
Bingley
Halifax
20 km

Morecambe Bay exCCG extended

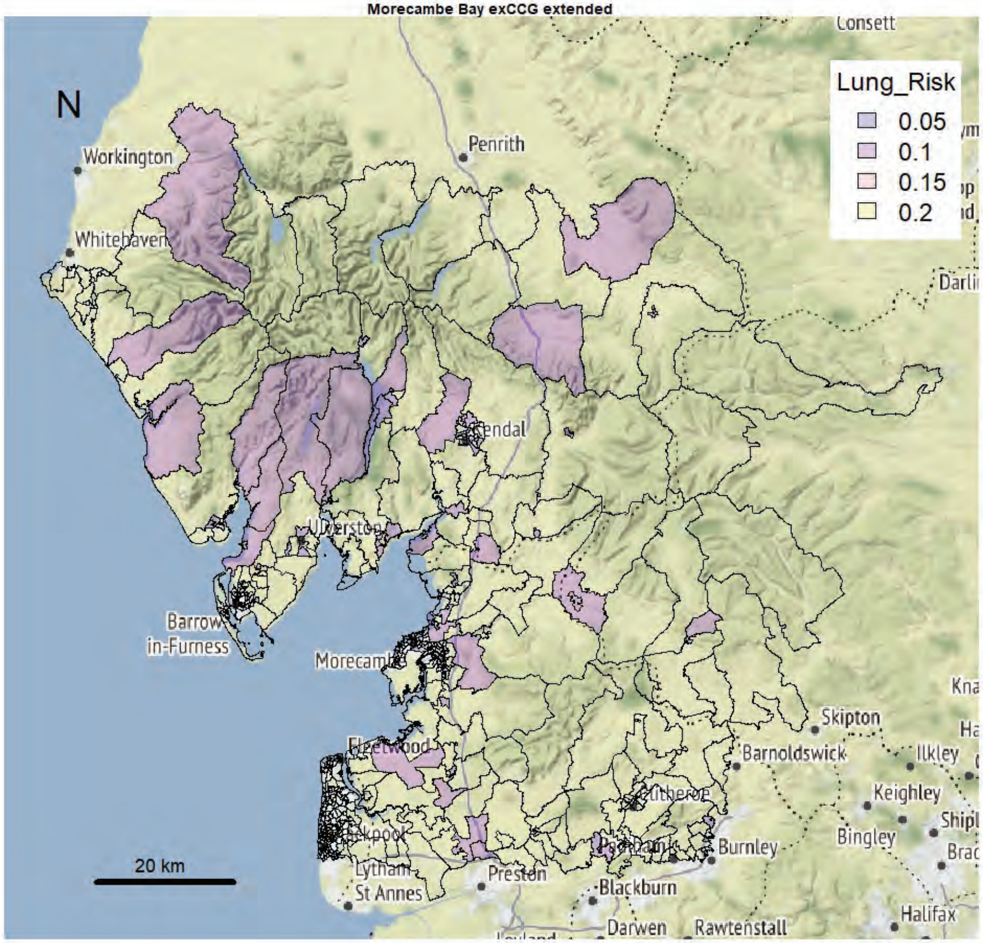


Morecambe Bay exCCG extended

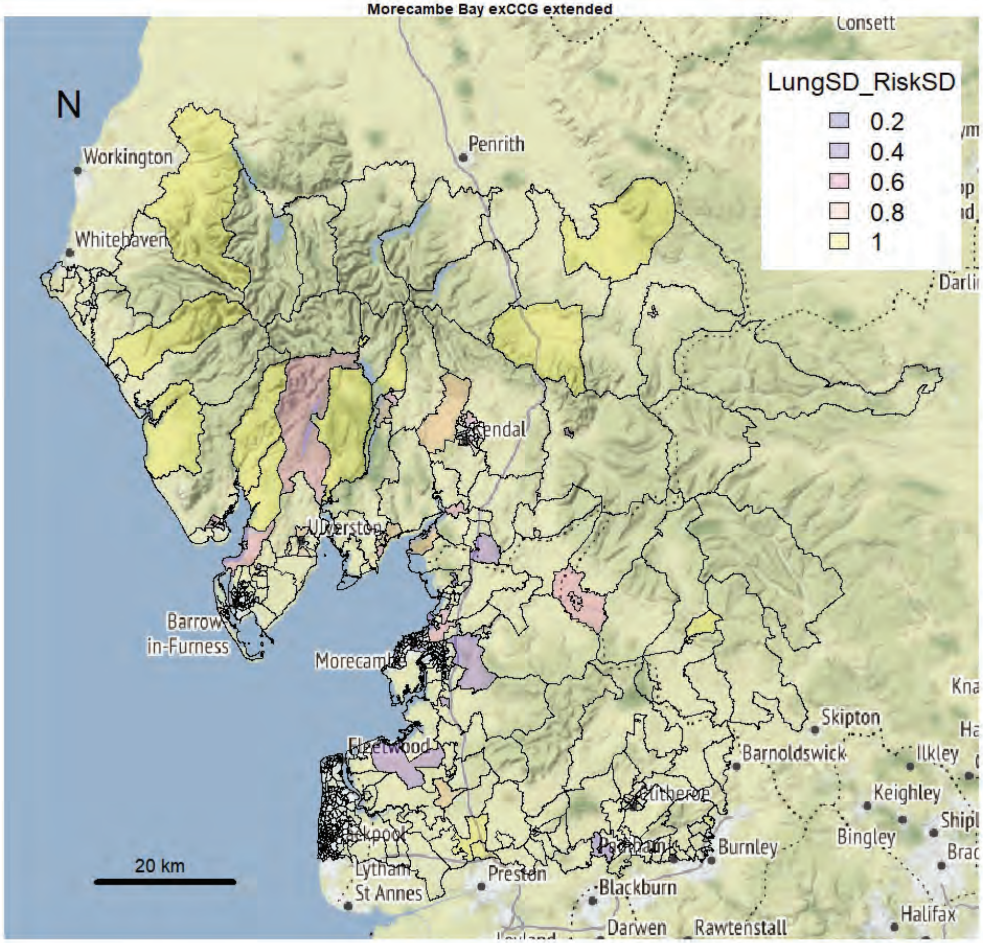

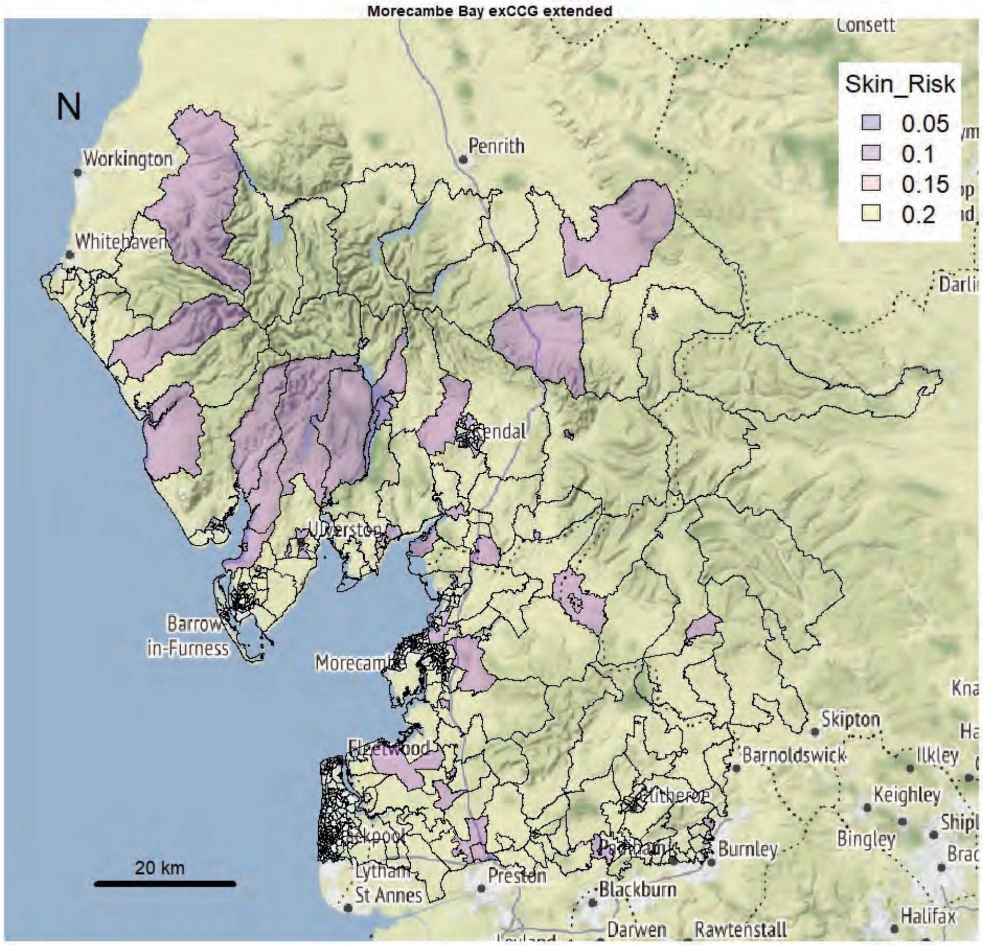
Morecambe Bay exCCG extended
N
Skin_Risk
0.05
0.1
0.15
0.2
Workington
Whitehaven
Penrith
Consett
Kendal
Barrow
in-Furness
Morecambe
Skipton
Barnoldswick
Ilkley
Keighley
Bingley
Burnley
Preston
Blackburn
Darwen
Rawtenstall
Halifax
St Annes
20 km
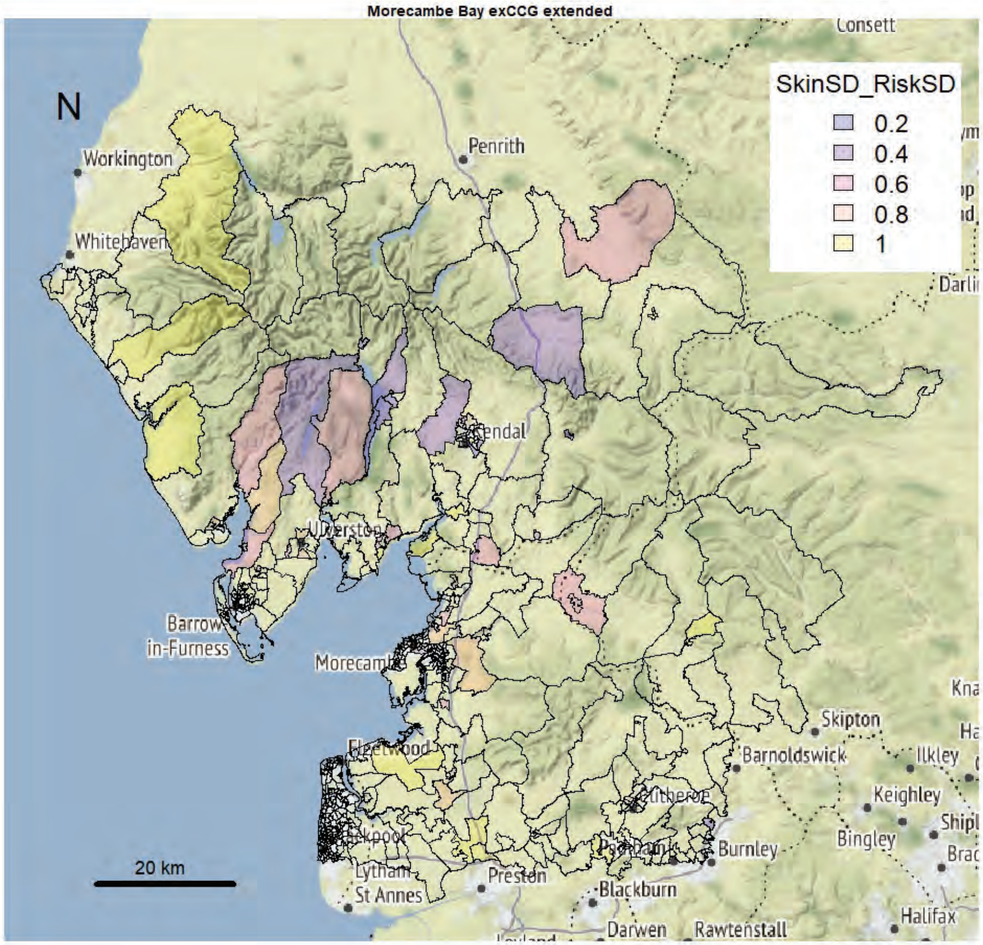
Morecambe Bay exCCG extended
N
SkinSD_RiskSD
0.2
0.4
0.6
0.8
1
Workington
Whitehaven
Penrith
Consett
Kendal
Barrow
in-Furness
Morecambe
Skipton
Barnoldswick
Ilkley
Keighley
Bingley
Burnley
Preston
Blackburn
Darwen
Rawtenstall
Halifax
St Annes
20 km

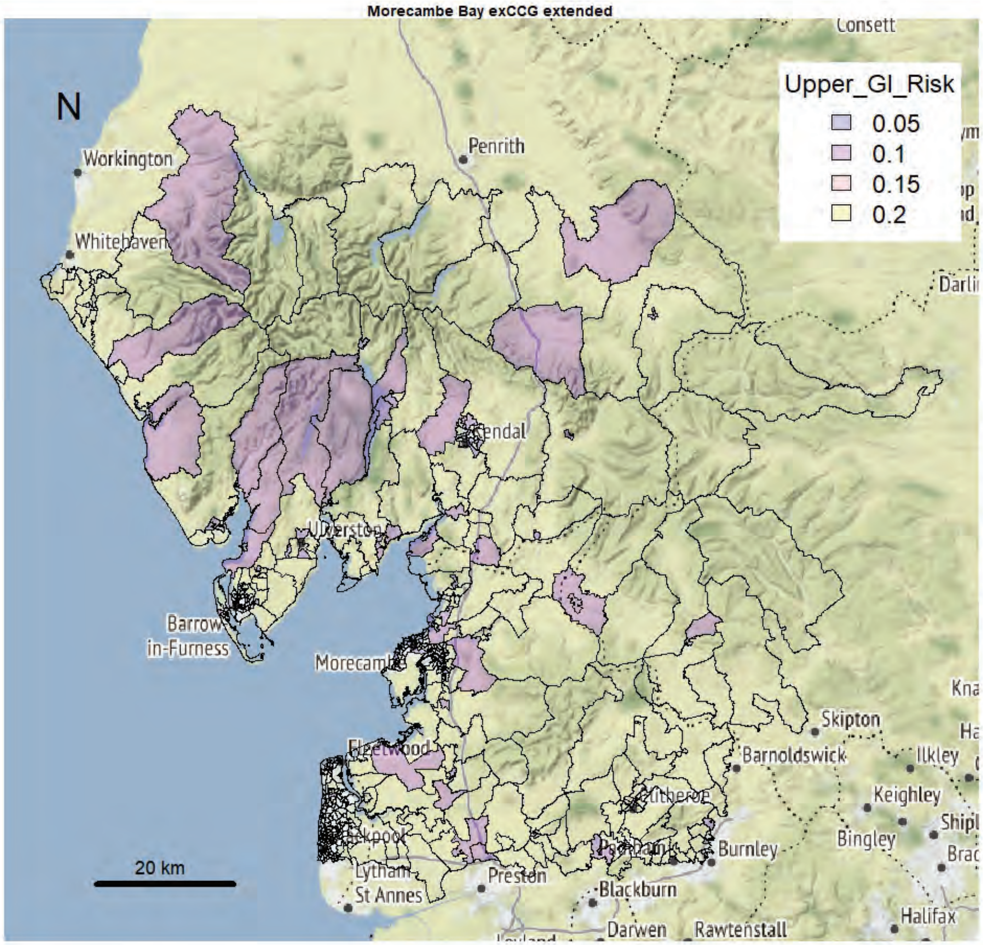

Morecambe Bay exCCG extended
Upper_GI_Risk
0.05
0.1
0.15
0.2
N
20 km


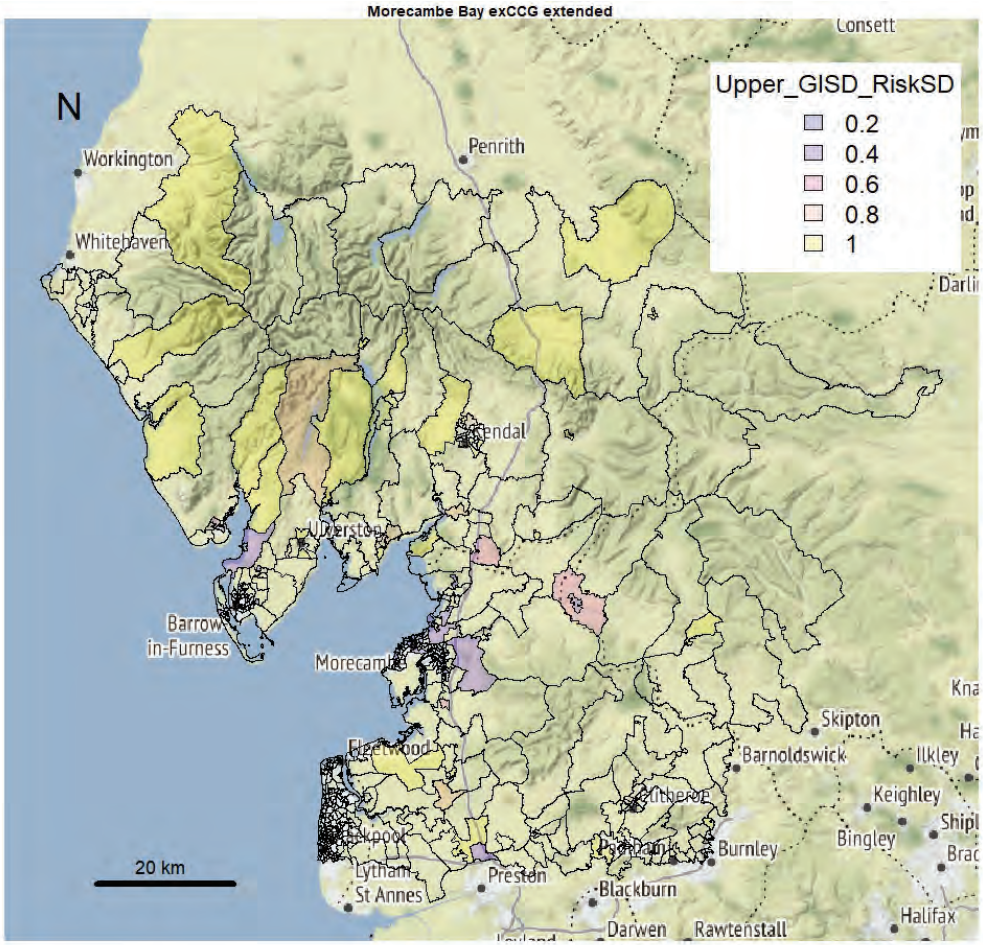

Morecambe Bay exCCG extended
Upper_GISD_RiskSD
0.2
0.4
0.6
0.8
1
N
20 km

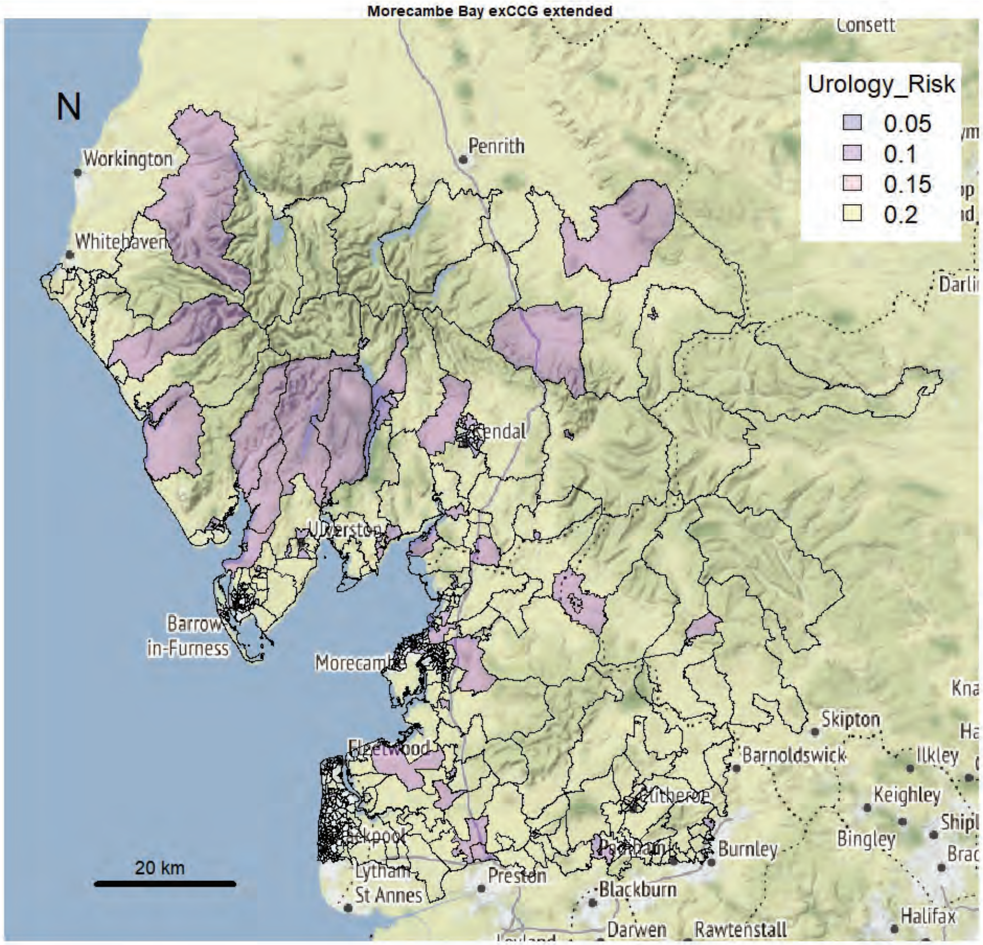
Morecambe Bay exCCG extended
N
Urology_Risk
0.05
0.1
0.15
0.2
20 km
Workington
Whitehaven
Penrith
Kendal
Ulverston
Barrow in-Furness
Morecambe
Fleetwood
Lytham St Annes
Preston
Blackburn
Burnley
Darwen
Rawtenstall
Skipton
Barnoldswick
Ilkley
Keighley
Bingley
Halifax
Consett

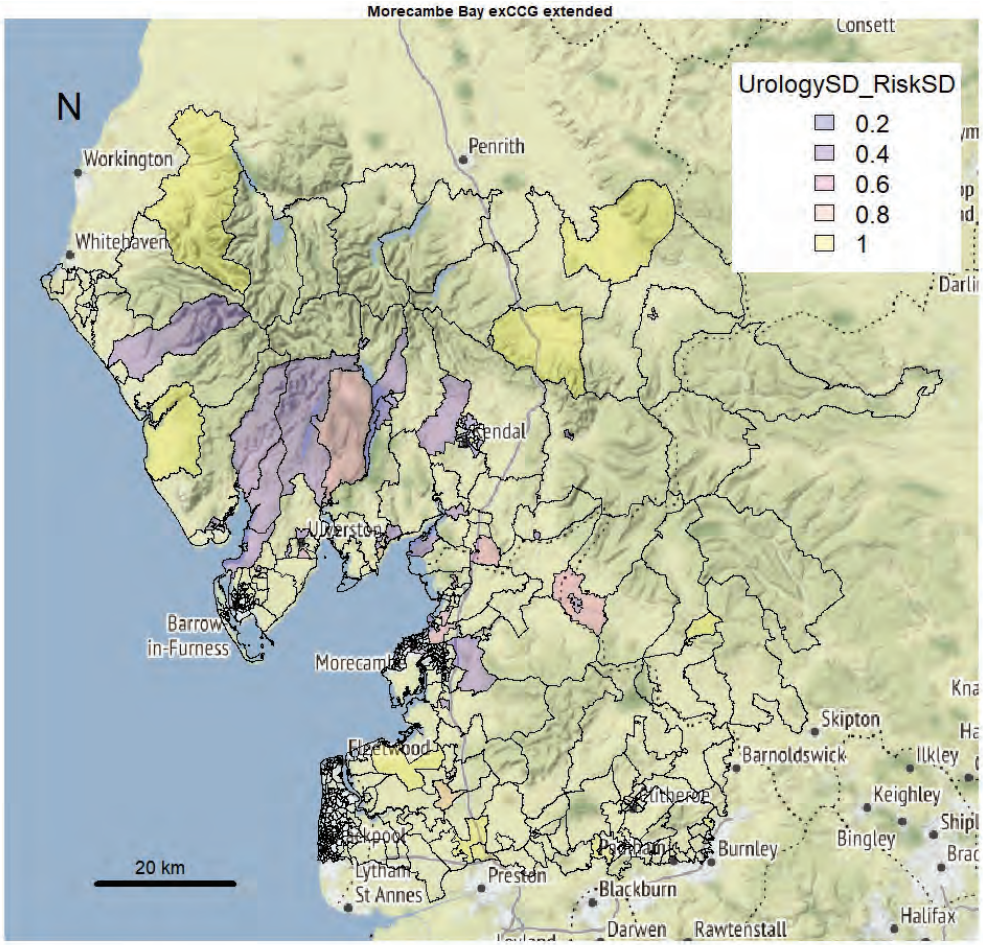
Morecambe Bay exCCG extended
N
UrologySD_RiskSD
0.2
0.4
0.6
0.8
1
20 km
Workington
Whitehaven
Penrith
Kendal
Ulverston
Barrow in-Furness
Morecambe
Fleetwood
Lytham St Annes
Preston
Blackburn
Burnley
Darwen
Rawtenstall
Skipton
Barnoldswick
Ilkley
Keighley
Bingley
Halifax
Consett

Lancaster and Morecambe

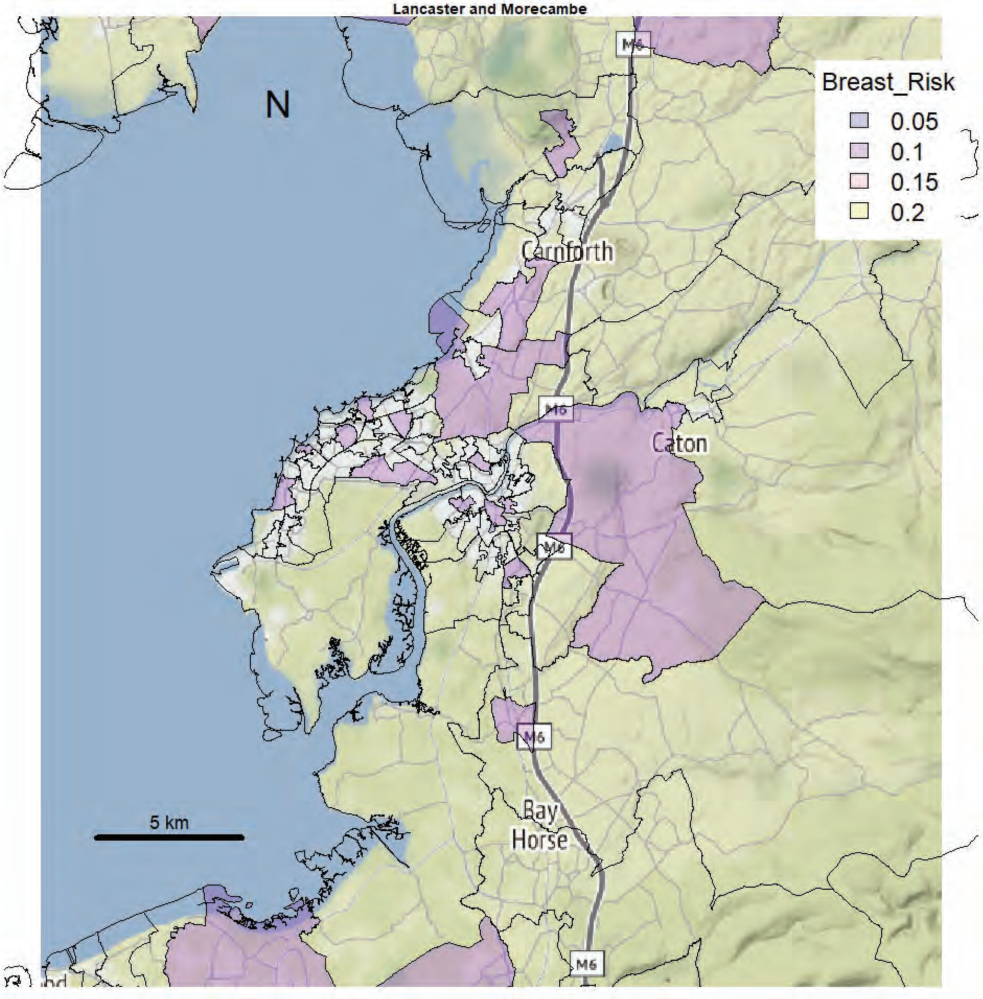


Lancaster and Morecambe

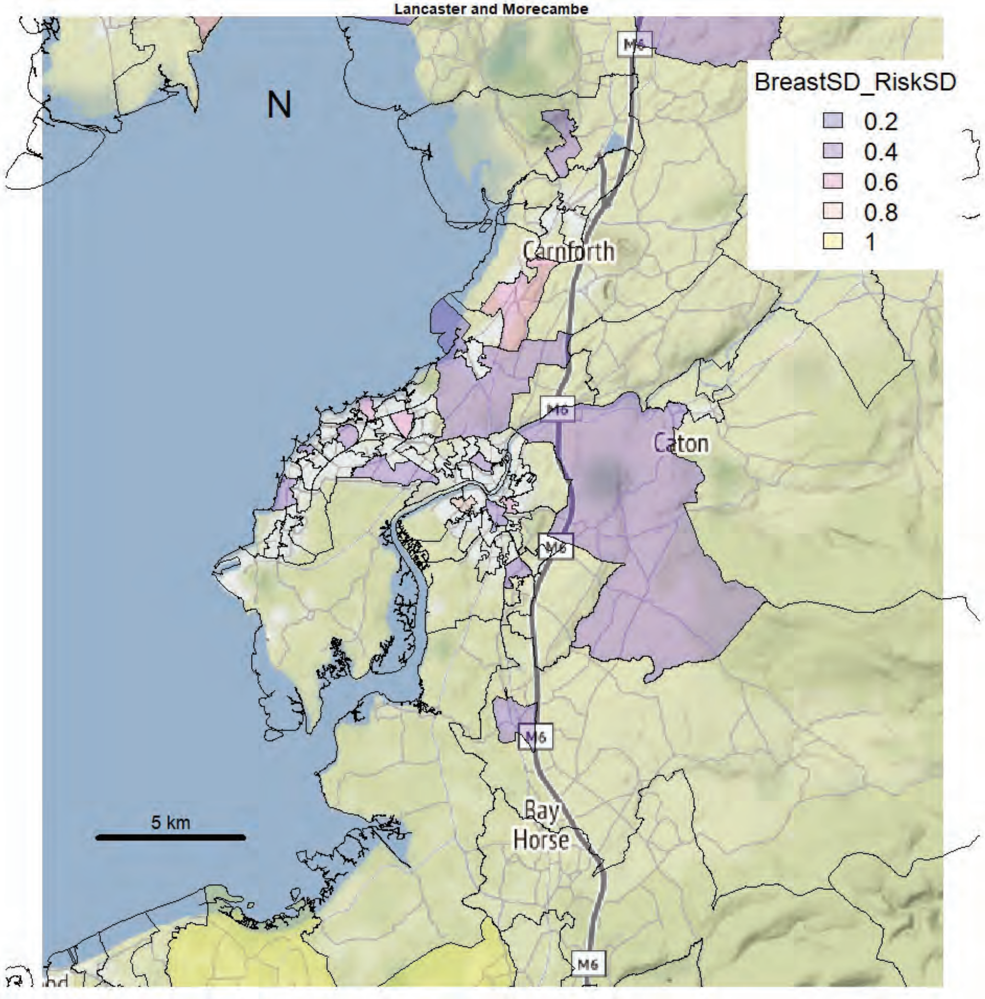

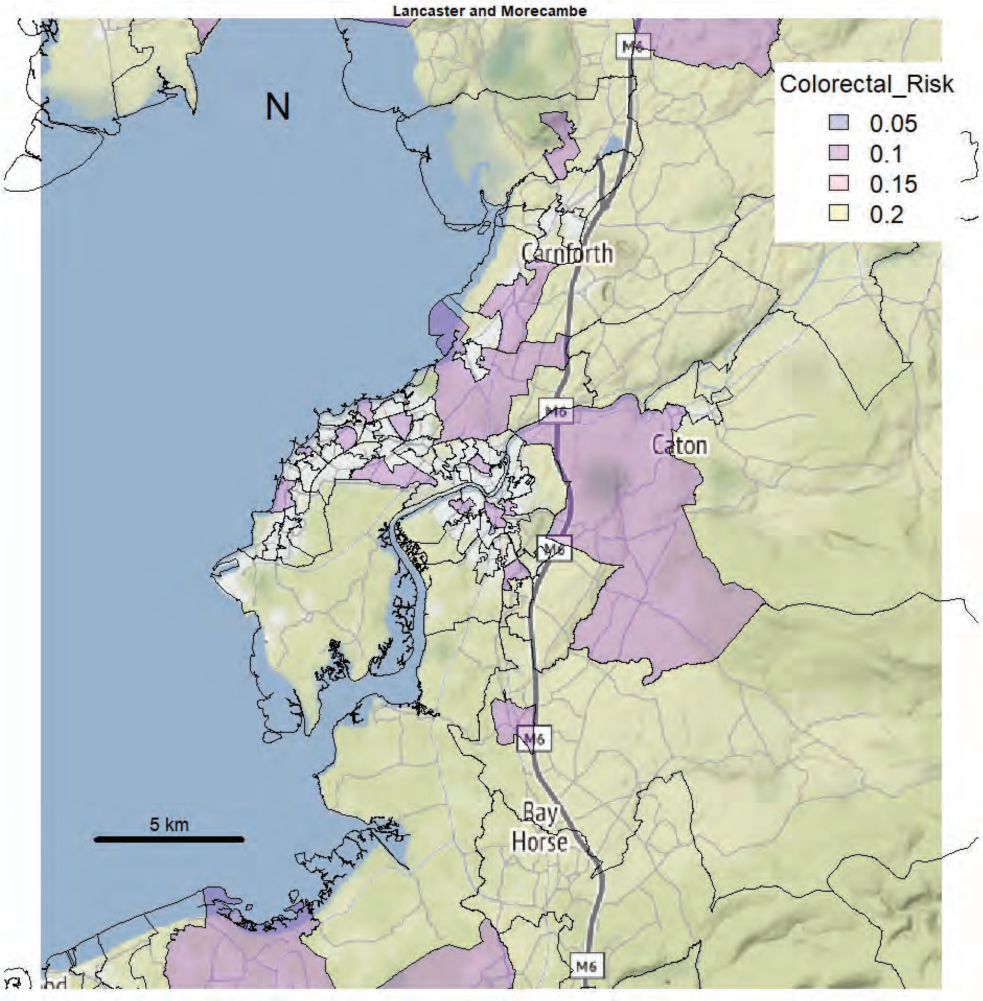
Lancaster and Morecambe
N
Colorectal_Risk
0.05
0.1
0.15
0.2
Carnforth
Caton
M6
Bay
Horse
5 km

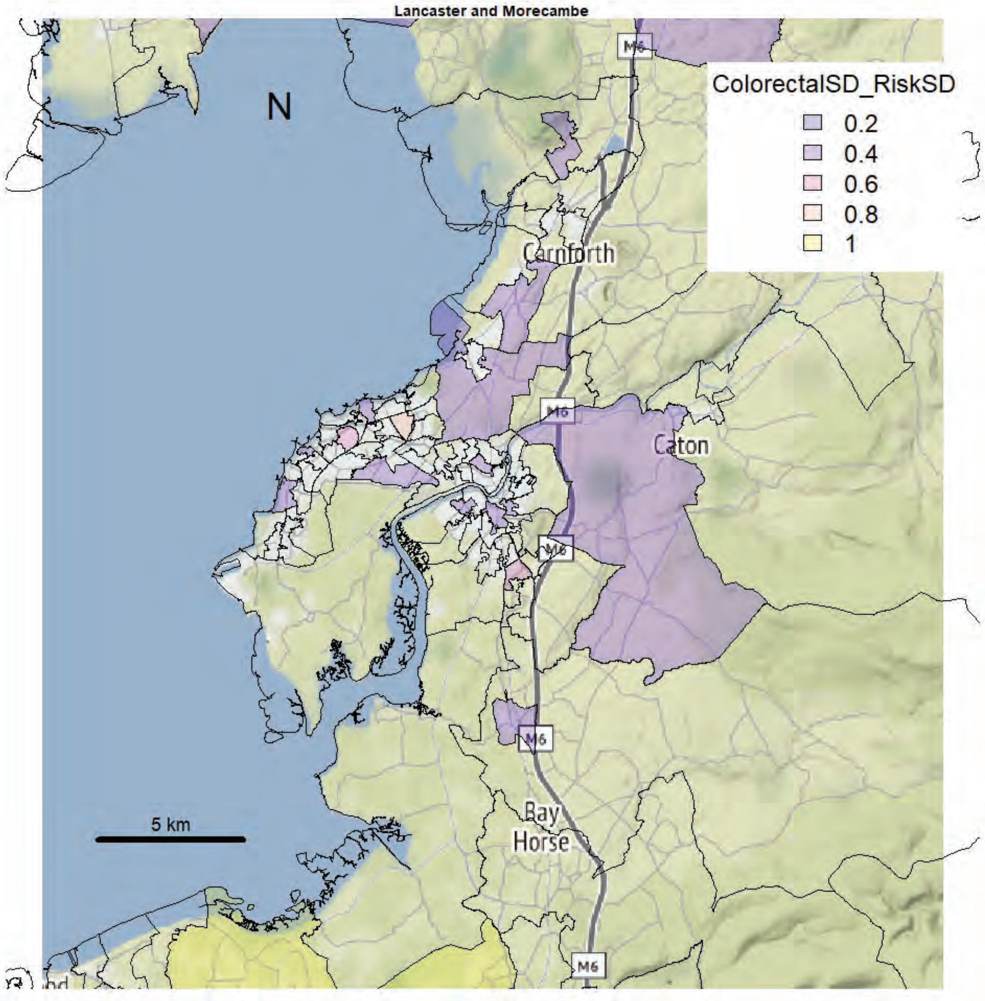
Lancaster and Morecambe
N
ColorectalSD_RiskSD
0.2
0.4
0.6
0.8
1
Carnforth
Caton
M6
Bay
Horse
5 km

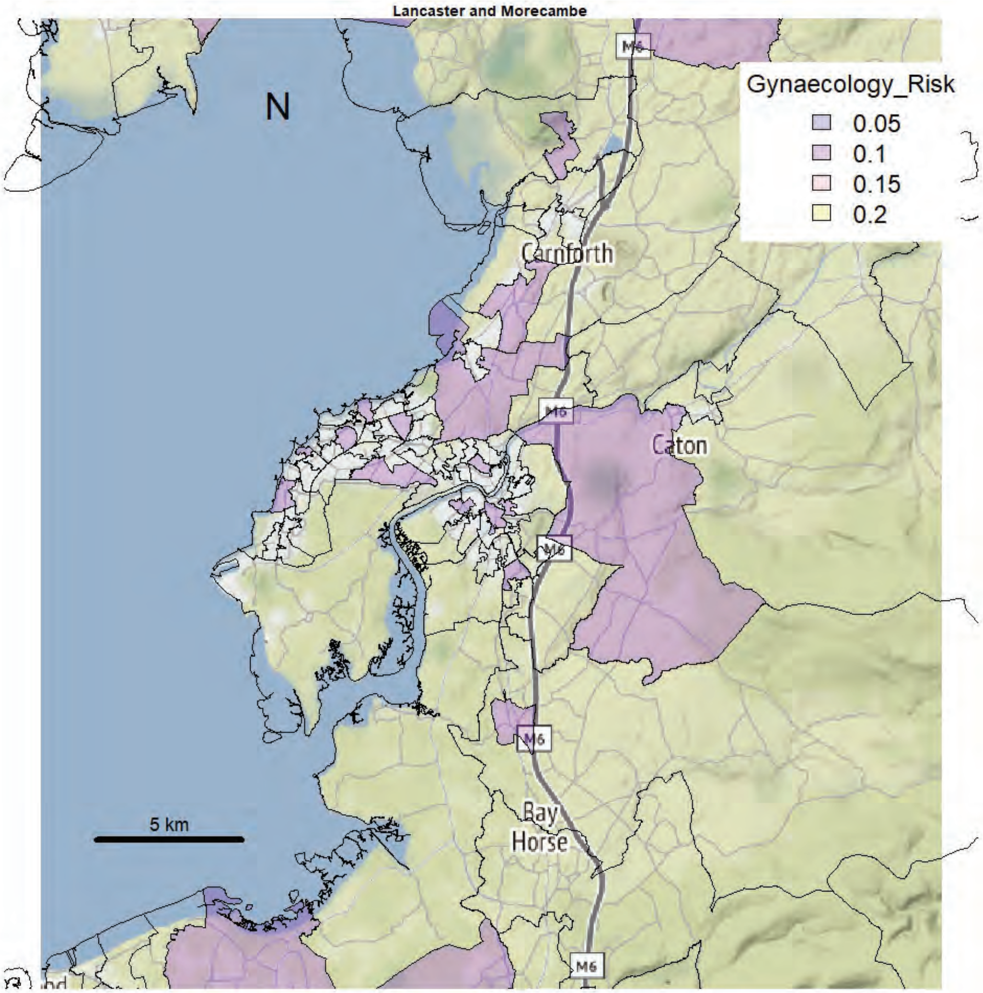
Lancaster and Morecambe
N
Gynaecology_Risk
0.05
0.1
0.15
0.2
Carnforth
Caton
M6
Bay
Horse
5 km

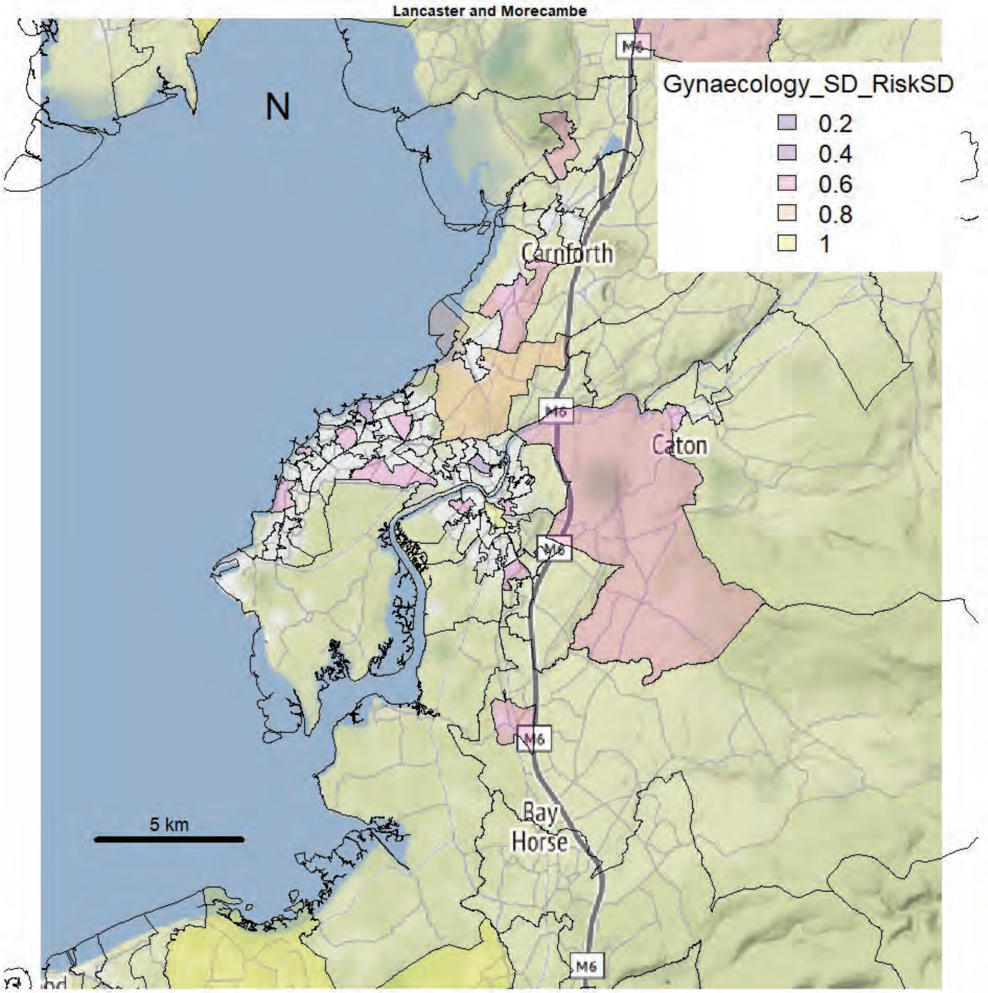
Lancaster and Morecambe
N
Gynaecology_SD_RiskSD
0.2
0.4
0.6
0.8
1
Carnforth
Caton
M6
Bay
Horse
5 km

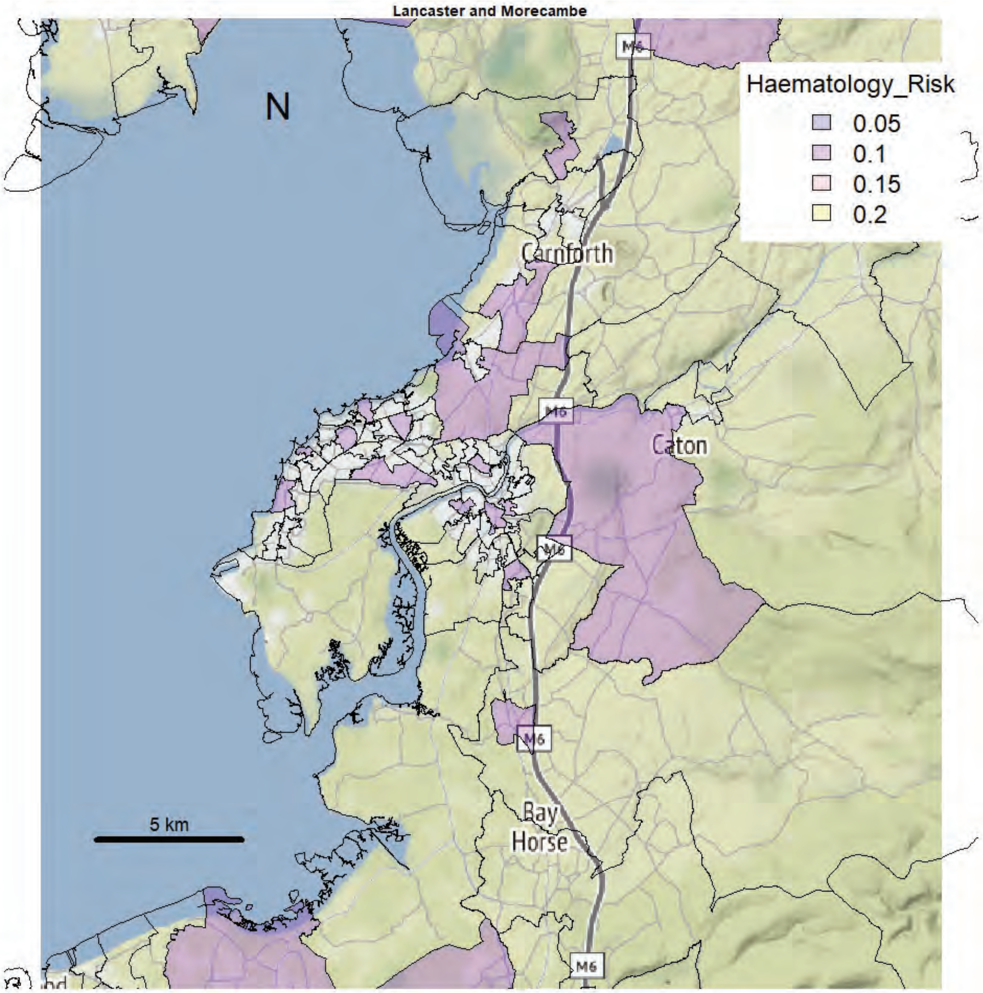
Lancaster and Morecambe
N
Haematology_Risk
0.05
0.1
0.15
0.2
Carnforth
Caton
Bay Horse
M6
5 km

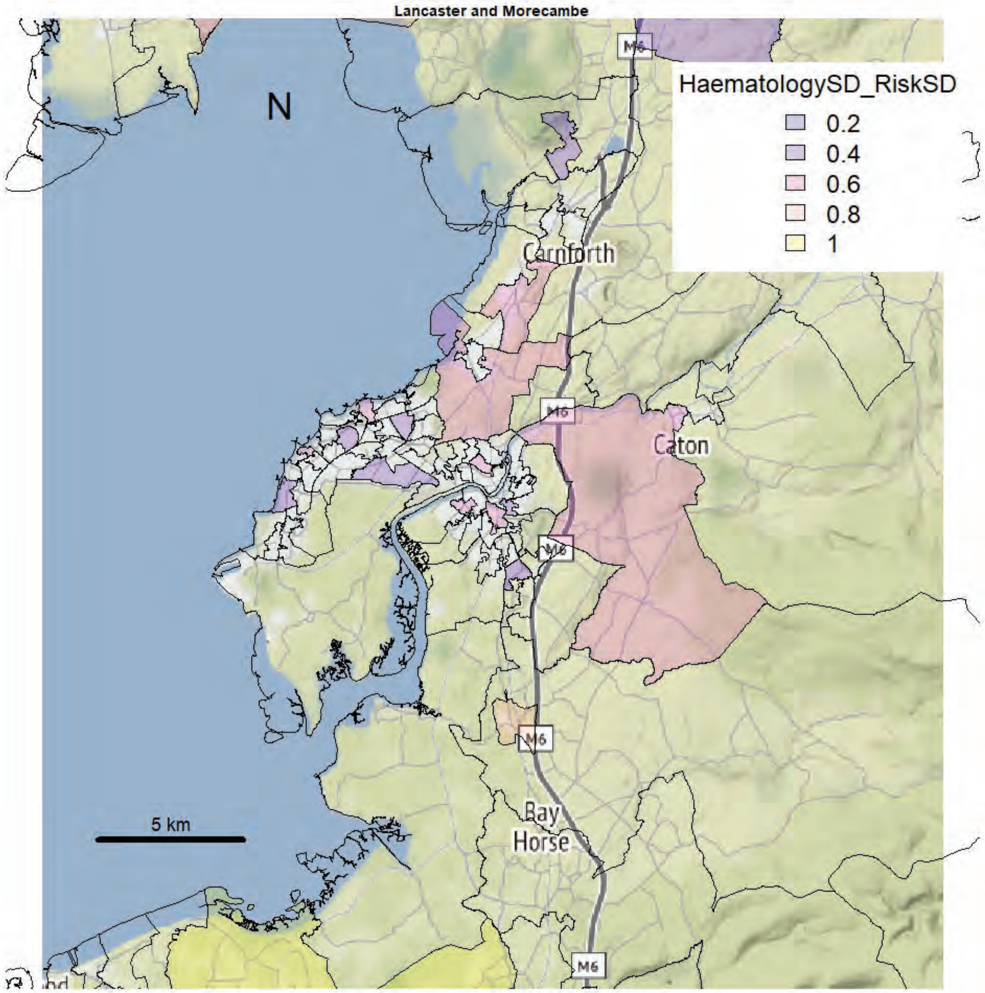
Lancaster and Morecambe
N
HaematologySD_RiskSD
0.2
0.4
0.6
0.8
1
Carnforth
Caton
Bay Horse
M6
5 km

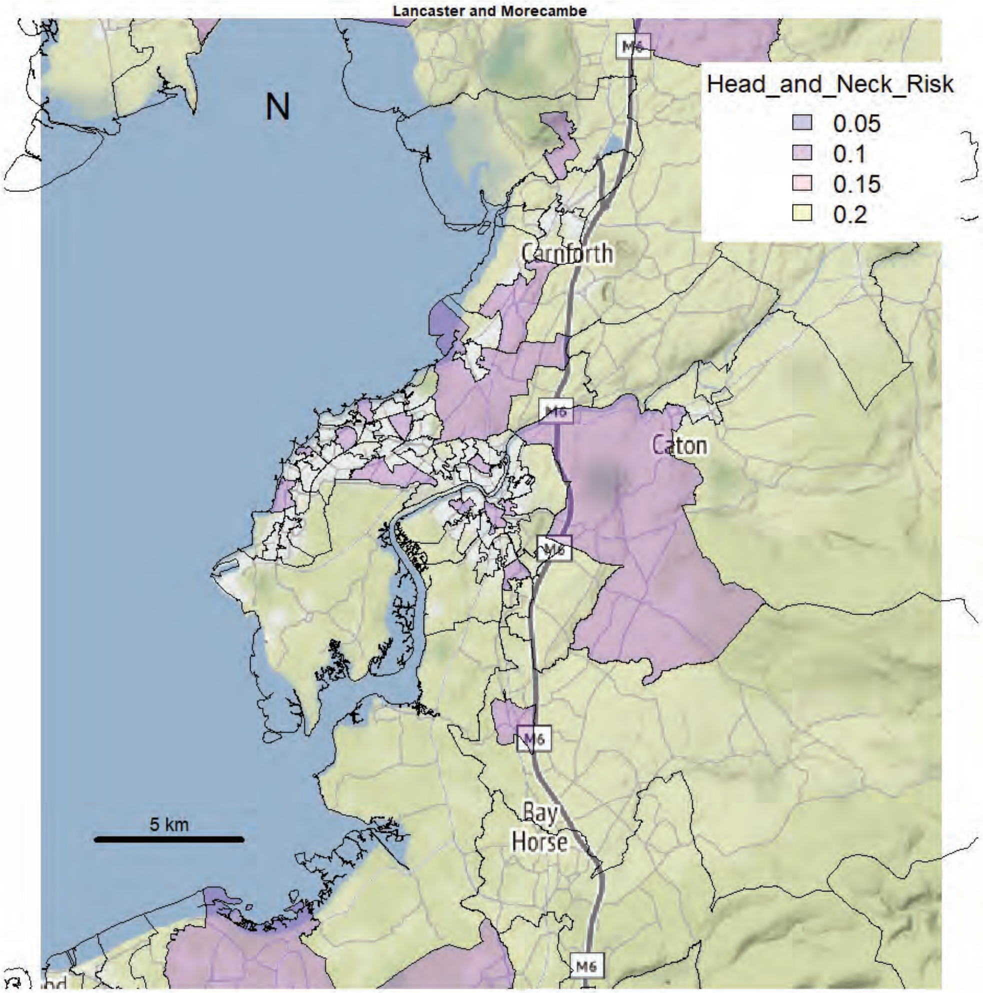
Lancaster and Morecambe
N
Head_and_Neck_Risk
0.05
0.1
0.15
0.2
Carnforth
Caton
Bay
Horse
M6
5 km

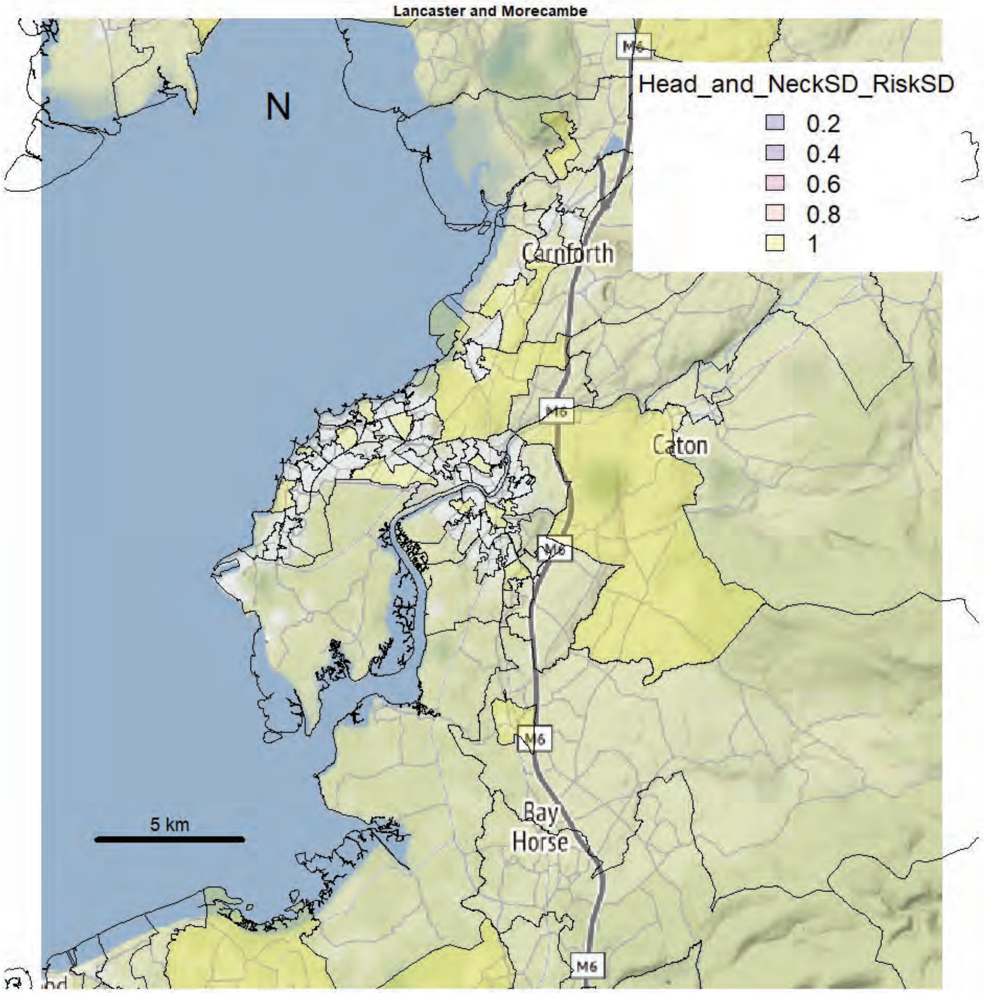
Lancaster and Morecambe
N
Head_and_NeckSD_RiskSD
0.2
0.4
0.6
0.8
1
Carnforth
Caton
Bay
Horse
M6
5 km

Lancaster and Morecambe

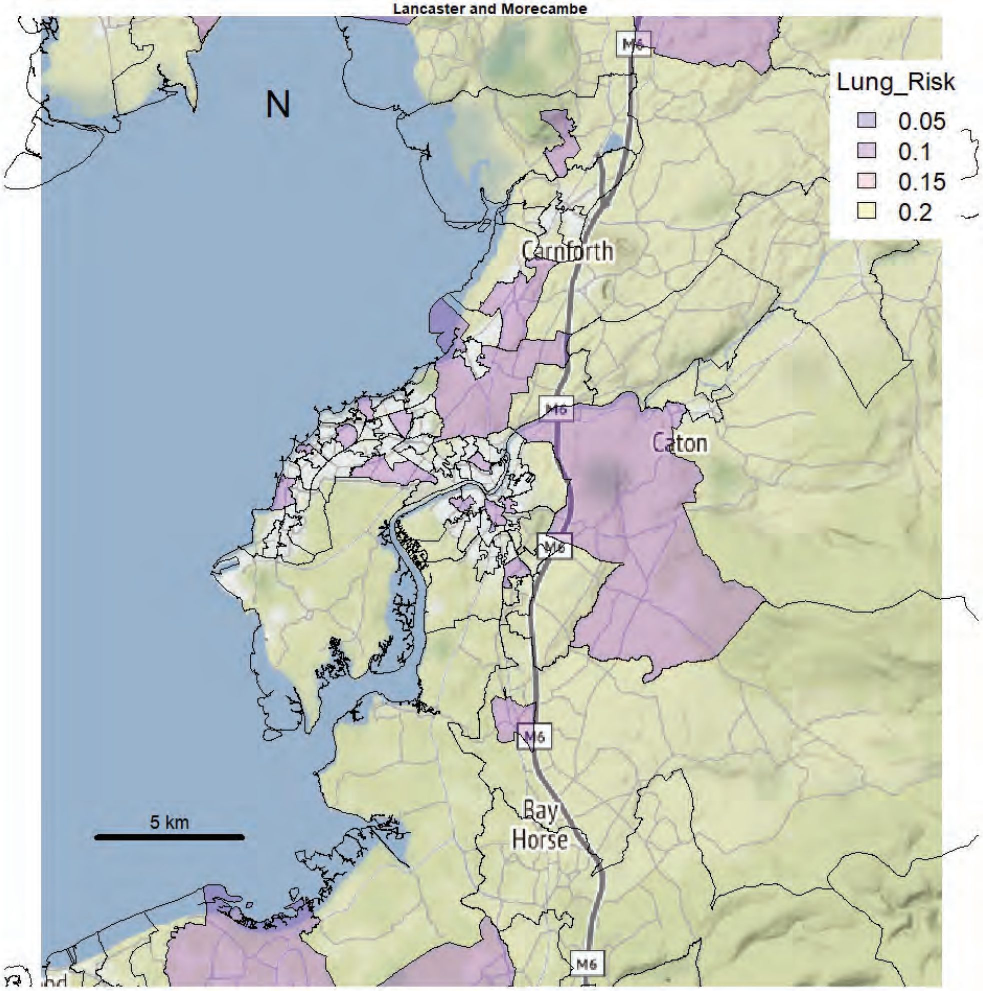


Lancaster and Morecambe

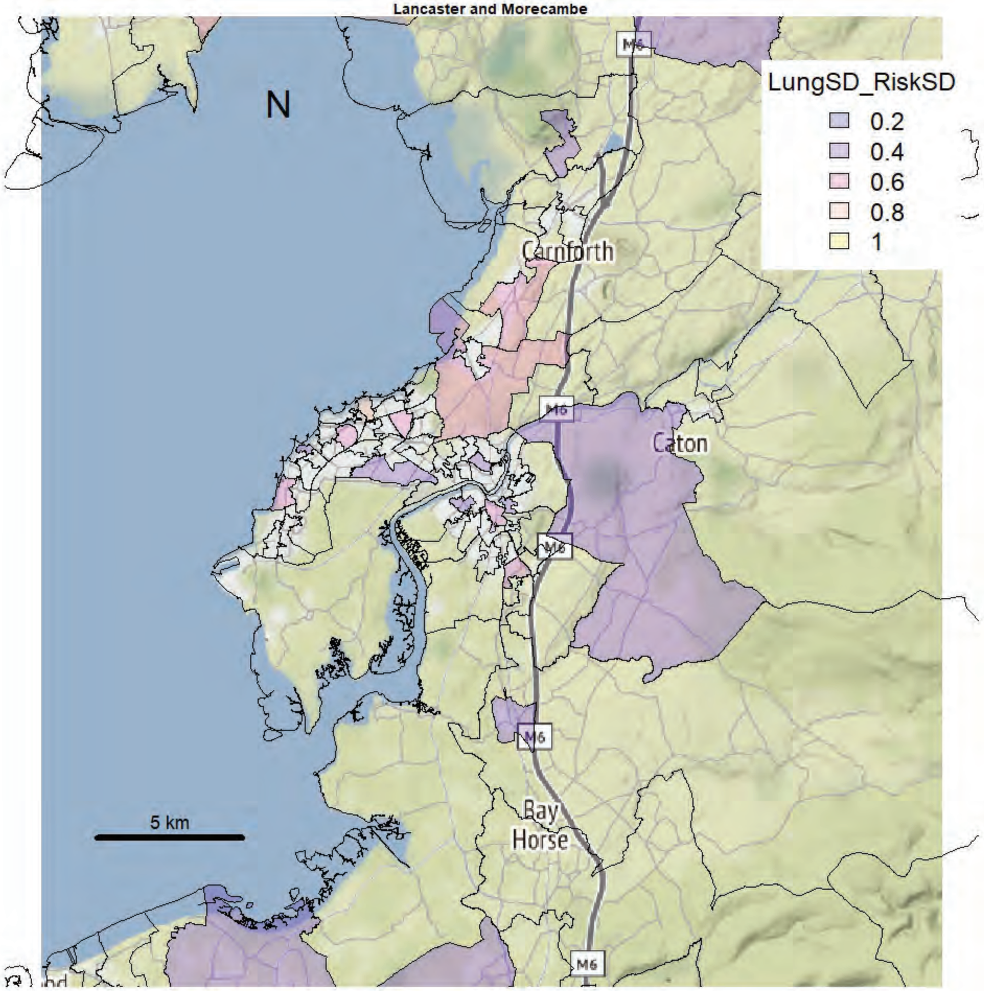

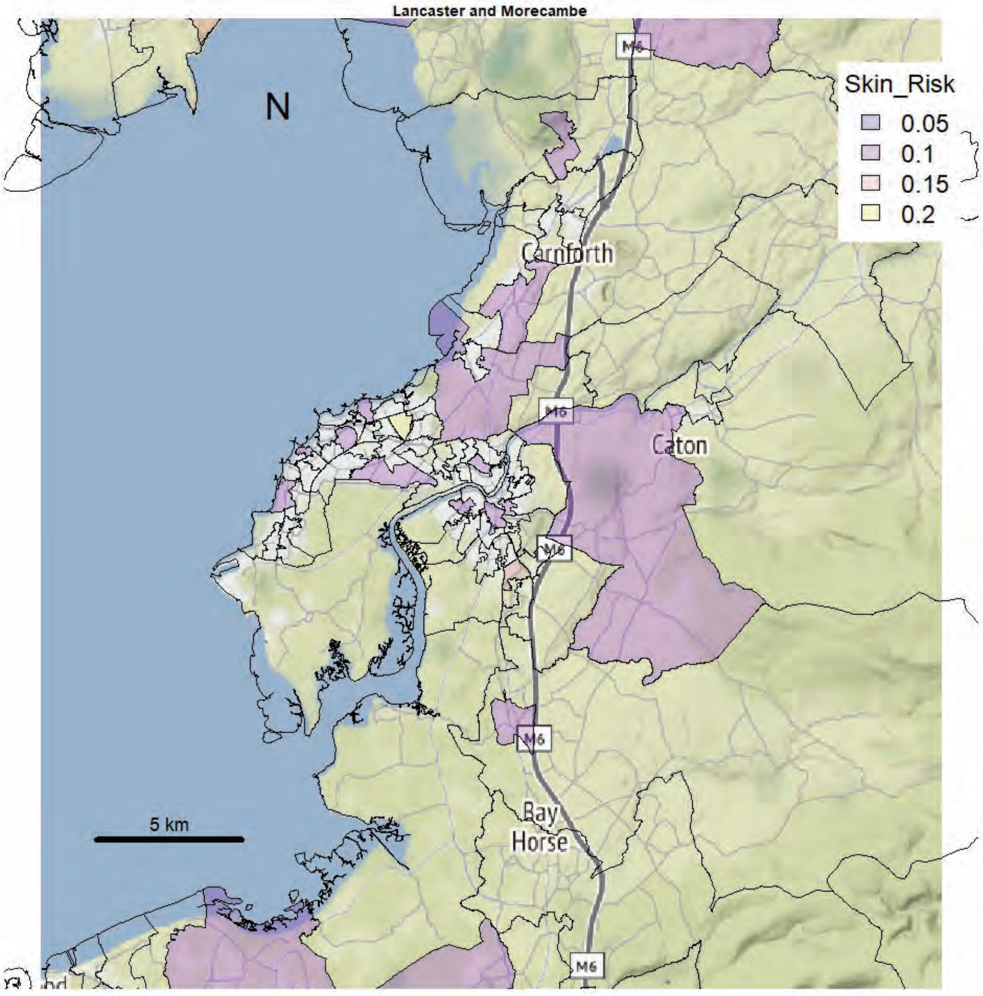
Lancaster and Morecambe
N
Skin_Risk
0.05
0.1
0.15
0.2
Carnforth
Caton
Bay
Horse
M6
5 km

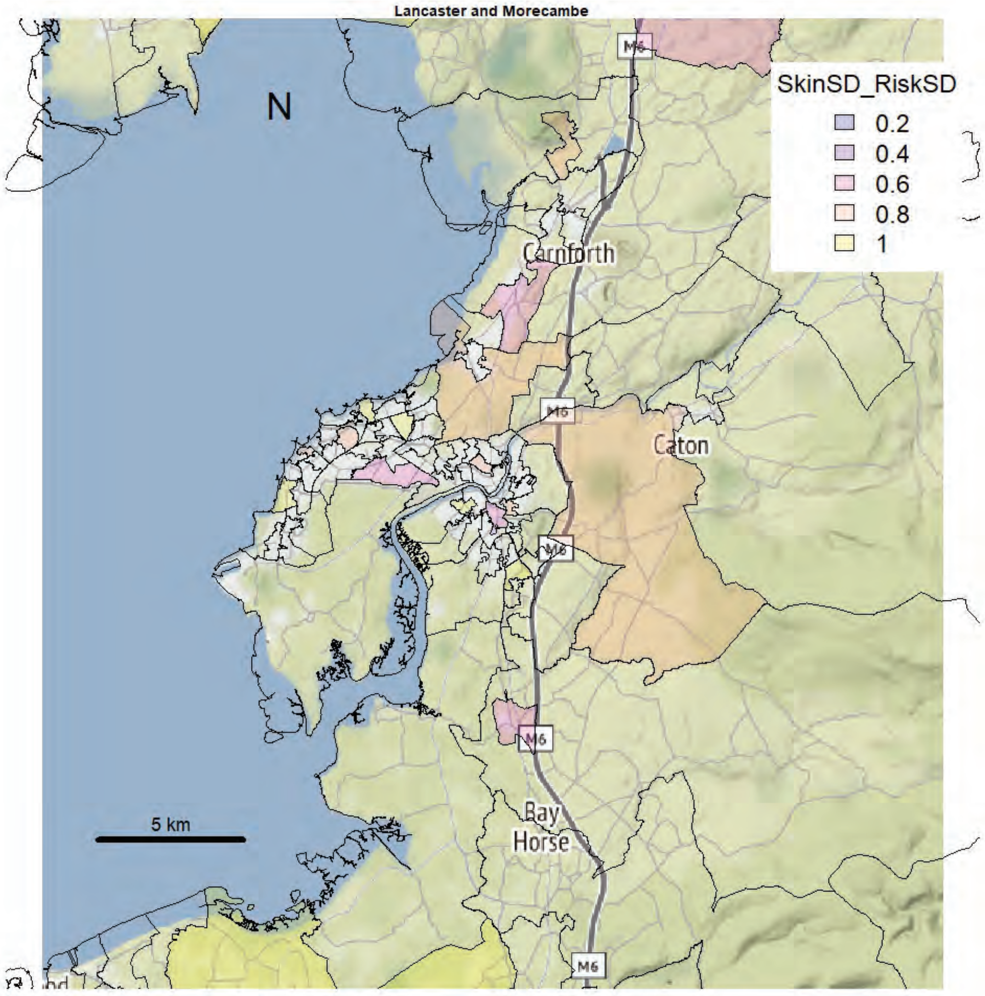
Lancaster and Morecambe
N
SkinSD_RiskSD
0.2
0.4
0.6
0.8
1
Carnforth
Caton
Bay
Horse
M6
5 km

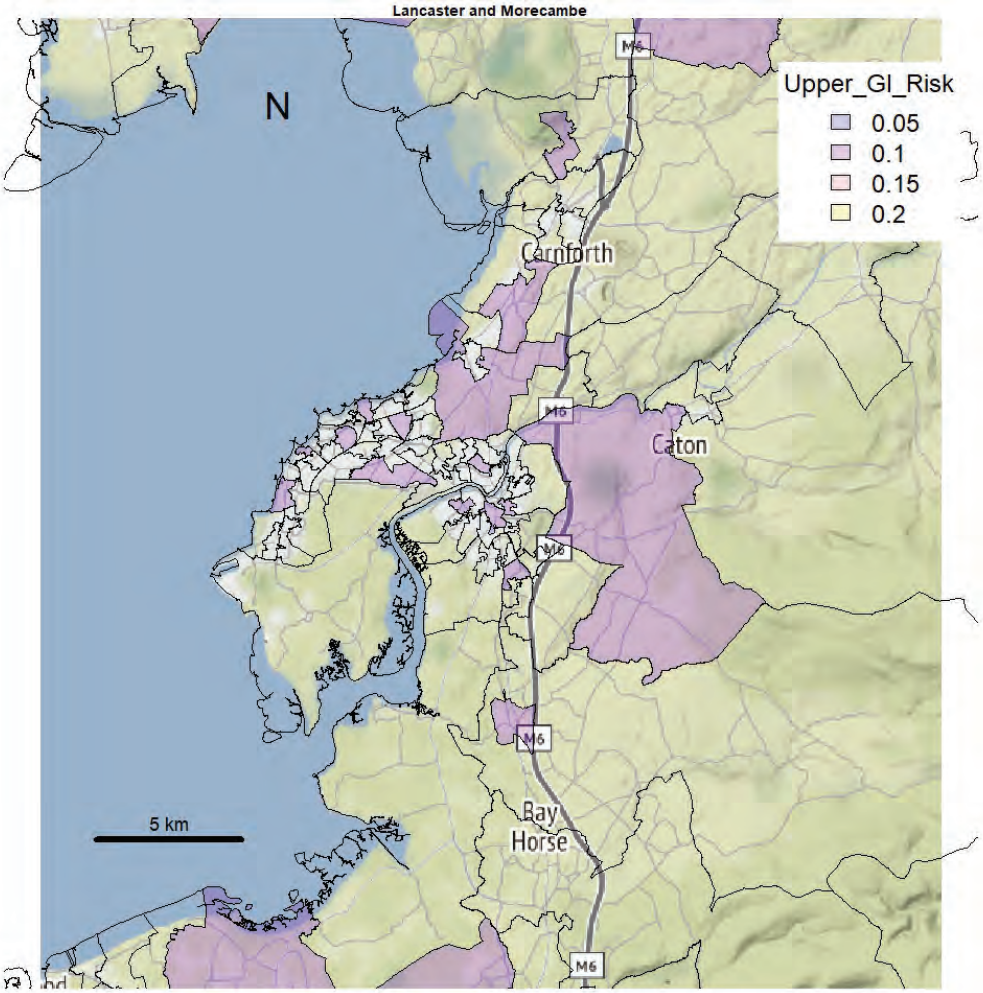
Lancaster and Morecambe
N
Upper_GI_Risk
0.05
0.1
0.15
0.2
Carnforth
Caton
M6
Bay Horse
5 km

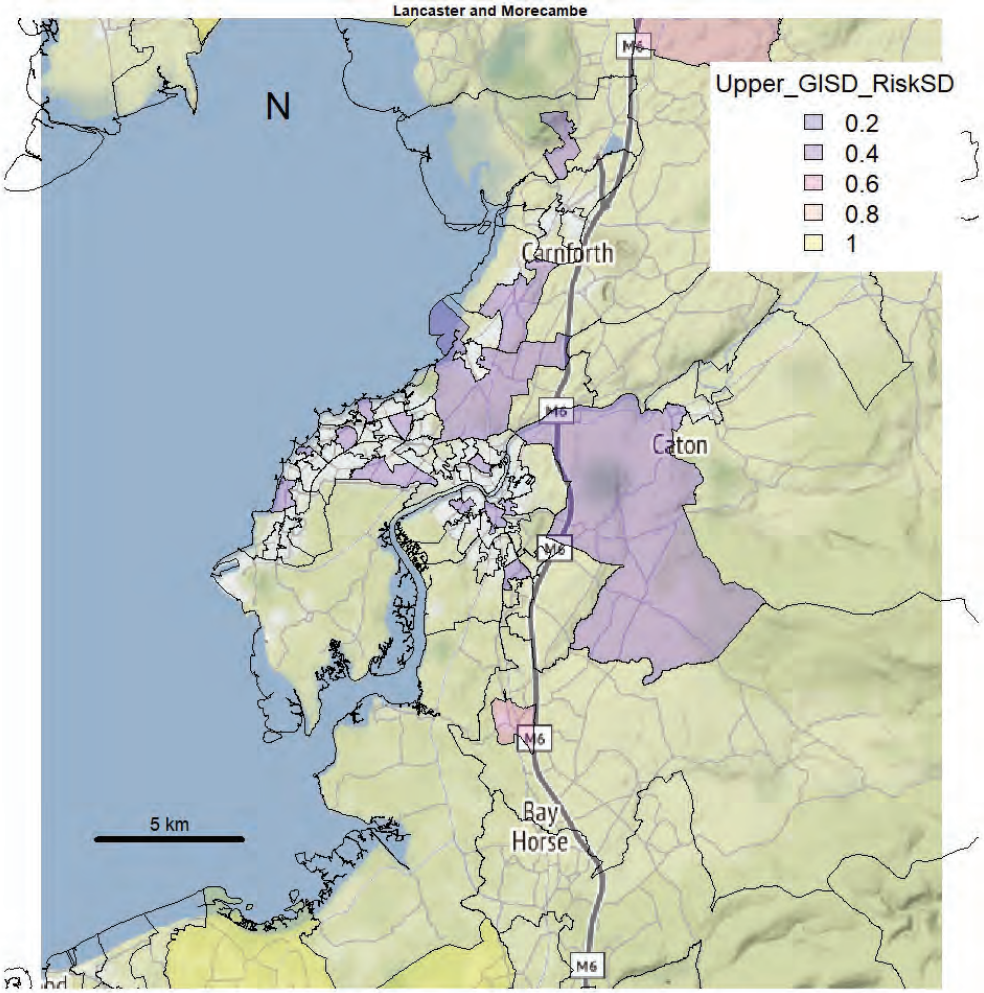
Lancaster and Morecambe
N
Upper_GISD_RiskSD
0.2
0.4
0.6
0.8
1
Carnforth
Caton
M6
Bay Horse
5 km

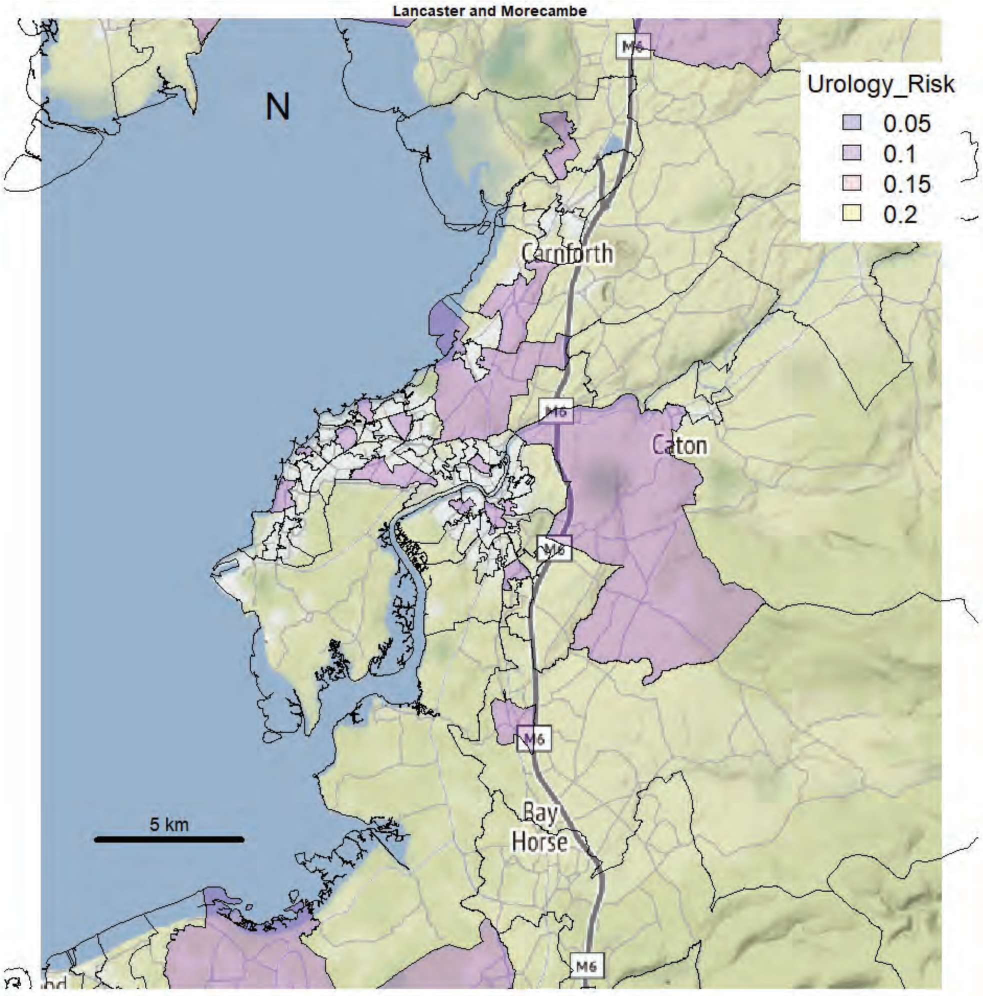
Lancaster and Morecambe
N
Urology_Risk
0.05
0.1
0.15
0.2
M6
Carnforth
Caton
Bay
Horse
5 km

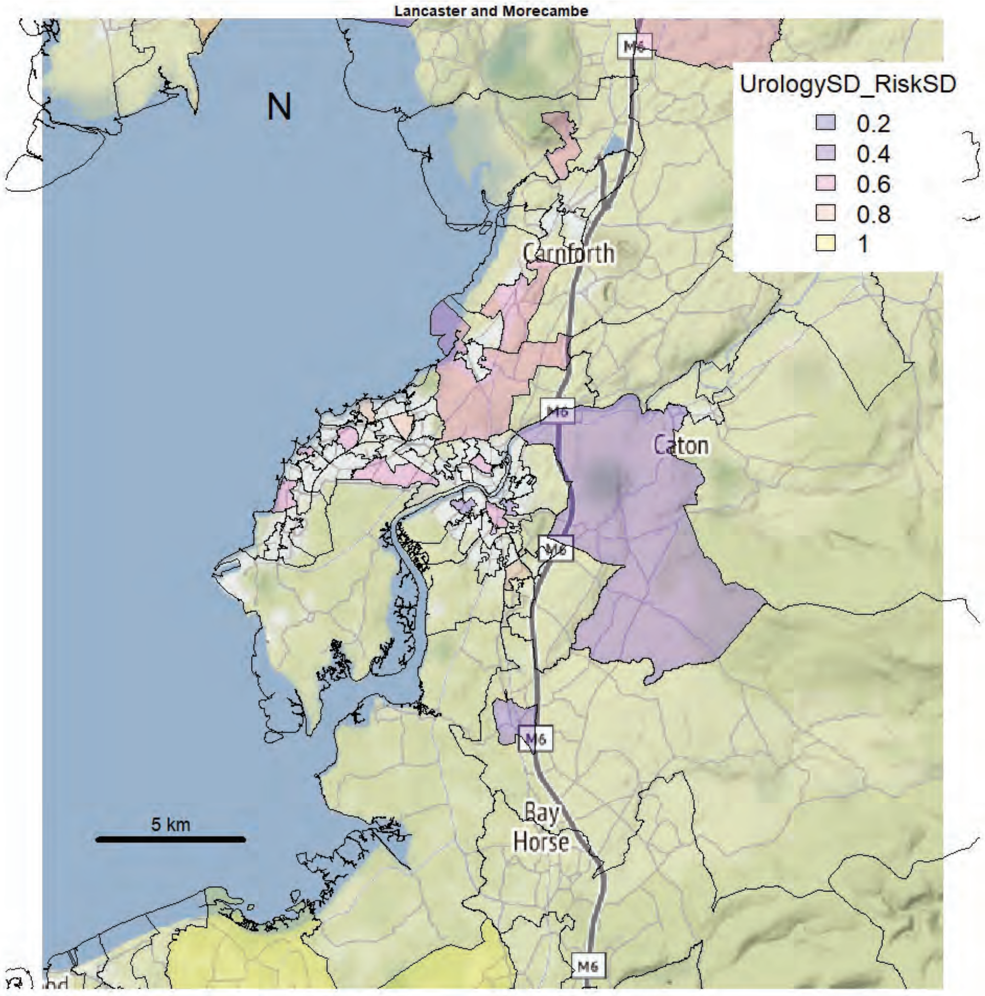
Lancaster and Morecambe
N
UrologySD_RiskSD
0.2
0.4
0.6
0.8
1
M6
Carnforth
Caton
Bay
Horse
5 km

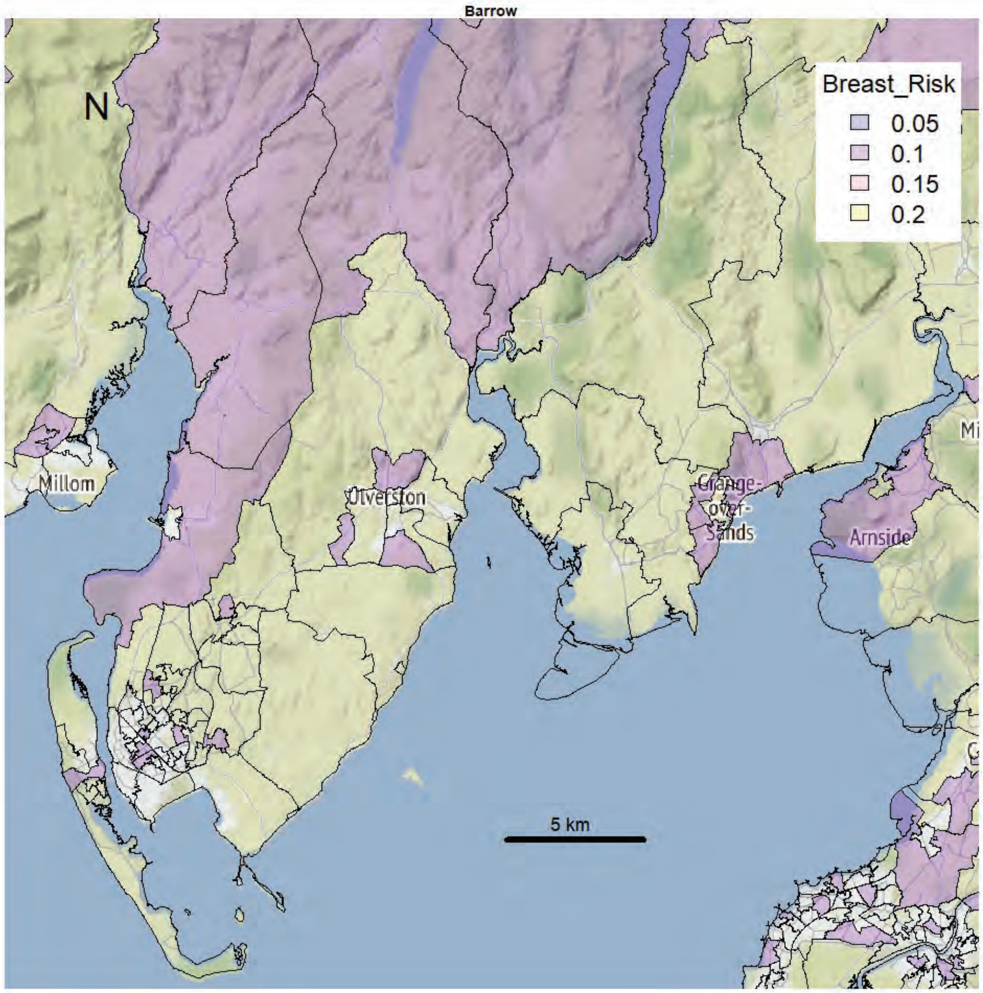
Barrow
N
Breast_Risk
0.05
0.1
0.15
0.2
Millom
Ulverston
Grange-over-Sands
Arnside
5 km

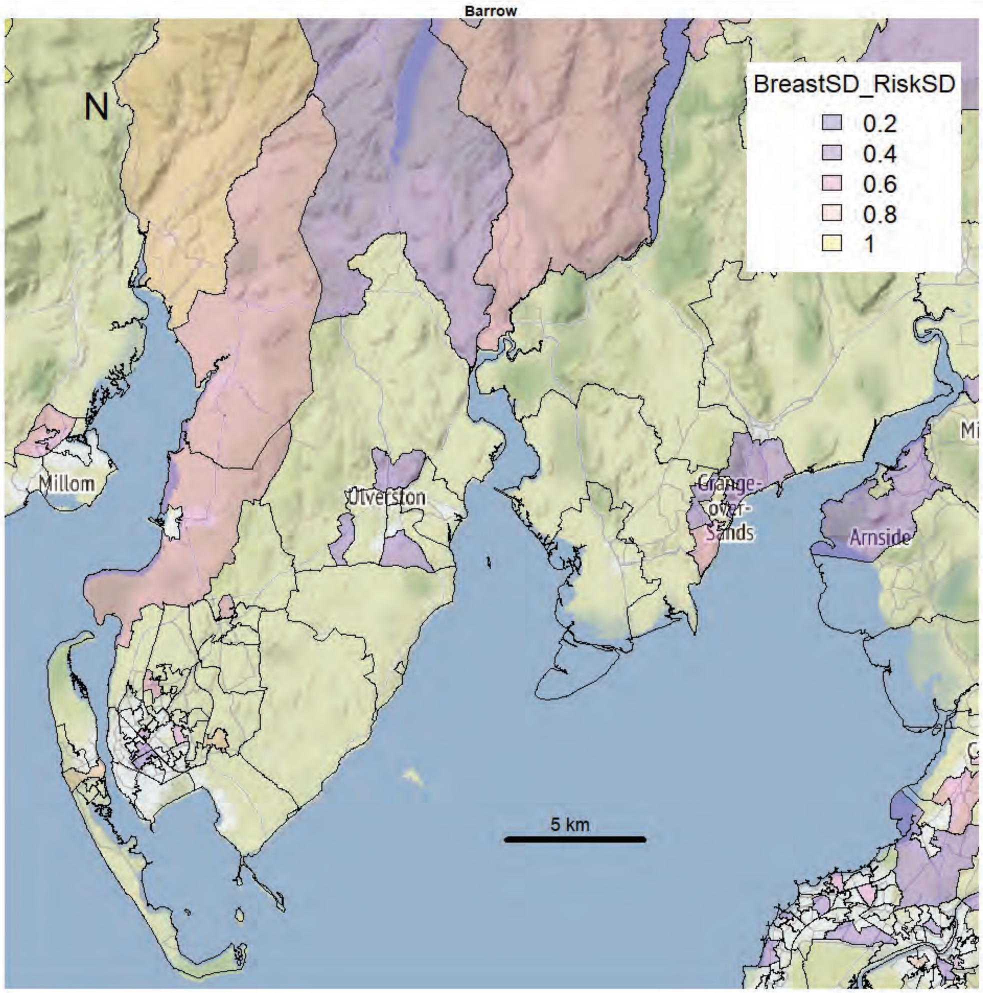
Barrow
N
BreastSD_RiskSD
0.2
0.4
0.6
0.8
1
Millom
Ulverston
Grange-over-Sands
Arnside
5 km

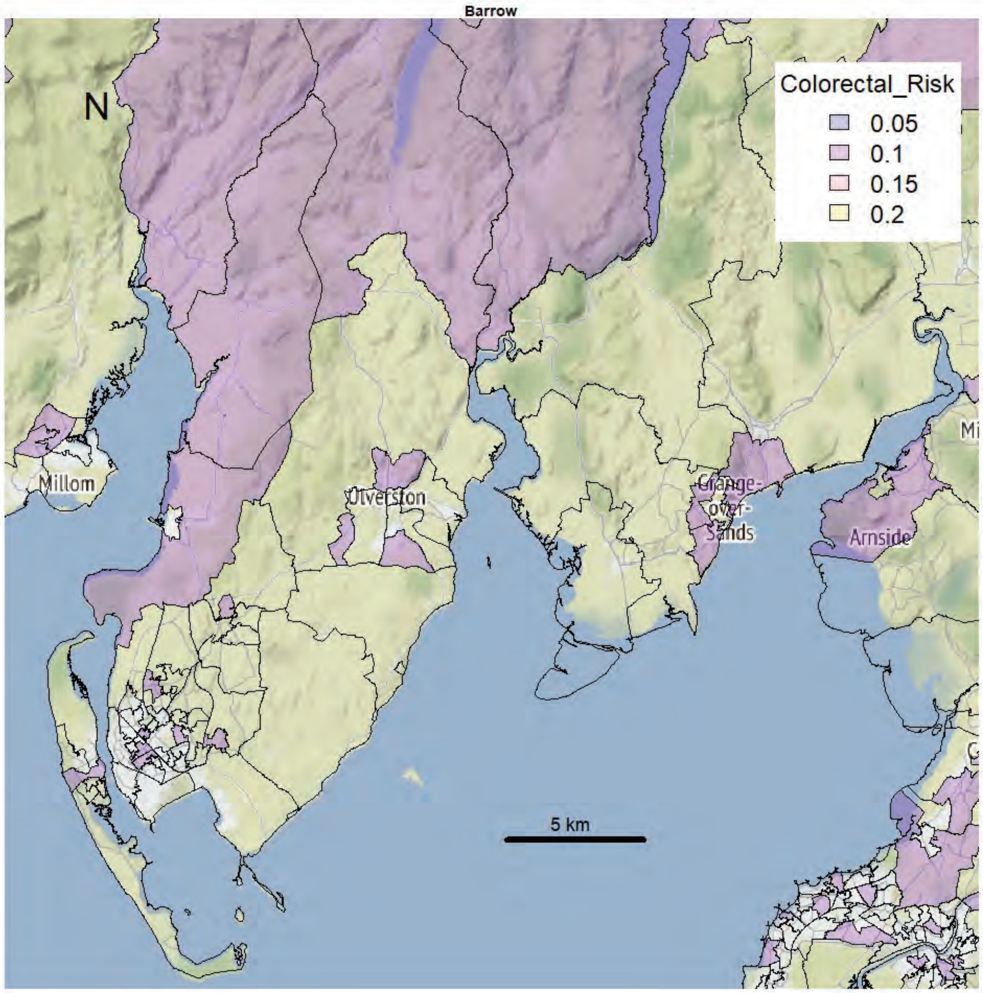
Barrow
N
Colorectal_Risk
0.05
0.1
0.15
0.2
Millom
Ulverston
Grange-over-Sands
Arnside
5 km

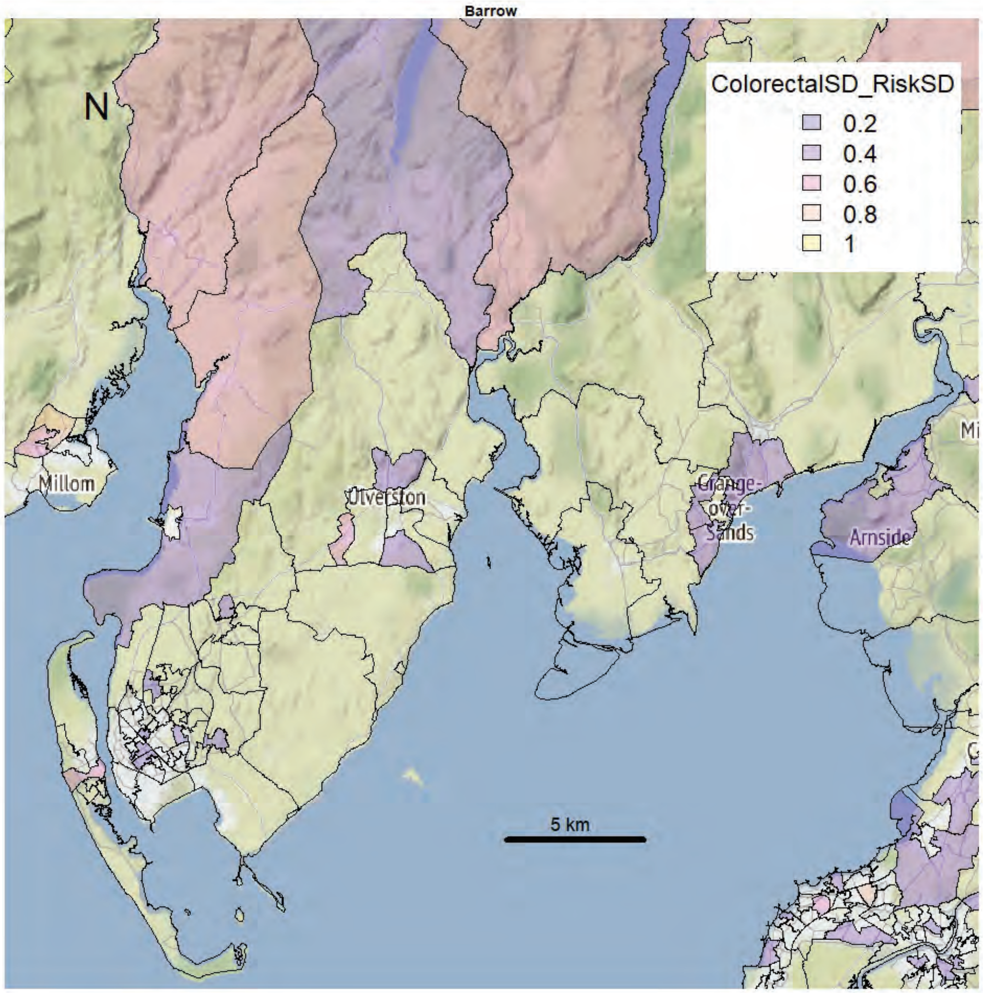
Barrow
N
ColorectalSD_RiskSD
0.2
0.4
0.6
0.8
1
Millom
Ulverston
Grange-over-Sands
Arnside
5 km

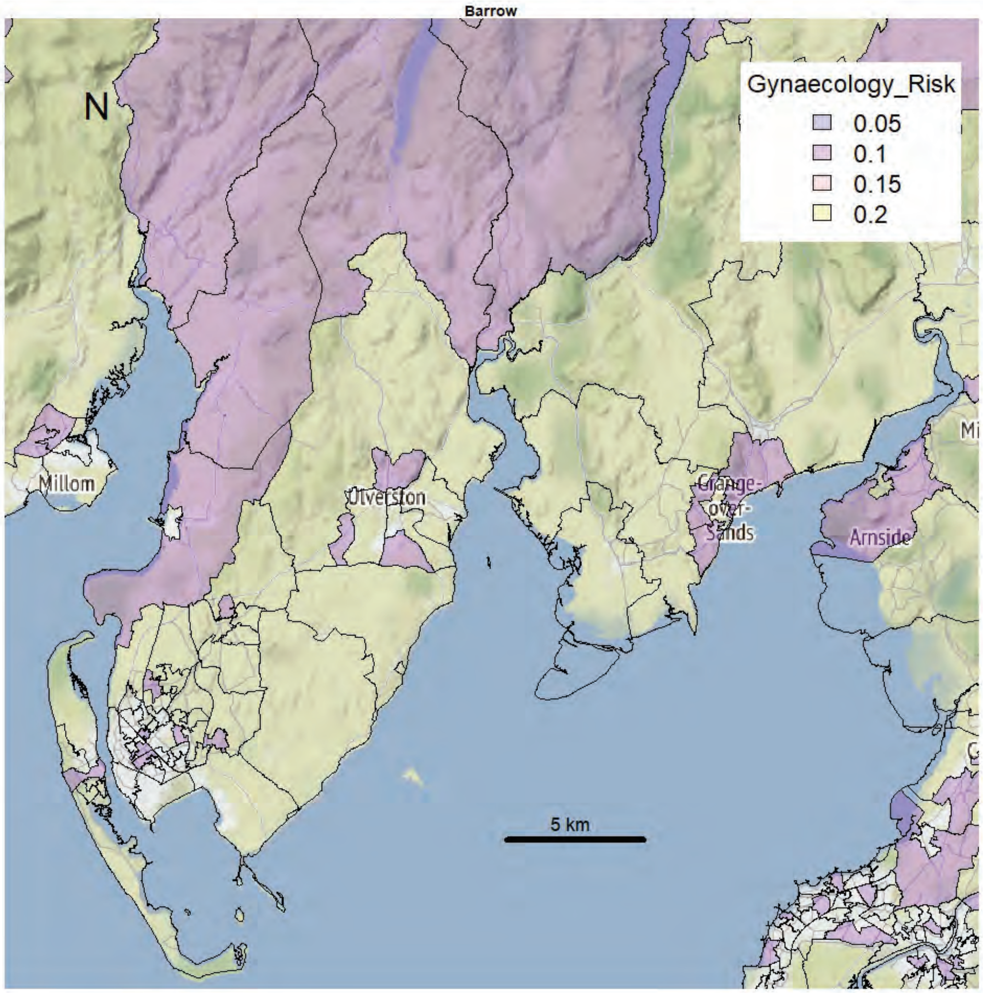
Barrow
N
Gynaecology_Risk
0.05
0.1
0.15
0.2
Millom
Ulverston
Grange-over-Sands
Arnside
5 km

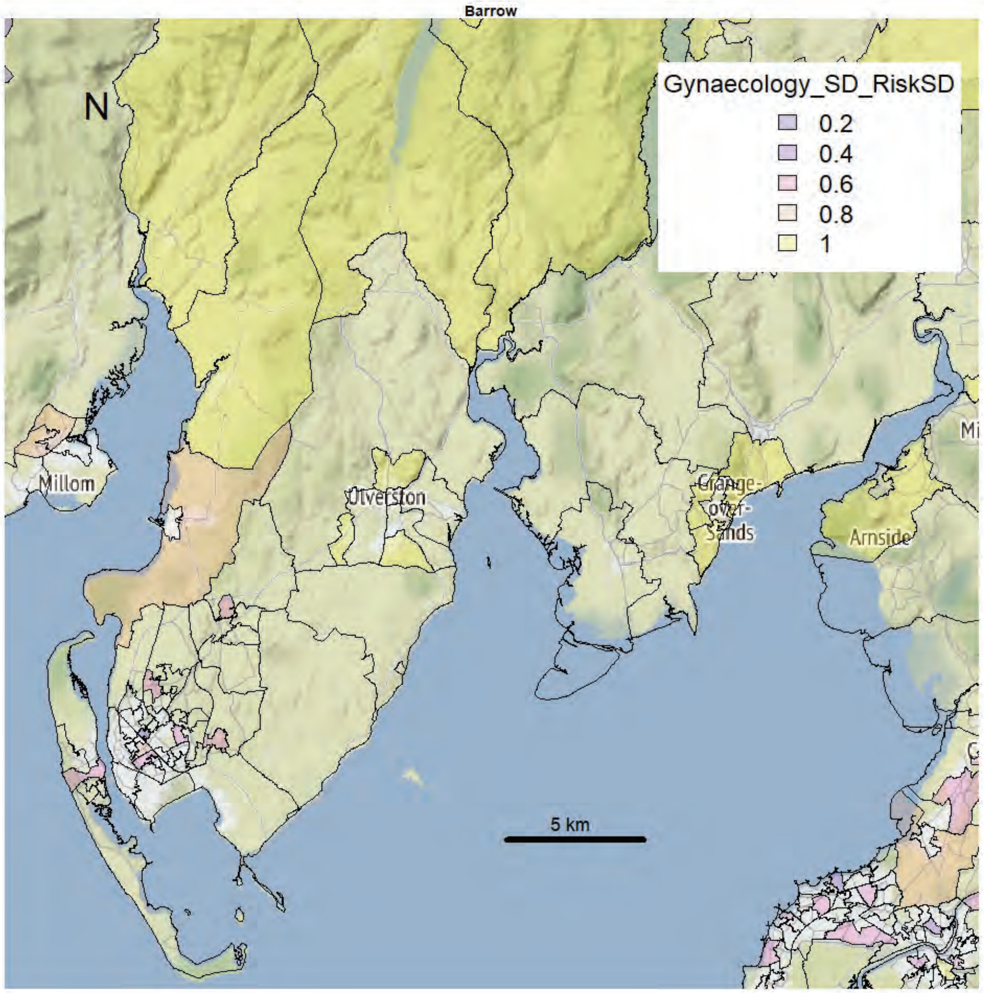
Barrow
N
Gynaecology_SD_RiskSD
0.2
0.4
0.6
0.8
1
Millom
Ulverston
Grange-over-Sands
Arnside
5 km

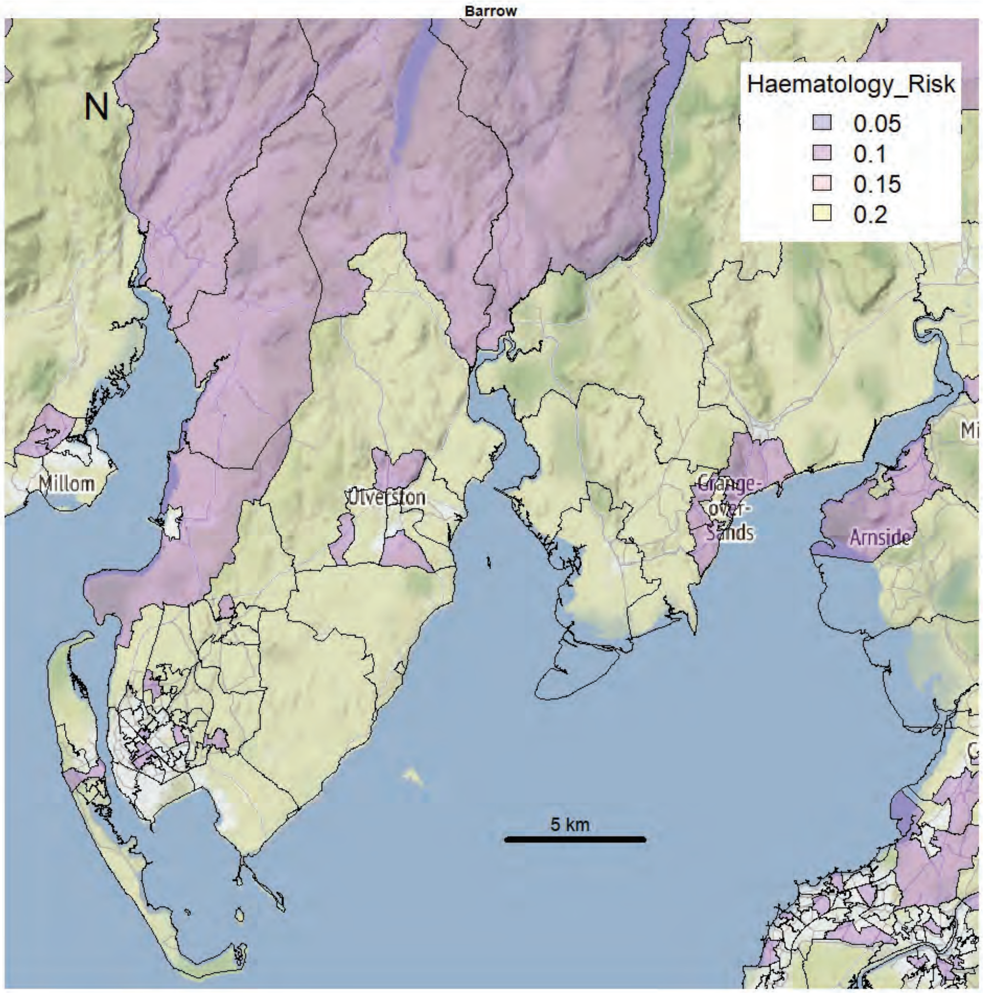
Barrow
N
Haematology_Risk
0.05
0.1
0.15
0.2
Millom
Ulverston
Grange-over-Sands
Arnside
5 km

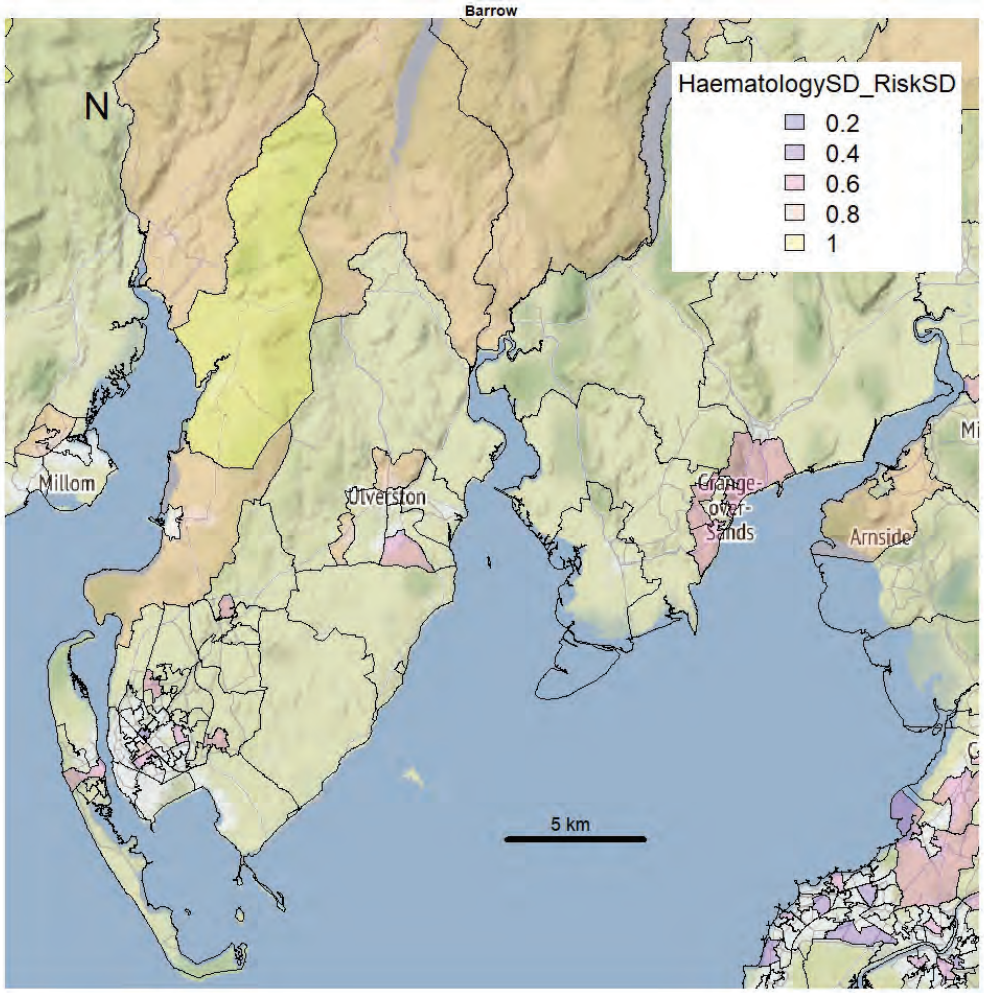
Barrow
N
HaematologySD_RiskSD
0.2
0.4
0.6
0.8
1
Millom
Ulverston
Grange-over-Sands
Arnside
5 km

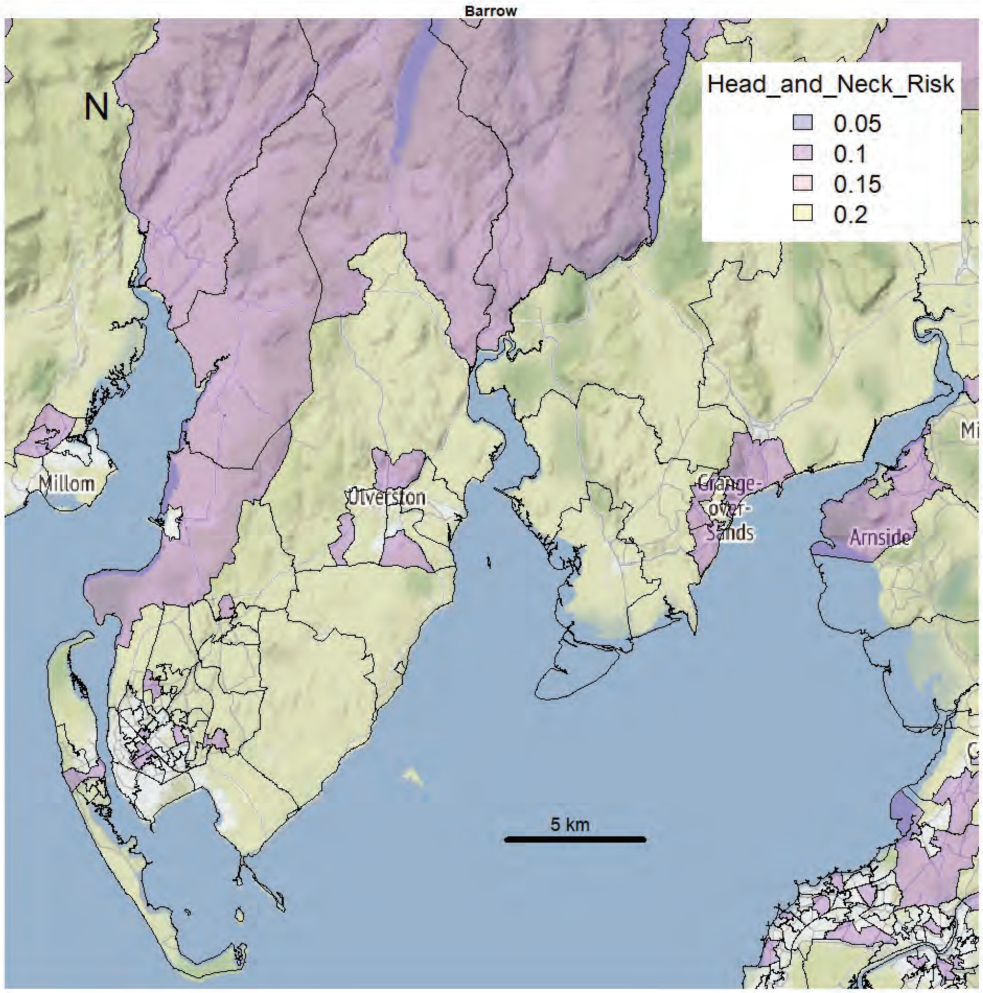
Barrow
N
Head_and_Neck_Risk
0.05
0.1
0.15
0.2
Millom
Ulverston
Grange-over-Sands
Arnside
5 km

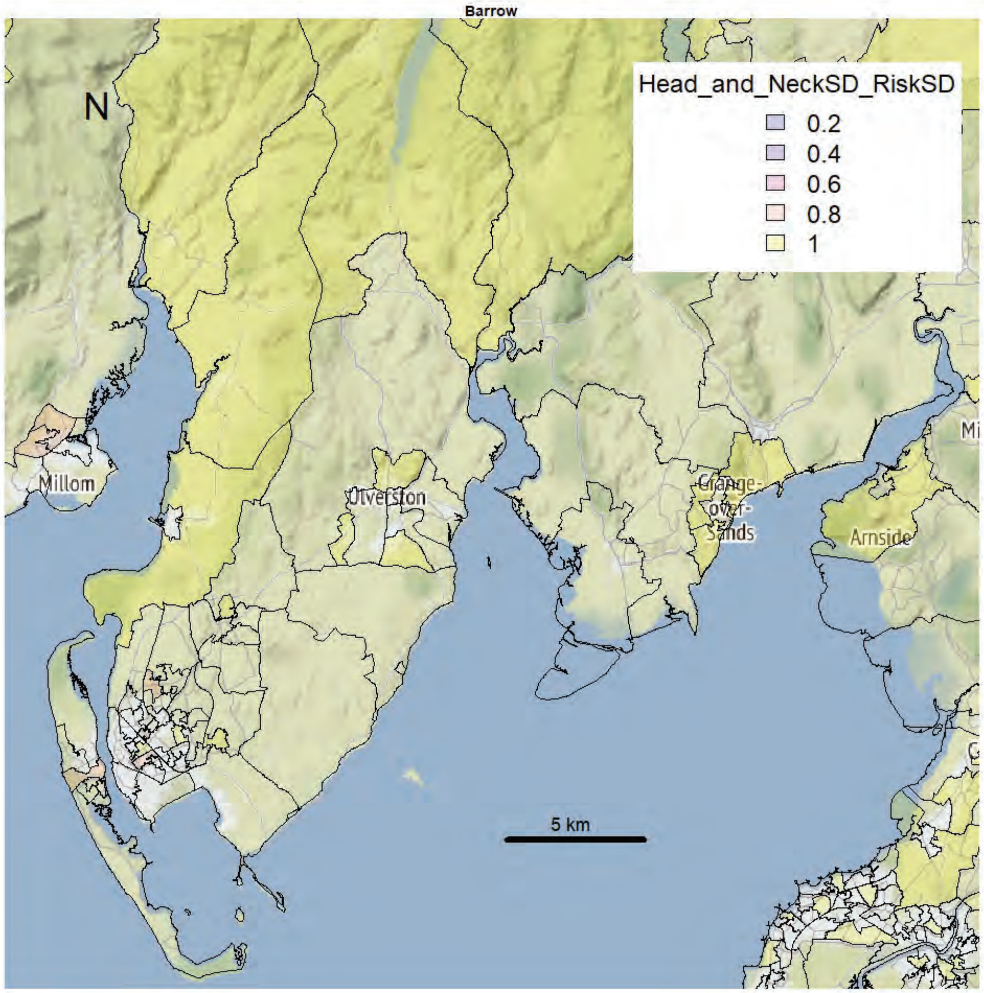
Barrow
N
Head_and_NeckSD_RiskSD
0.2
0.4
0.6
0.8
1
Millom
Ulverston
Grange-over-Sands
Arnside
5 km

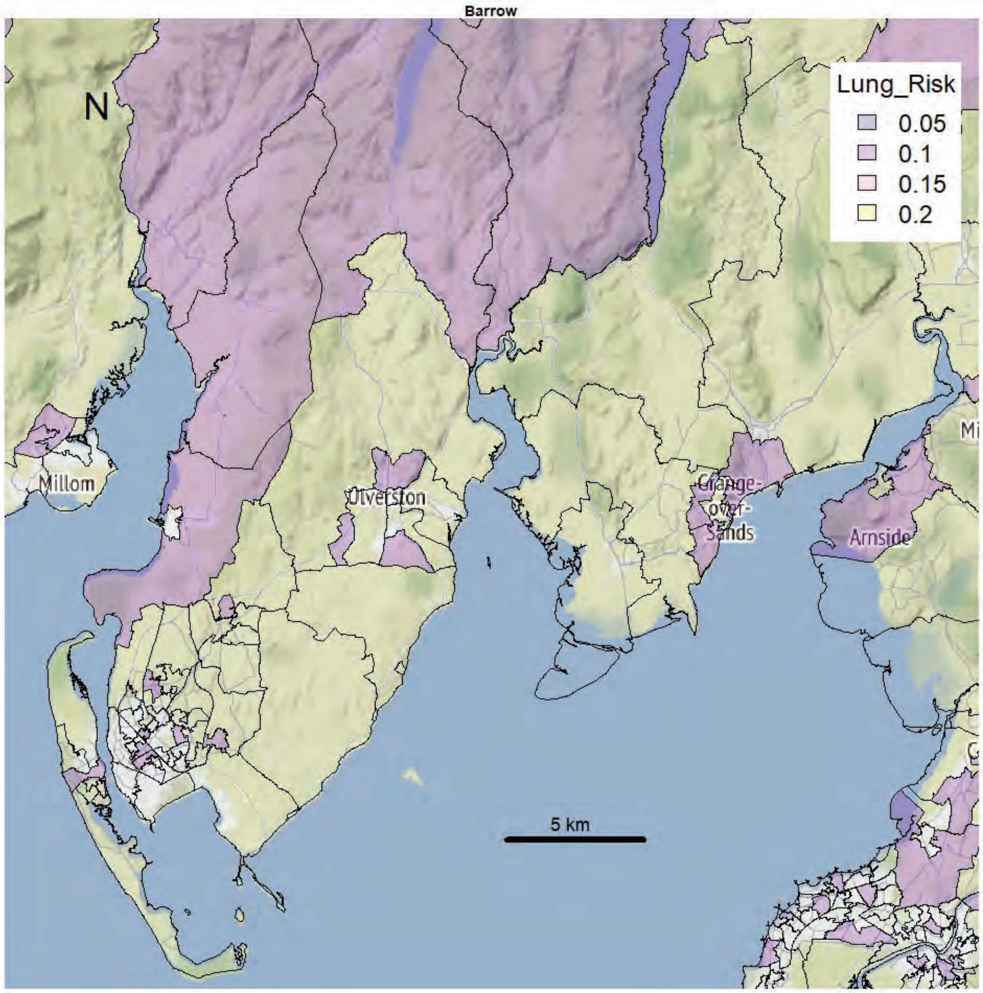
Barrow
N
Lung_Risk
0.05
0.1
0.15
0.2
Millom
Ulverston
Grange-over-Sands
Arnside
5 km

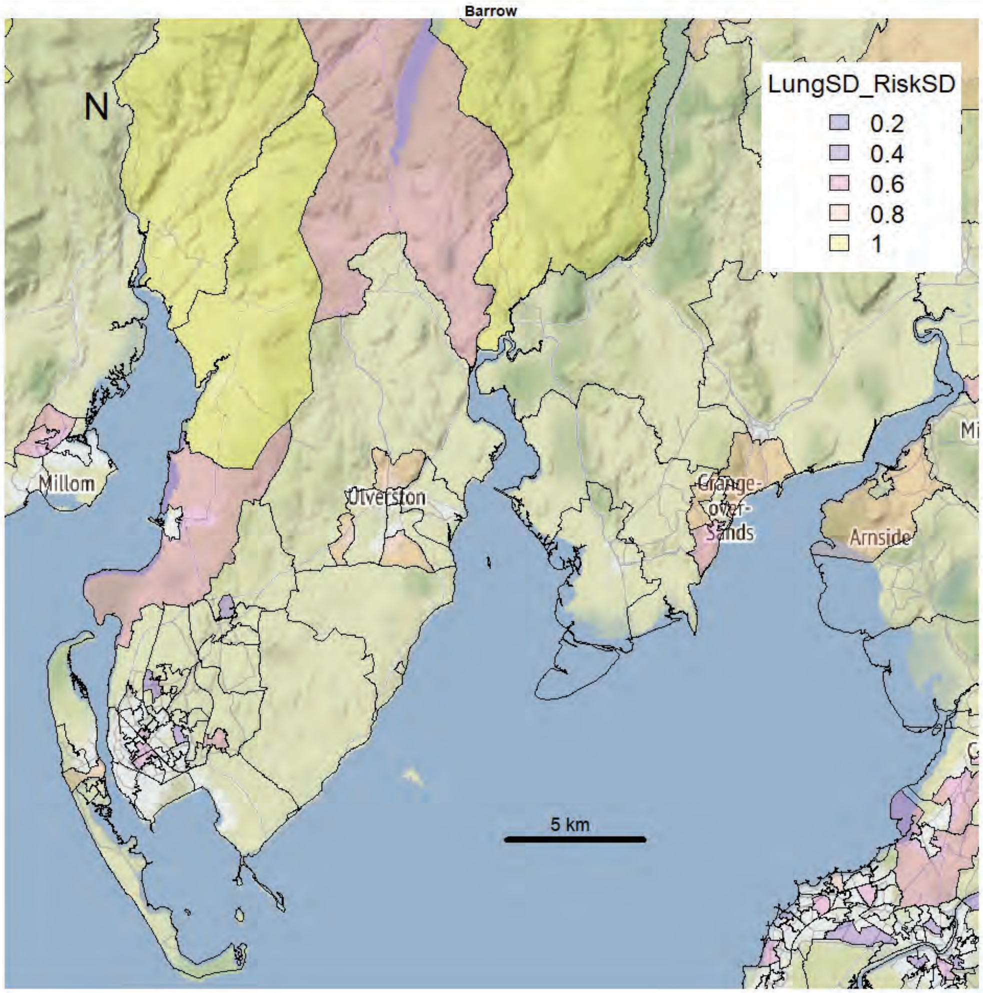
Barrow
N
LungSD_RiskSD
0.2
0.4
0.6
0.8
1
Millom
Ulverston
Grange-over-Sands
Arnside
5 km

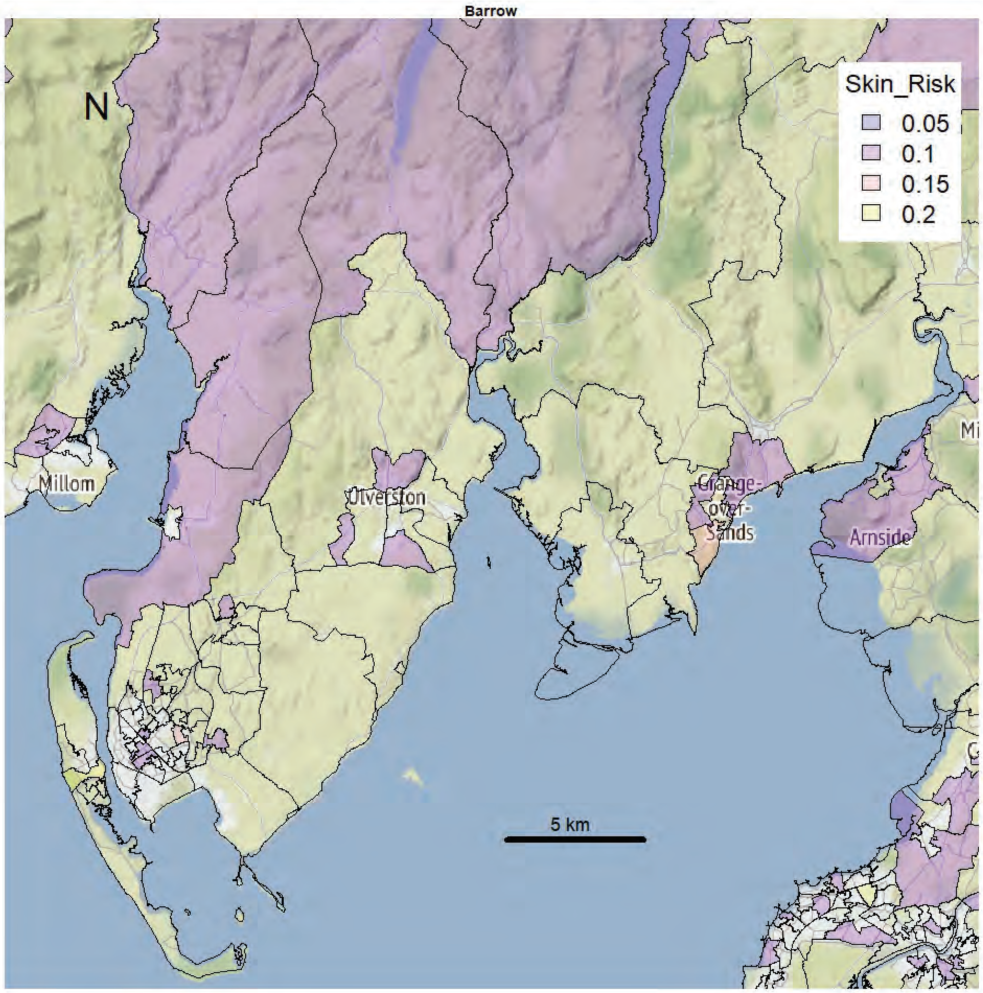
Barrow
N
Skin_Risk
0.05
0.1
0.15
0.2
Millom
Ulverston
Grange-over-Sands
Arnside
5 km

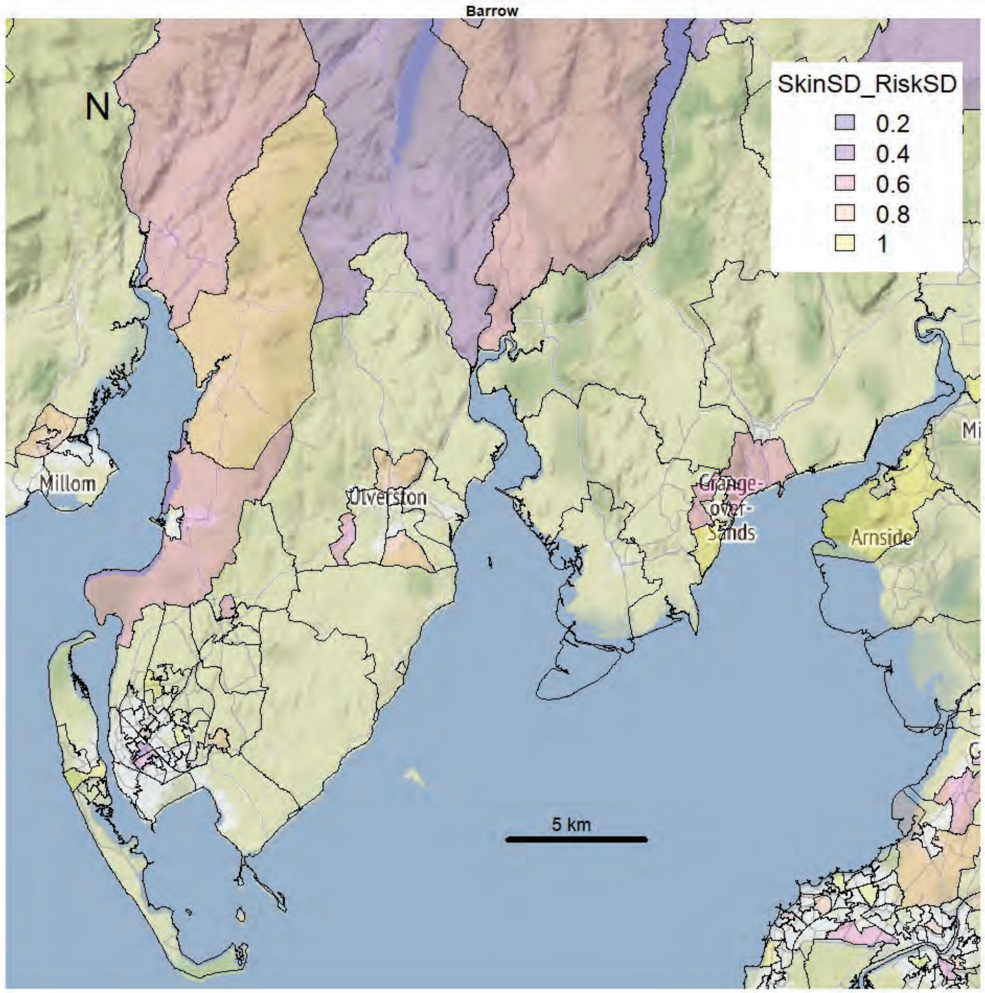
Barrow
N
SkinSD_RiskSD
0.2
0.4
0.6
0.8
1
Millom
Ulverston
Grange-over-Sands
Arnside
5 km

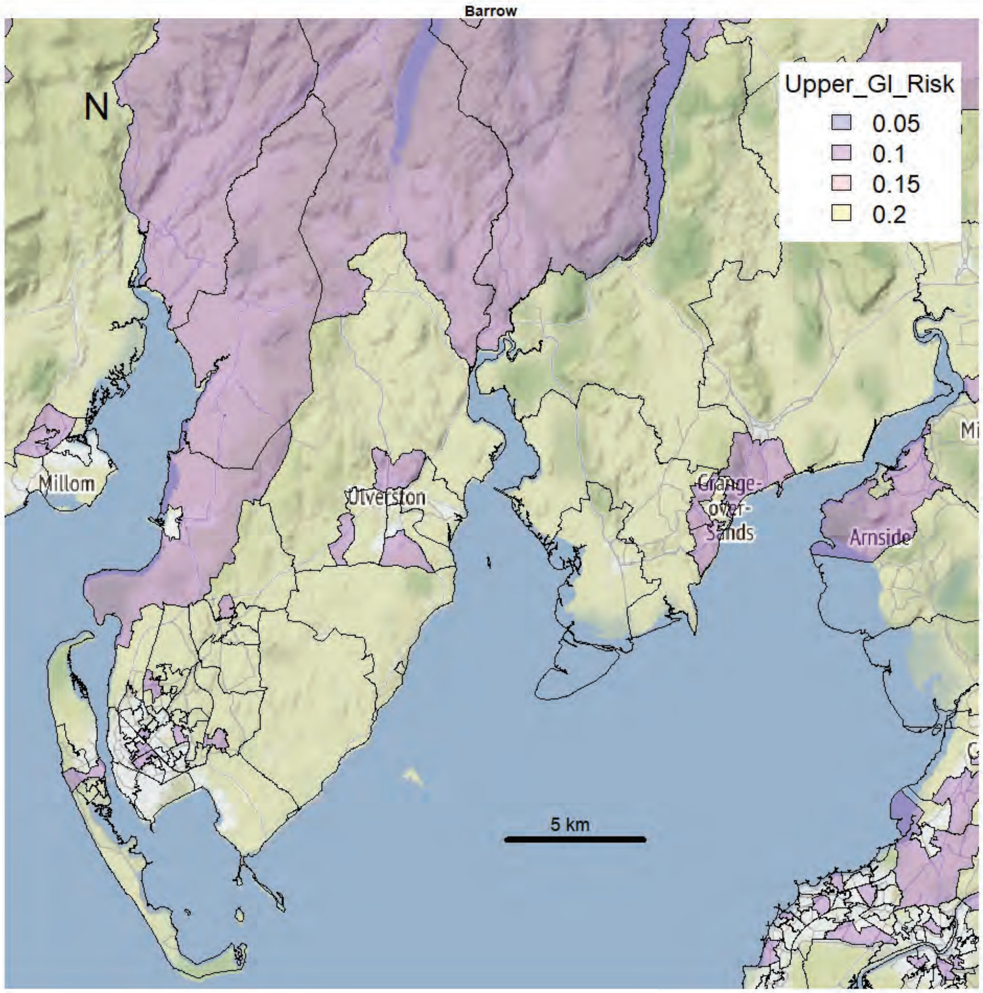
Barrow
N
Upper_GI_Risk
0.05
0.1
0.15
0.2
Millom
Ulverston
Grange-over-Sands
Arnside
5 km

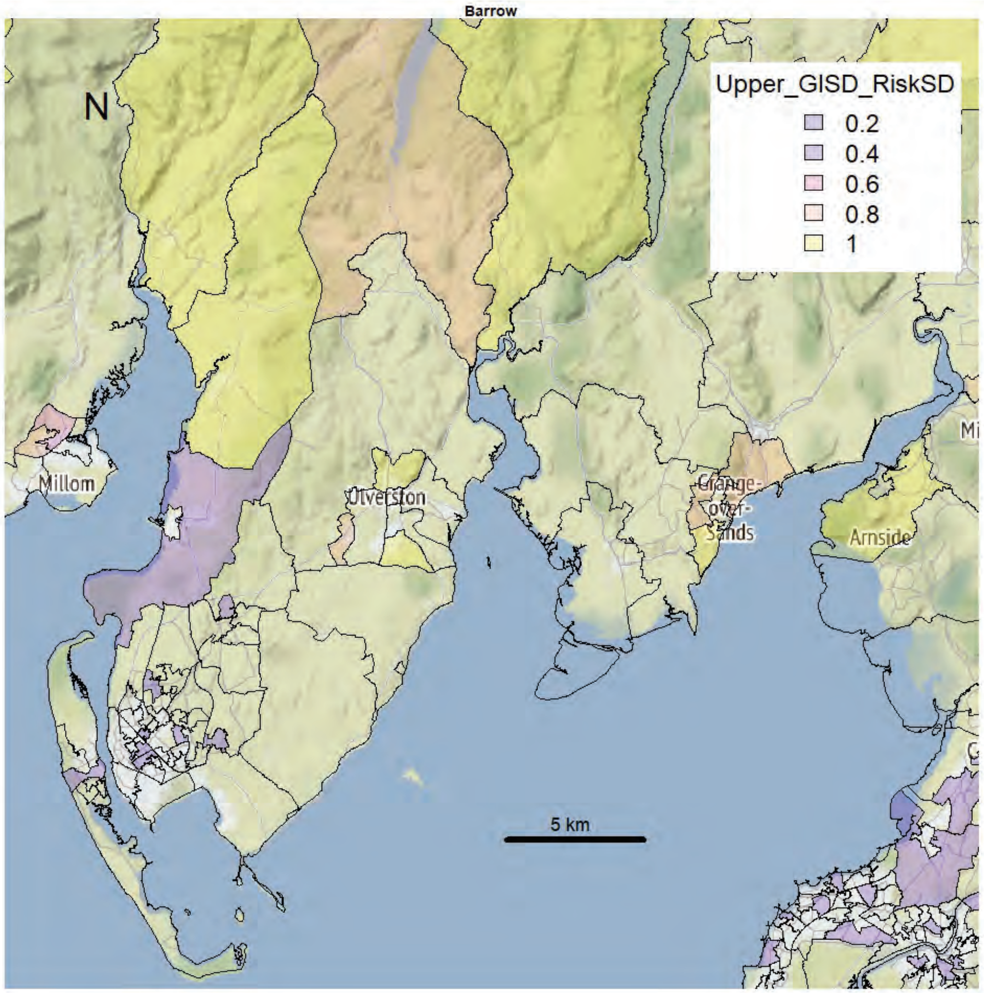
Barrow
N
Upper_GISD_RiskSD
0.2
0.4
0.6
0.8
1
Millom
Ulverston
Grange-over-Sands
Arnside
5 km

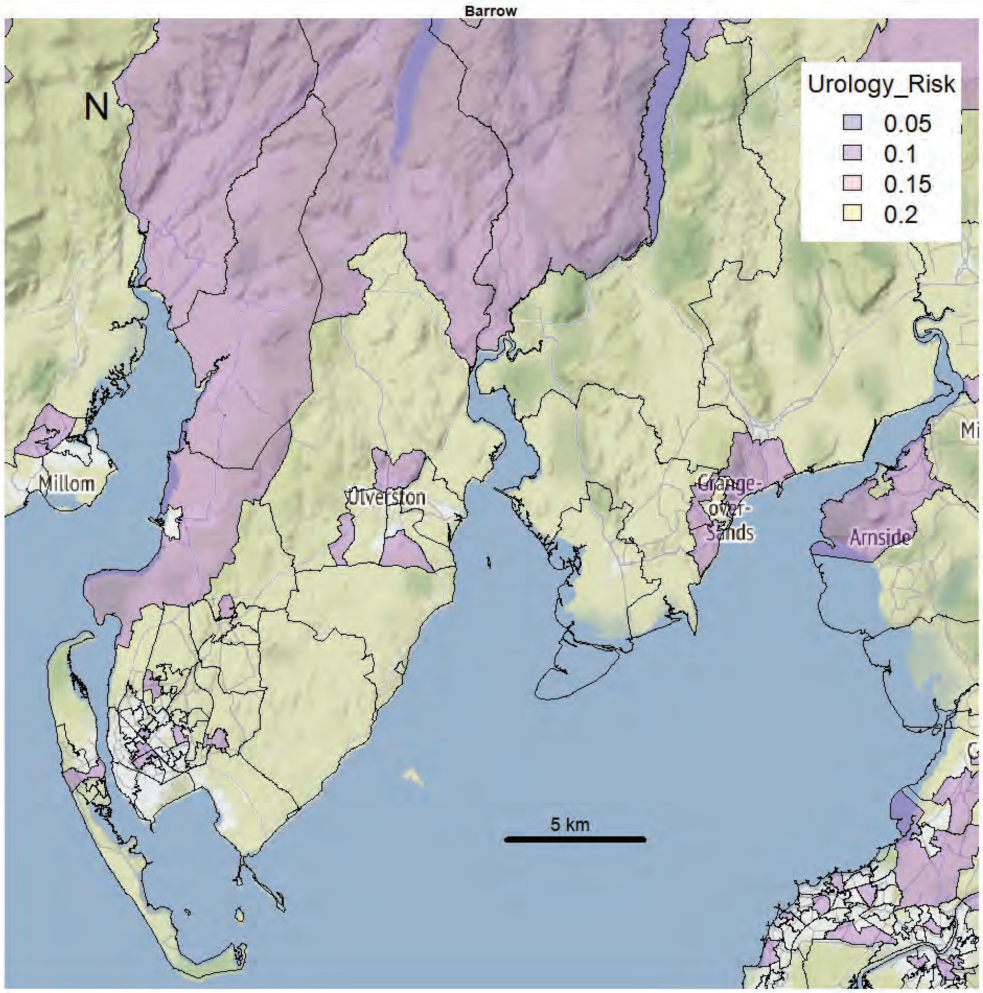
Barrow
N
Urology_Risk
0.05
0.1
0.15
0.2
Millom
Ulverston
Grange-over-Sands
Arnside
5 km

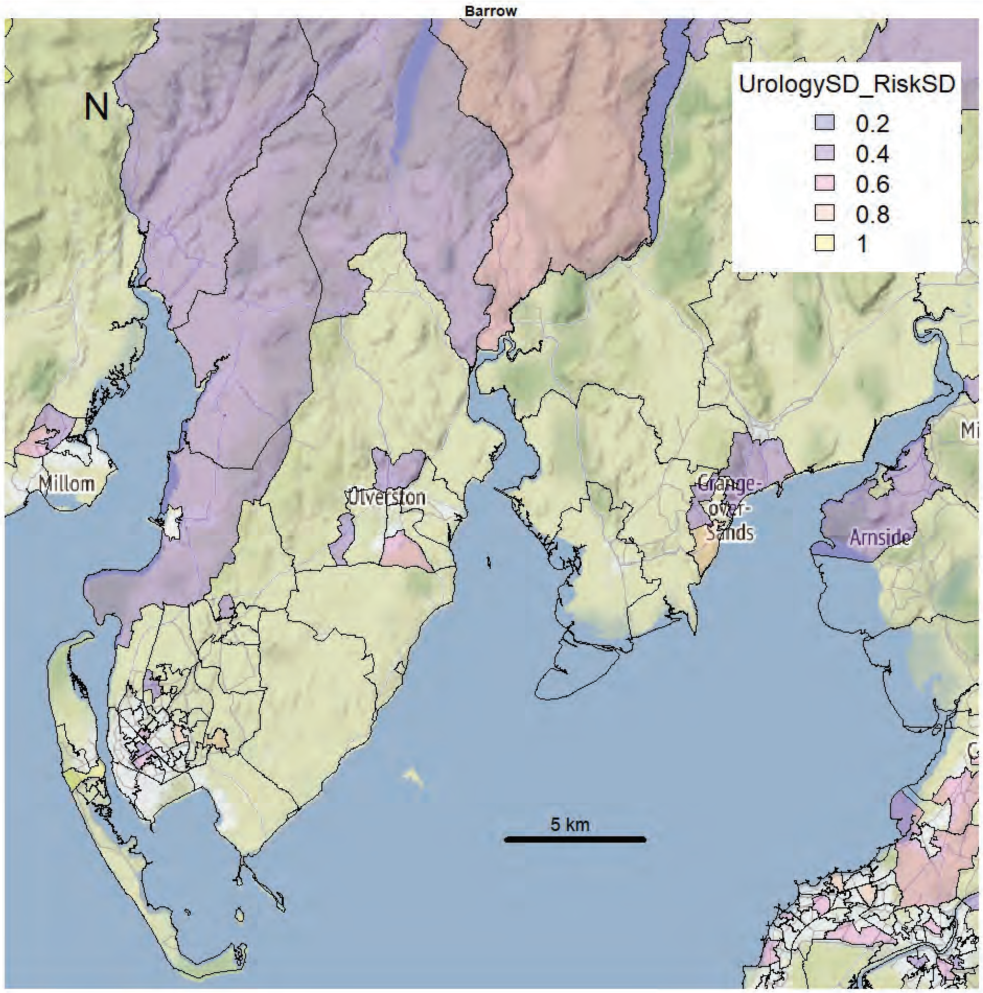
Barrow
N
UrologySD_RiskSD
0.2
0.4
0.6
0.8
1
Millom
Ulverston
Grange-over-Sands
Arnside
5 km

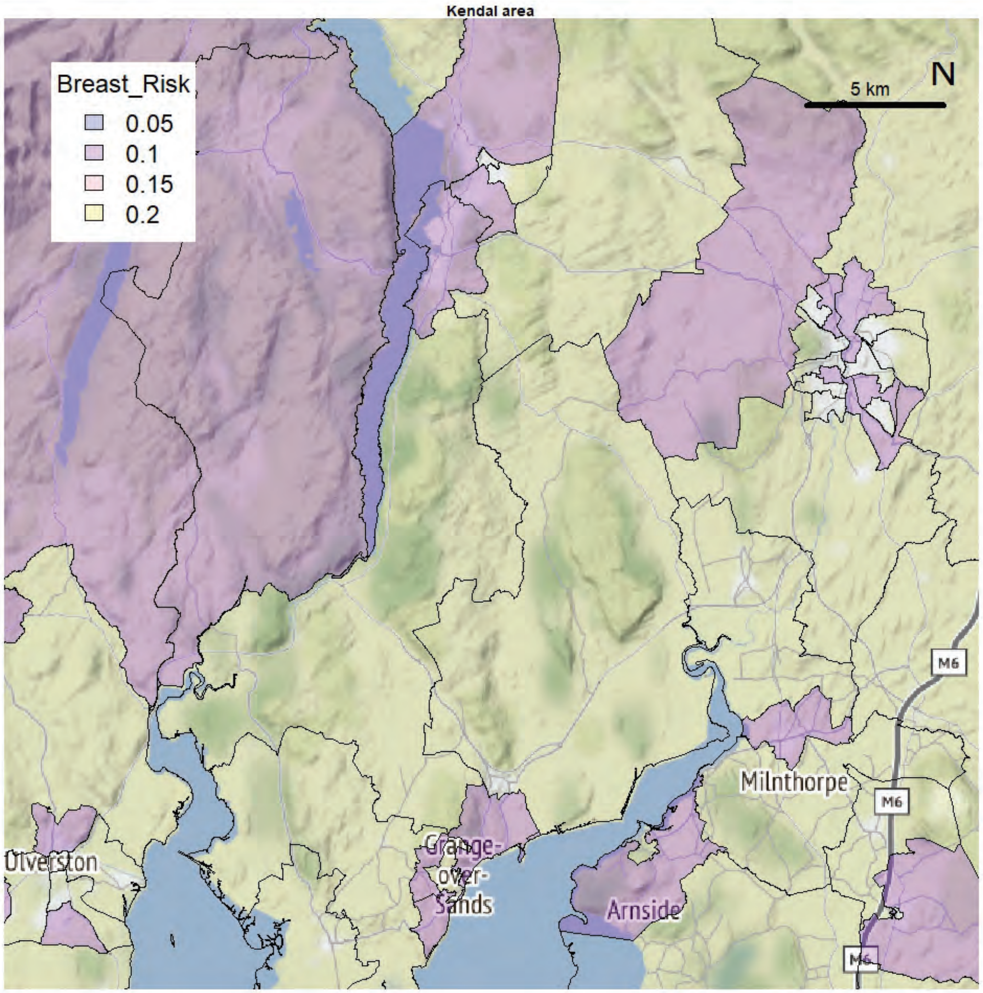
Kendal area
Breast_Risk
0.05
0.1
0.15
0.2
5 km
N
Milnthorpe
M6
Ulverston
Grange-over-Sands
Arnside

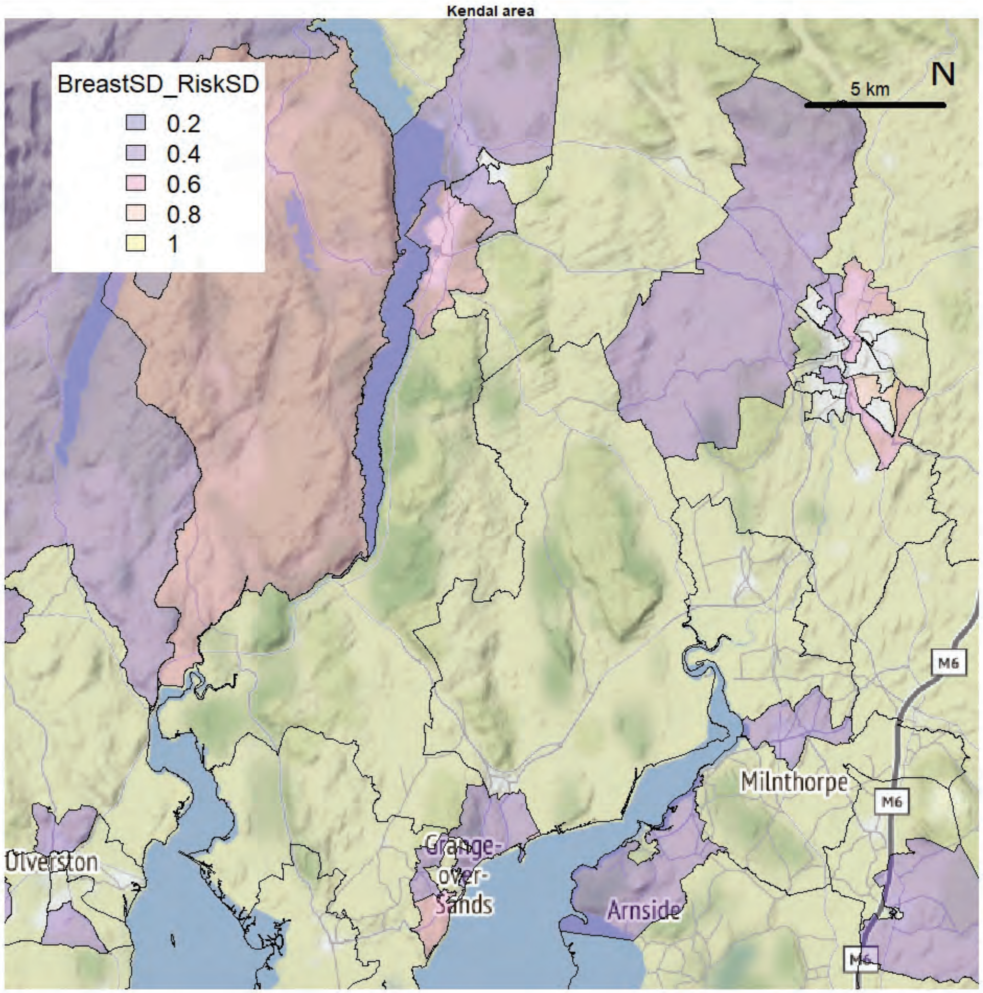
Kendal area
BreastSD_RiskSD
0.2
0.4
0.6
0.8
1
5 km
N
Milnthorpe
M6
Ulverston
Grange-over-Sands
Arnside

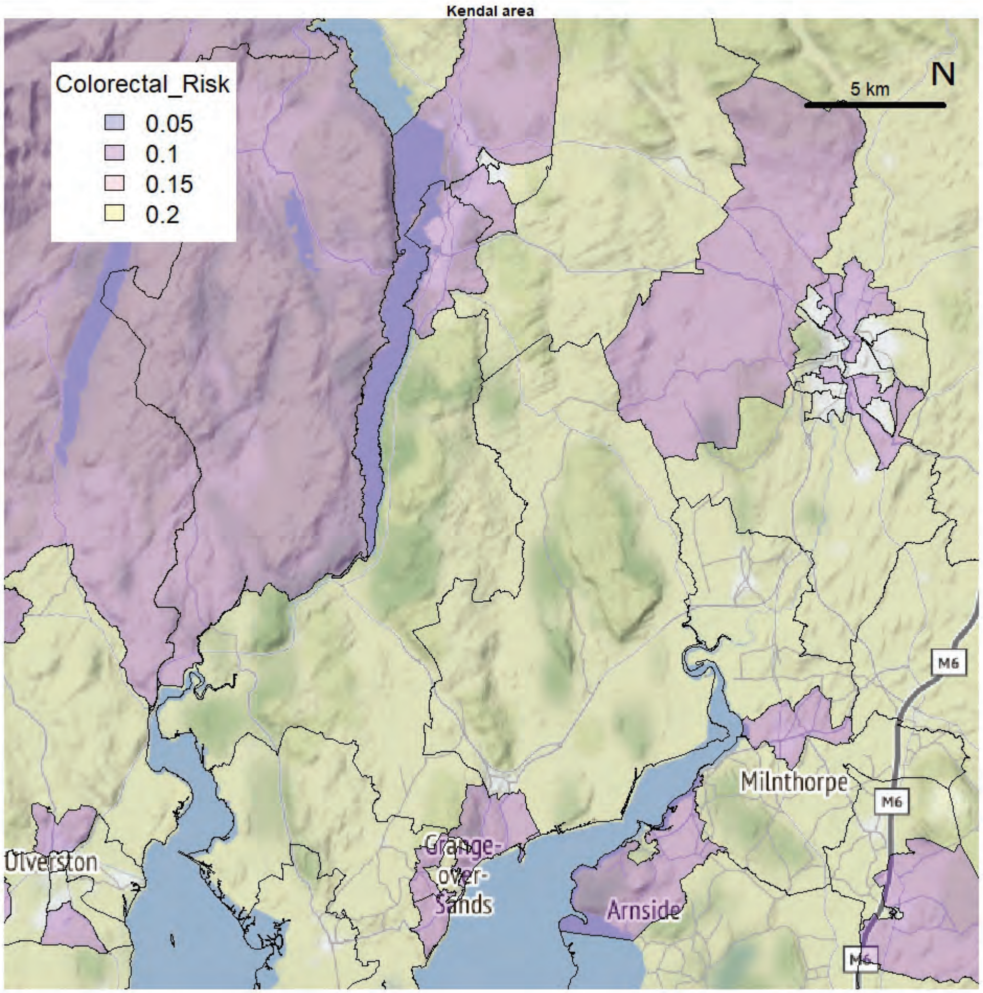
Kendal area
Colorectal_Risk
0.05
0.1
0.15
0.2
5 km
N
M6
Milnthorpe
Ulverston
Grange-over-Sands
Arnside

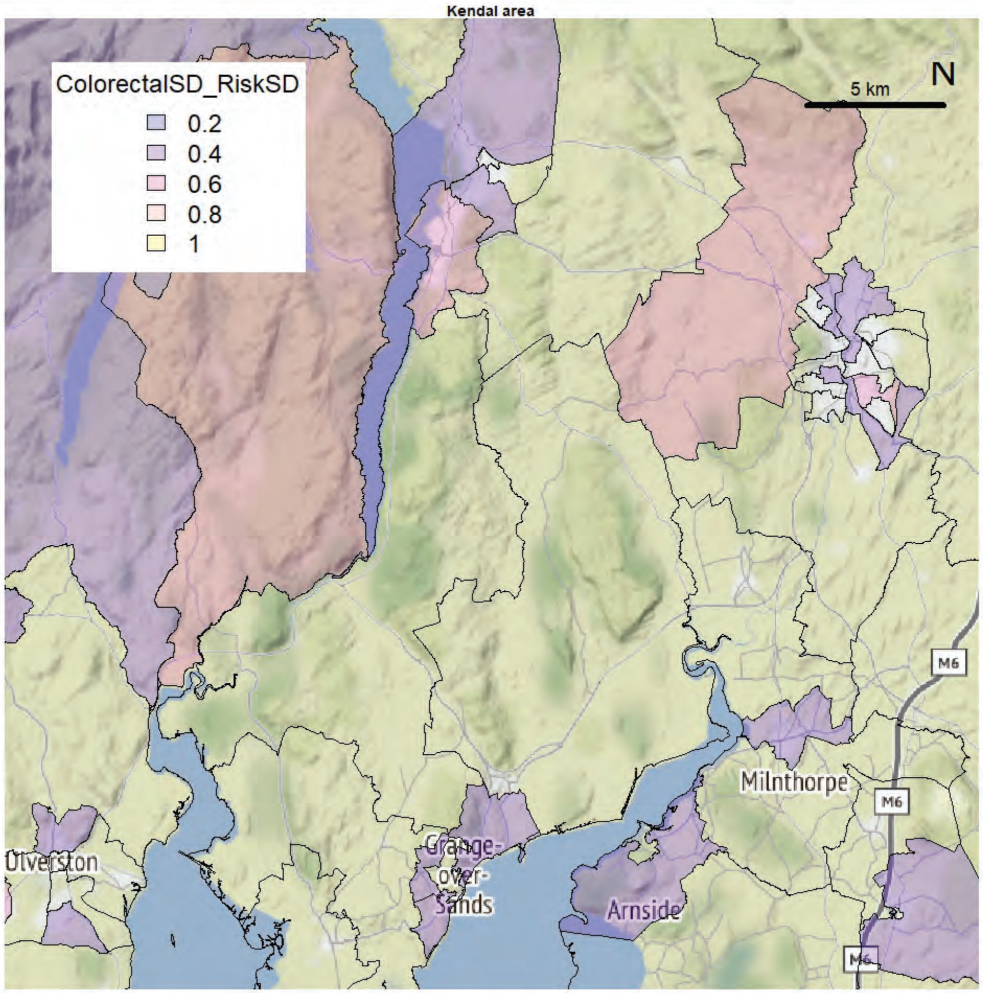
Kendal area
ColorectalSD_RiskSD
0.2
0.4
0.6
0.8
1
5 km
N
M6
Milnthorpe
Ulverston
Grange-over-Sands
Arnside

Kendal area

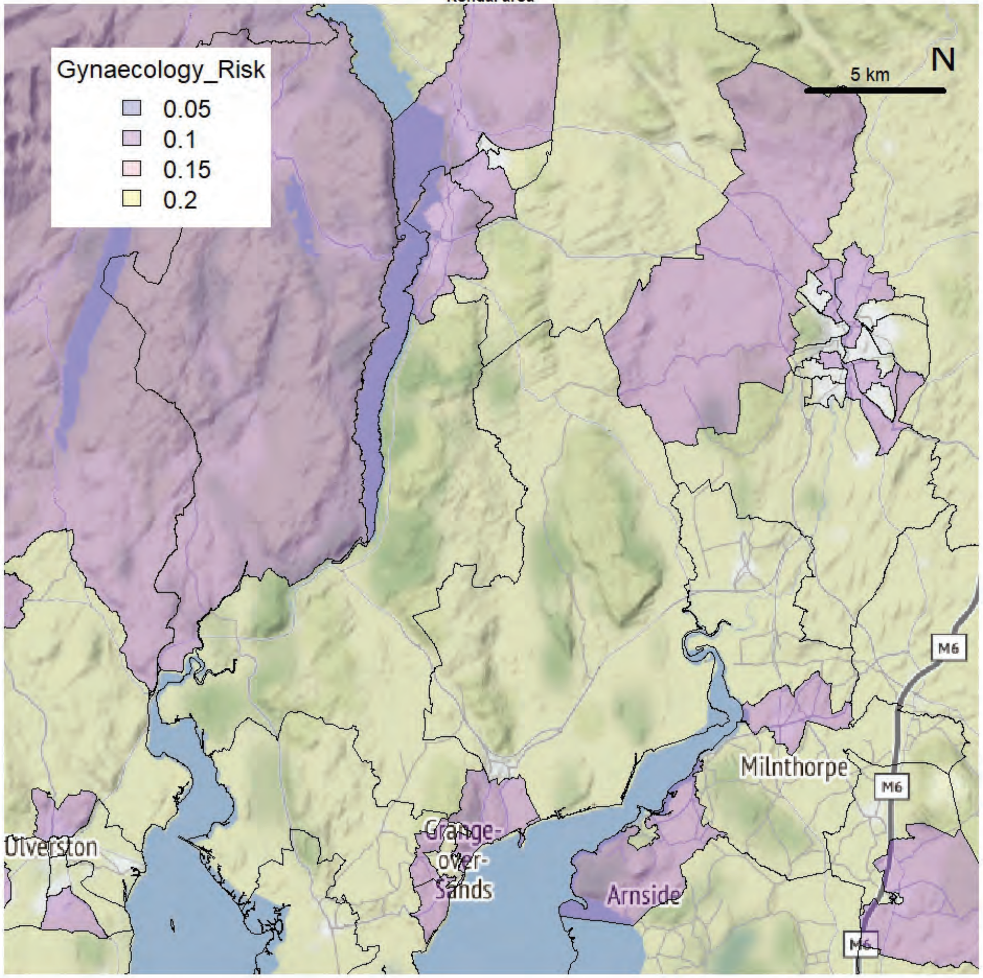


Kendal area

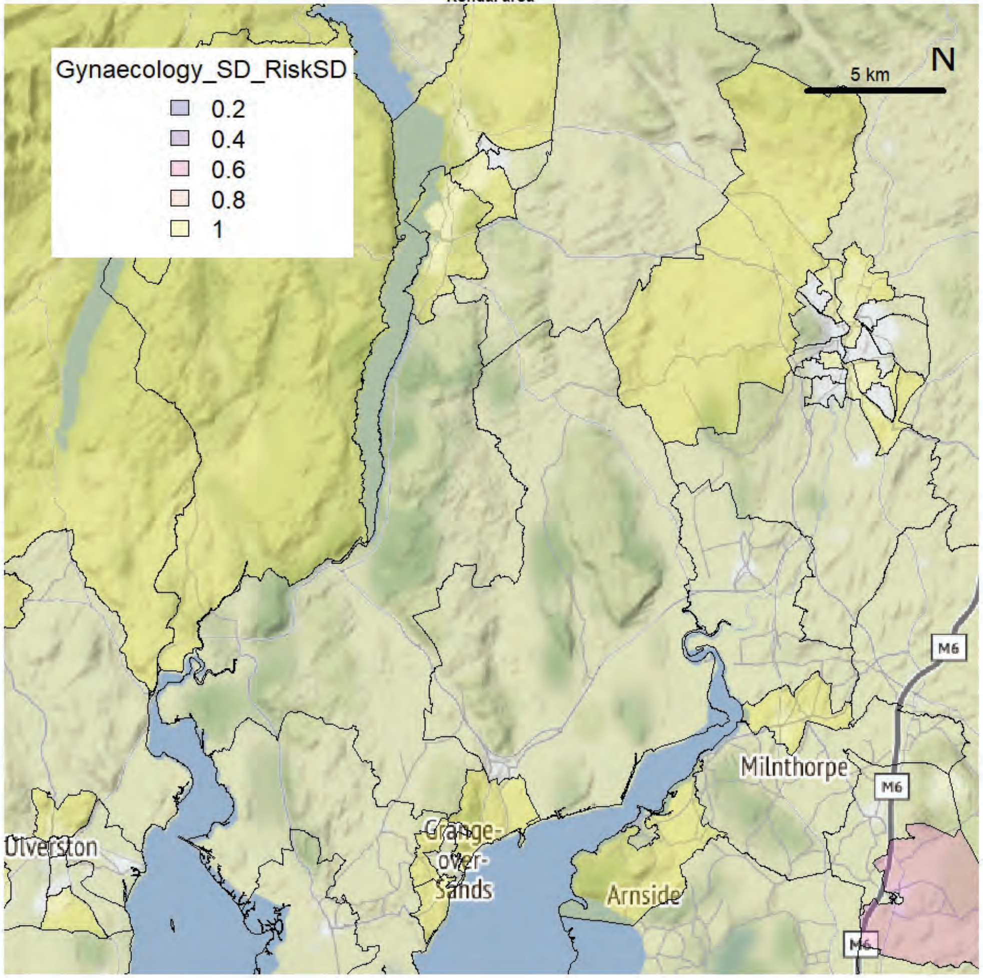

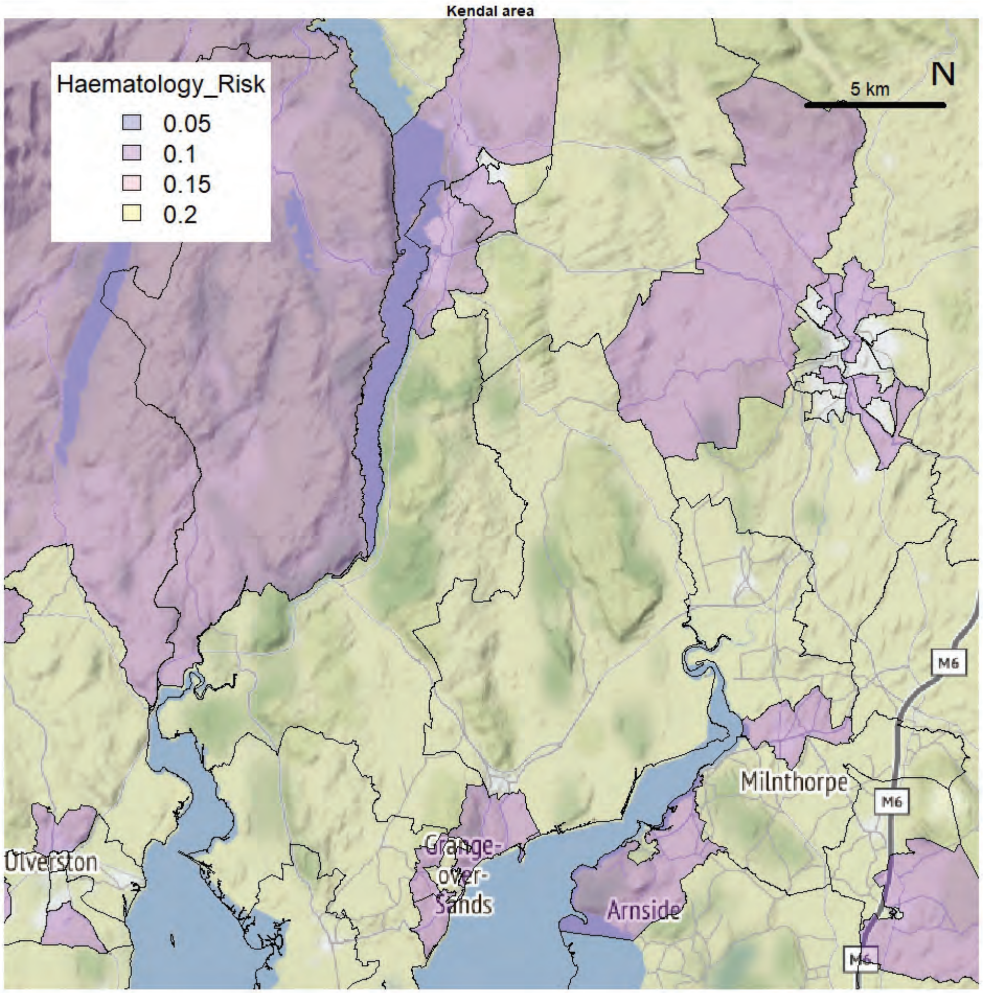
Kendal area
Haematology_Risk
0.05
0.1
0.15
0.2
5 km
N
M6
Milnthorpe
Ulverston
Grange-over-Sands
Arnside

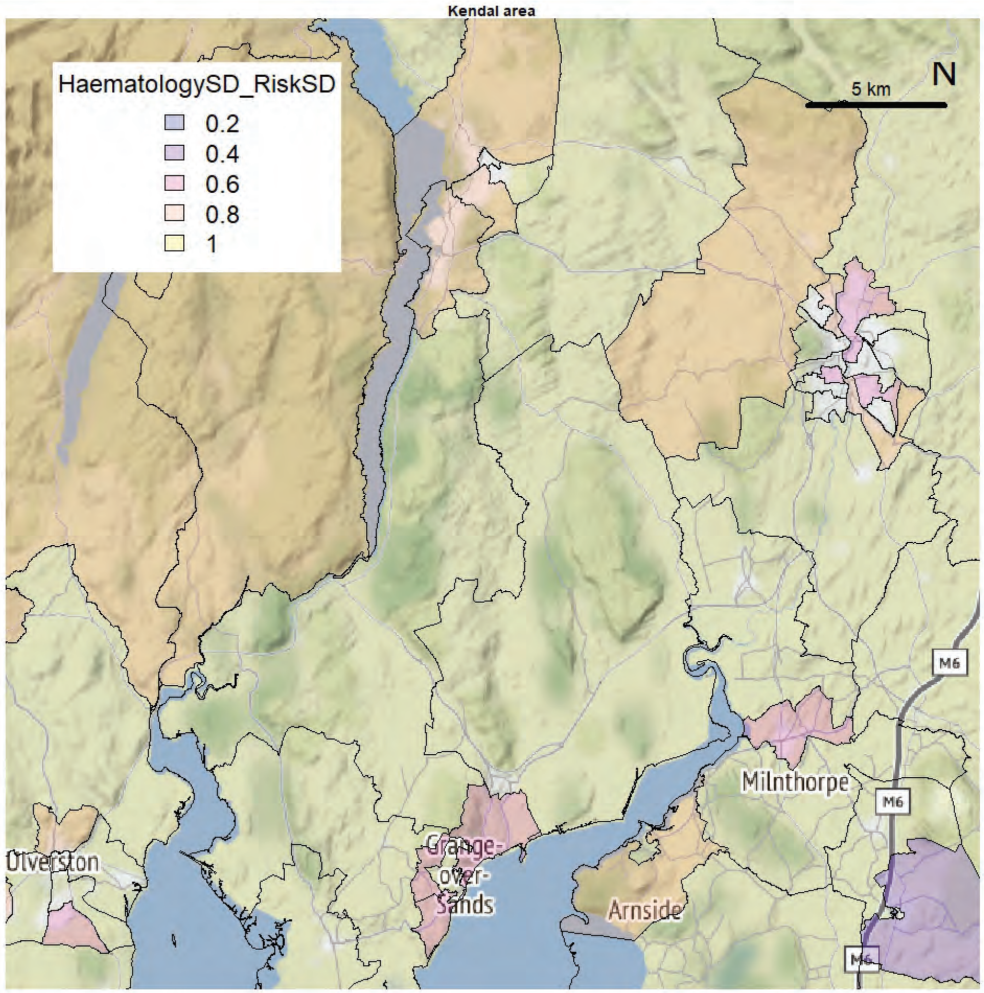
Kendal area
HaematologySD_RiskSD
0.2
0.4
0.6
0.8
1
5 km
N
M6
Milnthorpe
Ulverston
Grange-over-Sands
Arnside

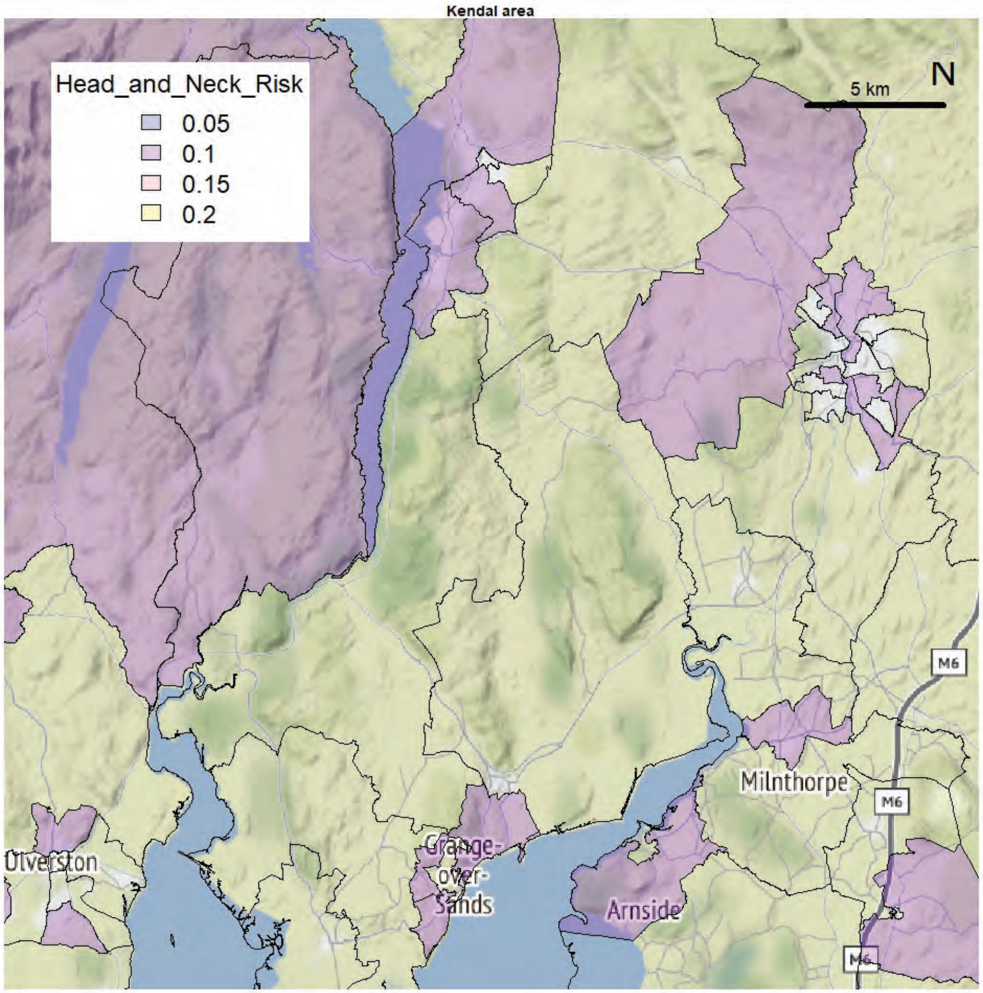
Kendal area
Head_and_Neck_Risk
0.05
0.1
0.15
0.2
5 km
N
Ulverston
Grange-over-Sands
Arnside
Milnthorpe
M6

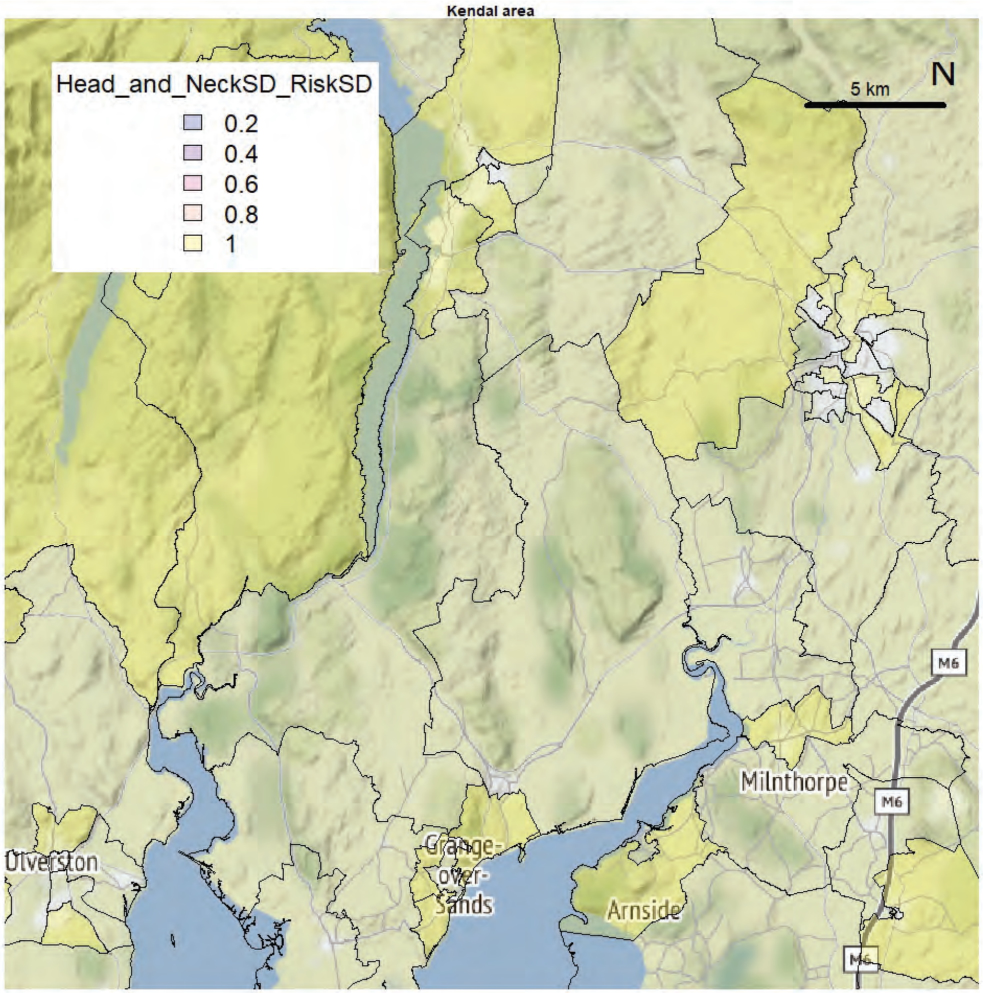
Kendal area
Head_and_NeckSD_RiskSD
0.2
0.4
0.6
0.8
1
5 km
N
Ulverston
Grange-over-Sands
Arnside
Milnthorpe
M6

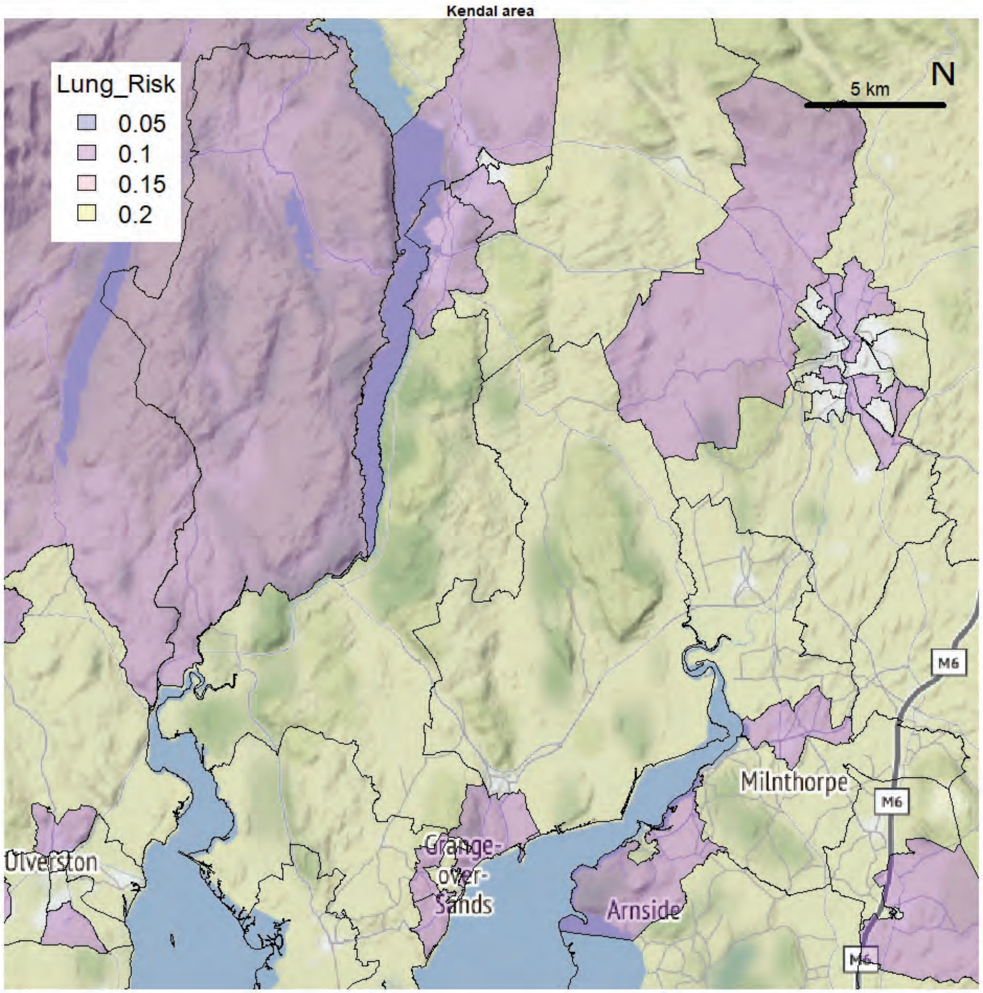
Kendal area
Lung_Risk
0.05
0.1
0.15
0.2
5 km
N
Milnthorpe
M6
Grange-over-Sands
Arnside
Ulverston

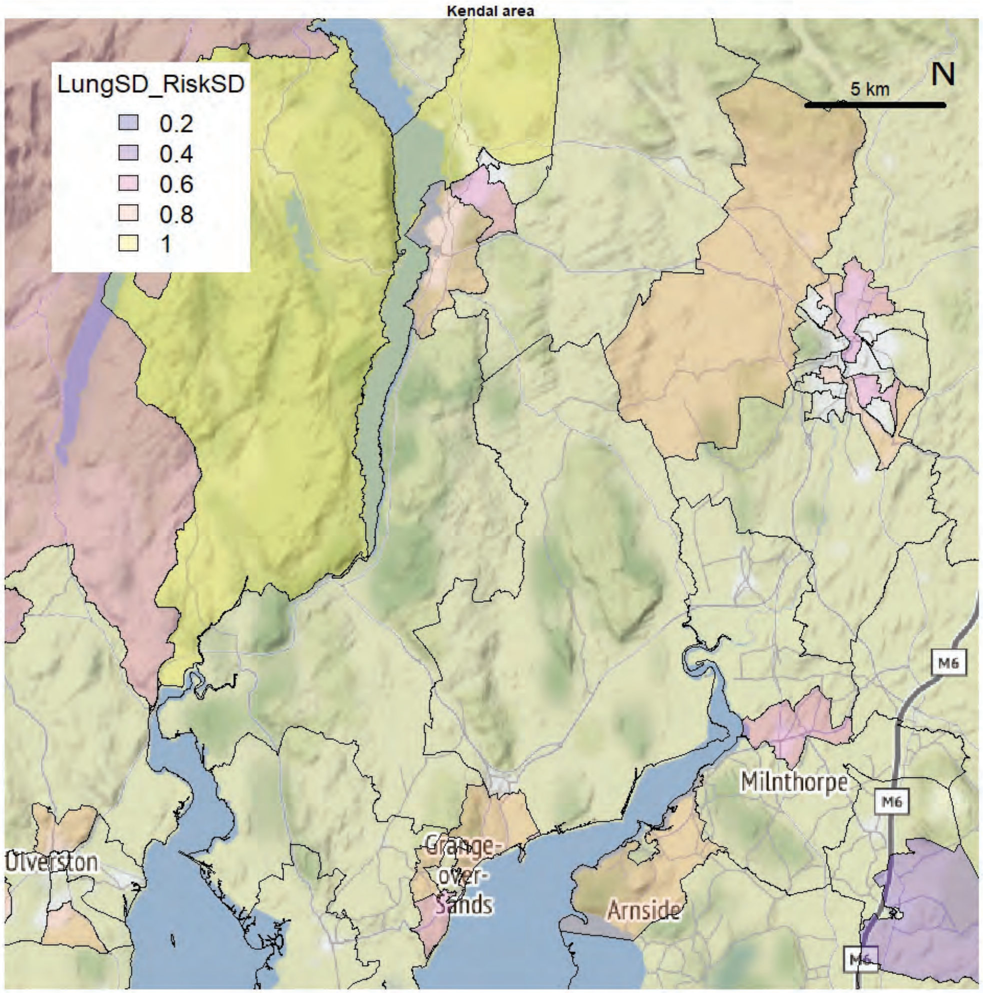
Kendal area
LungSD_RiskSD
0.2
0.4
0.6
0.8
1
5 km
N
Milnthorpe
M6
Grange-over-Sands
Arnside
Ulverston

Kendal area

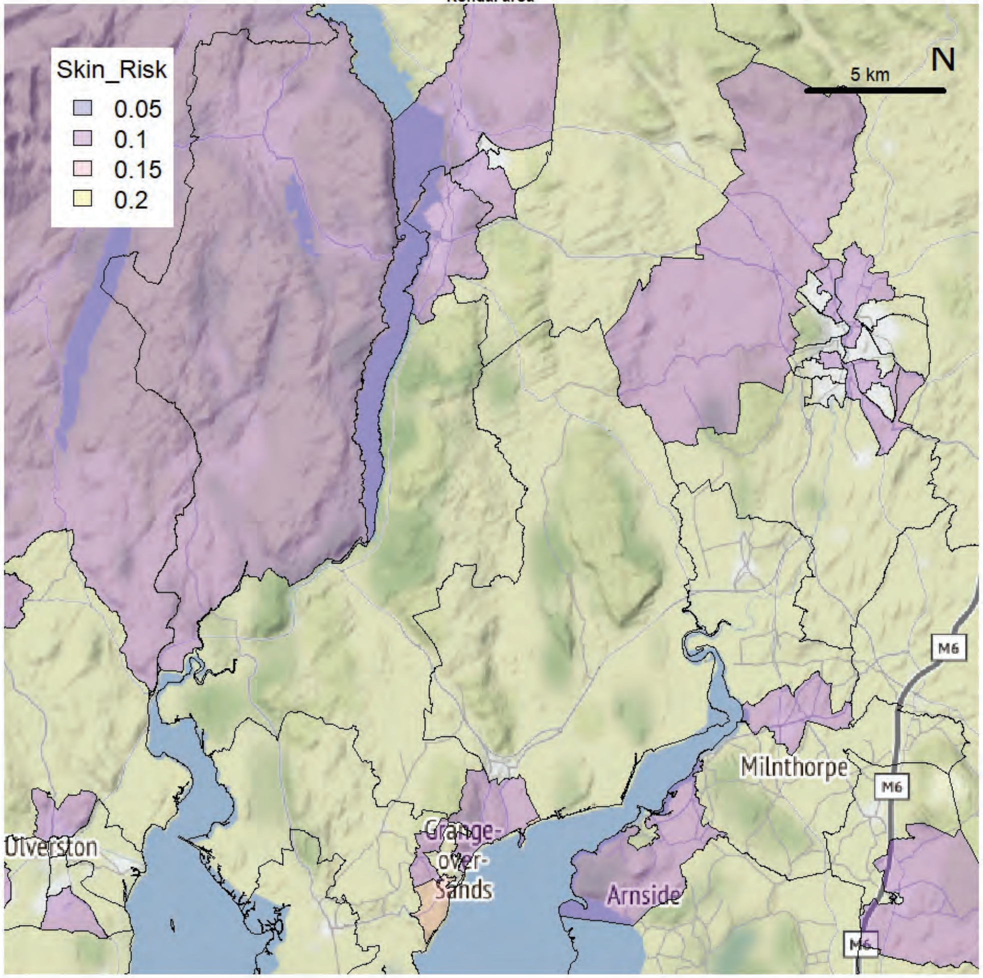


Kendal area

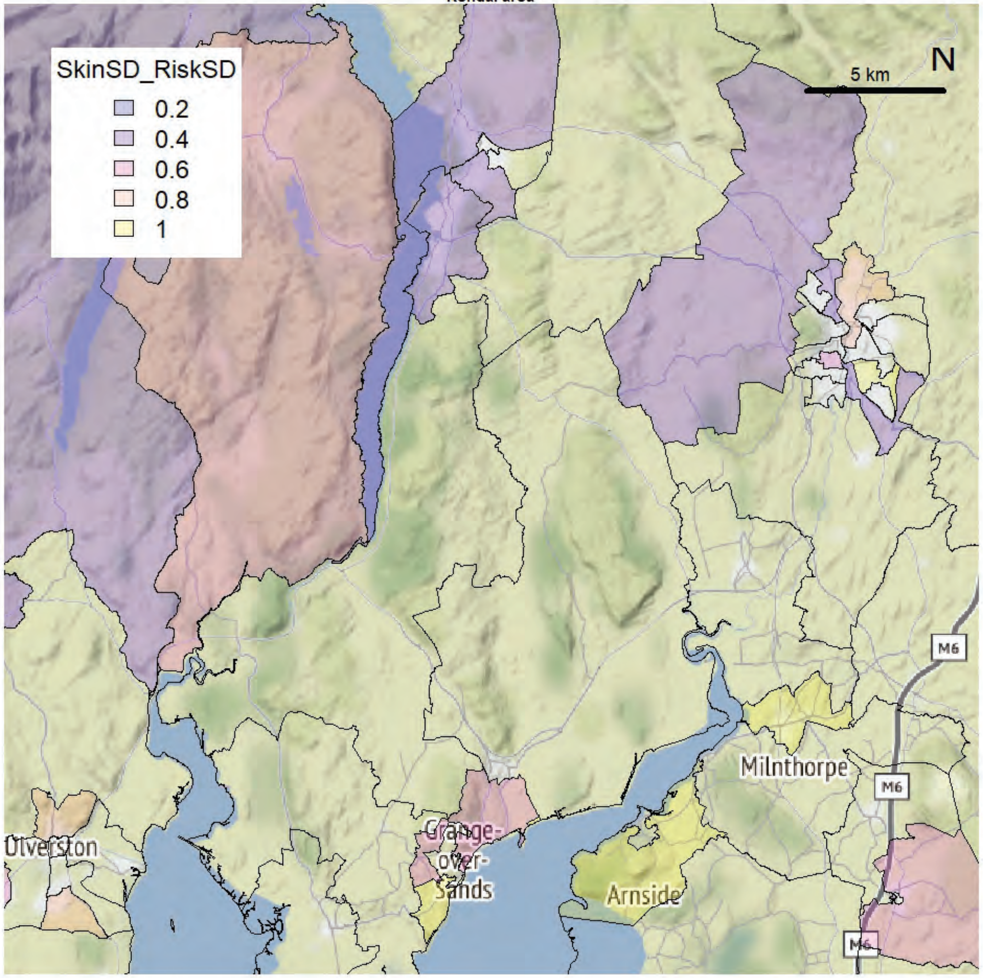

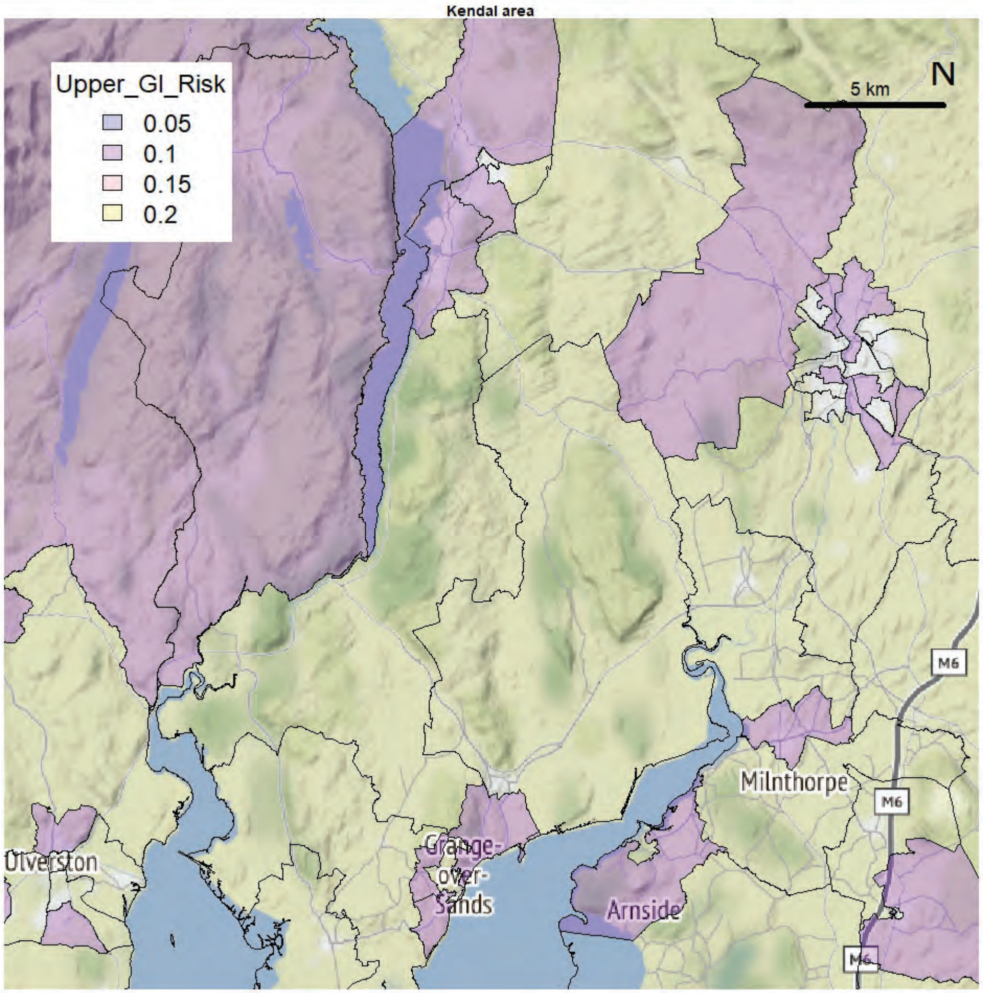
Kendal area
Upper_GI_Risk
0.05
0.1
0.15
0.2
5 km
N
Milnthorpe
Grange-over-Sands
Arnside
Ulverston
M6

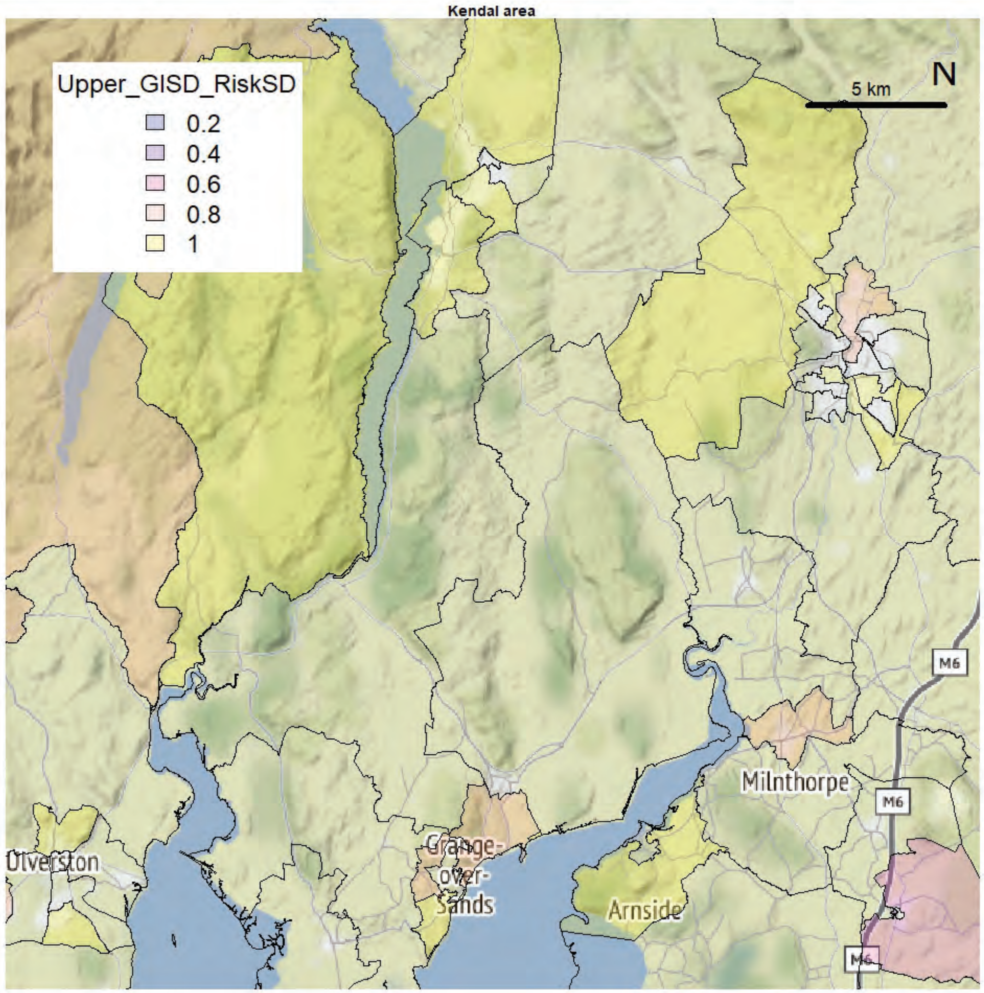
Kendal area
Upper_GISD_RiskSD
0.2
0.4
0.6
0.8
1
5 km
N
Milnthorpe
Grange-over-Sands
Arnside
Ulverston
M6

Kendal area

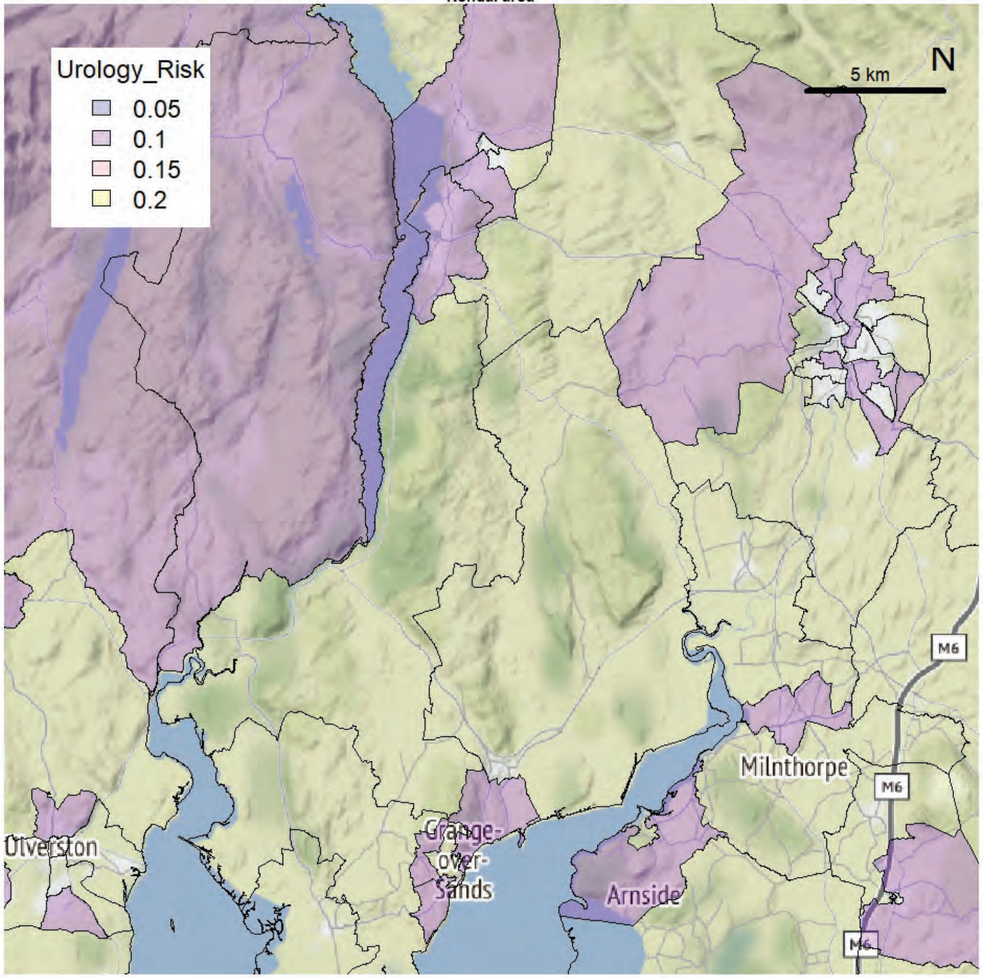


Kendal area

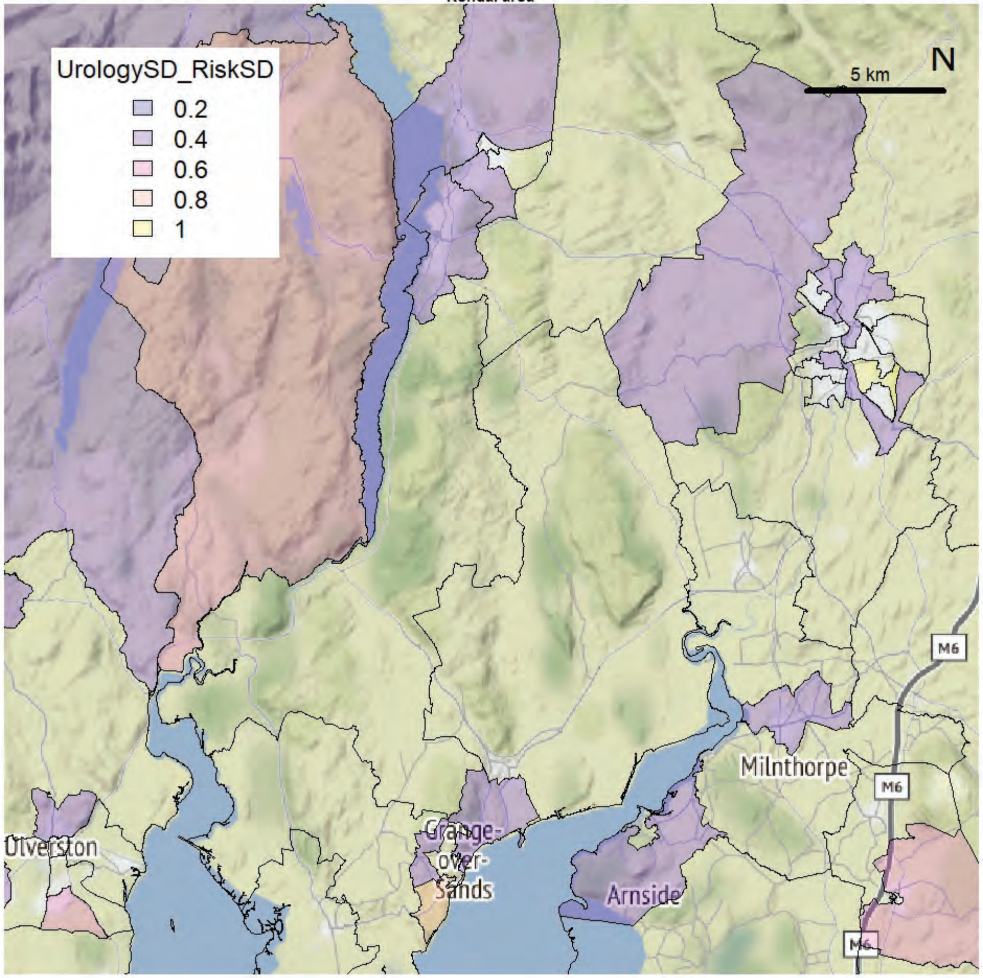

Morecambe Bay exCCG extended

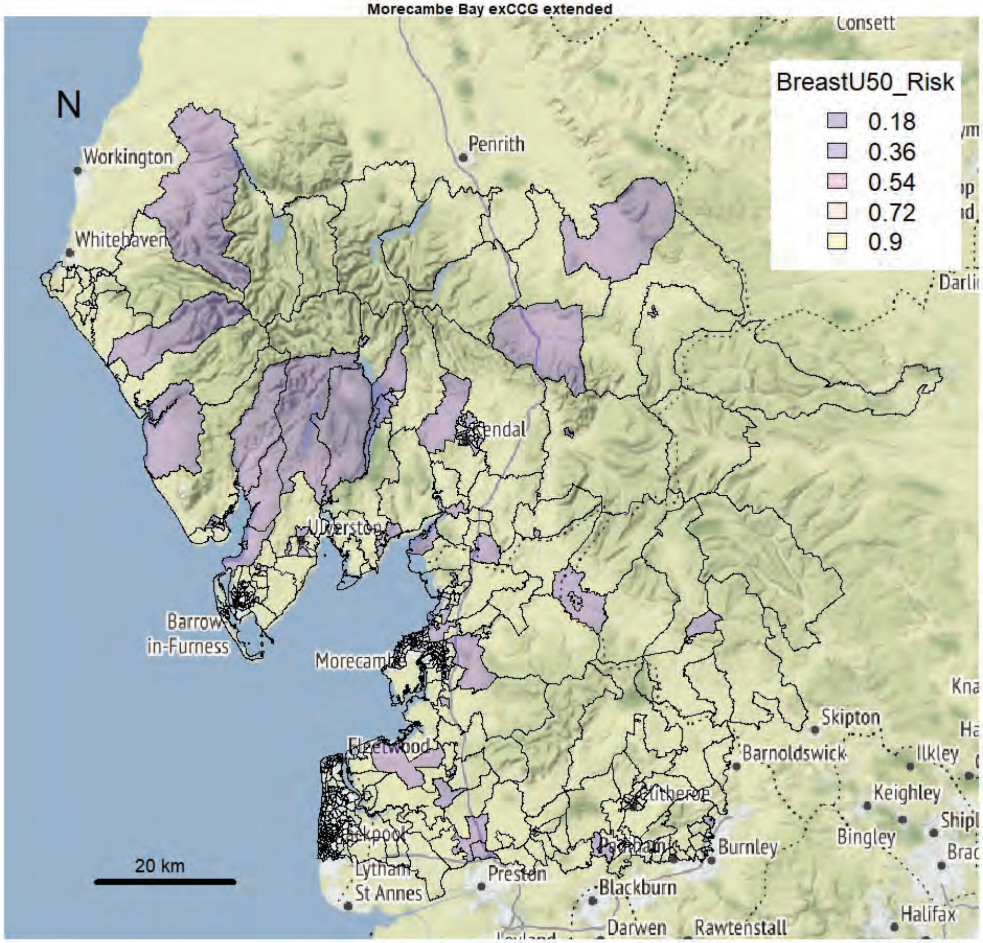


Morecambe Bay exCCG extended

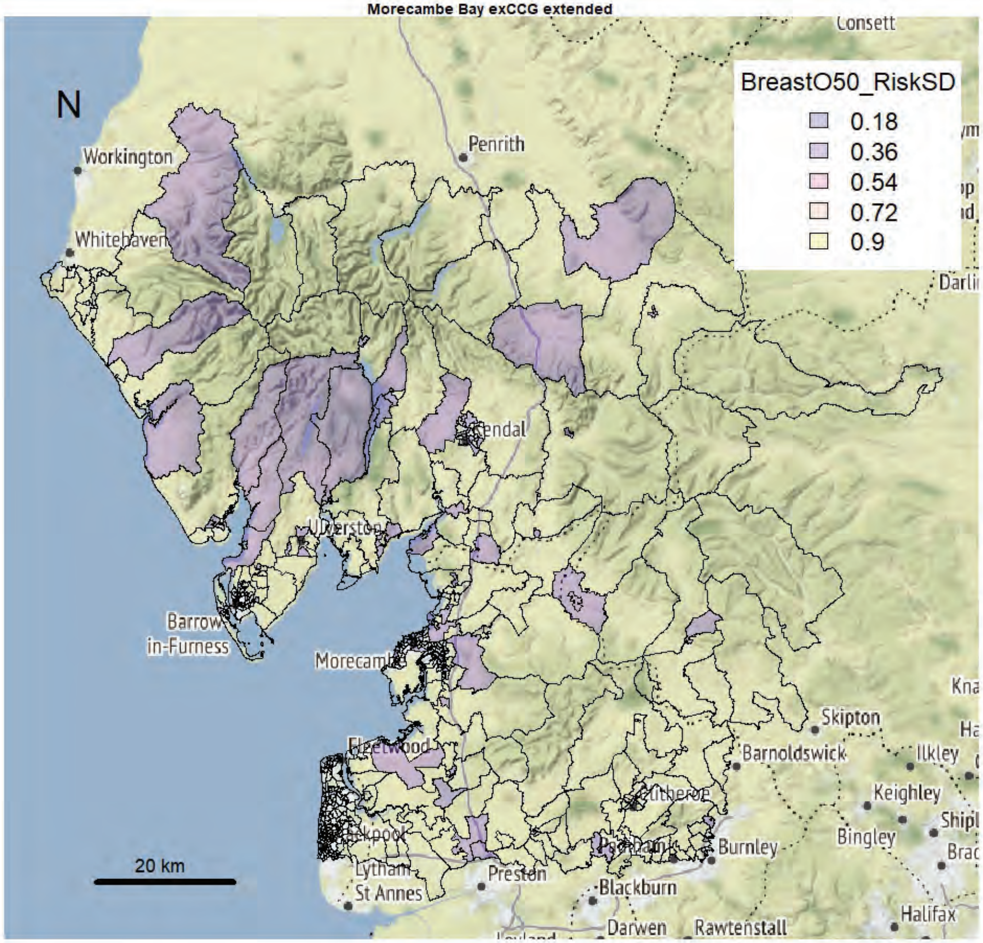

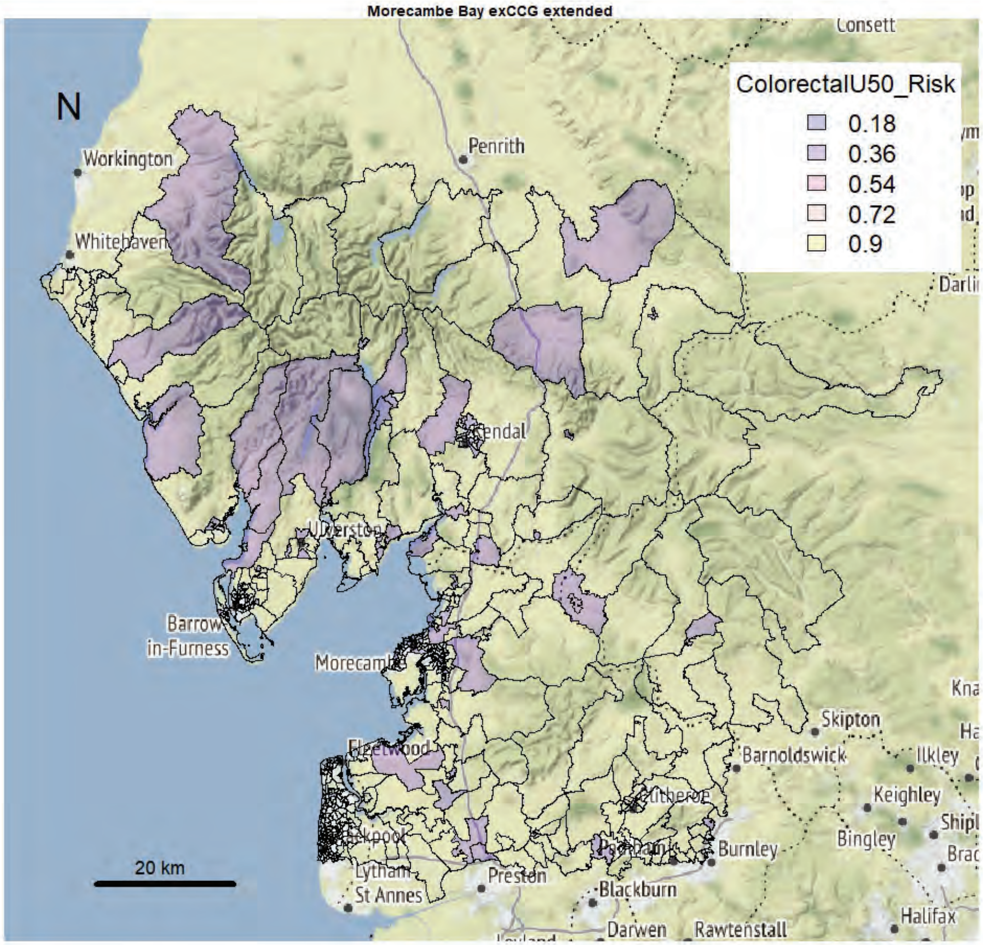
Morecambe Bay exCCG extended
N
ColorectalU50_Risk
0.18
0.36
0.54
0.72
0.9
Workington
Whitehaven
Penrith
Consett
Kendal
Ulverston
Barrow in-Furness
Morecambe
Fleetwood
Blackpool
Lytham St Annes
Preston
Blackburn
Darwen
Rawtenstall
Burnley
Clitheroe
Barnoldswick
Skipton
Ilkley
Keighley
Bingley
Halifax
20 km

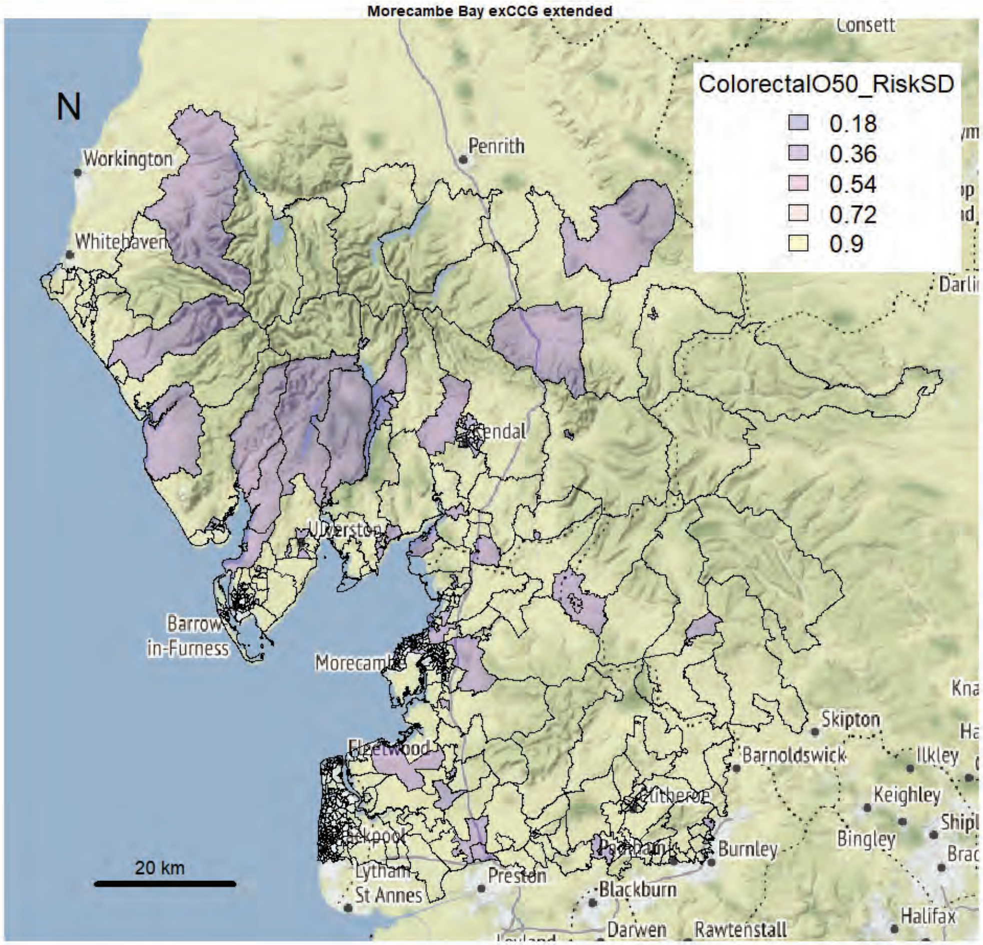
Morecambe Bay exCCG extended
N
ColorectalO50_RiskSD
0.18
0.36
0.54
0.72
0.9
Workington
Whitehaven
Penrith
Consett
Kendal
Ulverston
Barrow in-Furness
Morecambe
Fleetwood
Blackpool
Lytham St Annes
Preston
Blackburn
Darwen
Rawtenstall
Burnley
Clitheroe
Barnoldswick
Skipton
Ilkley
Keighley
Bingley
Halifax
20 km

Morecambe Bay exCCG extended

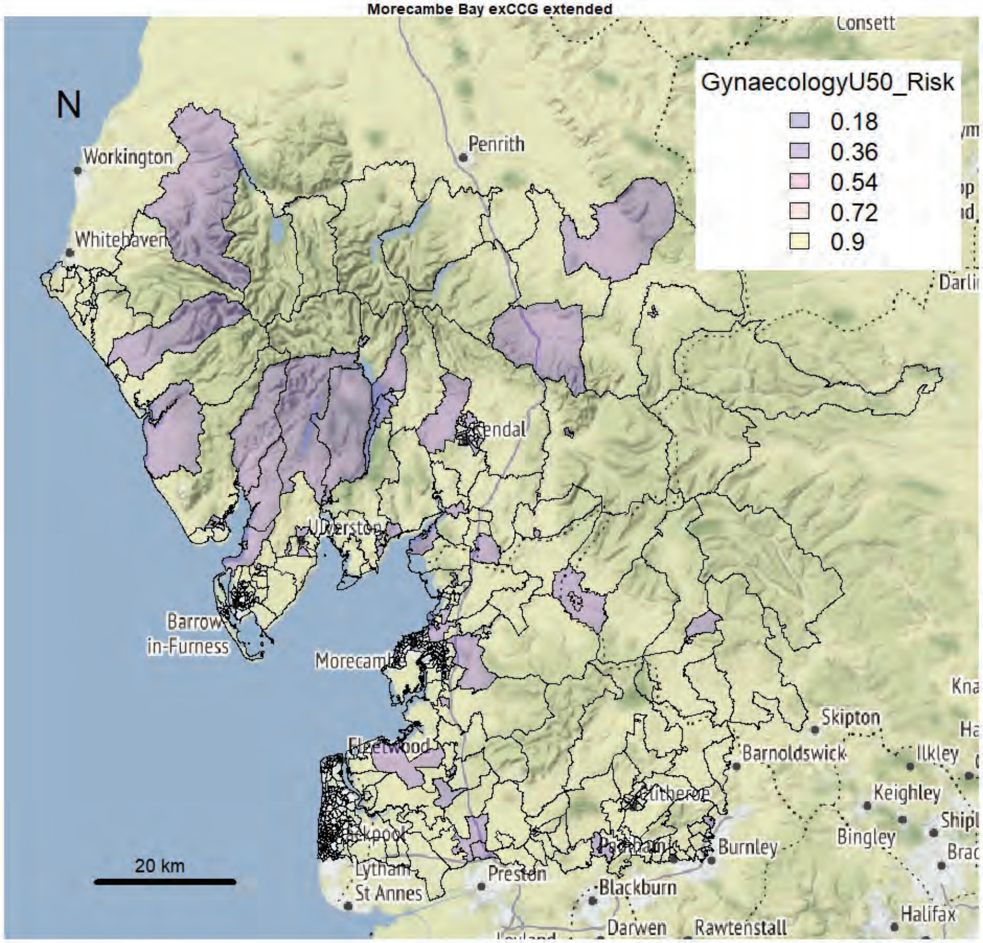


Morecambe Bay exCCG extended

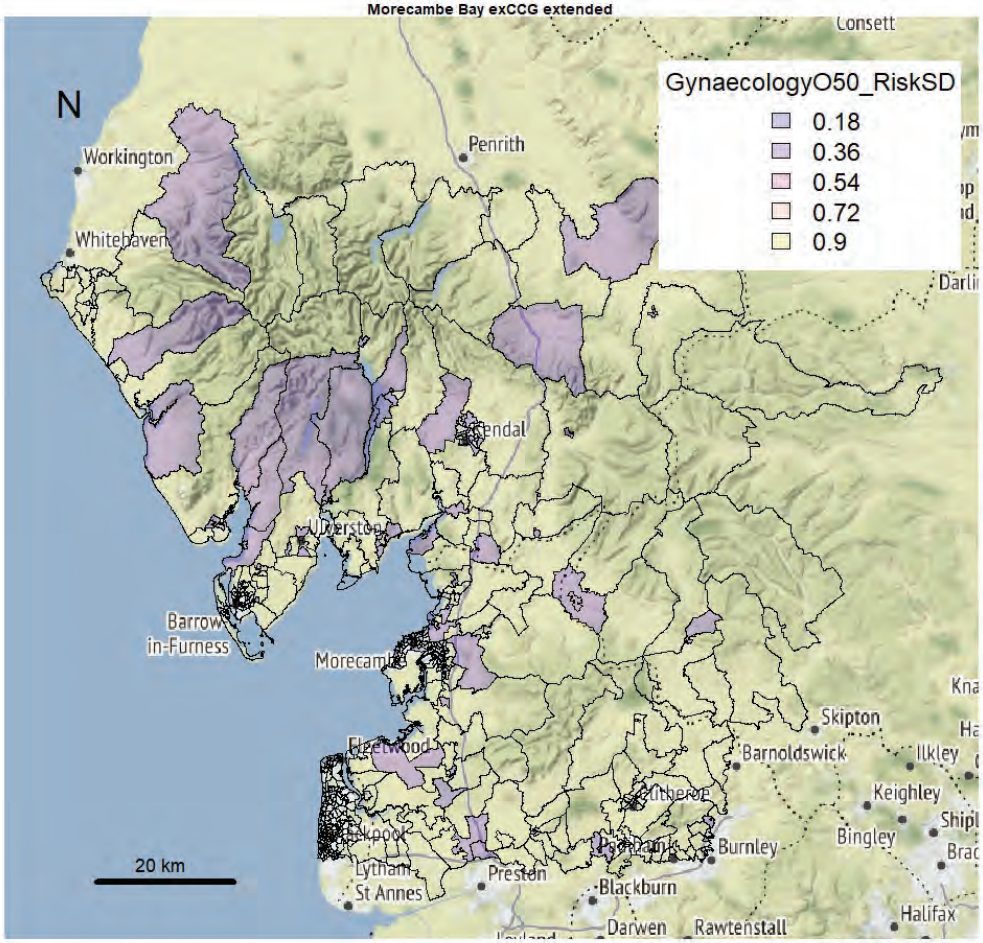

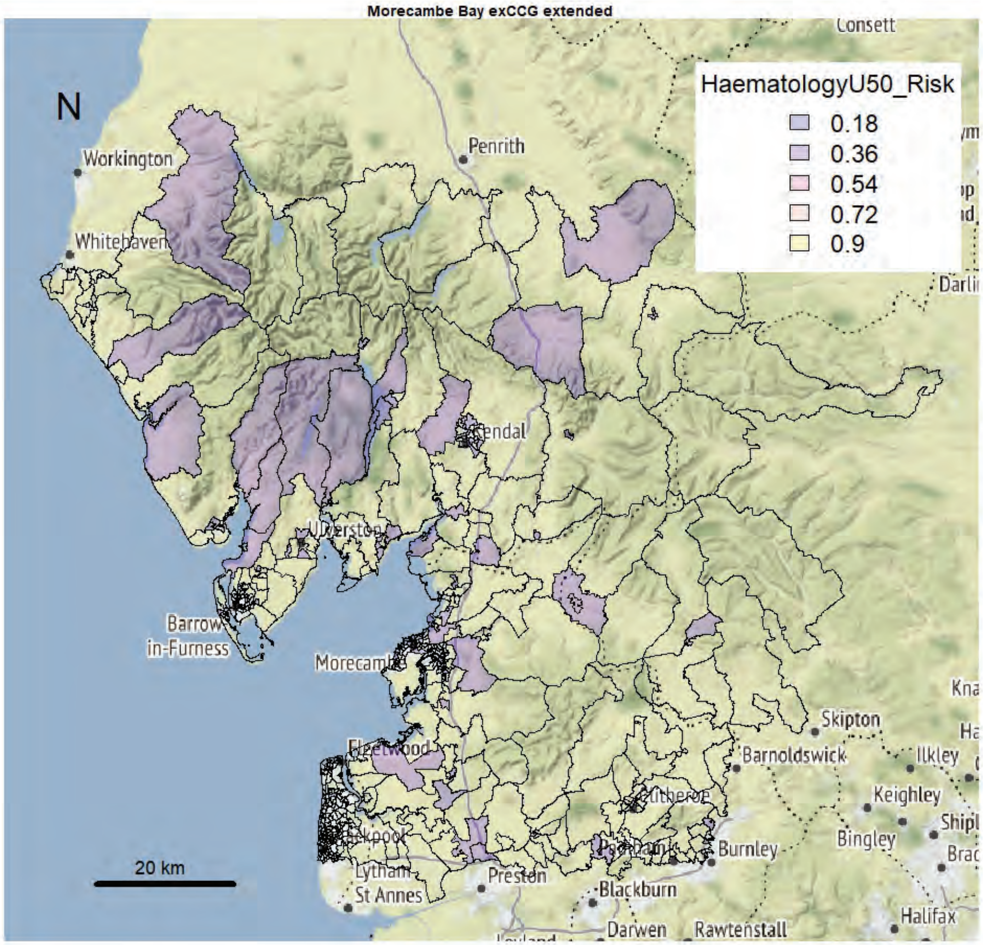
Morecambe Bay exCCG extended
N
HaematologyU50_Risk
0.18
0.36
0.54
0.72
0.9
Workington
Whitehaven
Penrith
Consett
Kendal
Ulverston
Barrow-in-Furness
Morecambe
Fleetwood
Blackpool
Lytham St Annes
Preston
Blackburn
Darwen
Rawtenstall
Burnley
Clitheroe
Barnoldswick
Skipton
Ilkley
Keighley
Bingley
Halifax
20 km

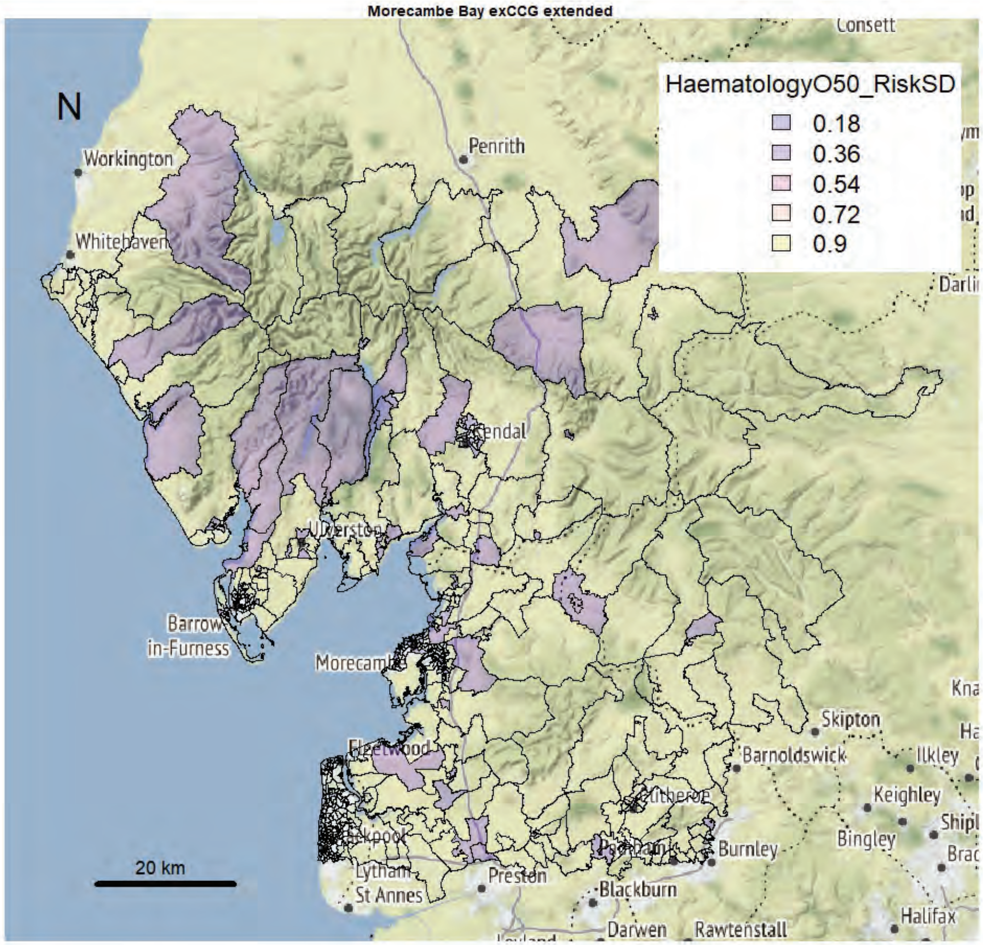
Morecambe Bay exCCG extended
N
HaematologyO50_RiskSD
0.18
0.36
0.54
0.72
0.9
Workington
Whitehaven
Penrith
Consett
Kendal
Ulverston
Barrow-in-Furness
Morecambe
Fleetwood
Blackpool
Lytham St Annes
Preston
Blackburn
Darwen
Rawtenstall
Burnley
Clitheroe
Barnoldswick
Skipton
Ilkley
Keighley
Bingley
Halifax
20 km

Morecambe Bay exCCG extended

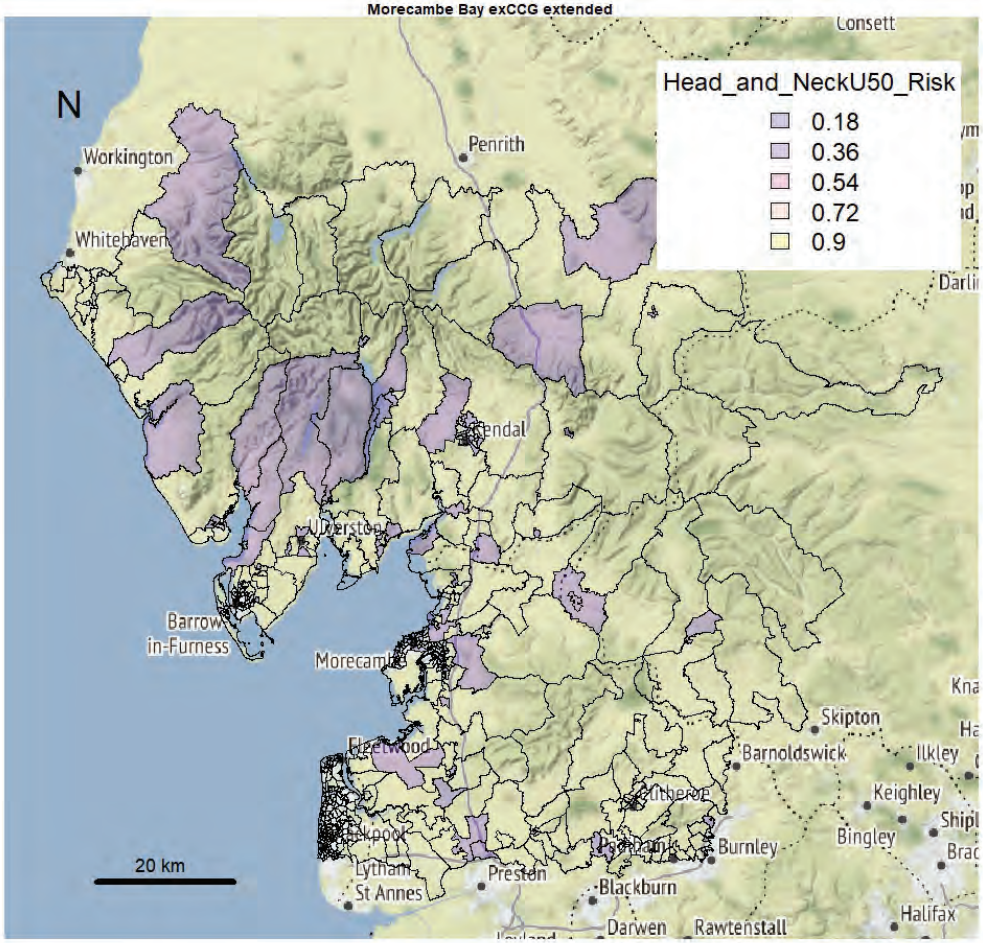


Morecambe Bay exCCG extended

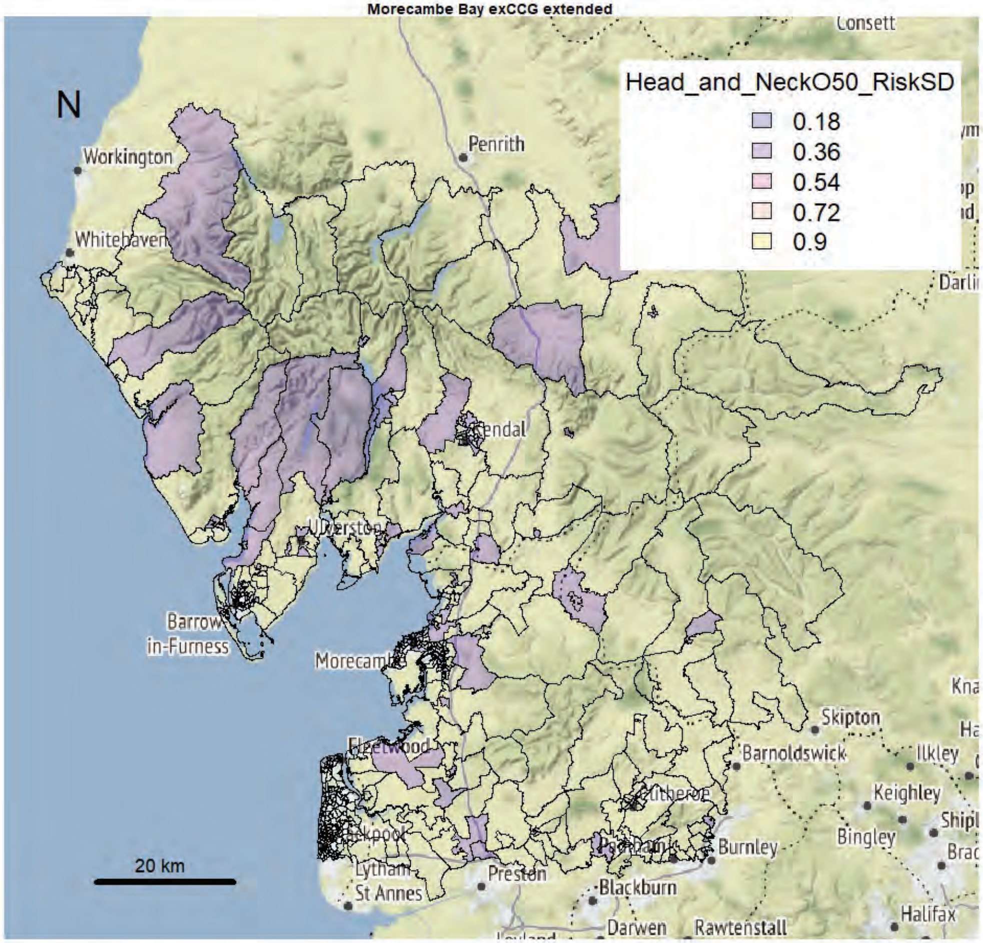

Morecambe Bay exCCG extended

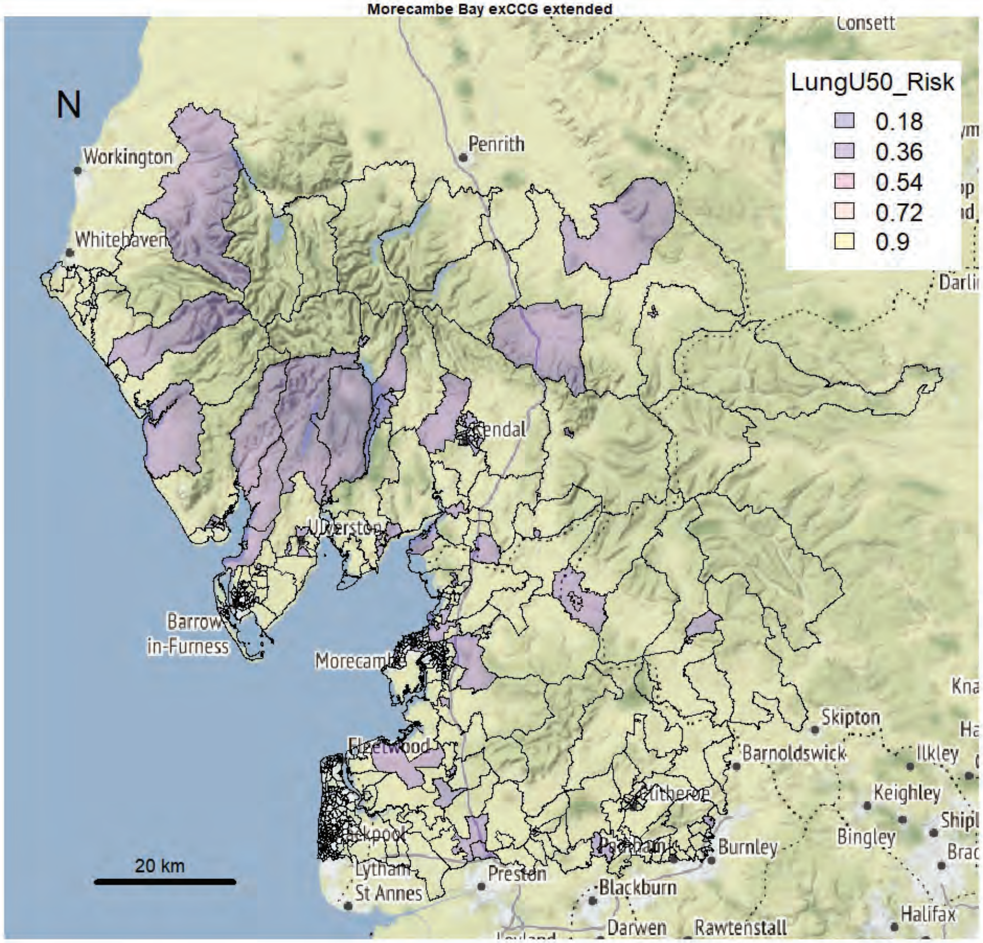


Morecambe Bay exCCG extended

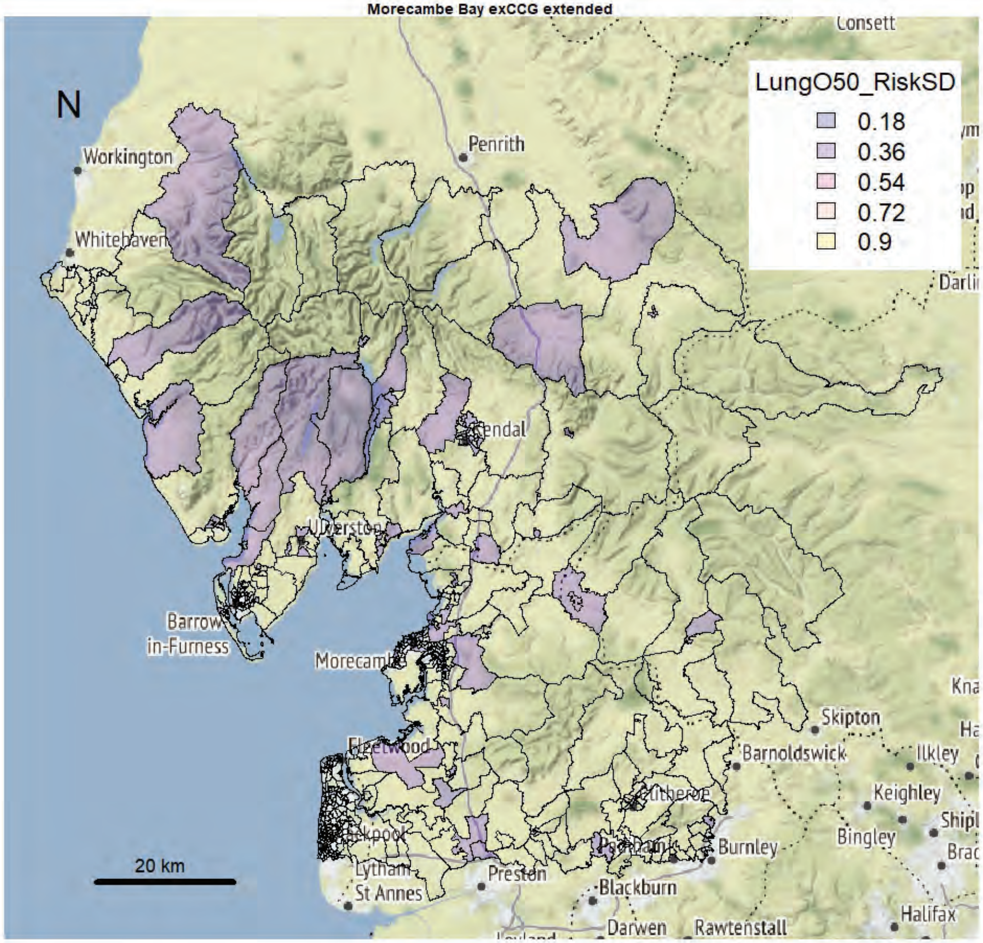

Morecambe Bay exCCG extended

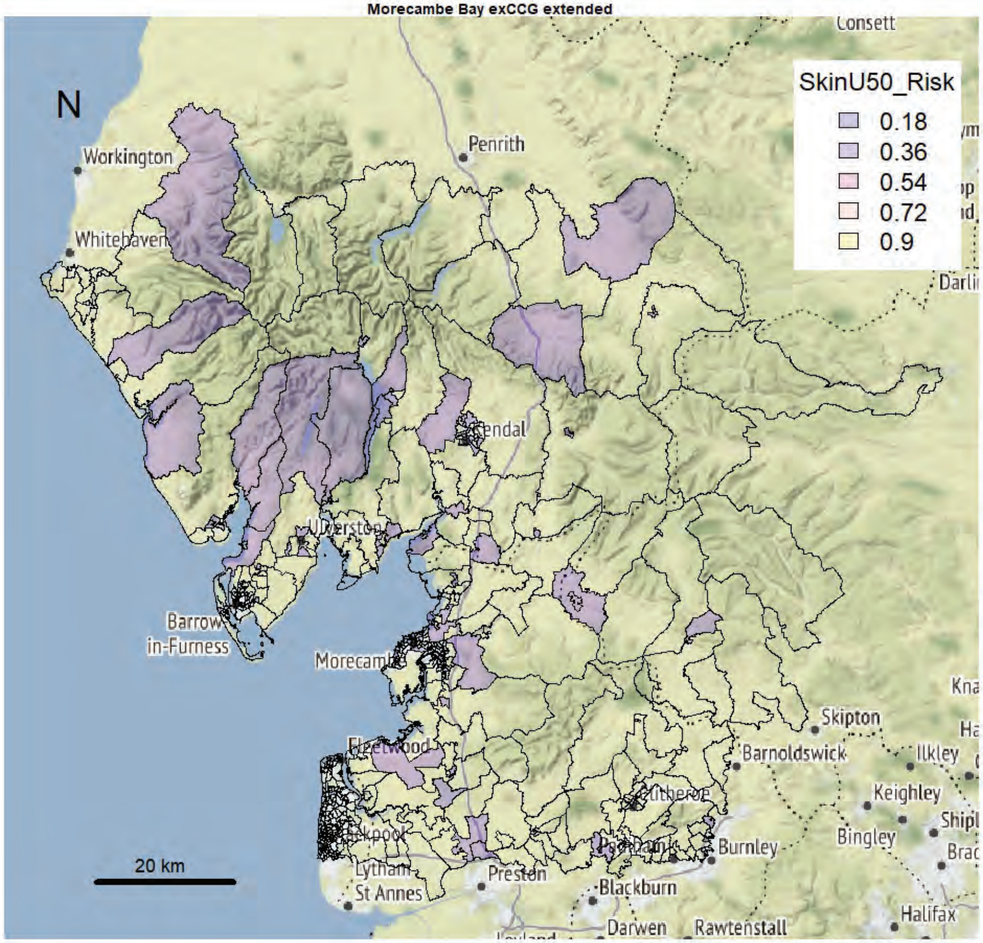


Morecambe Bay exCCG extended

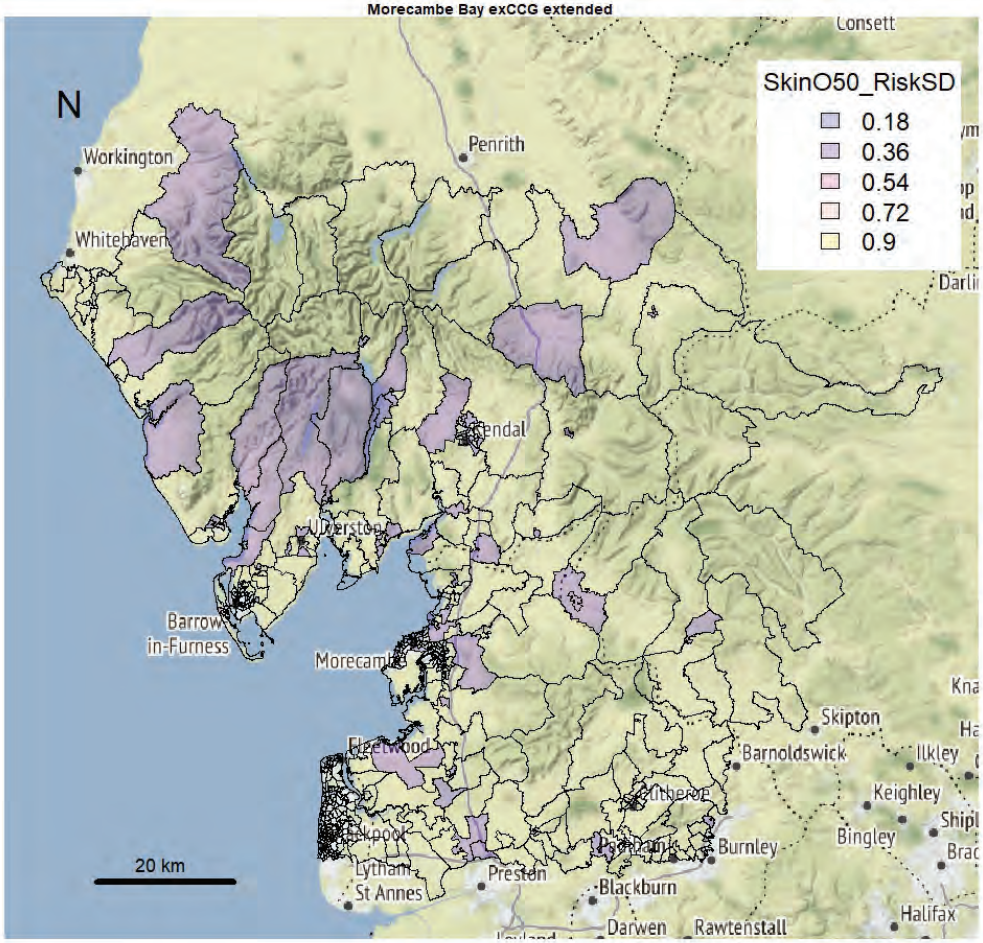

Morecambe Bay exCCG extended

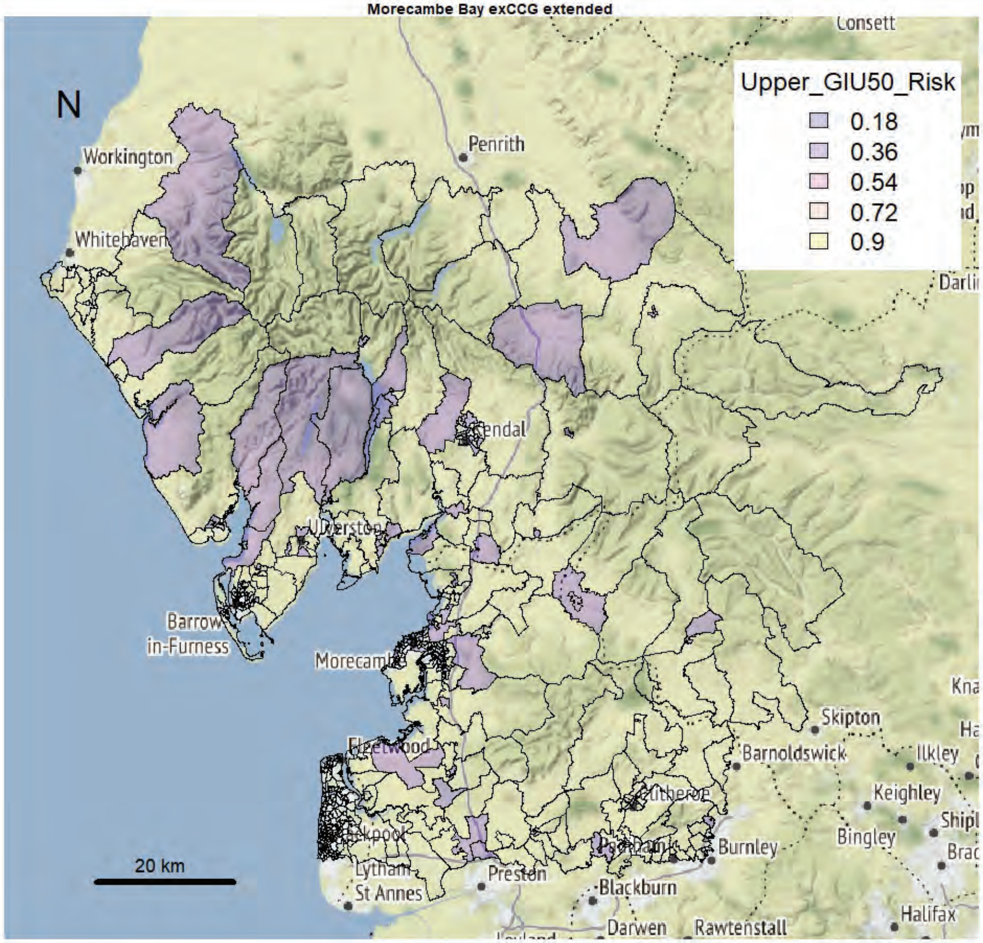


Morecambe Bay exCCG extended

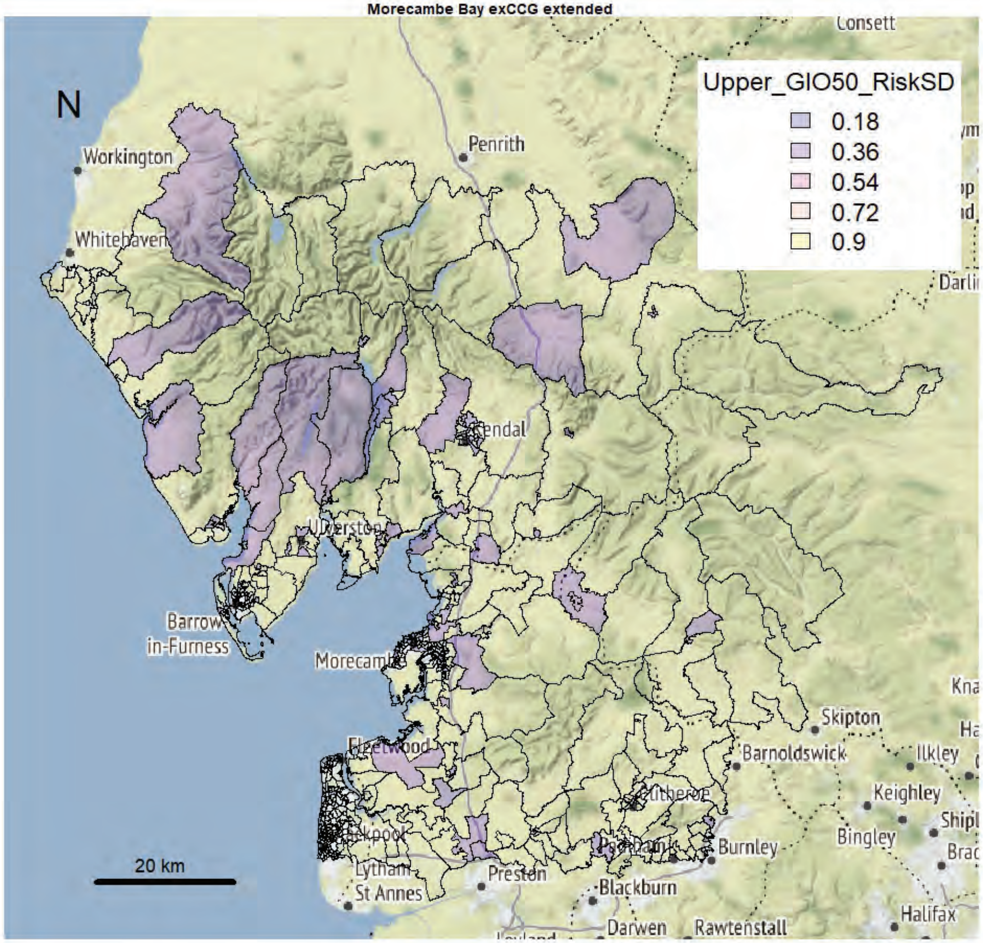

Morecambe Bay exCCG extended

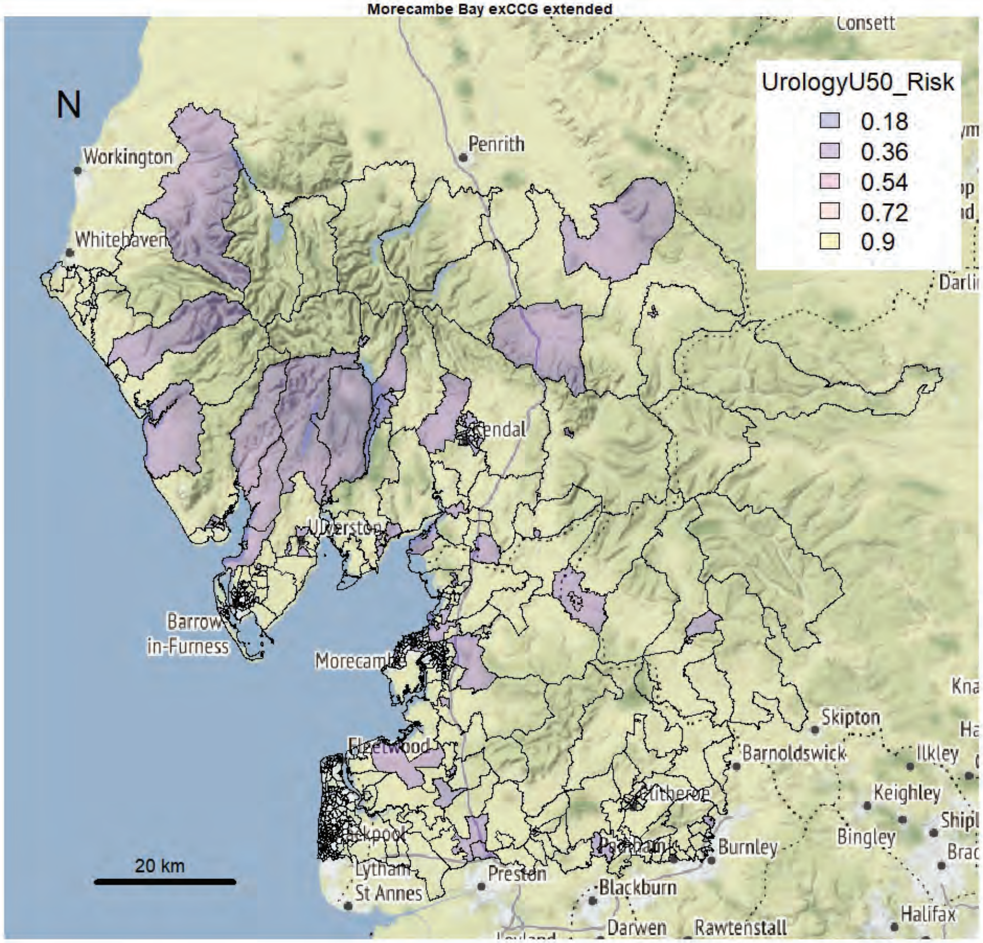


Morecambe Bay exCCG extended

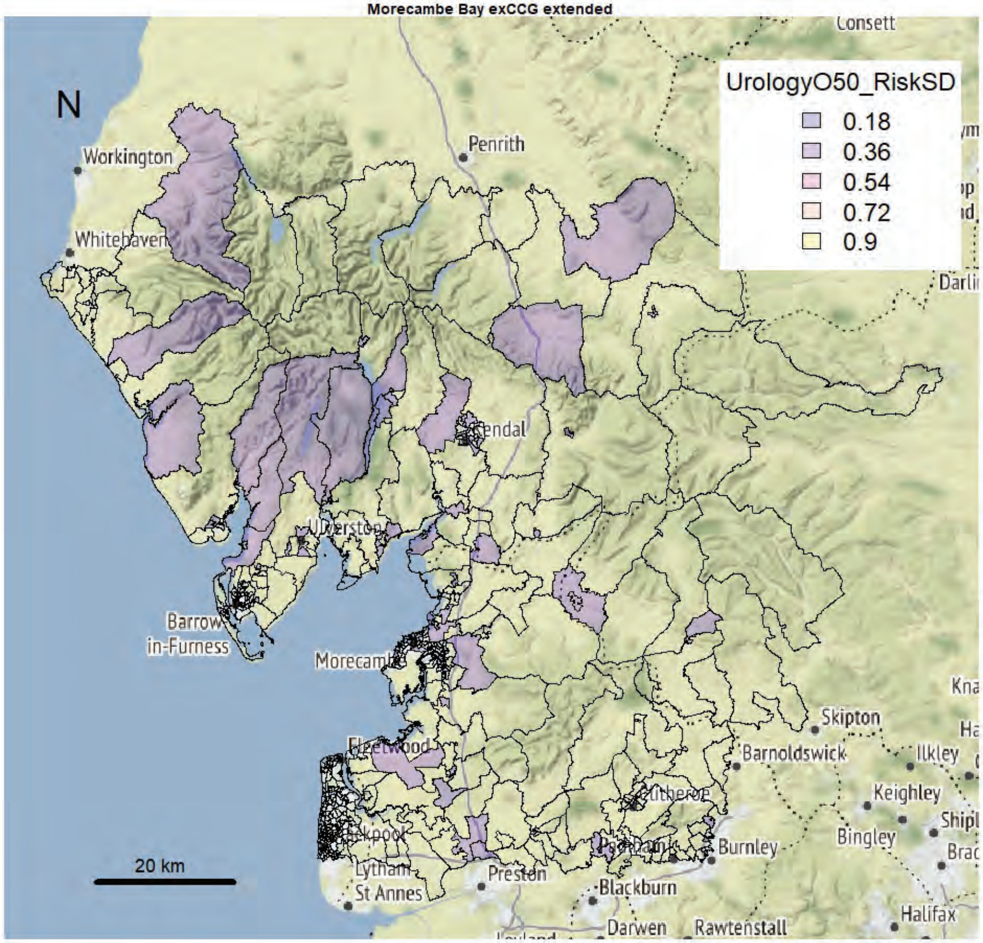

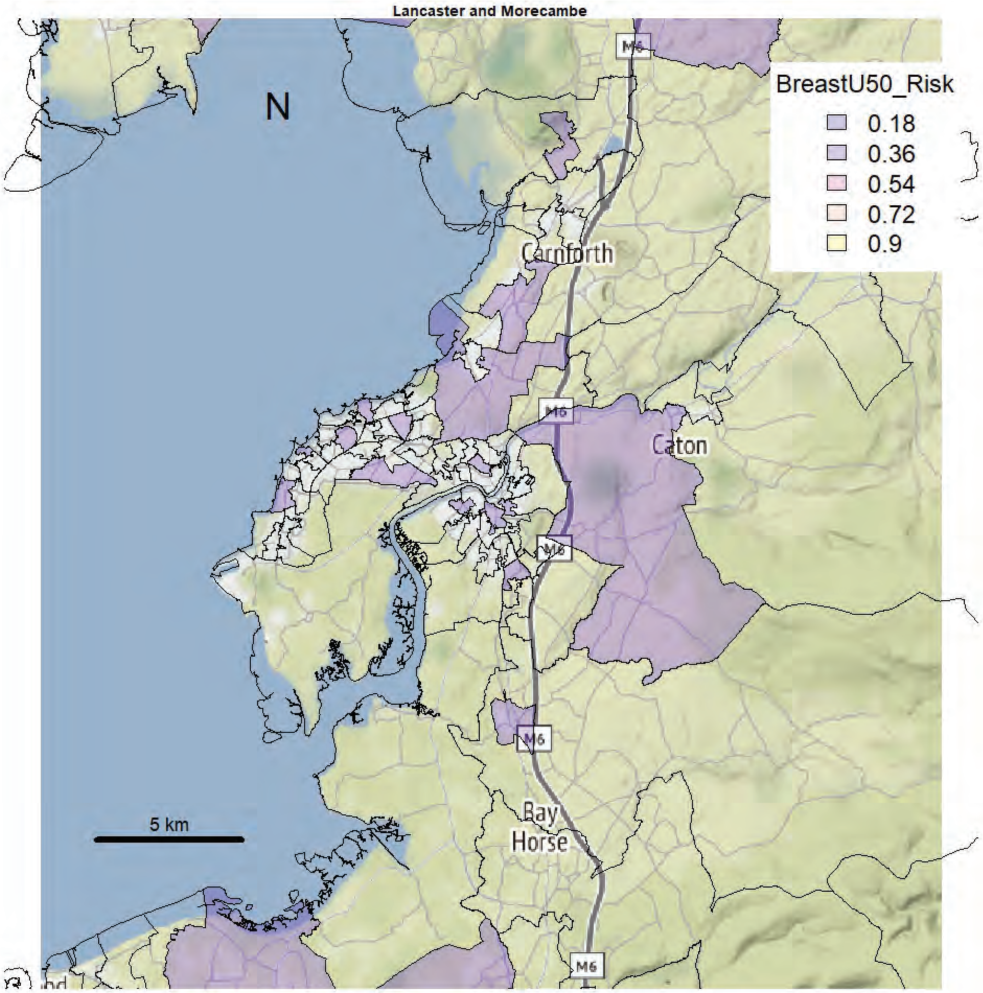
Lancaster and Morecambe
N
BreastU50_Risk
0.18
0.36
0.54
0.72
0.9
Carnforth
Caton
Bay
Horse
M6
5 km

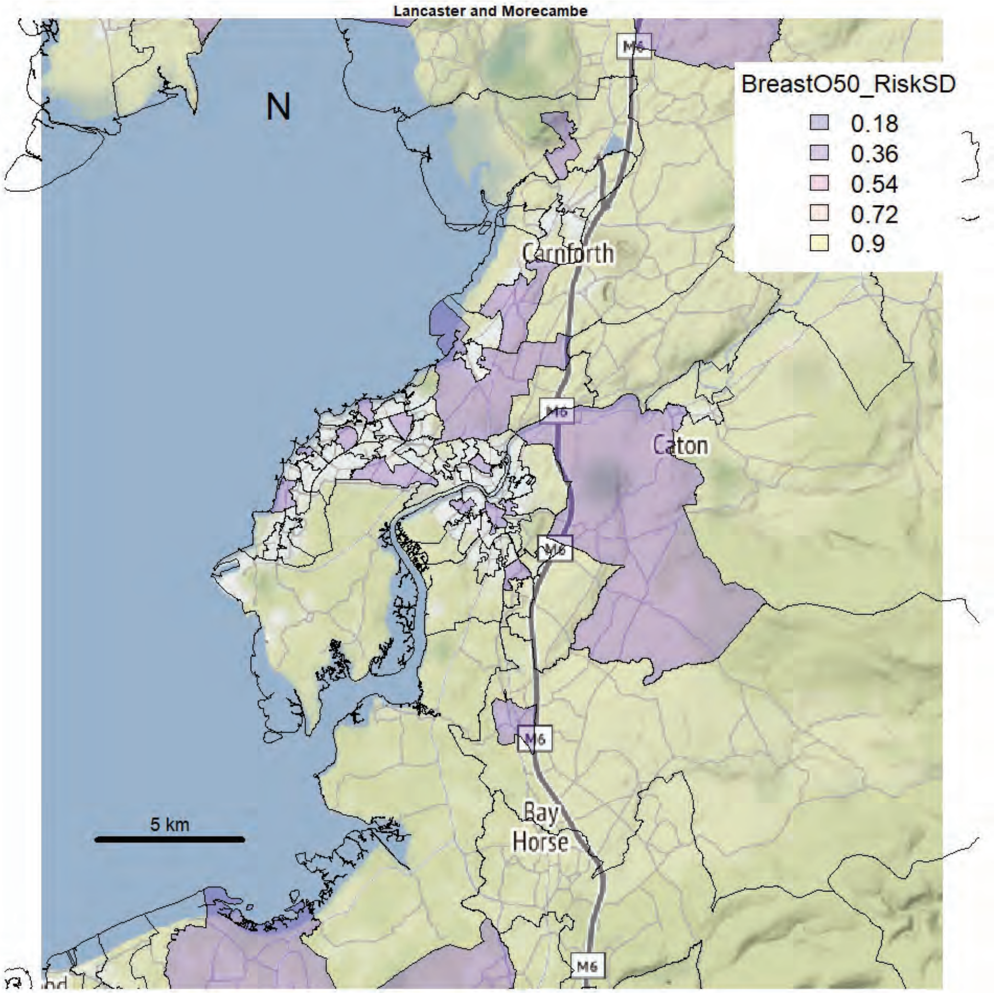
Lancaster and Morecambe
N
BreastO50_RiskSD
0.18
0.36
0.54
0.72
0.9
Carnforth
Caton
Bay
Horse
M6
5 km

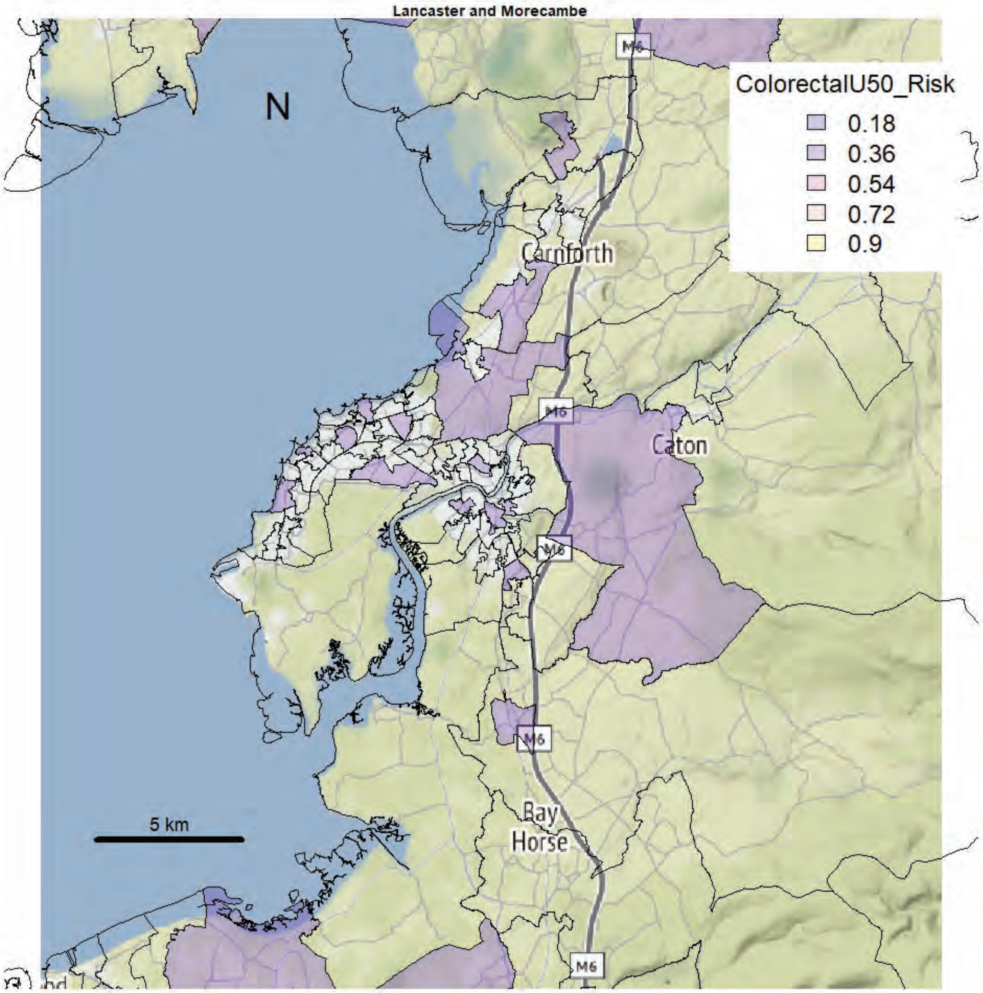
Lancaster and Morecambe
N
ColorectalU50_Risk
0.18
0.36
0.54
0.72
0.9
Carnforth
Caton
M6
Bay Horse
5 km

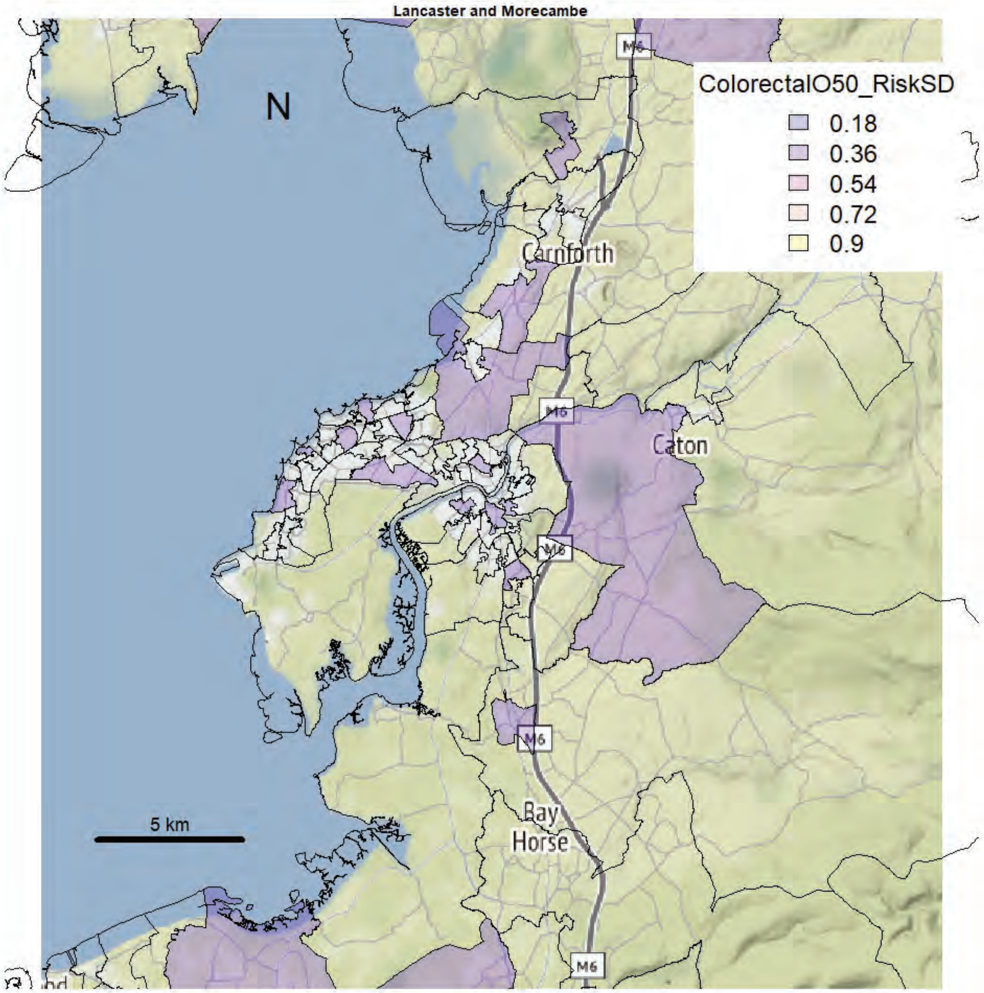
Lancaster and Morecambe
N
ColorectalO50_RiskSD
0.18
0.36
0.54
0.72
0.9
Carnforth
Caton
M6
Bay Horse
5 km

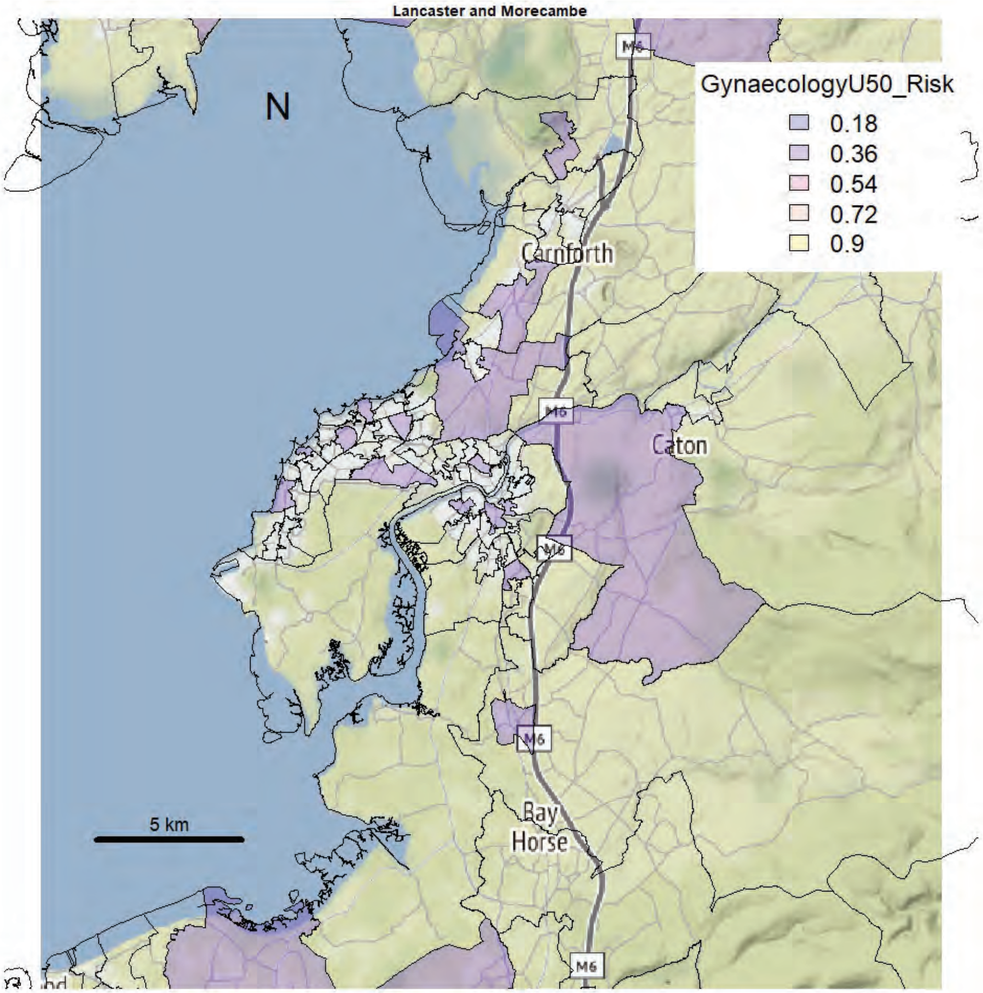
Lancaster and Morecambe
N
GynaecologyU50_Risk
0.18
0.36
0.54
0.72
0.9
Carnforth
Caton
M6
Bay Horse
5 km

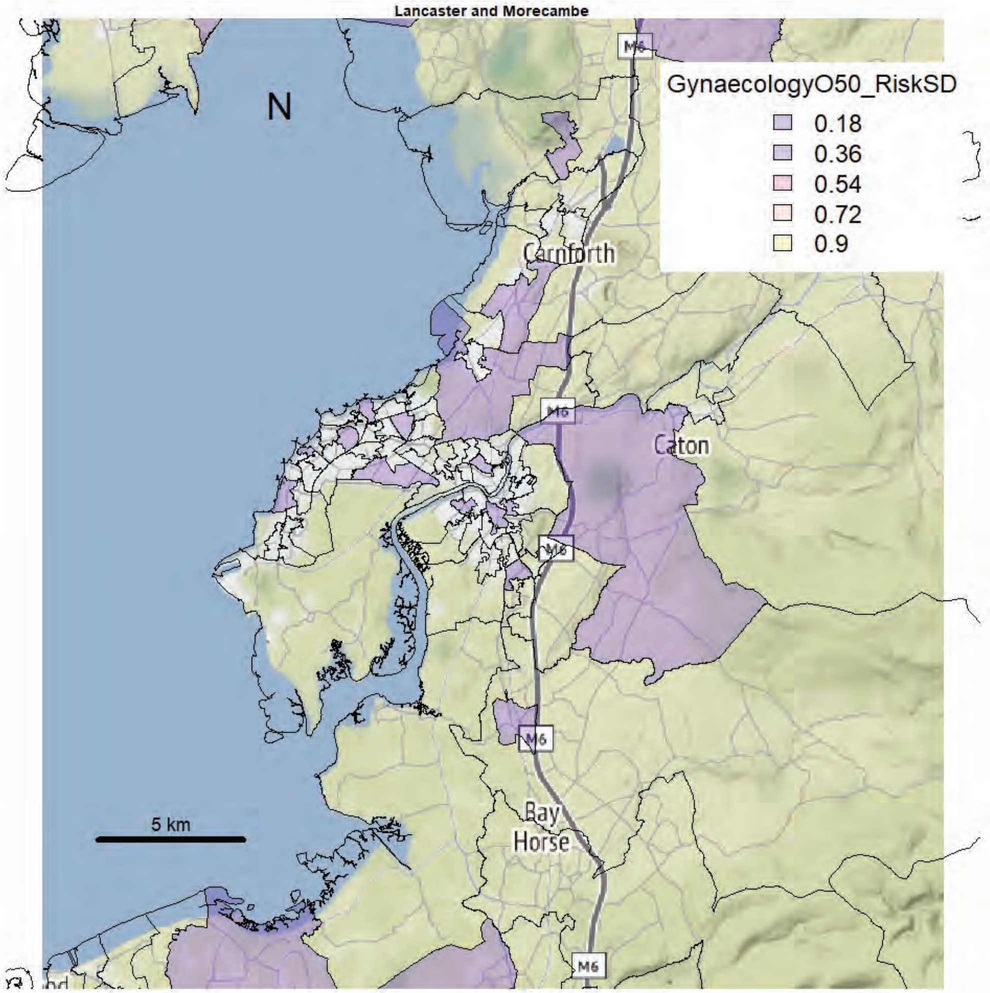
Lancaster and Morecambe
N
GynaecologyO50_RiskSD
0.18
0.36
0.54
0.72
0.9
Carnforth
Caton
M6
Bay Horse
5 km

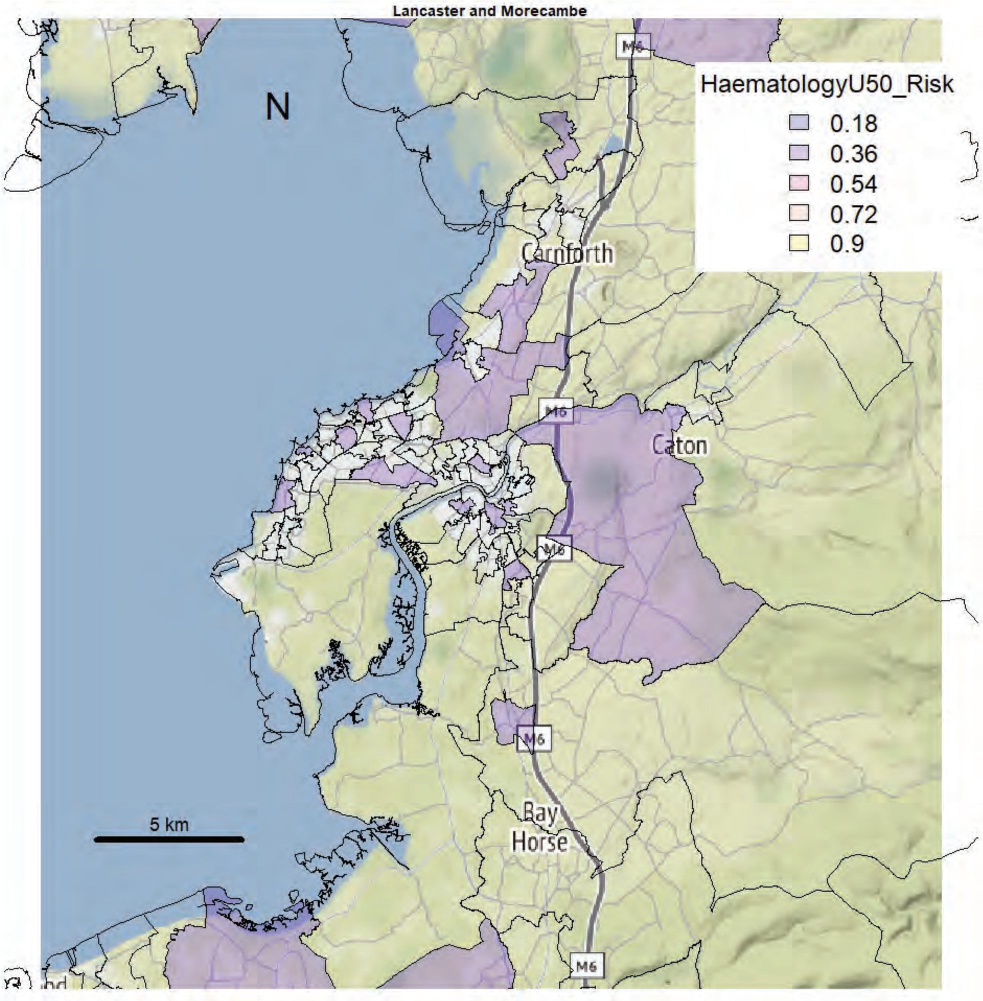
Lancaster and Morecambe
N
HaematologyU50_Risk
0.18
0.36
0.54
0.72
0.9
Carnforth
Caton
M6
Bay
Horse
5 km

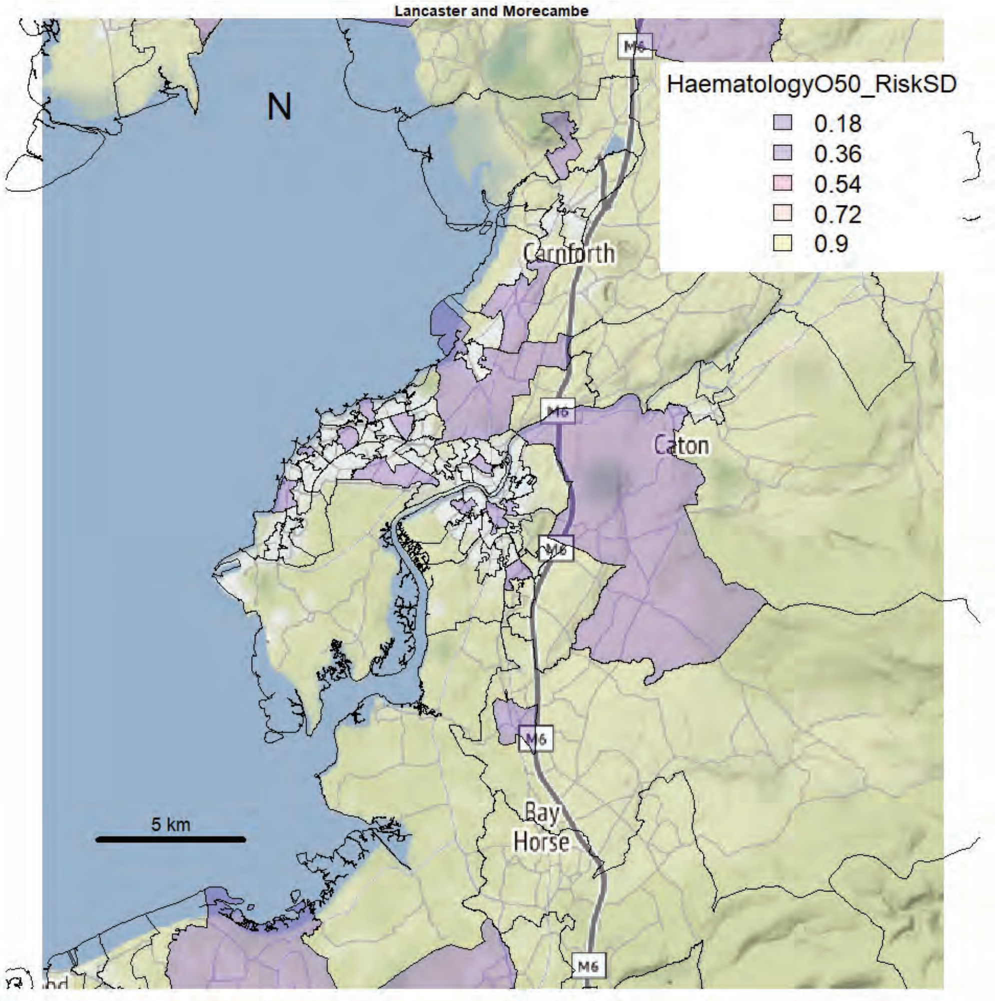
Lancaster and Morecambe
N
HaematologyO50_RiskSD
0.18
0.36
0.54
0.72
0.9
Carnforth
Caton
M6
Bay
Horse
5 km

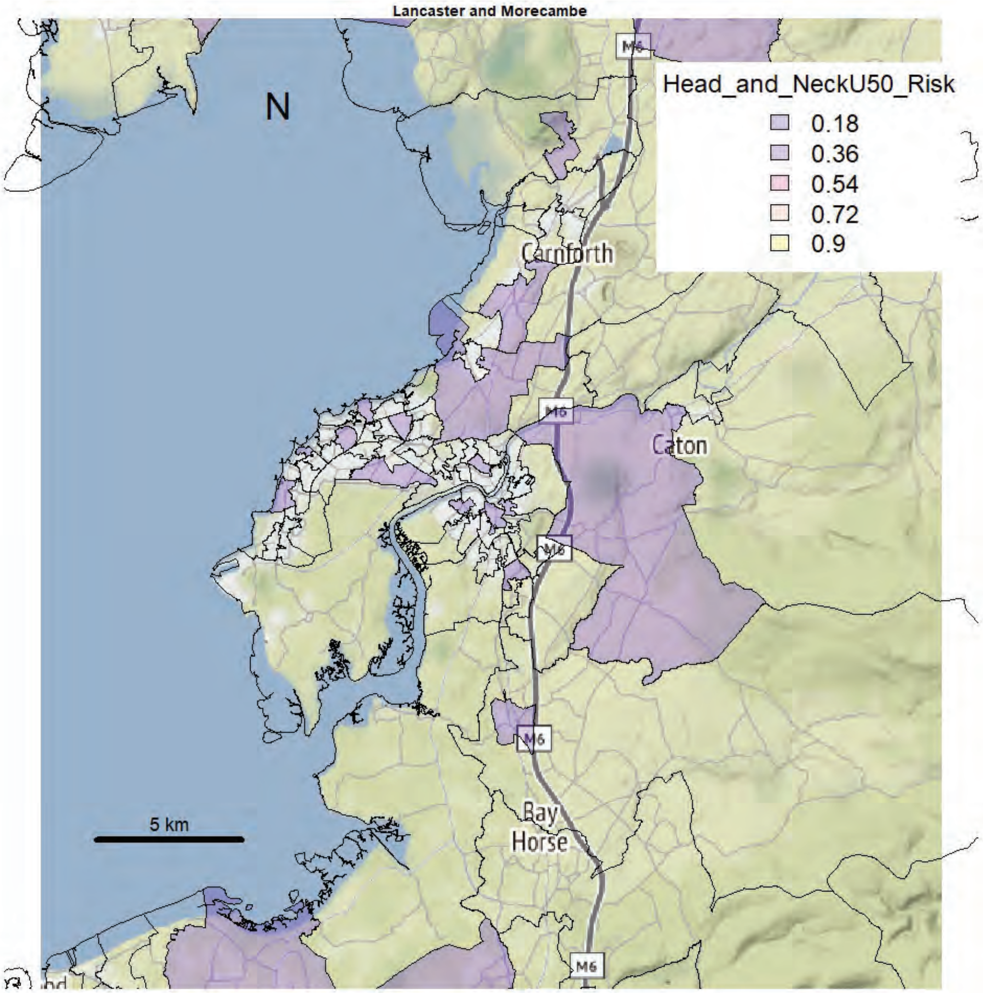
Lancaster and Morecambe
N
Head_and_NeckU50_Risk
0.18
0.36
0.54
0.72
0.9
Carnforth
Caton
M6
Bay
Horse
5 km

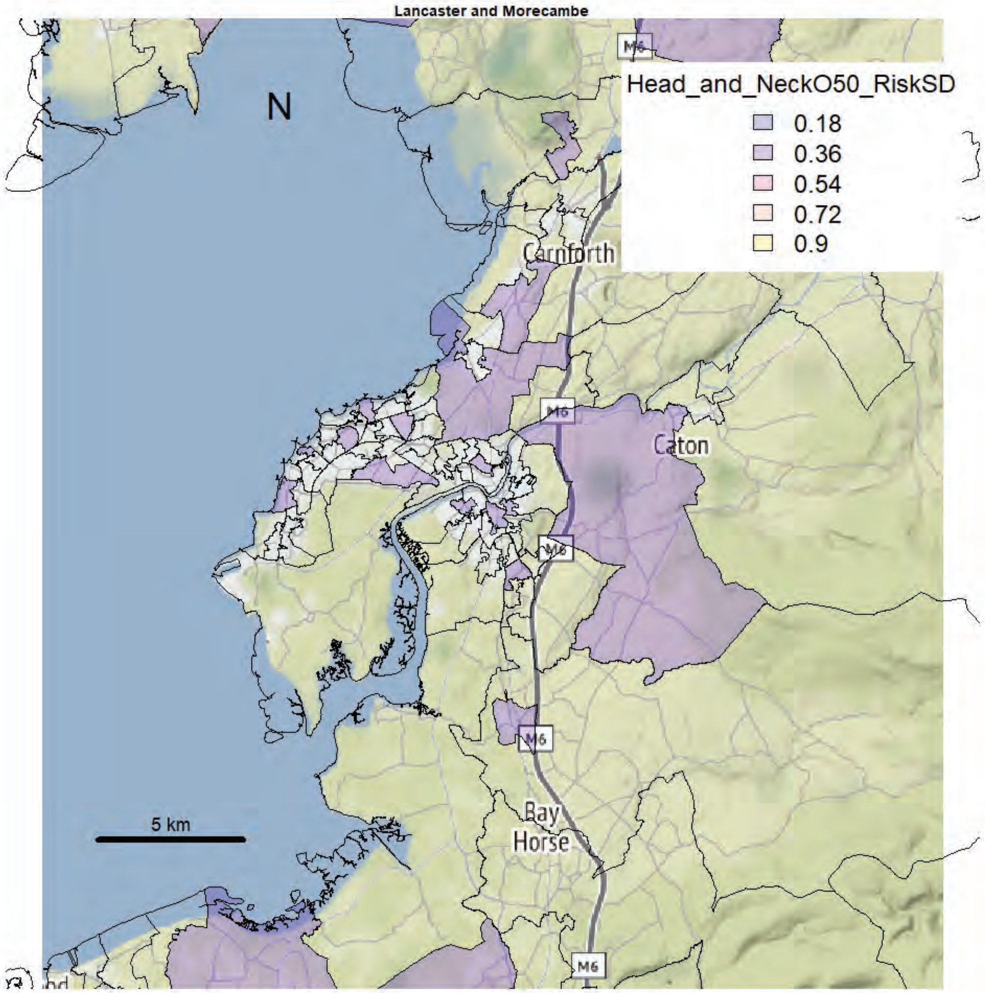
Lancaster and Morecambe
N
Head_and_NeckO50_RiskSD
0.18
0.36
0.54
0.72
0.9
Carnforth
Caton
M6
Bay
Horse
5 km

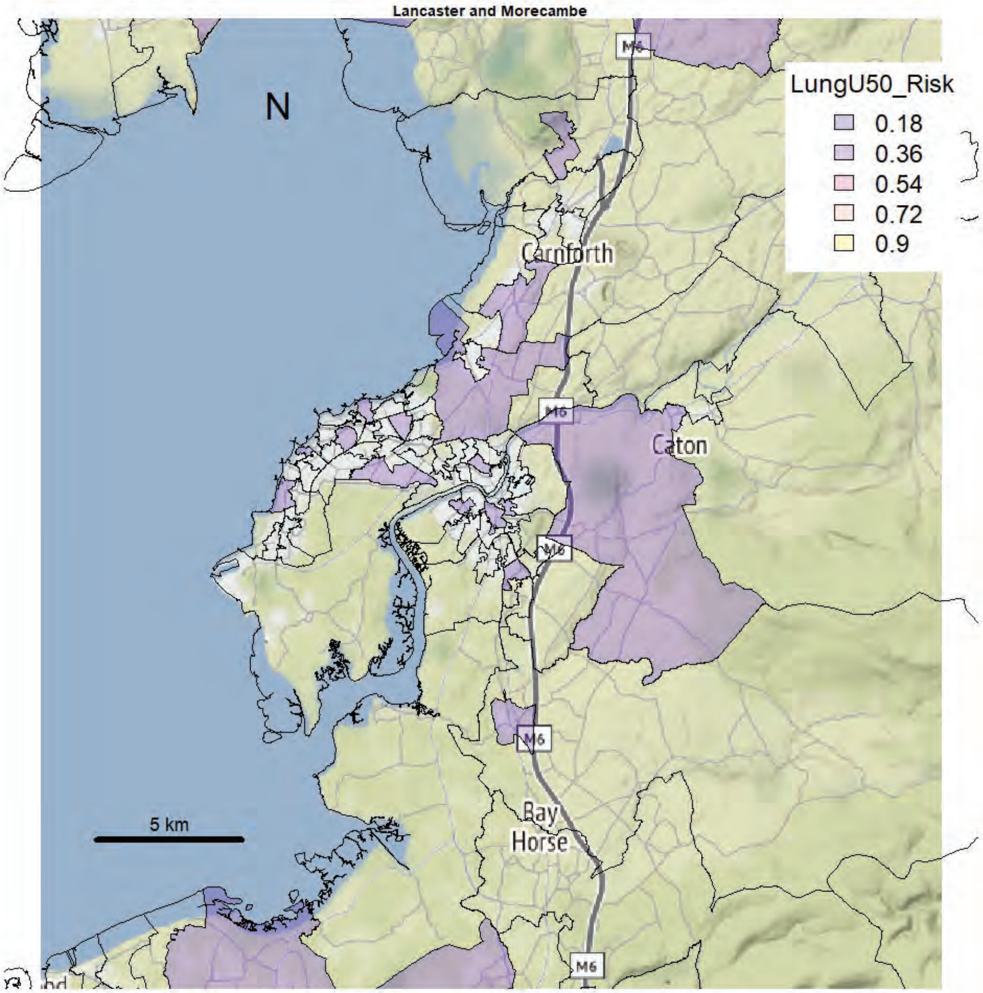
Lancaster and Morecambe
N
LungU50_Risk
0.18
0.36
0.54
0.72
0.9
Carnforth
Caton
M6
Bay
Horse
5 km

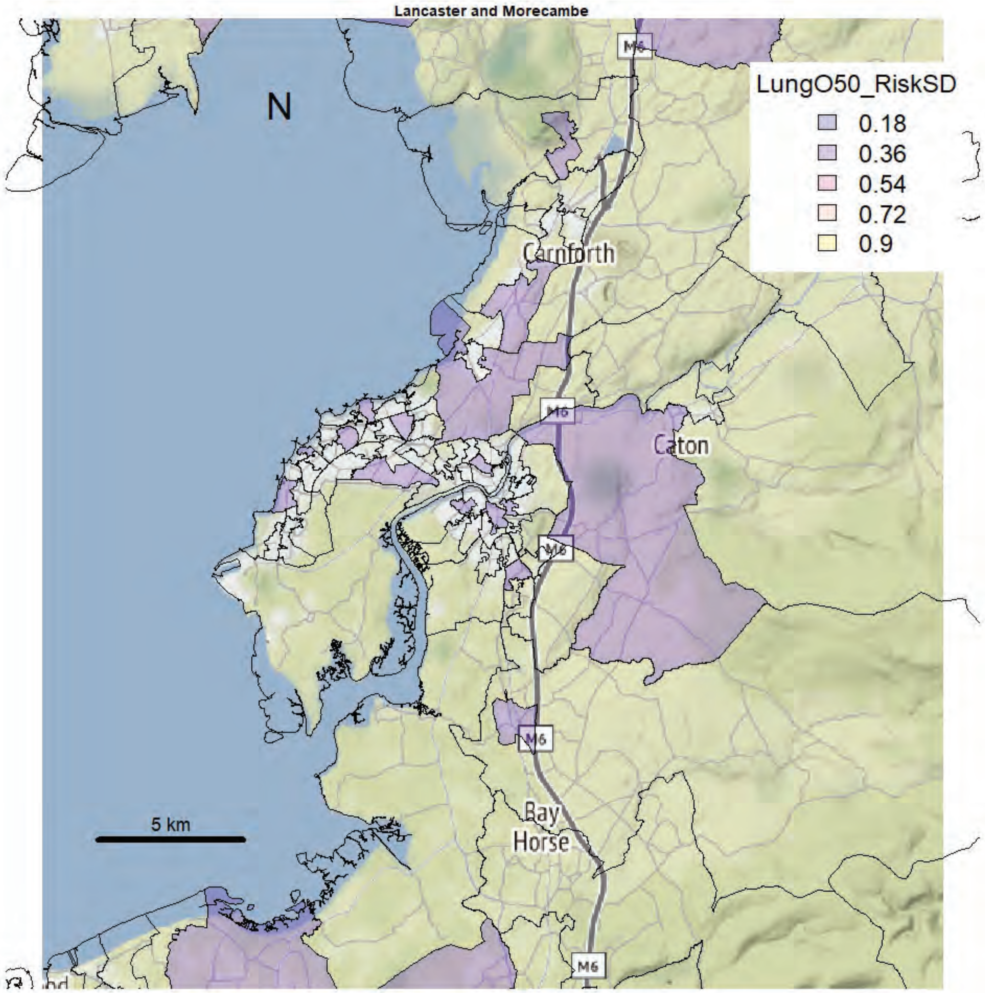
Lancaster and Morecambe
N
LungO50_RiskSD
0.18
0.36
0.54
0.72
0.9
Carnforth
Caton
M6
Bay
Horse
5 km

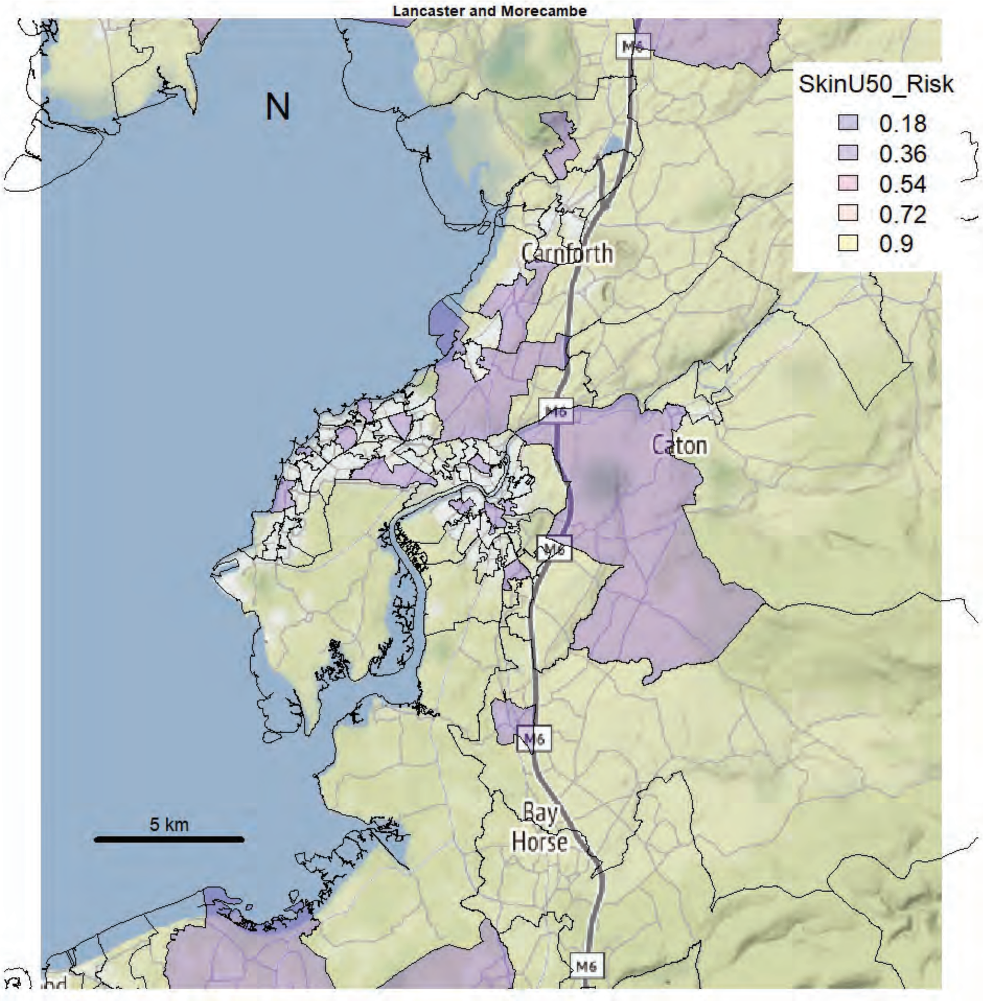
Lancaster and Morecambe
N
SkinU50_Risk
0.18
0.36
0.54
0.72
0.9
Carnforth
Caton
Bay
Horse
M6
5 km

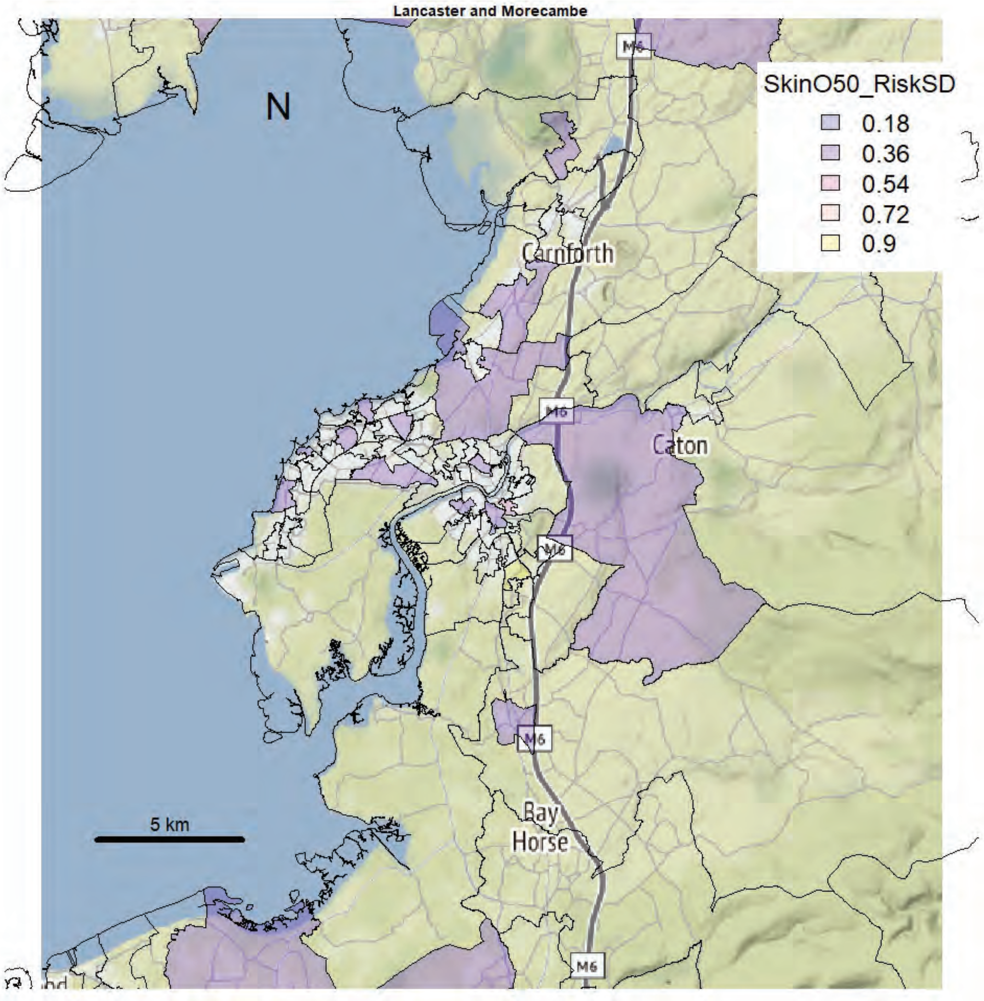
Lancaster and Morecambe
N
SkinO50_RiskSD
0.18
0.36
0.54
0.72
0.9
Carnforth
Caton
Bay
Horse
M6
5 km

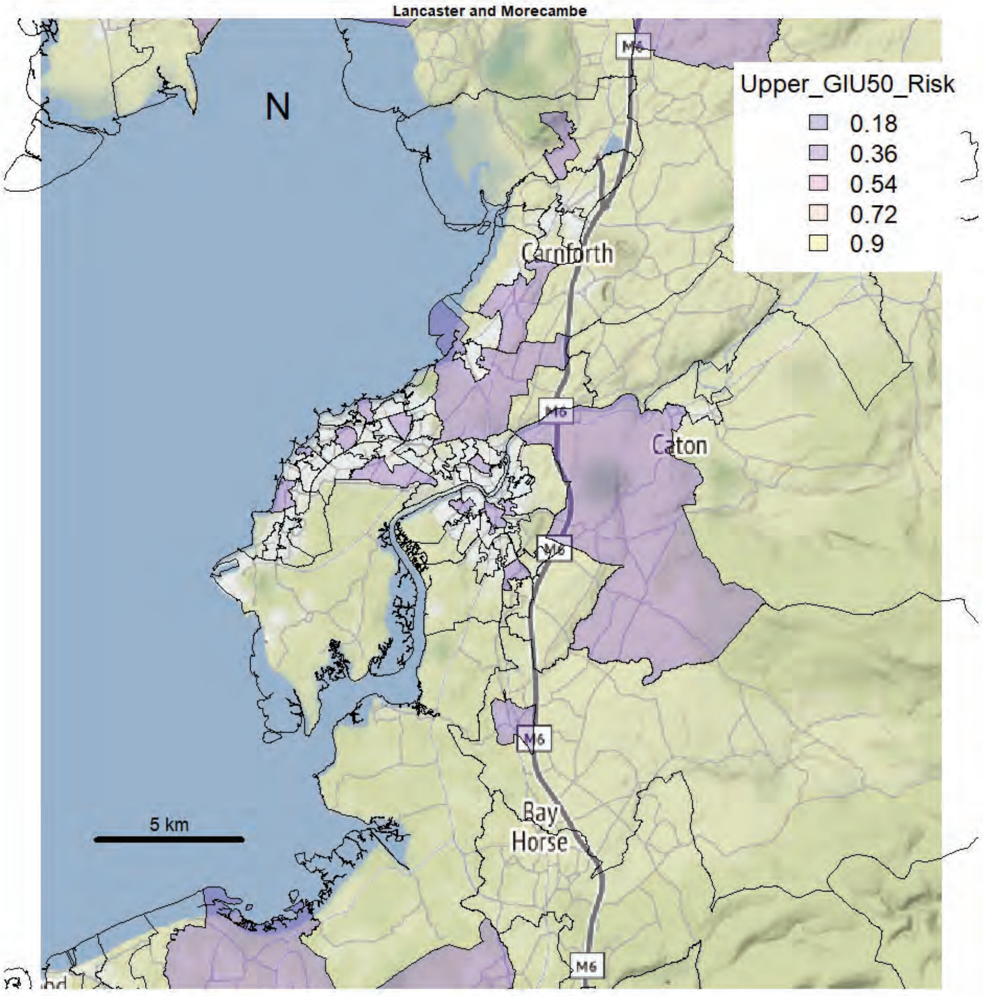

Lancaster and Morecambe
Upper_GIU50_Risk
0.18
0.36
0.54
0.72
0.9
N
Carnforth
Caton
Bay
Horse
M6
5 km


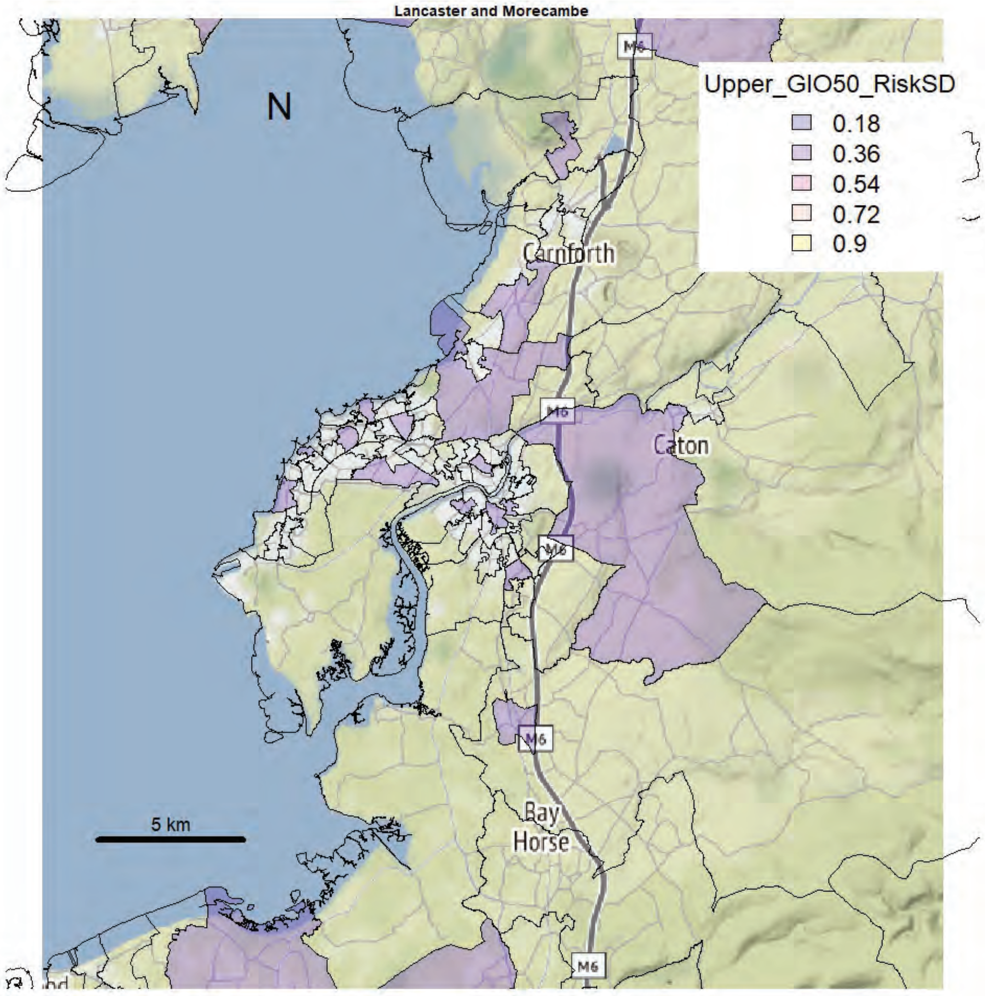

Lancaster and Morecambe
Upper_GIO50_RiskSD
0.18
0.36
0.54
0.72
0.9
N
Carnforth
Caton
Bay
Horse
M6
5 km

Lancaster and Morecambe

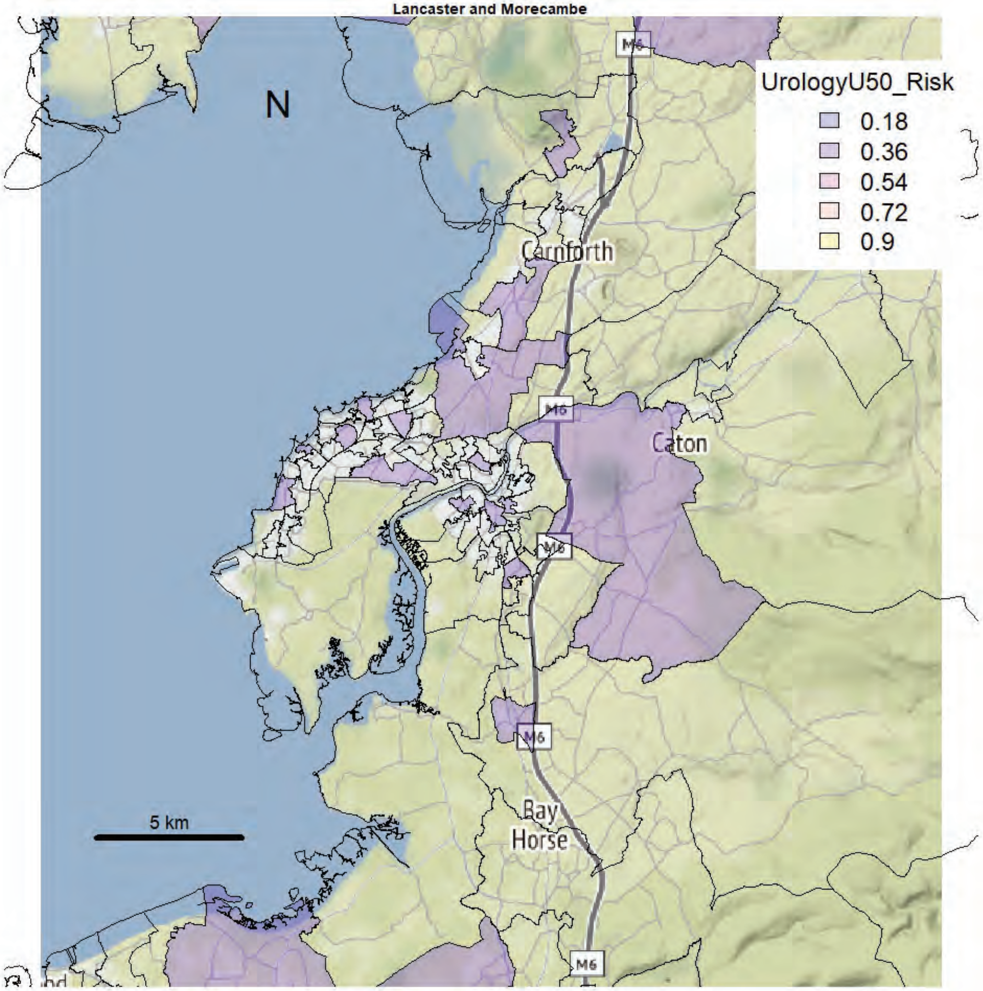


Lancaster and Morecambe

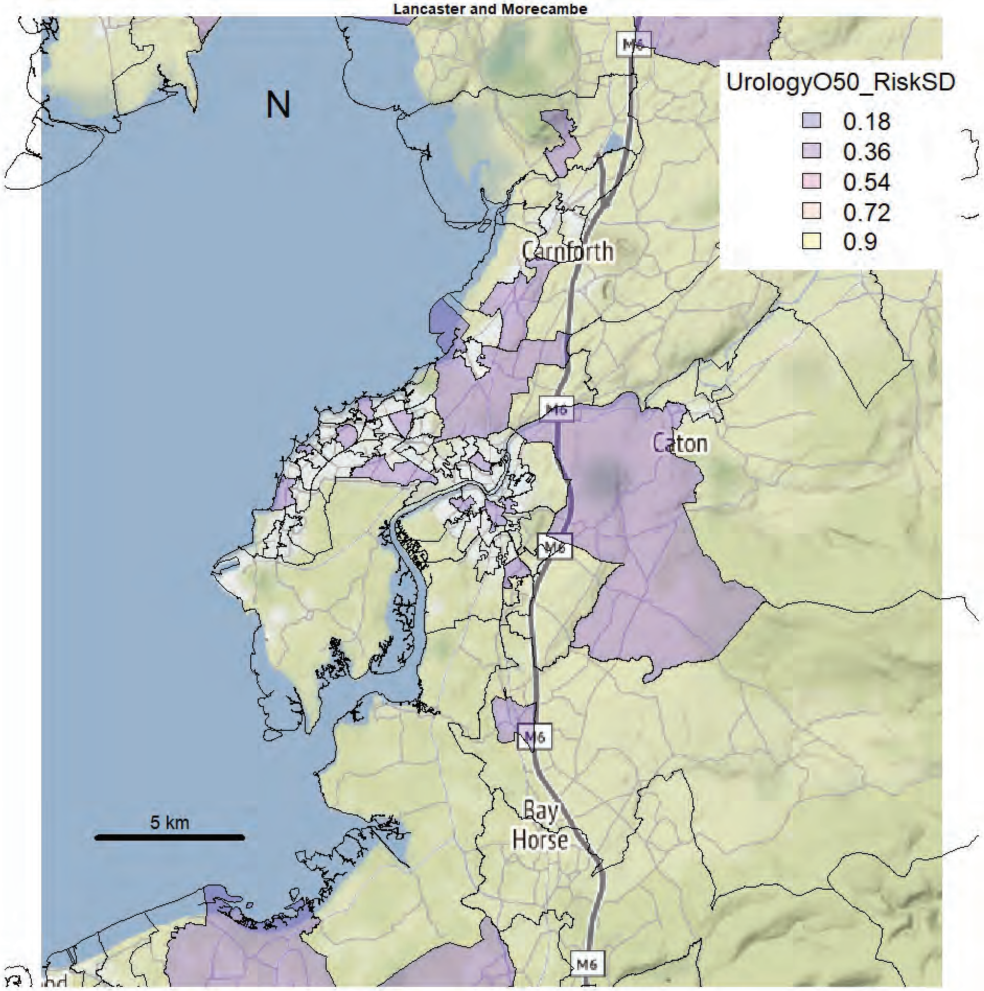

Barrow

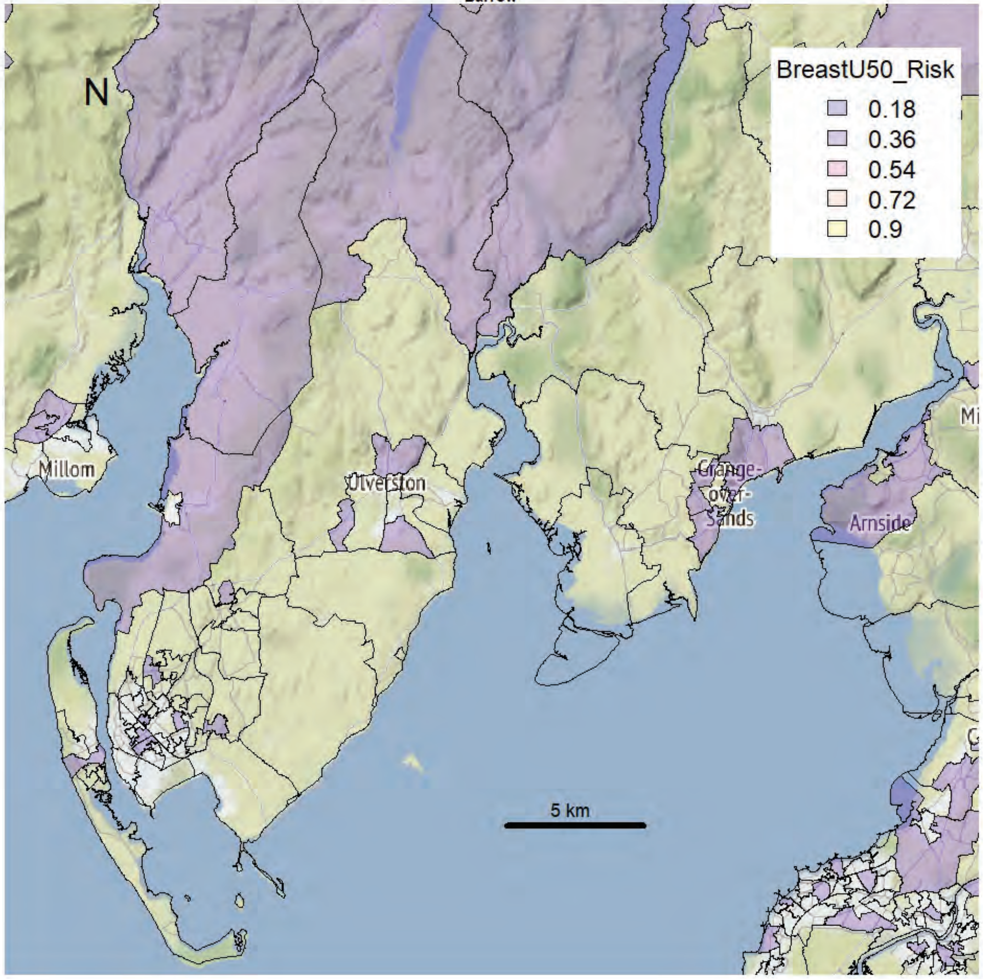


Barrow

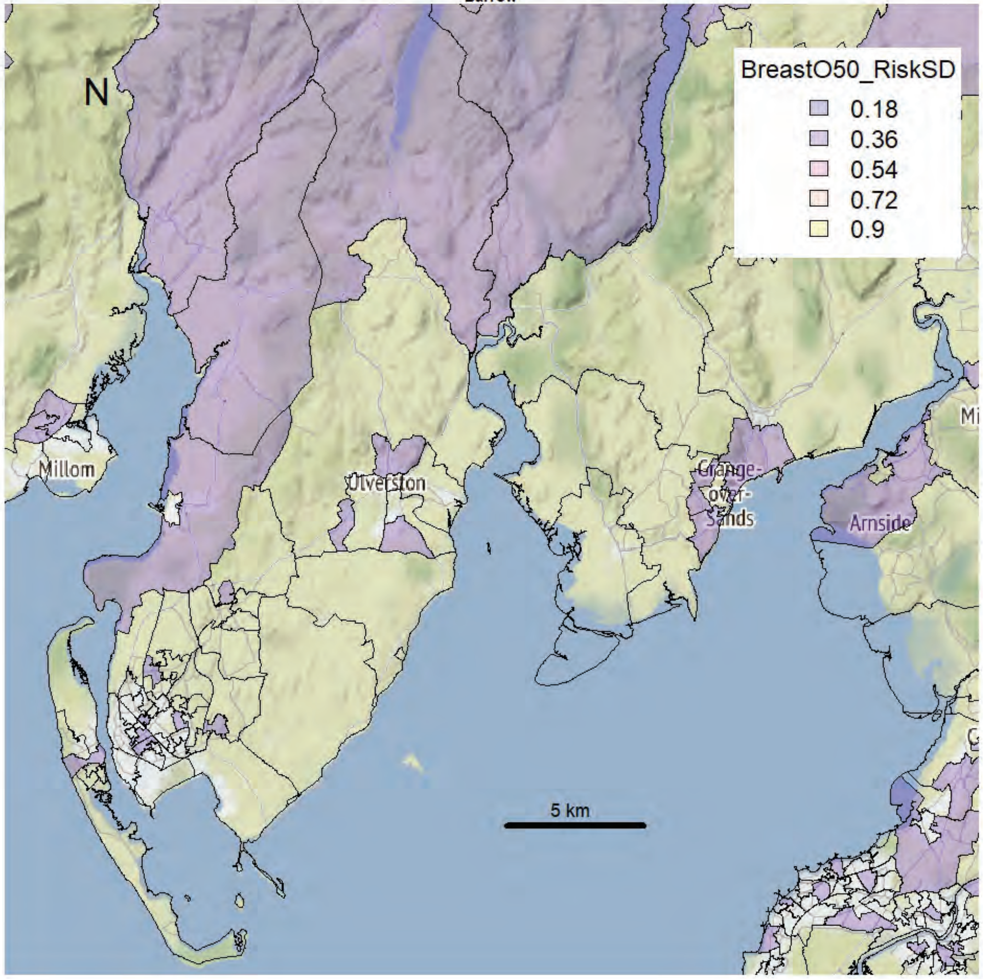

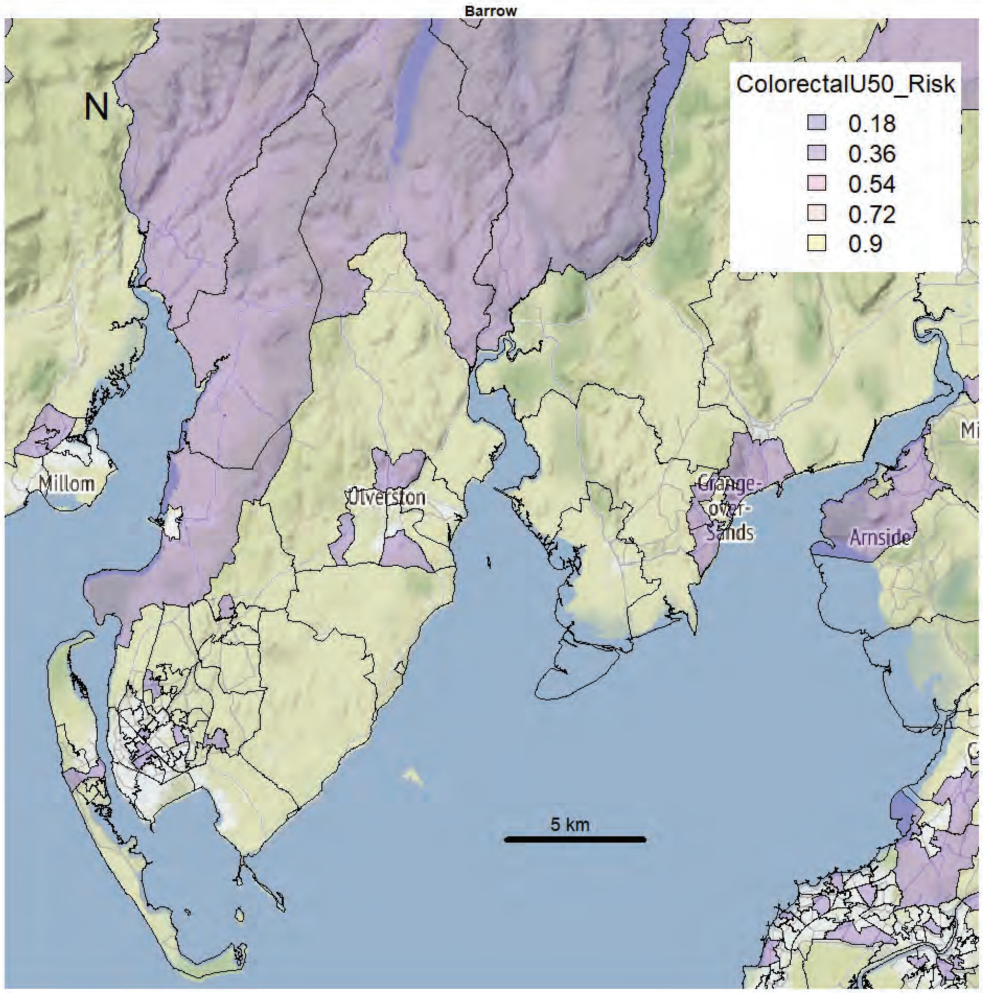
Barrow
N
ColorectalU50_Risk
0.18
0.36
0.54
0.72
0.9
Millom
Ulverston
Grange-over-Sands
Arnside
5 km

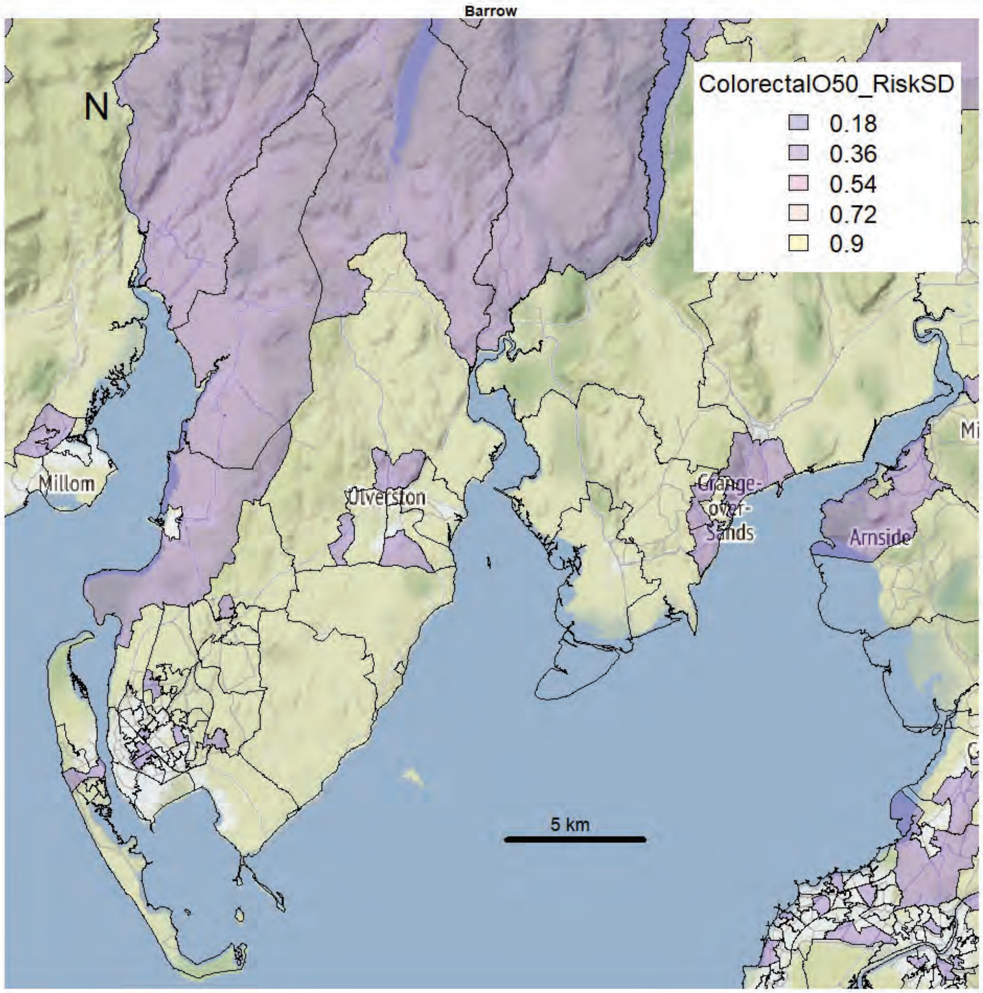
Barrow
N
ColorectalO50_RiskSD
0.18
0.36
0.54
0.72
0.9
Millom
Ulverston
Grange-over-Sands
Arnside
5 km

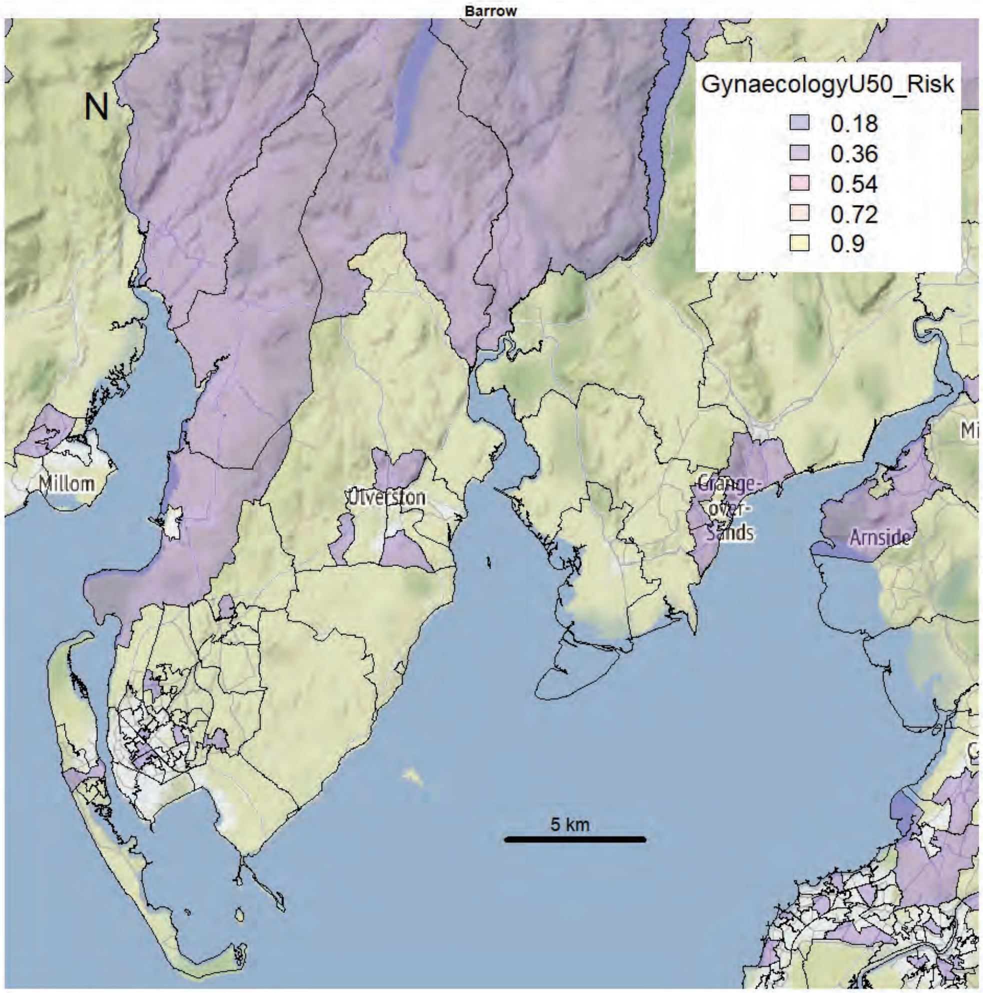
Barrow
N
GynaecologyU50_Risk
0.18
0.36
0.54
0.72
0.9
Millom
Ulverston
Grange-over-Sands
Arnside
5 km

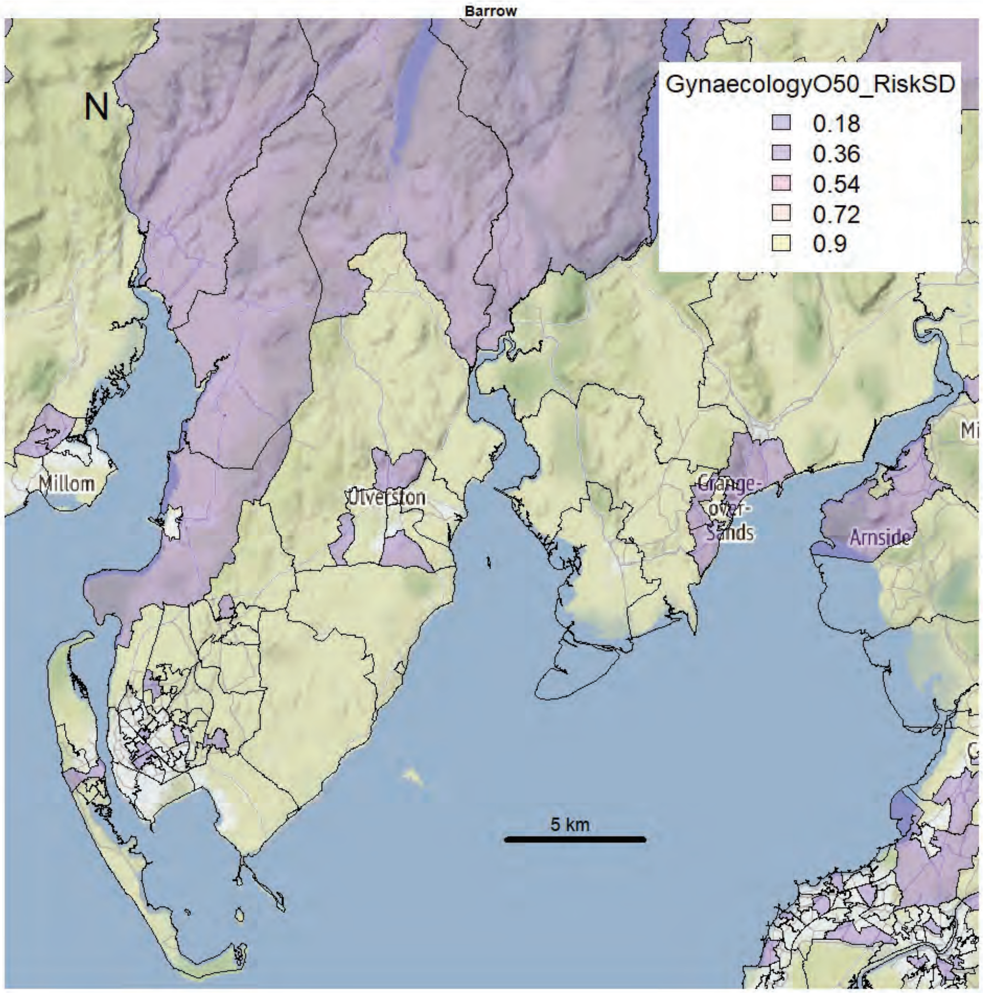
Barrow
N
GynaecologyO50_RiskSD
0.18
0.36
0.54
0.72
0.9
Millom
Ulverston
Grange-over-Sands
Arnside
5 km

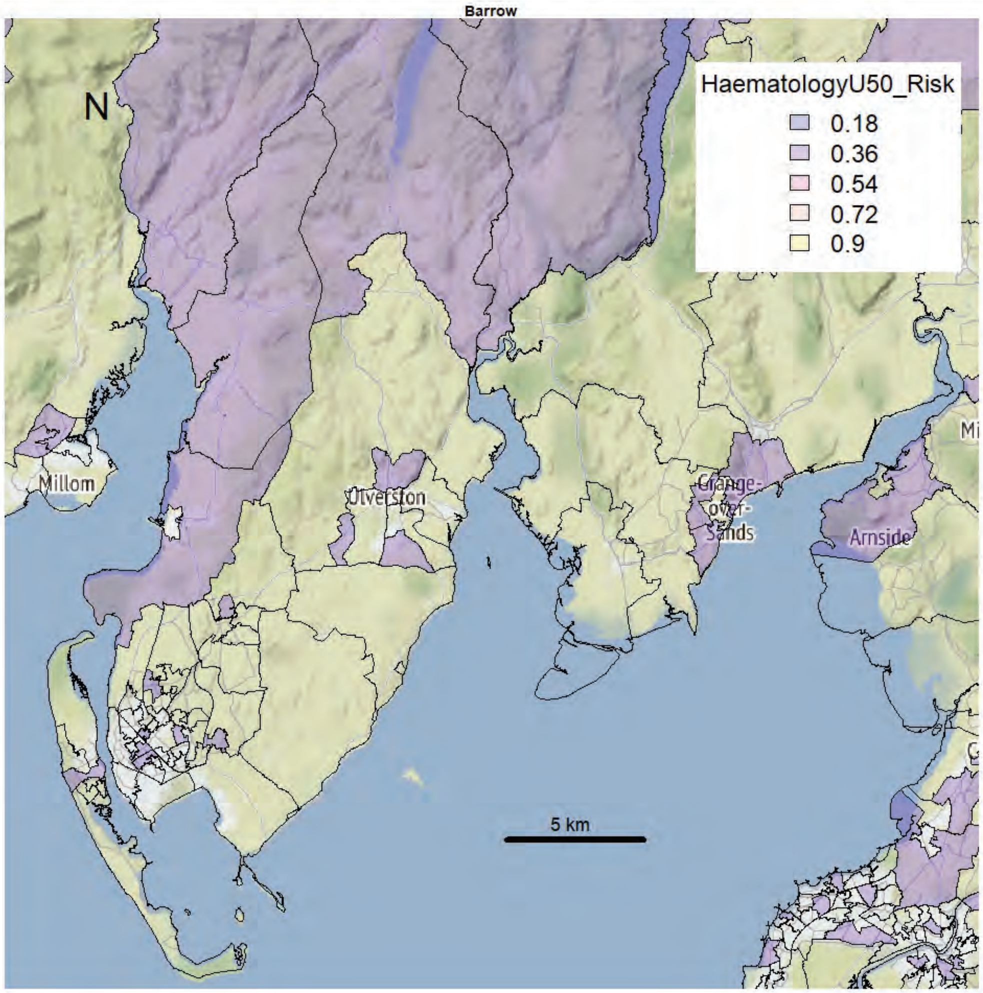
Barrow
N
HaematologyU50_Risk
0.18
0.36
0.54
0.72
0.9
Millom
Ulverston
Grange-over-Sands
Arnside
5 km

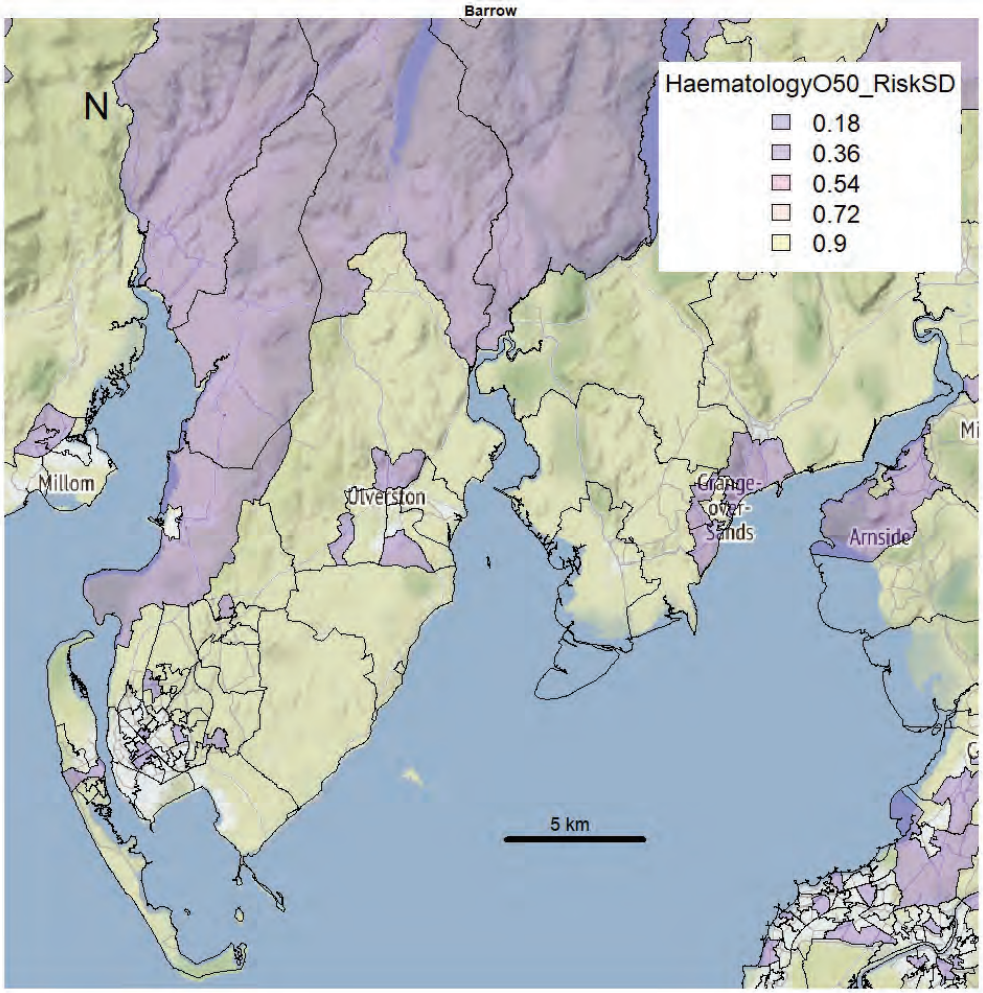
Barrow
N
HaematologyO50_RiskSD
0.18
0.36
0.54
0.72
0.9
Millom
Ulverston
Grange-over-Sands
Arnside
5 km

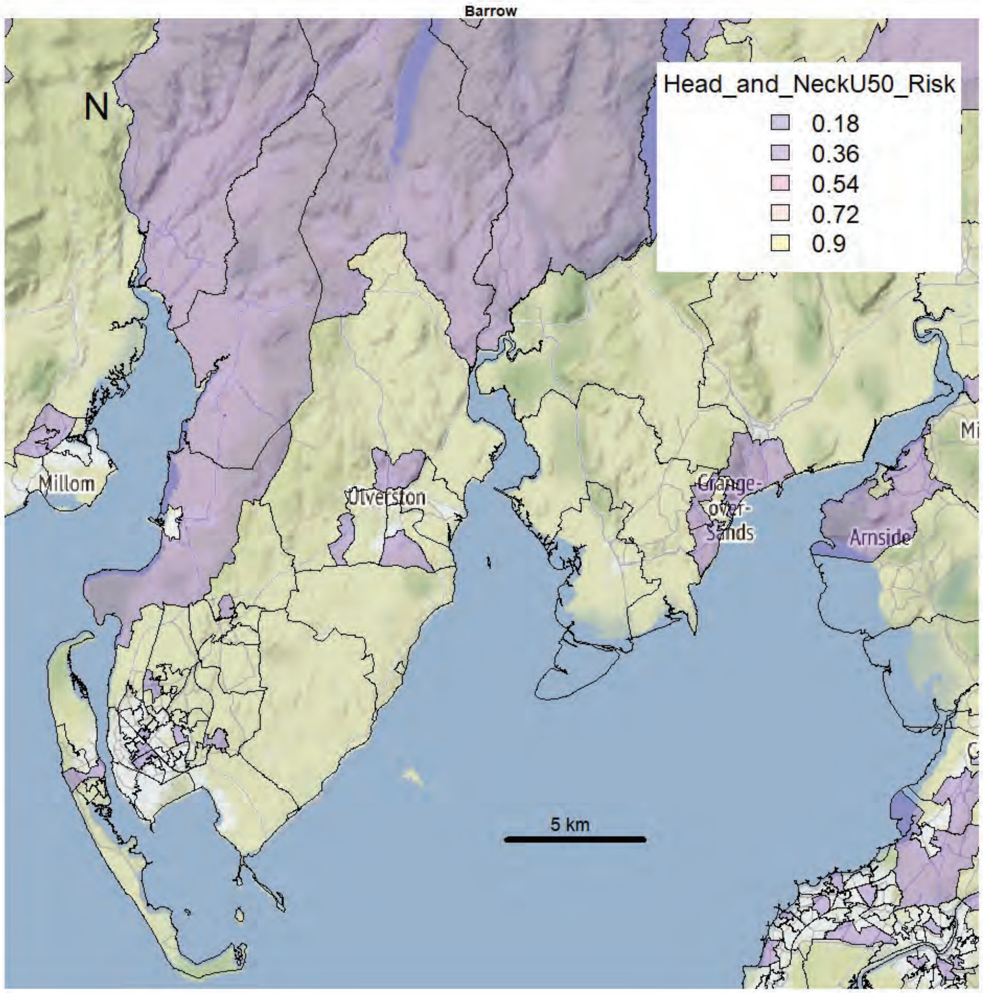
Barrow
N
Head_and_NeckU50_Risk
0.18
0.36
0.54
0.72
0.9
Millom
Ulverston
Grange-over-Sands
Arnside
5 km

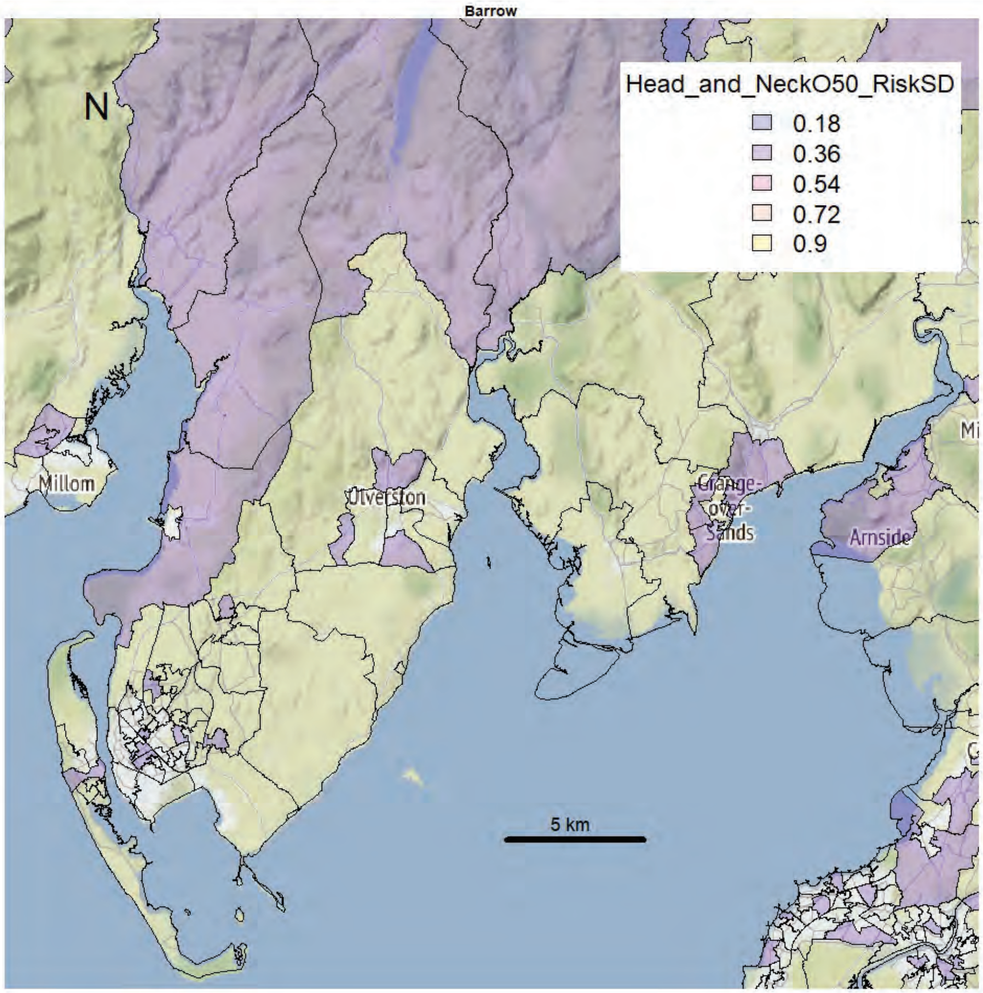
Barrow
N
Head_and_NeckO50_RiskSD
0.18
0.36
0.54
0.72
0.9
Millom
Ulverston
Grange-over-Sands
Arnside
5 km

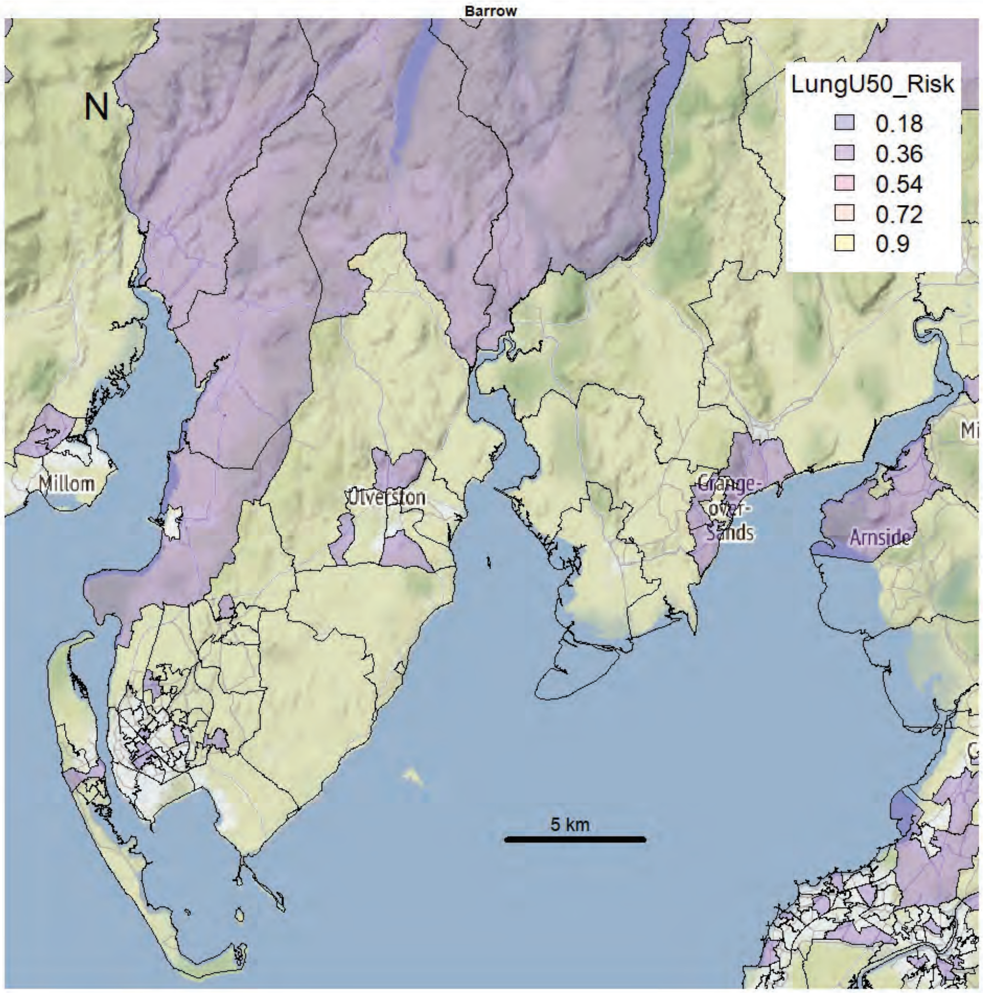
Barrow
N
LungU50_Risk
0.18
0.36
0.54
0.72
0.9
Millom
Ulverston
Grange-over-Sands
Arnside
5 km

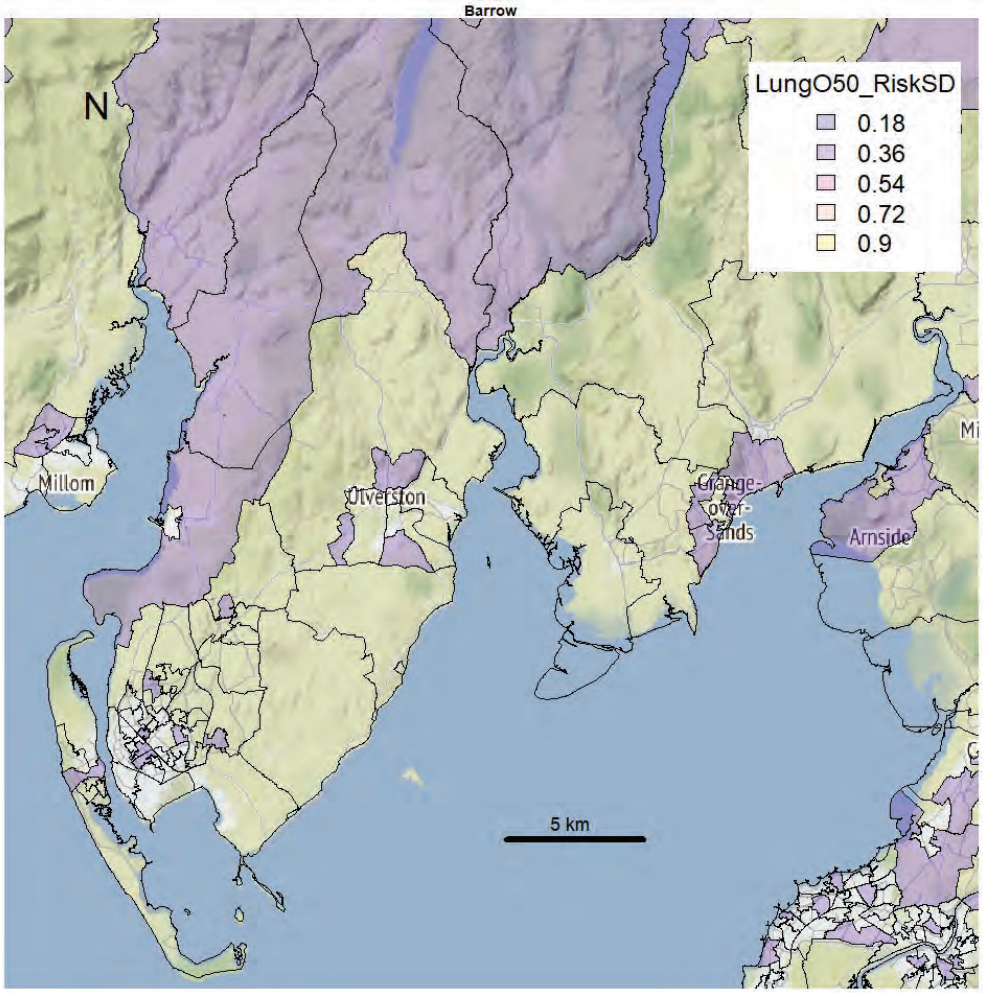
Barrow
N
LungO50_RiskSD
0.18
0.36
0.54
0.72
0.9
Millom
Ulverston
Grange-over-Sands
Arnside
5 km

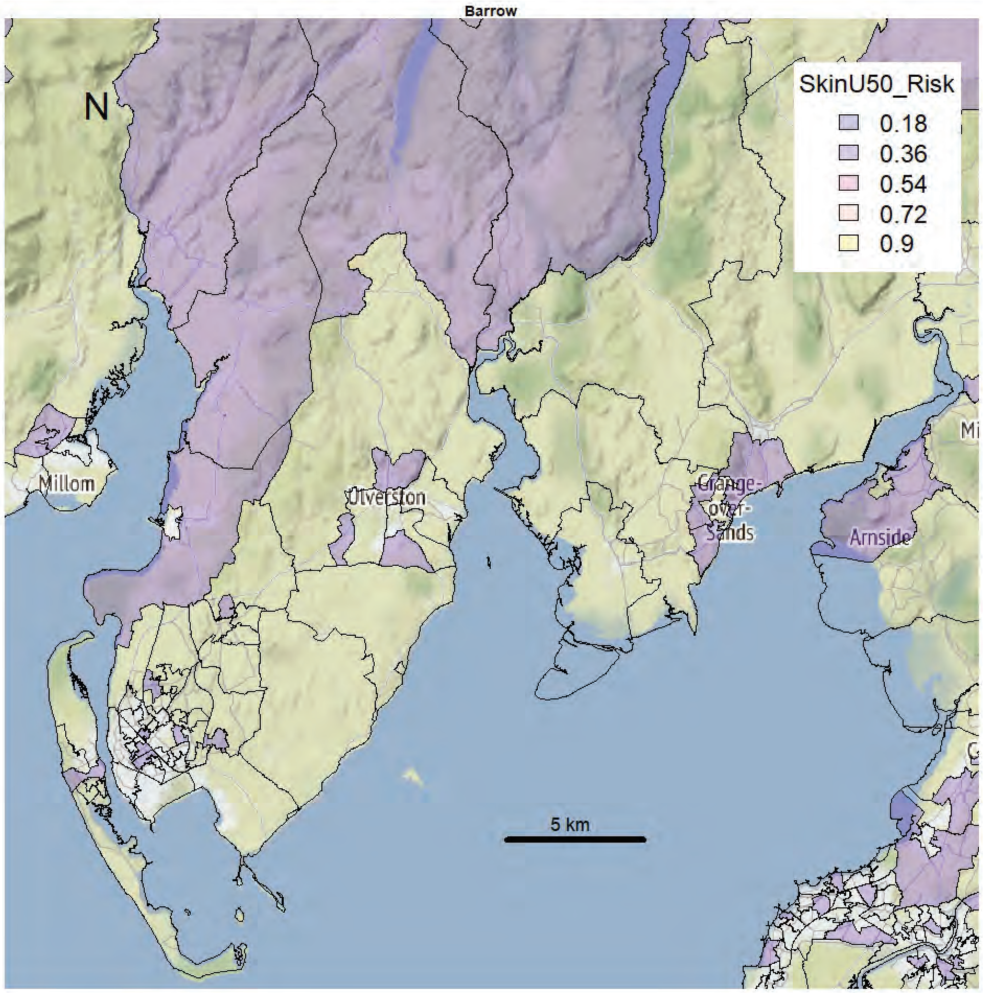
Barrow
N
SkinU50_Risk
0.18
0.36
0.54
0.72
0.9
Millom
Ulverston
Grange-over-Sands
Arnside
5 km

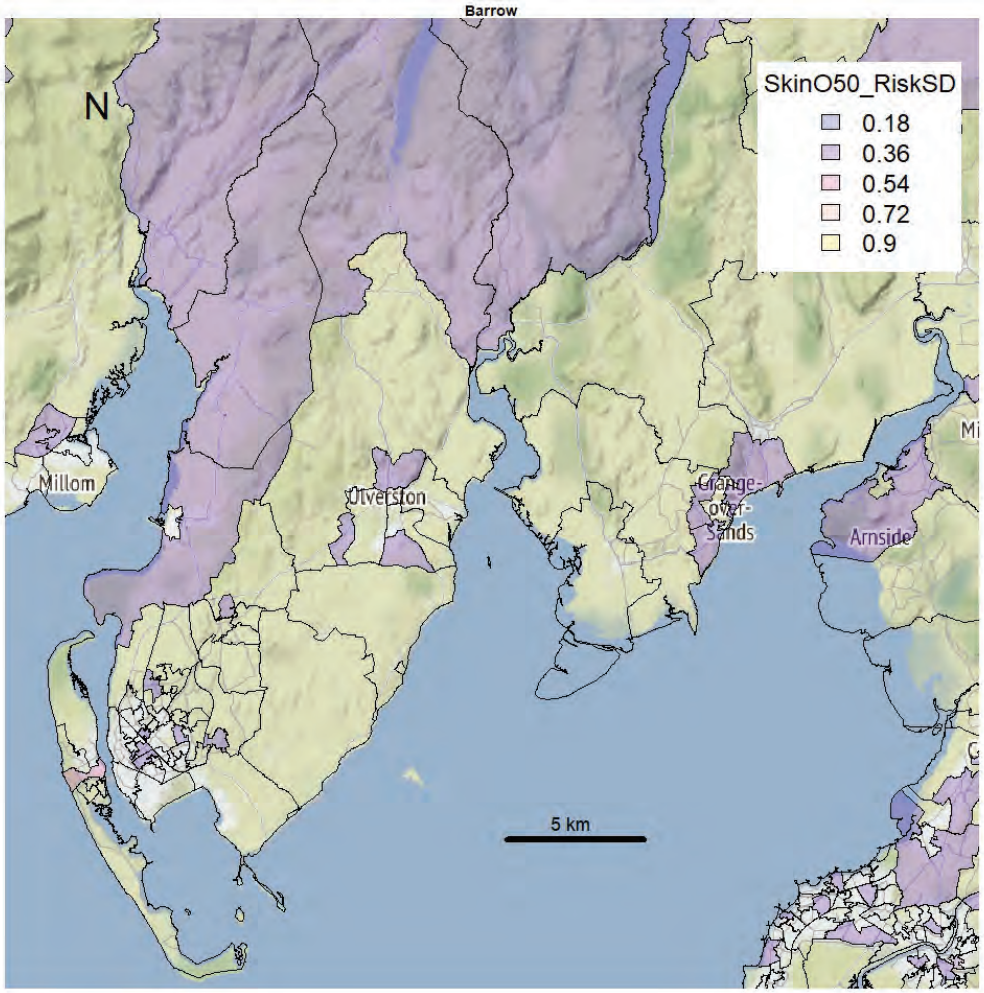
Barrow
N
SkinO50_RiskSD
0.18
0.36
0.54
0.72
0.9
Millom
Ulverston
Grange-over-Sands
Arnside
5 km

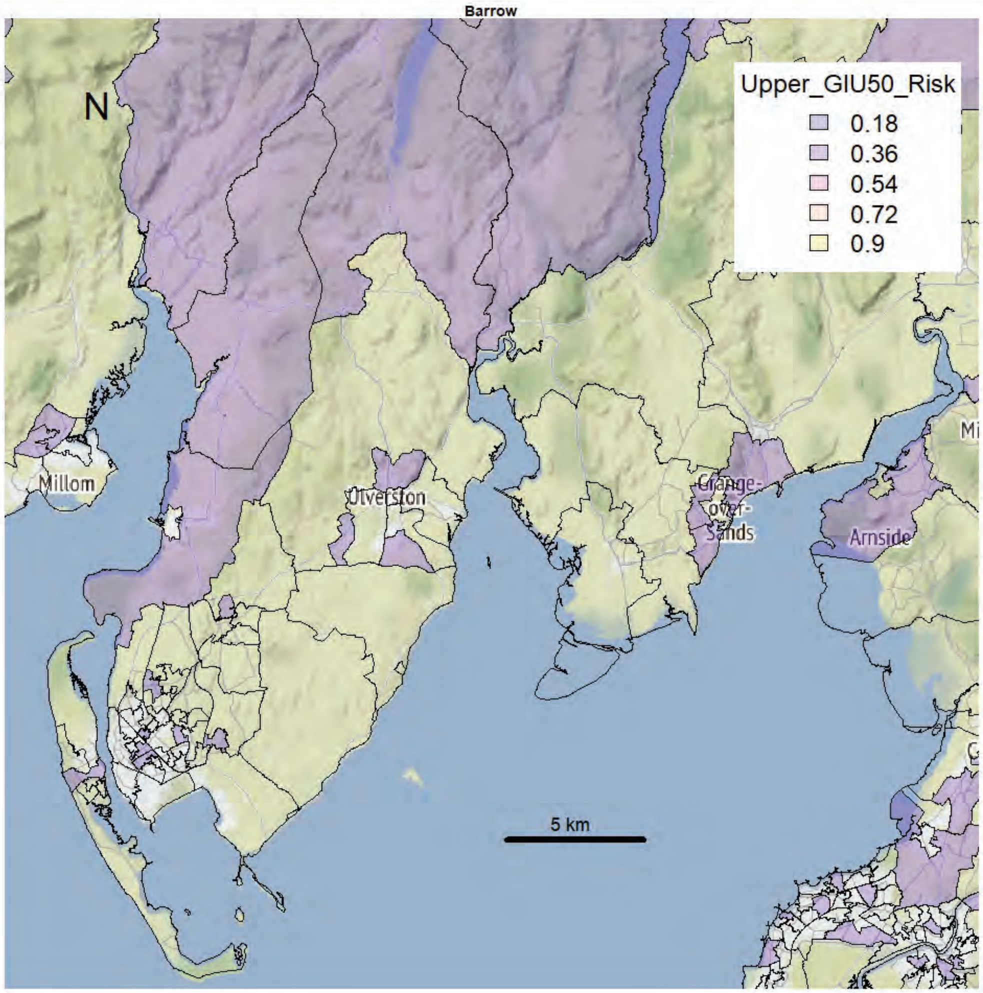
Barrow
N
Upper_GIU50_Risk
0.18
0.36
0.54
0.72
0.9
Millom
Ulverston
Grange-over-Sands
Arnside
5 km

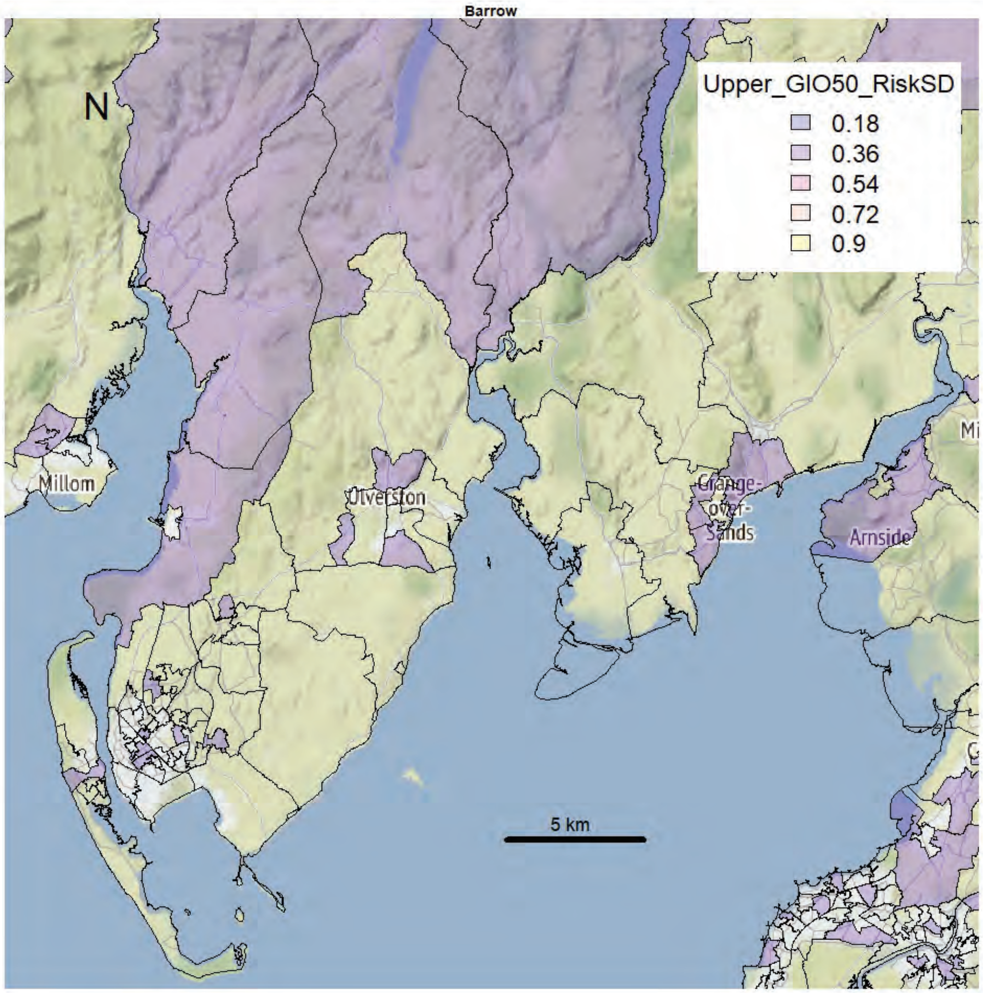
Barrow
N
Upper_GIO50_RiskSD
0.18
0.36
0.54
0.72
0.9
Millom
Ulverston
Grange-over-Sands
Arnside
5 km

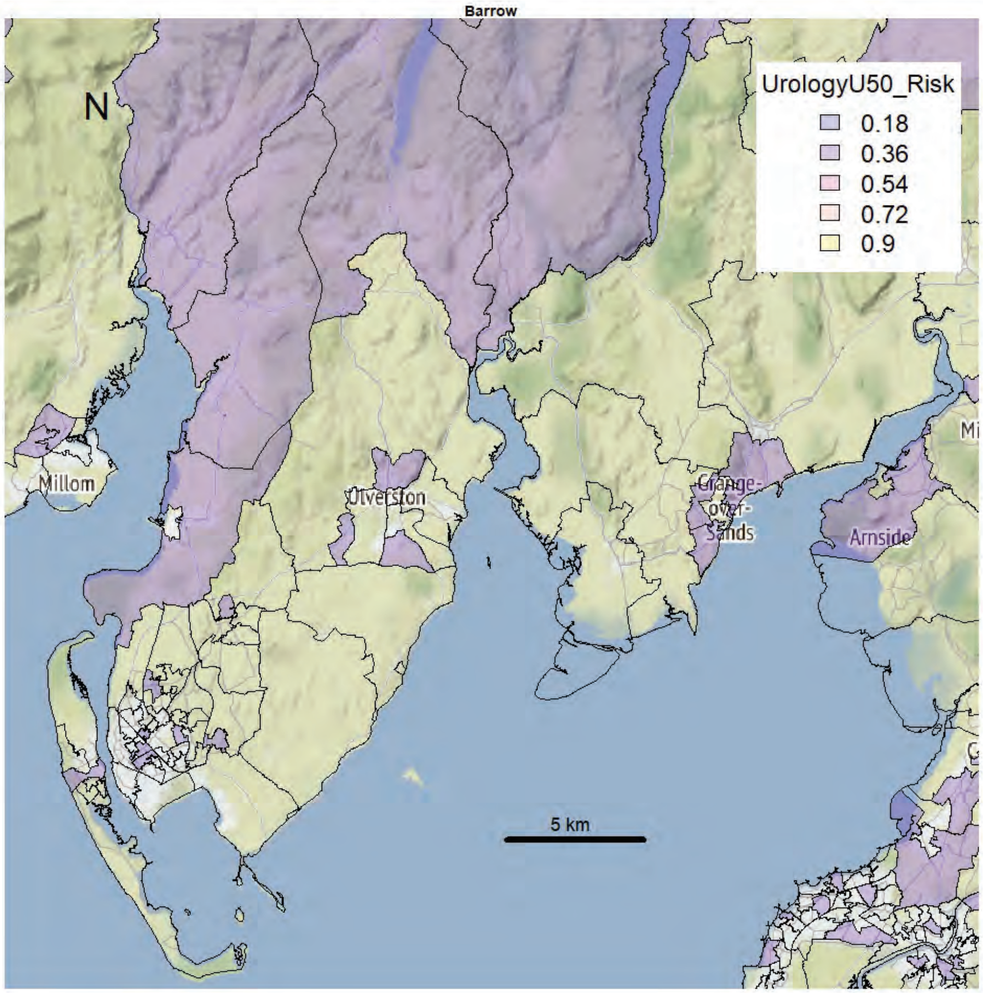
Barrow
N
UrologyU50_Risk
0.18
0.36
0.54
0.72
0.9
Millom
Ulverston
Grange-over-Sands
Arnside
5 km

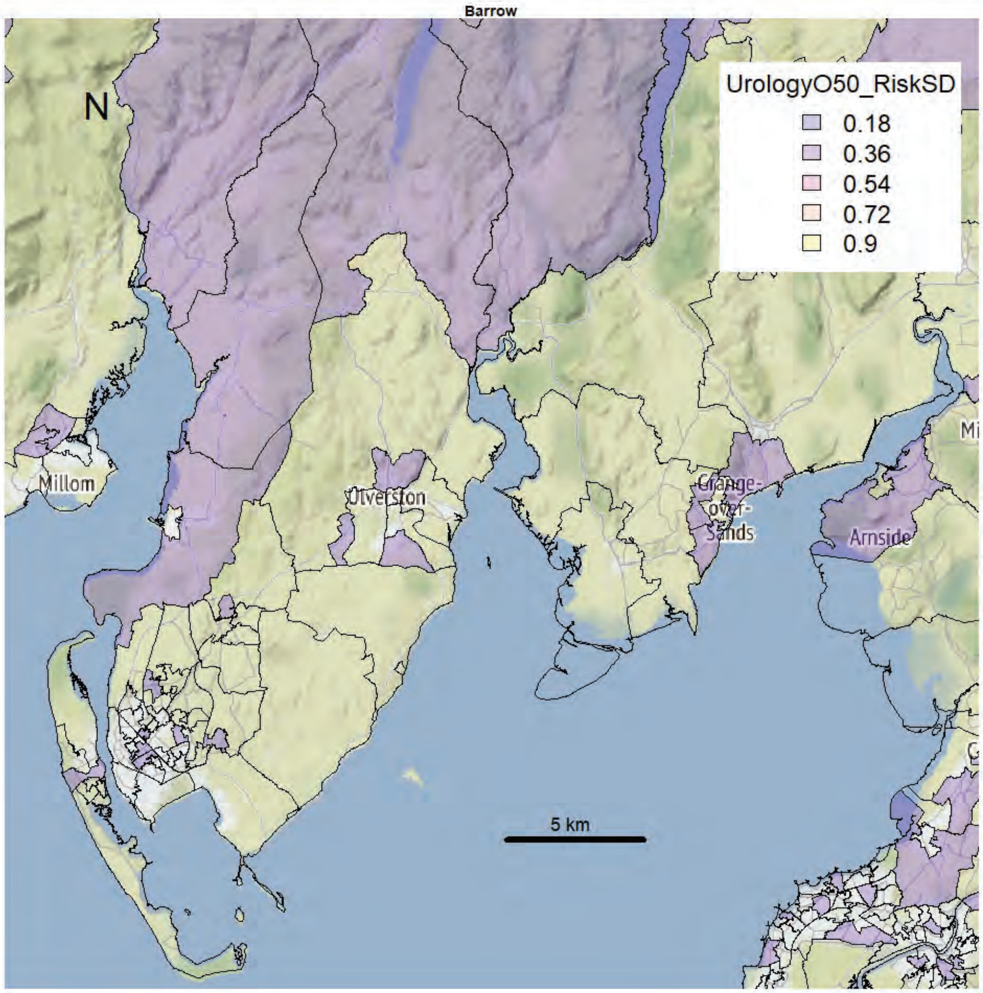
Barrow
N
UrologyO50_RiskSD
0.18
0.36
0.54
0.72
0.9
Millom
Ulverston
Grange-over-Sands
Arnside
5 km

Kendal area

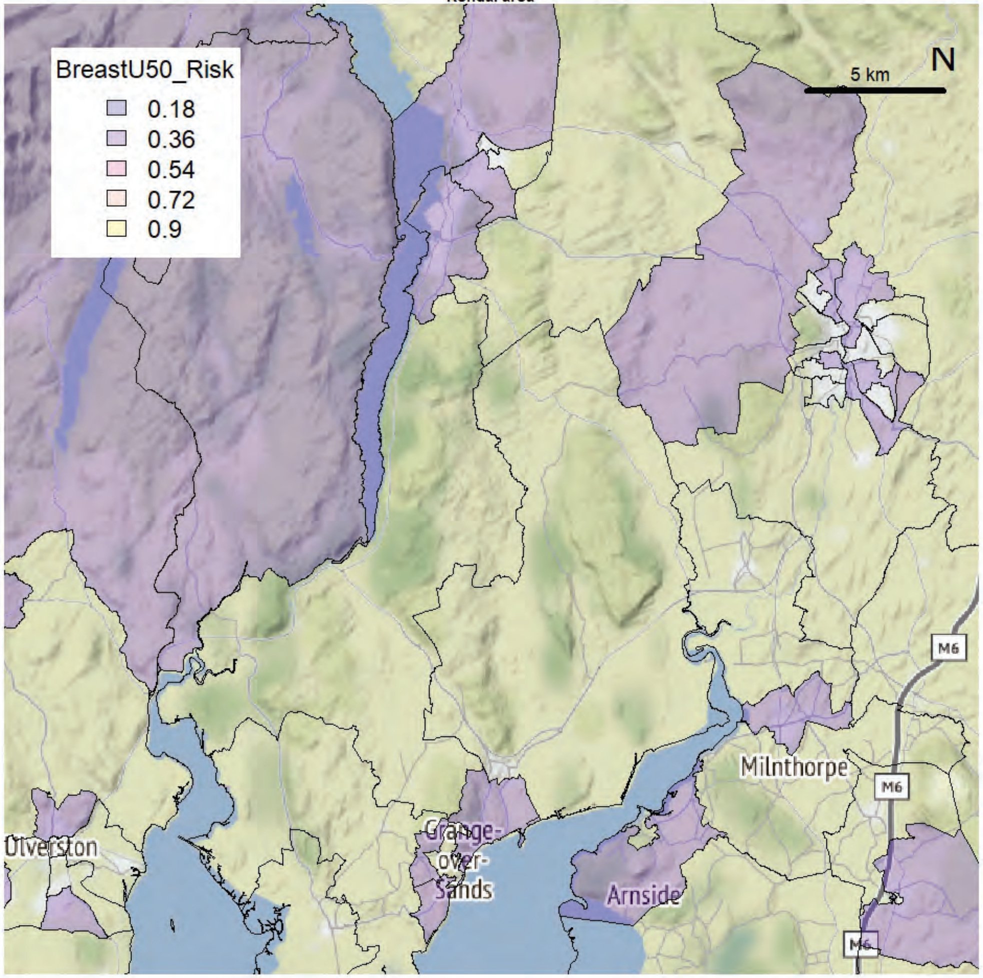


Kendal area

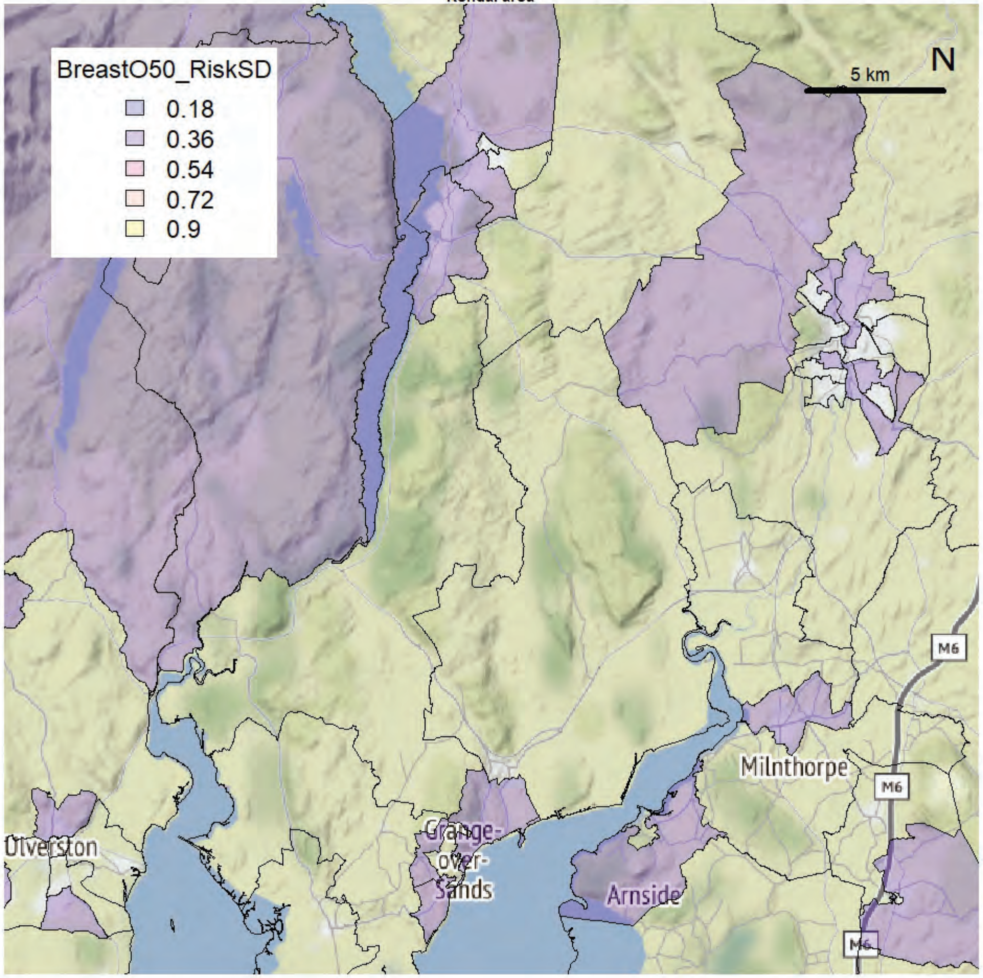

Kendal area

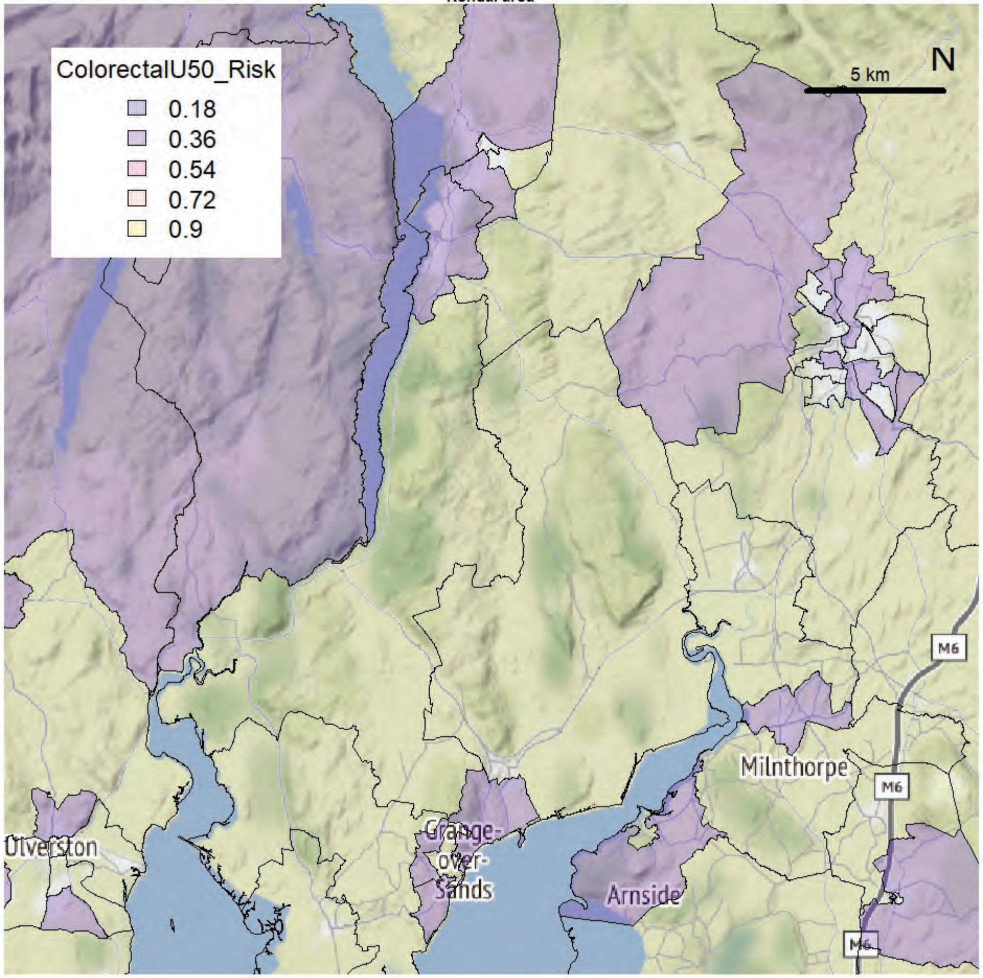


Kendal area

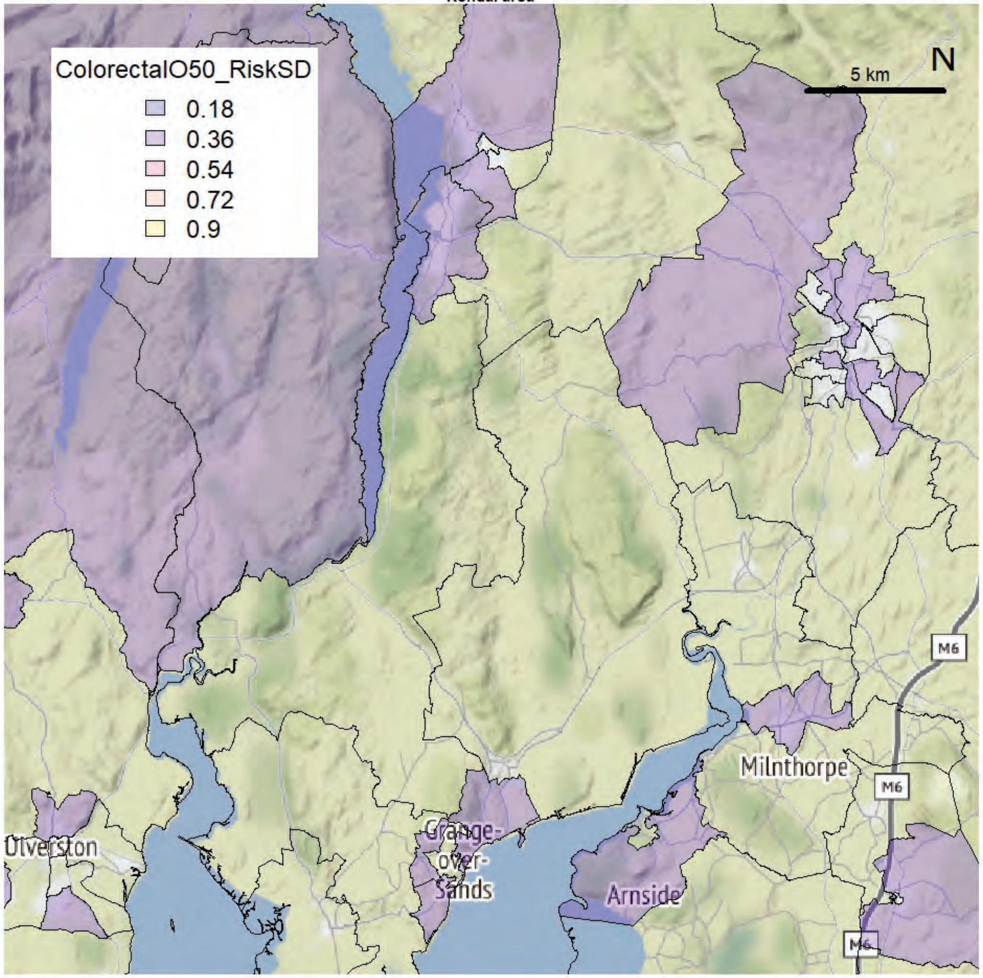

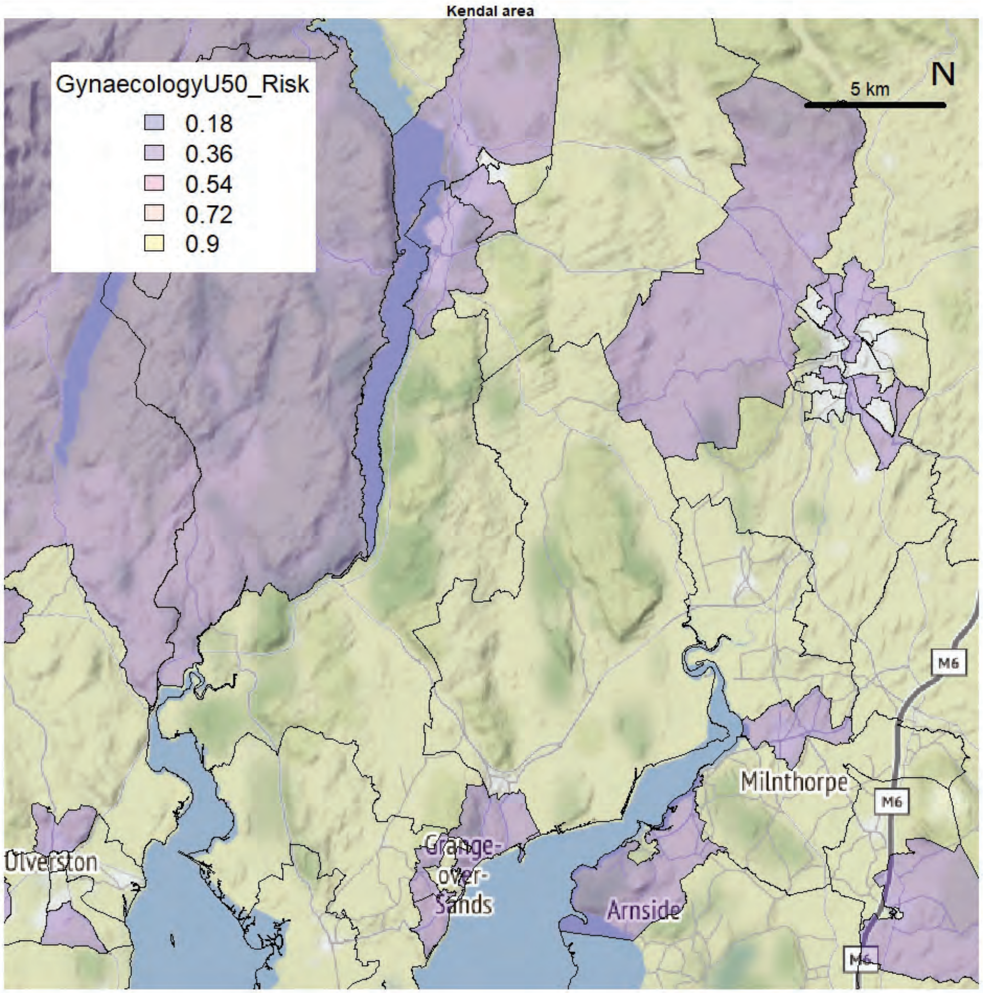
Kendal area
GynaecologyU50_Risk
0.18
0.36
0.54
0.72
0.9
5 km
N
Ulverston
Grange-over-Sands
Arnside
Milnthorpe
M6

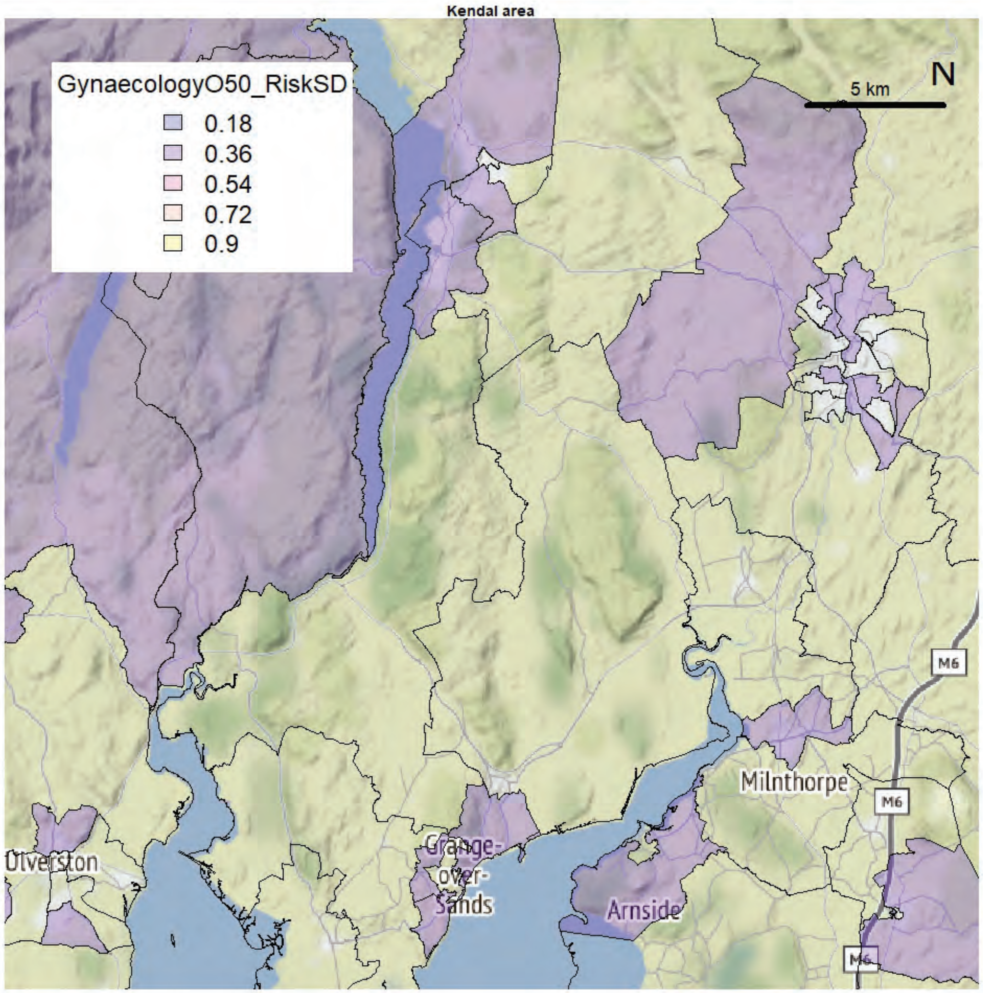
Kendal area
GynaecologyO50_RiskSD
0.18
0.36
0.54
0.72
0.9
5 km
N
Ulverston
Grange-over-Sands
Arnside
Milnthorpe
M6

Kendal area

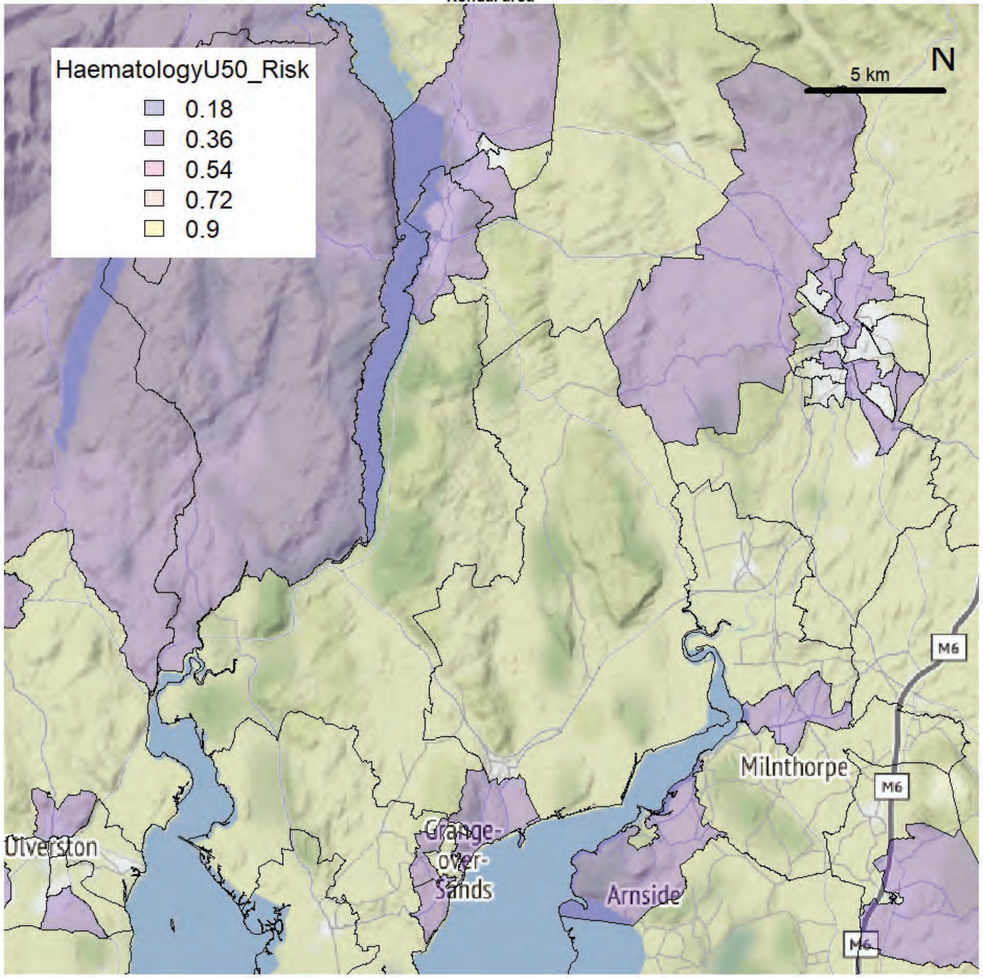


Kendal area

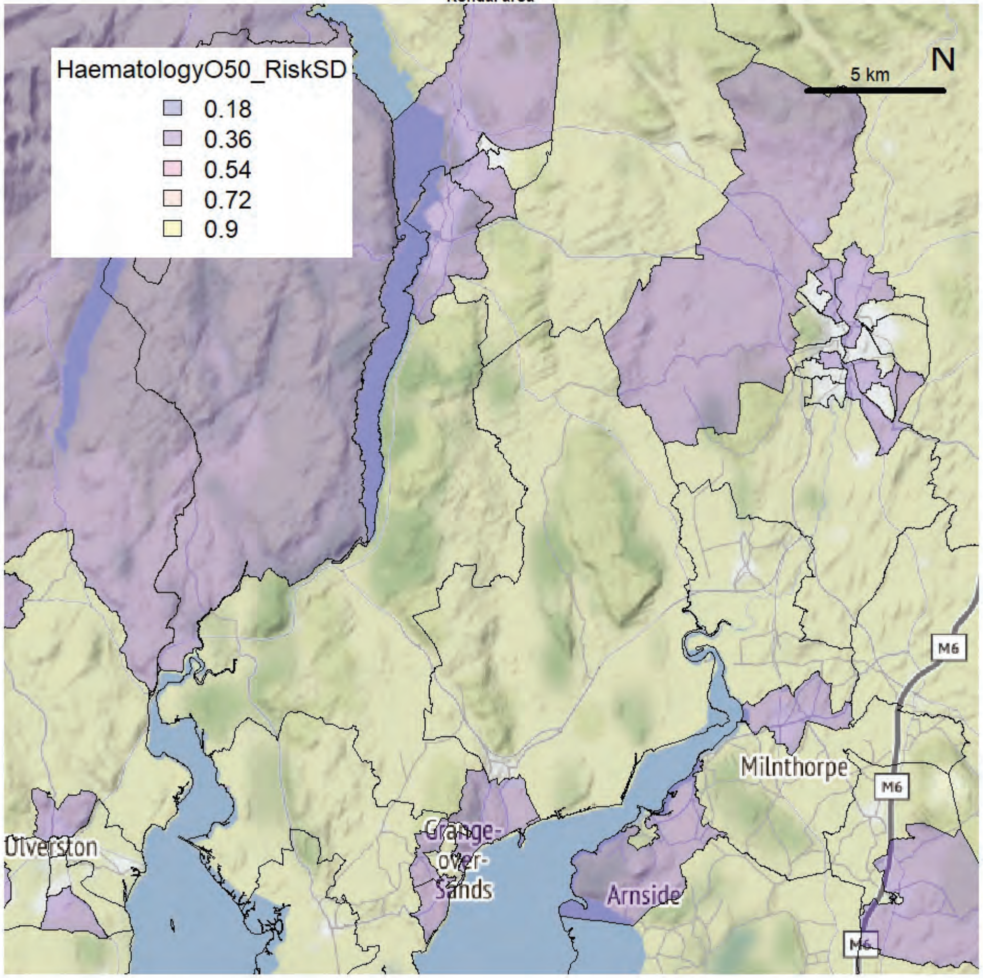

Kendal area

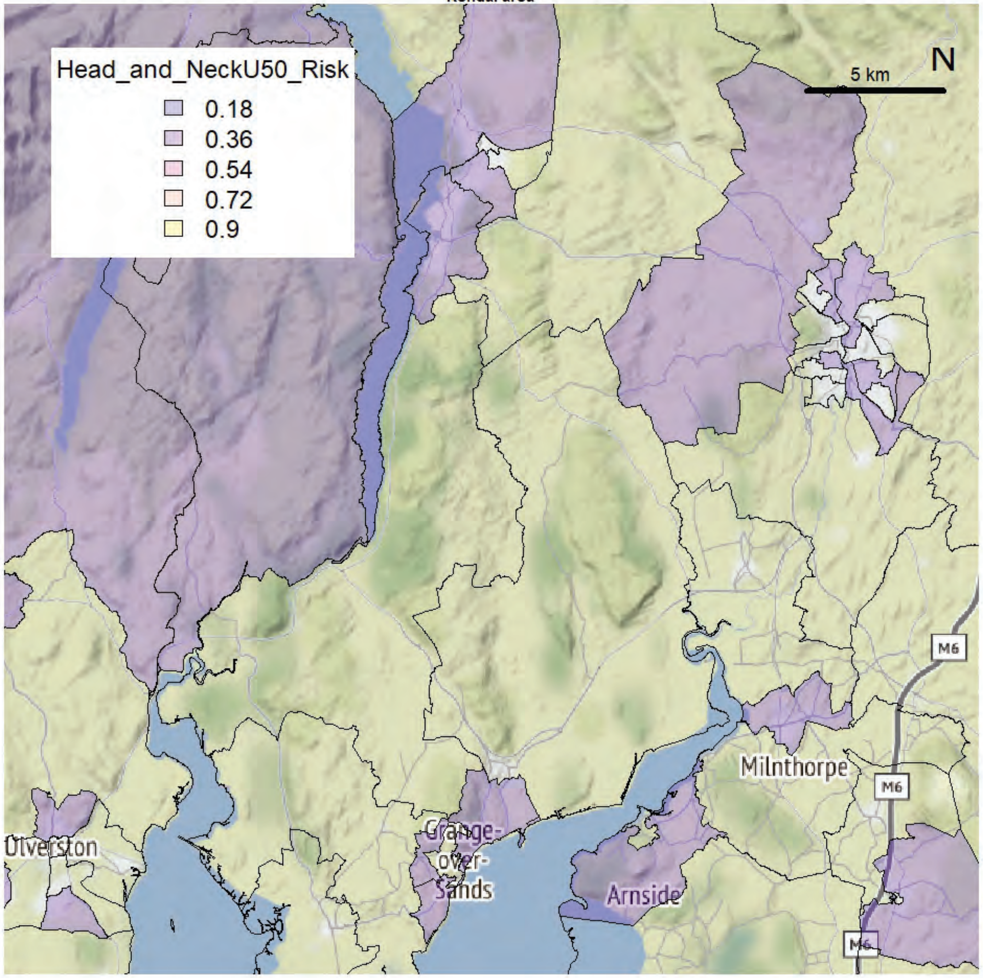


Kendal area

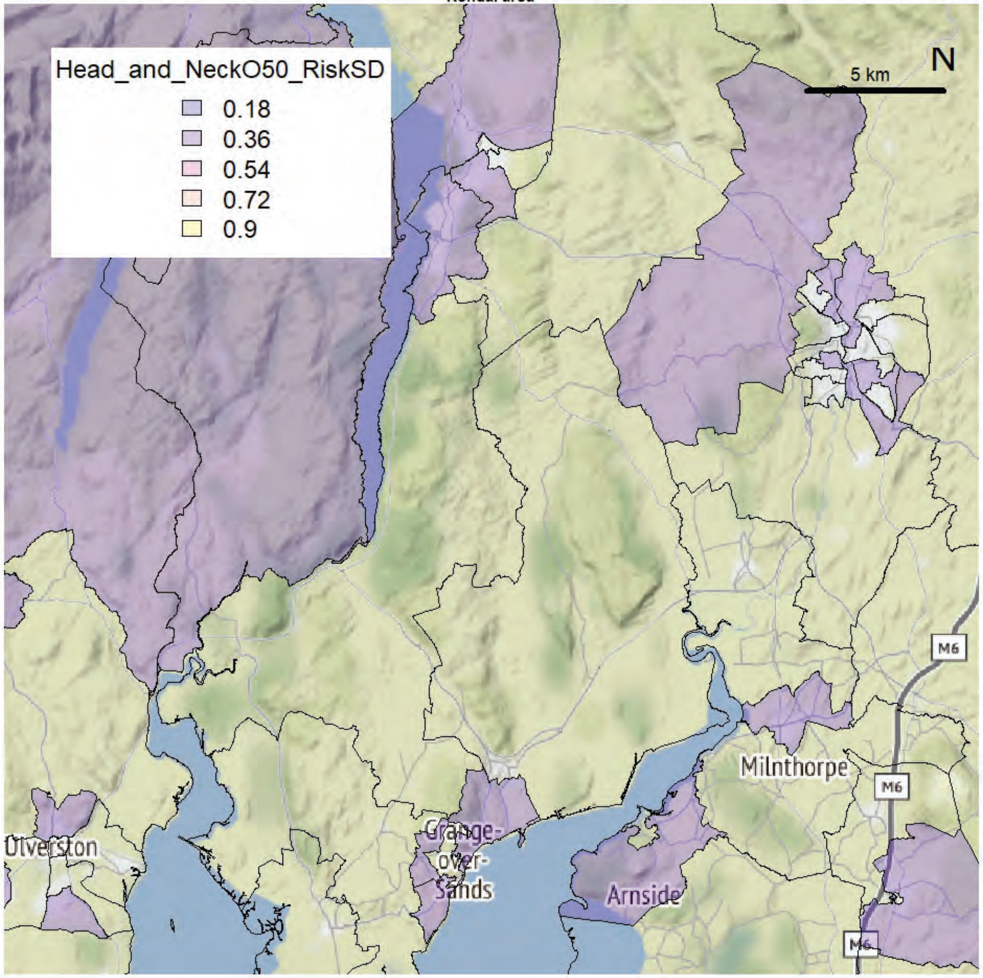

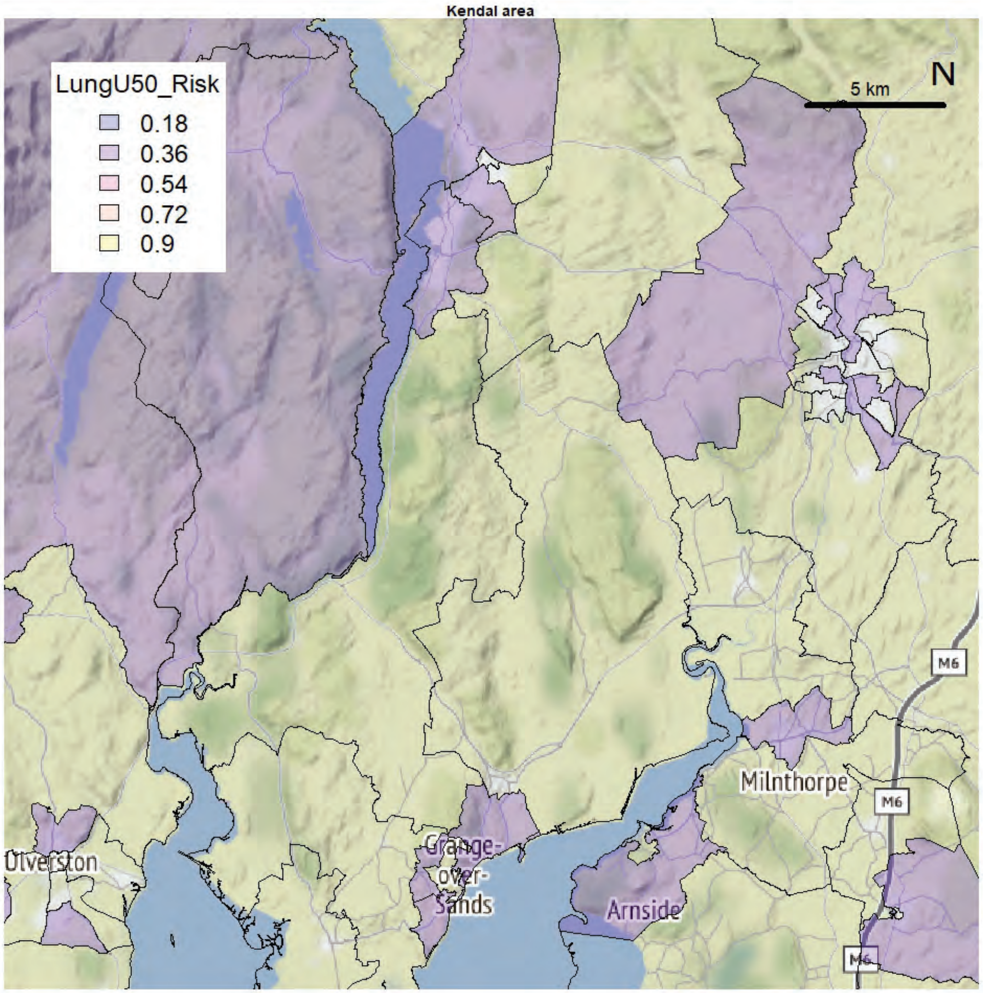
Kendal area
LungU50_Risk
0.18
0.36
0.54
0.72
0.9
5 km
N
M6
Milnthorpe
Ulverston
Grange-over-Sands
Arnside

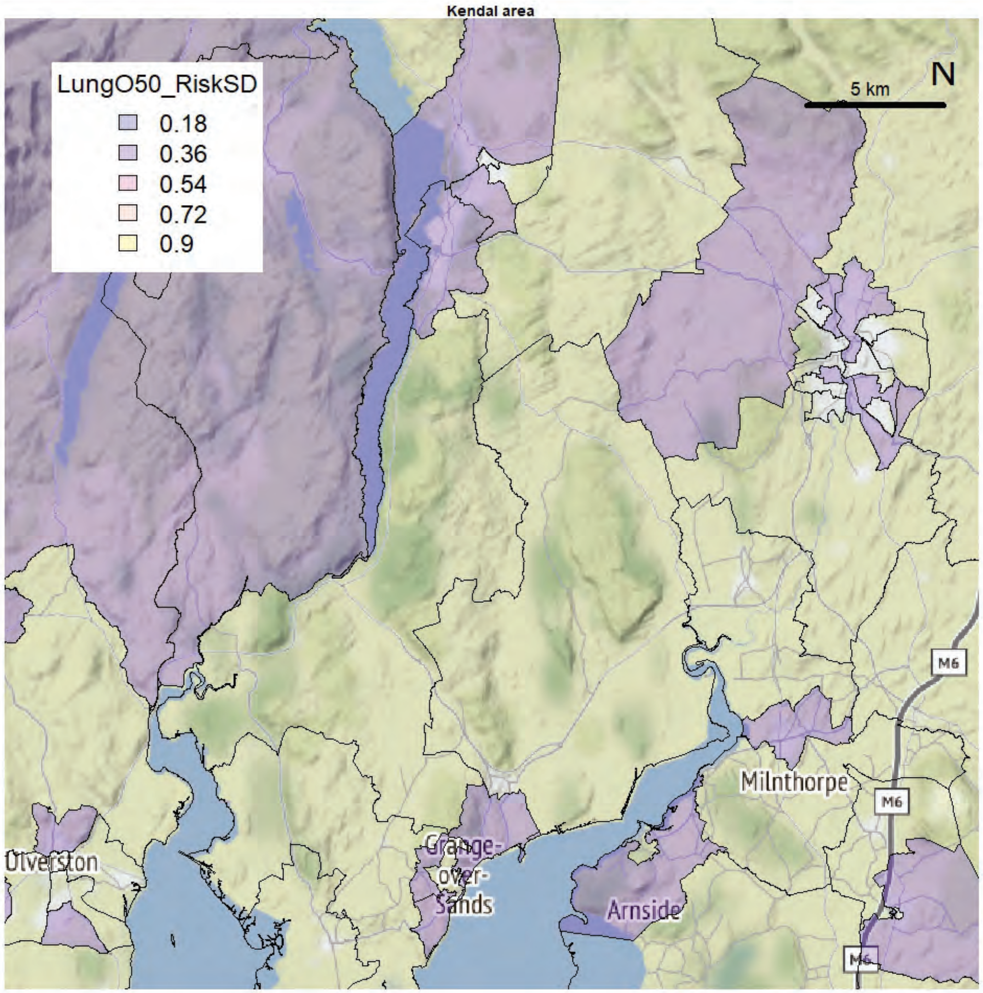
Kendal area
LungO50_RiskSD
0.18
0.36
0.54
0.72
0.9
5 km
N
M6
Milnthorpe
Ulverston
Grange-over-Sands
Arnside

Kendal area

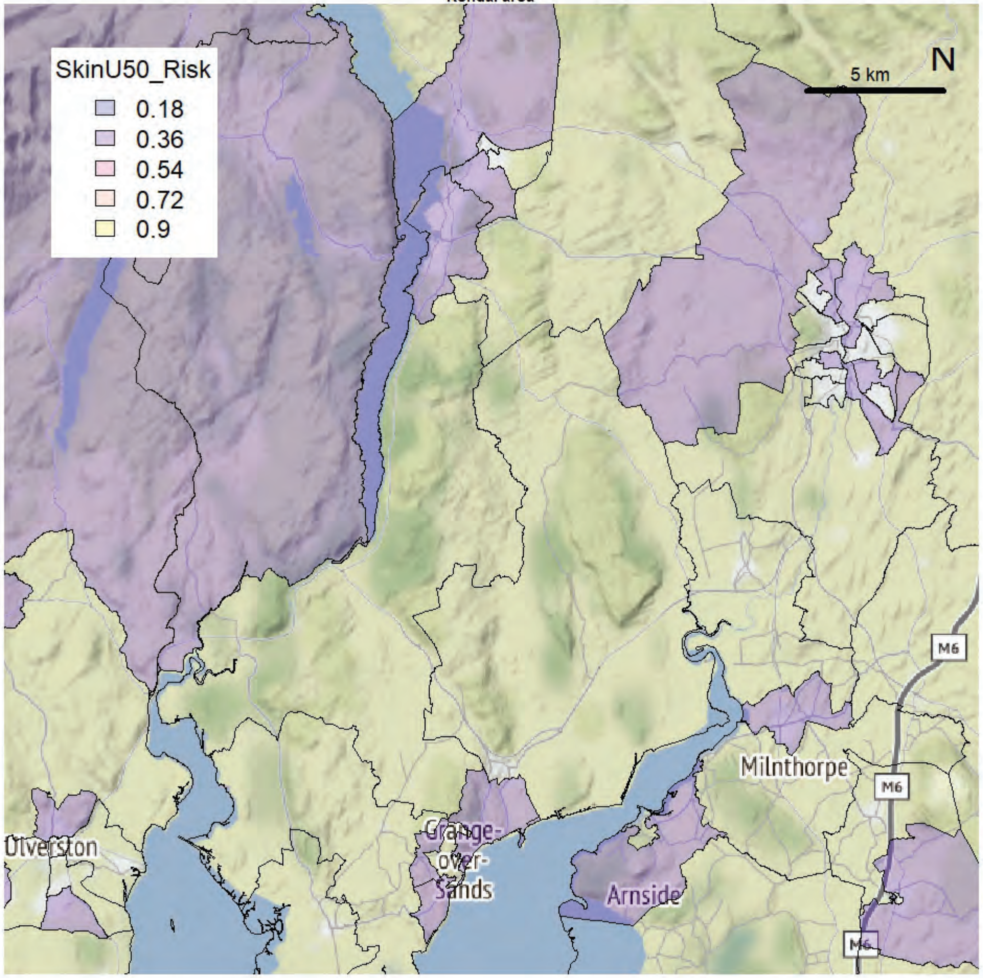


Kendal area

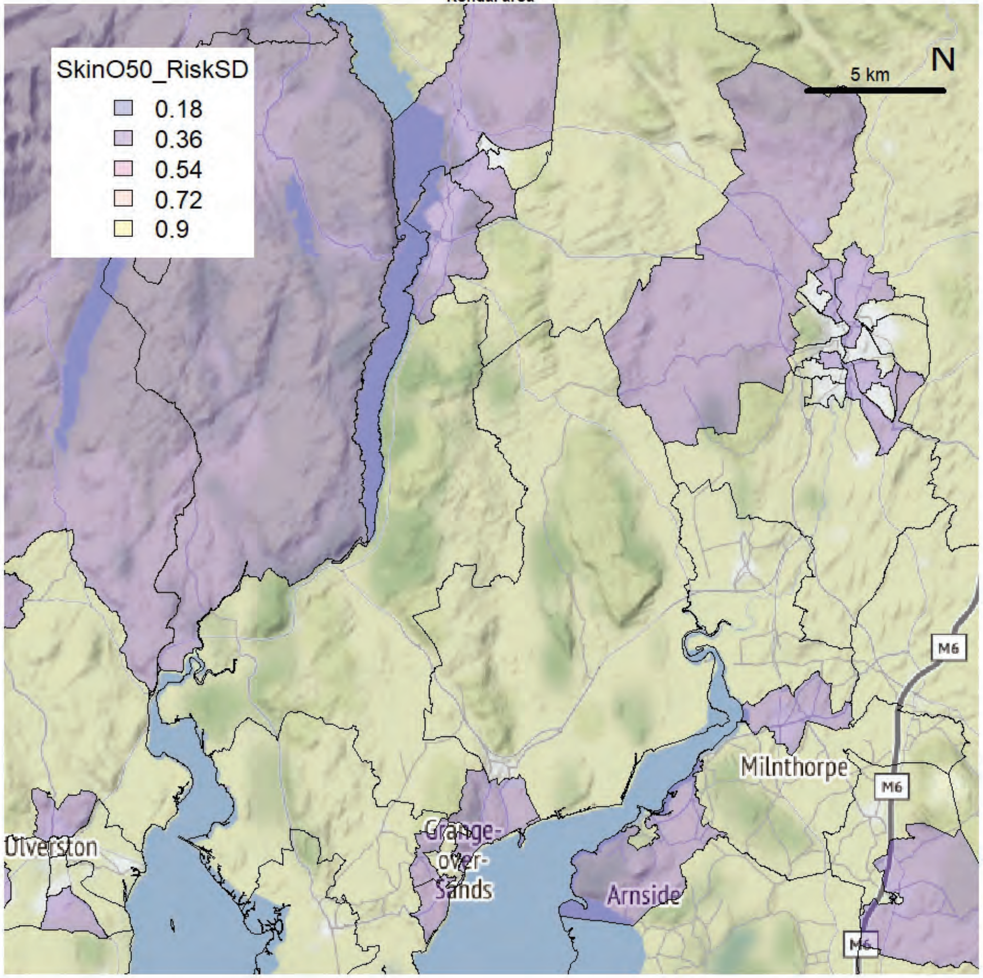

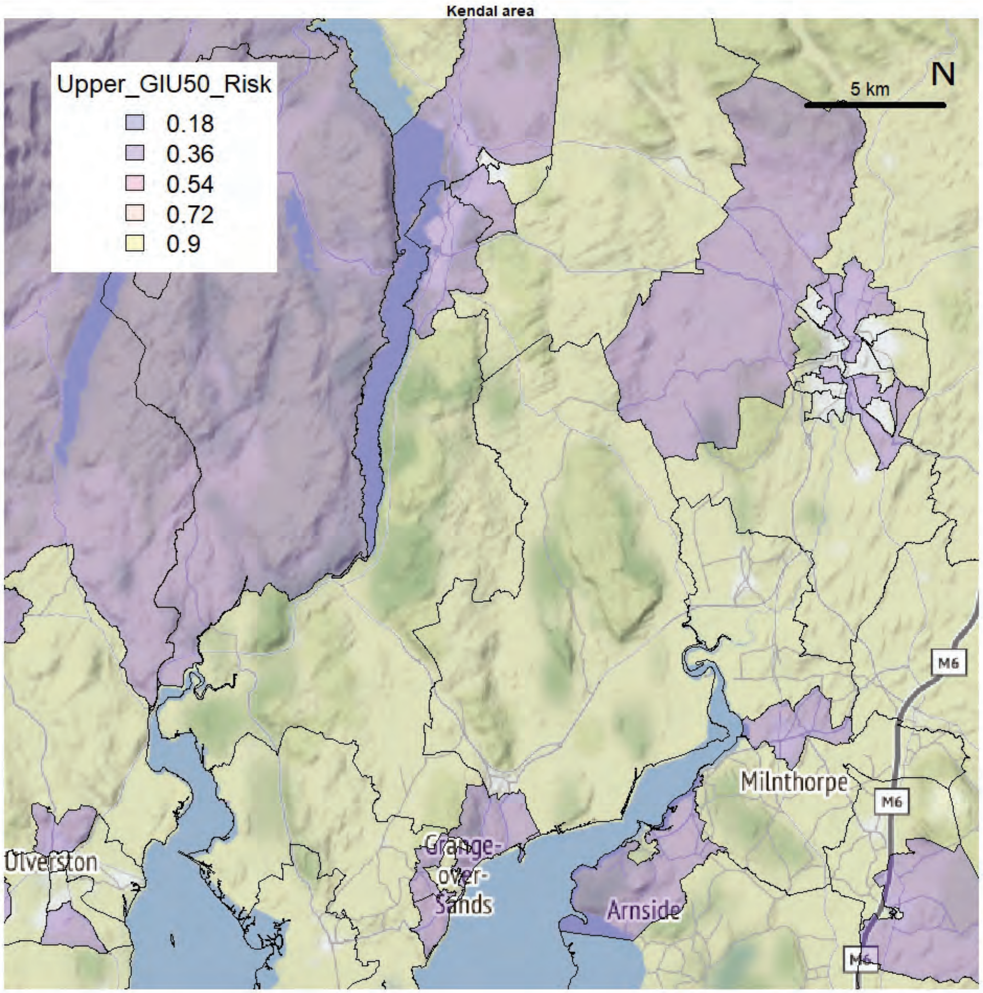
Kendal area
Upper_GIU50_Risk
0.18
0.36
0.54
0.72
0.9
5 km
N
Ulverston
Grange-over-Sands
Arnside
Milnthorpe
M6

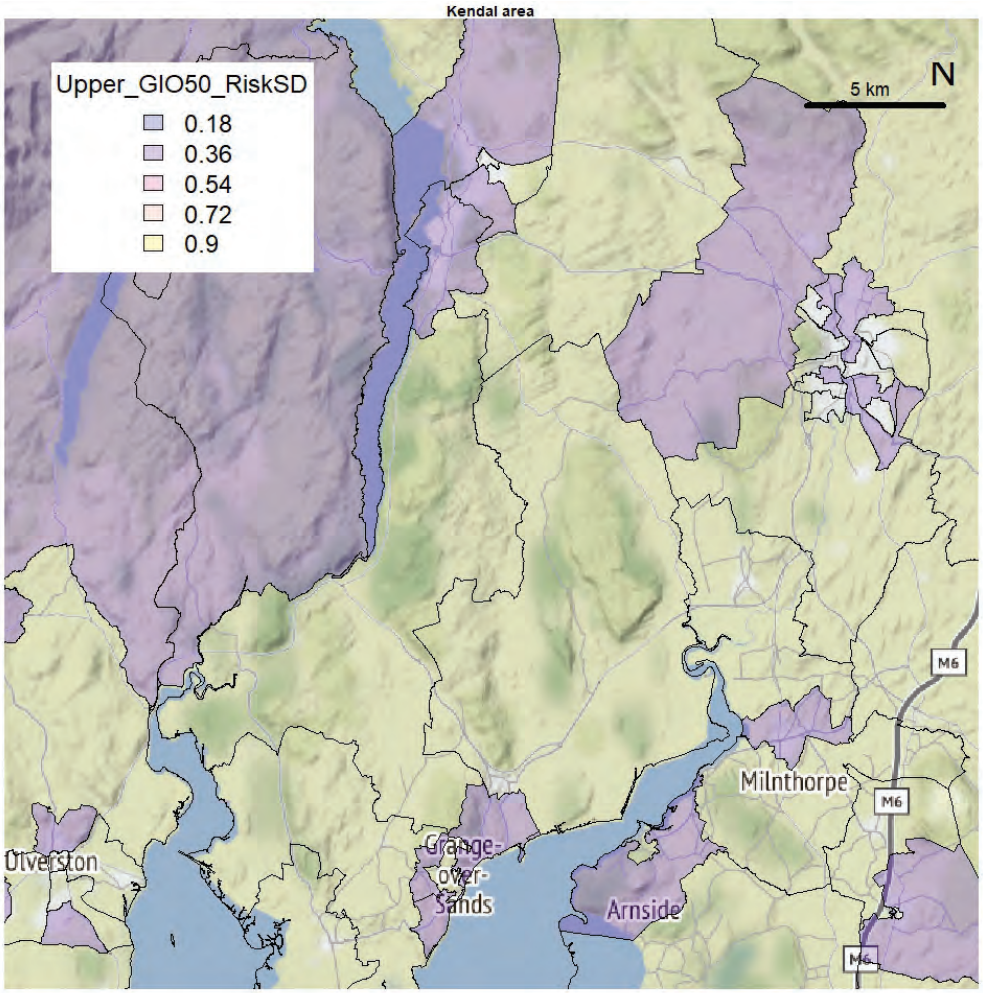
Kendal area
Upper_GIO50_RiskSD
0.18
0.36
0.54
0.72
0.9
5 km
N
Ulverston
Grange-over-Sands
Arnside
Milnthorpe
M6

Kendal area

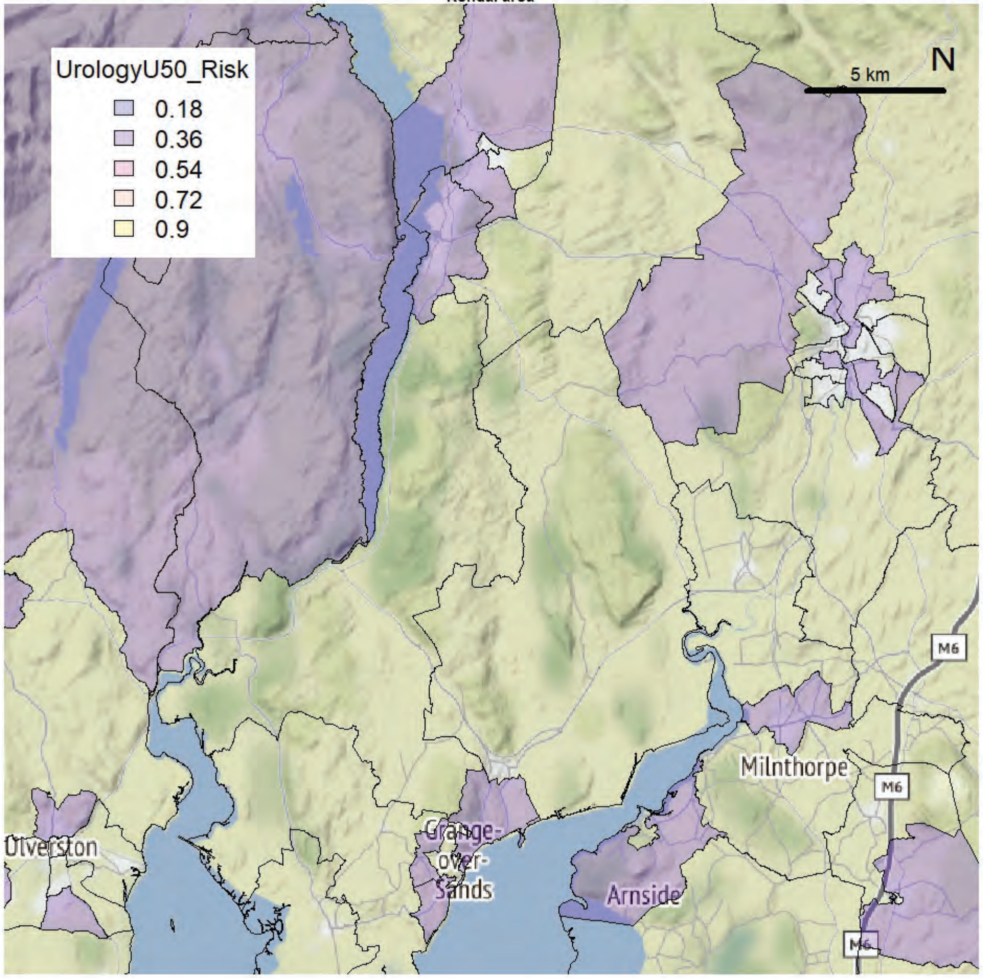


Kendal area

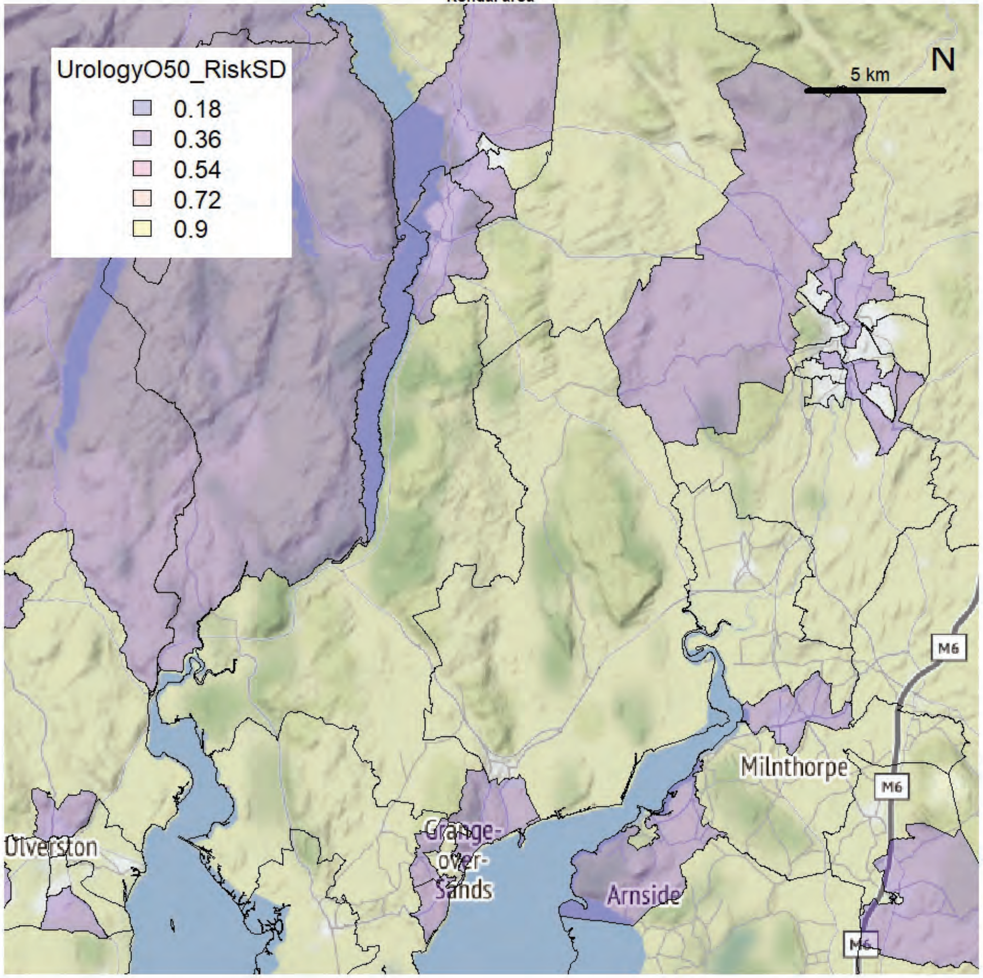

Morecambe Bay exCCG extended

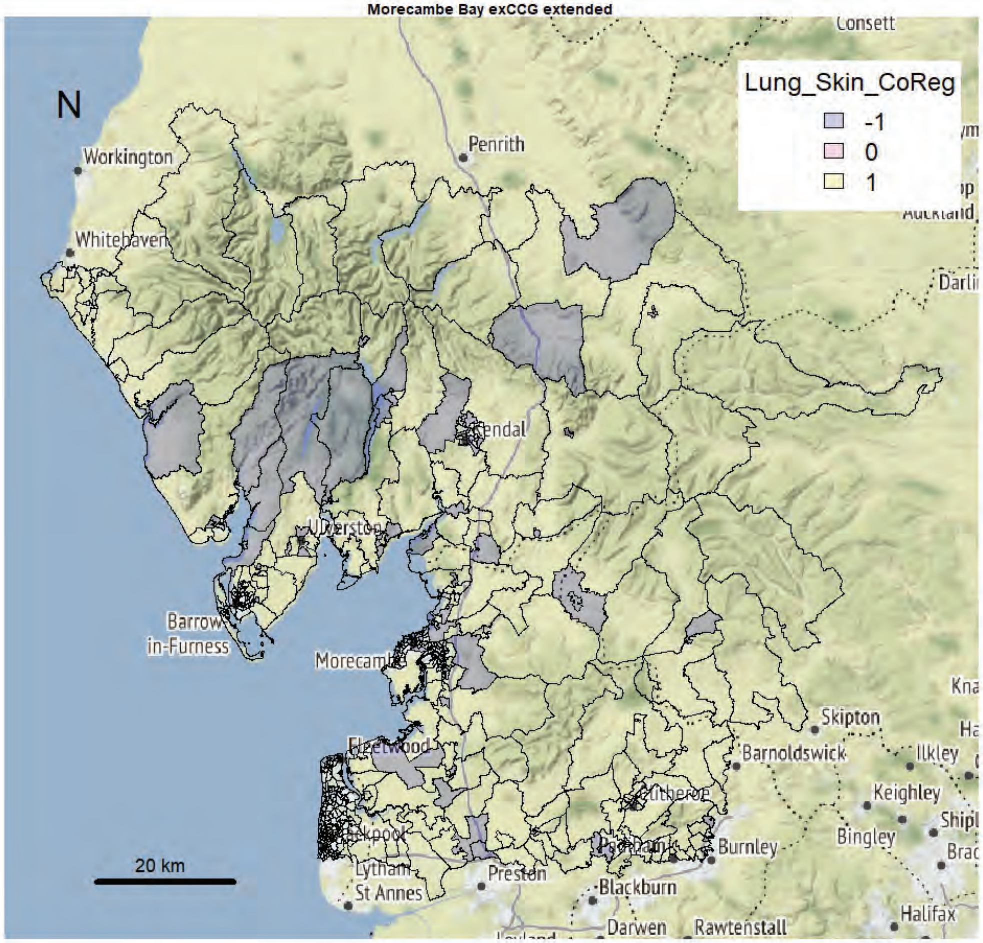


Morecambe Bay exCCG extended

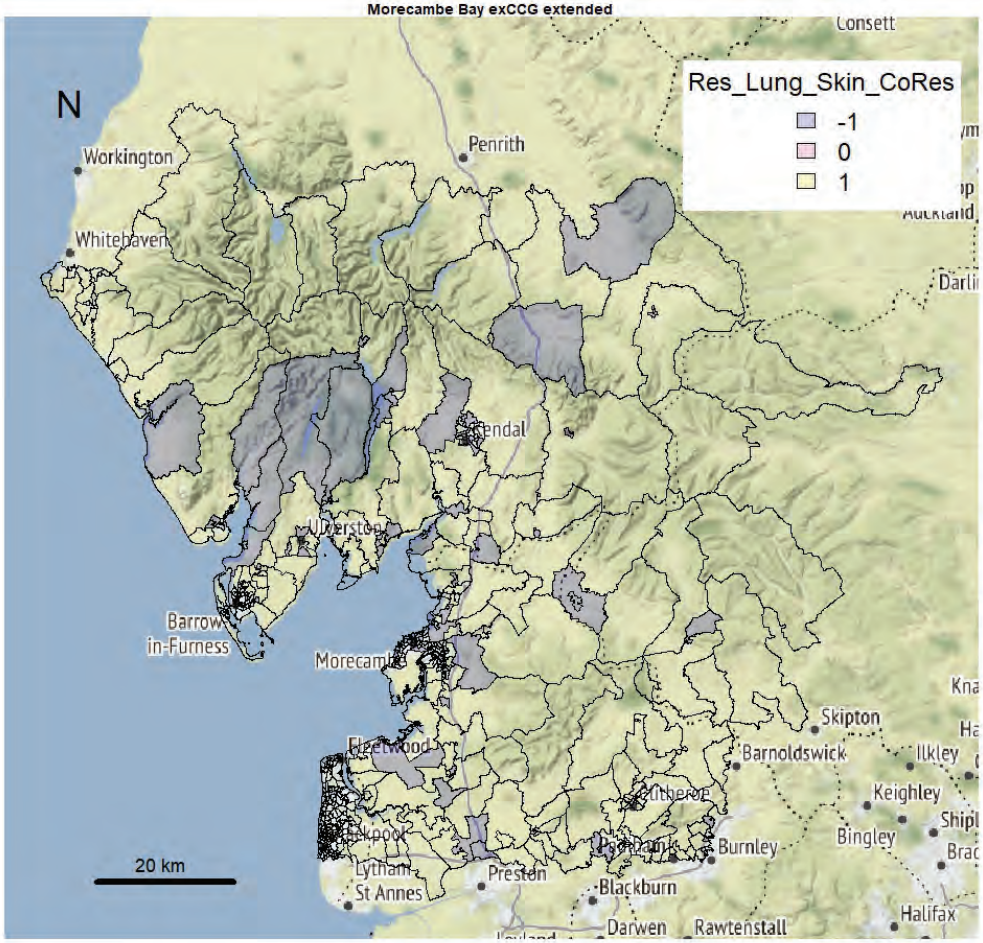

Morecambe Bay exCCG extended

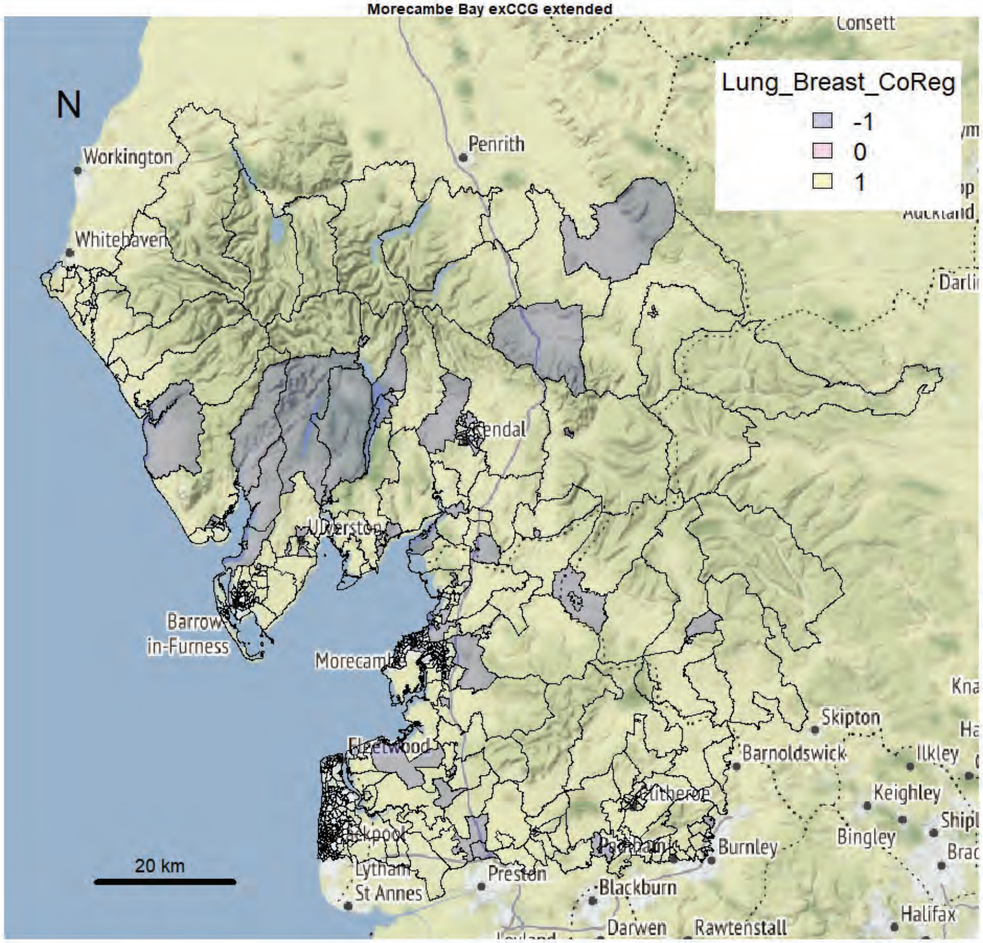


Morecambe Bay exCCG extended

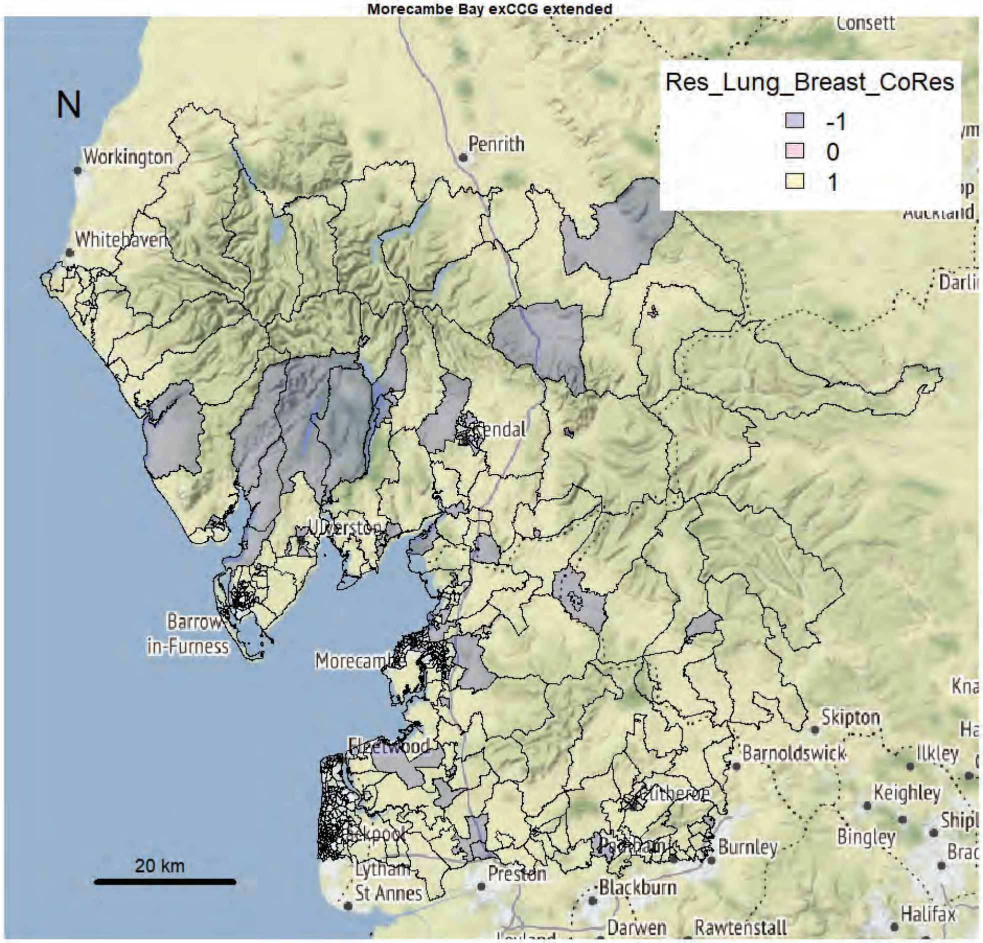

Morecambe Bay exCCG extended

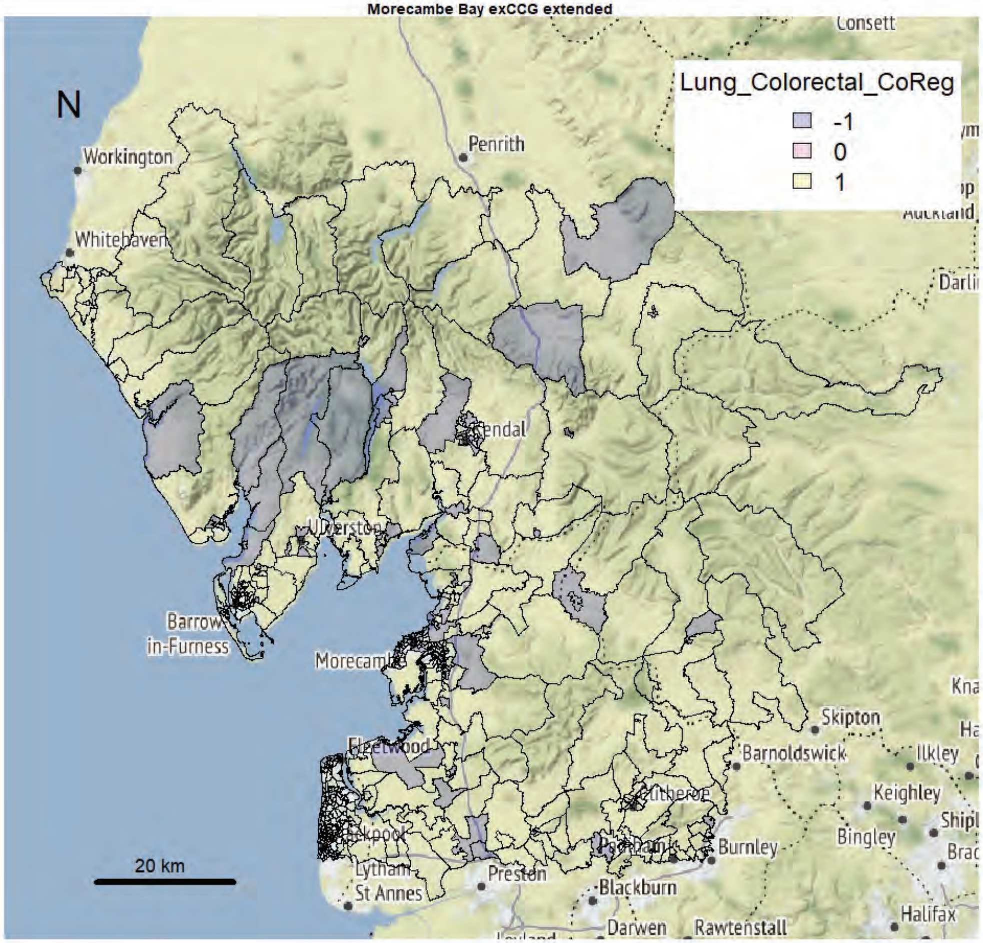


Morecambe Bay exCCG extended

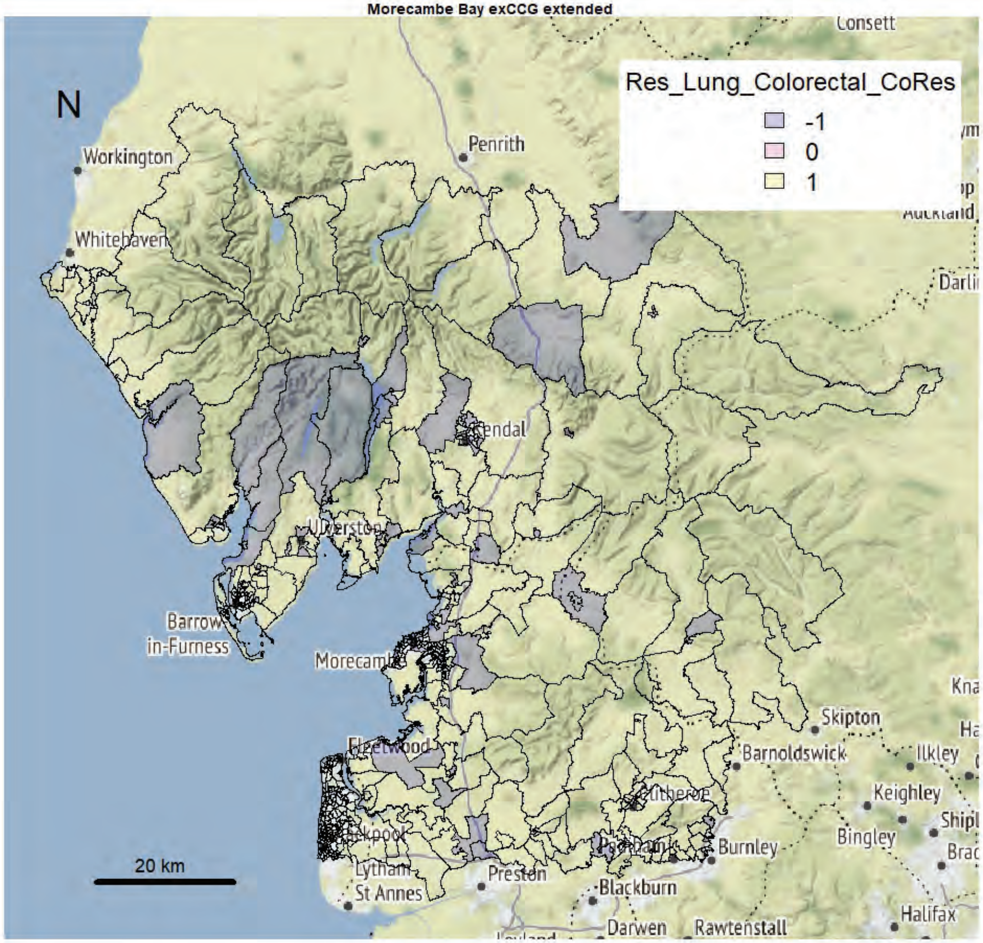

Morecambe Bay exCCG extended

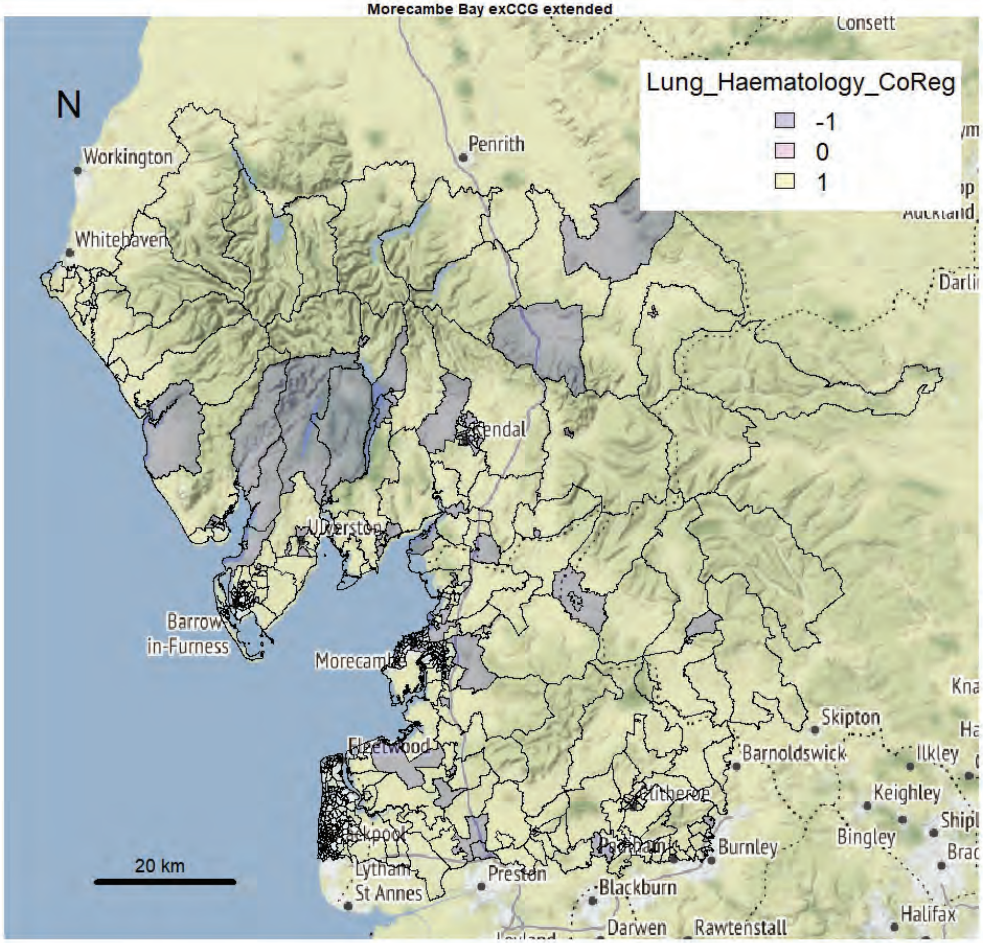


Morecambe Bay exCCG extended

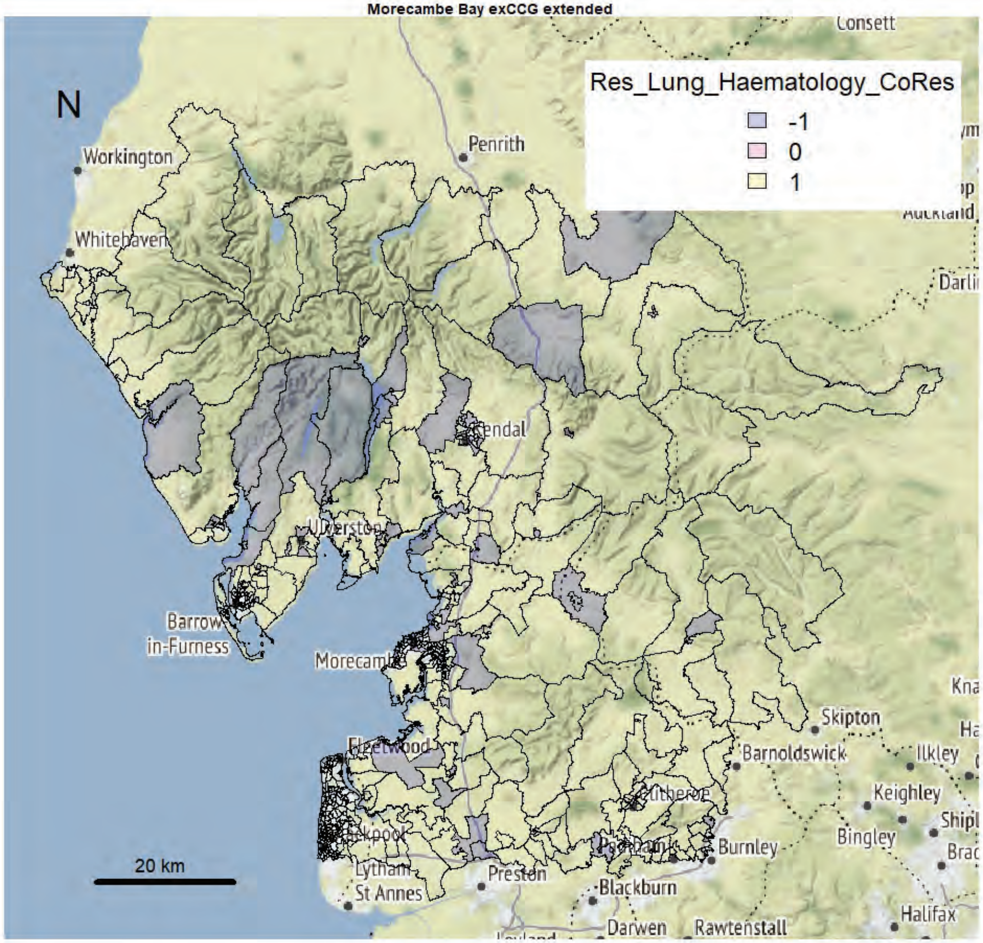

Morecambe Bay exCCG extended

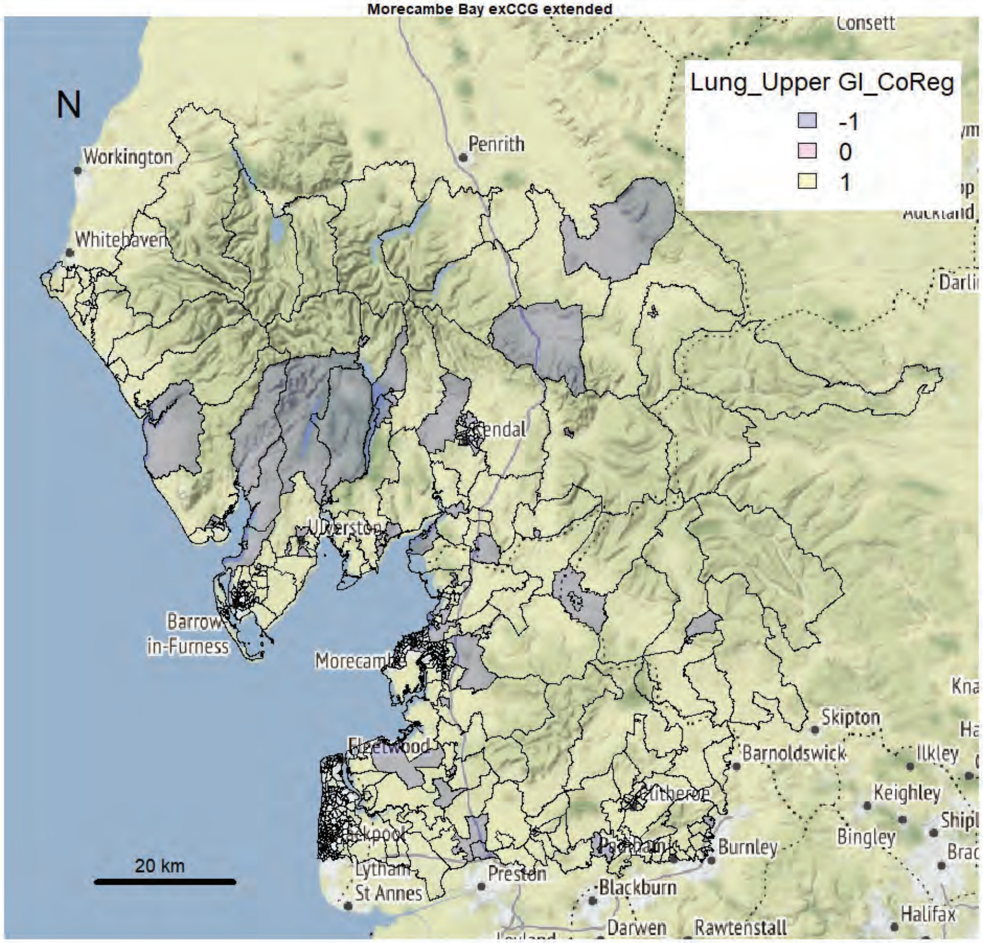


Morecambe Bay exCCG extended

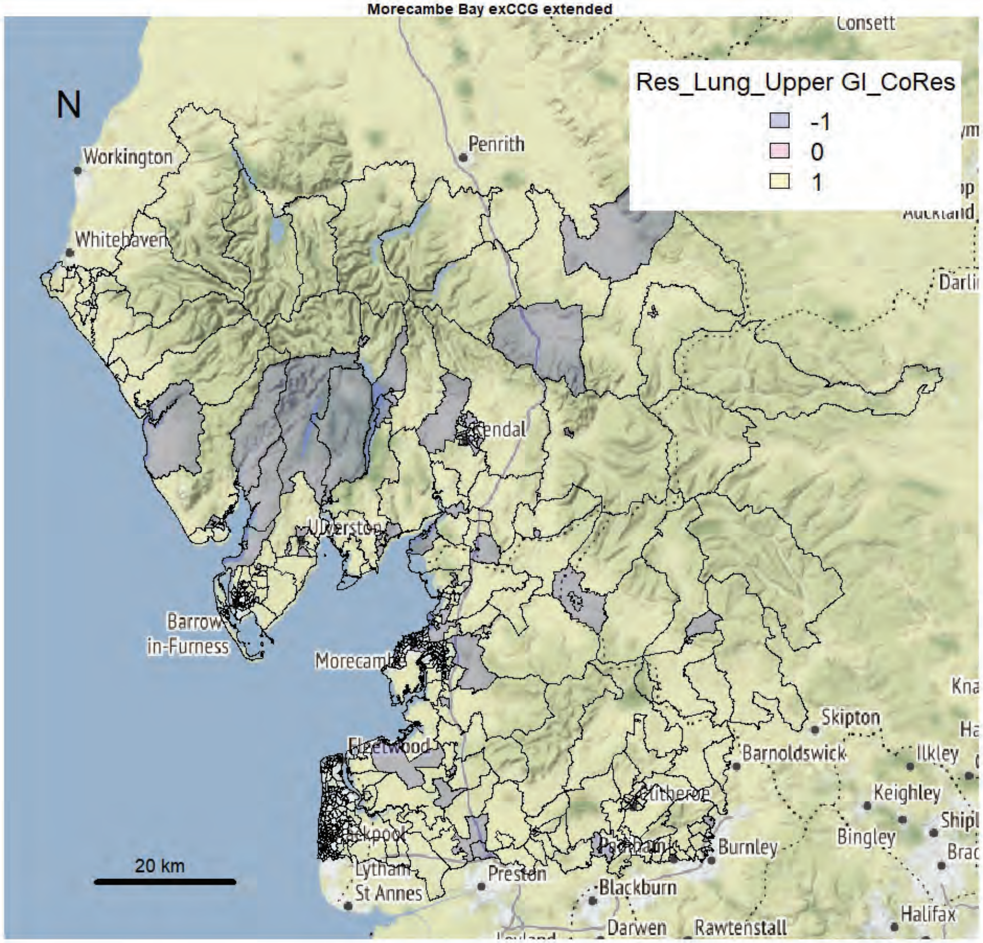

Morecambe Bay exCCG extended

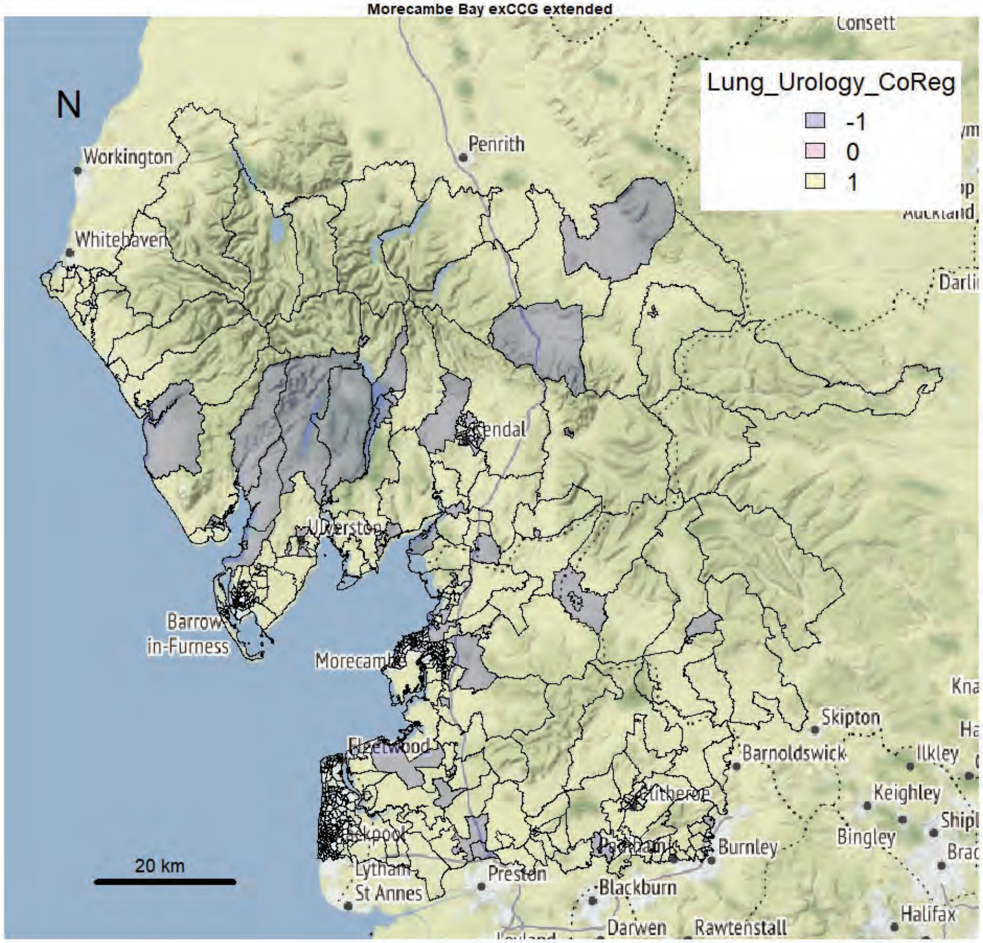


Morecambe Bay exCCG extended

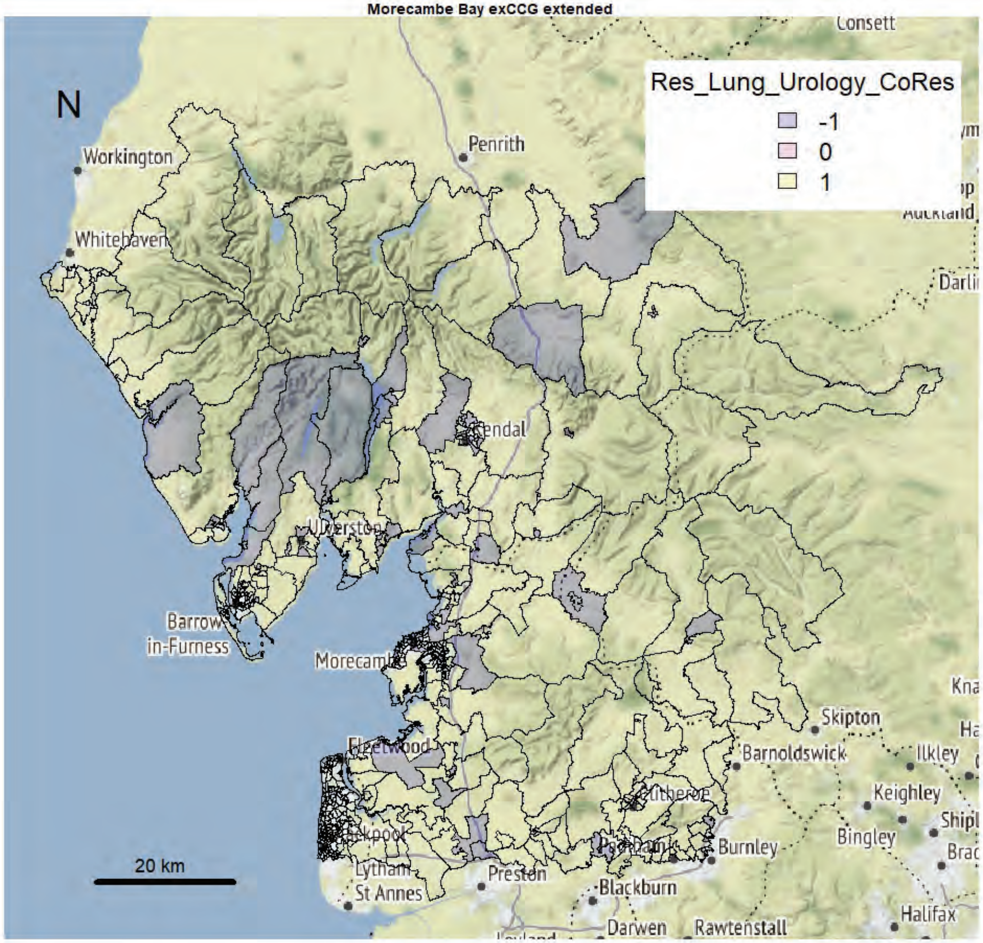

Morecambe Bay exCCG extended

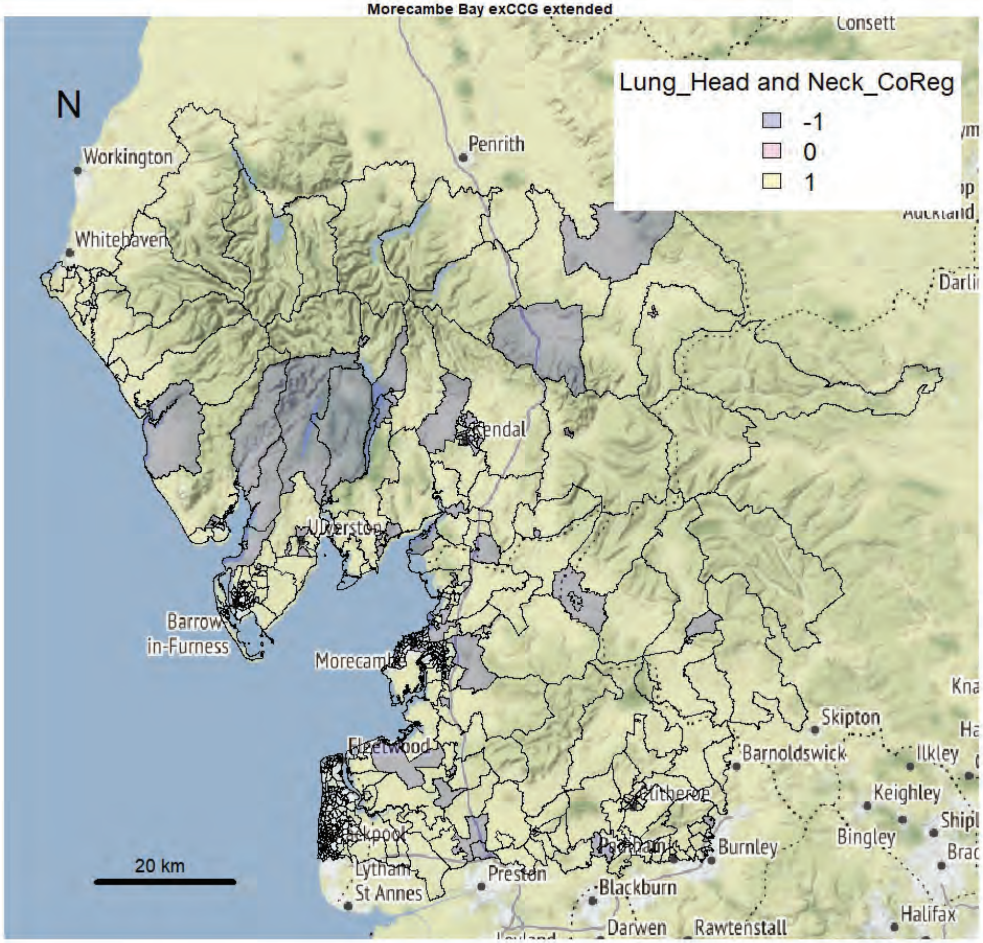


Morecambe Bay exCCG extended

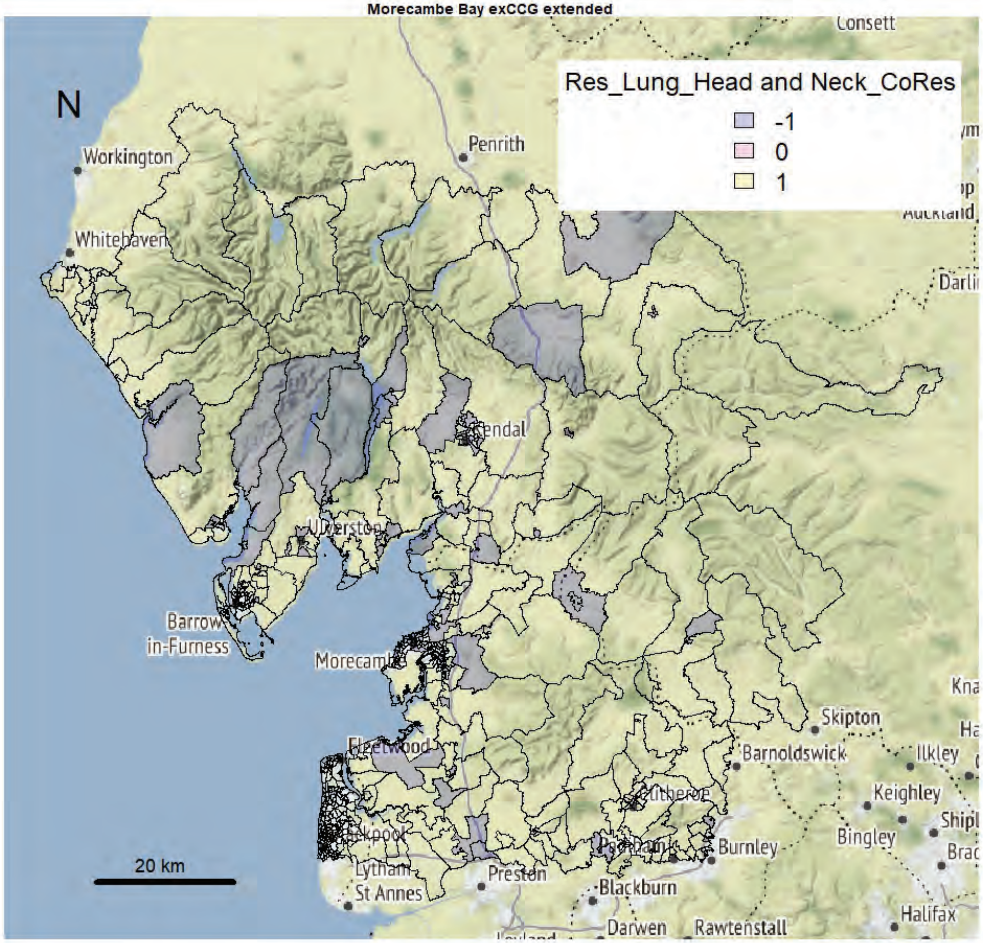

Morecambe Bay exCCG extended

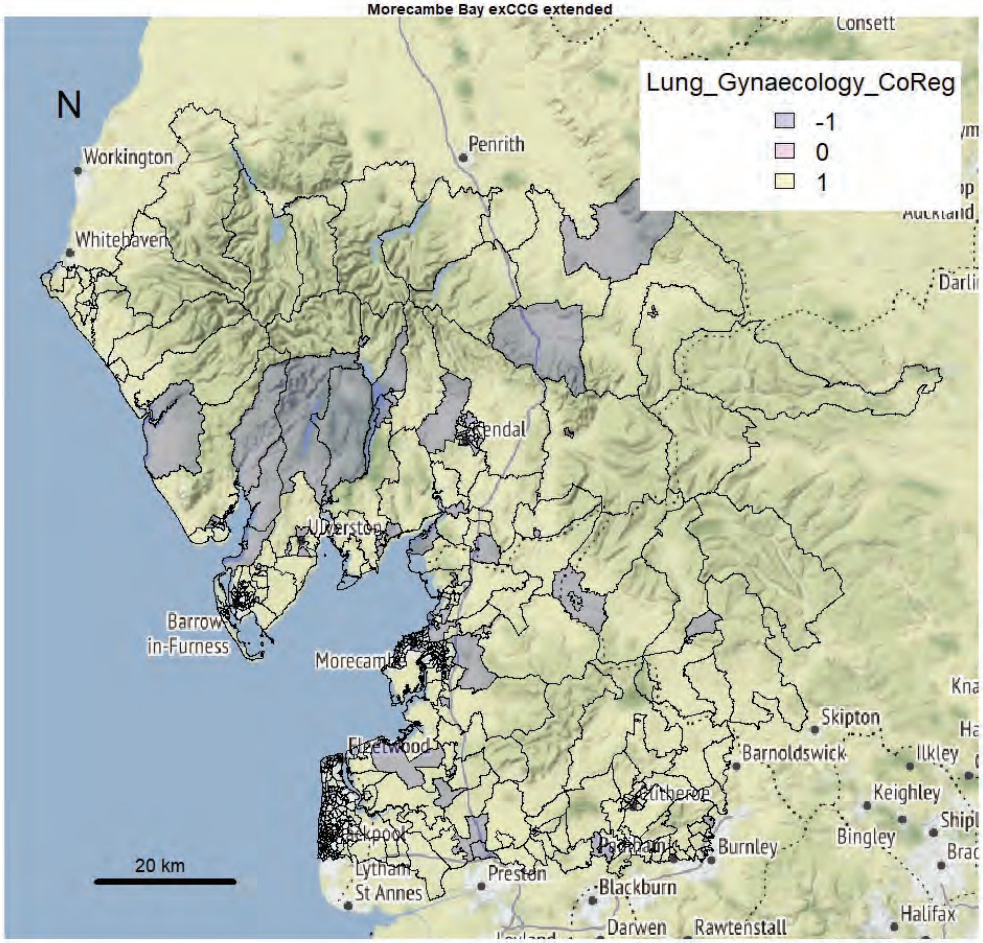


Morecambe Bay exCCG extended

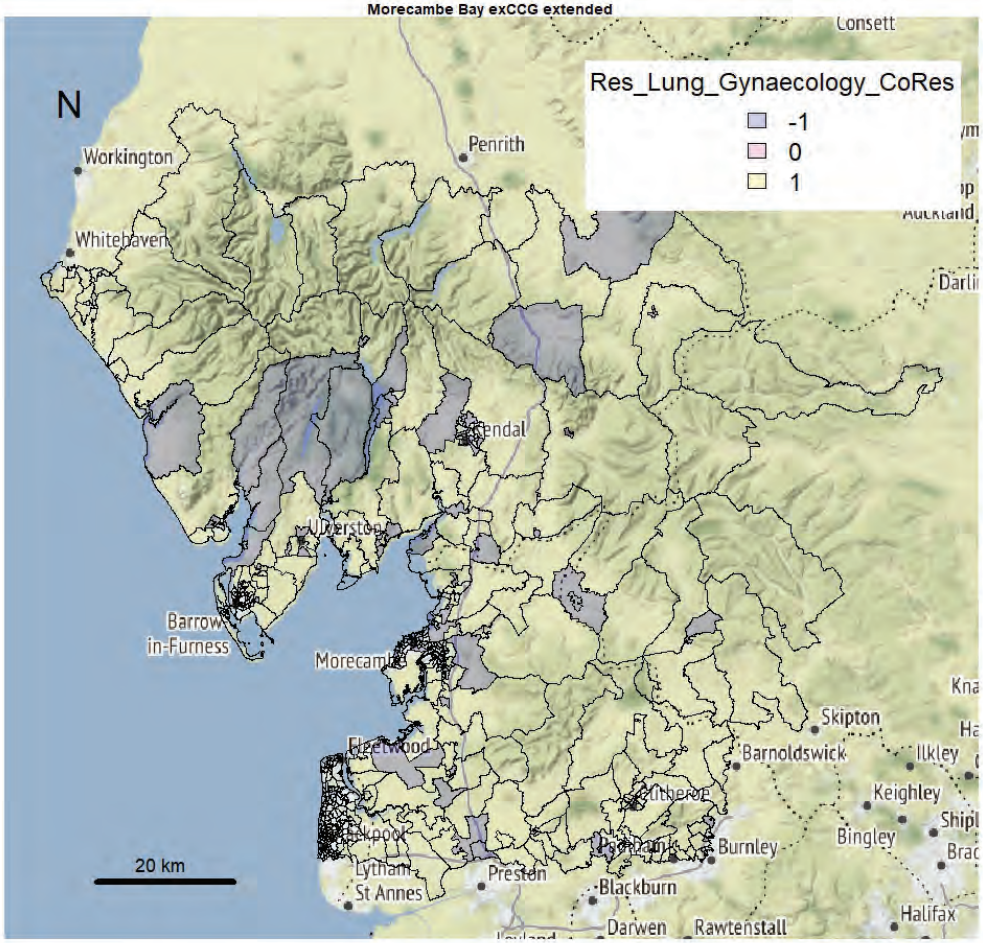

Morecambe Bay exCCG extended

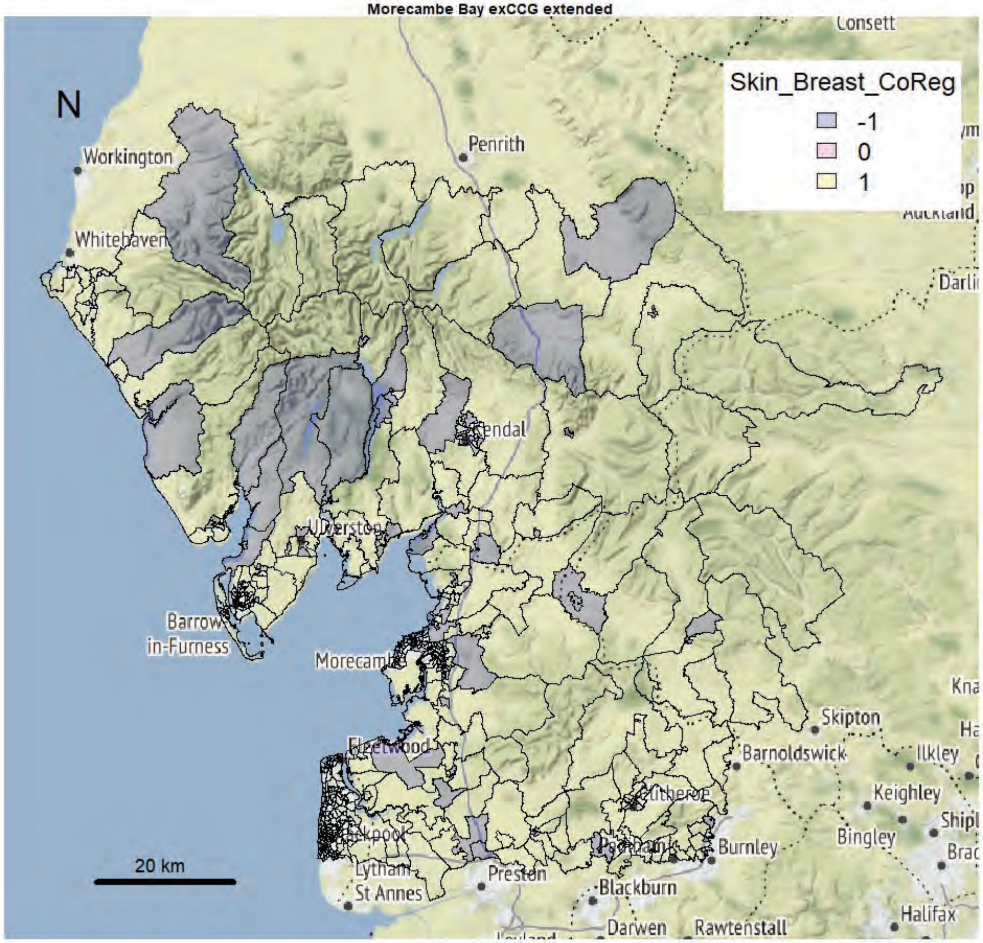


Morecambe Bay exCCG extended

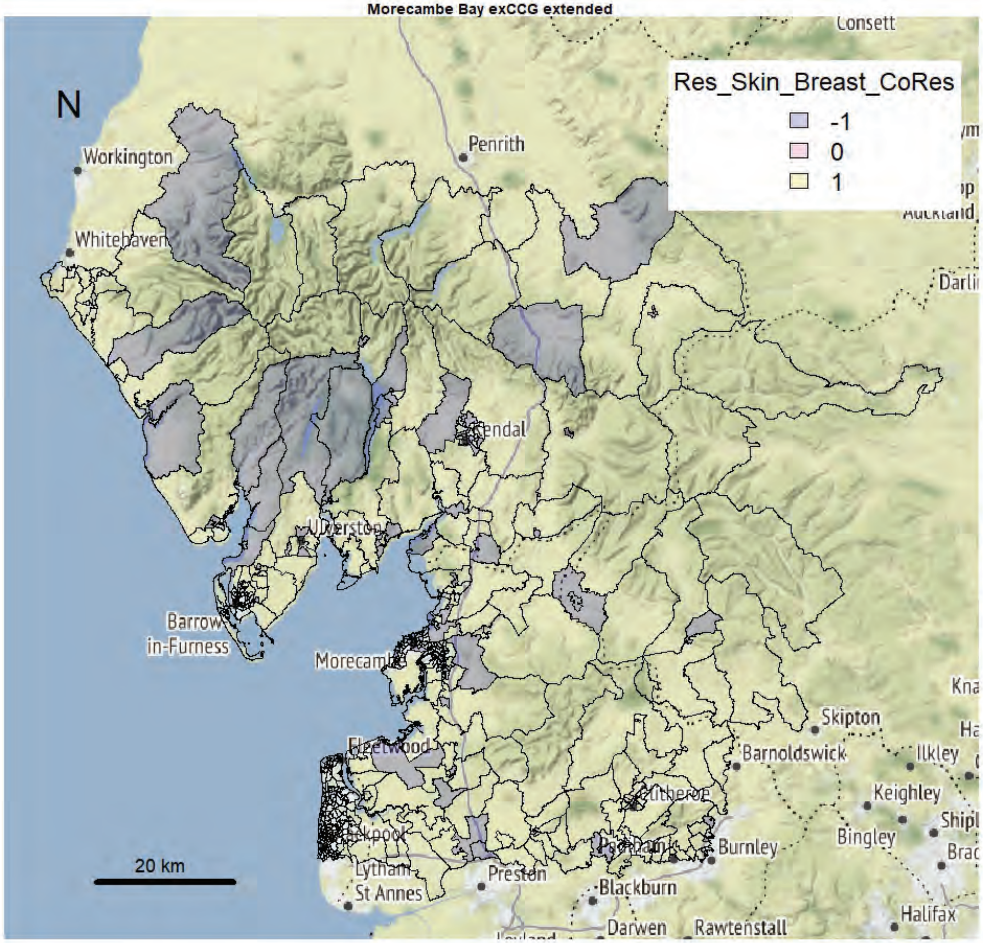

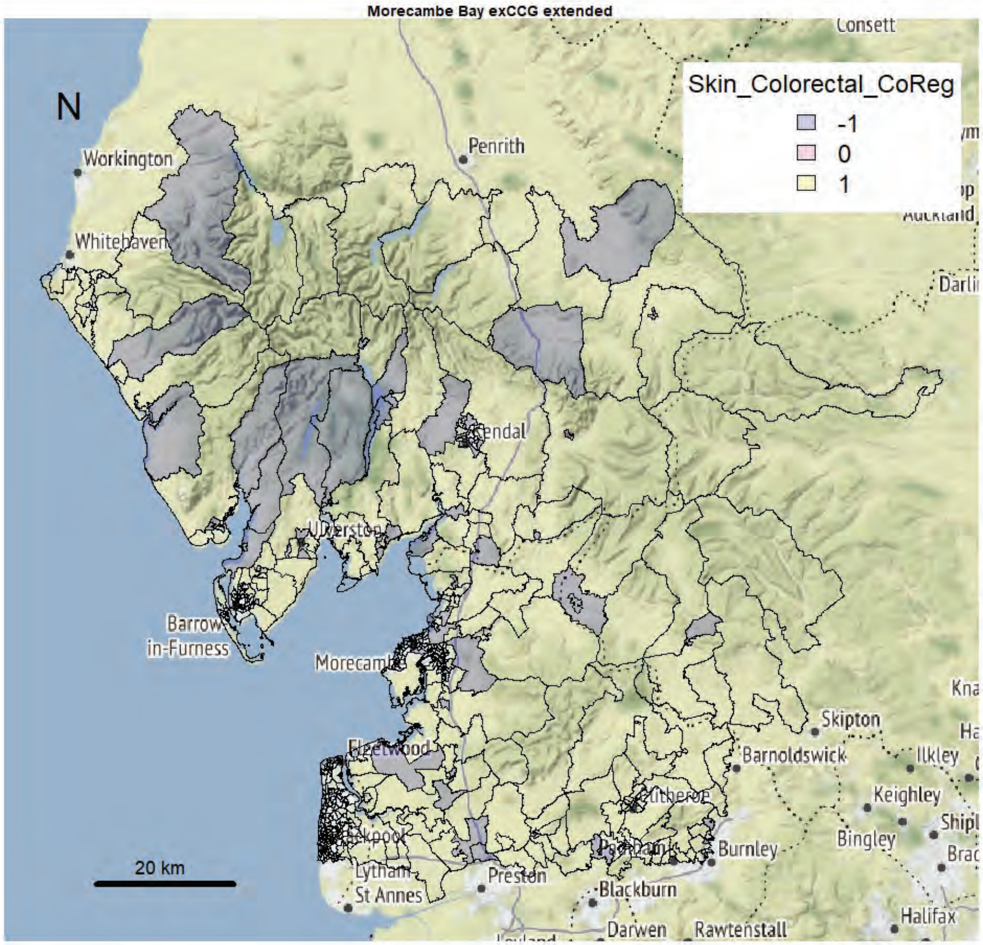
Morecambe Bay exCCG extended
N
Skin_Colorectal_CoReg
-1
0
1
Workington
Whitehaven
Penrith
Consett
Kendal
Barrow-in-Furness
Morecambe
Fleetwood
Lytham St Annes
Preston
Blackburn
Darwen
Rawtenstall
Burnley
Barnoldswick
Skipton
Ilkley
Keighley
Bingley
Halifax
20 km

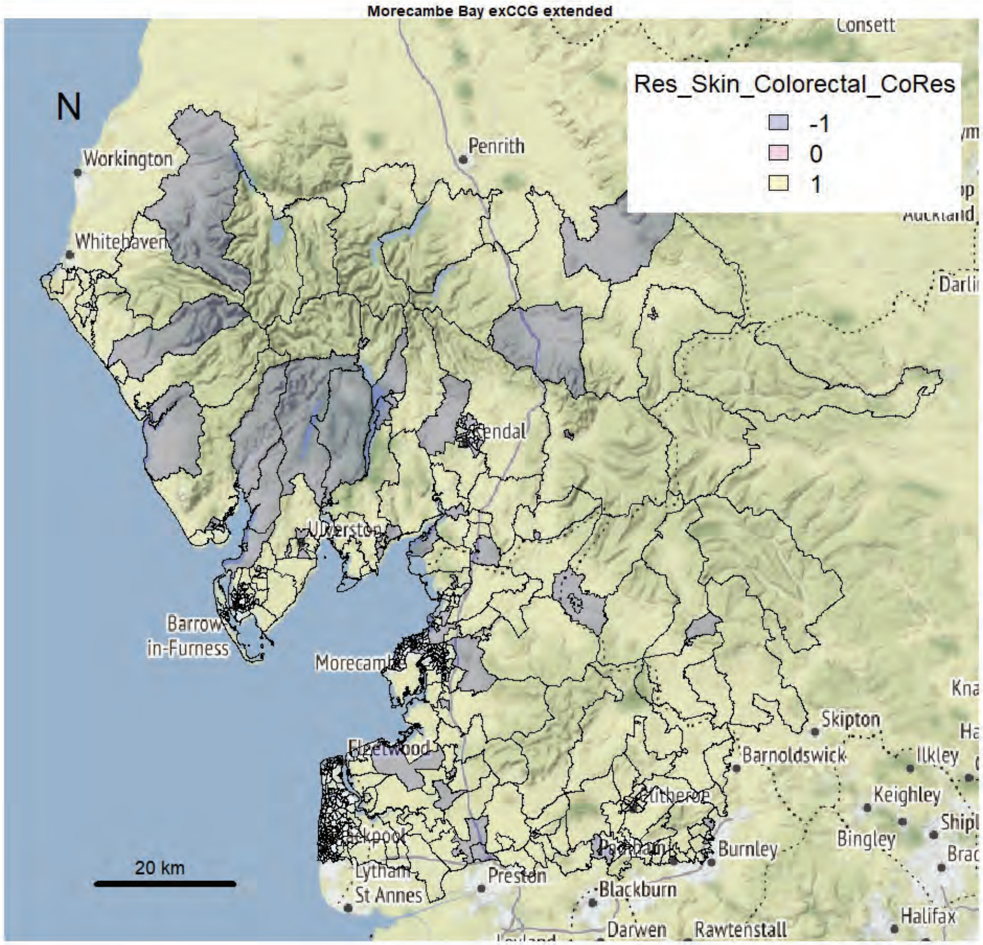
Morecambe Bay exCCG extended
N
Res_Skin_Colorectal_CoRes
-1
0
1
Workington
Whitehaven
Penrith
Consett
Kendal
Barrow-in-Furness
Morecambe
Fleetwood
Lytham St Annes
Preston
Blackburn
Darwen
Rawtenstall
Burnley
Barnoldswick
Skipton
Ilkley
Keighley
Bingley
Halifax
20 km

Morecambe Bay exCCG extended

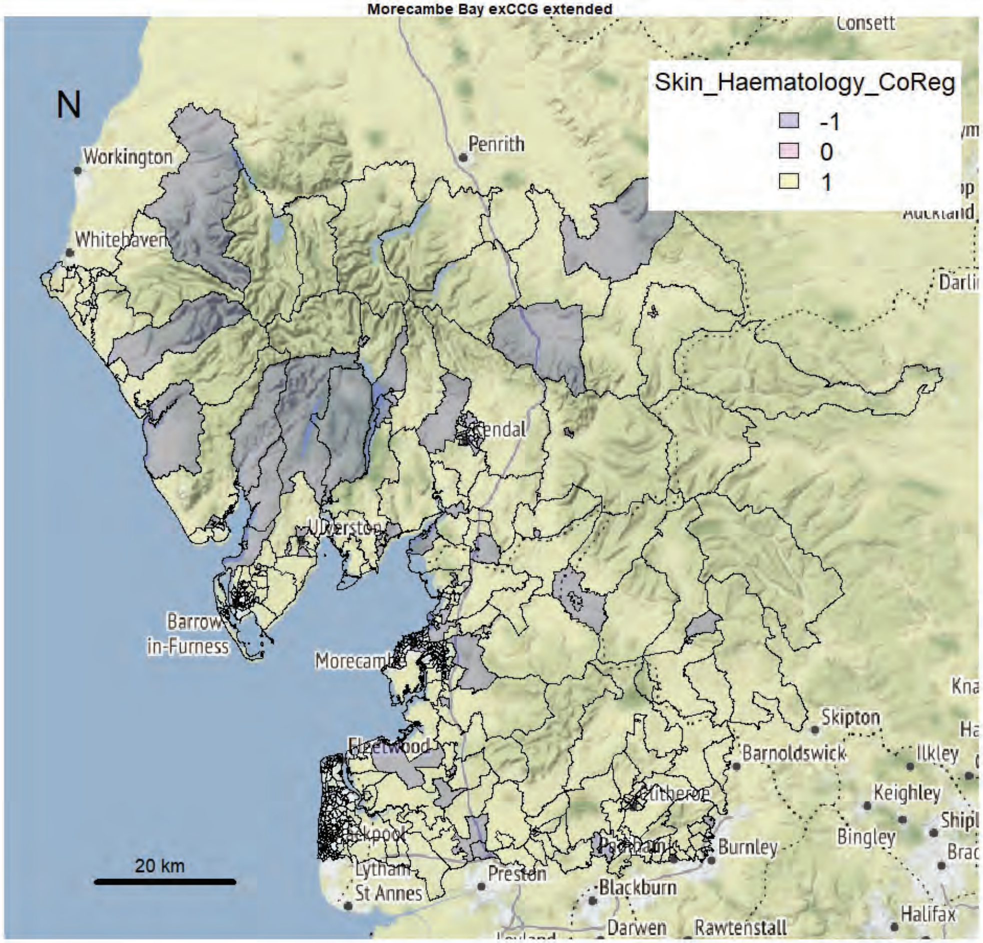


Morecambe Bay exCCG extended

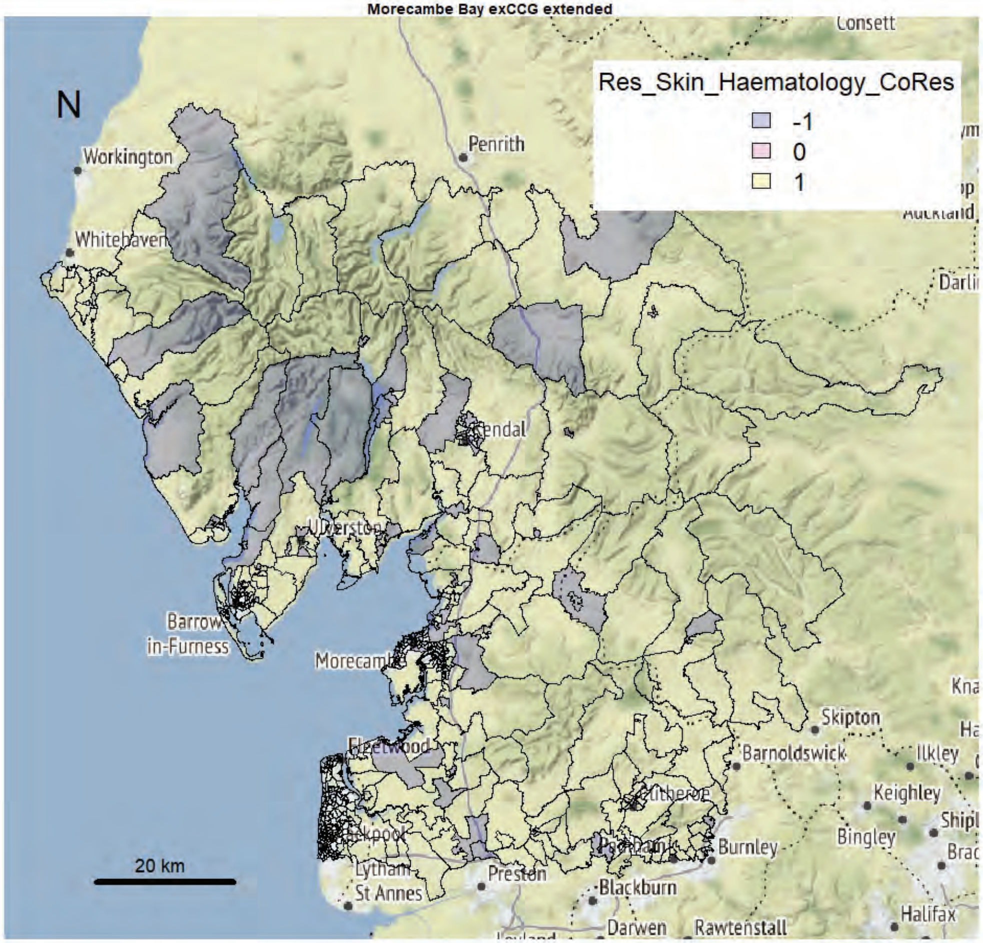

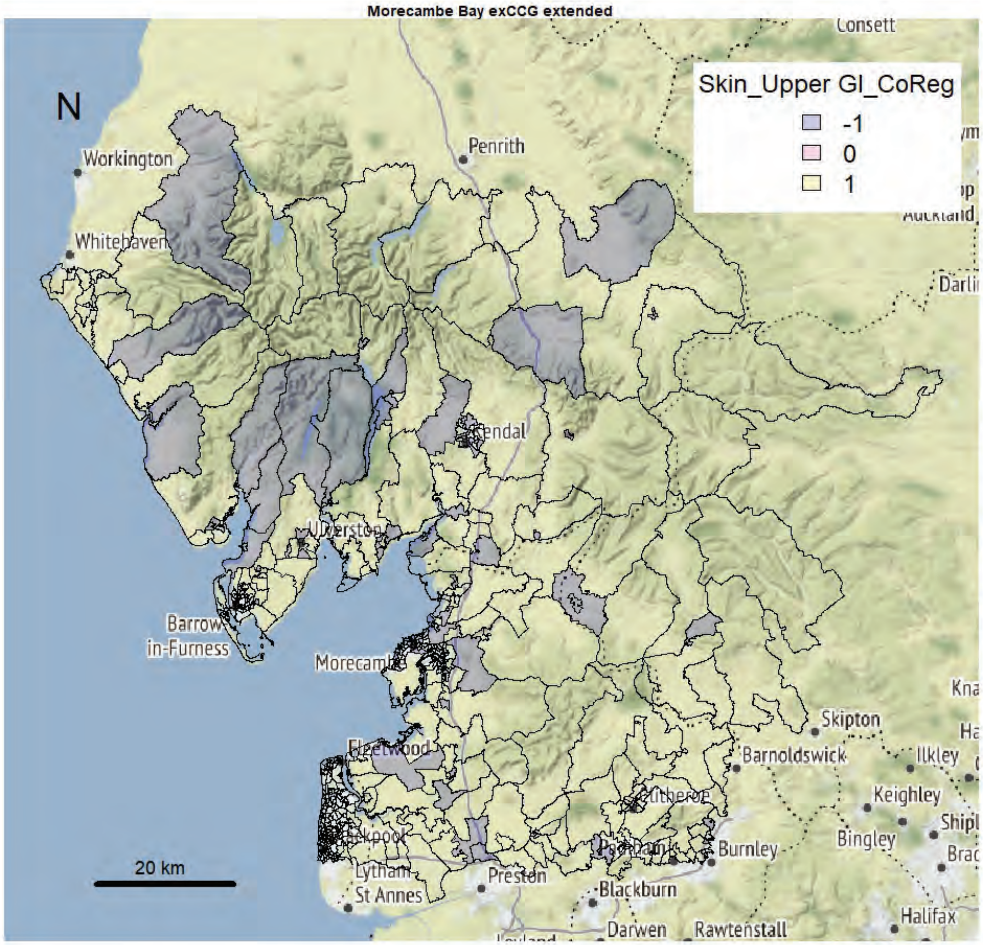
Morecambe Bay exCCG extended
N
Skin_Upper GI_CoReg
-1
0
1
Workington
Whitehaven
Penrith
Consett
Kendal
Ulverston
Barrow-in-Furness
Morecambe
Fleetwood
Blackpool
Lytham St Annes
Preston
Blackburn
Darwen
Rawtenstall
Burnley
Barnoldswick
Skipton
Ilkley
Keighley
Bingley
Halifax
Darlington
20 km

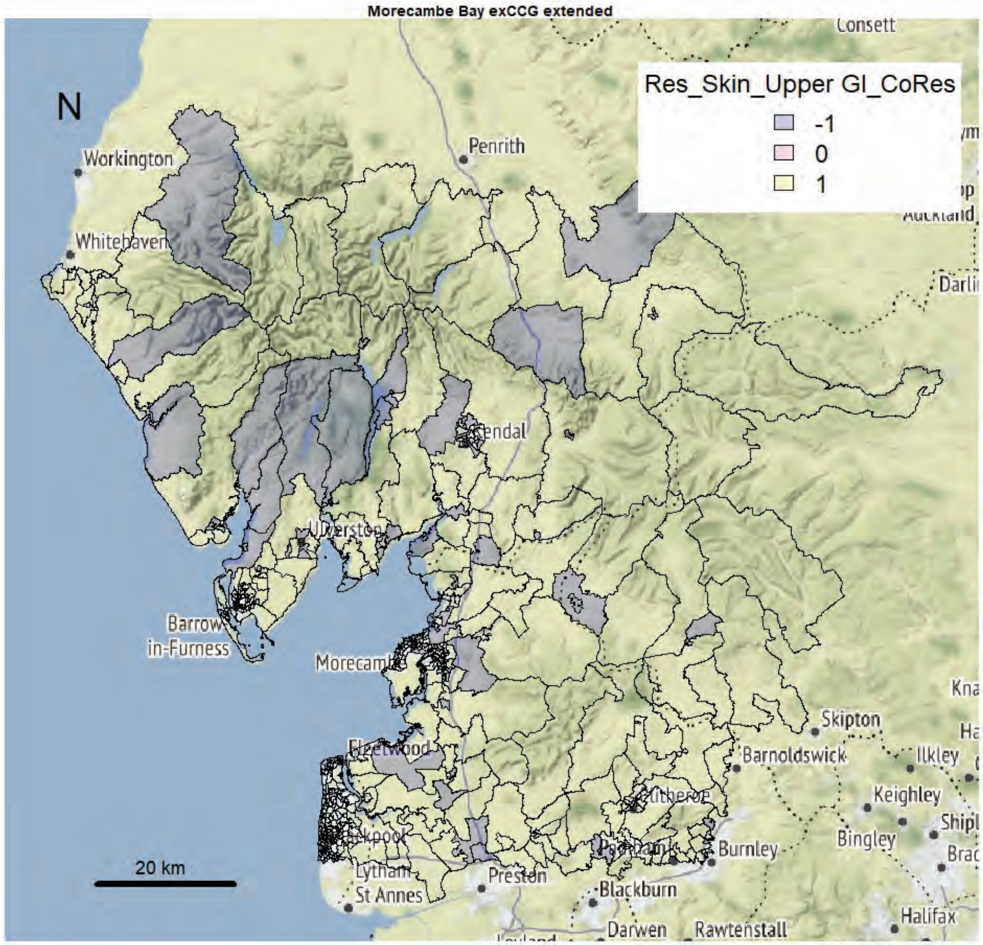
Morecambe Bay exCCG extended
N
Res_Skin_Upper GI_CoRes
-1
0
1
Workington
Whitehaven
Penrith
Consett
Kendal
Ulverston
Barrow-in-Furness
Morecambe
Fleetwood
Blackpool
Lytham St Annes
Preston
Blackburn
Darwen
Rawtenstall
Burnley
Barnoldswick
Skipton
Ilkley
Keighley
Bingley
Halifax
Darlington
20 km

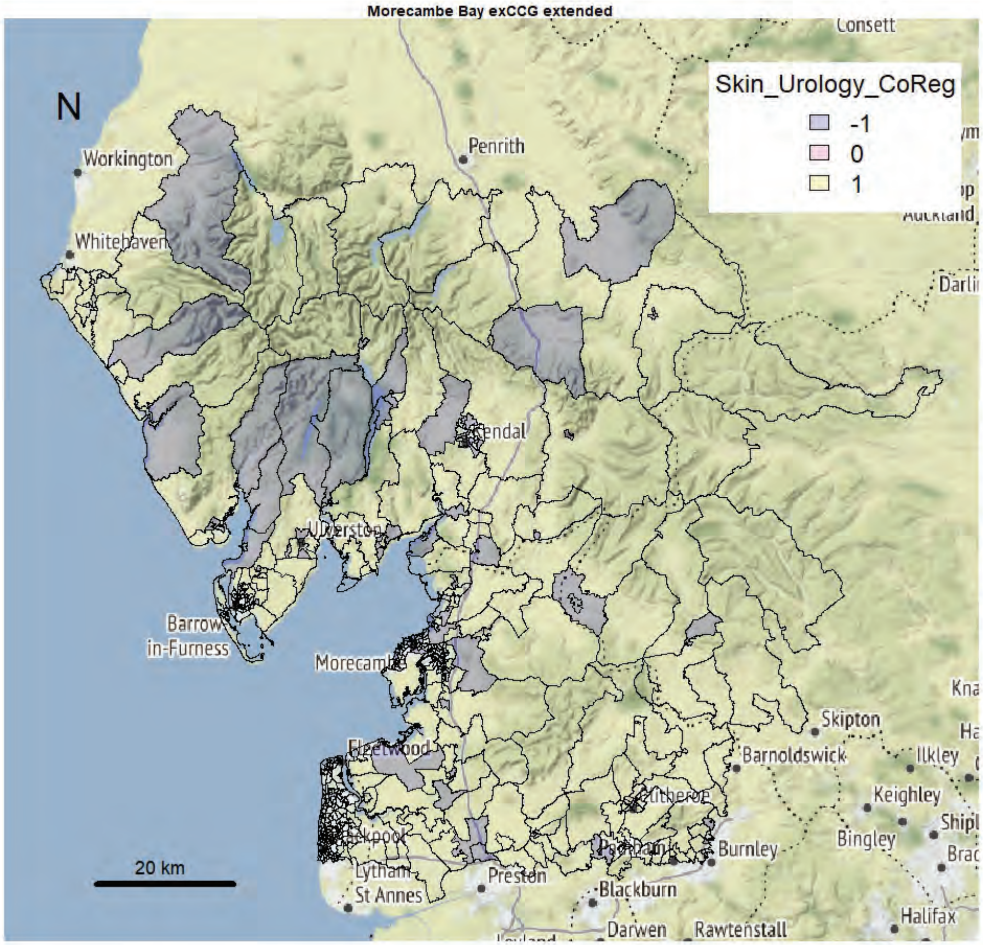
Morecambe Bay exCCG extended
Skin_Urology_CoReg
-1
0
1
N
20 km
Workington
Whitehaven
Penrith
Consett
Barrow-in-Furness
Morecambe
Skipton
Barnoldswick
Ilkley
Keighley
Bingley
Burnley
Preston
Blackburn
Darwen
Rawtenstall
Halifax
St Annes

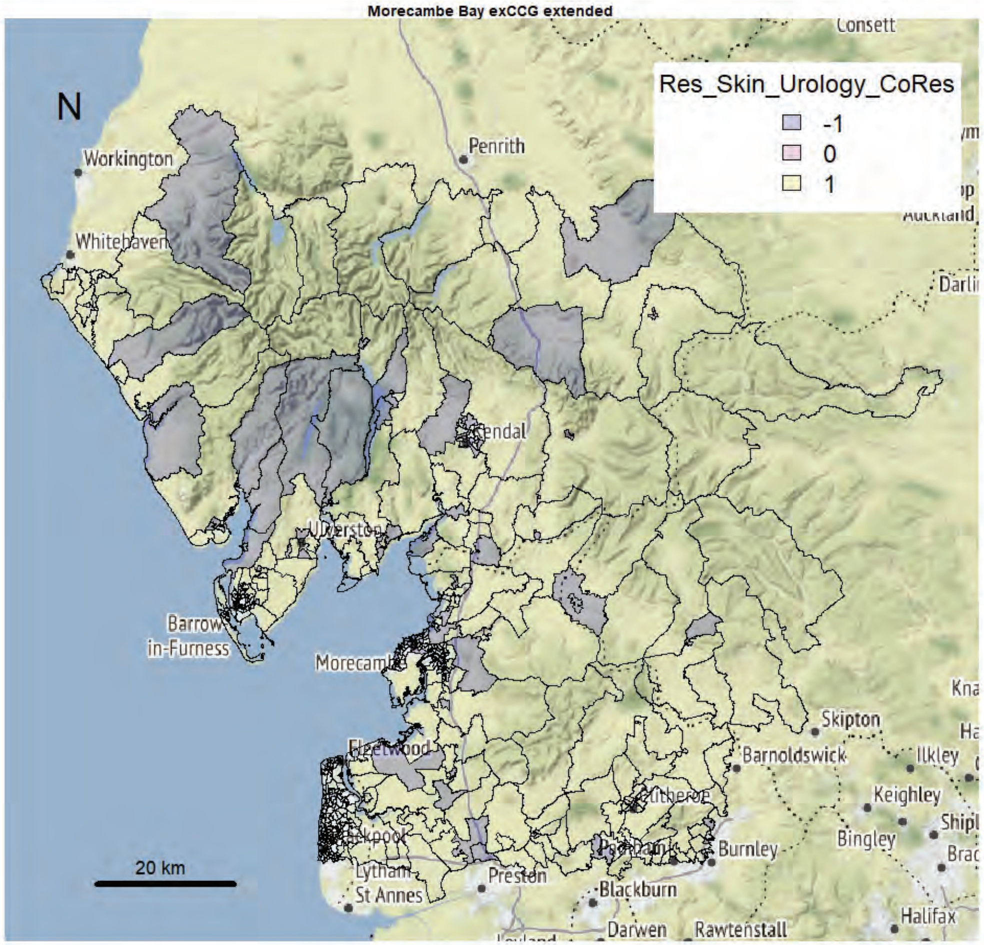
Morecambe Bay exCCG extended
Res_Skin_Urology_CoRes
-1
0
1
N
20 km
Workington
Whitehaven
Penrith
Consett
Barrow-in-Furness
Morecambe
Skipton
Barnoldswick
Ilkley
Keighley
Bingley
Burnley
Preston
Blackburn
Darwen
Rawtenstall
Halifax
St Annes

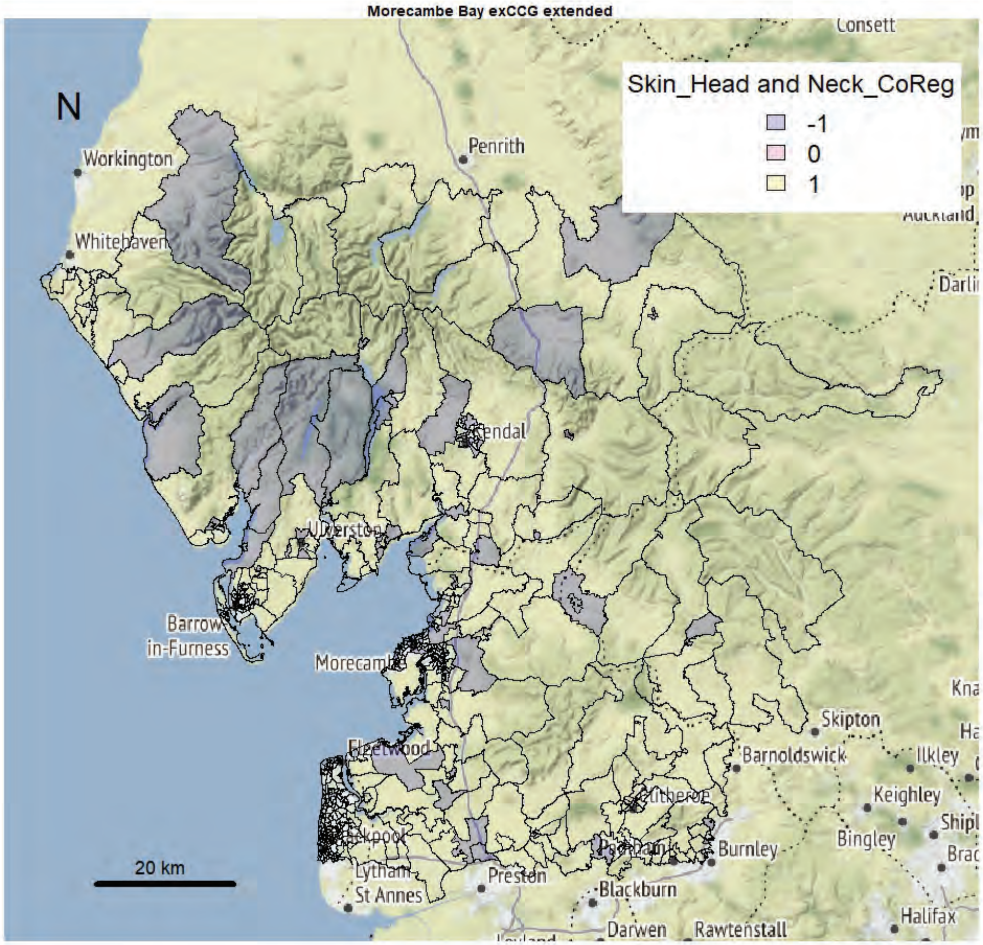
Morecambe Bay exCCG extended
N
Skin_Head and Neck_CoReg
-1
0
1
20 km
Workington
Whitehaven
Penrith
Consett
Darli
Barrow
in-Furness
Morecamb
Skipton
Barnoldswick
Ilkley
Keighley
Bingley
Burnley
Preston
Blackburn
Darwen
Rawtenstall
Halifax
Lytham
St Annes

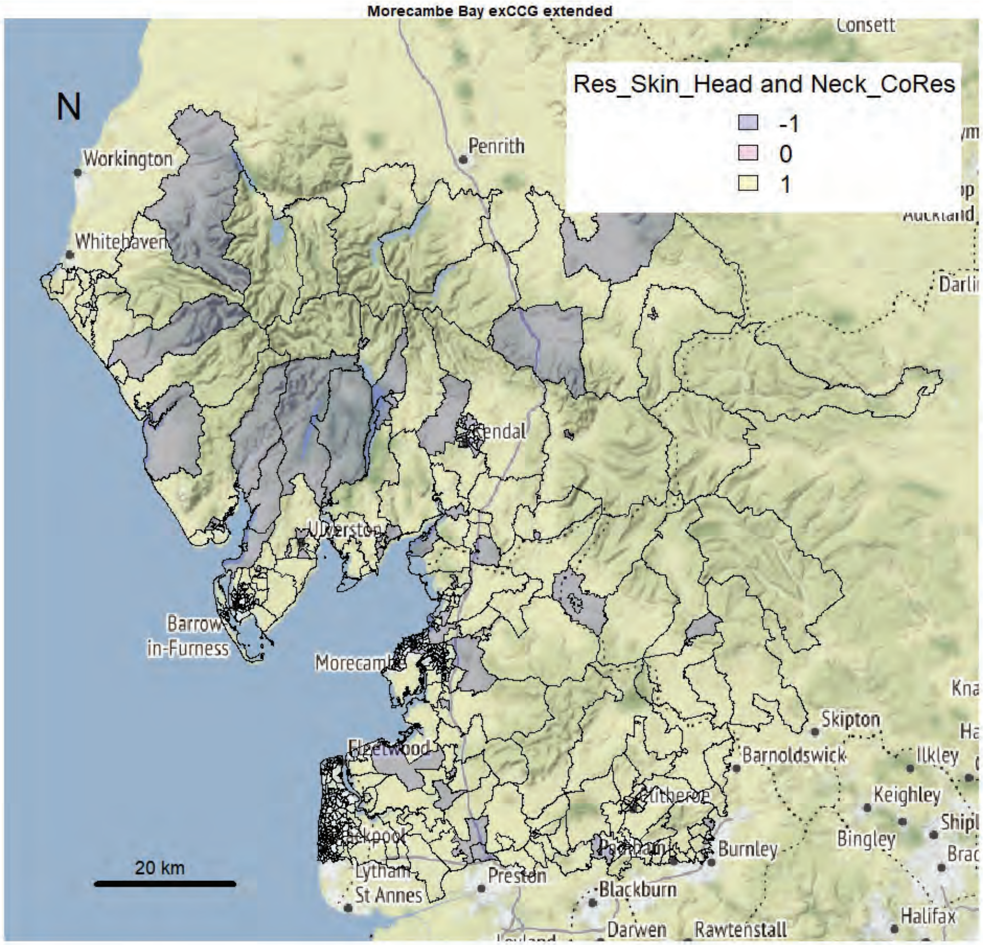
Morecambe Bay exCCG extended
N
Res_Skin_Head and Neck_CoRes
-1
0
1
20 km
Workington
Whitehaven
Penrith
Consett
Darli
Barrow
in-Furness
Morecamb
Skipton
Barnoldswick
Ilkley
Keighley
Bingley
Burnley
Preston
Blackburn
Darwen
Rawtenstall
Halifax
Lytham
St Annes

Morecambe Bay exCCG extended

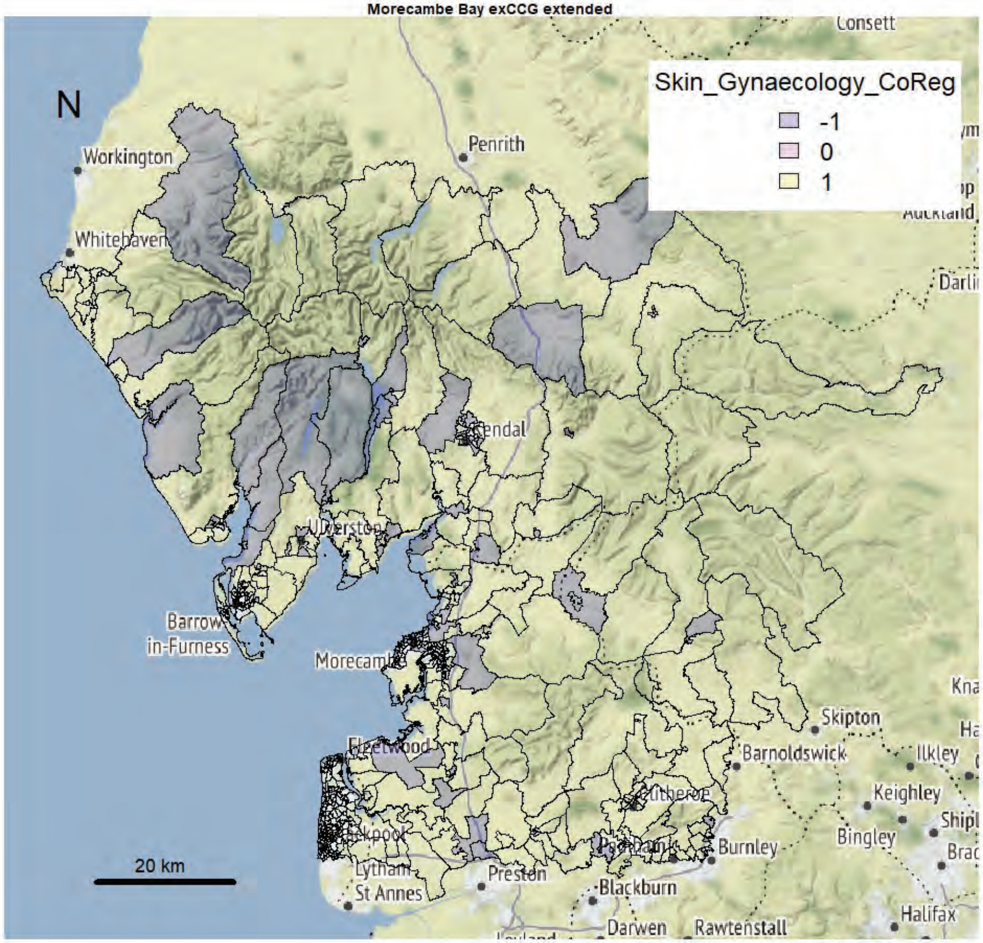


Morecambe Bay exCCG extended

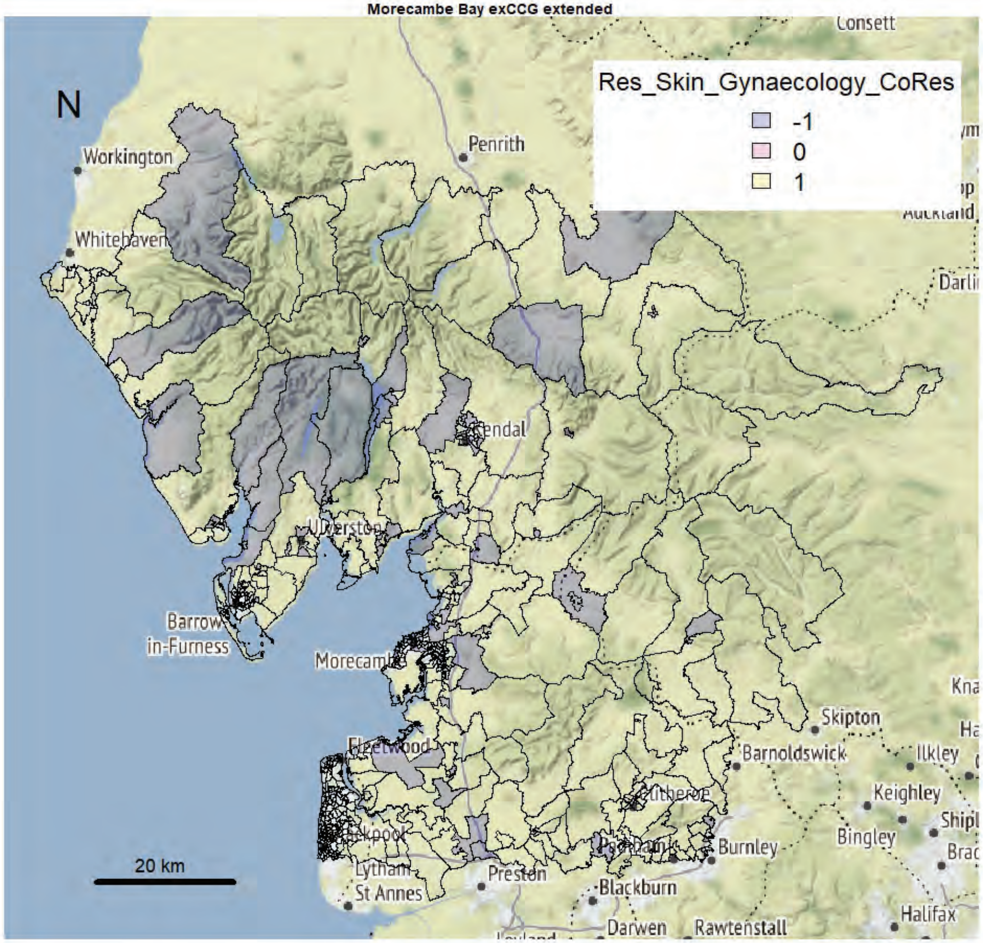

Morecambe Bay exCCG extended

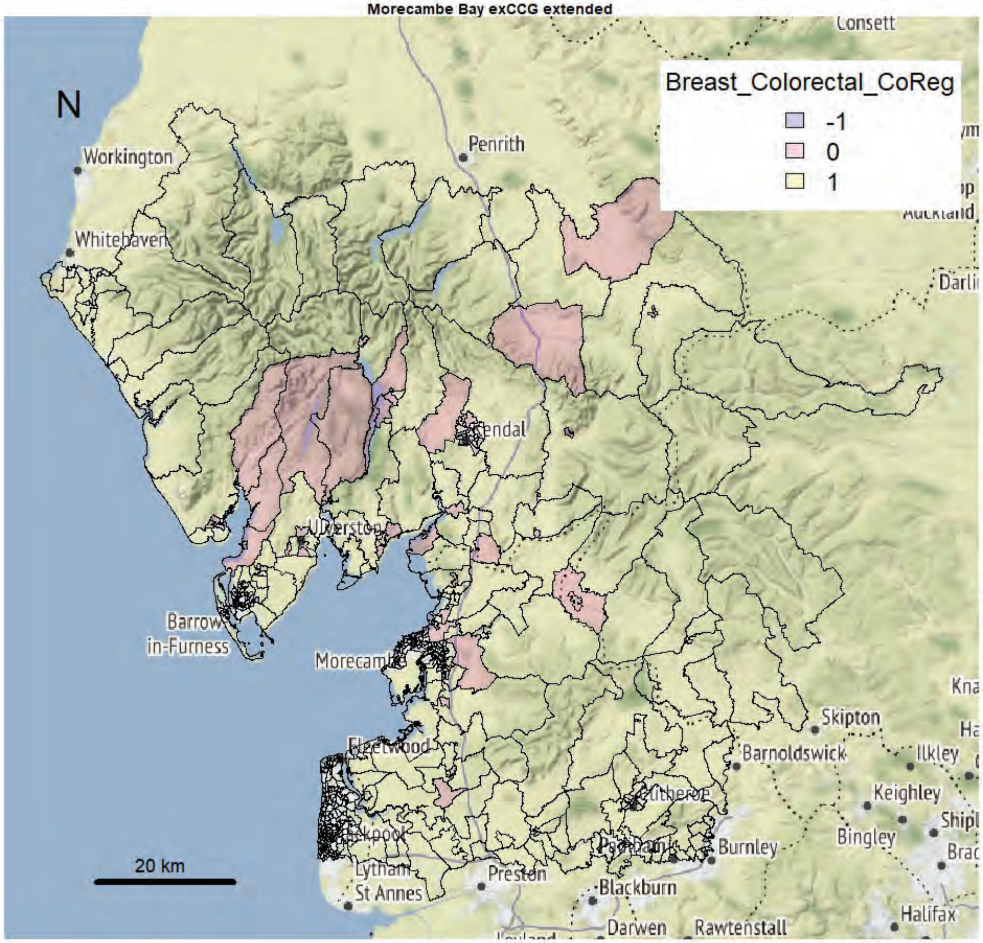


Morecambe Bay exCCG extended

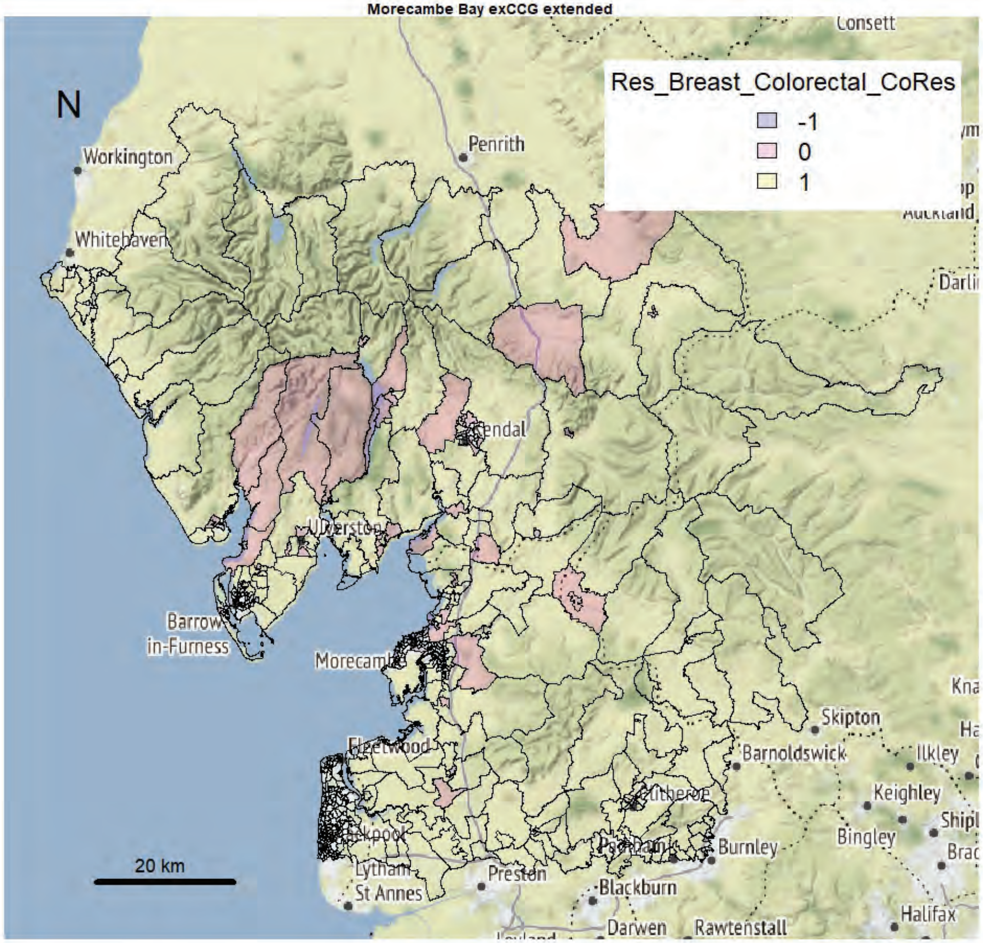

Morecambe Bay exCCG extended

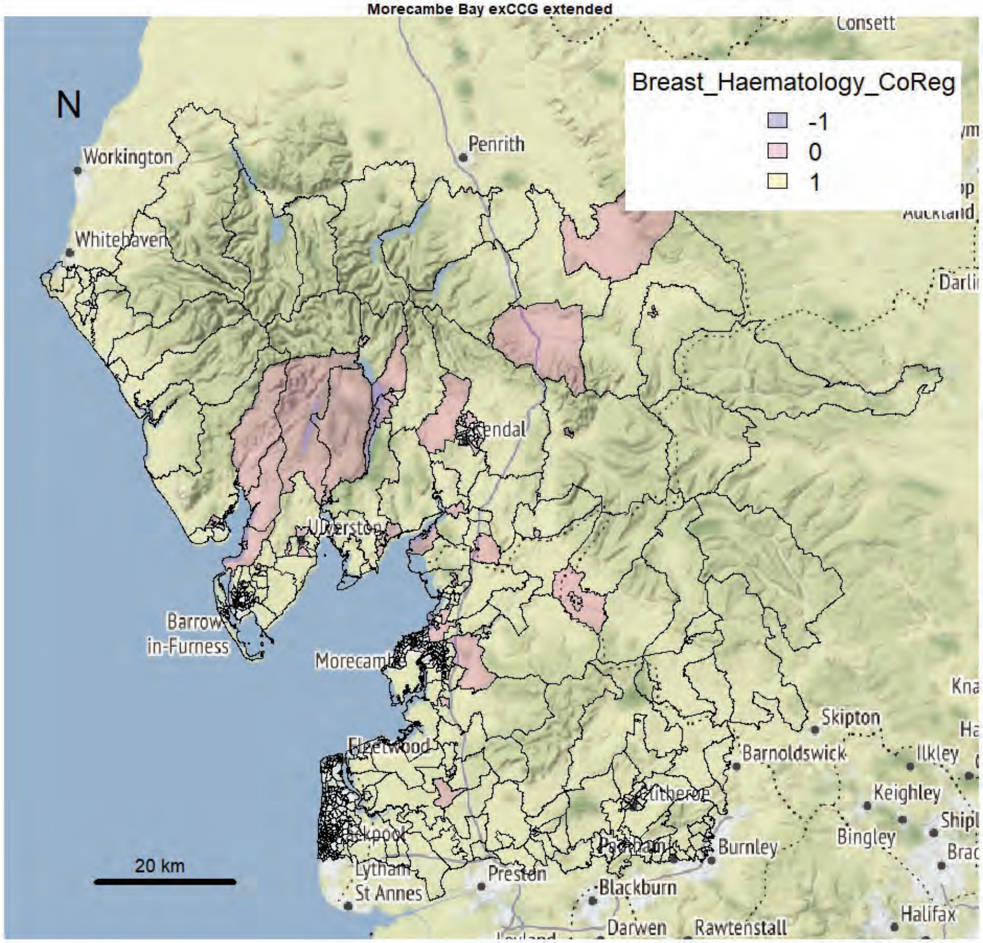


Morecambe Bay exCCG extended

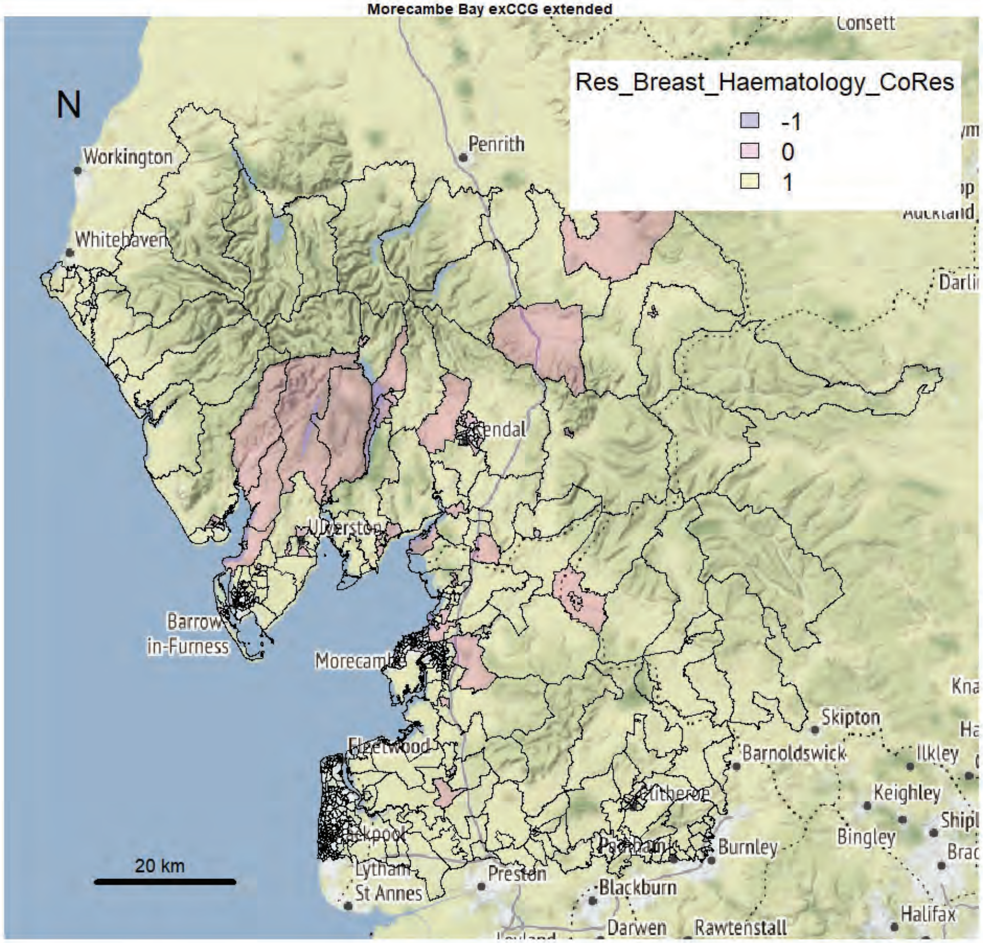

Morecambe Bay exCCG extended

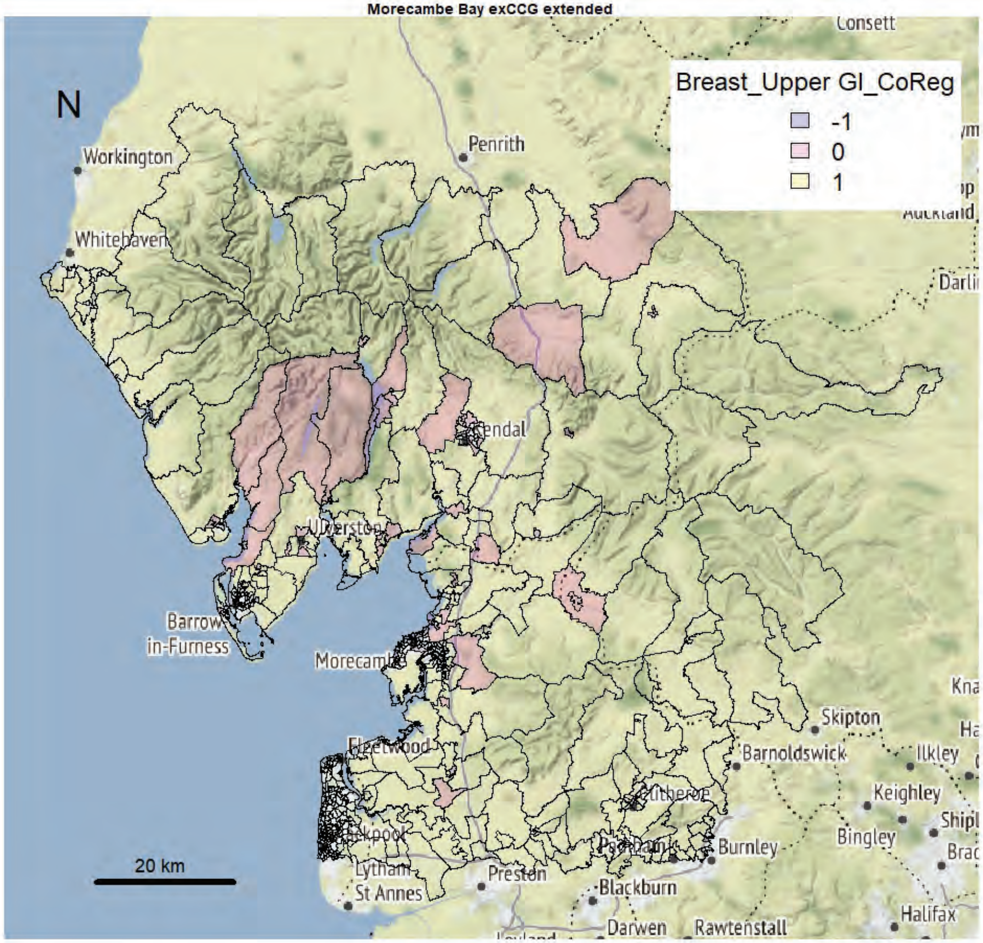


Morecambe Bay exCCG extended

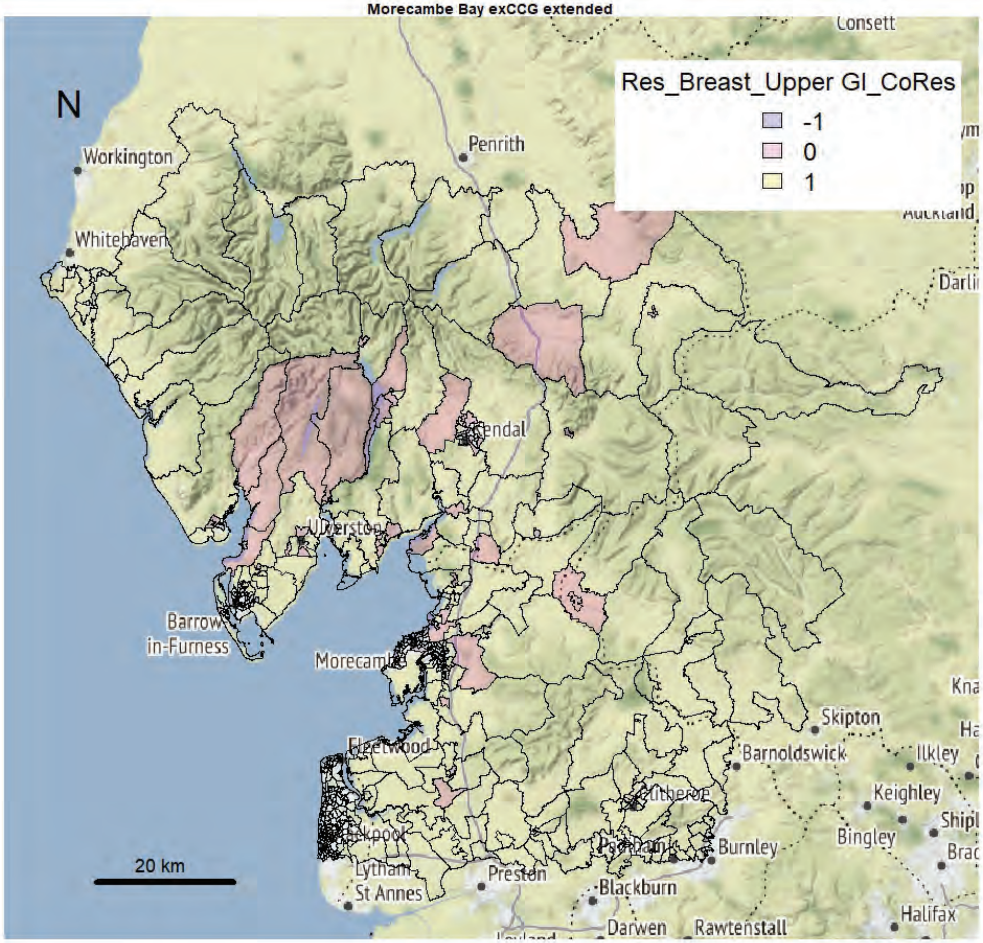

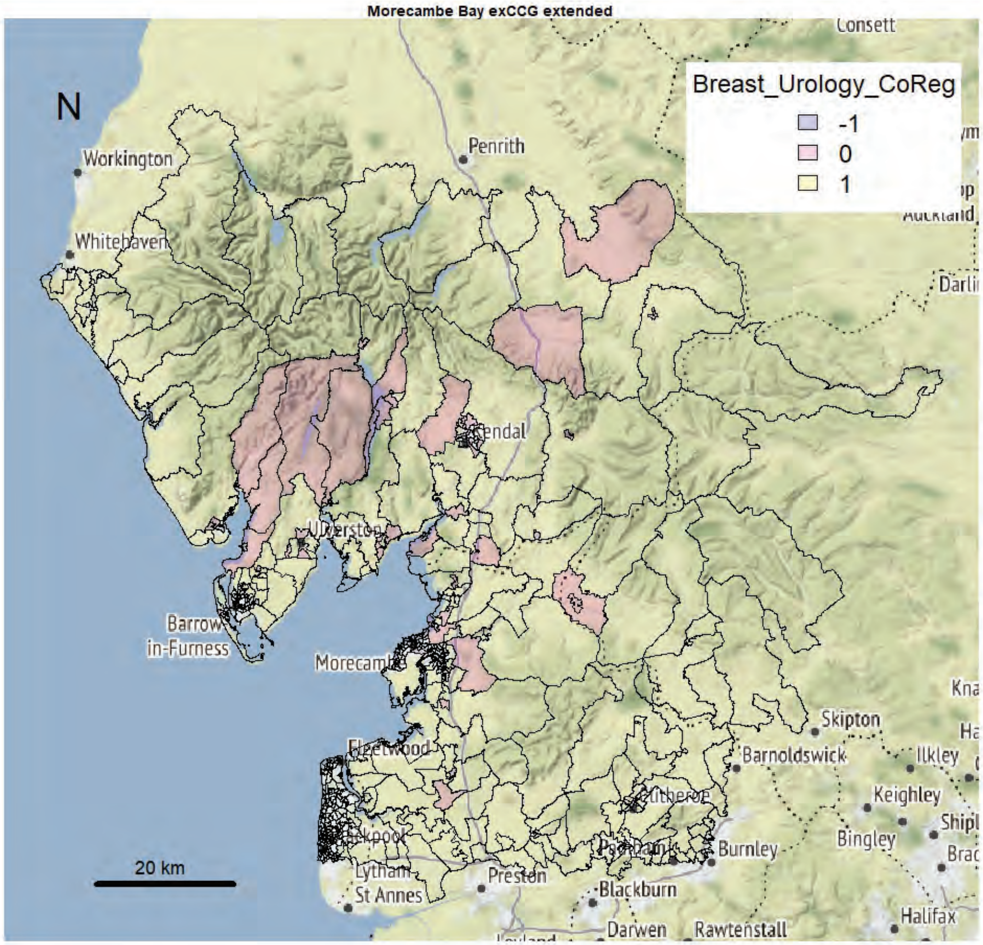
Morecambe Bay exCCG extended
N
Breast_Urology_CoReg
-1
0
1
Workington
Whitehaven
Penrith
Kendal
Barrow-in-Furness
Morecambe
Fleetwood
Lytham St Annes
Preston
Blackburn
Darwen
Rawtenstall
Burnley
Barnoldswick
Skipton
Ilkley
Keighley
Bingley
Halifax
Consett
20 km

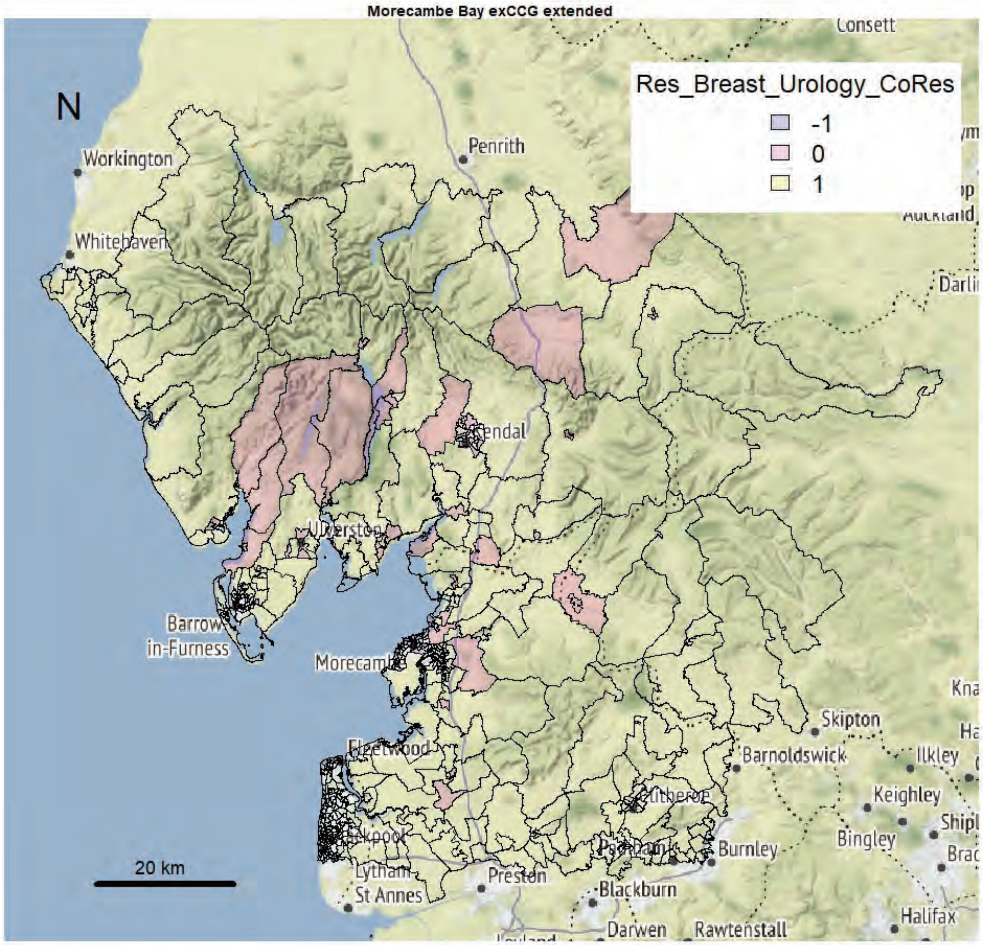
Morecambe Bay exCCG extended
N
Res_Breast_Urology_CoRes
-1
0
1
Workington
Whitehaven
Penrith
Kendal
Barrow-in-Furness
Morecambe
Fleetwood
Lytham St Annes
Preston
Blackburn
Darwen
Rawtenstall
Burnley
Barnoldswick
Skipton
Ilkley
Keighley
Bingley
Halifax
Consett
20 km

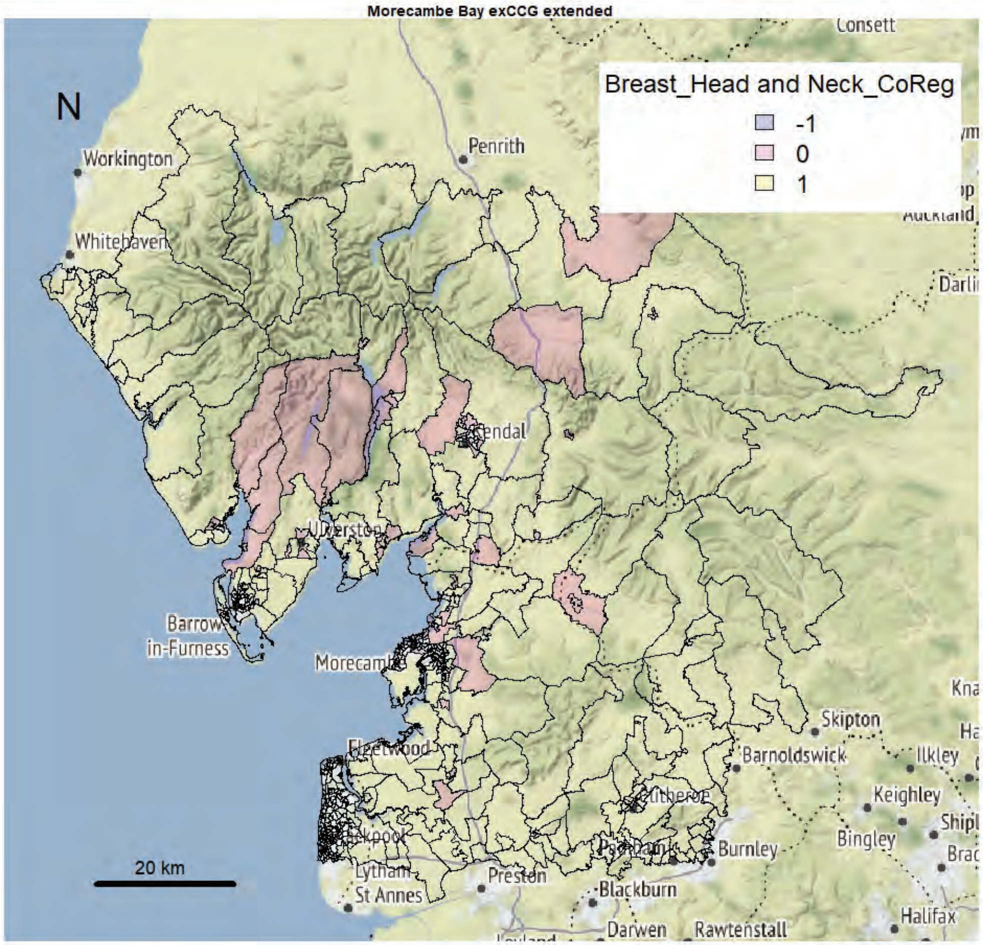
Morecambe Bay exCCG extended
N
Breast_Head and Neck_CoReg
-1
0
1
20 km
Workington
Whitehaven
Penrith
Consett
Kendal
Barrow-in-Furness
Morecambe
Fleetwood
Lytham St Annes
Preston
Blackburn
Darwen
Rawtenstall
Burnley
Barnoldswick
Skipton
Ilkley
Keighley
Bingley
Halifax

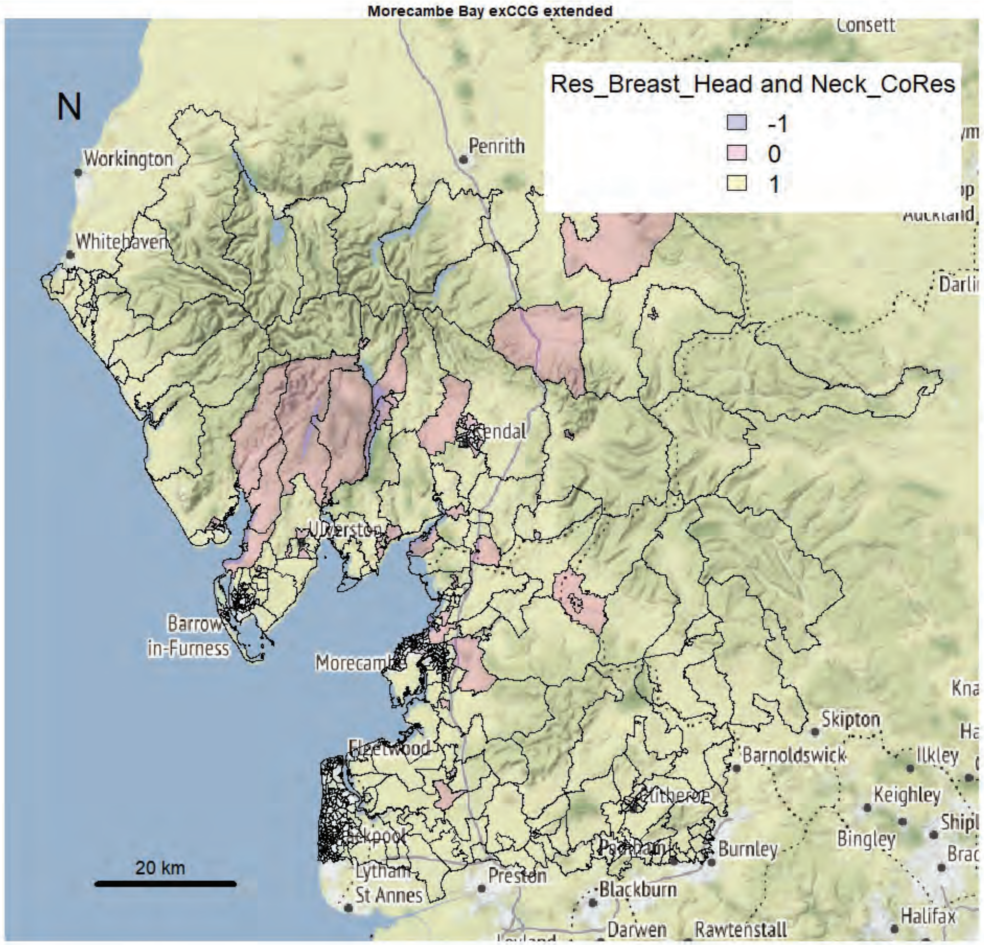
Morecambe Bay exCCG extended
N
Res_Breast_Head and Neck_CoRes
-1
0
1
20 km
Workington
Whitehaven
Penrith
Consett
Kendal
Barrow-in-Furness
Morecambe
Fleetwood
Lytham St Annes
Preston
Blackburn
Darwen
Rawtenstall
Burnley
Barnoldswick
Skipton
Ilkley
Keighley
Bingley
Halifax

Morecambe Bay exCCG extended

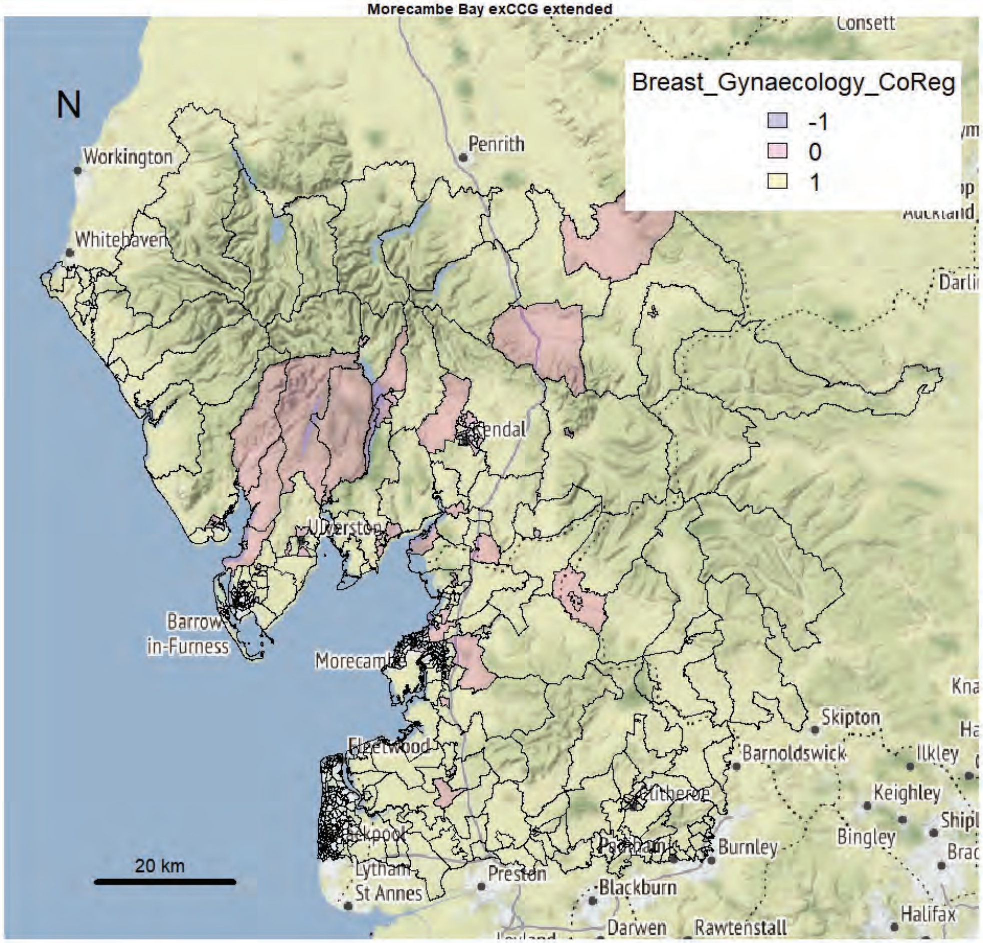


Morecambe Bay exCCG extended

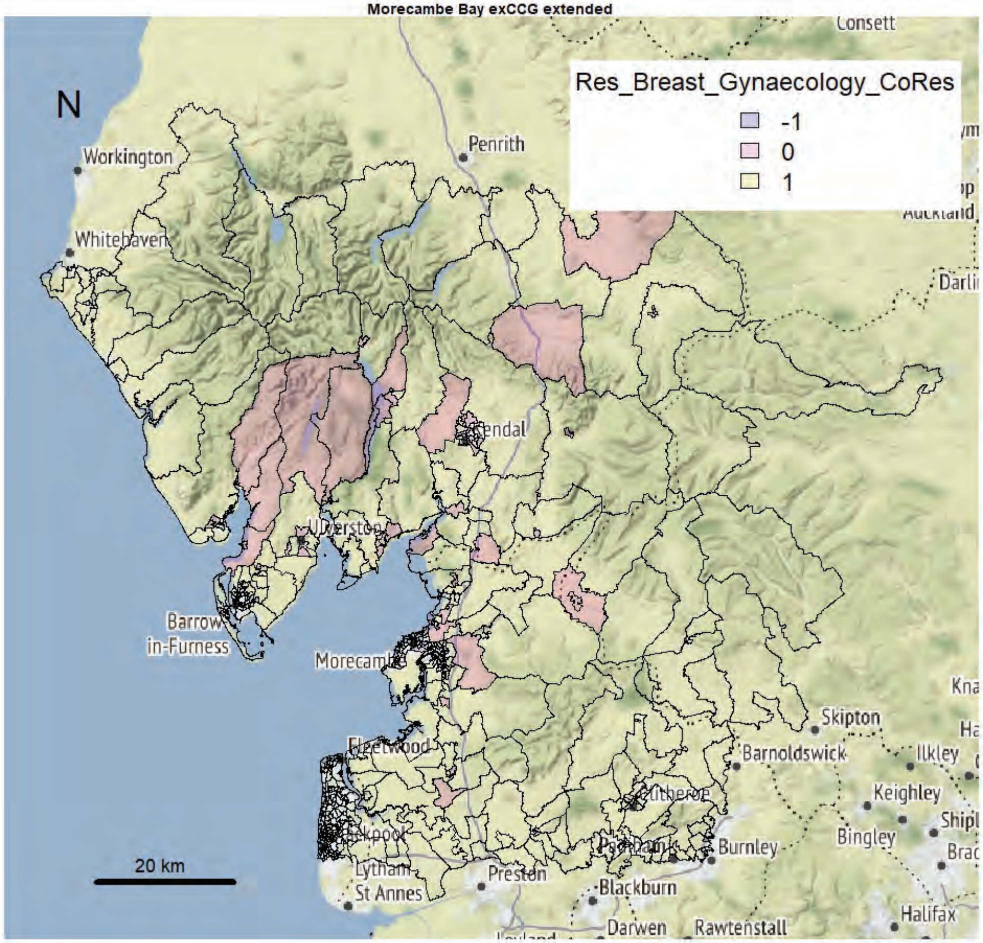

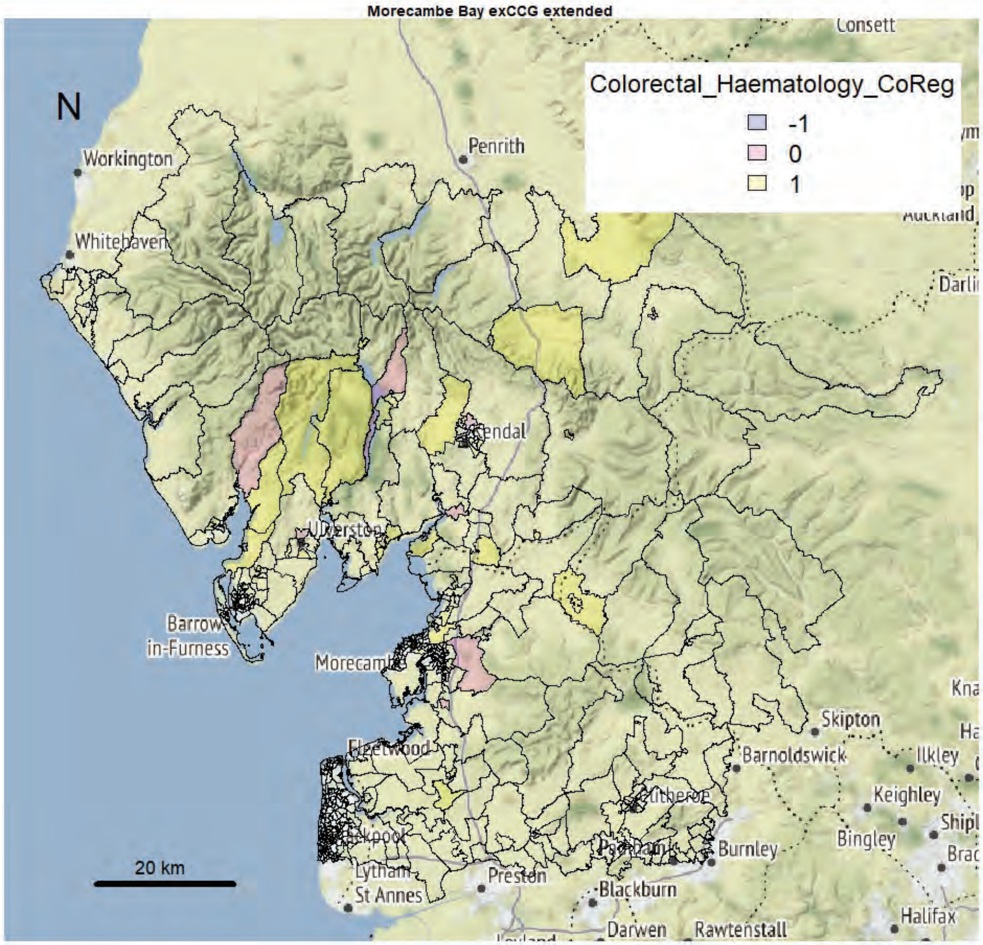
Morecambe Bay exCCG extended
N
Colorectal_Haematology_CoReg
-1
0
1
Workington
Whitehaven
Penrith
Consett
Kendal
Ulverston
Barrow-in-Furness
Morecambe
Fleetwood
Lytham St Annes
Preston
Blackburn
Darwen
Rawtenstall
Burnley
Barnoldswick
Skipton
Ilkley
Keighley
Bingley
Halifax
20 km

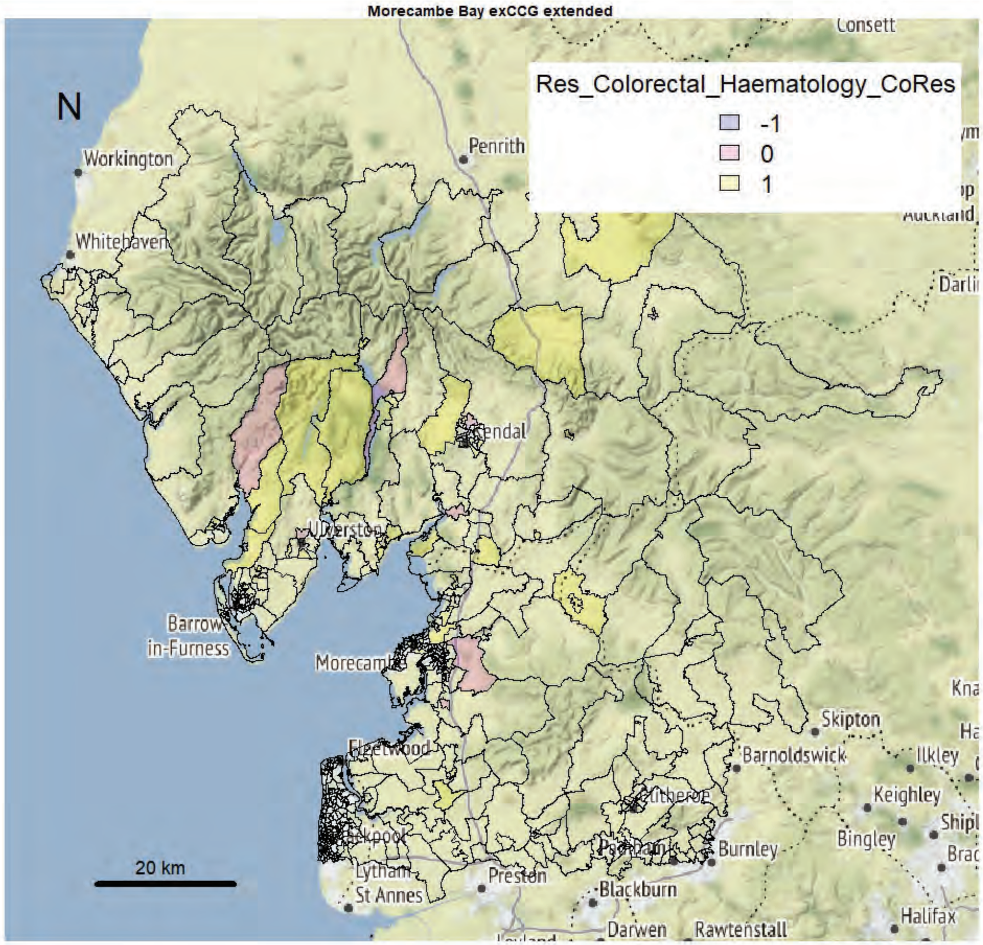
Morecambe Bay exCCG extended
N
Res_Colorectal_Haematology_CoRes
-1
0
1
Workington
Whitehaven
Penrith
Consett
Kendal
Ulverston
Barrow-in-Furness
Morecambe
Fleetwood
Lytham St Annes
Preston
Blackburn
Darwen
Rawtenstall
Burnley
Barnoldswick
Skipton
Ilkley
Keighley
Bingley
Halifax
20 km

Morecambe Bay exCCG extended

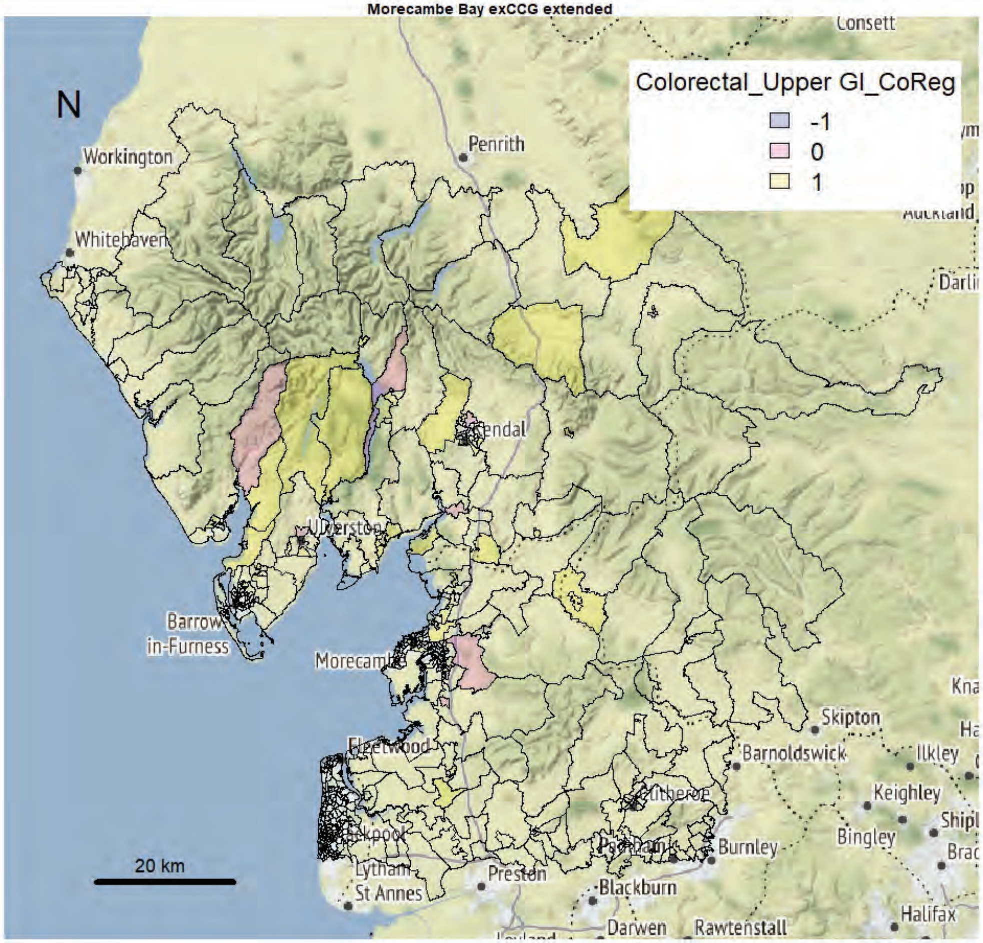


Morecambe Bay exCCG extended

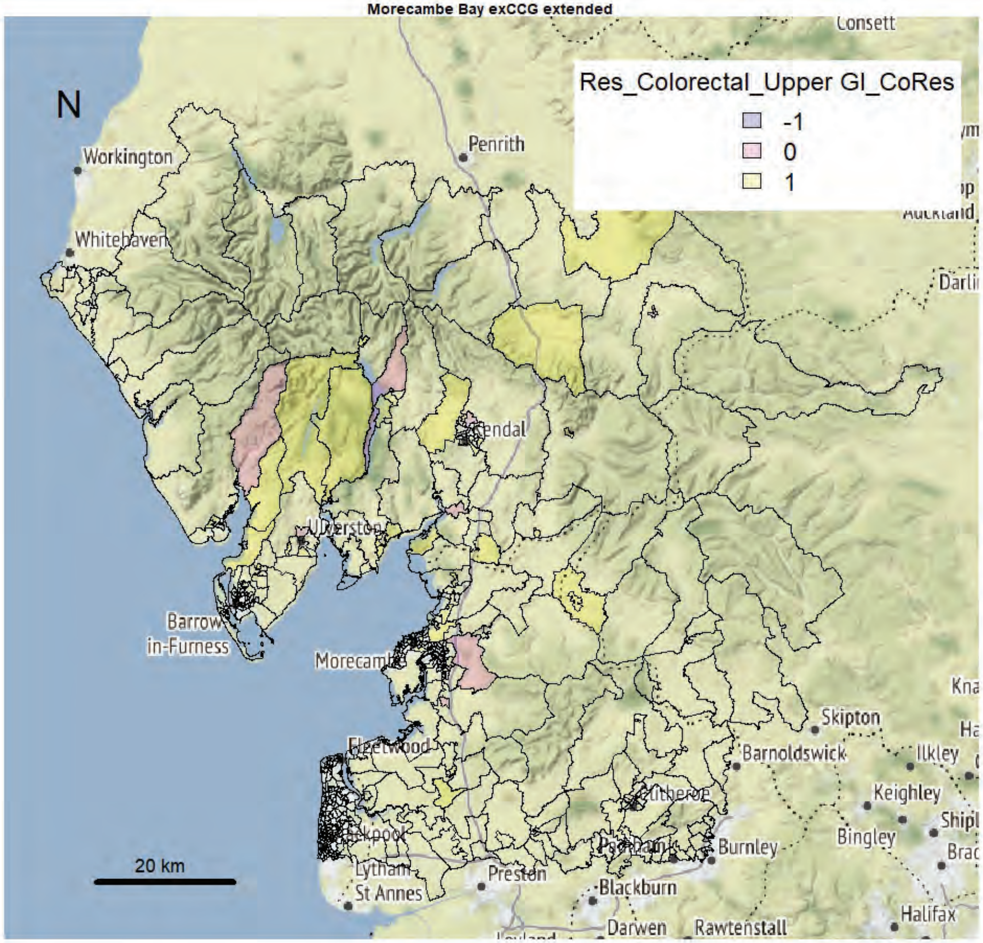

Morecambe Bay exCCG extended

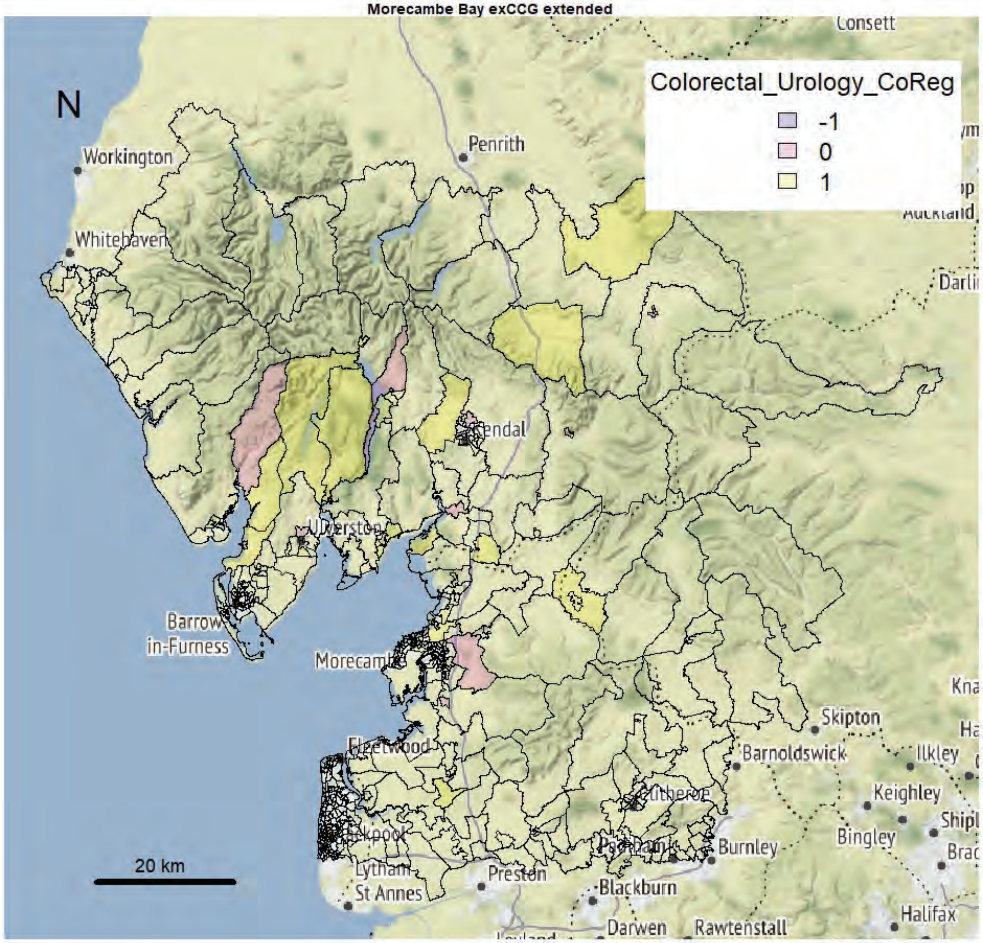


Morecambe Bay exCCG extended

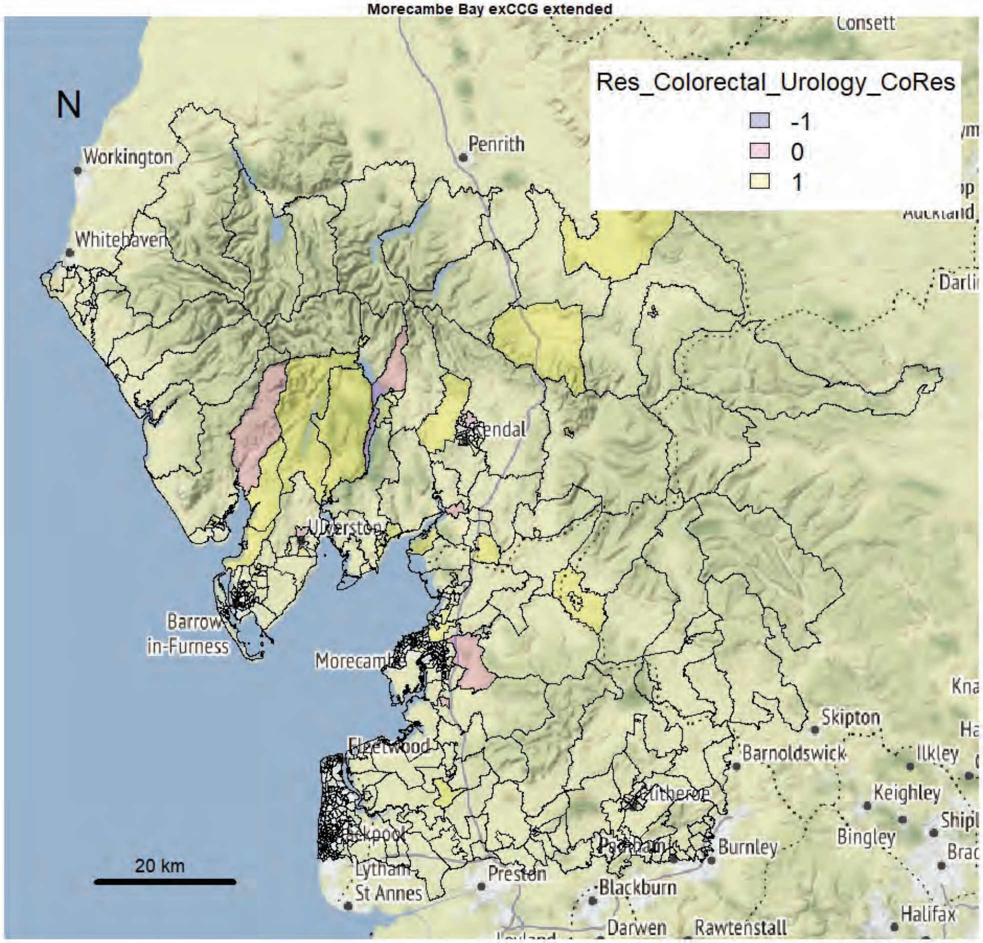

Morecambe Bay exCCG extended

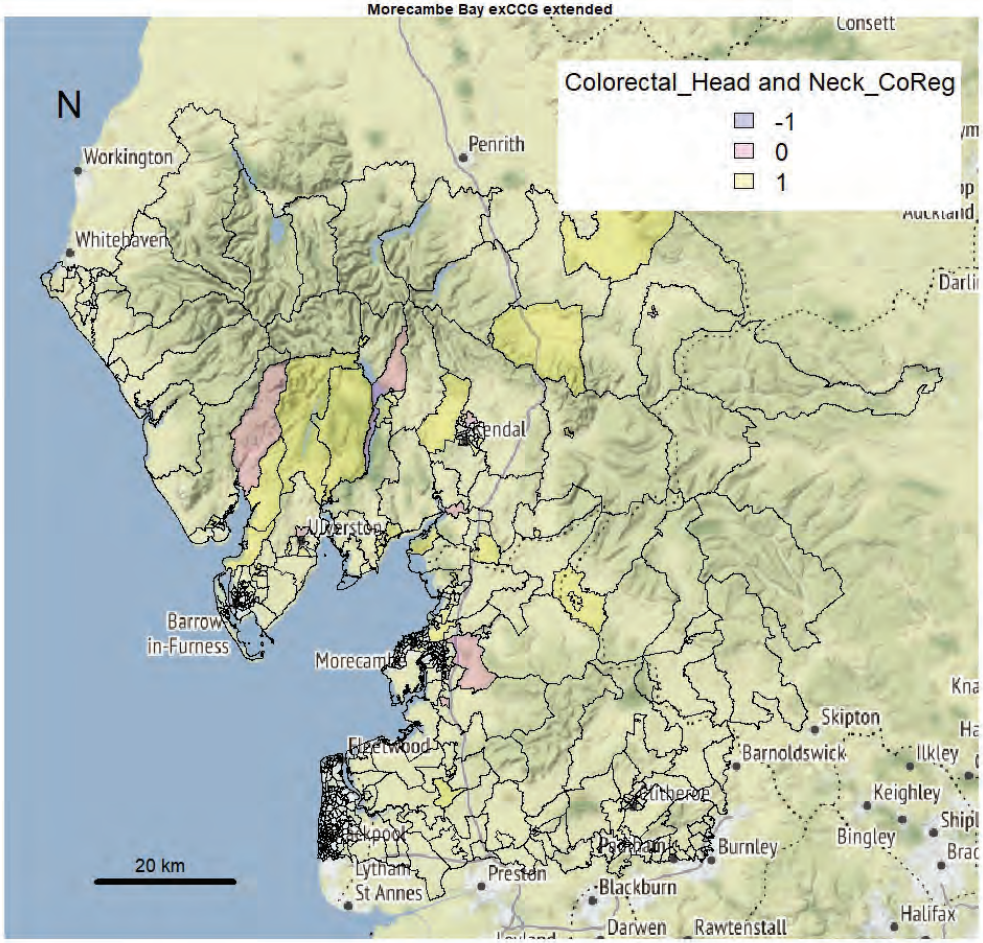


Morecambe Bay exCCG extended

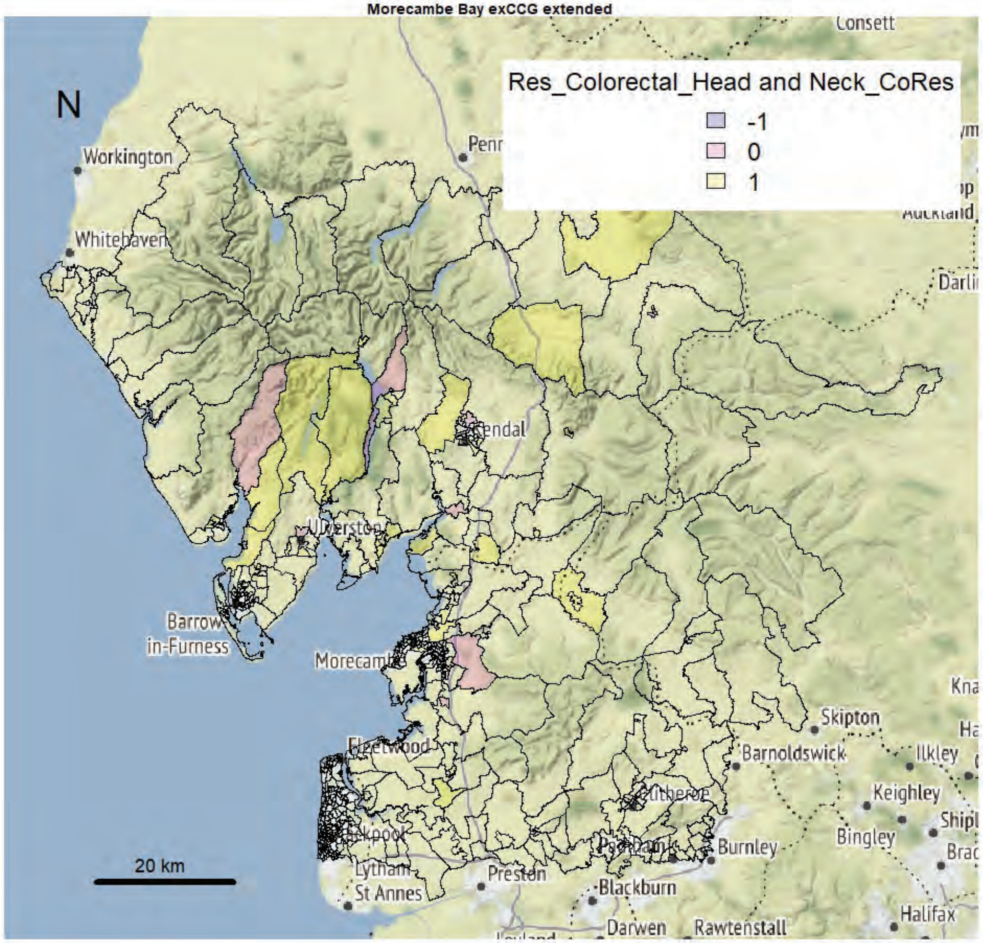

Morecambe Bay exCCG extended

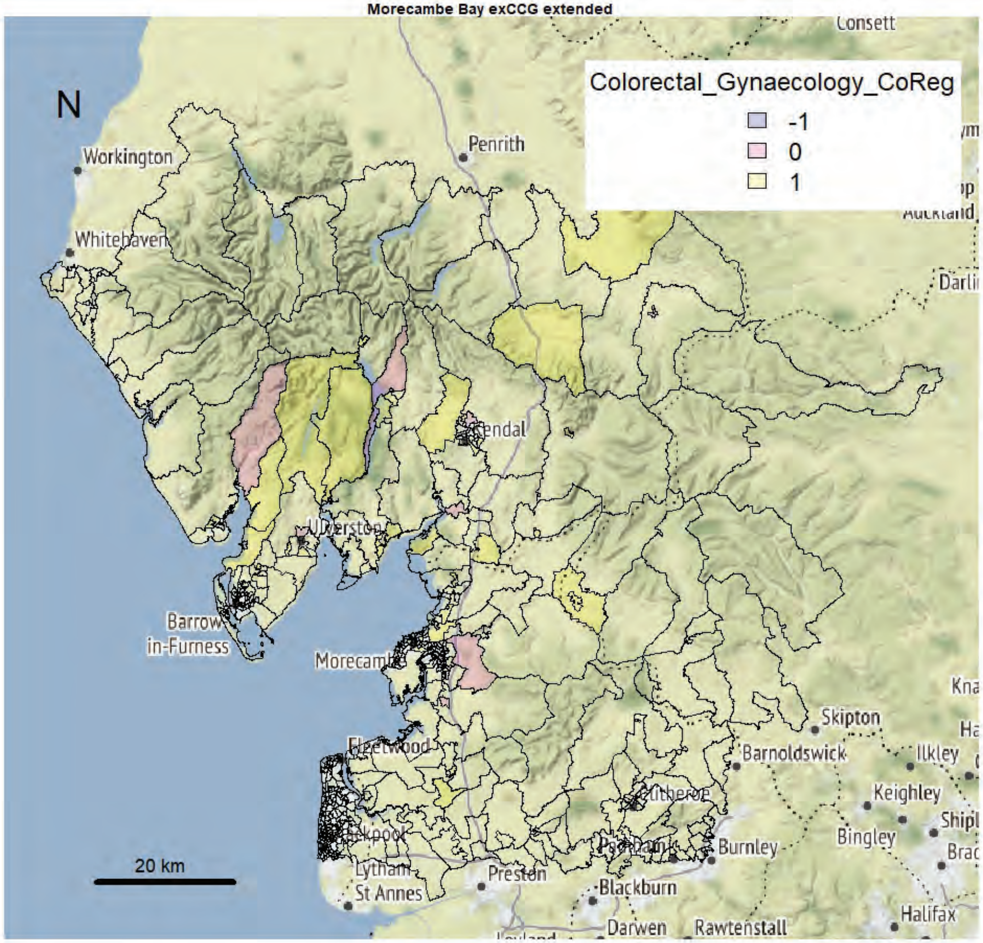


Morecambe Bay exCCG extended

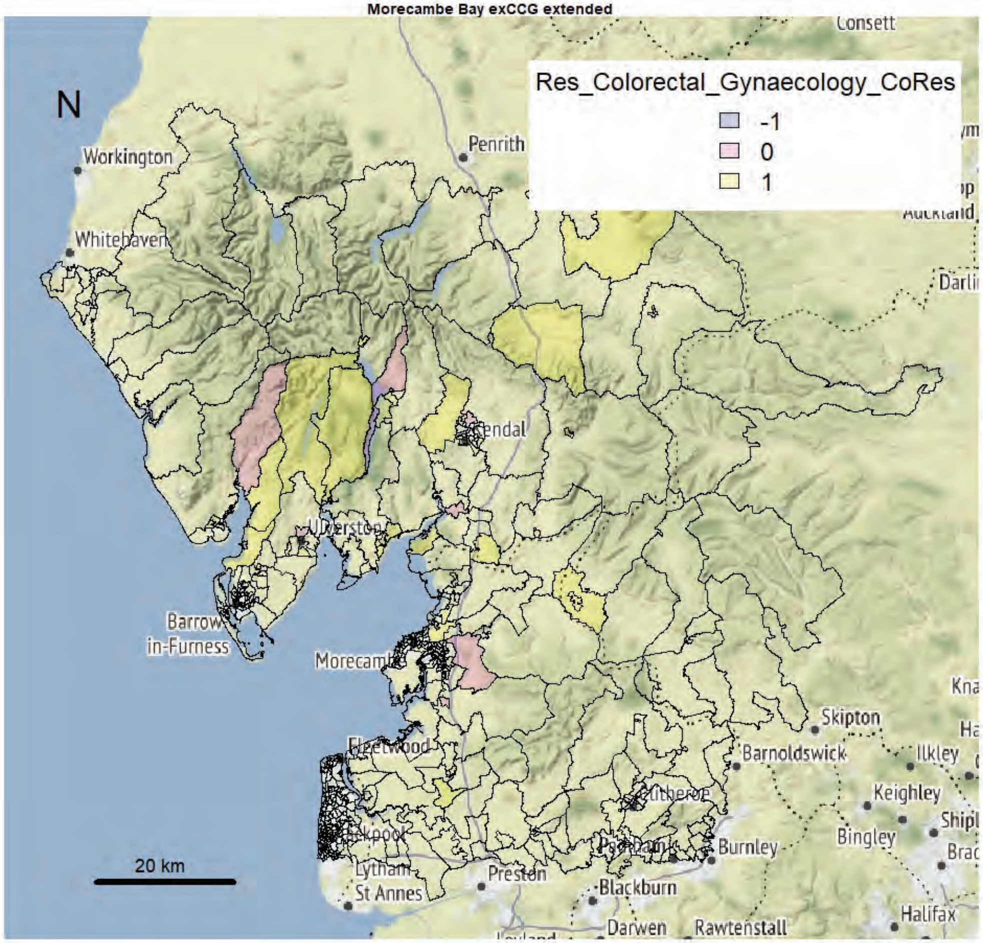

Morecambe Bay exCCG extended

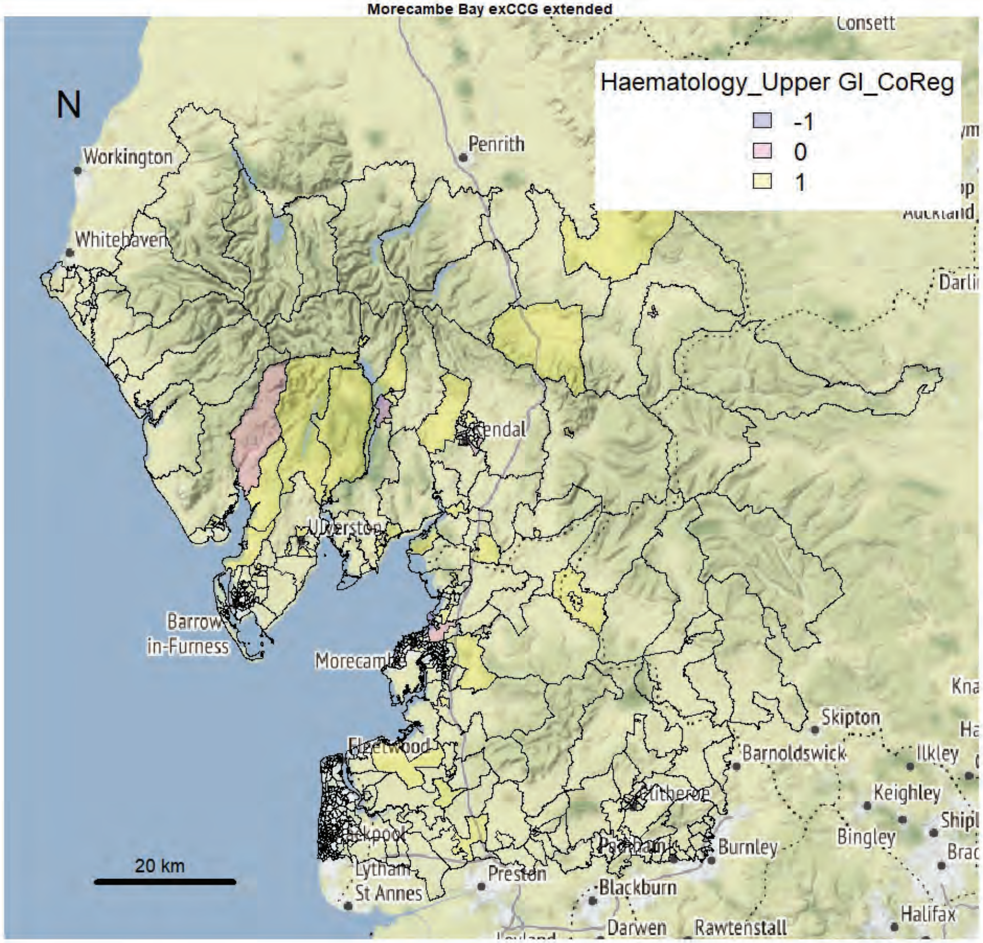


Morecambe Bay exCCG extended

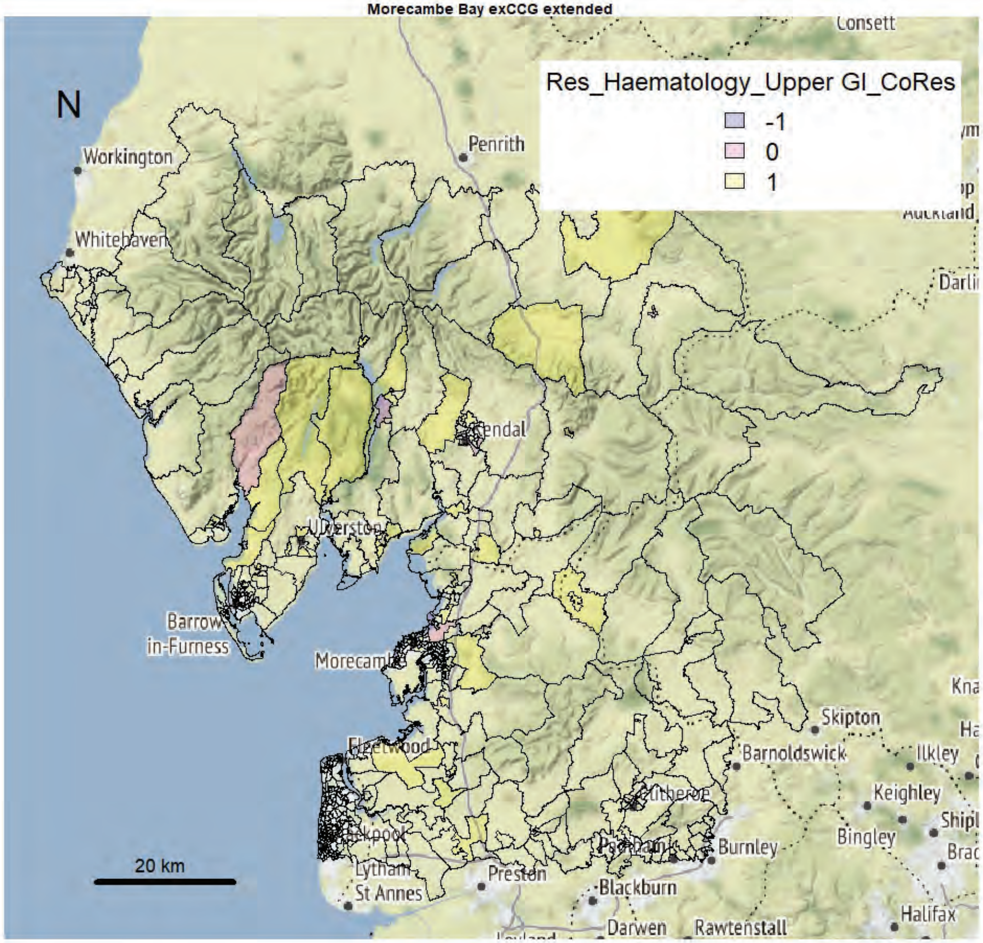

Morecambe Bay exCCG extended

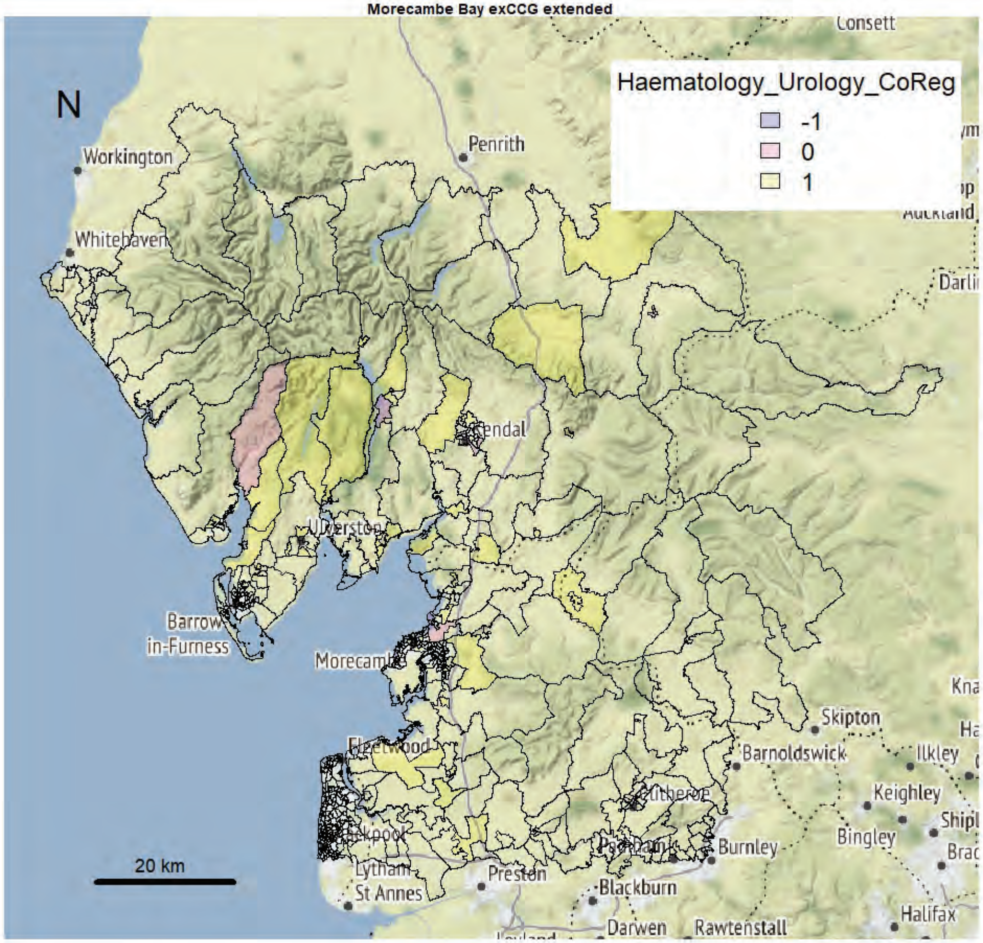


Morecambe Bay exCCG extended

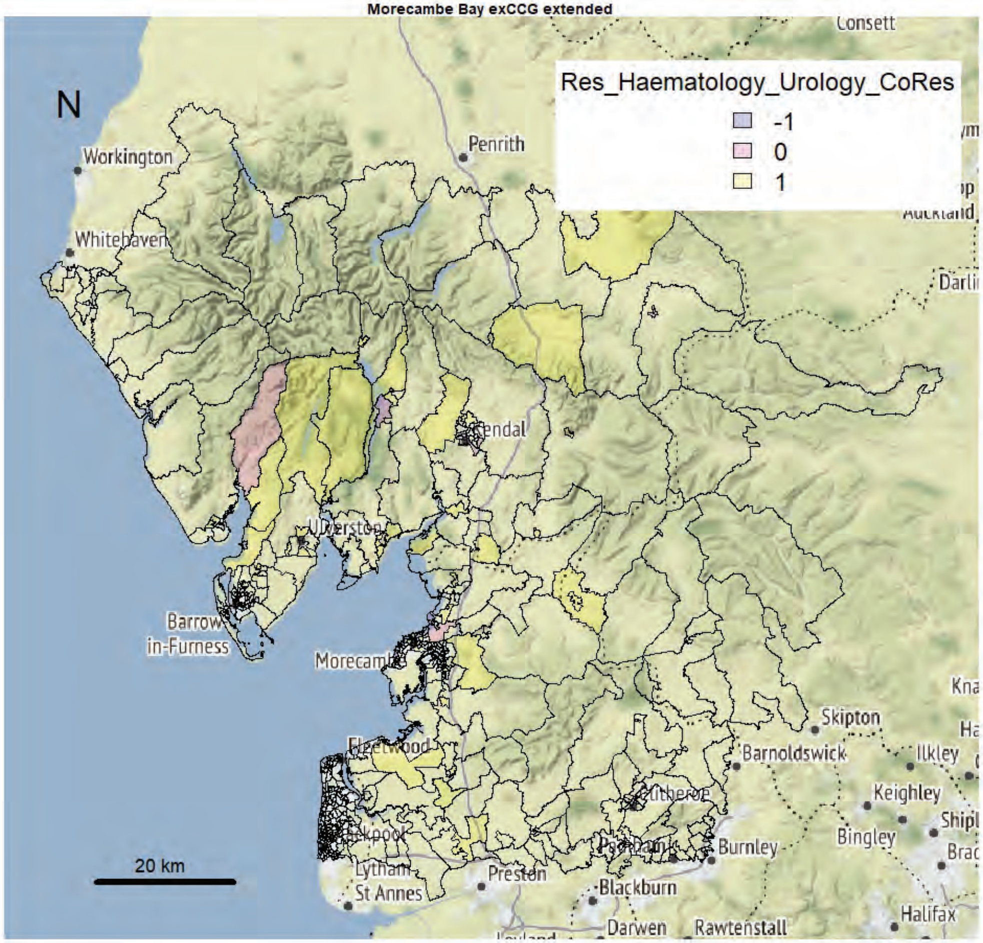

Morecambe Bay exCCG extended

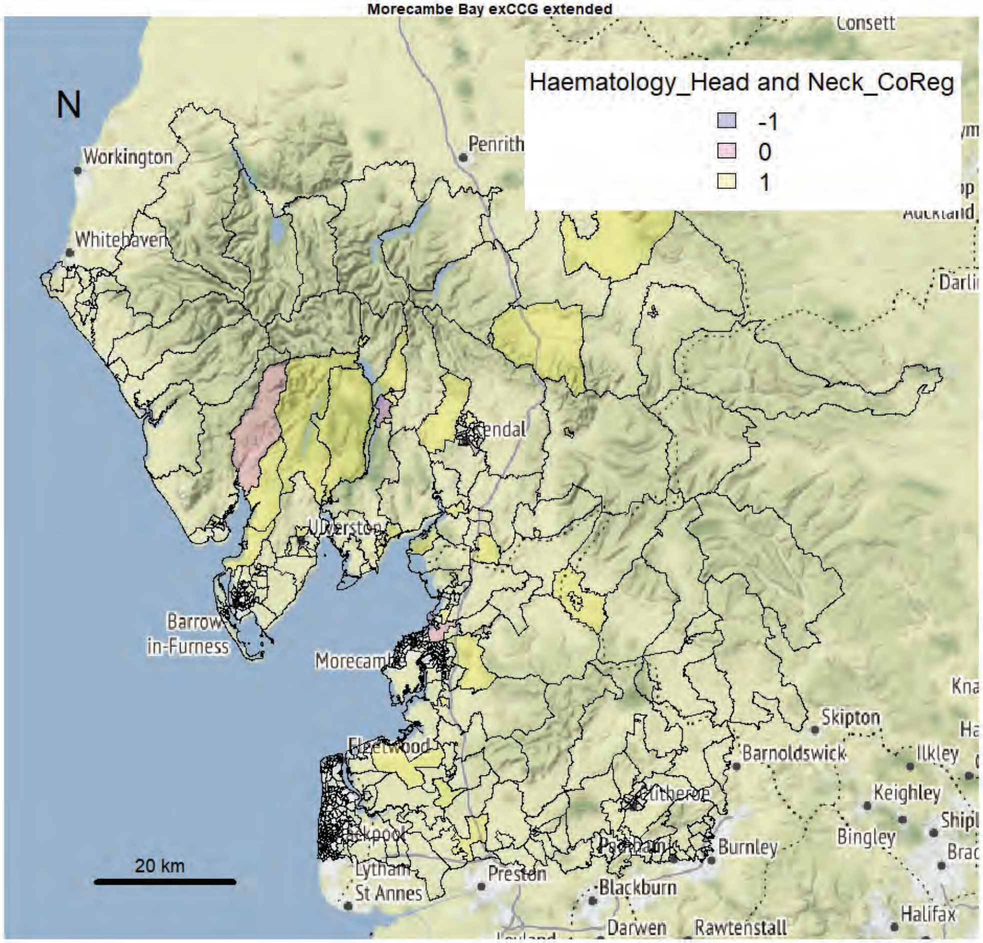


Morecambe Bay exCCG extended

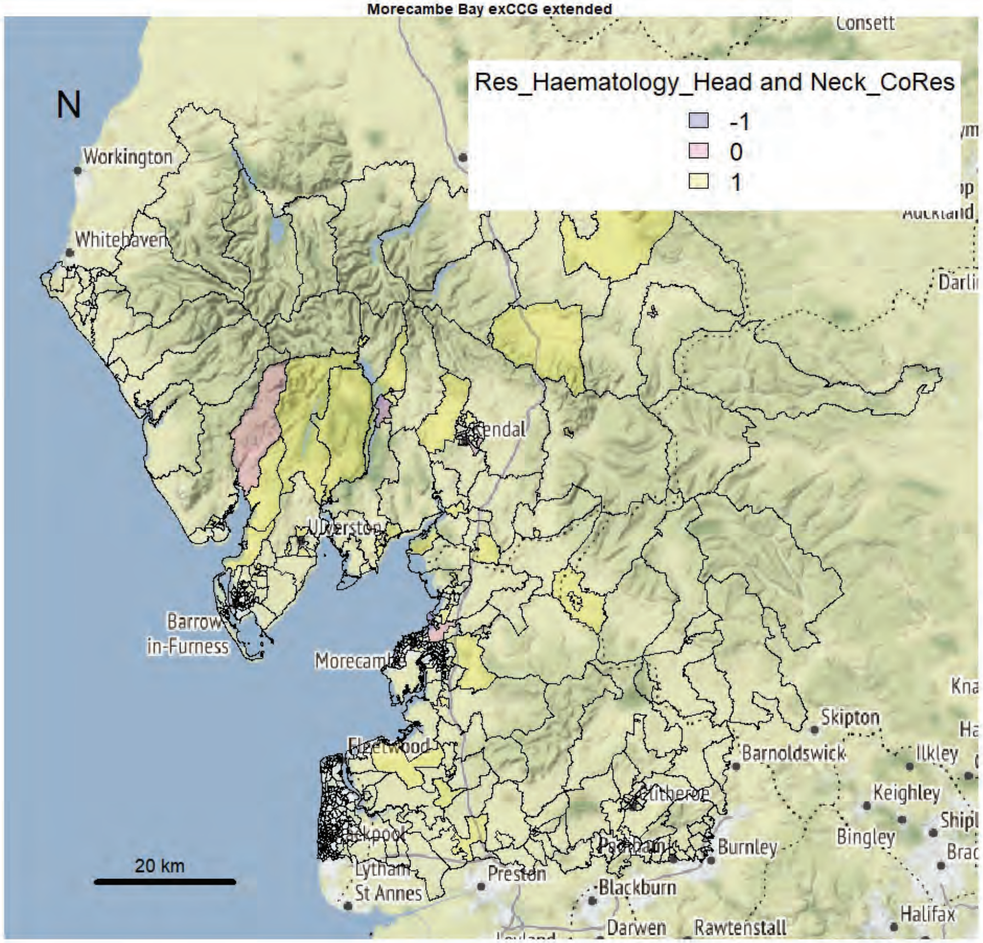

Morecambe Bay exCCG extended

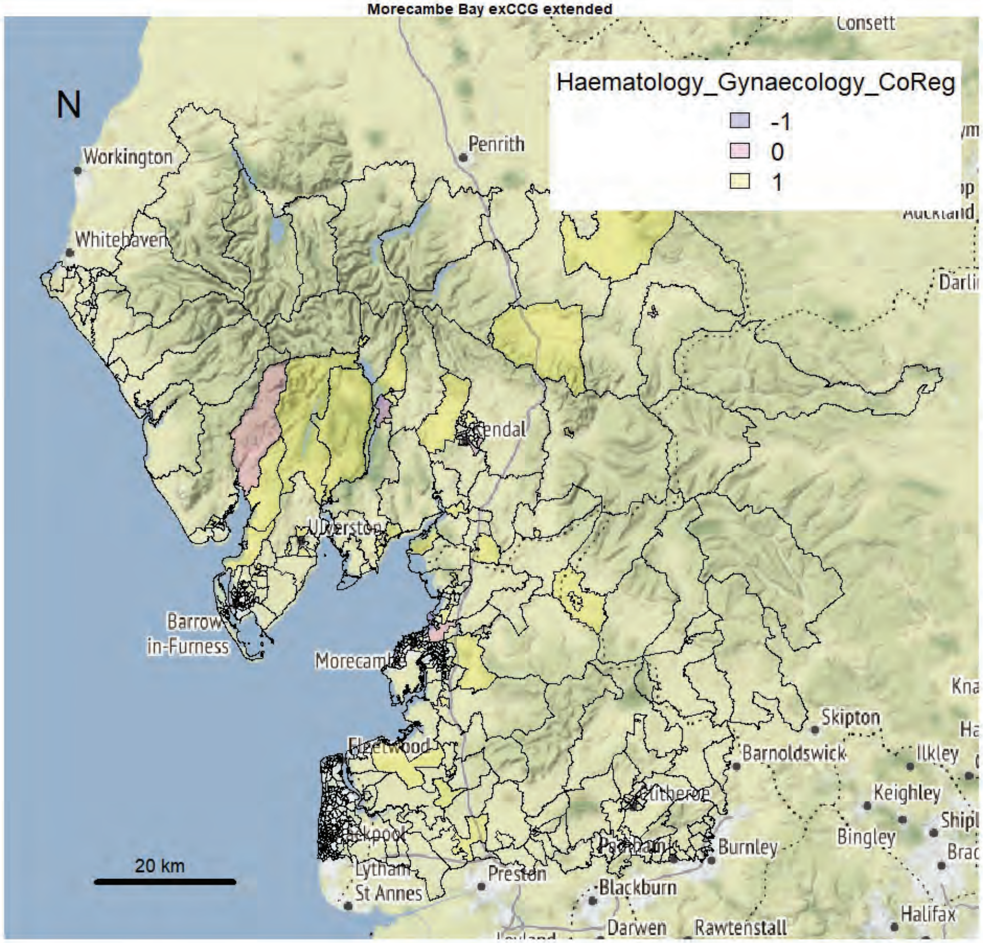


Morecambe Bay exCCG extended

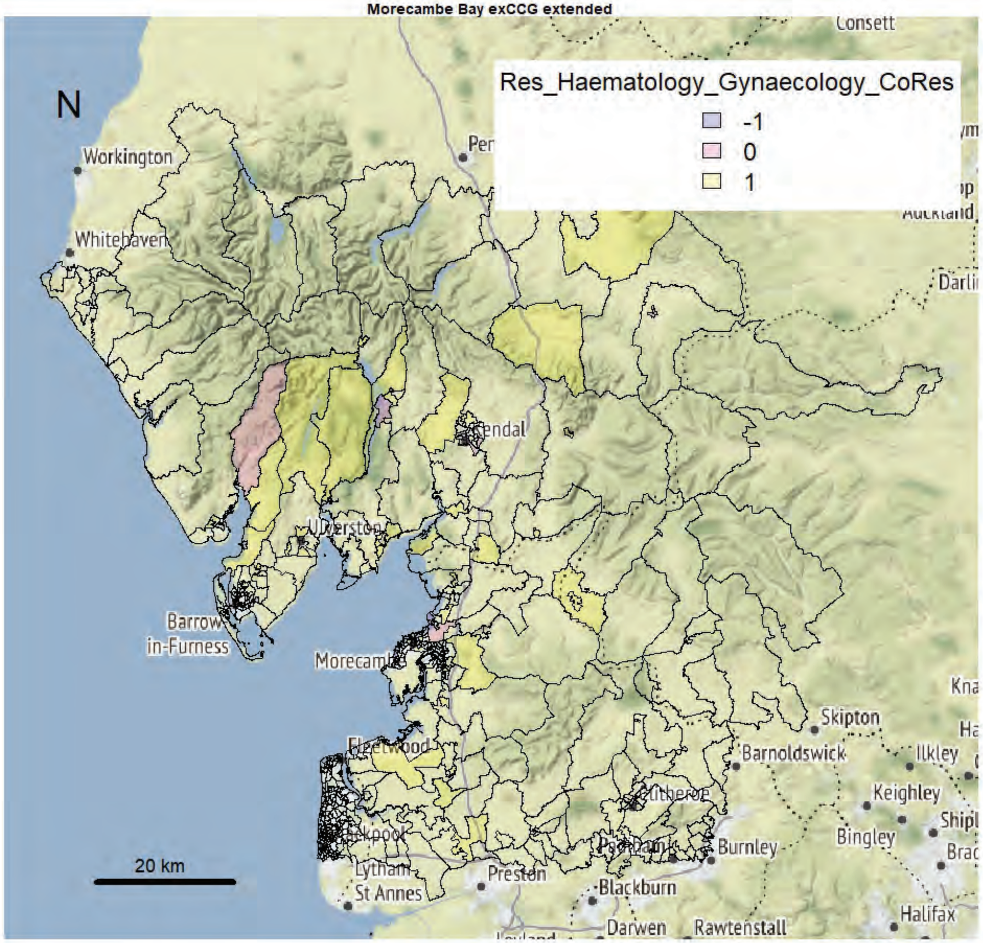

Morecambe Bay exCCG extended

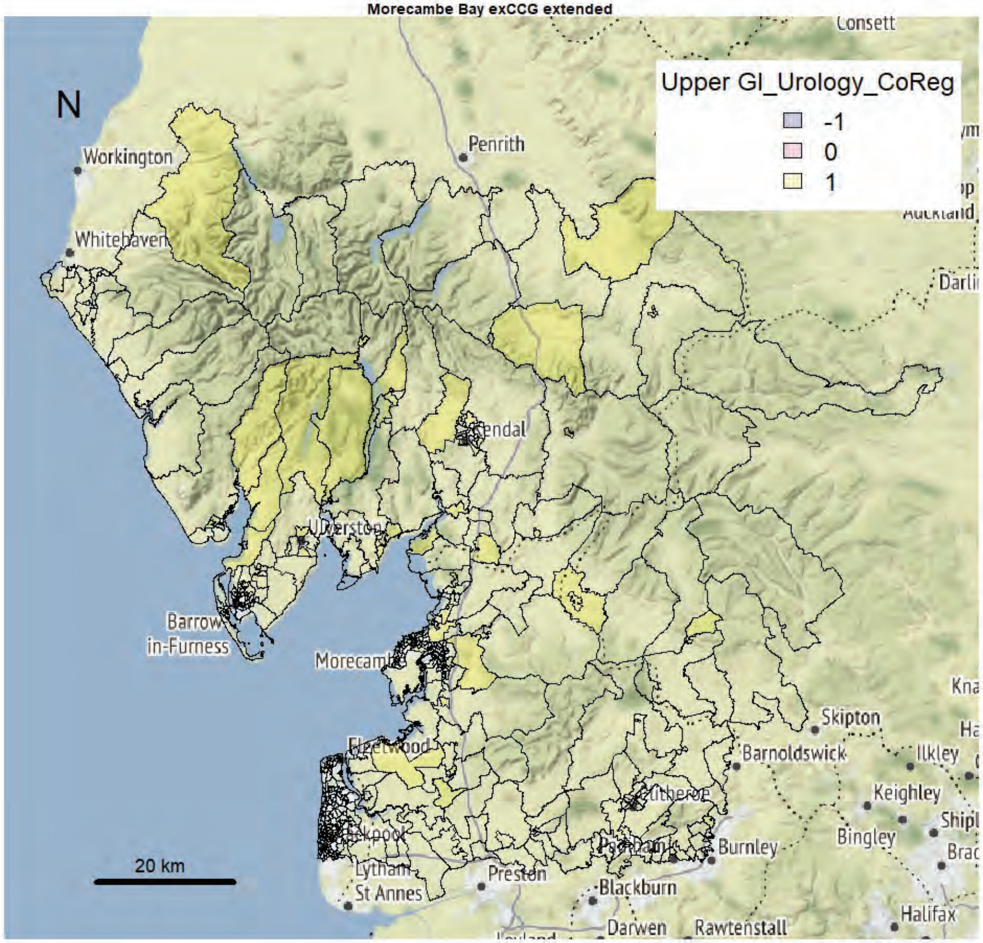


Morecambe Bay exCCG extended

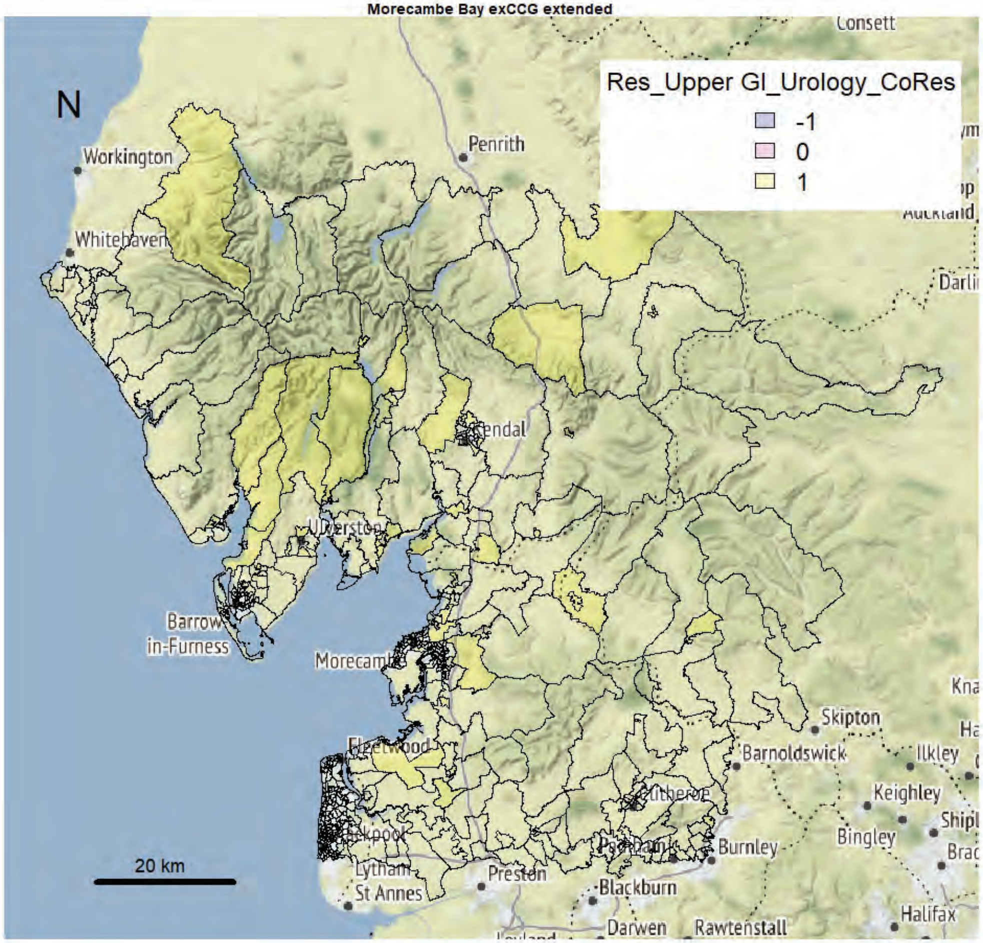

Morecambe Bay exCCG extended

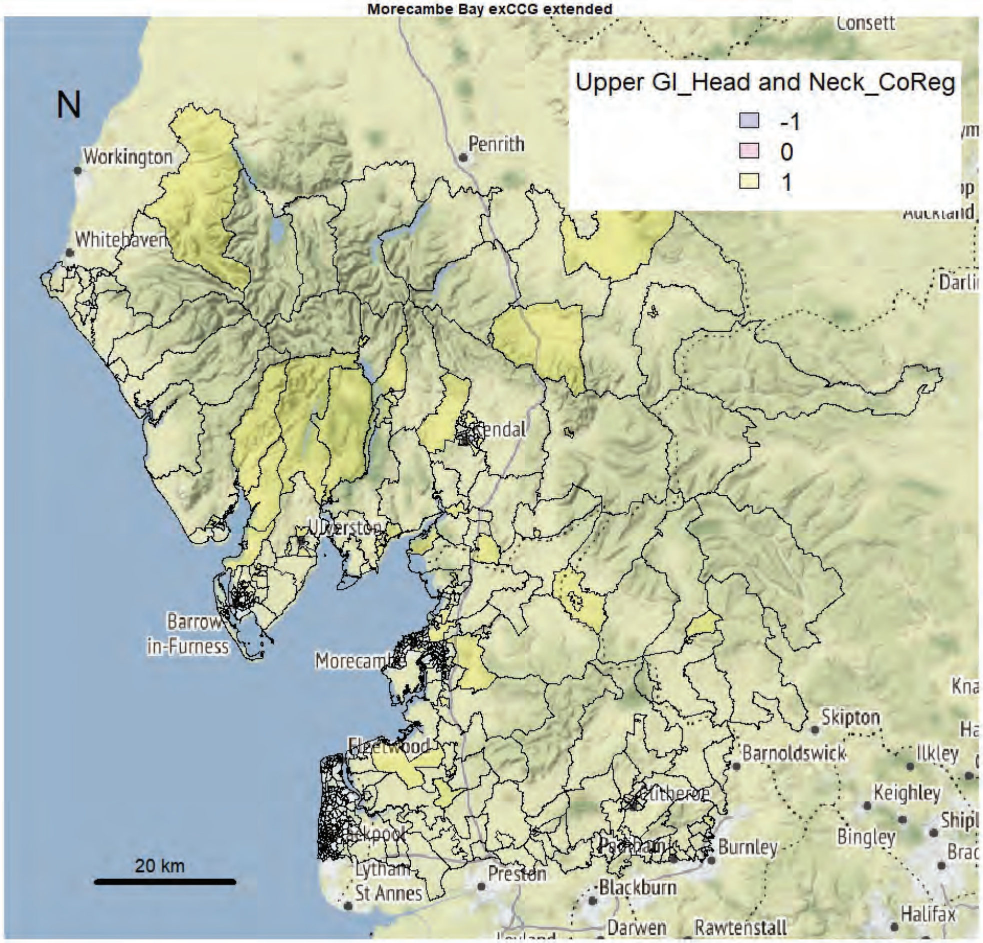


Morecambe Bay exCCG extended

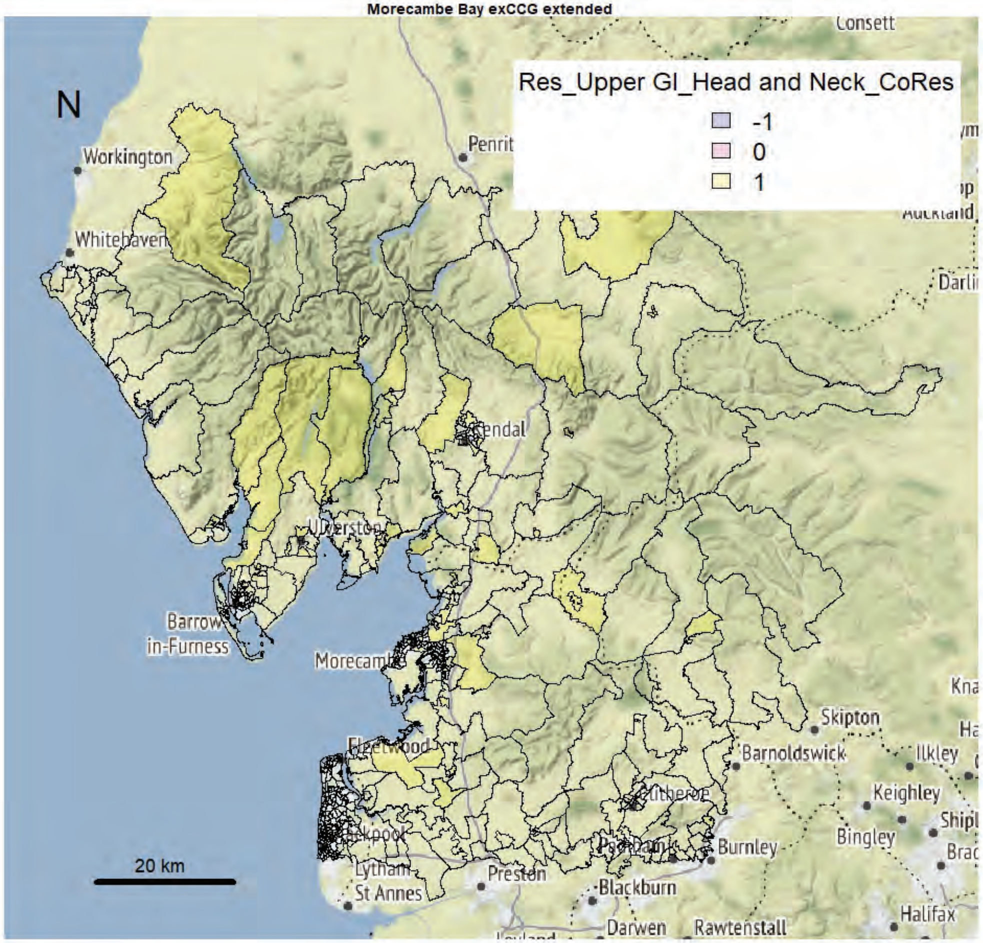

Morecambe Bay exCCG extended

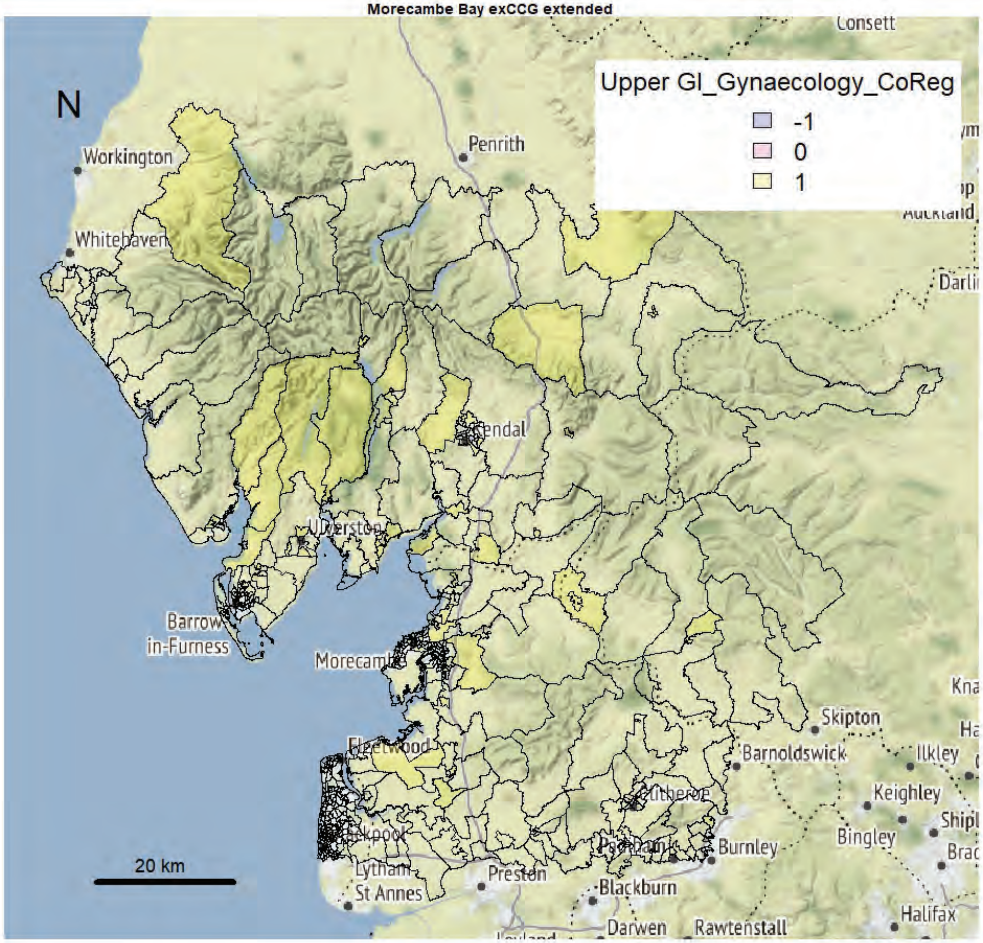


Morecambe Bay exCCG extended

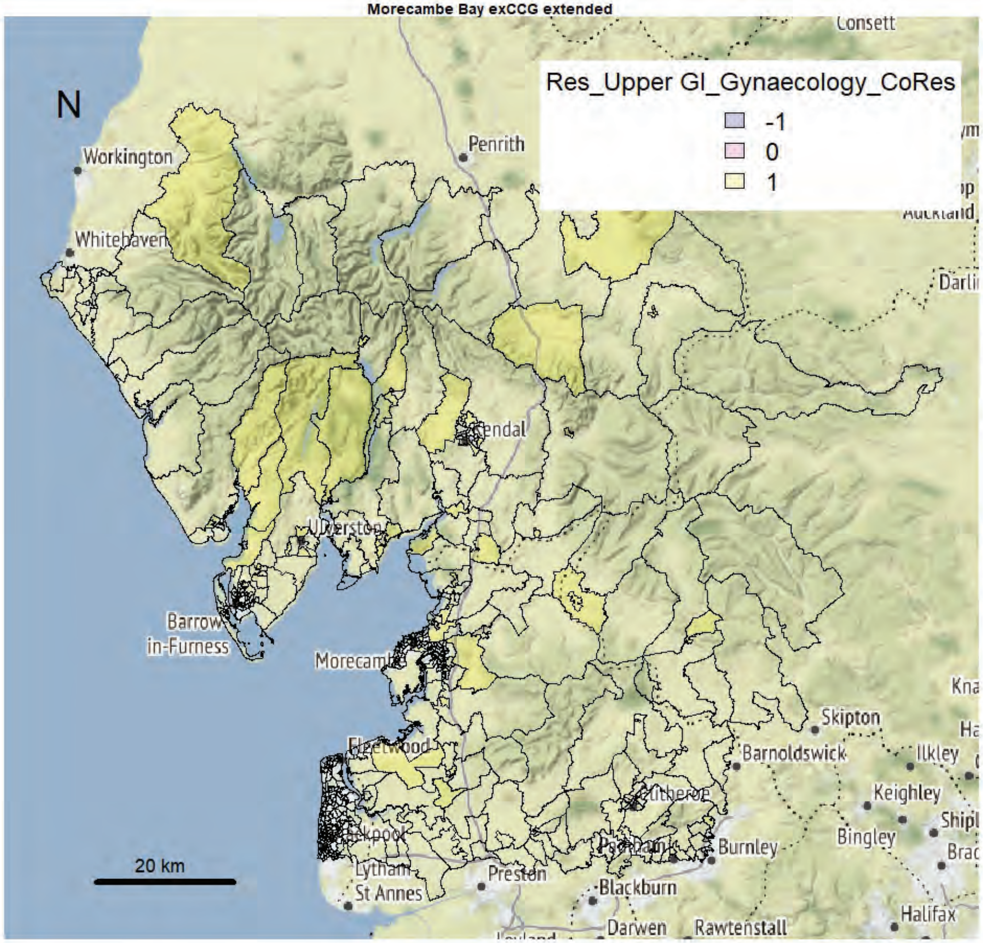

Morecambe Bay exCCG extended

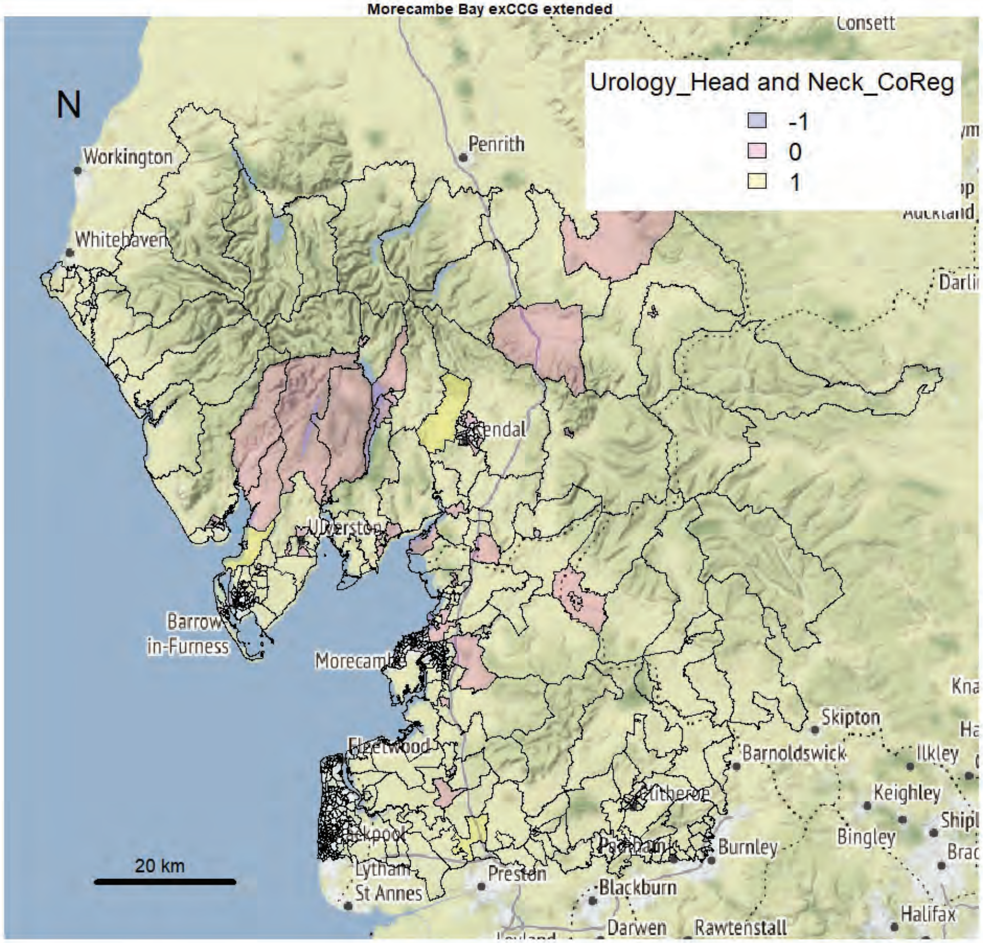


Morecambe Bay exCCG extended

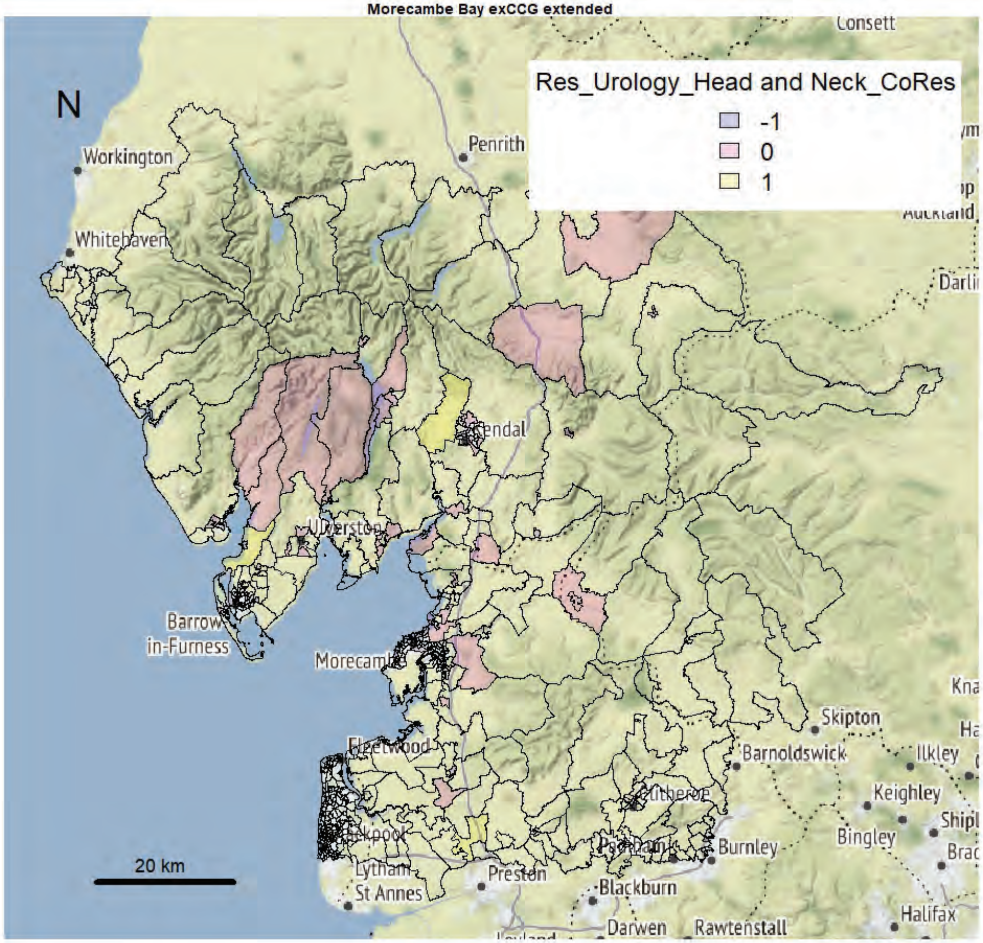

Morecambe Bay exCCG extended

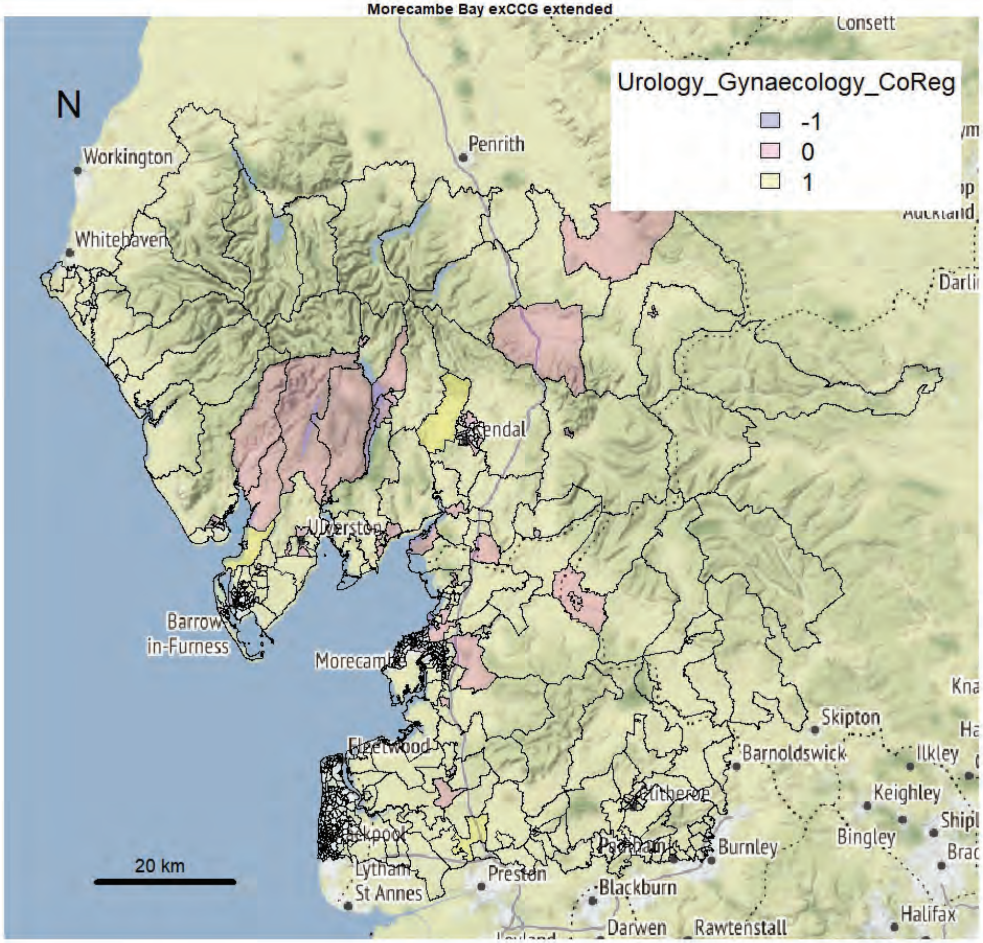


Morecambe Bay exCCG extended

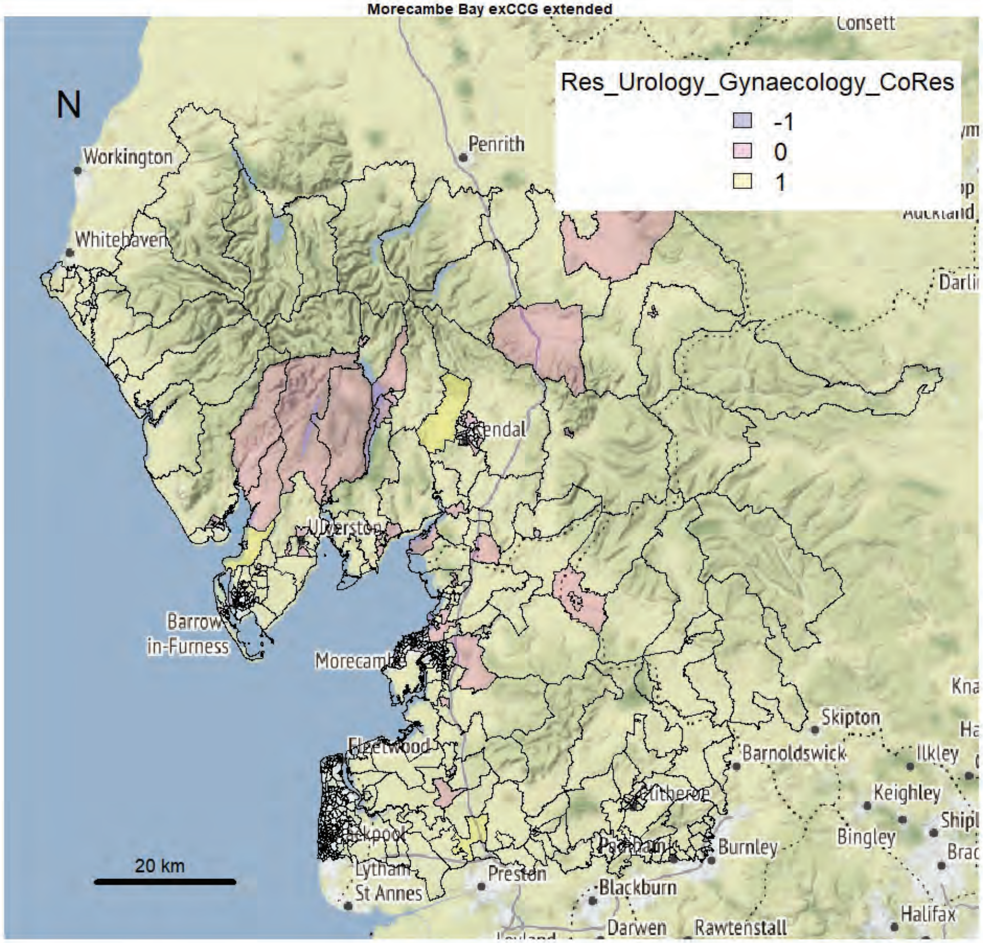

Morecambe Bay exCCG extended

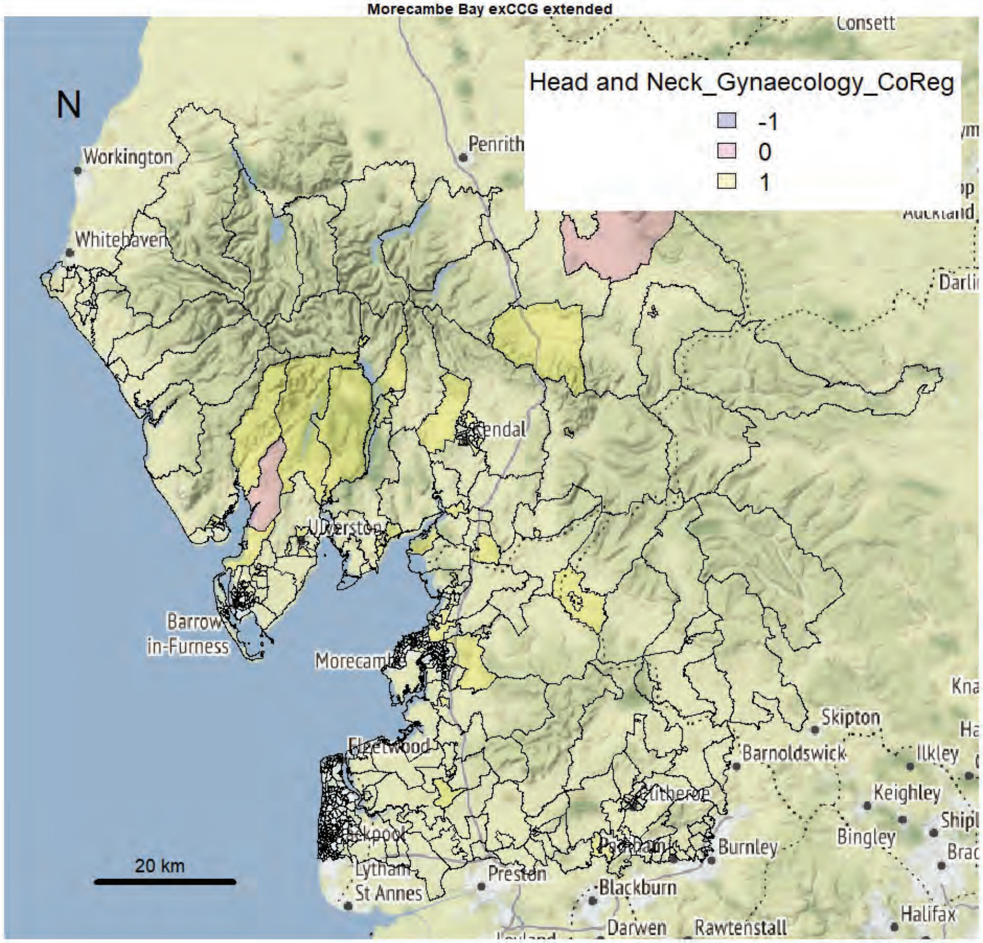


Morecambe Bay exCCG extended

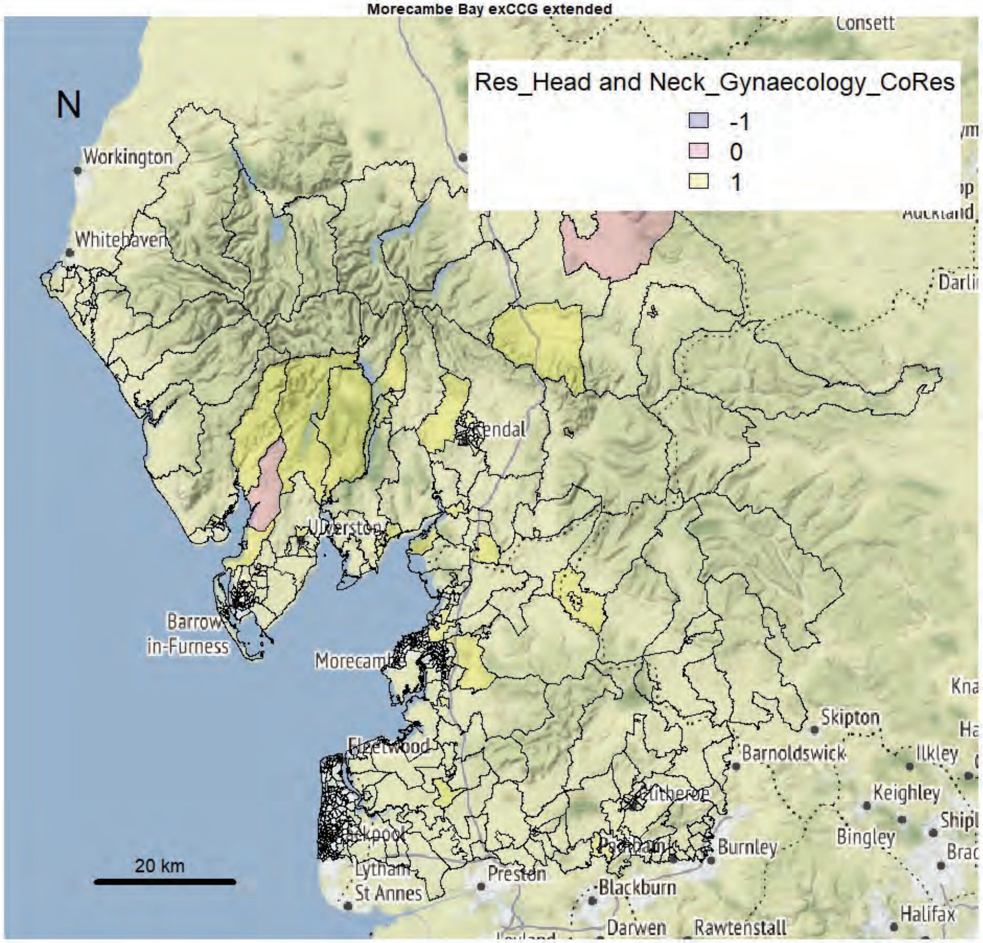

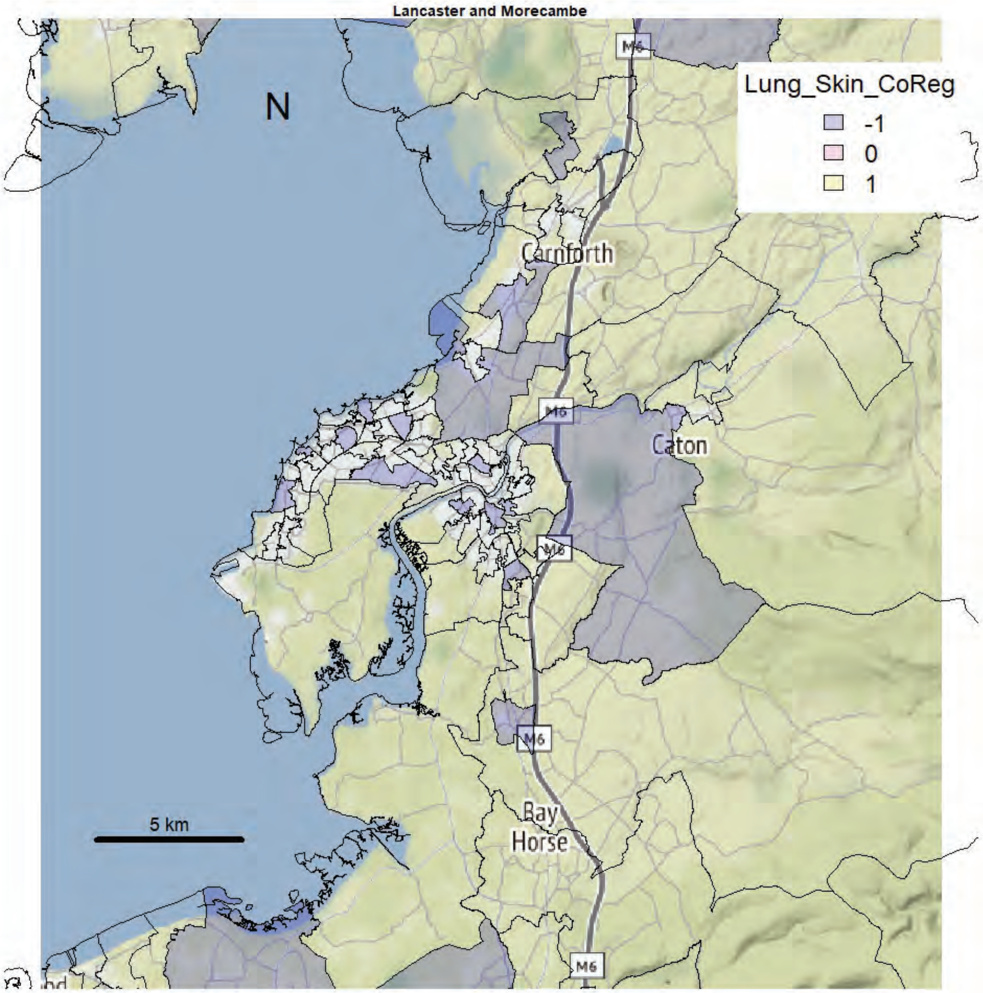
Lancaster and Morecambe
N
Lung_Skin_CoReg
-1
0
1
M6
Carnforth
Caton
Bay
Horse
5 km

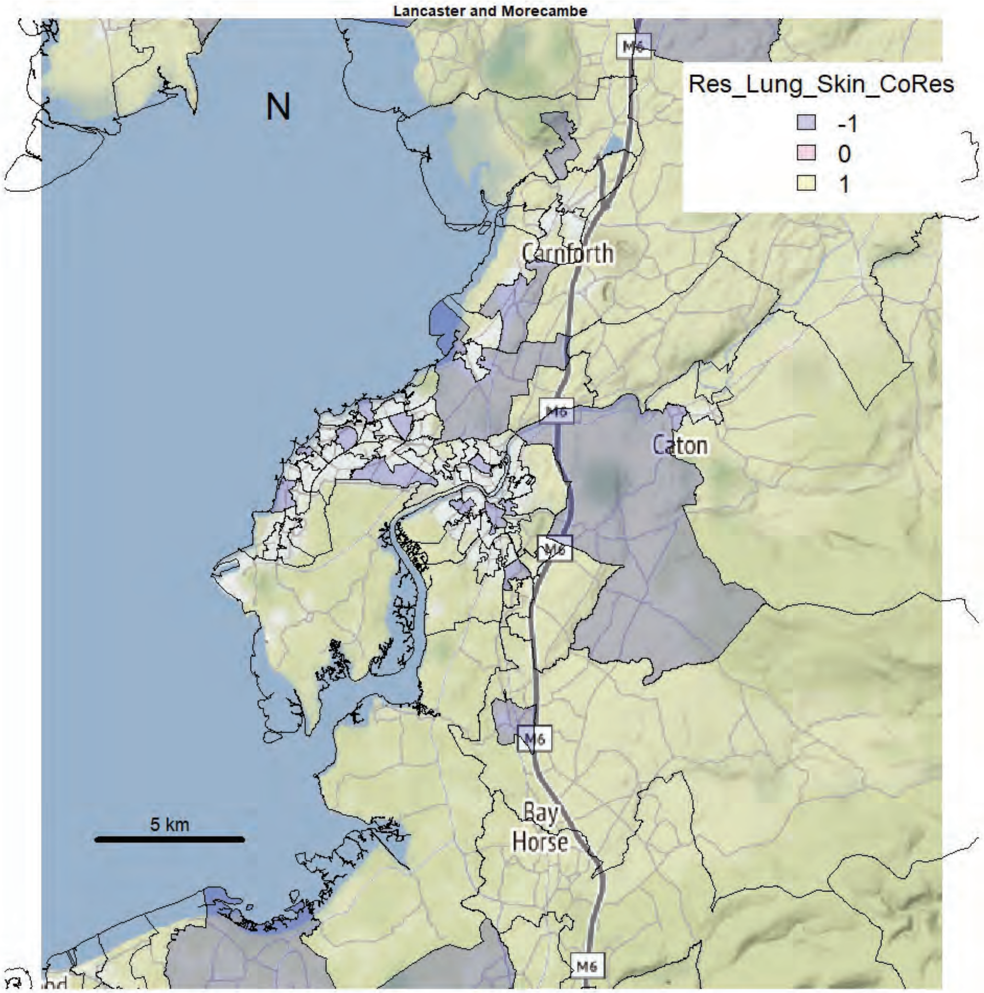
Lancaster and Morecambe
N
Res_Lung_Skin_CoRes
-1
0
1
M6
Carnforth
Caton
Bay
Horse
5 km

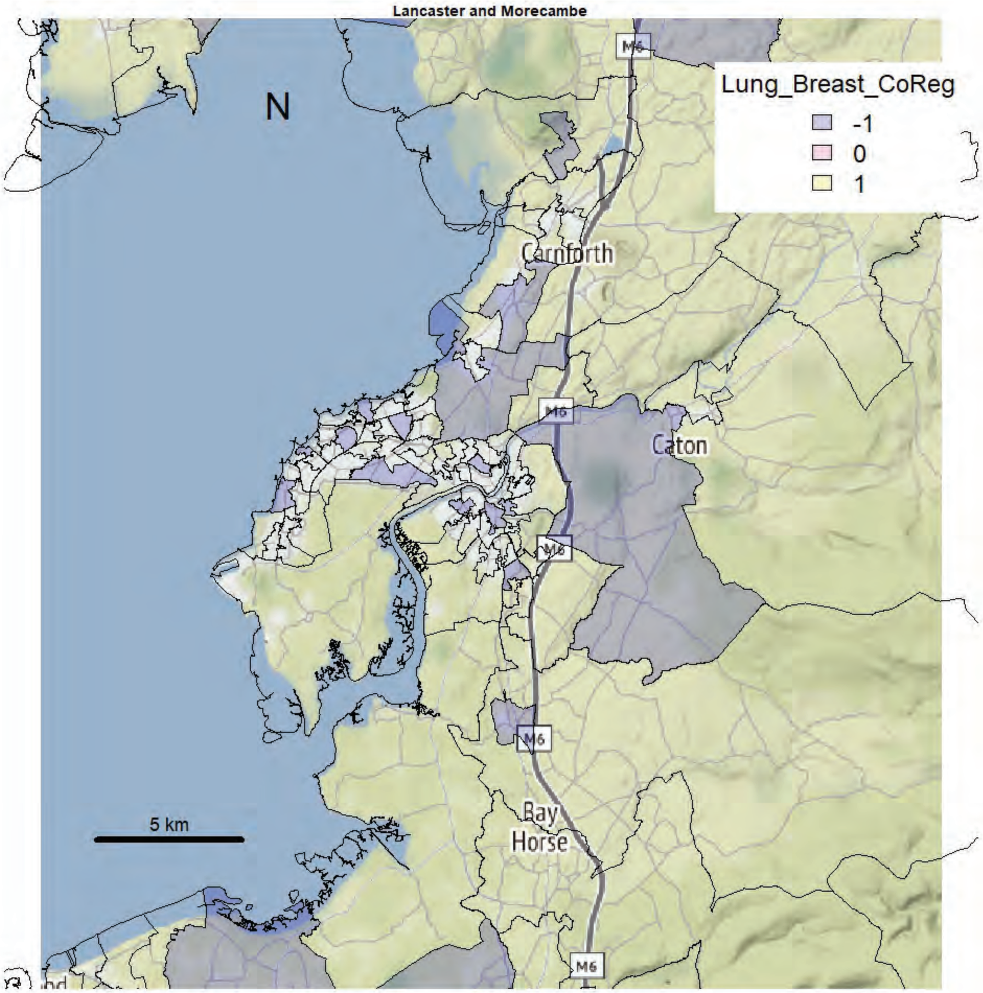
Lancaster and Morecambe
N
Lung_Breast_CoReg
-1
0
1
Carnforth
Caton
M6
Bay Horse
5 km

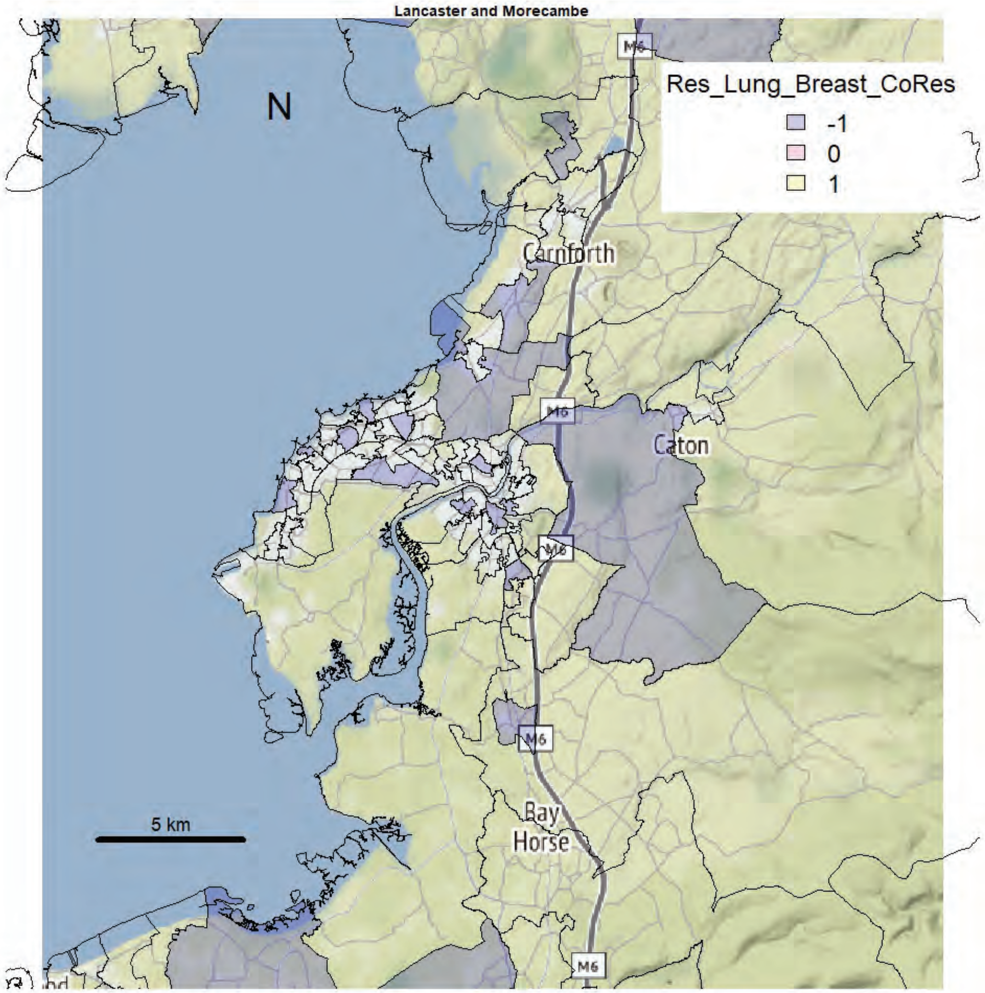
Lancaster and Morecambe
N
Res_Lung_Breast_CoRes
-1
0
1
Carnforth
Caton
M6
Bay Horse
5 km

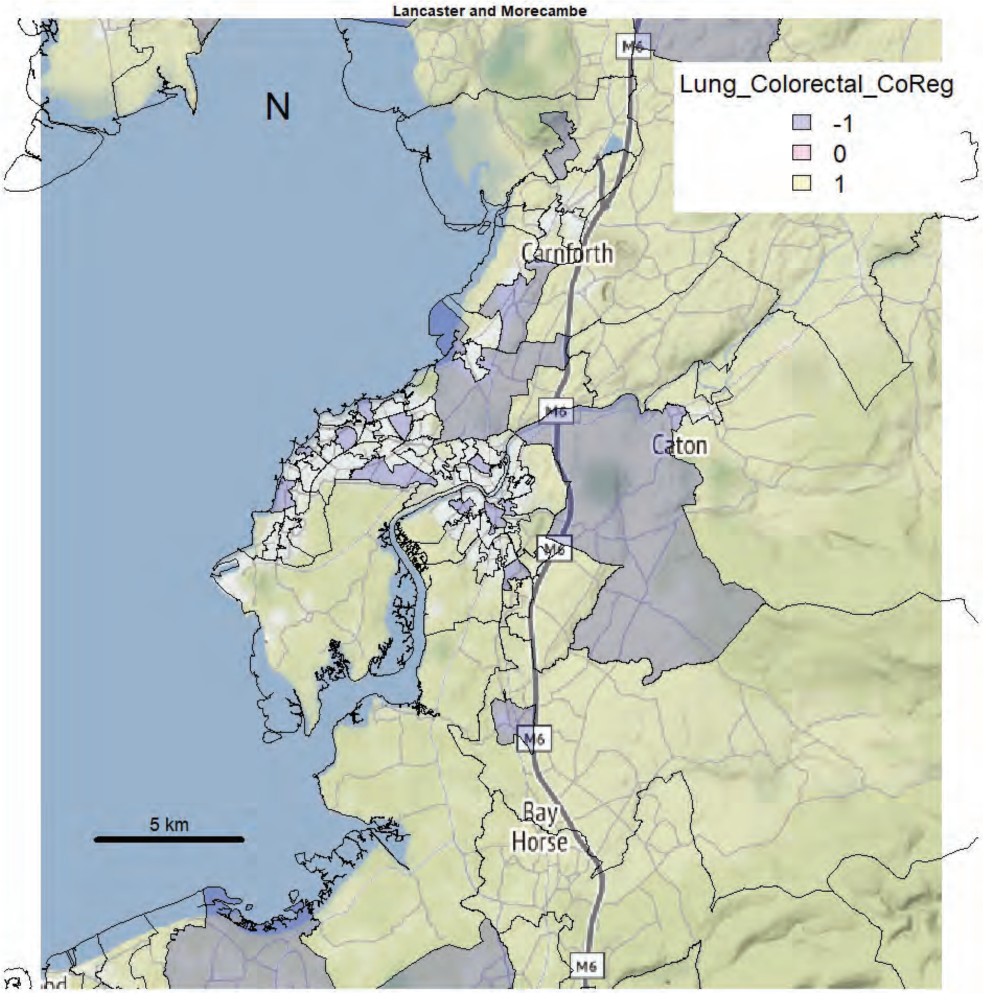

Lancaster and Morecambe
Lung_Colorectal_CoReg
-1
0
1
N
Carnforth
Caton
Bay Horse
M6
5 km


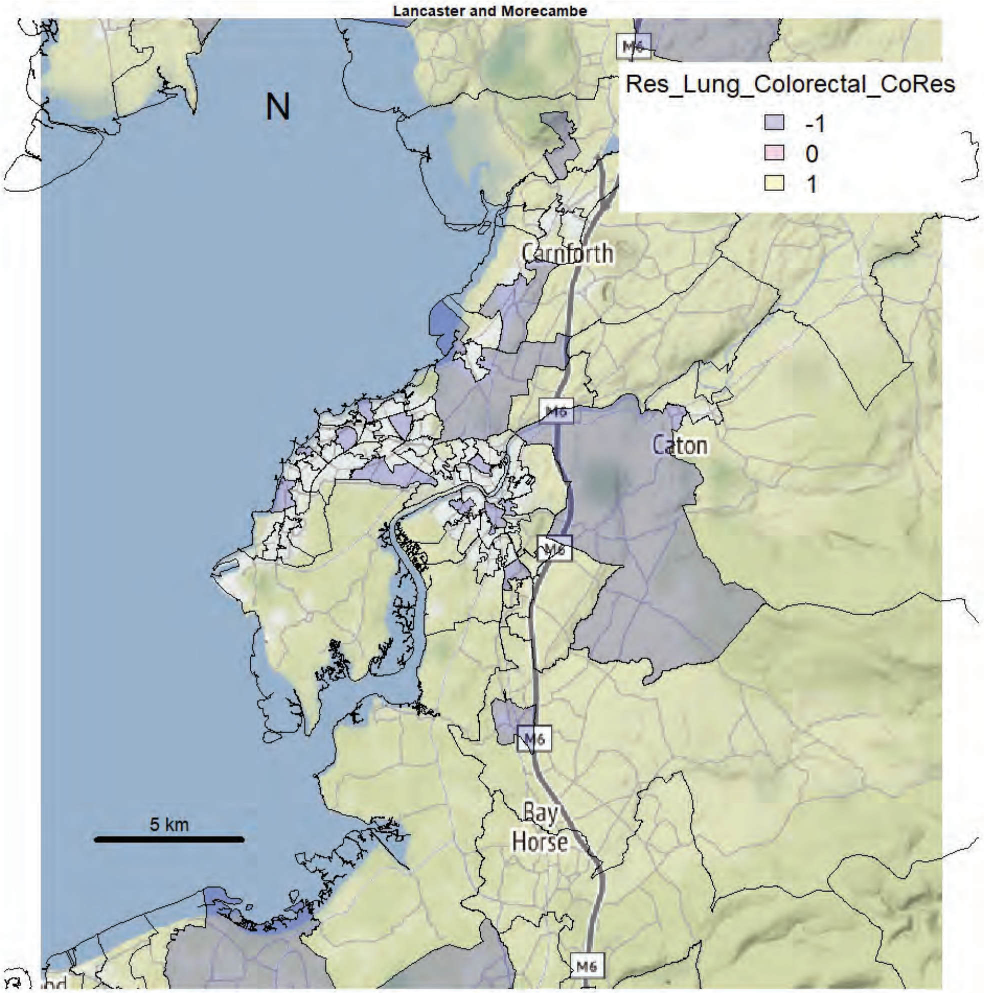

Lancaster and Morecambe
Res_Lung_Colorectal_CoRes
-1
0
1
N
Carnforth
Caton
Bay Horse
M6
5 km

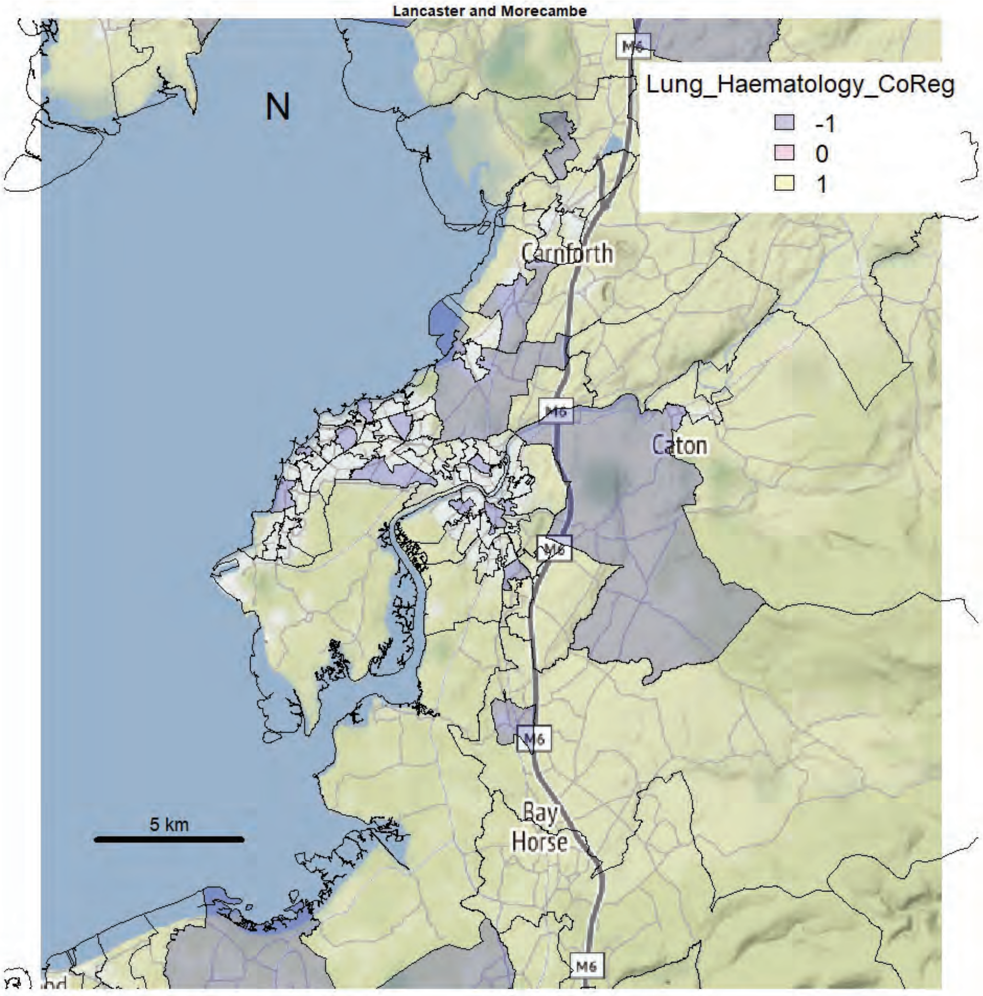

Lancaster and Morecambe
N
Lung_Haematology_CoReg
-1
0
1
Carnforth
Caton
Bay Horse
M6
5 km


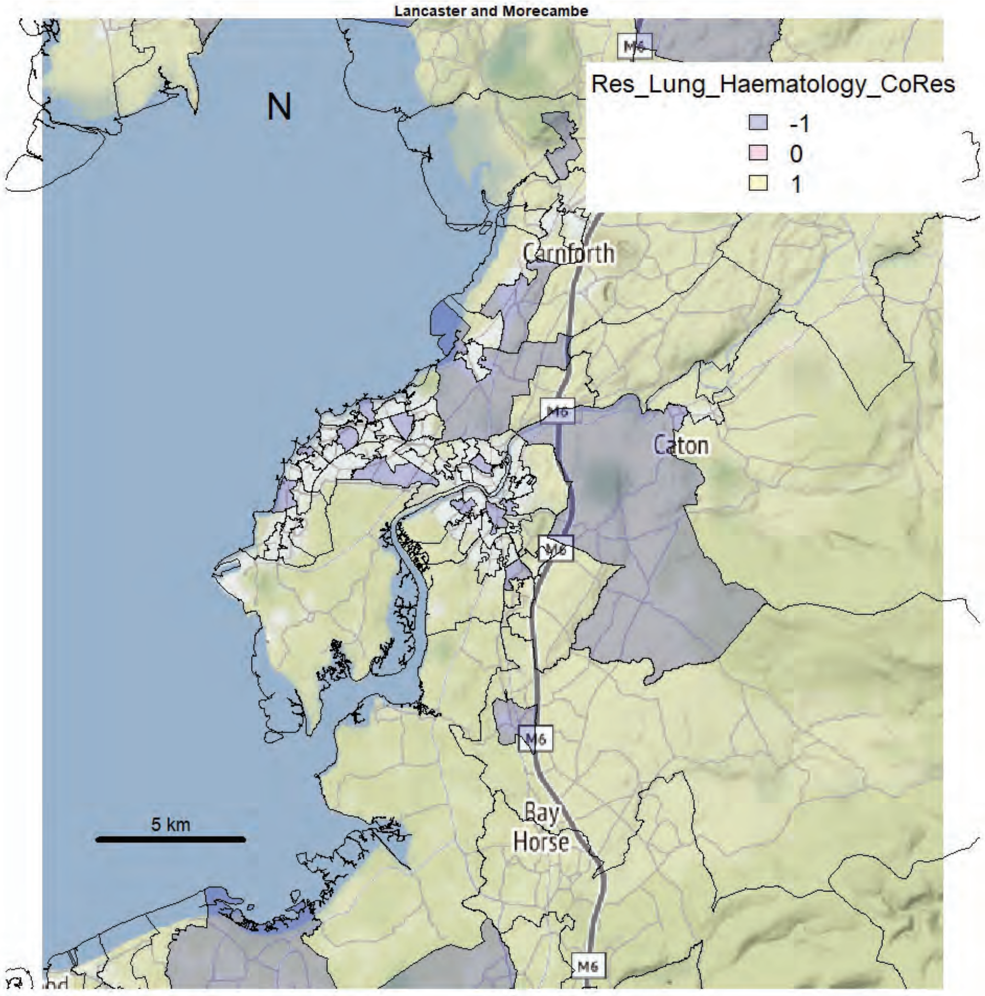

Lancaster and Morecambe
N
Res_Lung_Haematology_CoRes
-1
0
1
Carnforth
Caton
Bay Horse
M6
5 km

Lancaster and Morecambe

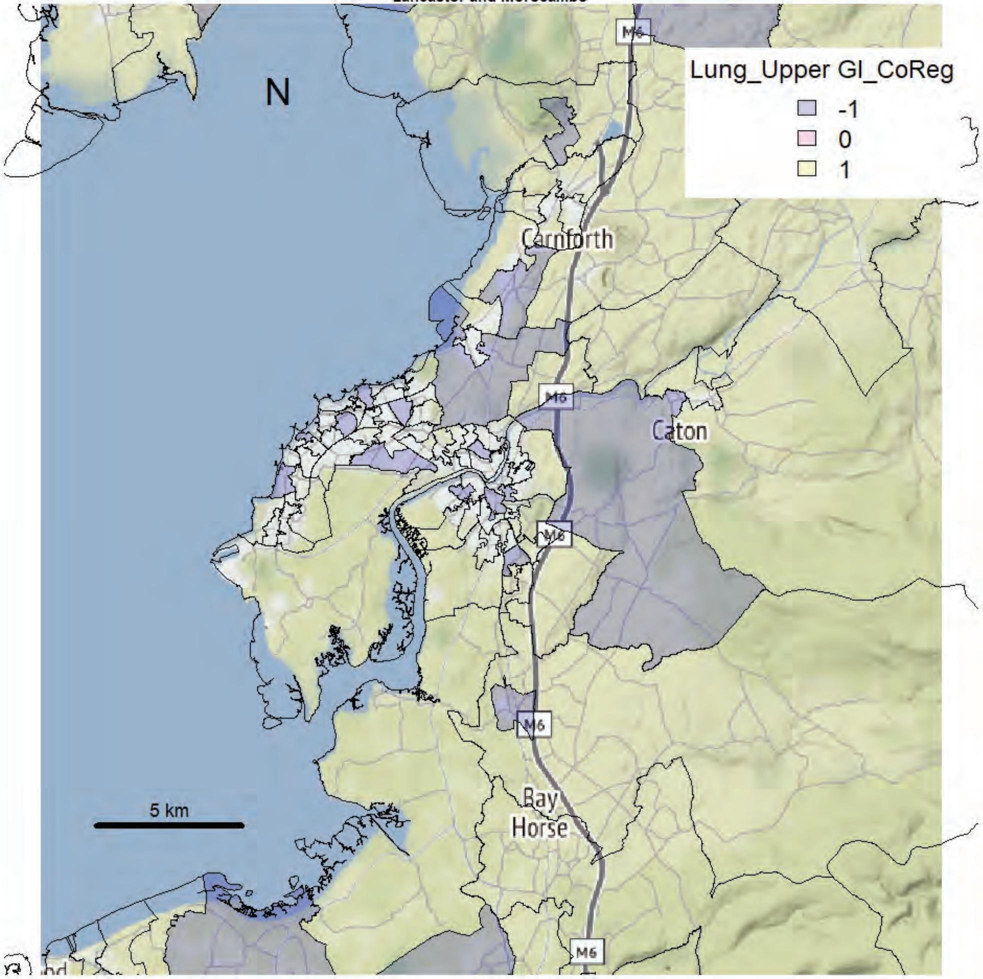


Lancaster and Morecambe

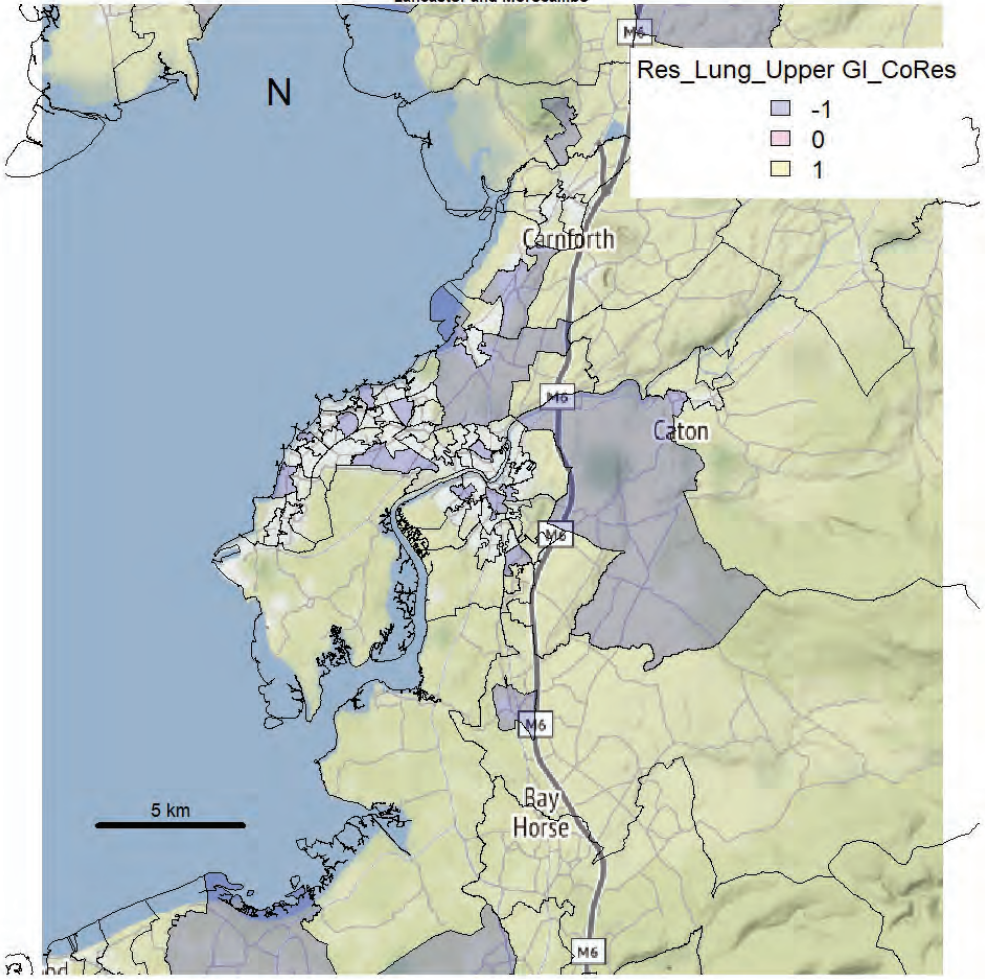

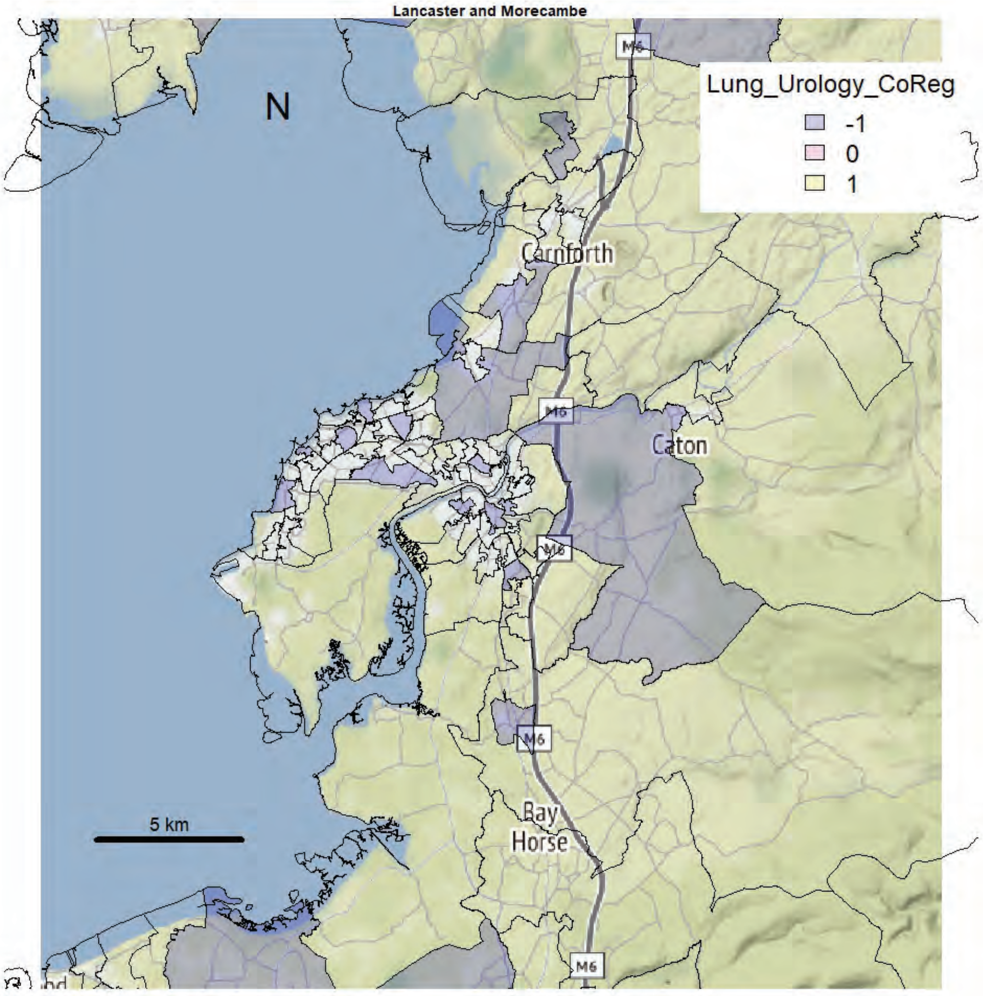
Lancaster and Morecambe
N
Lung_Urology_CoReg
-1
0
1
Carnforth
Caton
M6
Bay
Horse
5 km

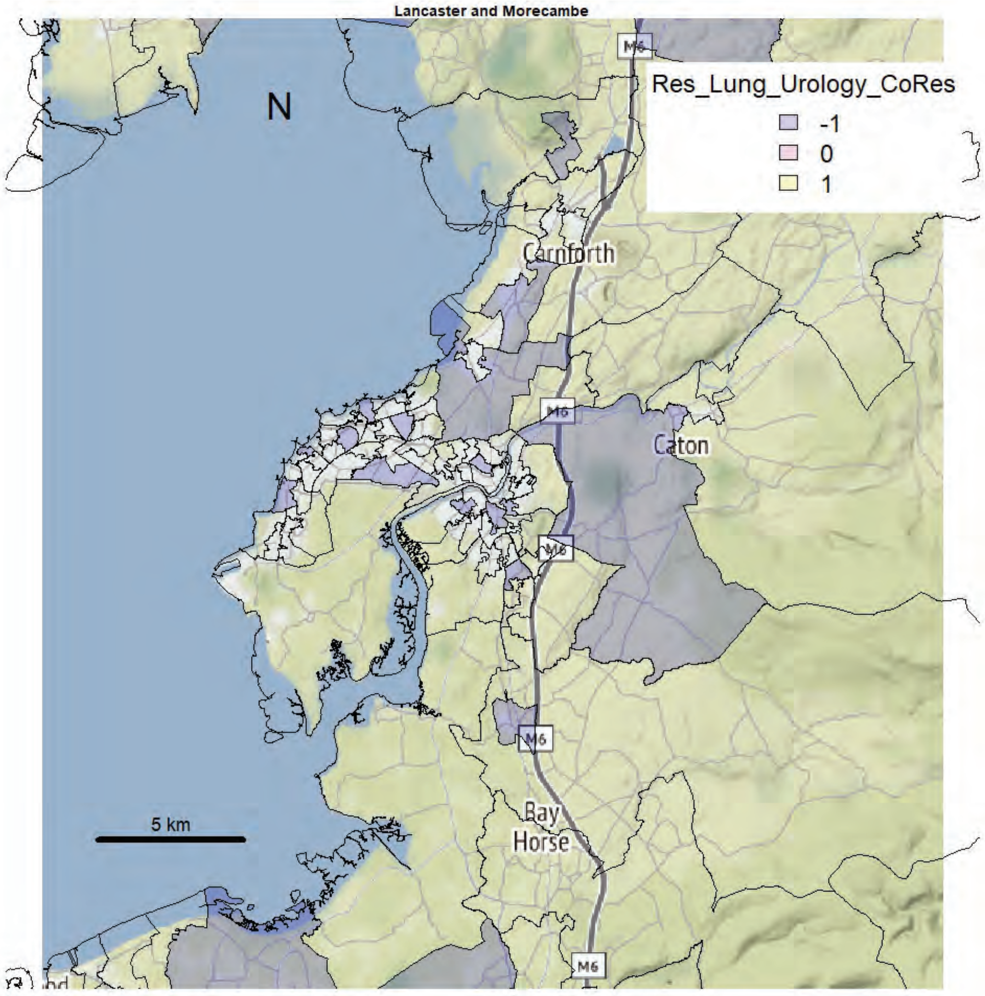
Lancaster and Morecambe
N
Res_Lung_Urology_CoRes
-1
0
1
Carnforth
Caton
M6
Bay
Horse
5 km

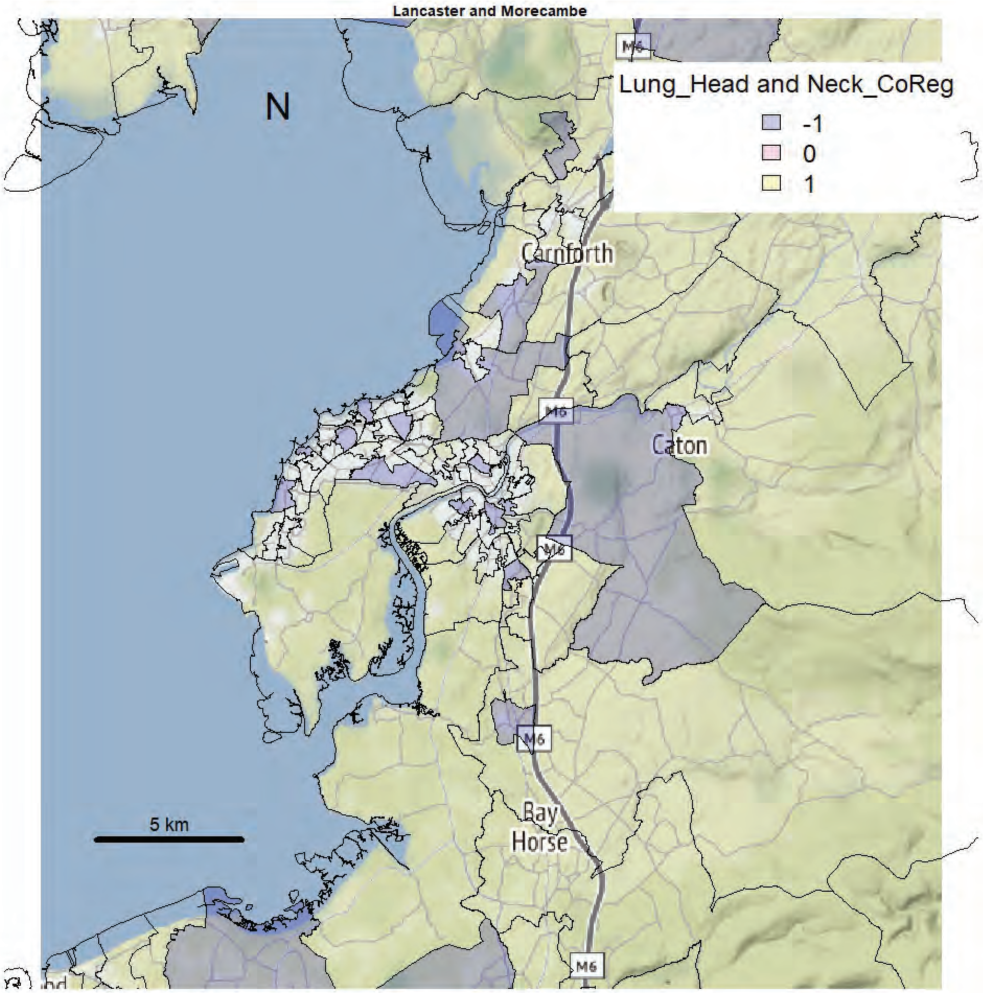
Lancaster and Morecambe
N
Lung_Head and Neck_CoReg
-1
0
1
Carnforth
Caton
Bay
Horse
M6
5 km

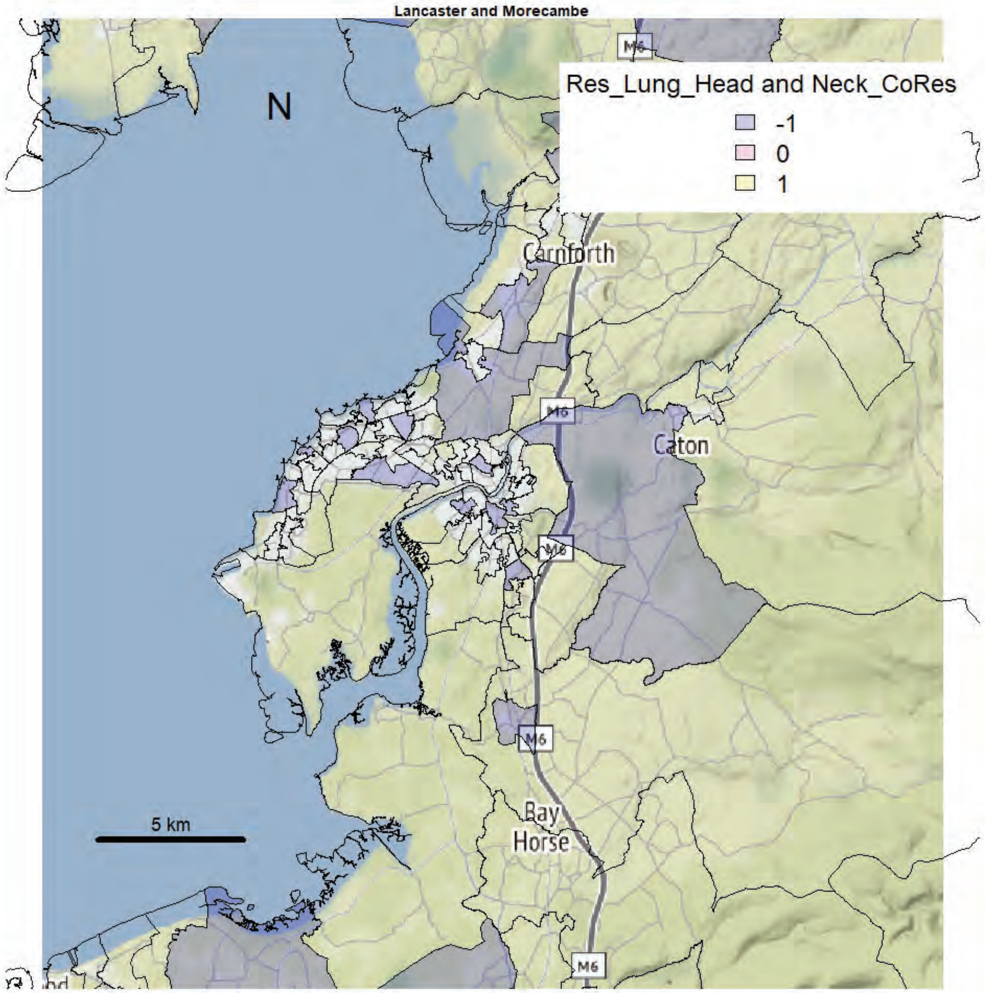
Lancaster and Morecambe
N
Res_Lung_Head and Neck_CoRes
-1
0
1
Carnforth
Caton
Bay
Horse
M6
5 km

Lancaster and Morecambe

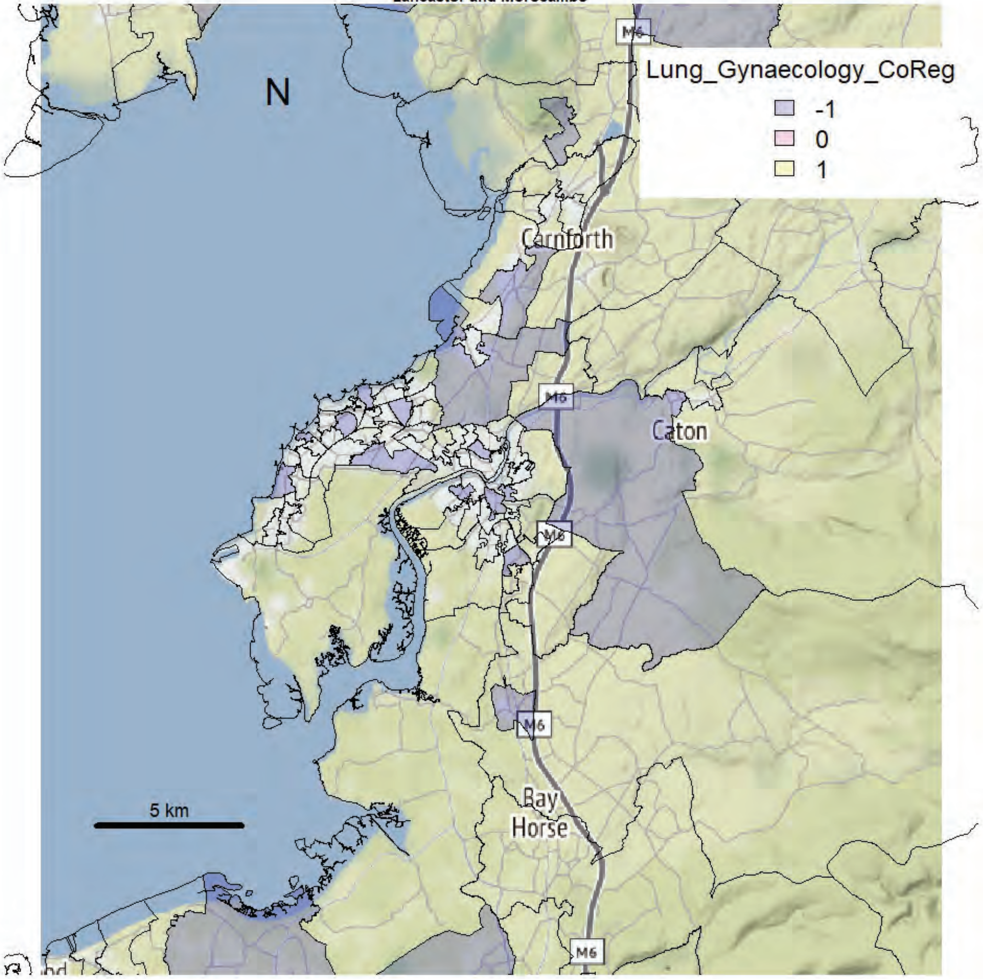


Lancaster and Morecambe

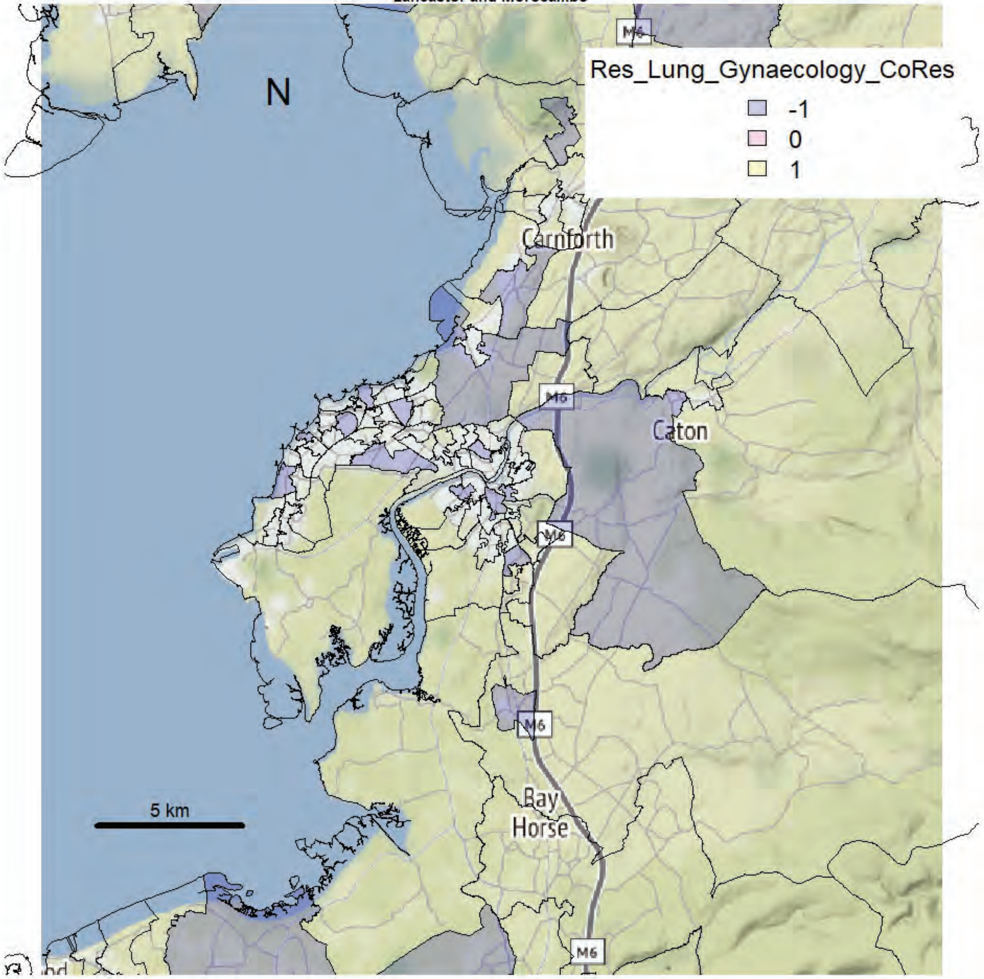

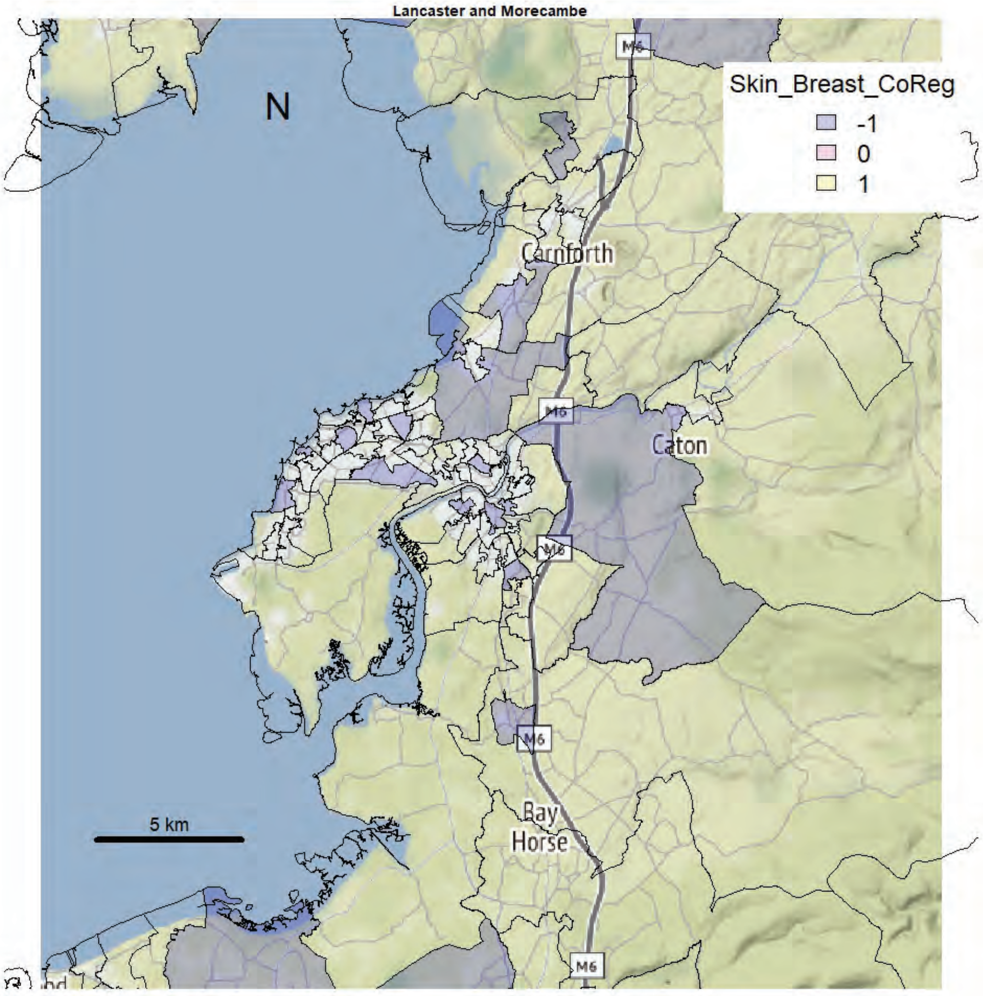
Lancaster and Morecambe
N
Skin_Breast_CoReg
-1
0
1
Carnforth
Caton
M6
Bay
Horse
5 km

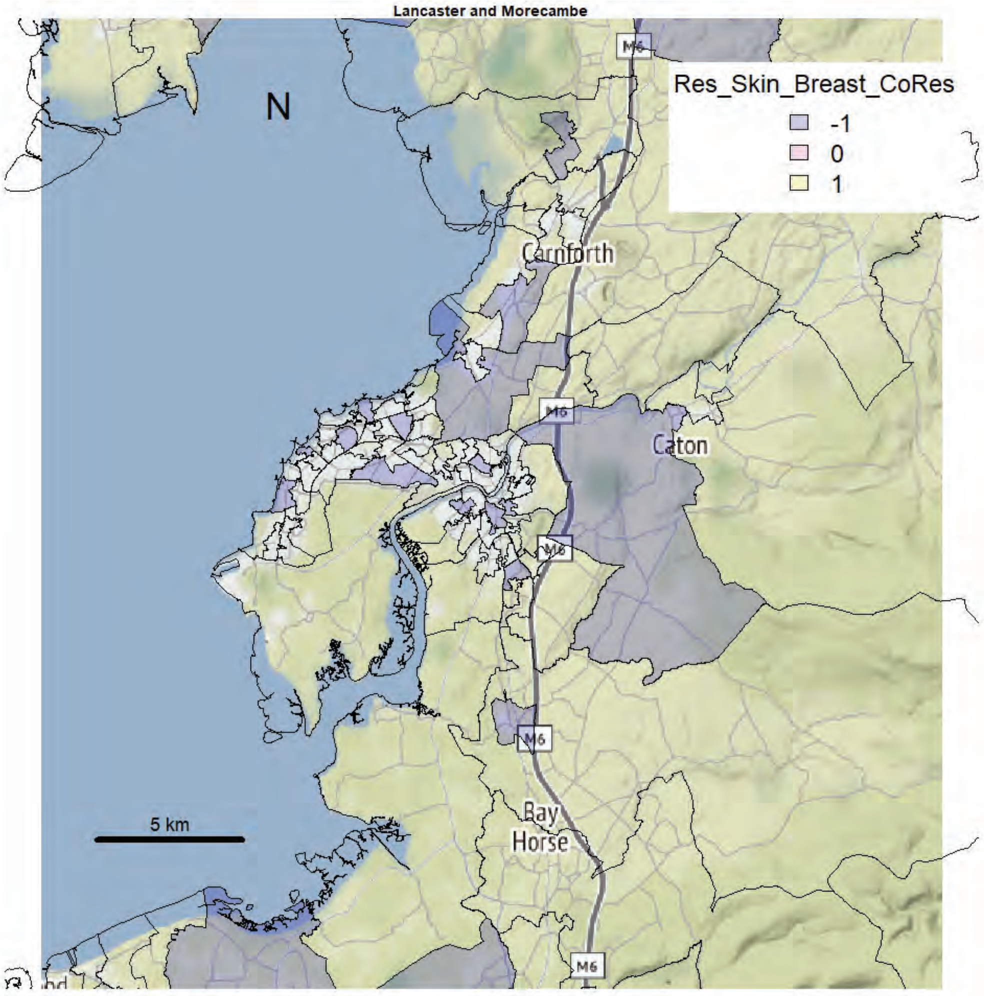
Lancaster and Morecambe
N
Res_Skin_Breast_CoRes
-1
0
1
Carnforth
Caton
M6
Bay
Horse
5 km

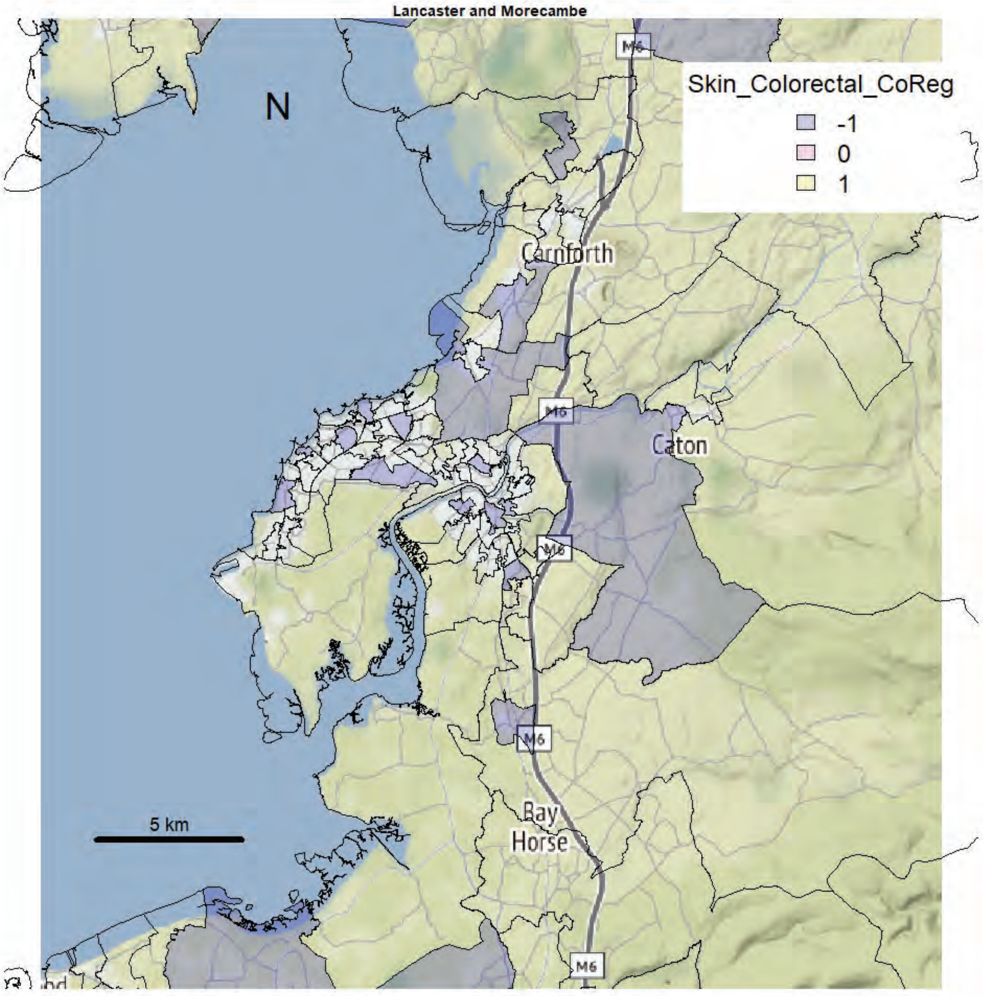
Lancaster and Morecambe
N
Skin_Colorectal_CoReg
-1
0
1
Carnforth
Caton
Bay
Horse
M6
5 km

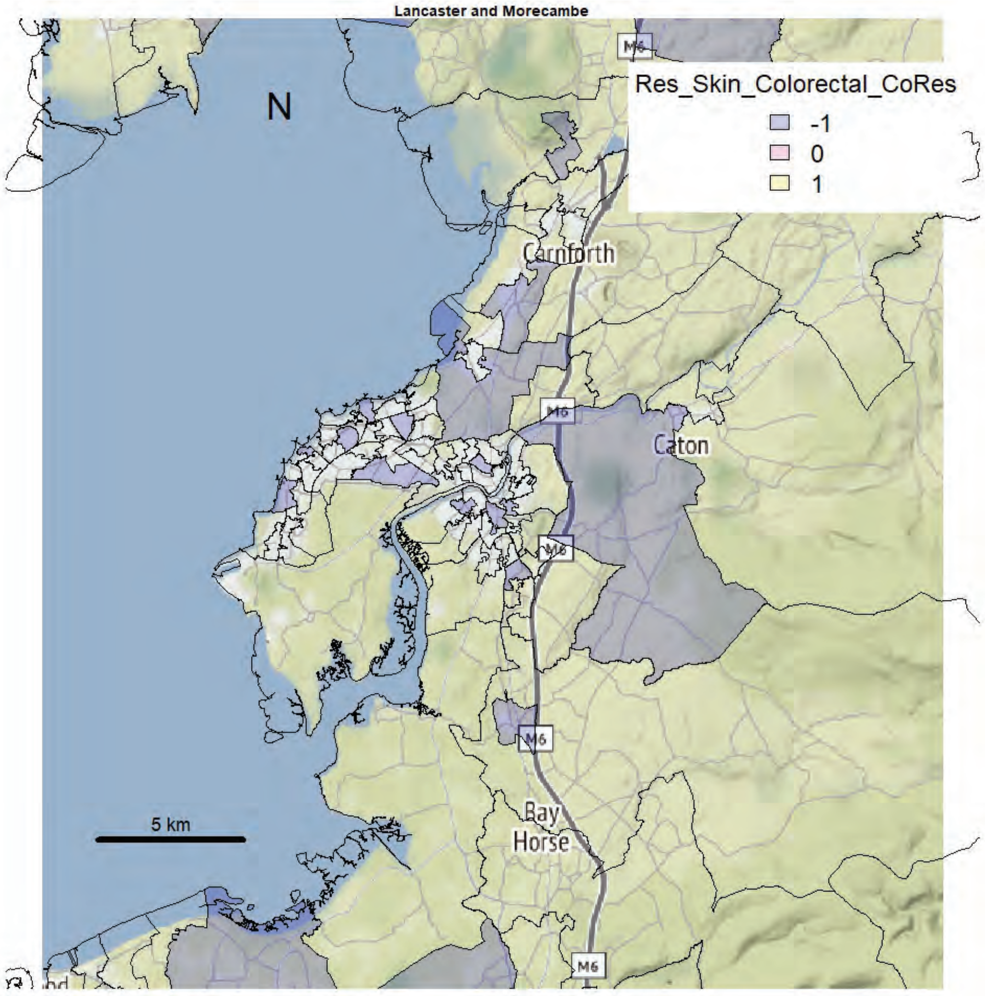
Lancaster and Morecambe
N
Res_Skin_Colorectal_CoRes
-1
0
1
Carnforth
Caton
Bay
Horse
M6
5 km

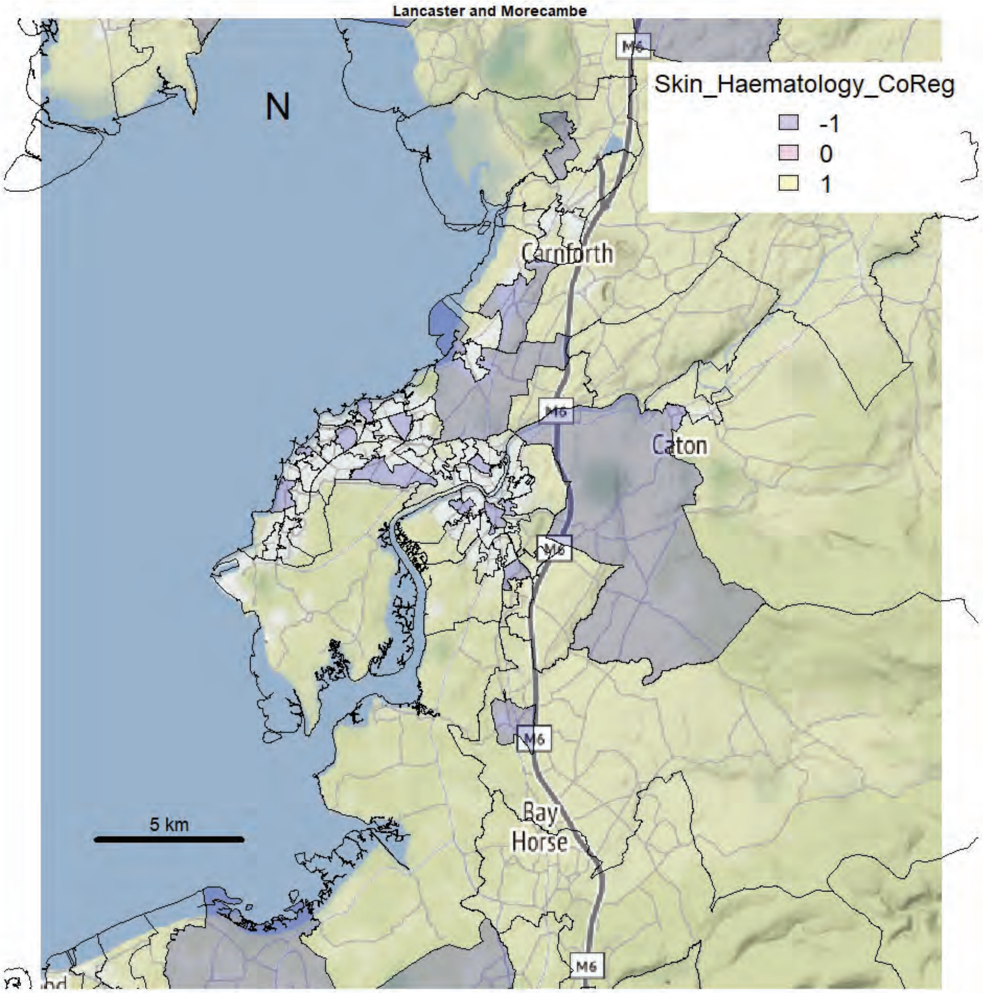
Lancaster and Morecambe
N
Skin_Haematology_CoReg
-1
0
1
Carnforth
Caton
Bay Horse
M6
5 km

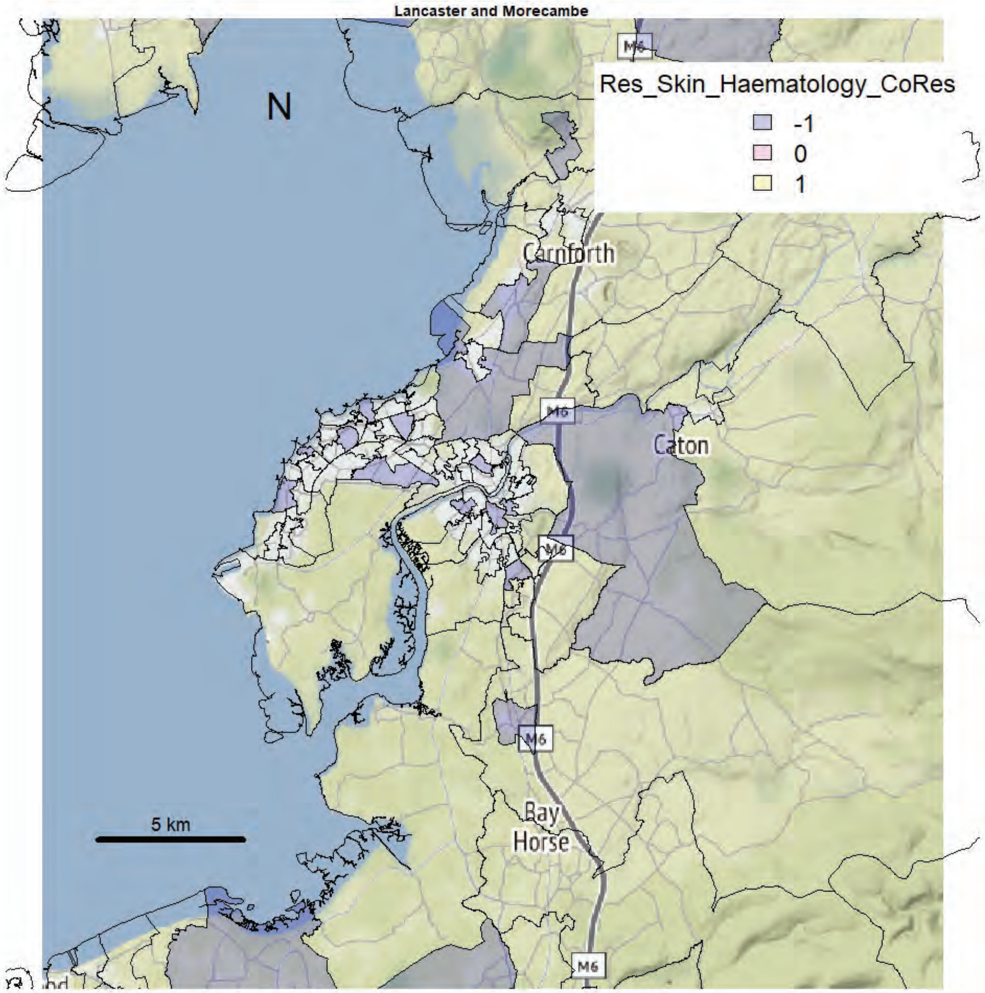
Lancaster and Morecambe
N
Res_Skin_Haematology_CoRes
-1
0
1
Carnforth
Caton
Bay Horse
M6
5 km

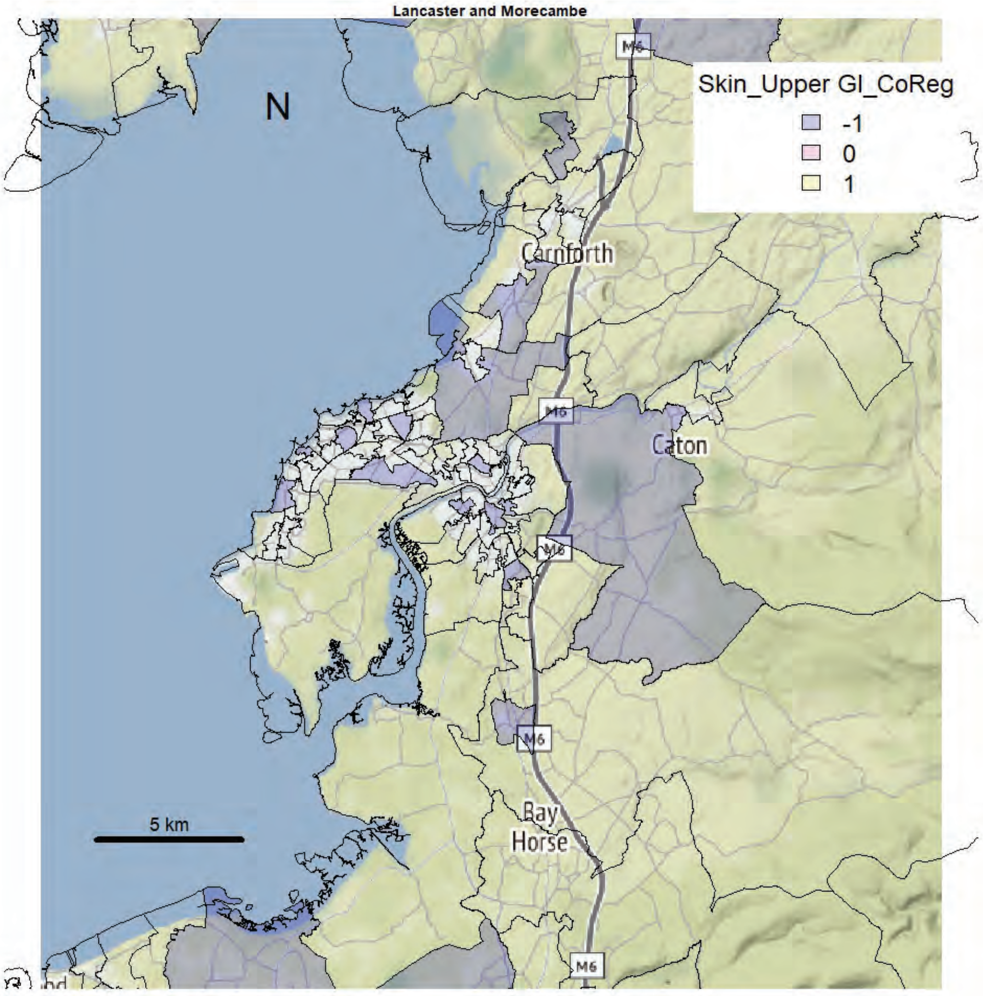
Lancaster and Morecambe
N
Skin_Upper GI_CoReg
-1
0
1
Carnforth
Caton
Bay
Horse
M6
5 km

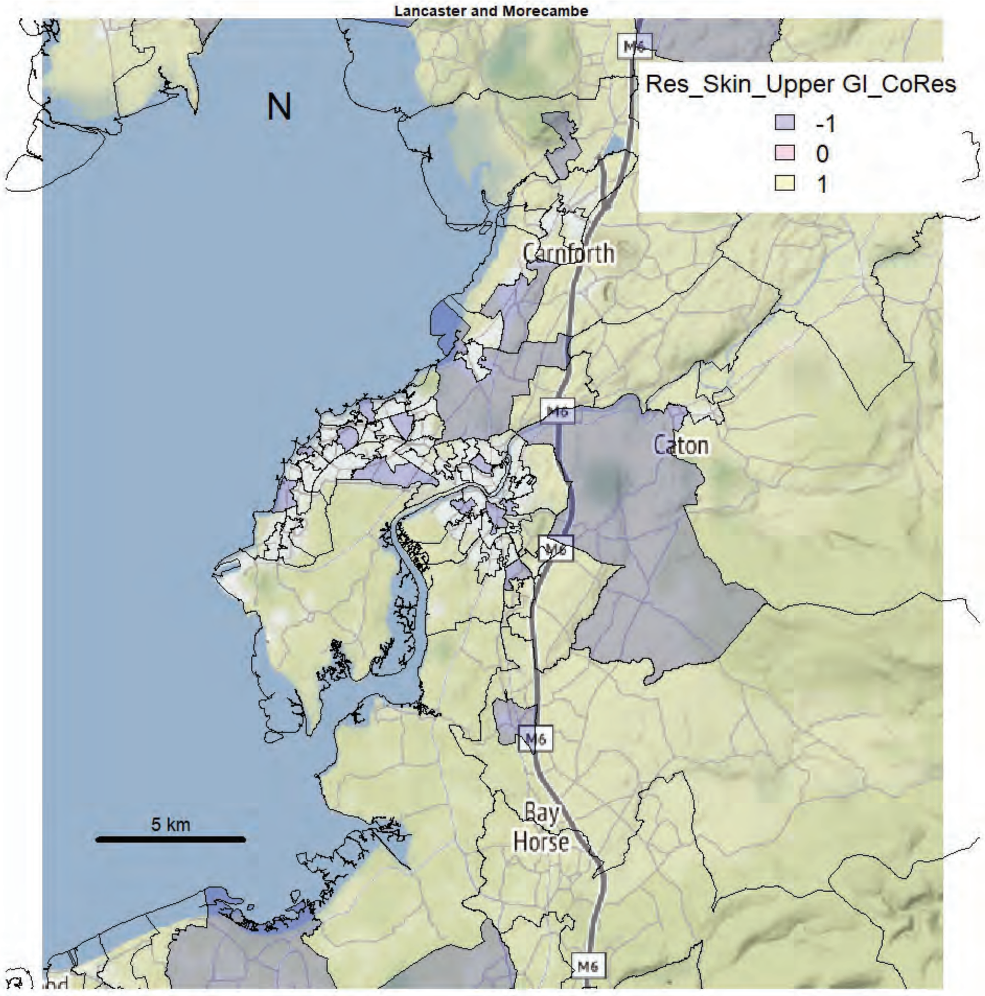
Lancaster and Morecambe
N
Res_Skin_Upper GI_CoRes
-1
0
1
Carnforth
Caton
Bay
Horse
M6
5 km

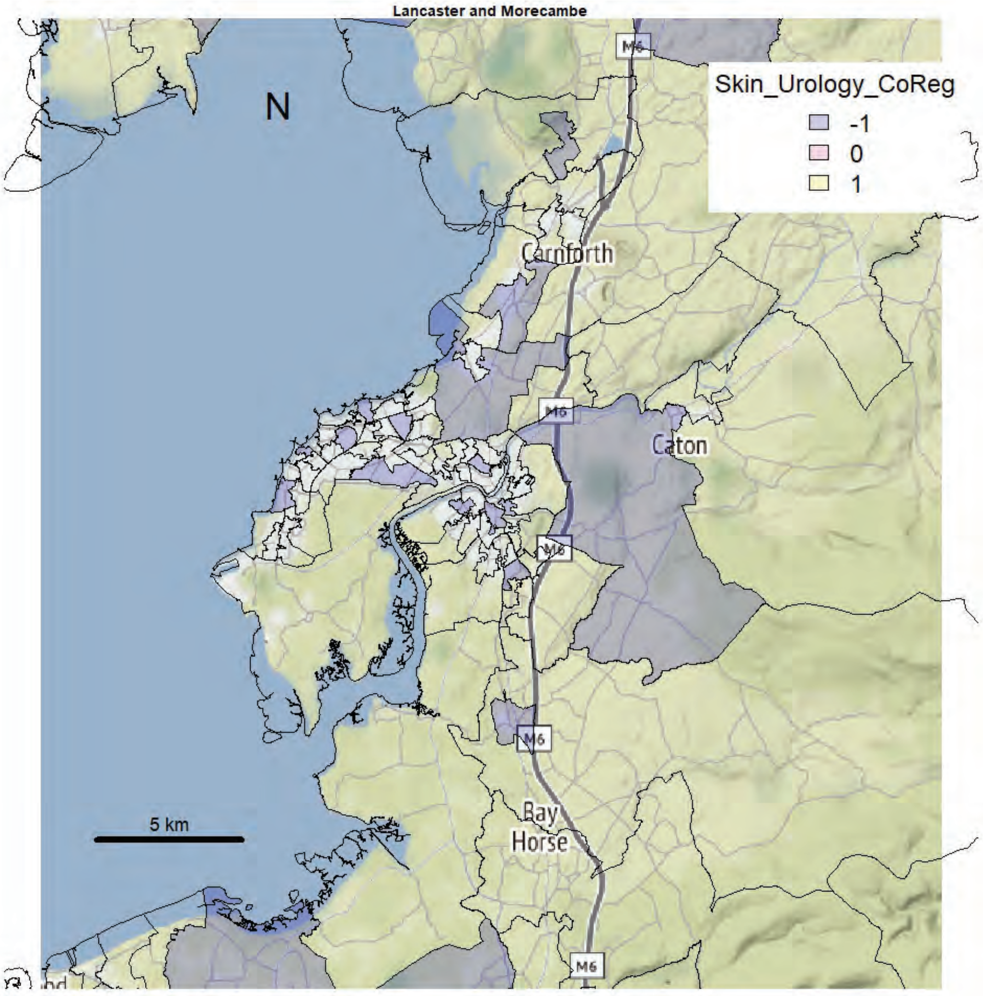
Lancaster and Morecambe
N
Skin_Urology_CoReg
-1
0
1
Carnforth
M6
Caton
Bay
Horse
5 km

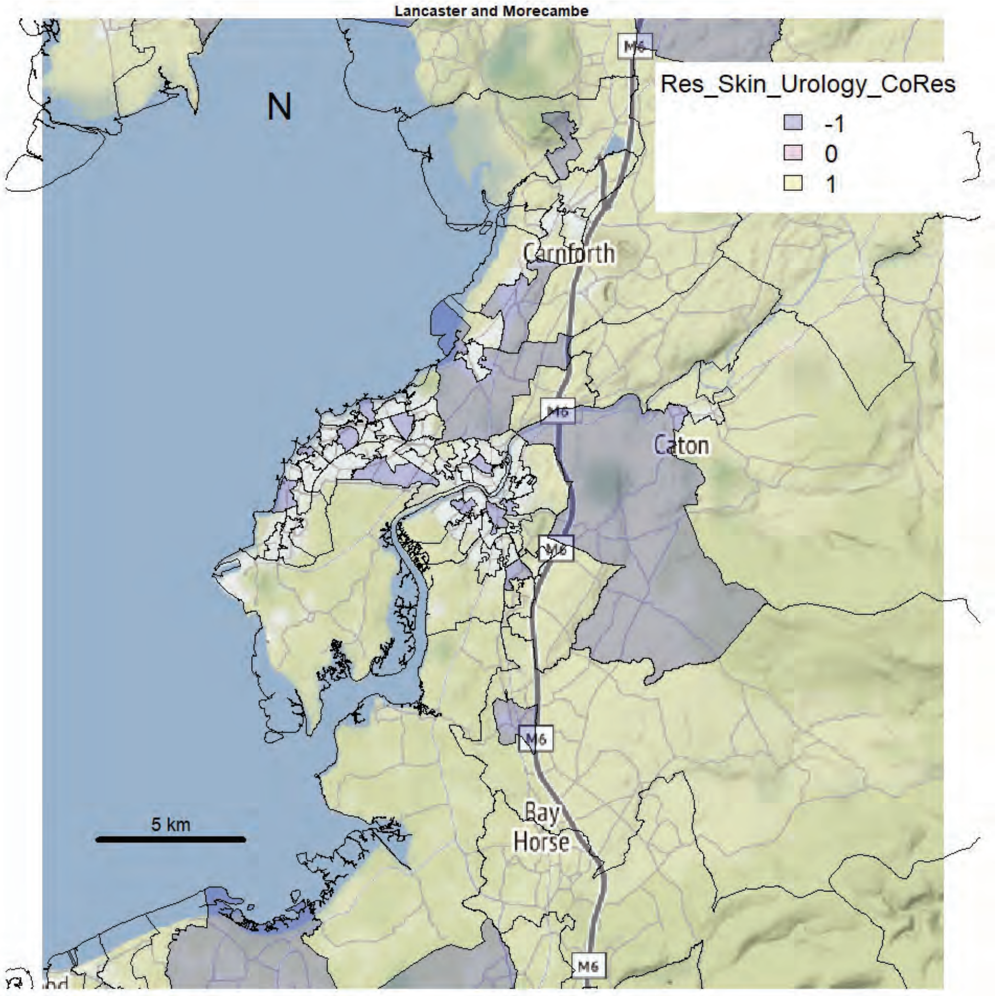
Lancaster and Morecambe
N
Res_Skin_Urology_CoRes
-1
0
1
Carnforth
M6
Caton
Bay
Horse
5 km

Lancaster and Morecambe

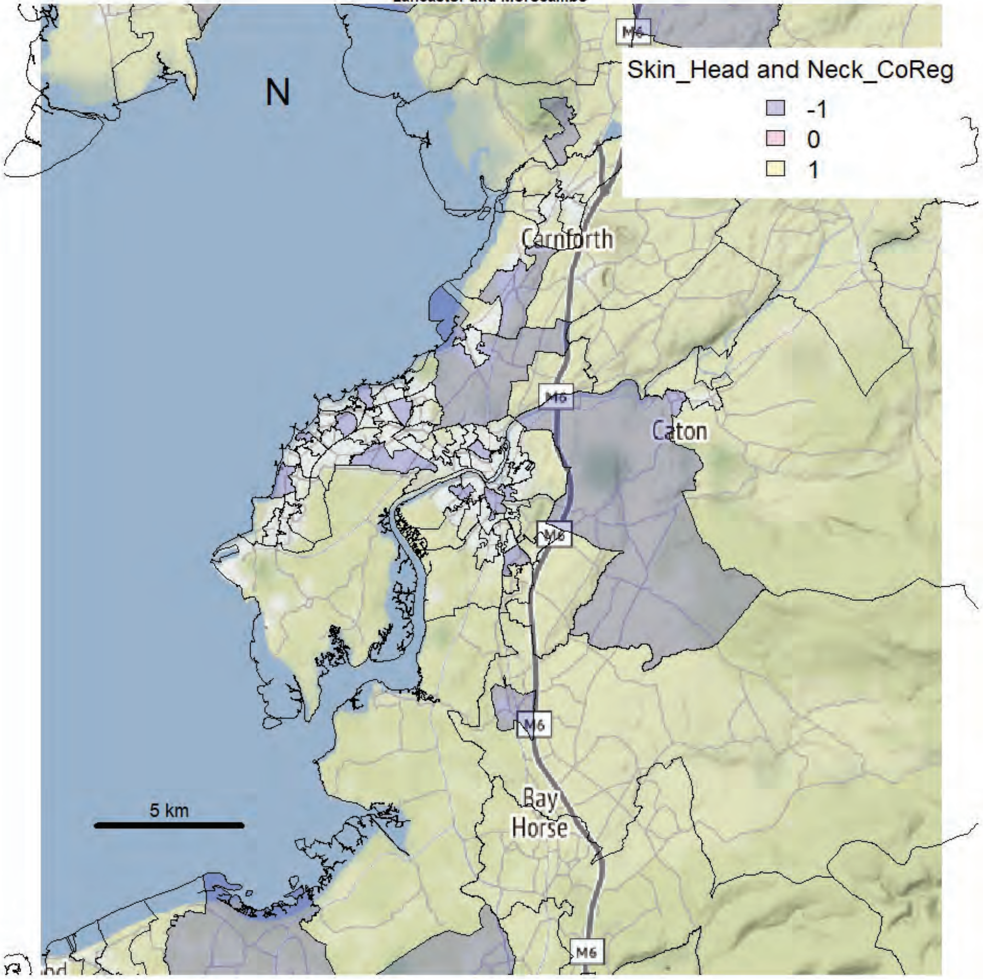


Lancaster and Morecambe

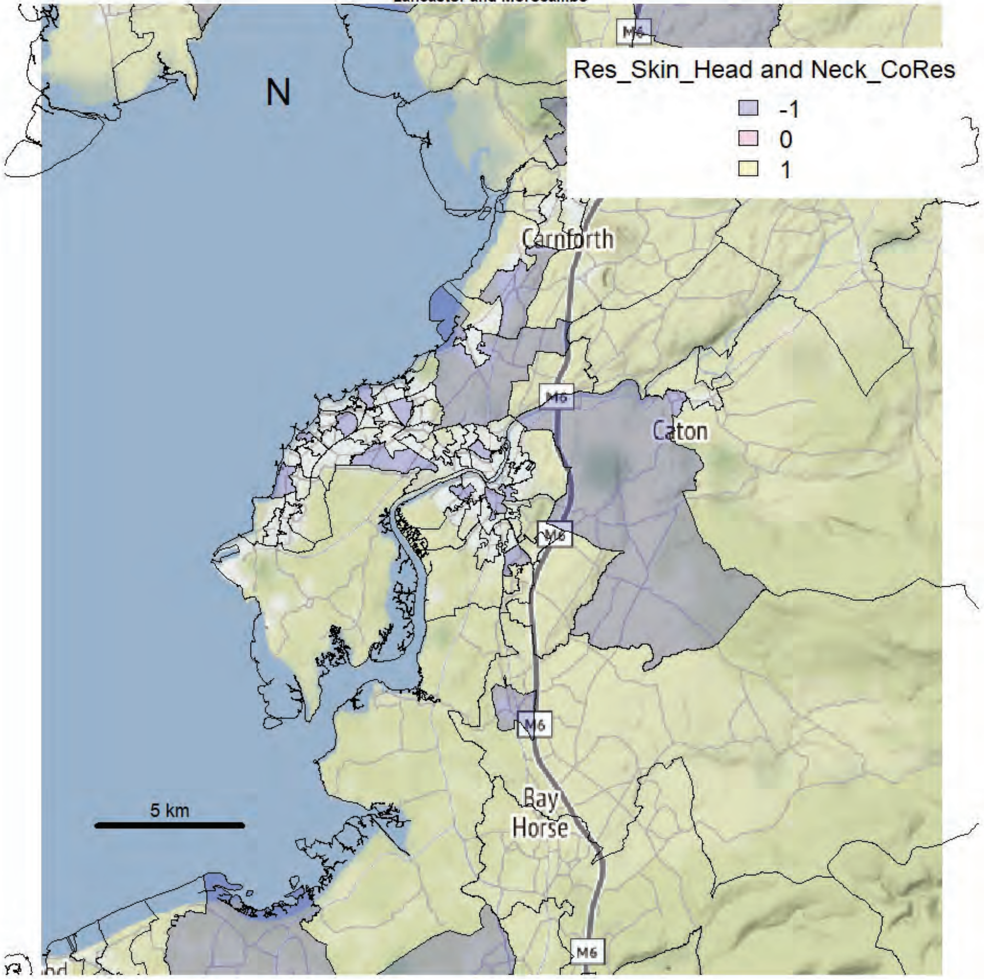

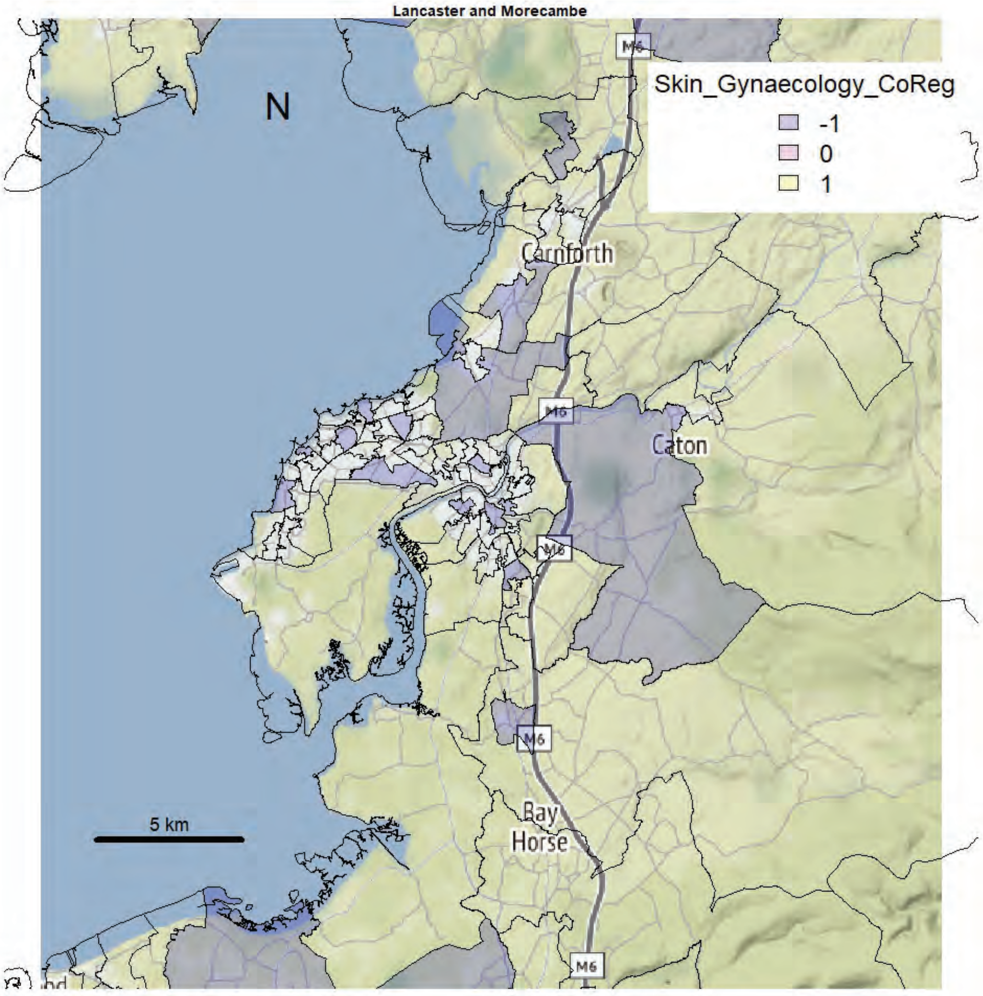
Lancaster and Morecambe
N
Skin_Gynaecology_CoReg
-1
0
1
Carnforth
Caton
Bay Horse
M6
5 km

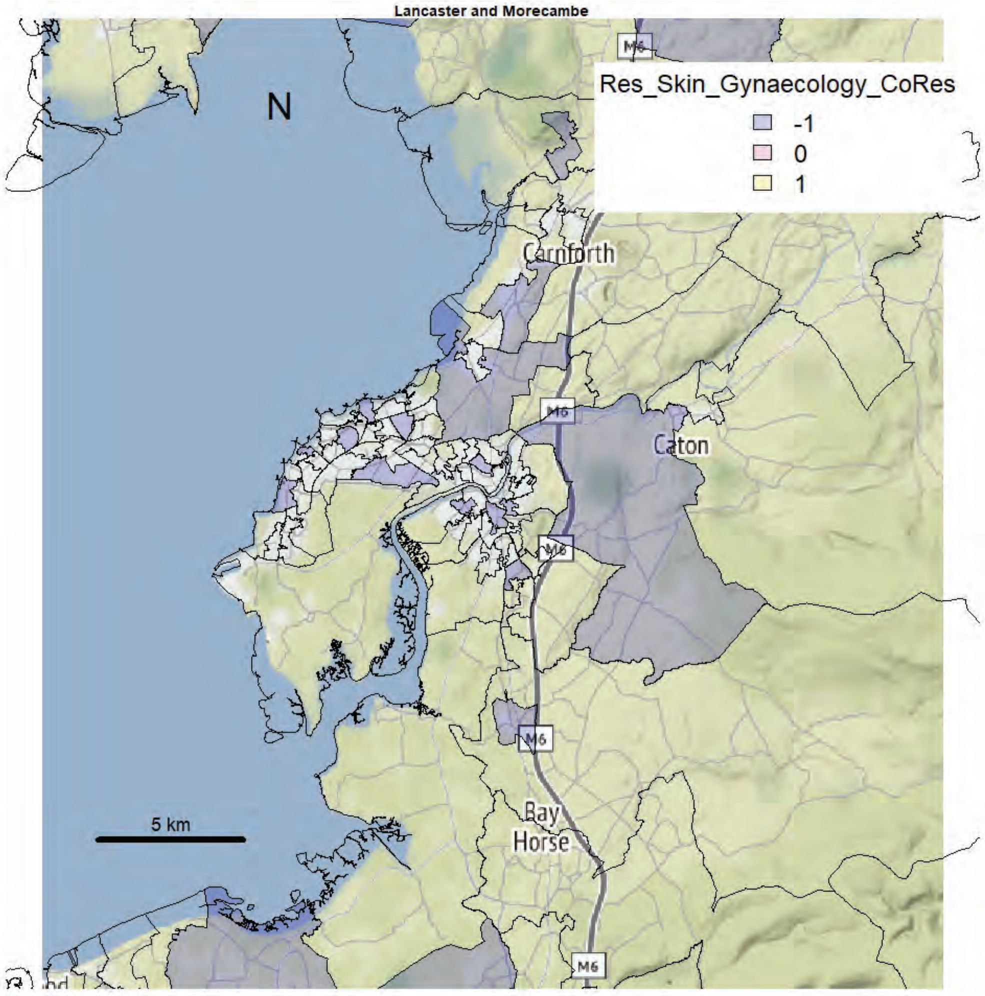
Lancaster and Morecambe
N
Res_Skin_Gynaecology_CoRes
-1
0
1
Carnforth
Caton
Bay Horse
M6
5 km

Lancaster and Morecambe

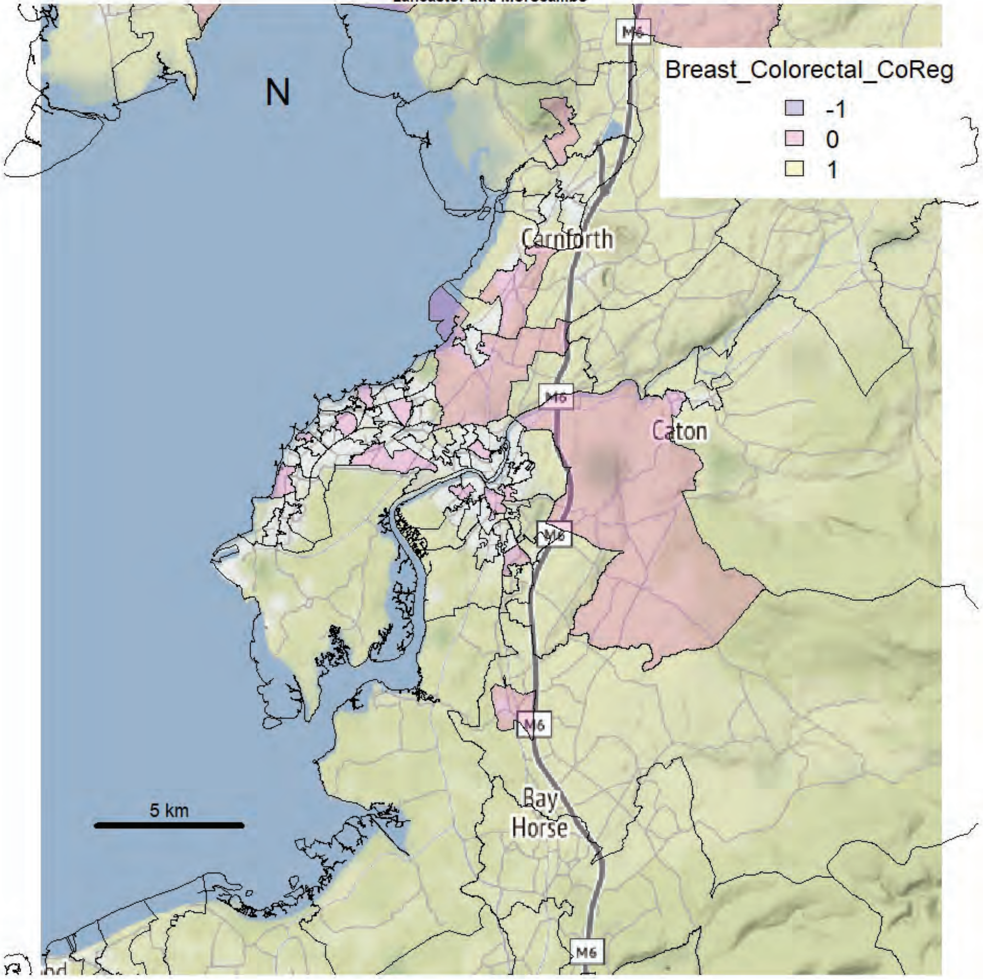


Lancaster and Morecambe

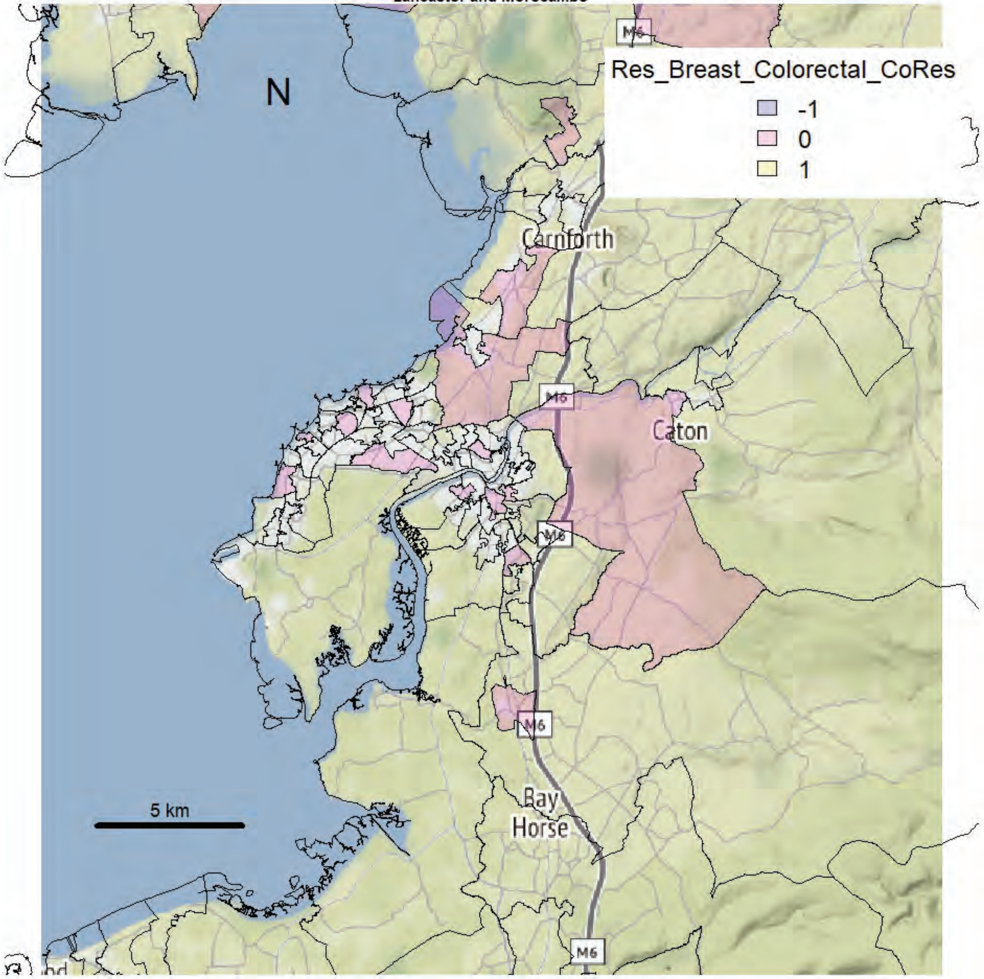

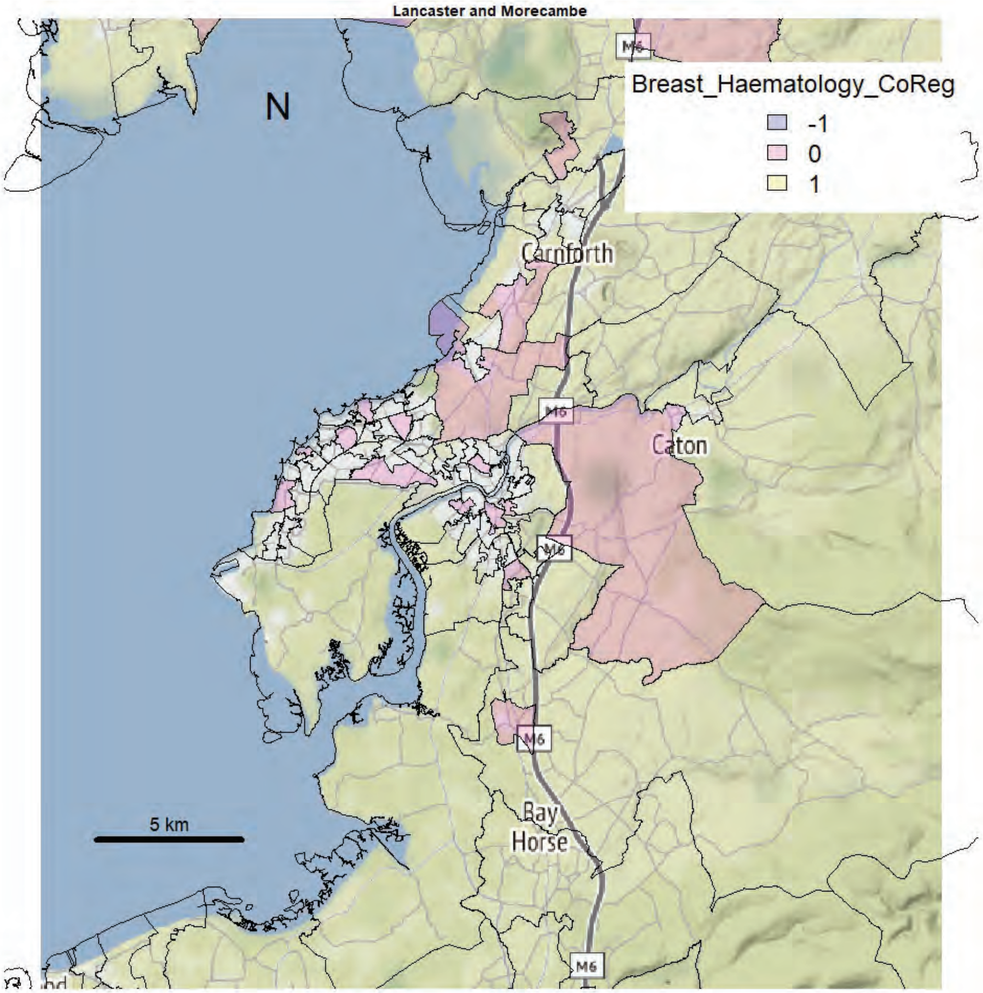
Lancaster and Morecambe
Breast_Haematology_CoReg
-1
0
1
N
Carnforth
Caton
Bay
Horse
M6
5 km

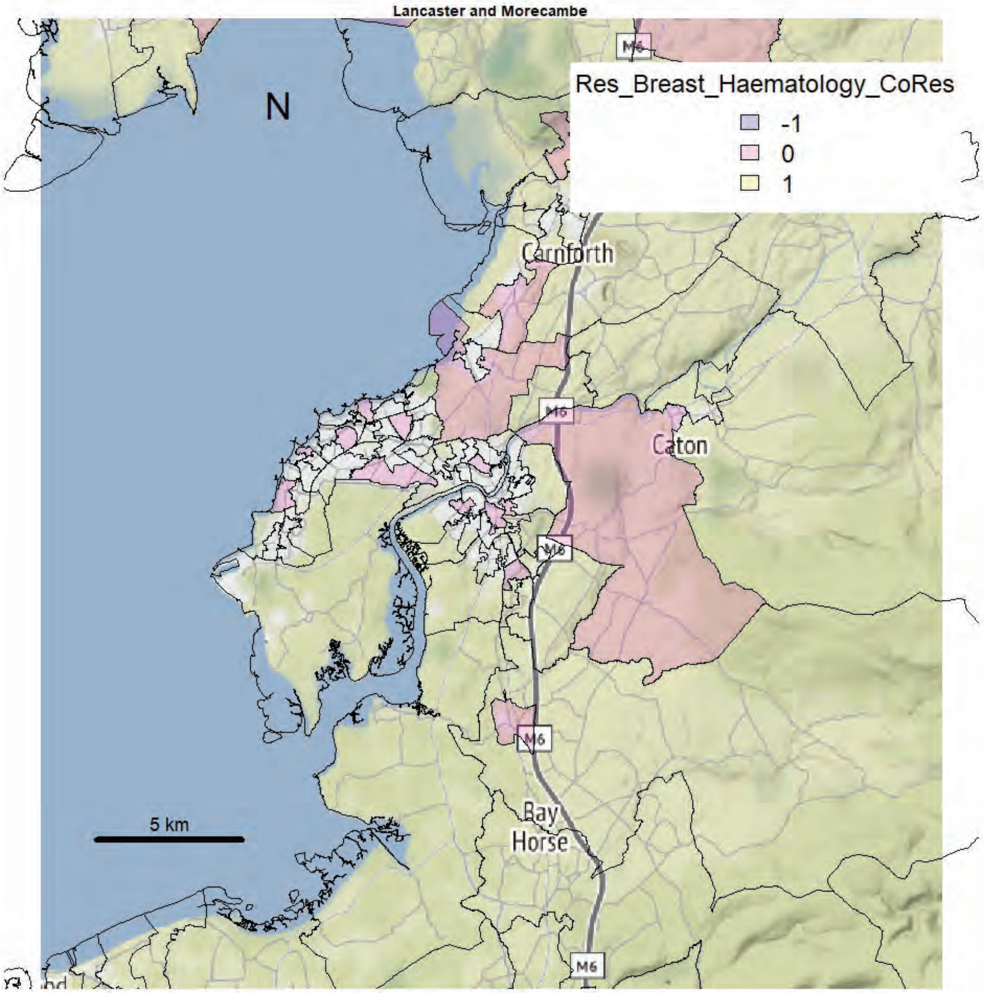
Lancaster and Morecambe
Res_Breast_Haematology_CoRes
-1
0
1
N
Carnforth
Caton
Bay
Horse
M6
5 km

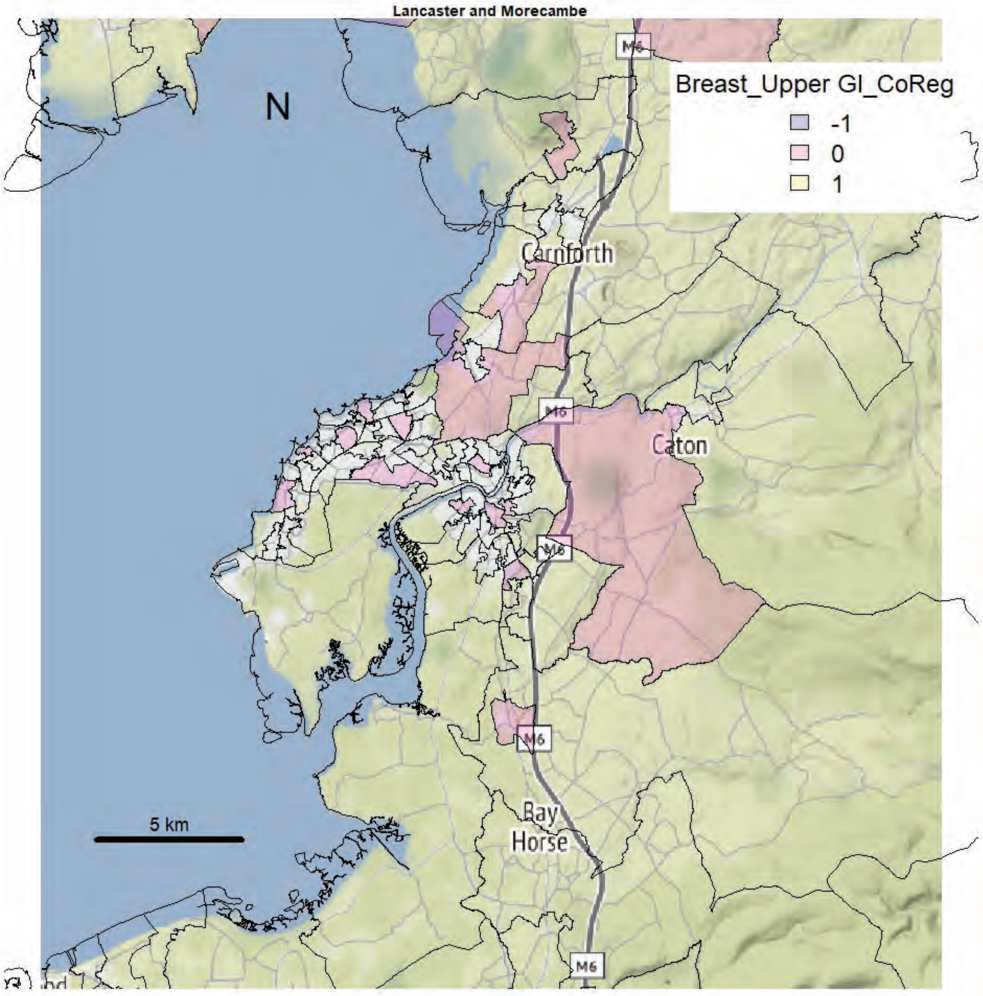
Lancaster and Morecambe
N
Breast_Upper GI_CoReg
-1
0
1
Carnforth
Caton
Bay
Horse
M6
5 km

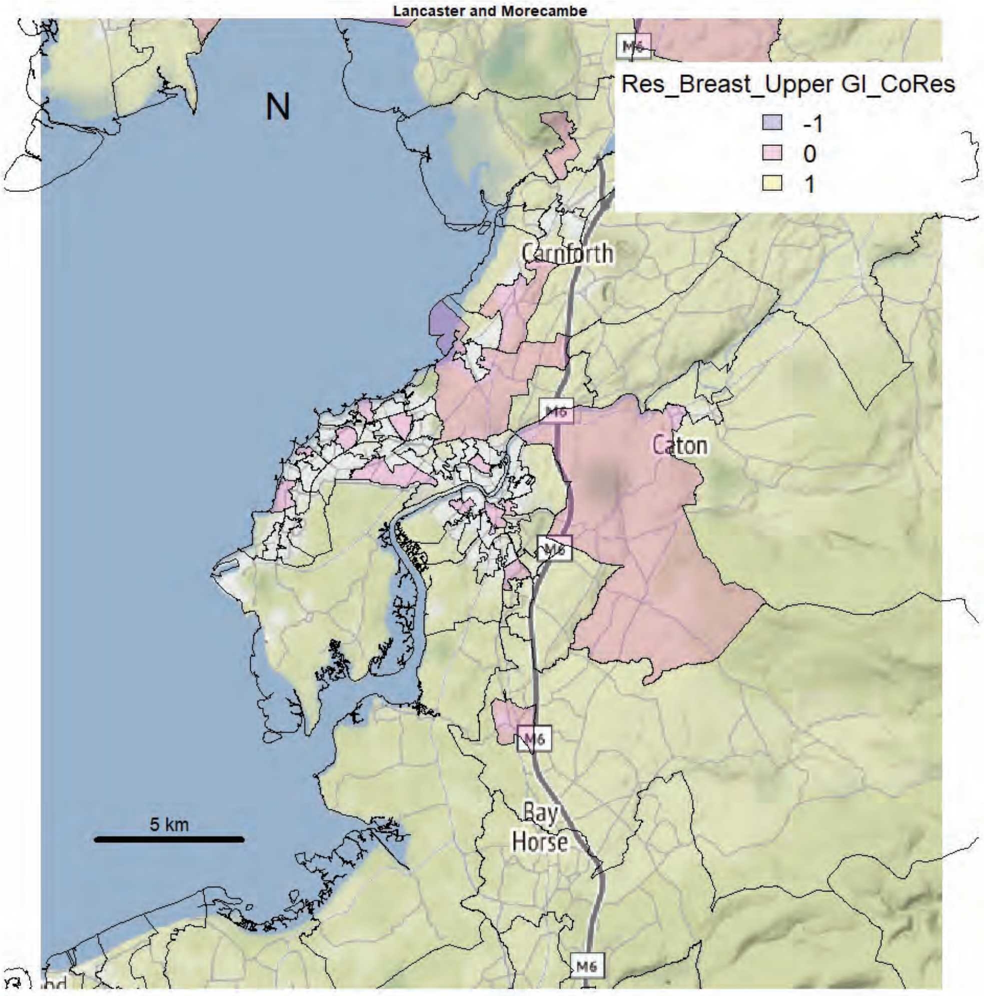
Lancaster and Morecambe
N
Res_Breast_Upper GI_CoRes
-1
0
1
Carnforth
Caton
Bay
Horse
M6
5 km

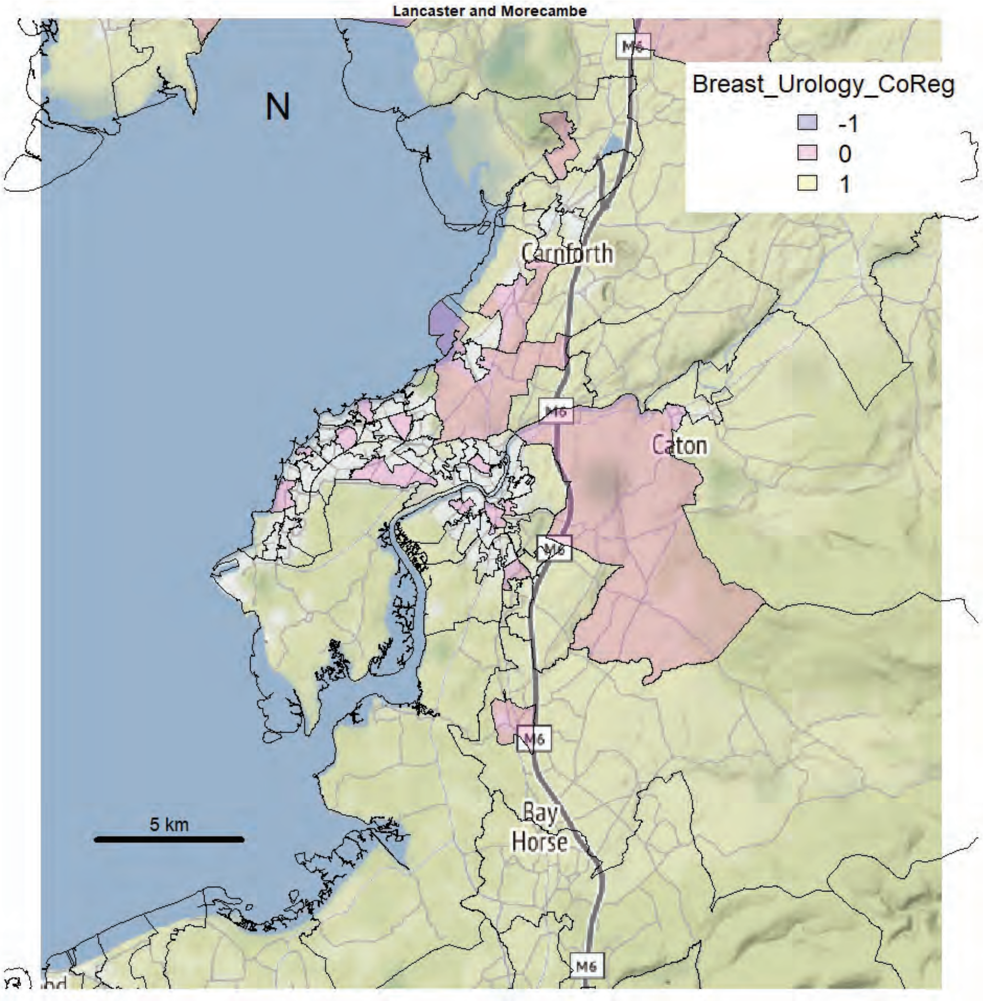
Lancaster and Morecambe
N
Breast_Urology_CoReg
-1
0
1
Carnforth
Caton
M6
Bay
Horse
5 km

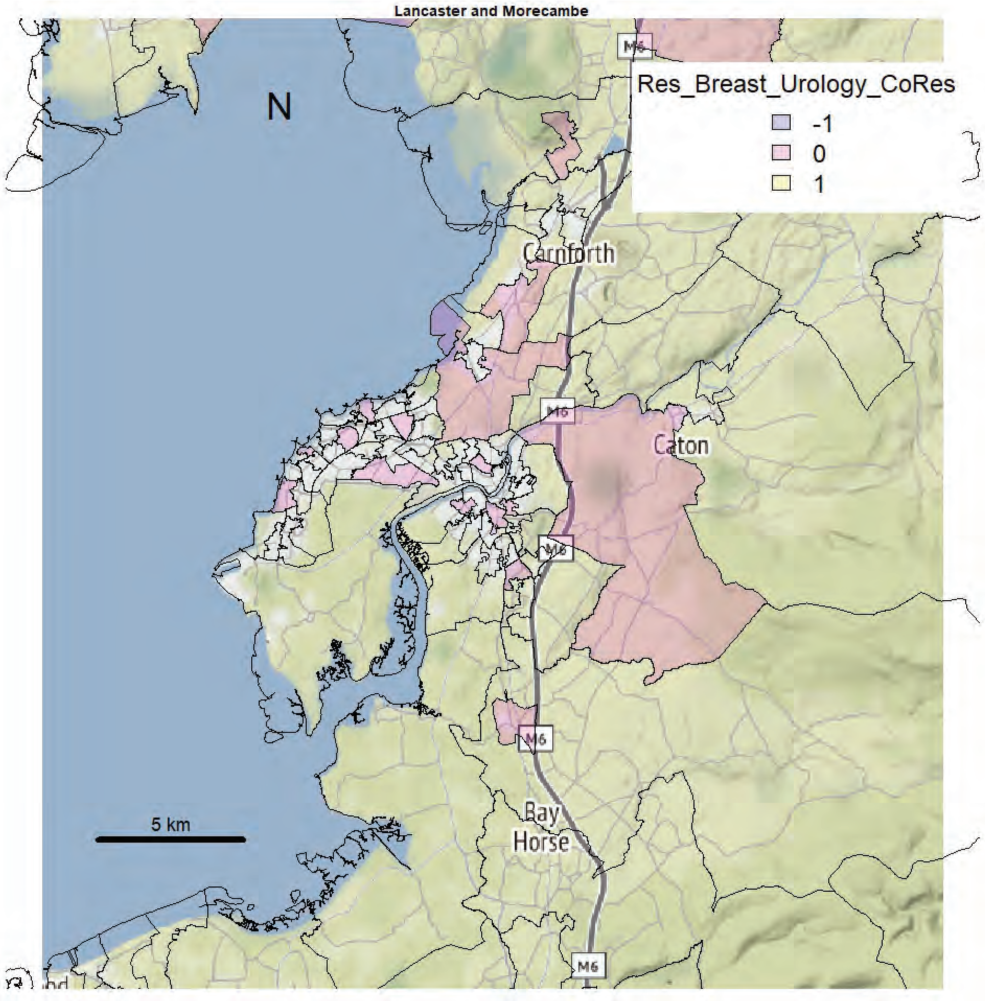
Lancaster and Morecambe
N
Res_Breast_Urology_CoRes
-1
0
1
Carnforth
Caton
M6
Bay
Horse
5 km

Lancaster and Morecambe

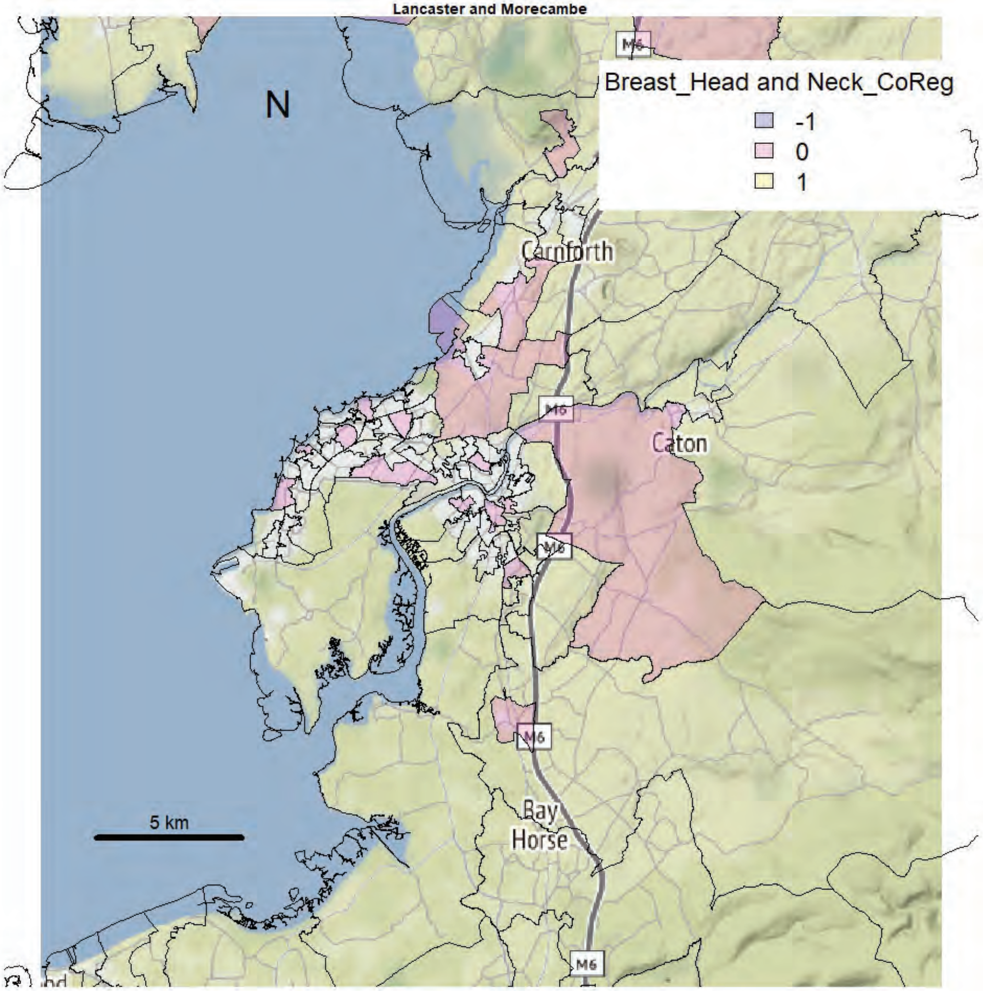


Lancaster and Morecambe

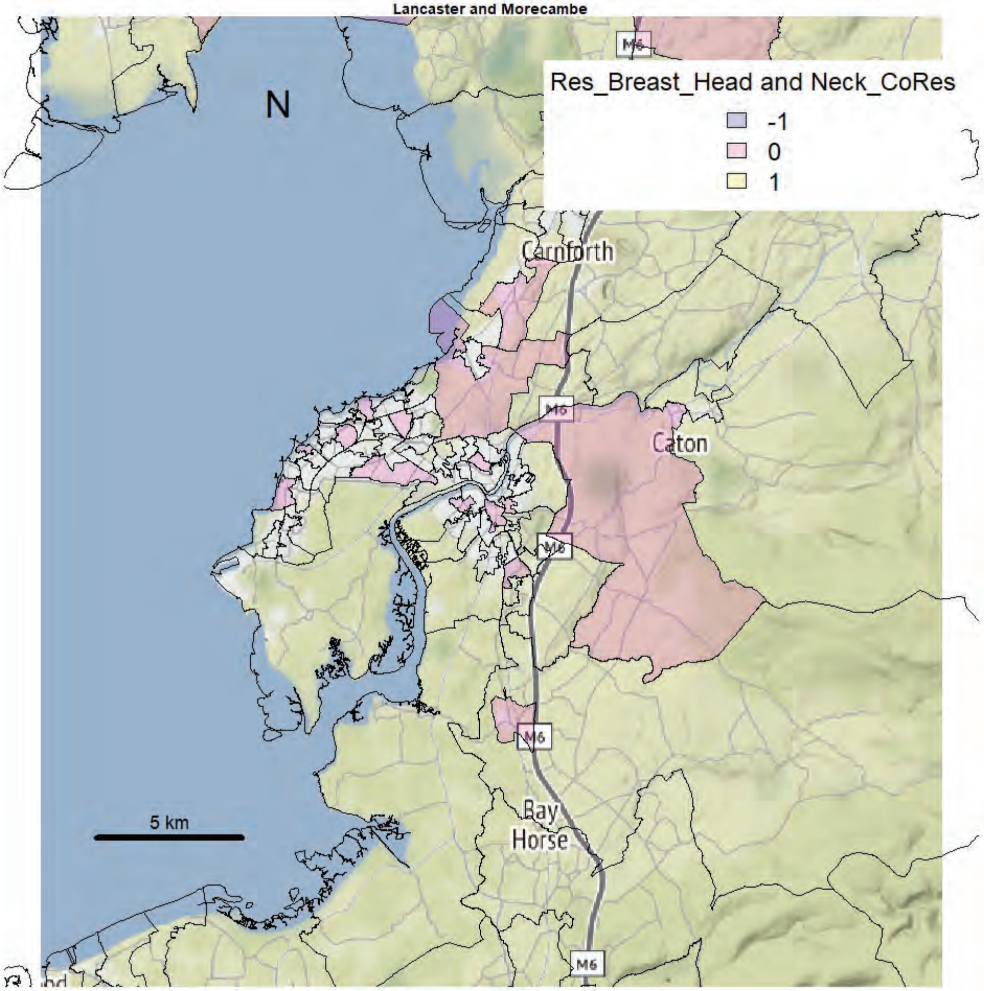

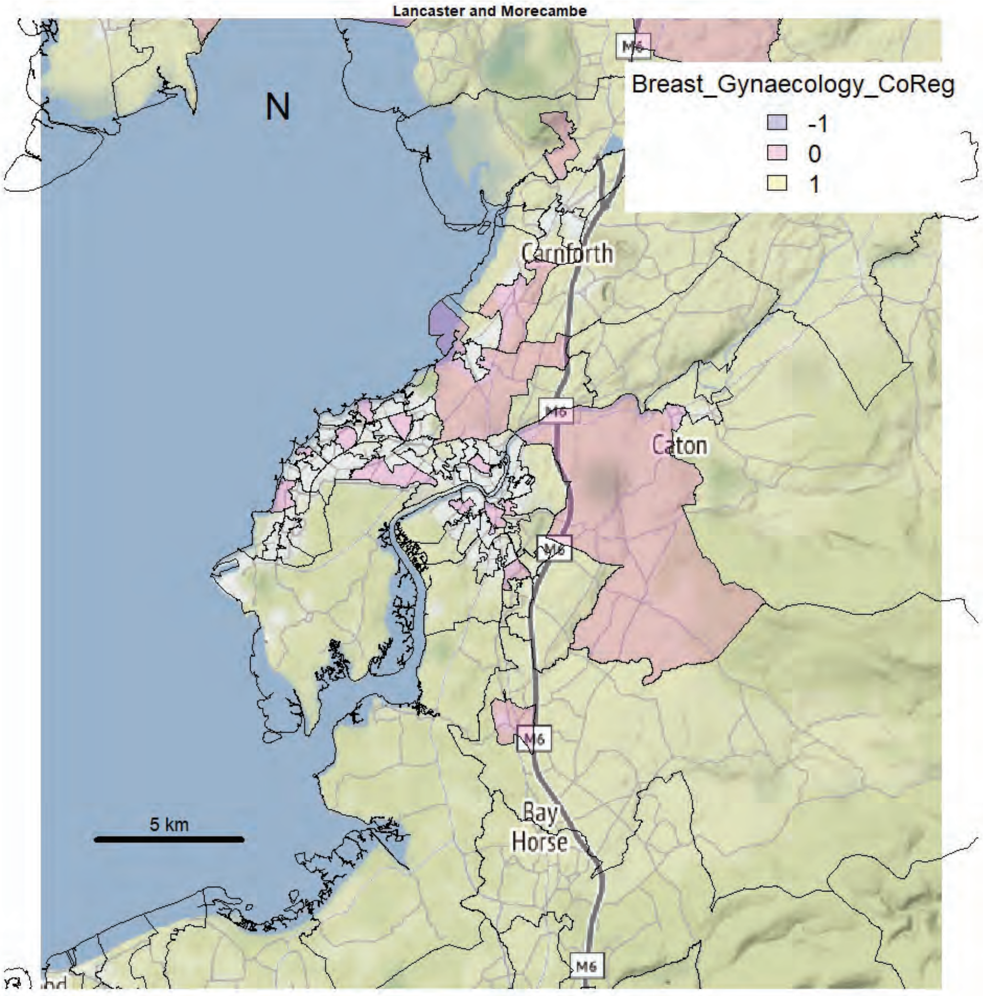
Lancaster and Morecambe
Breast_Gynaecology_CoReg
-1
0
1
N
Carnforth
Caton
M6
Bay
Horse
5 km

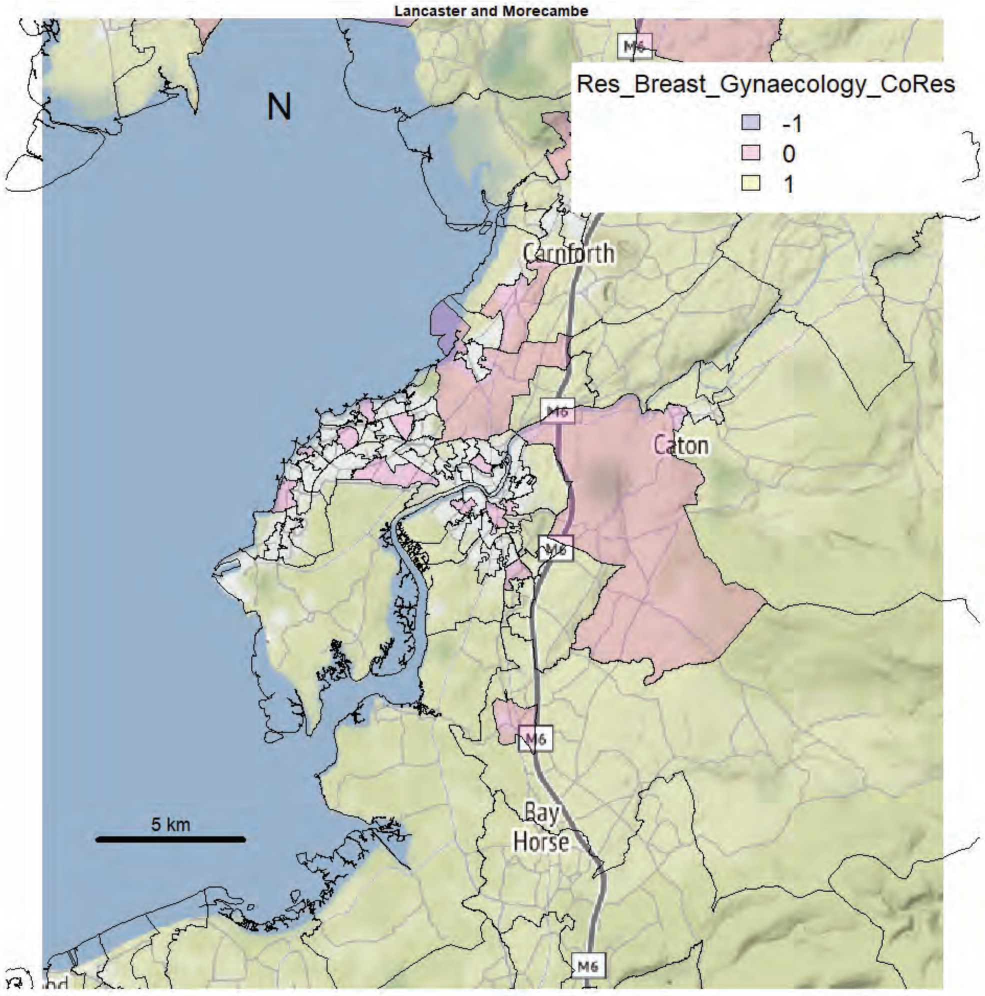
Lancaster and Morecambe
Res_Breast_Gynaecology_CoRes
-1
0
1
N
Carnforth
Caton
M6
Bay
Horse
5 km

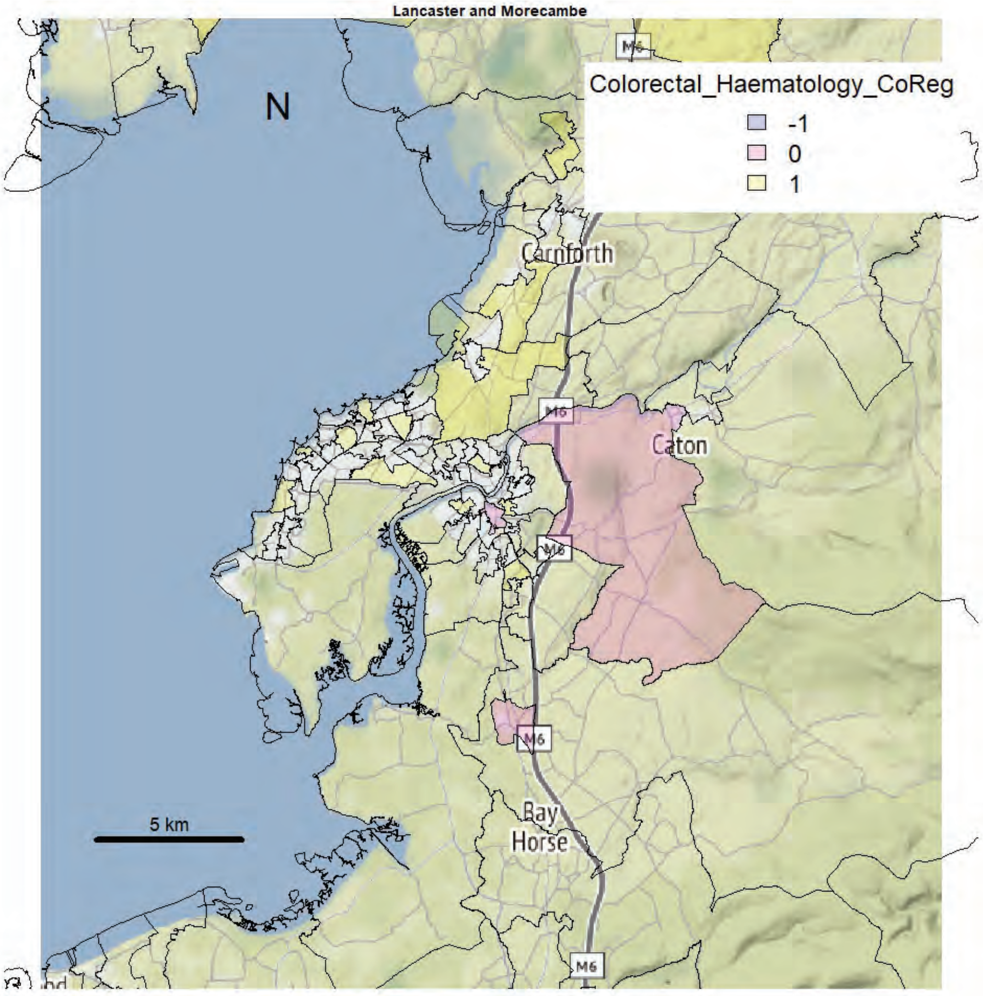
Lancaster and Morecambe
N
Colorectal_Haematology_CoReg
-1
0
1
Carnforth
Caton
Bay Horse
M6
5 km

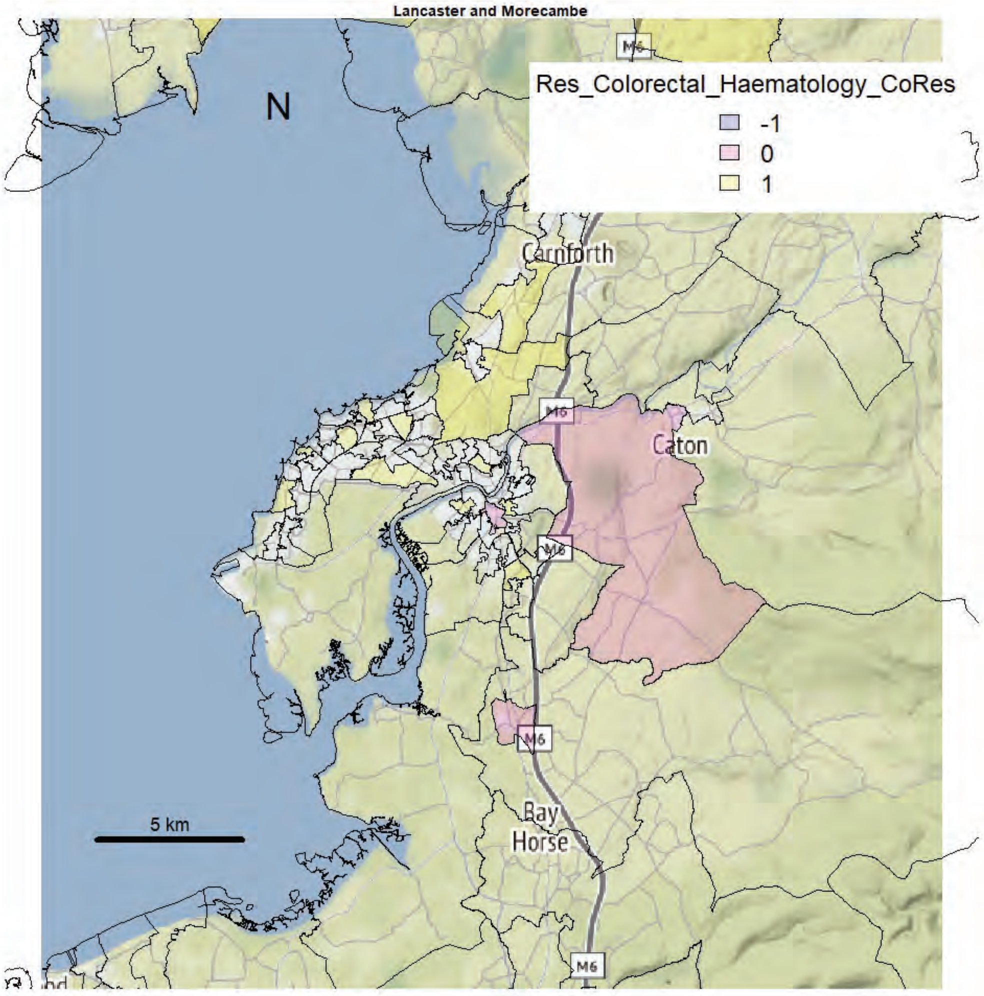
Lancaster and Morecambe
N
Res_Colorectal_Haematology_CoRes
-1
0
1
Carnforth
Caton
Bay Horse
M6
5 km

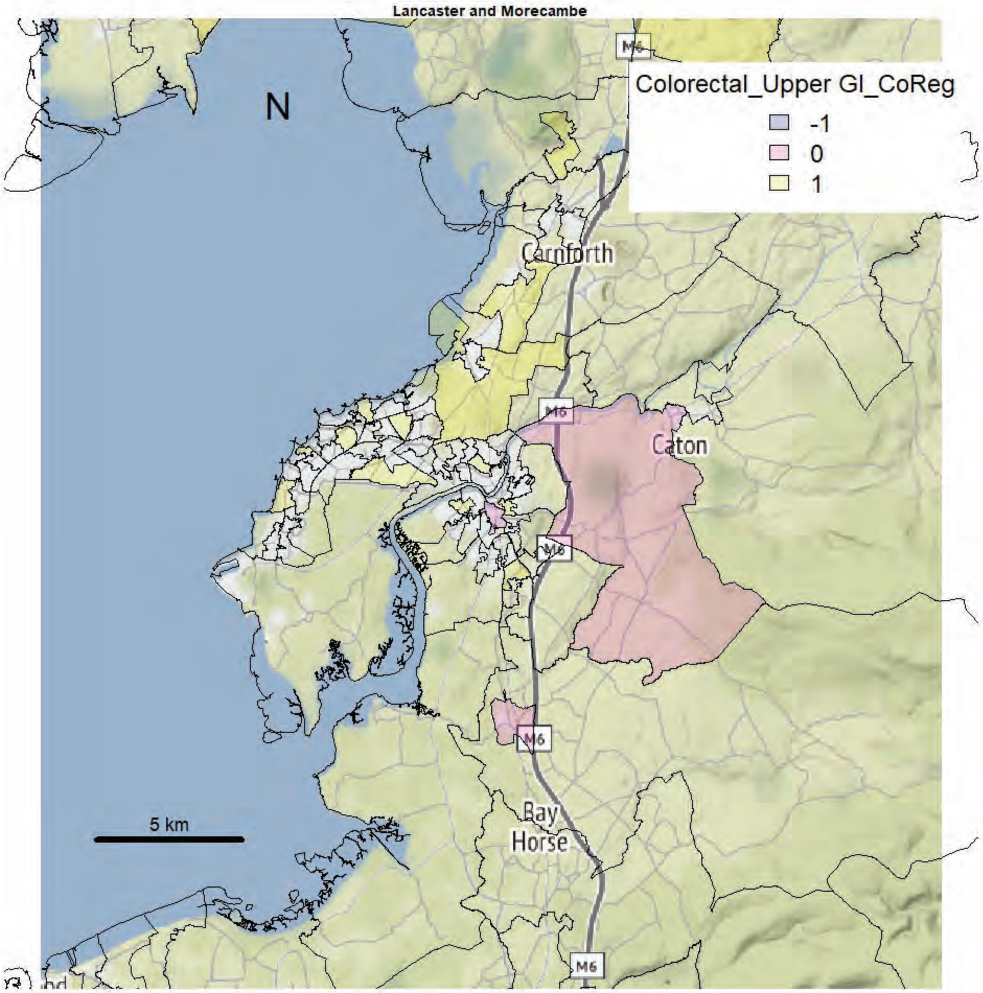
Lancaster and Morecambe
N
Colorectal_Upper GI_CoReg
-1
0
1
Carnforth
Caton
M6
Bay
Horse
5 km

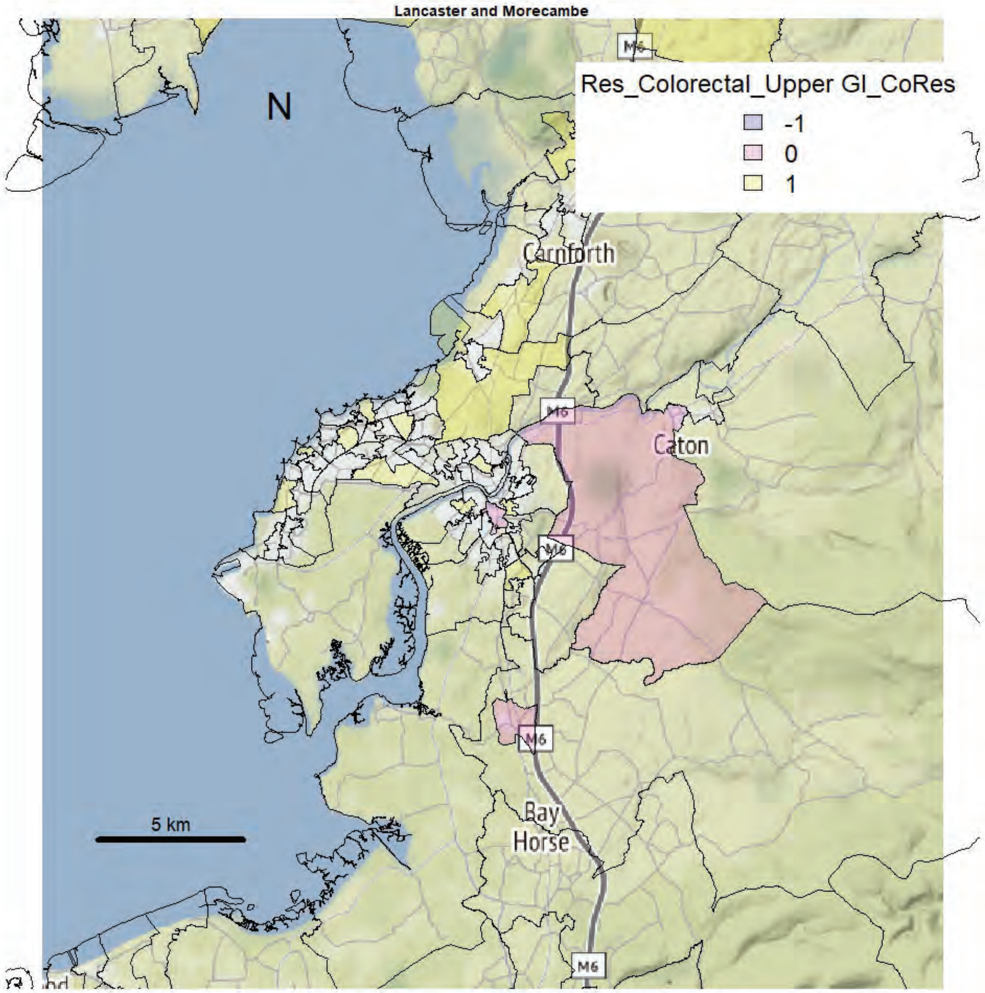
Lancaster and Morecambe
N
Res_Colorectal_Upper GI_CoRes
-1
0
1
Carnforth
Caton
M6
Bay
Horse
5 km

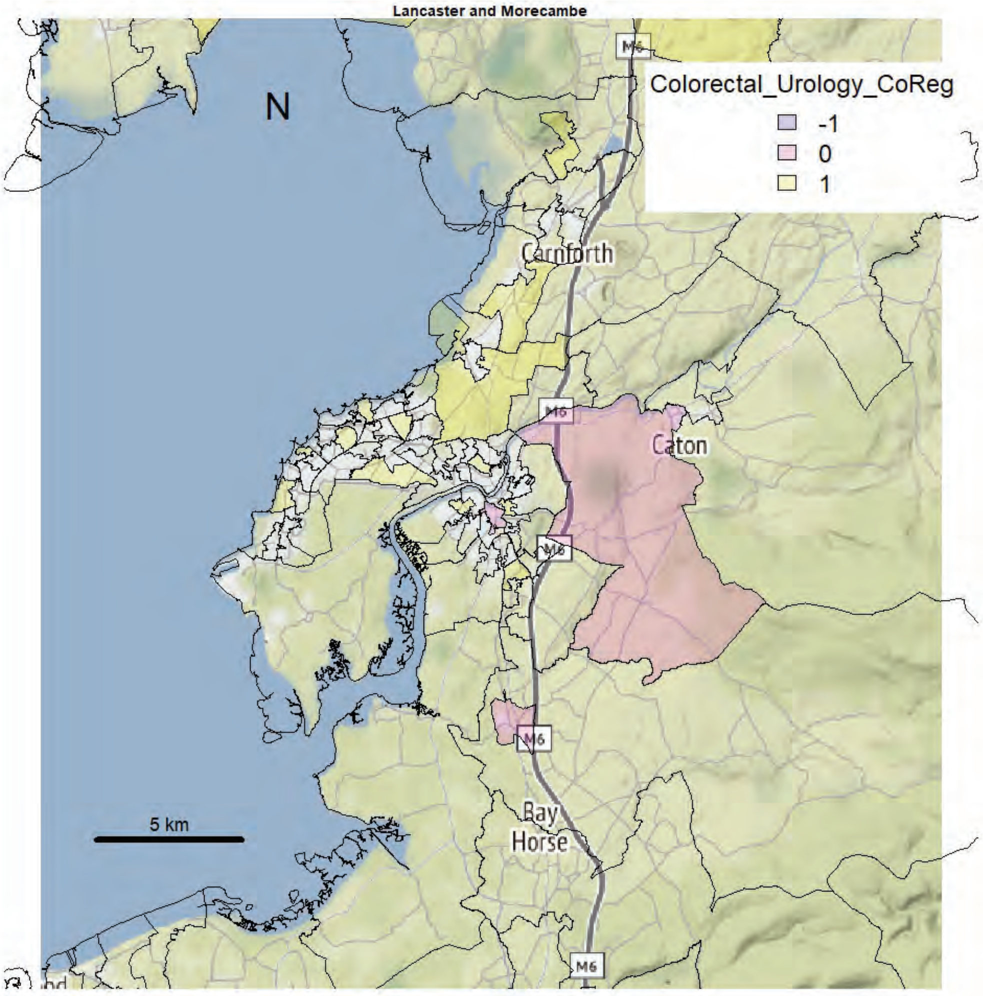
Lancaster and Morecambe
Colorectal_Urology_CoReg
-1
0
1
N
Carnforth
Caton
Bay
Horse
5 km
M6

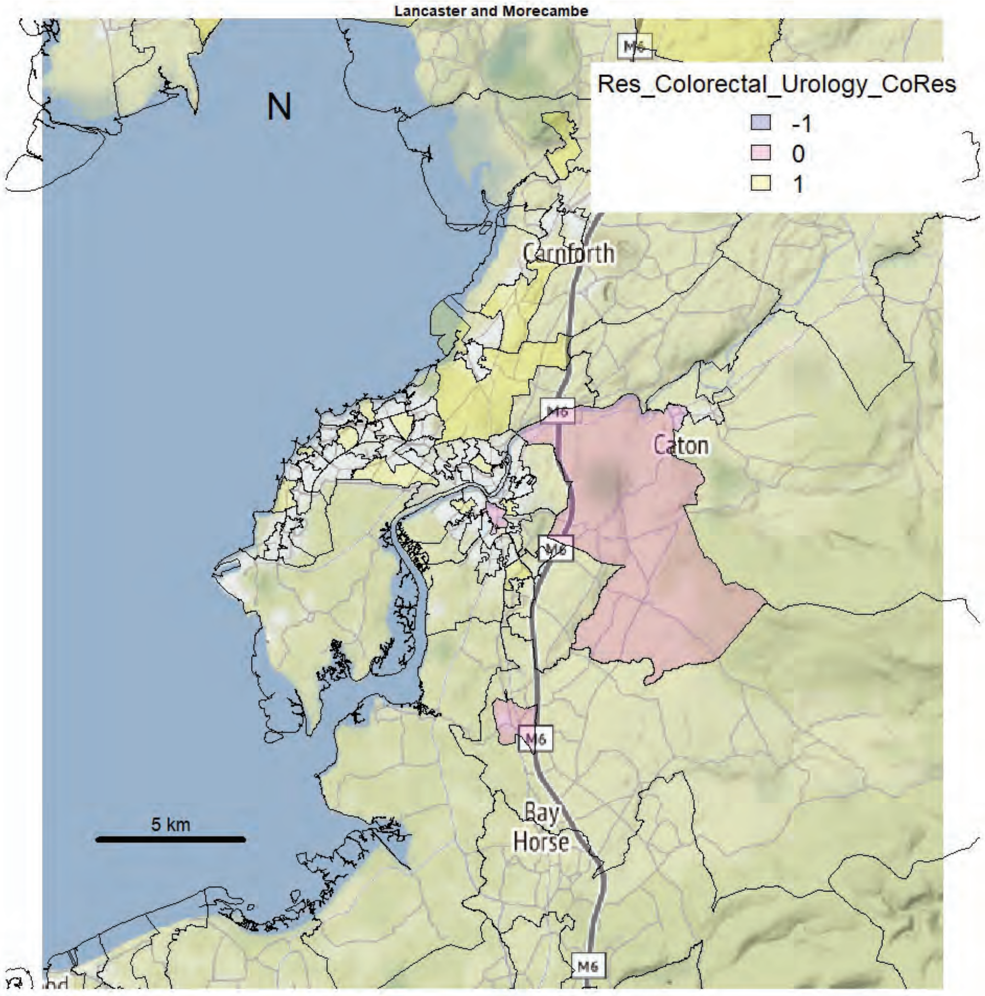
Lancaster and Morecambe
Res_Colorectal_Urology_CoRes
-1
0
1
N
Carnforth
Caton
Bay
Horse
5 km
M6

Lancaster and Morecambe

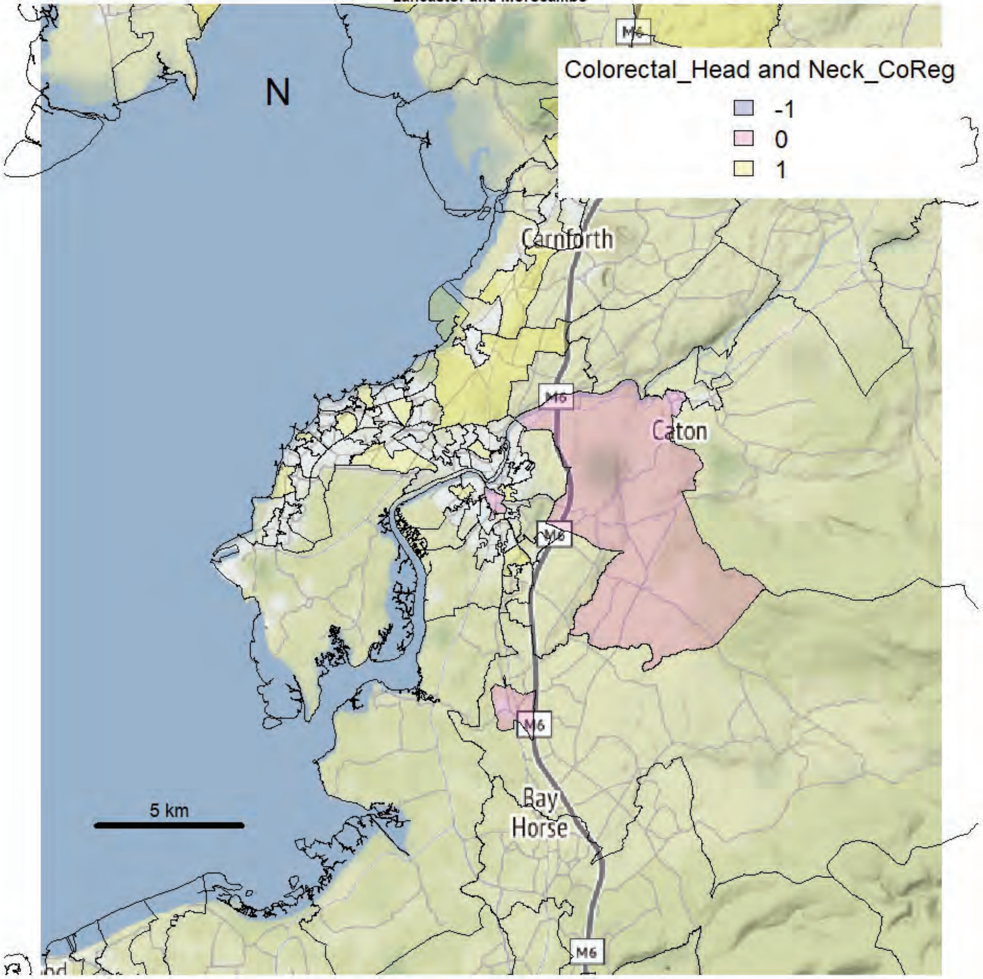


Lancaster and Morecambe

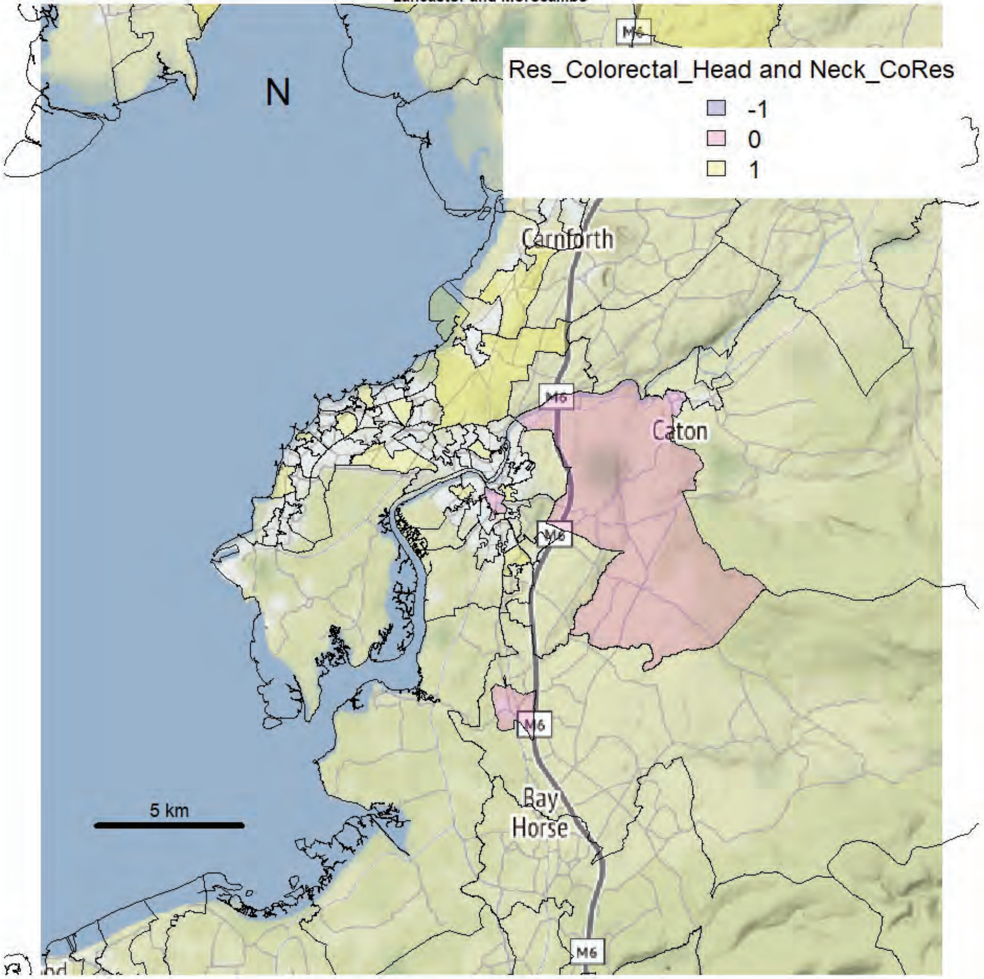

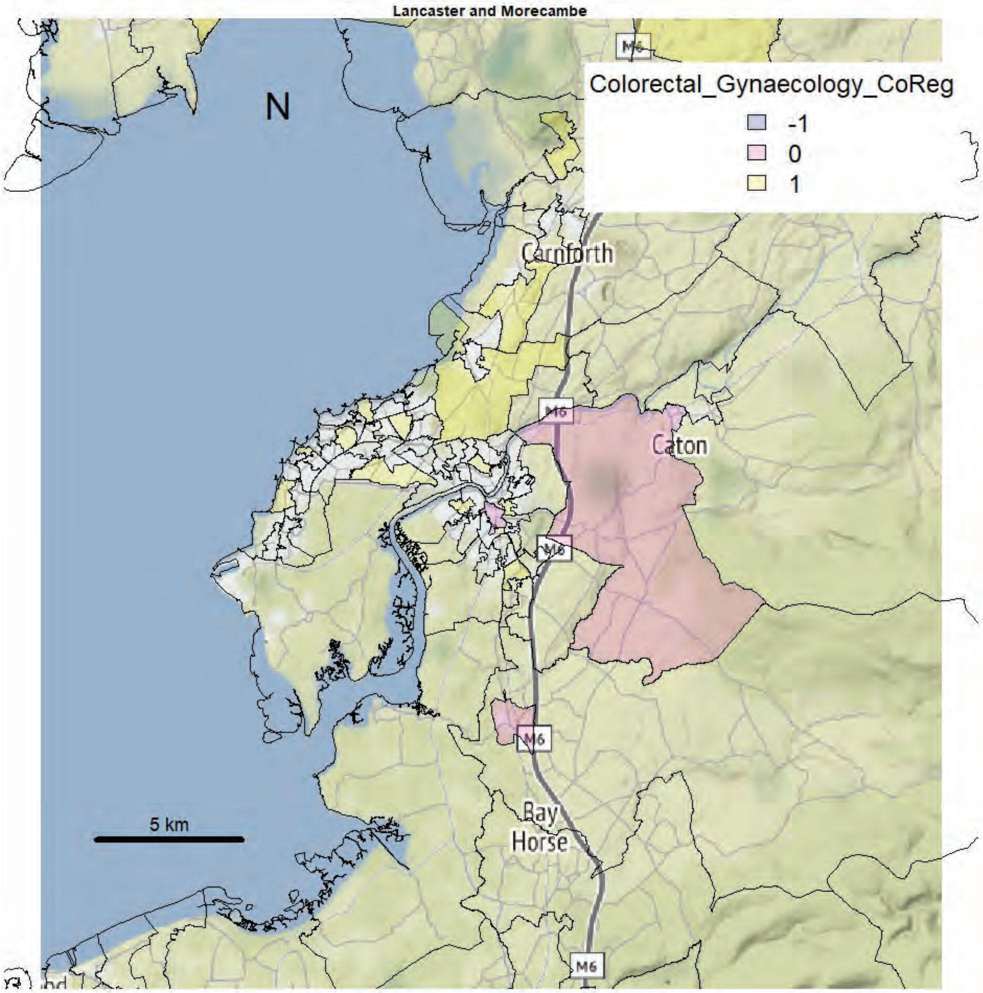
Lancaster and Morecambe
N
Colorectal_Gynaecology_CoReg
-1
0
1
Carnforth
Caton
Bay Horse
M6
5 km

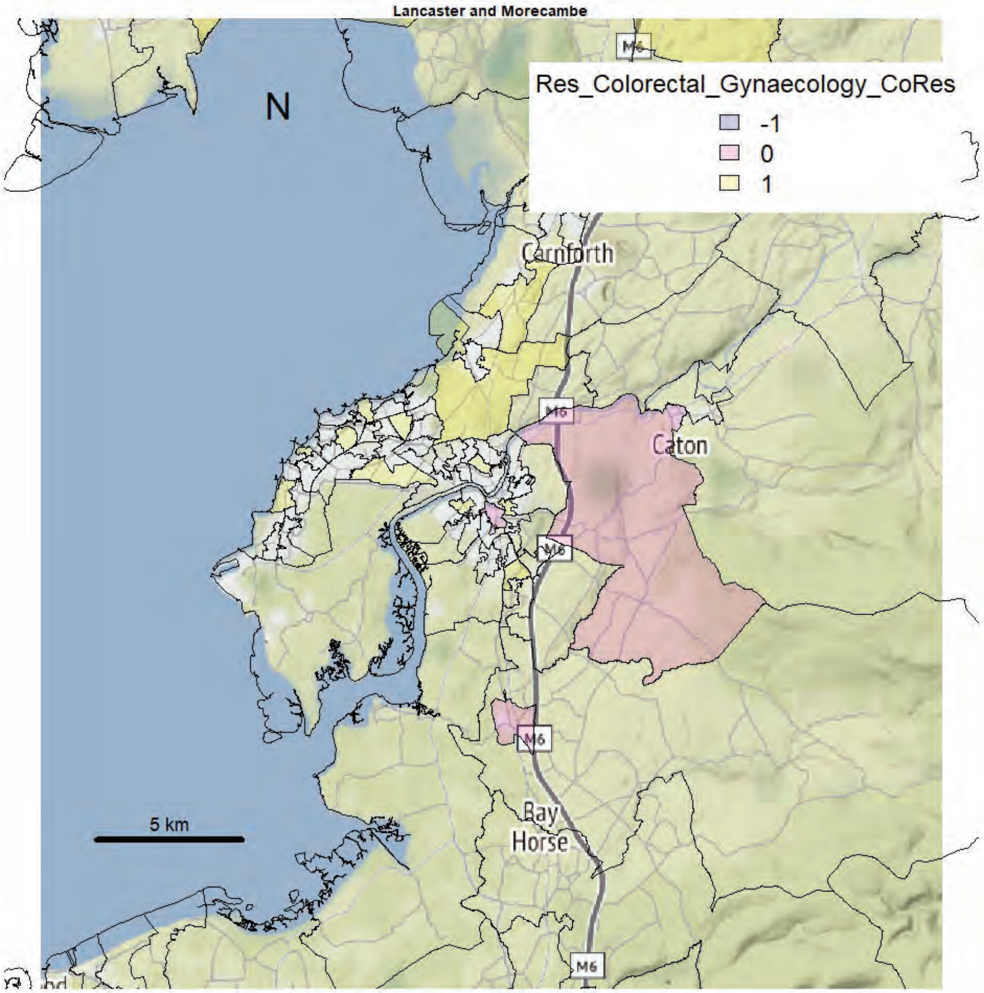
Lancaster and Morecambe
N
Res_Colorectal_Gynaecology_CoRes
-1
0
1
Carnforth
Caton
Bay Horse
M6
5 km

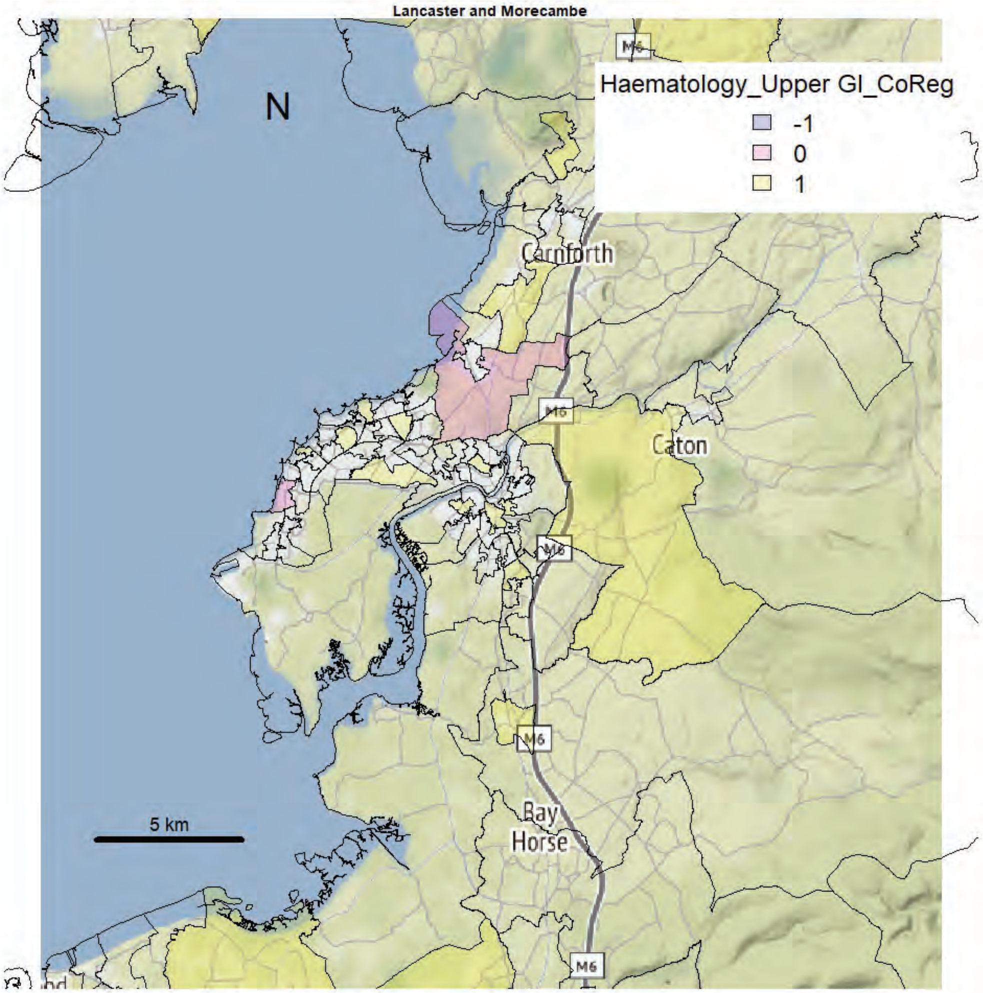
Lancaster and Morecambe
Haematology_Upper GI_CoReg
-1
0
1
N
Carnforth
Caton
M6
Bay
Horse
5 km

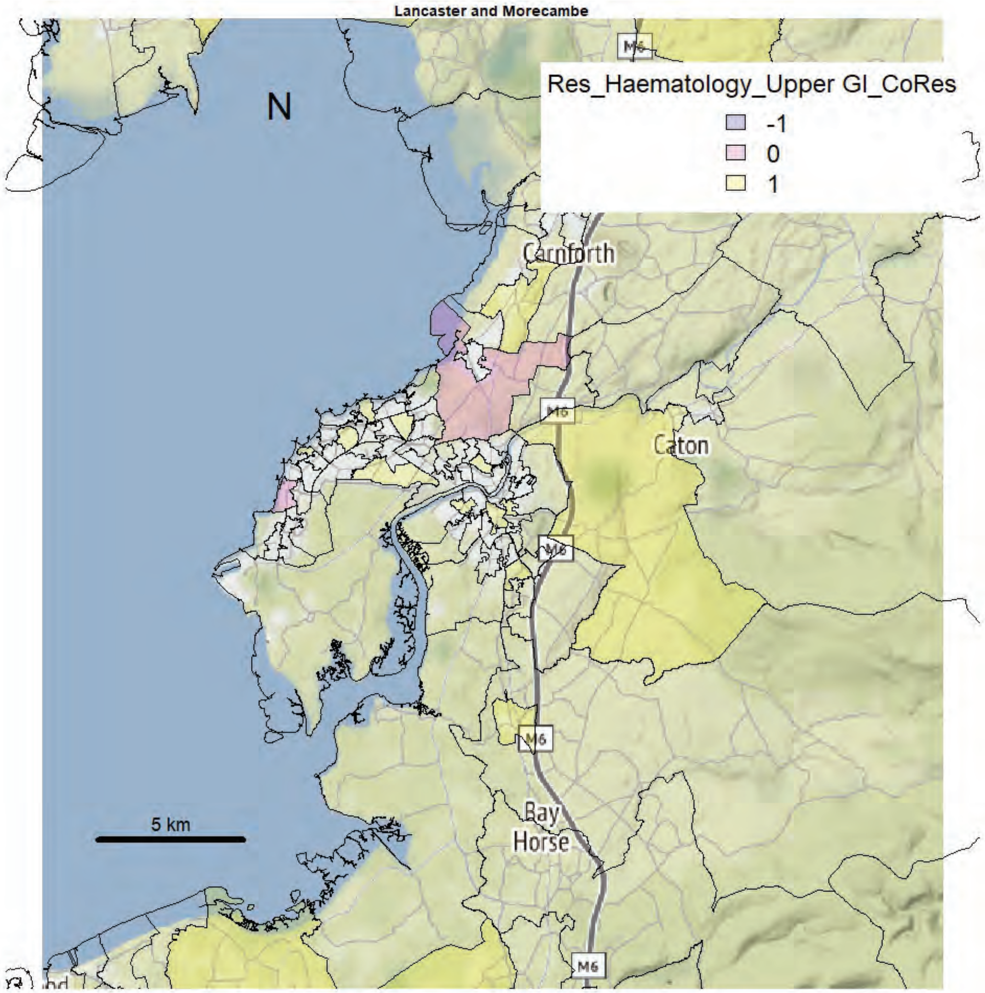
Lancaster and Morecambe
Res_Haematology_Upper GI_CoRes
-1
0
1
N
Carnforth
Caton
M6
Bay
Horse
5 km

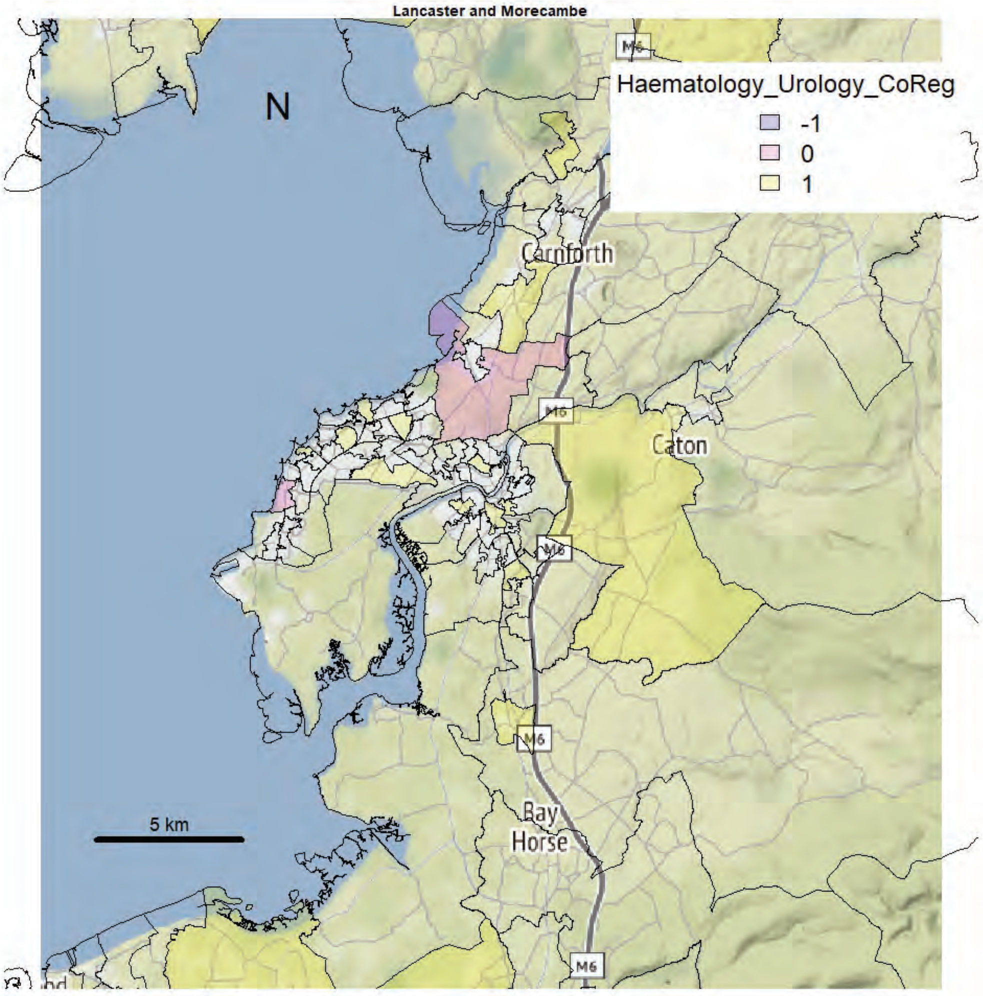
Lancaster and Morecambe
Haematology_Urology_CoReg
-1
0
1
N
Carnforth
Caton
M6
Bay Horse
5 km

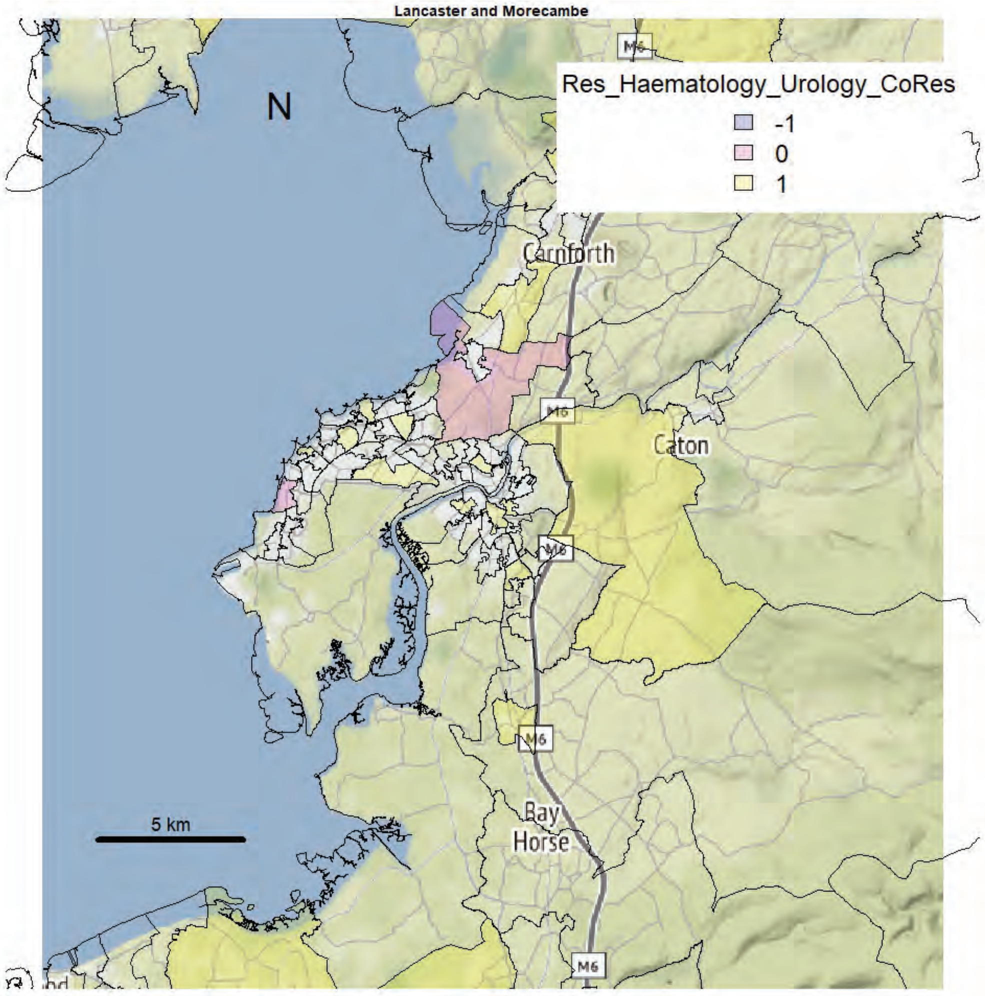
Lancaster and Morecambe
Res_Haematology_Urology_CoRes
-1
0
1
N
Carnforth
Caton
M6
Bay Horse
5 km

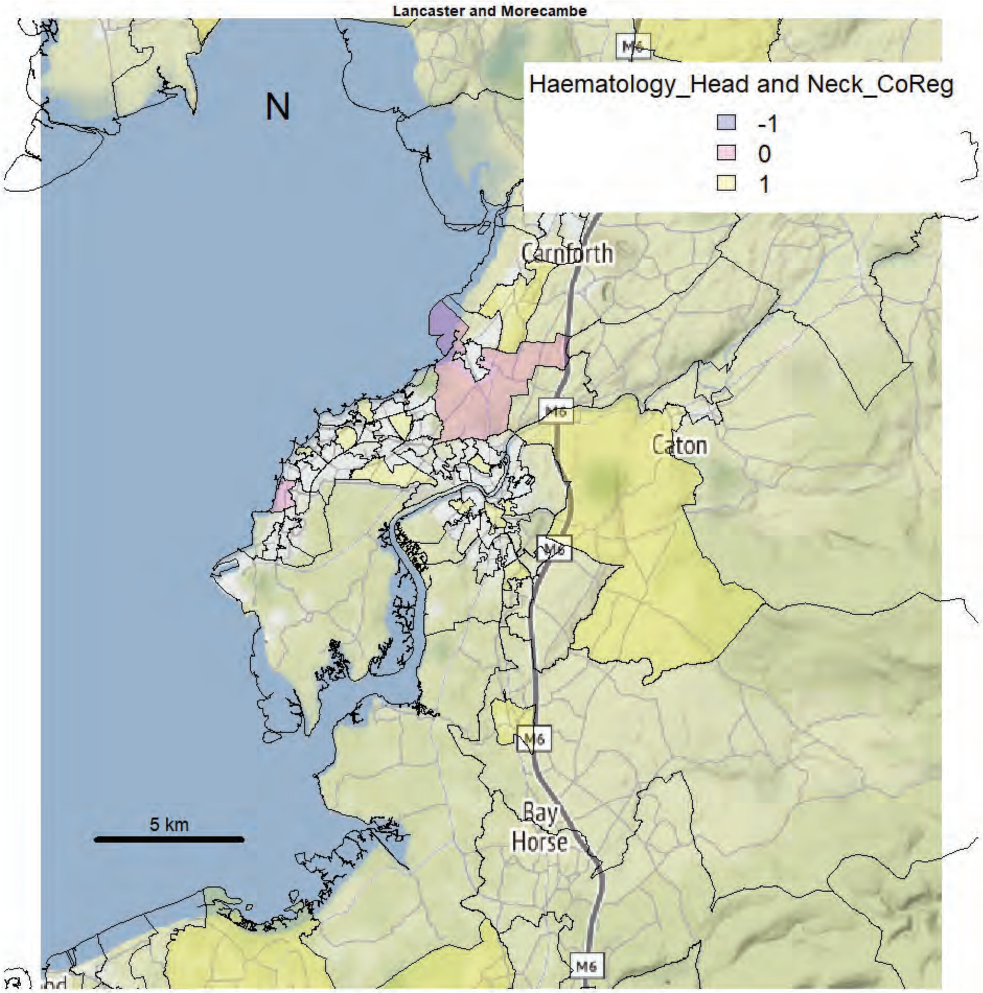
Lancaster and Morecambe
Haematology_Head and Neck_CoReg
-1
0
1
N
Carnforth
Caton
M6
Bay
Horse
5 km

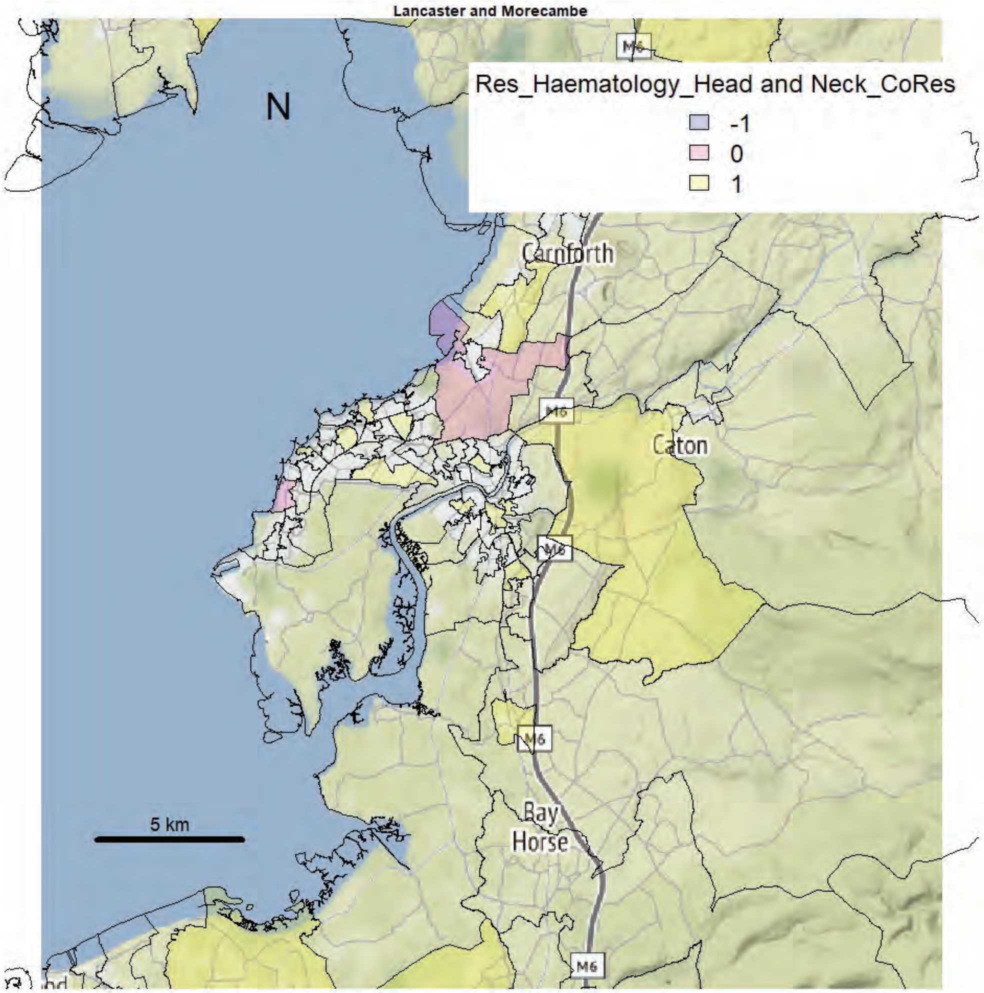
Lancaster and Morecambe
Res_Haematology_Head and Neck_CoRes
-1
0
1
N
Carnforth
Caton
M6
Bay
Horse
5 km

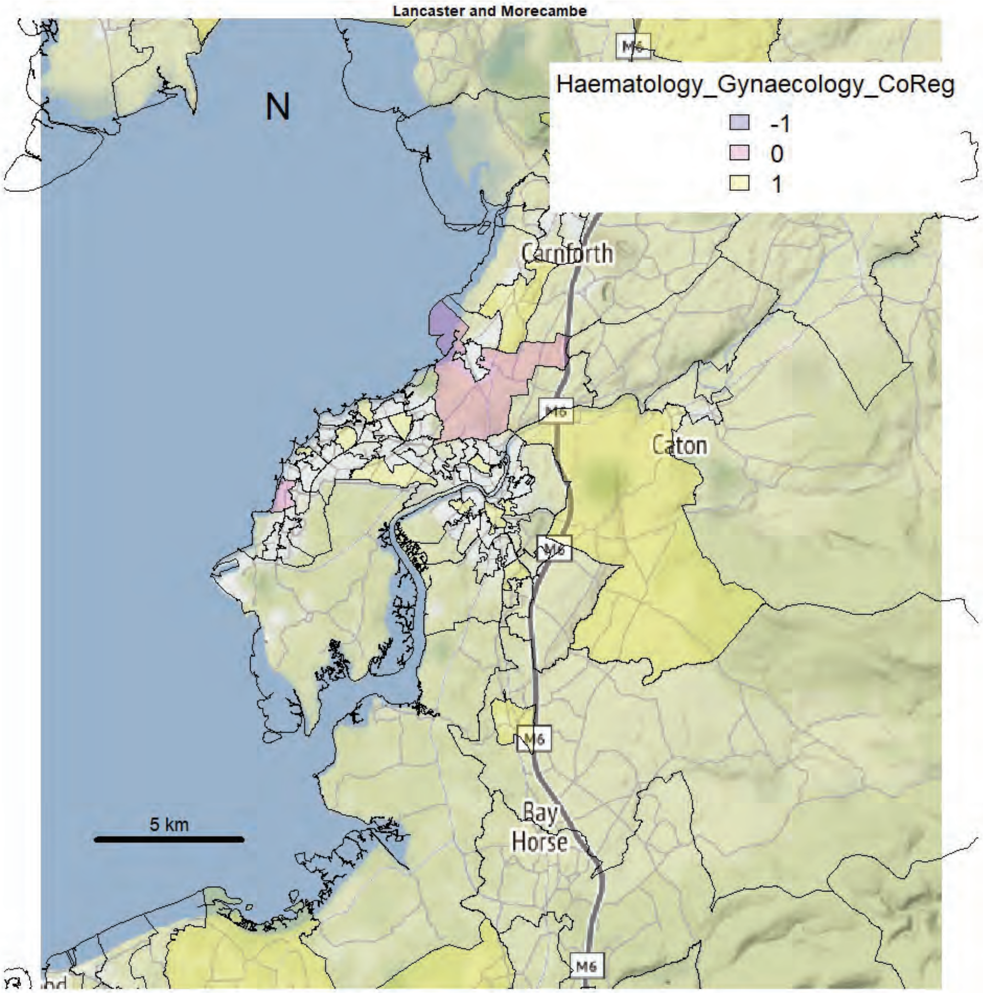

Lancaster and Morecambe
Haematology_Gynaecology_CoReg
-1
0
1
N
5 km
Carnforth
Caton
Bay Horse
M6


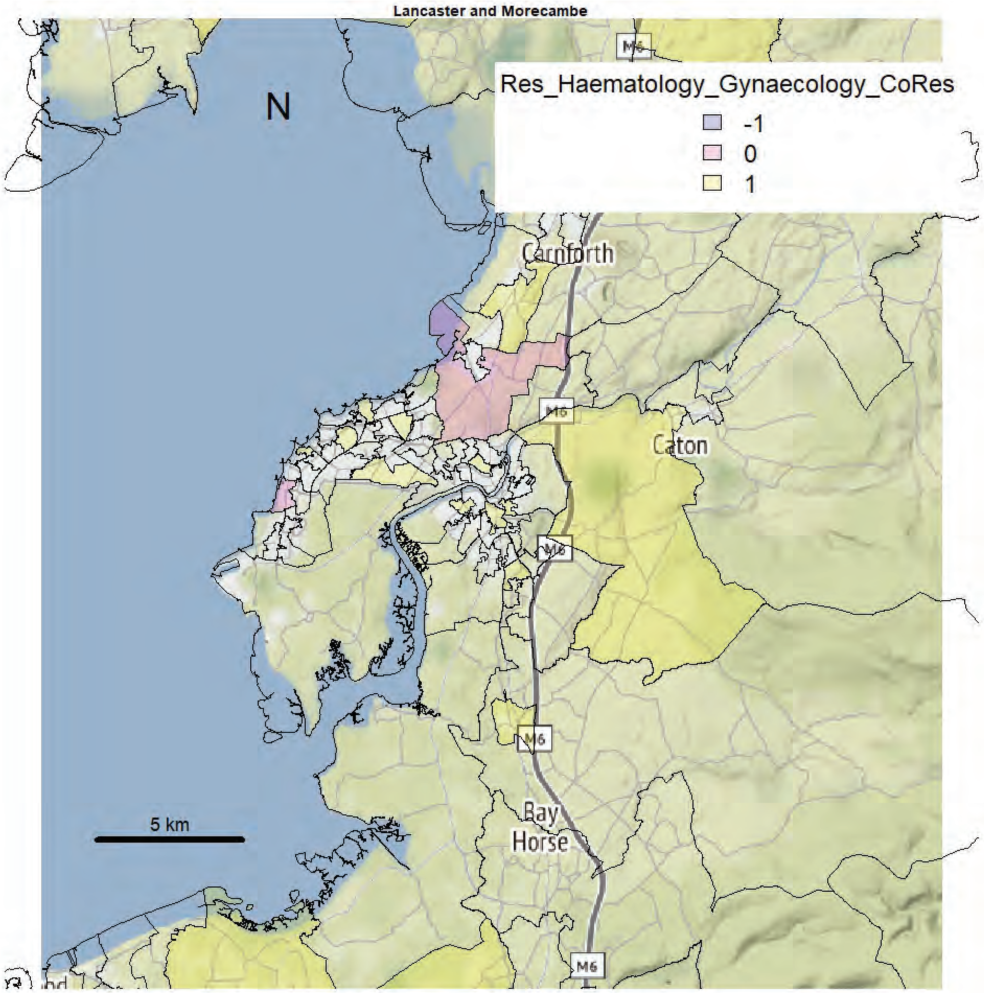

Lancaster and Morecambe
Res_Haematology_Gynaecology_CoRes
-1
0
1
N
5 km
Carnforth
Caton
Bay Horse
M6

Lancaster and Morecambe

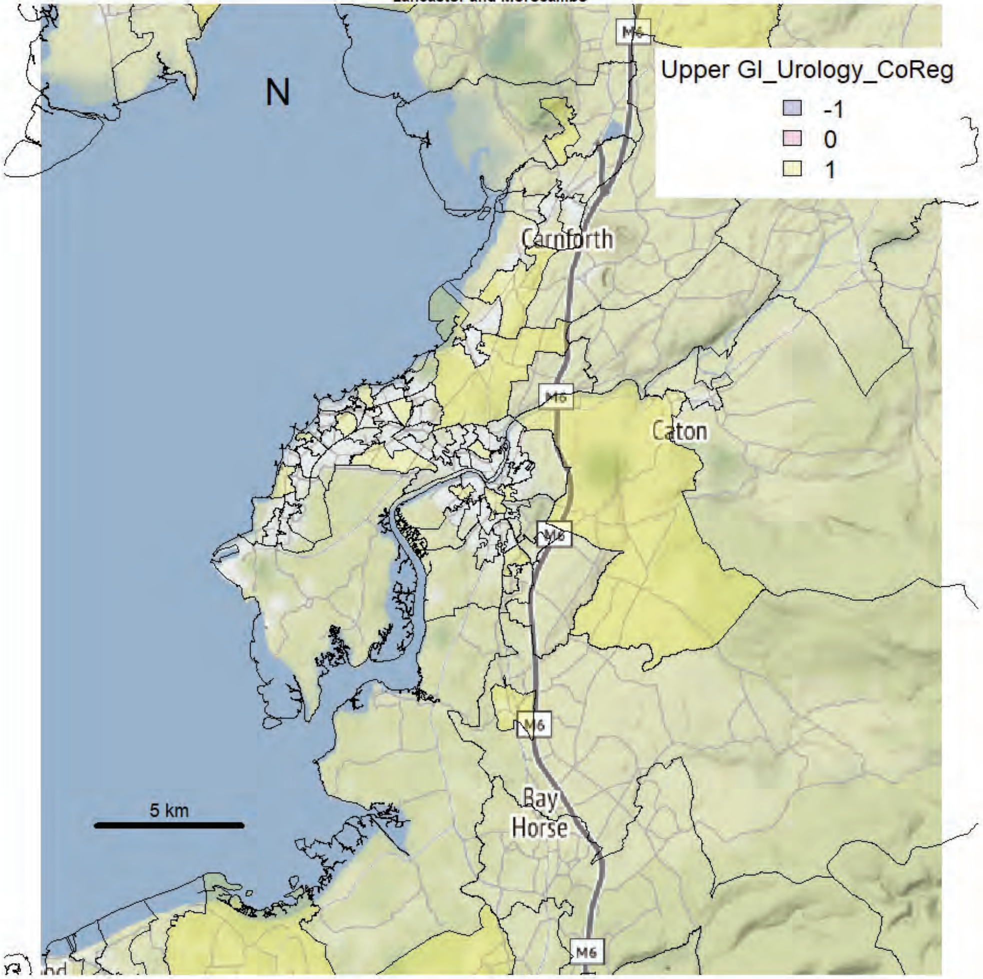


Lancaster and Morecambe

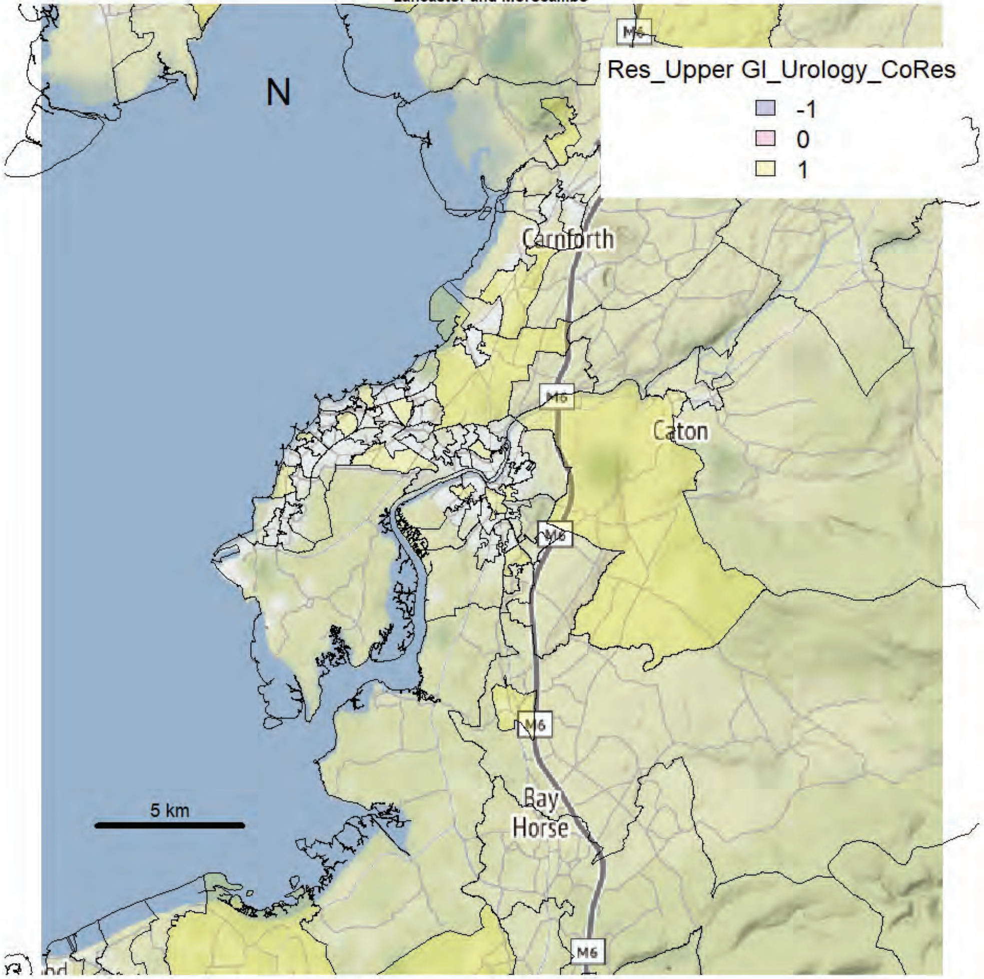

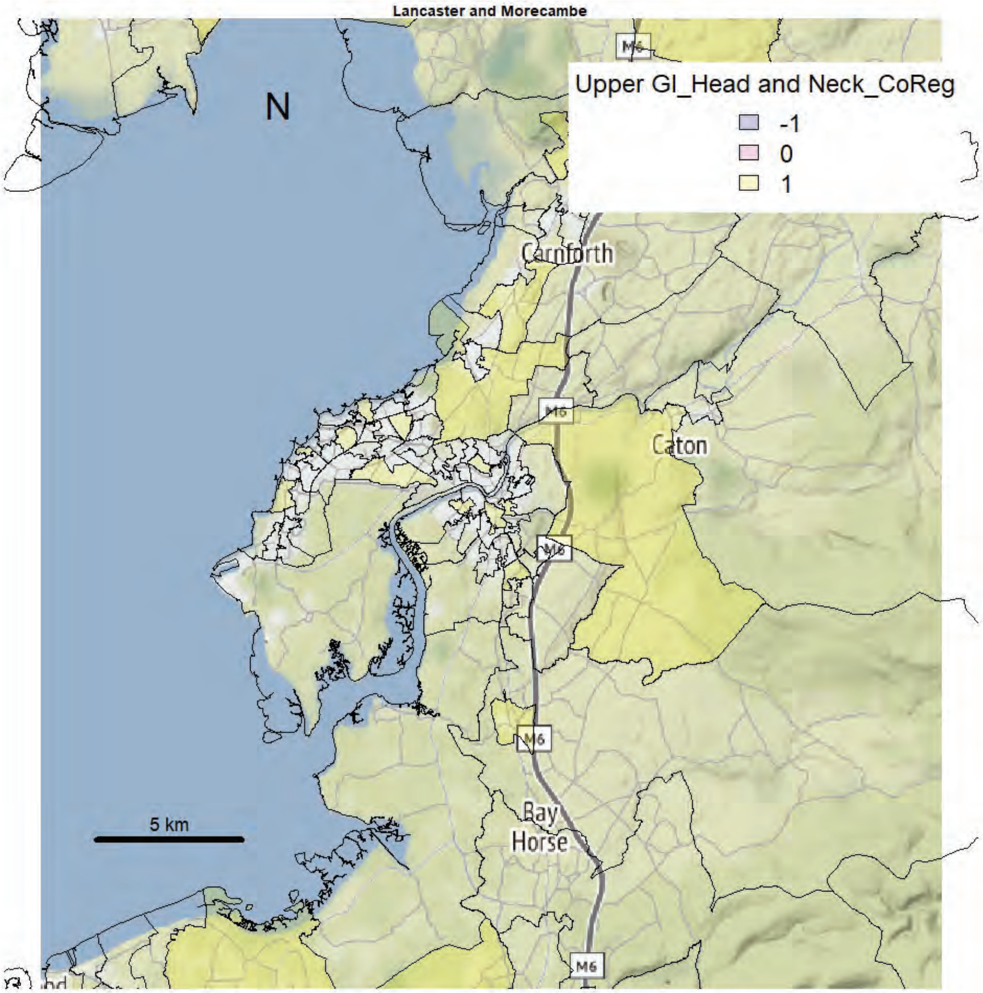
Lancaster and Morecambe
Upper GI_Head and Neck_CoReg
-1
0
1
N
Carnforth
Caton
Bay Horse
M6
5 km

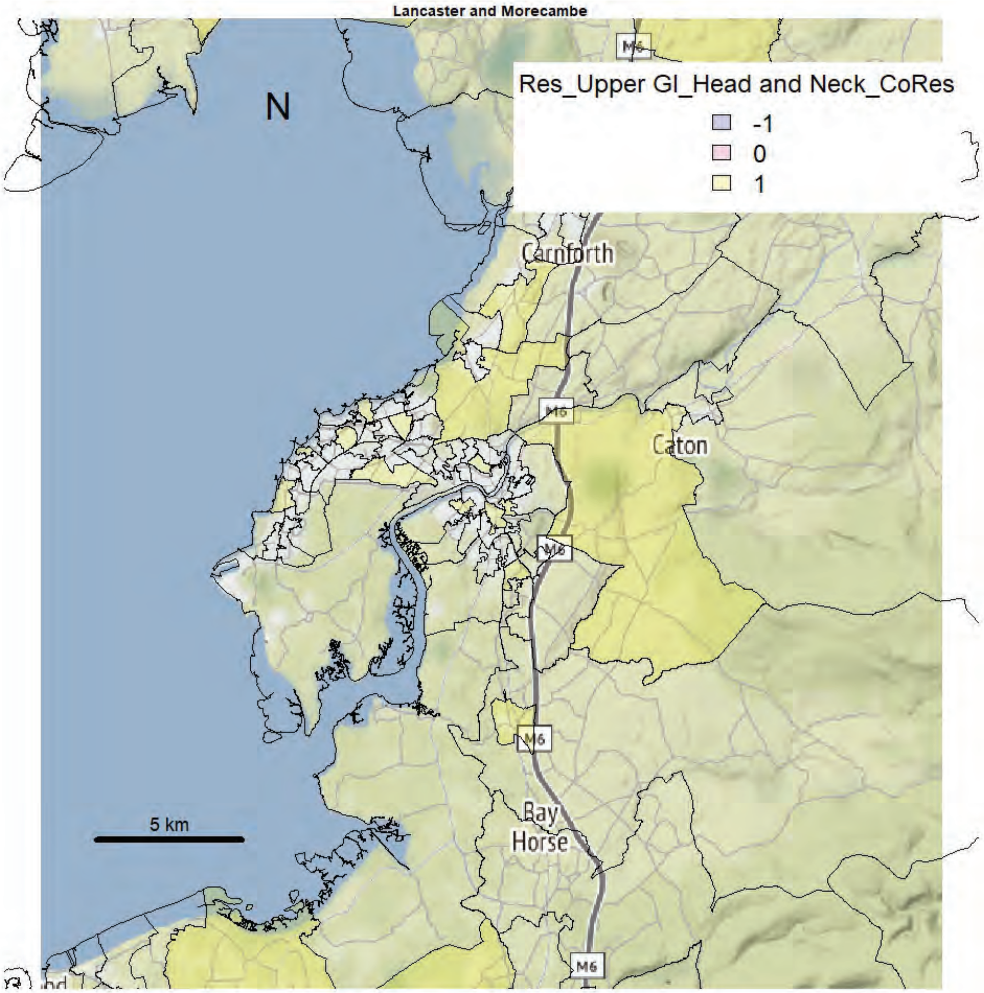
Lancaster and Morecambe
Res_Upper GI_Head and Neck_CoRes
-1
0
1
N
Carnforth
Caton
Bay Horse
M6
5 km

Lancaster and Morecambe

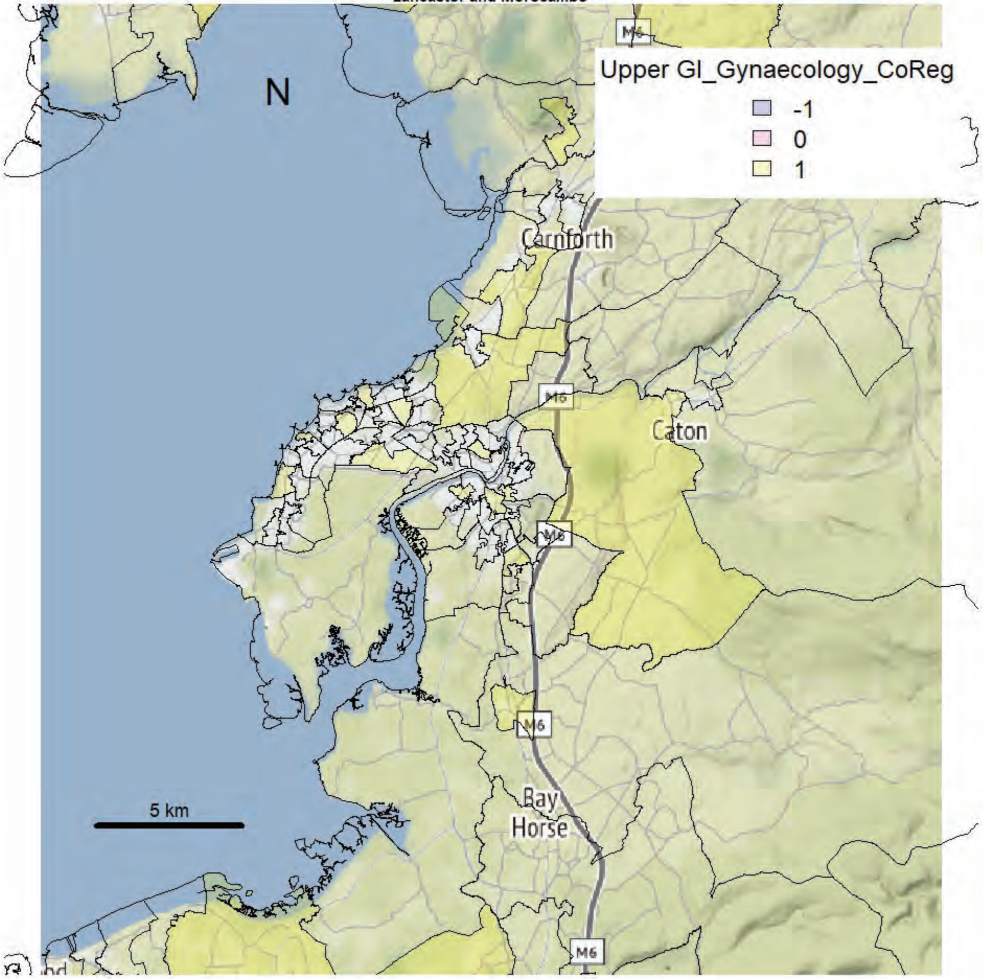


Lancaster and Morecambe

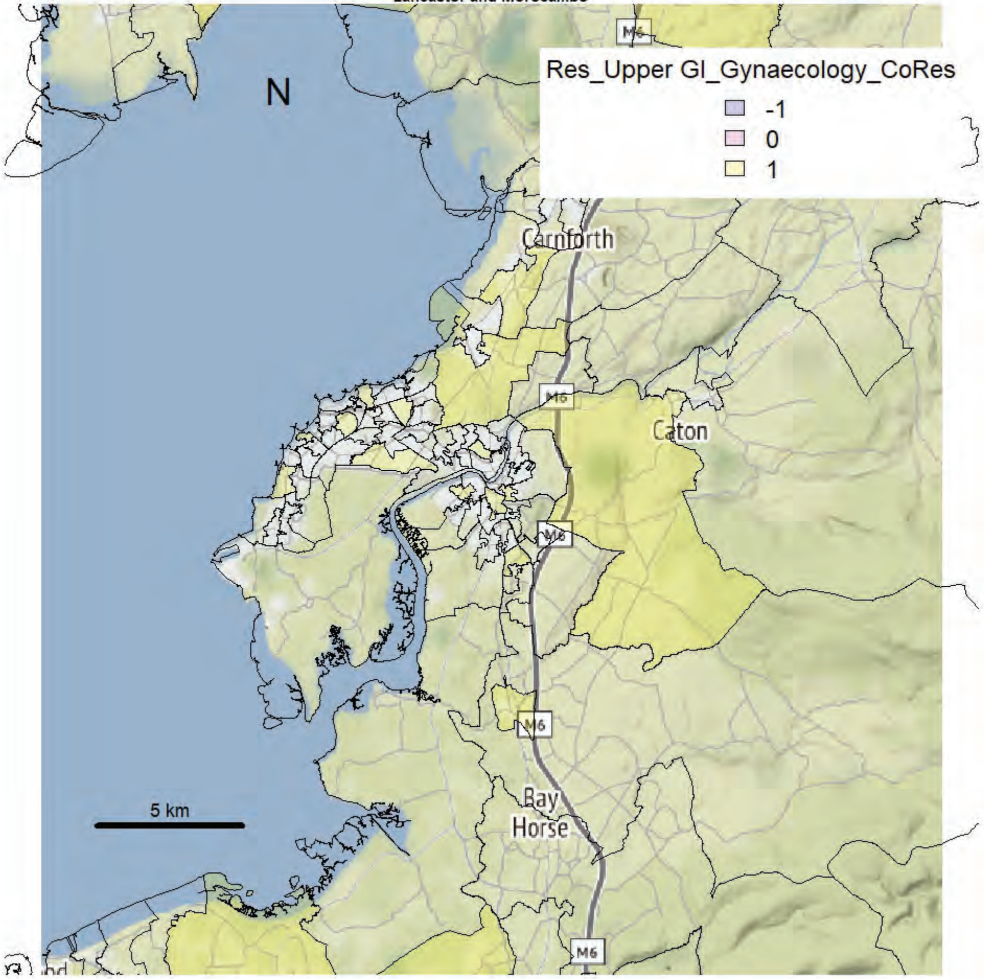

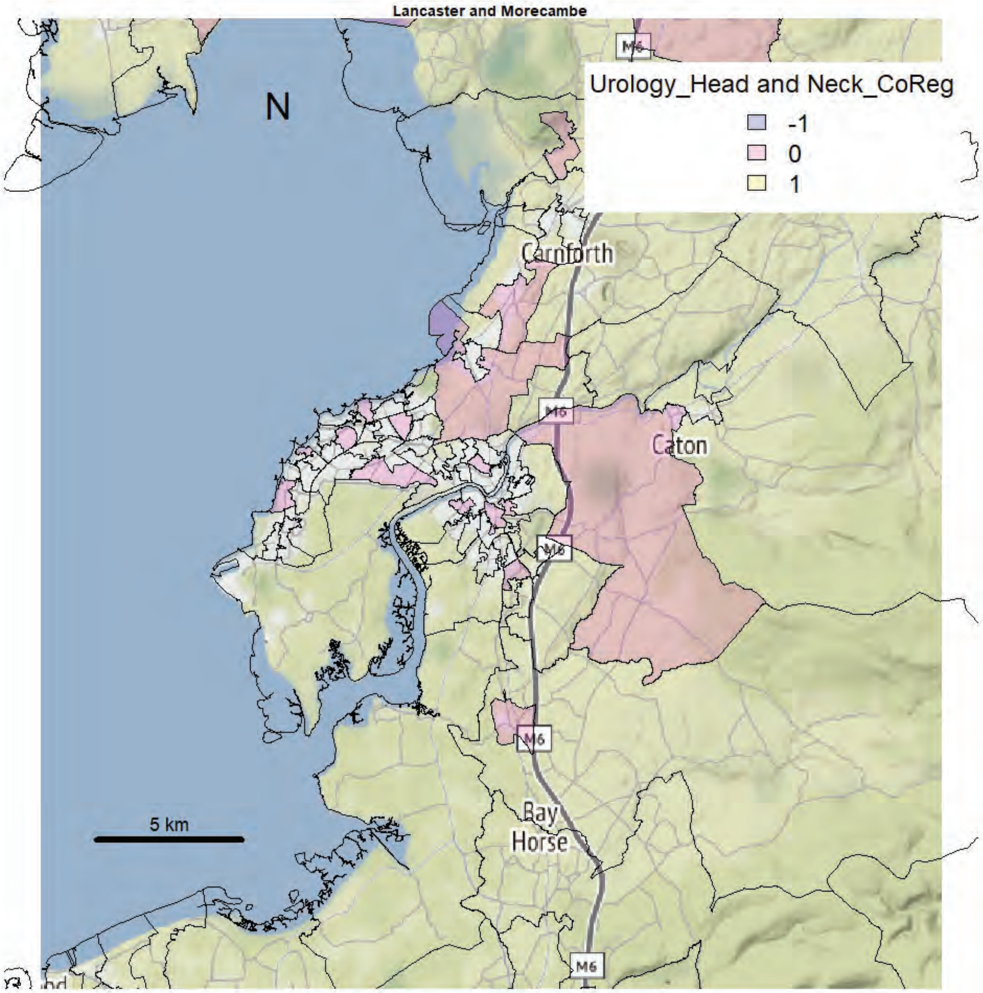
Lancaster and Morecambe
N
Urology_Head and Neck_CoReg
-1
0
1
Carnforth
Caton
M6
Bay
Horse
5 km

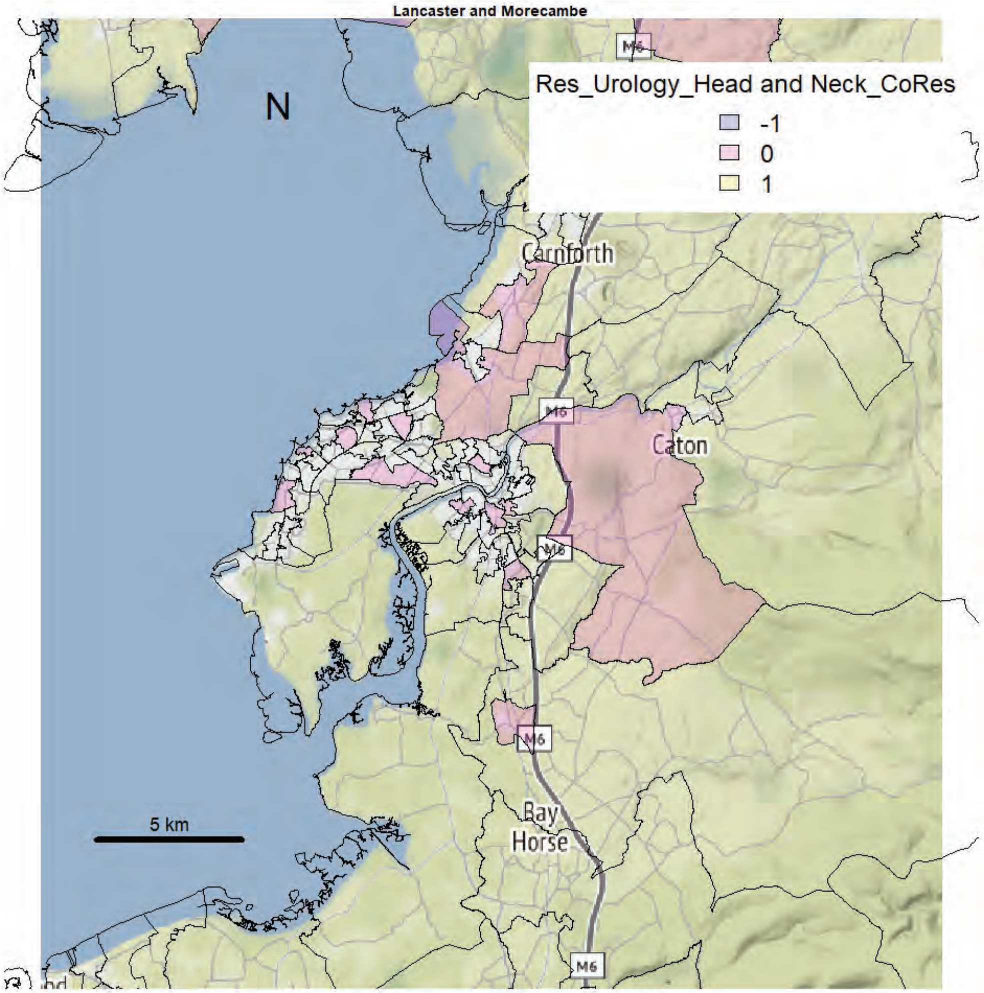
Lancaster and Morecambe
N
Res_Urology_Head and Neck_CoRes
-1
0
1
Carnforth
Caton
M6
Bay
Horse
5 km

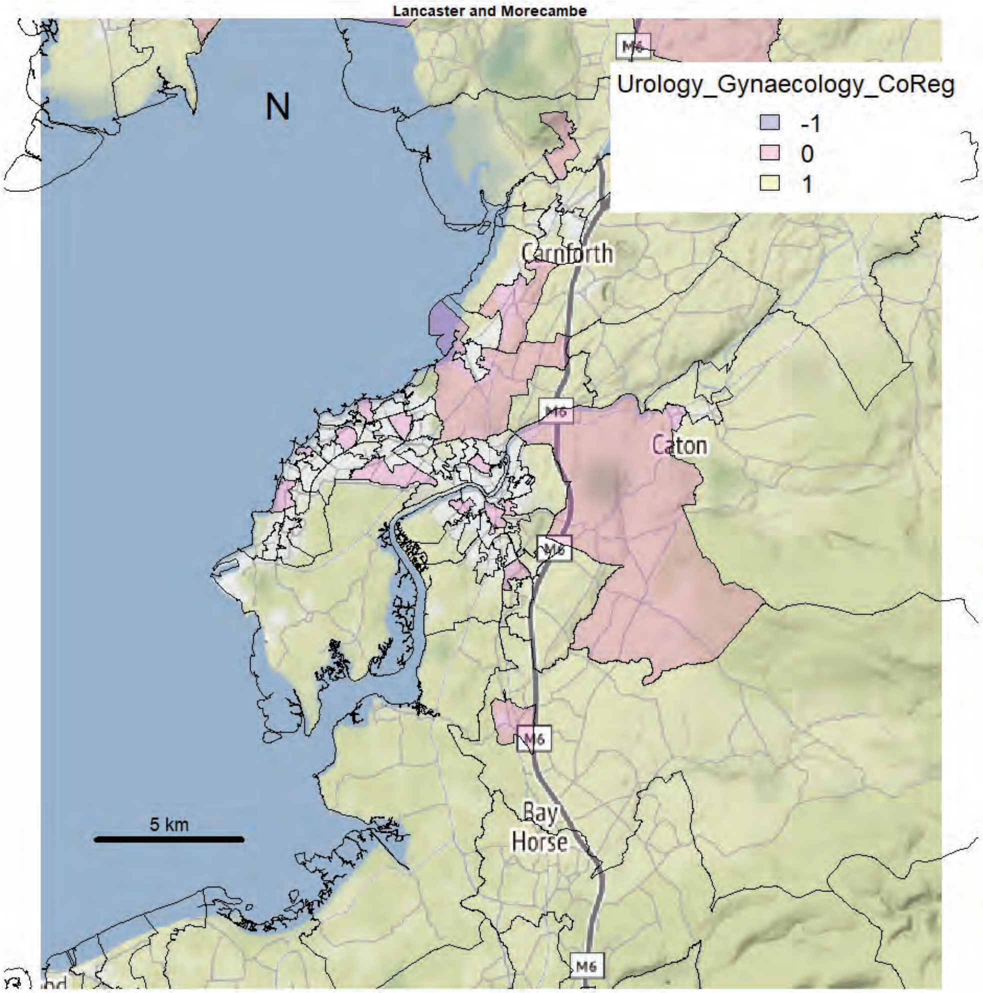
Lancaster and Morecambe
N
Urology_Gynaecology_CoReg
-1
0
1
Carnforth
Caton
M6
Bay
Horse
5 km

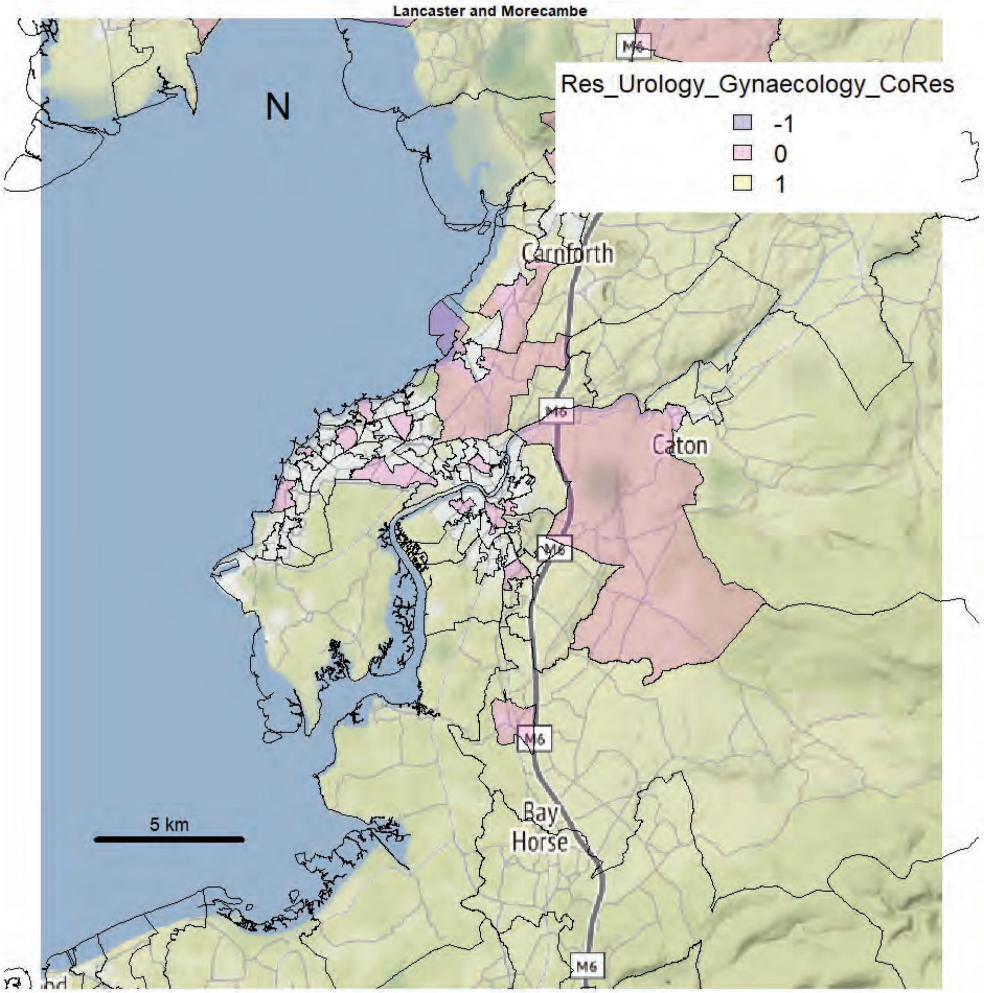
Lancaster and Morecambe
N
Res_Urology_Gynaecology_CoRes
-1
0
1
Carnforth
Caton
M6
Bay
Horse
5 km

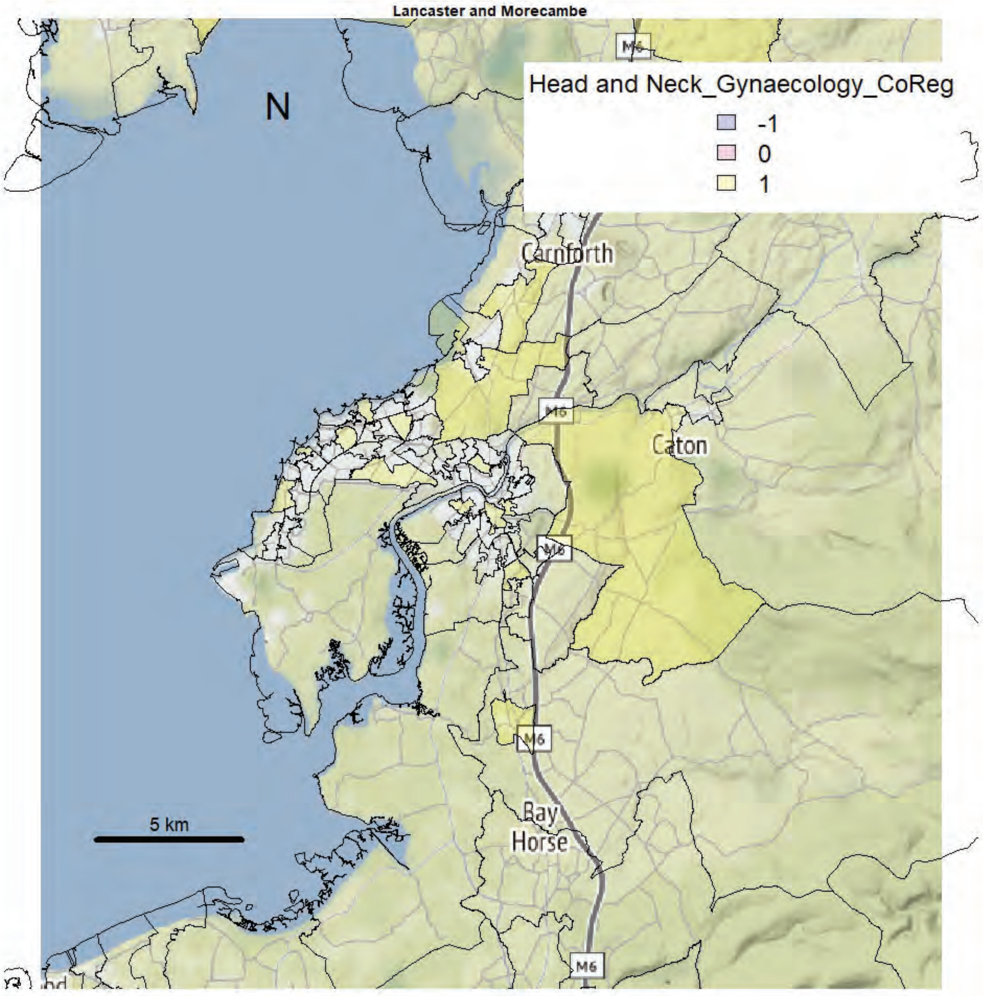
Lancaster and Morecambe
N
Head and Neck_Gynaecology_CoReg
-1
0
1
Carnforth
Caton
Bay Horse
M6
5 km

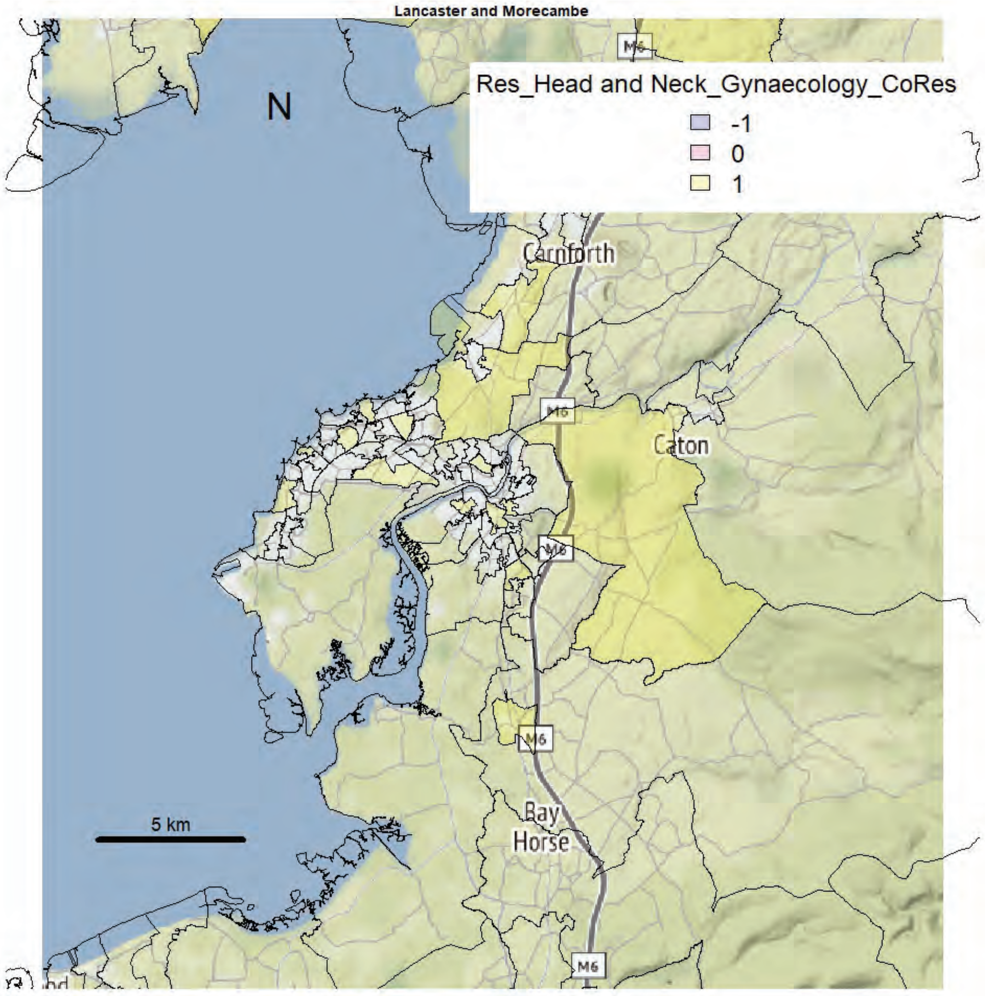
Lancaster and Morecambe
N
Res_Head and Neck_Gynaecology_CoRes
-1
0
1
Carnforth
Caton
Bay Horse
M6
5 km

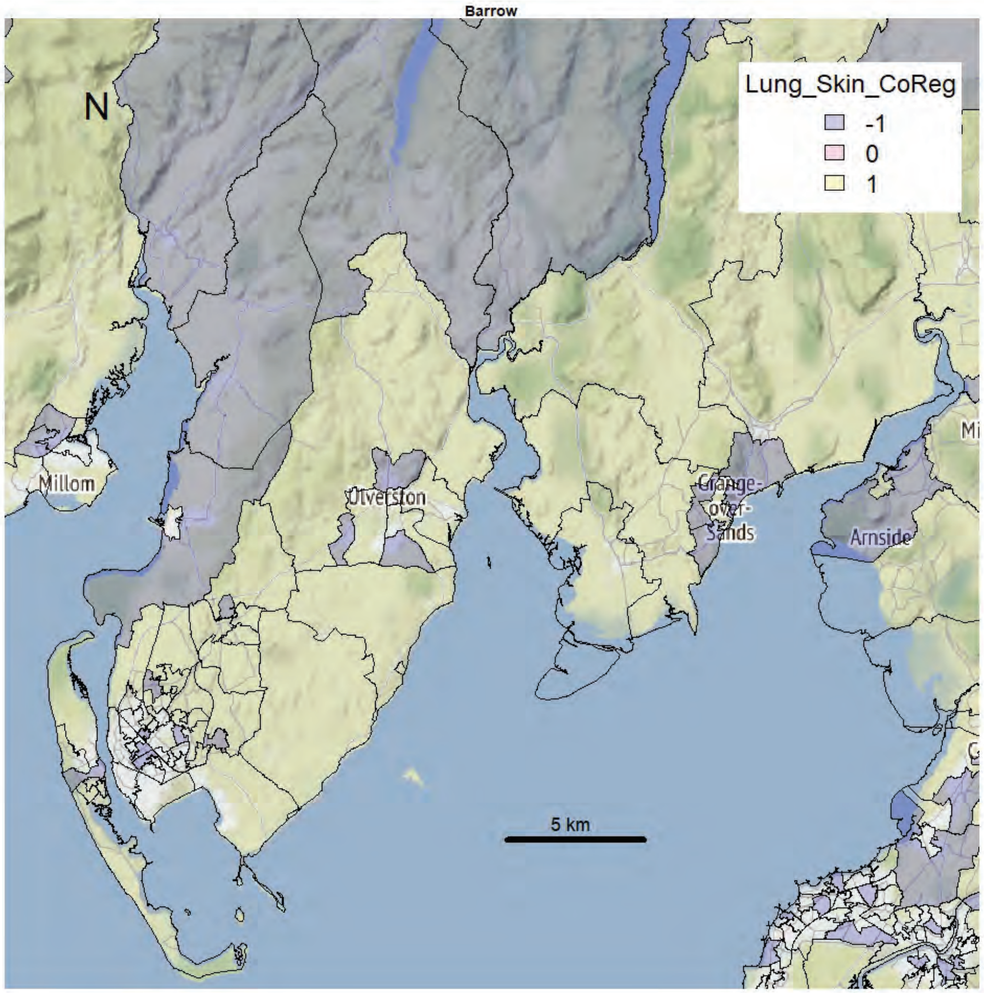
Barrow
N
Lung_Skin_CoReg
-1
0
1
Millom
Ulverston
Grange-over-Sands
Arnside
5 km

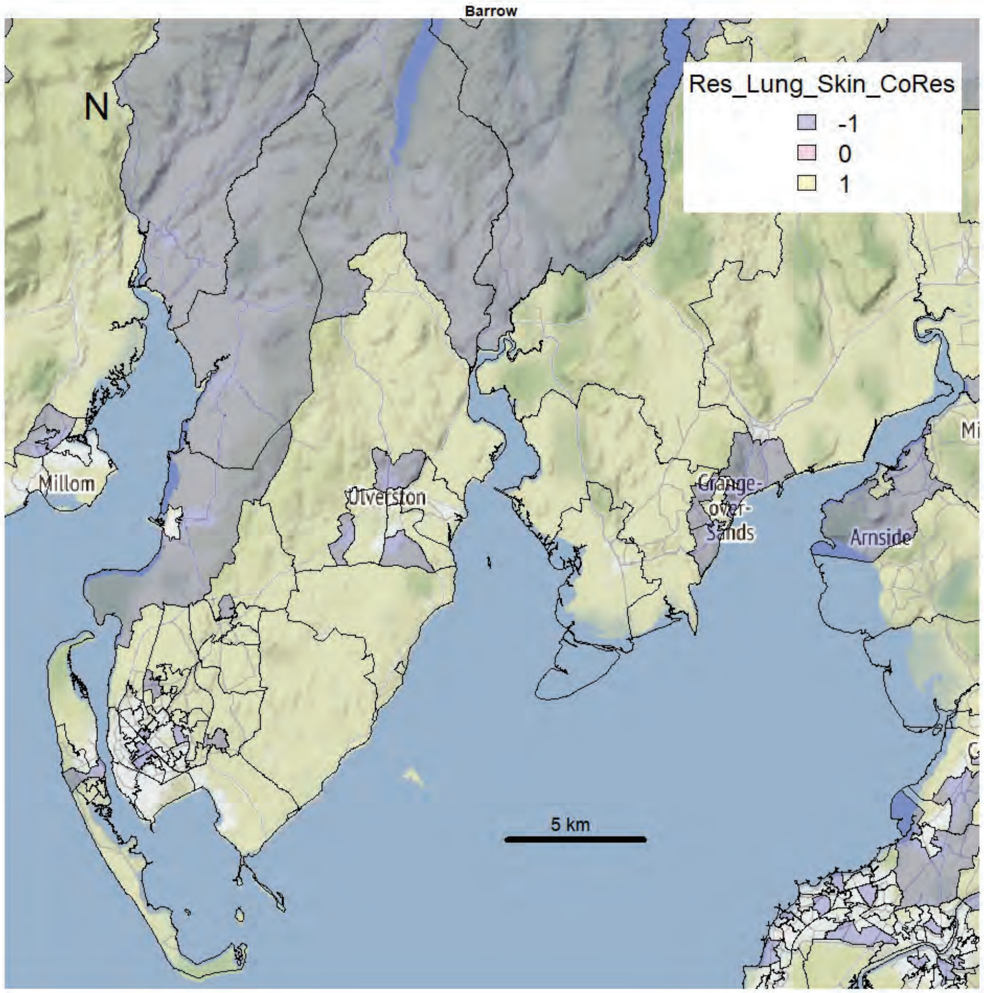
Barrow
N
Res_Lung_Skin_CoRes
-1
0
1
Millom
Ulverston
Grange-over-Sands
Arnside
5 km

Barrow

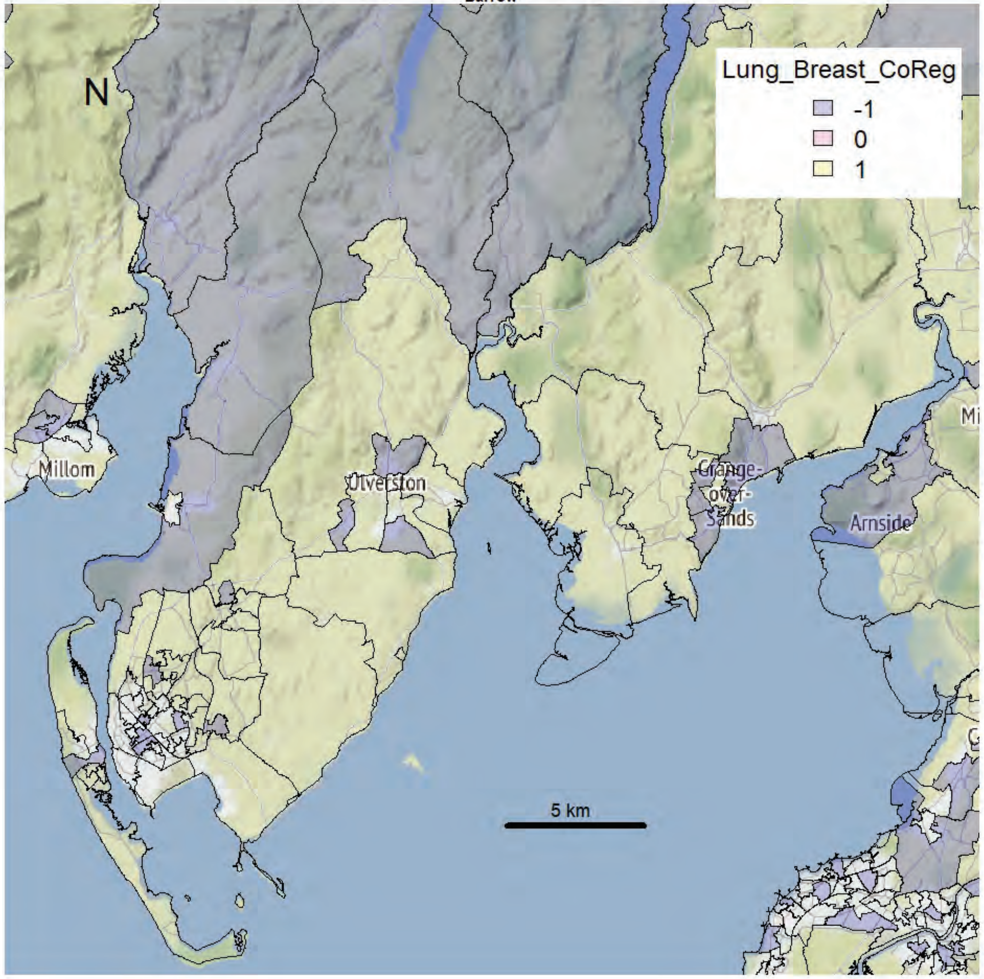


Barrow

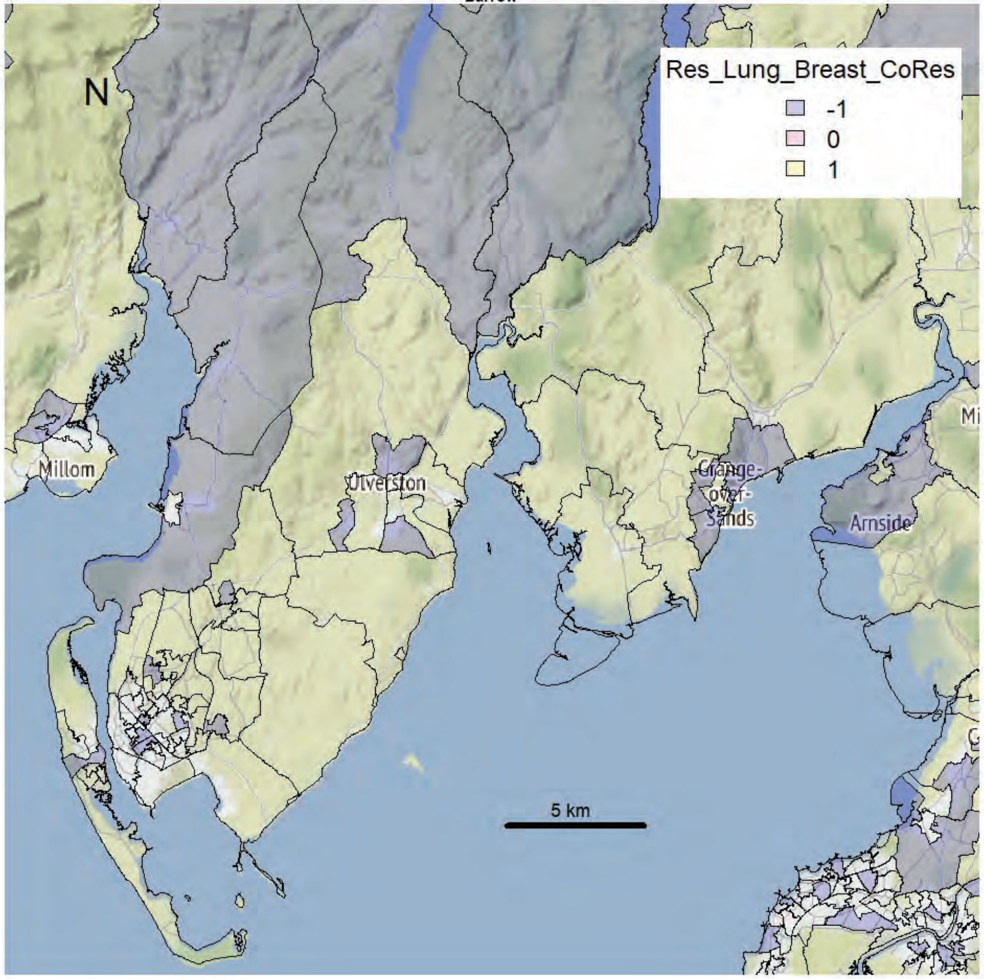

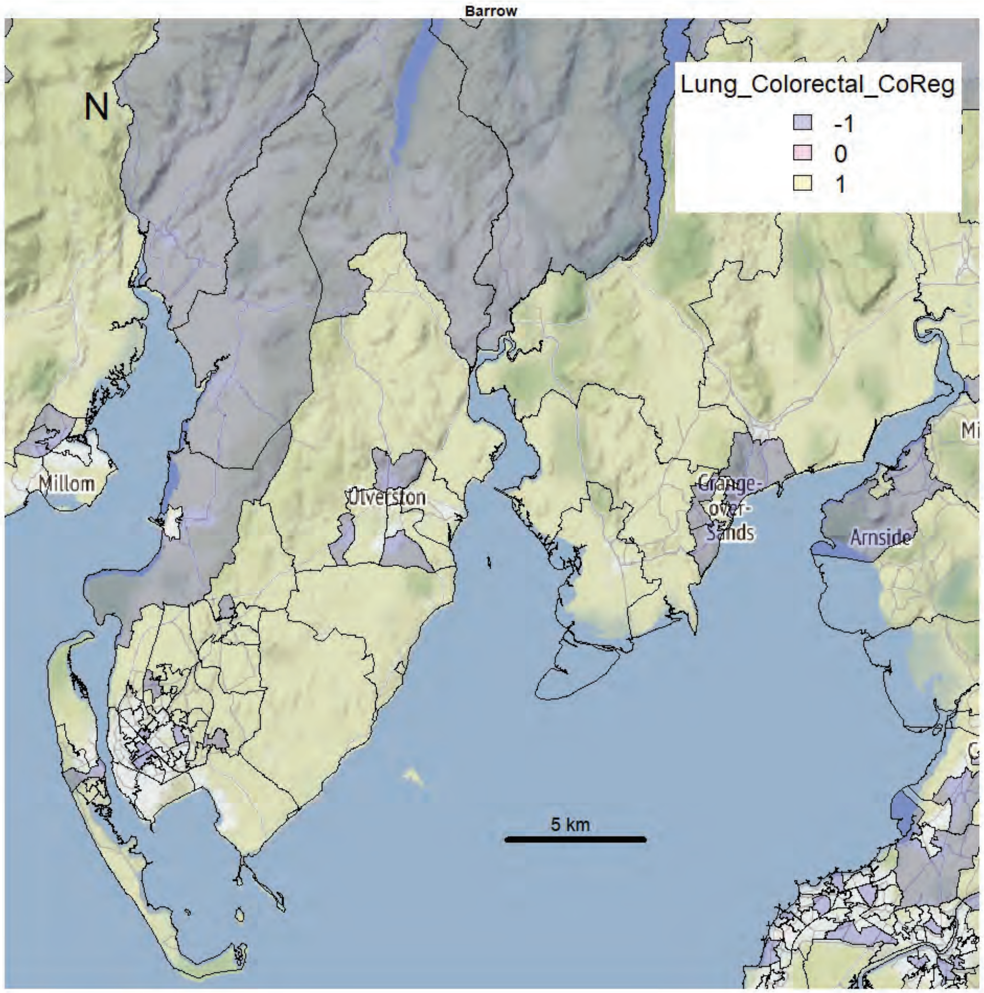
Barrow
N
Lung_Colorectal_CoReg
-1
0
1
Millom
Ulverston
Grange-over-Sands
Arnside
5 km

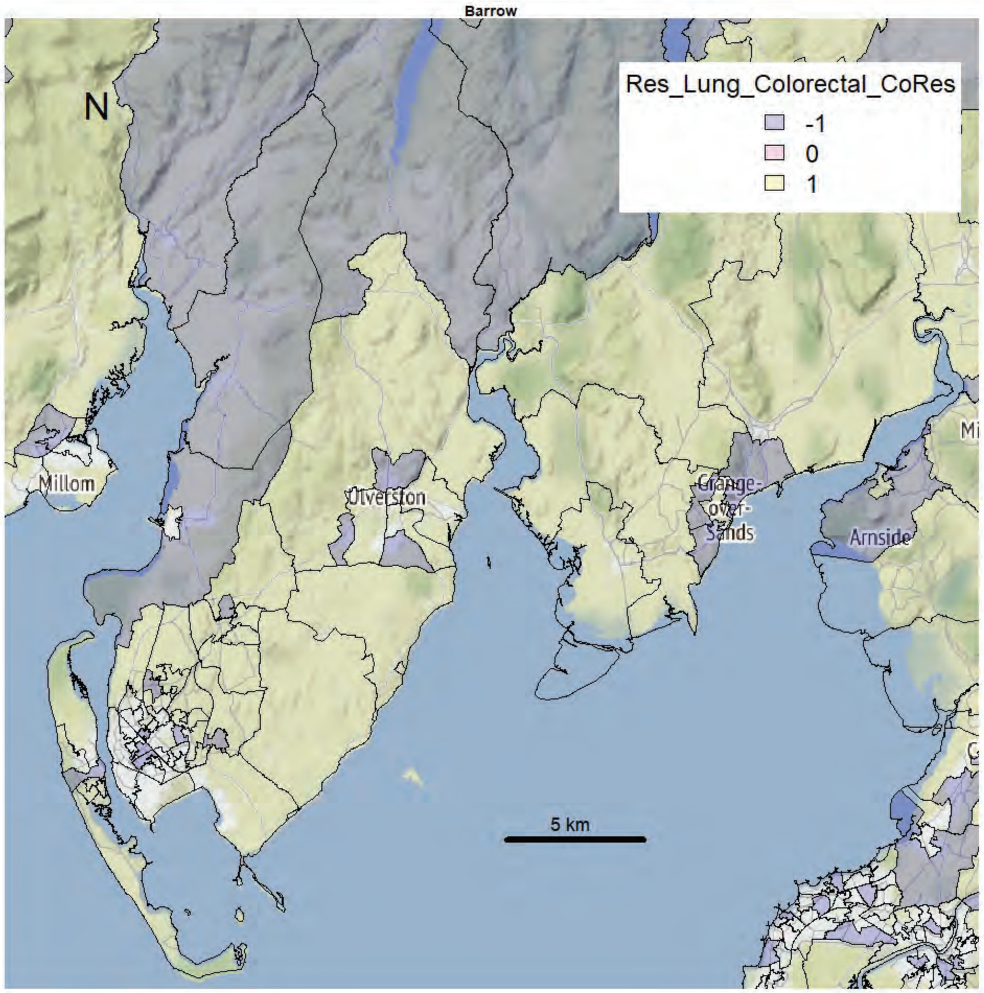
Barrow
N
Res_Lung_Colorectal_CoRes
-1
0
1
Millom
Ulverston
Grange-over-Sands
Arnside
5 km

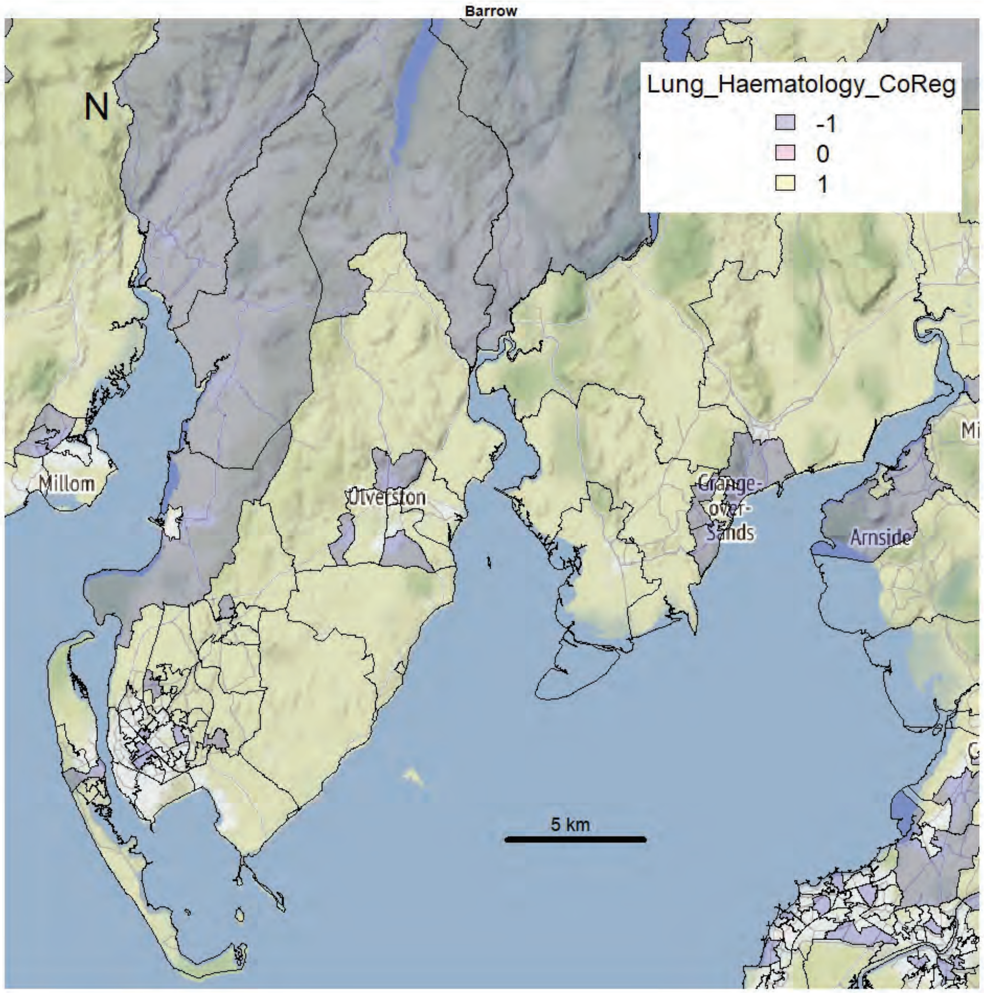
Barrow
N
Lung_Haematology_CoReg
-1
0
1
Millom
Ulverston
Grange-over-Sands
Arnside
5 km

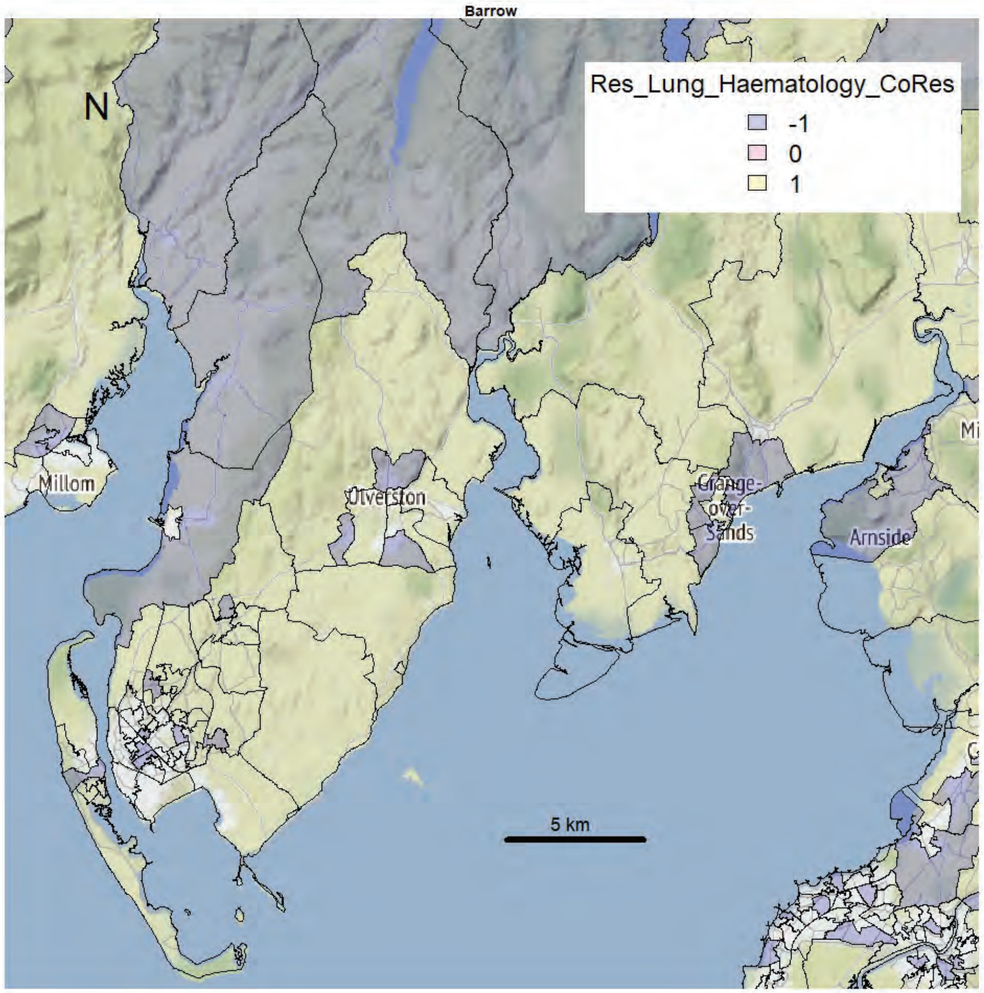
Barrow
N
Res_Lung_Haematology_CoRes
-1
0
1
Millom
Ulverston
Grange-over-Sands
Arnside
5 km

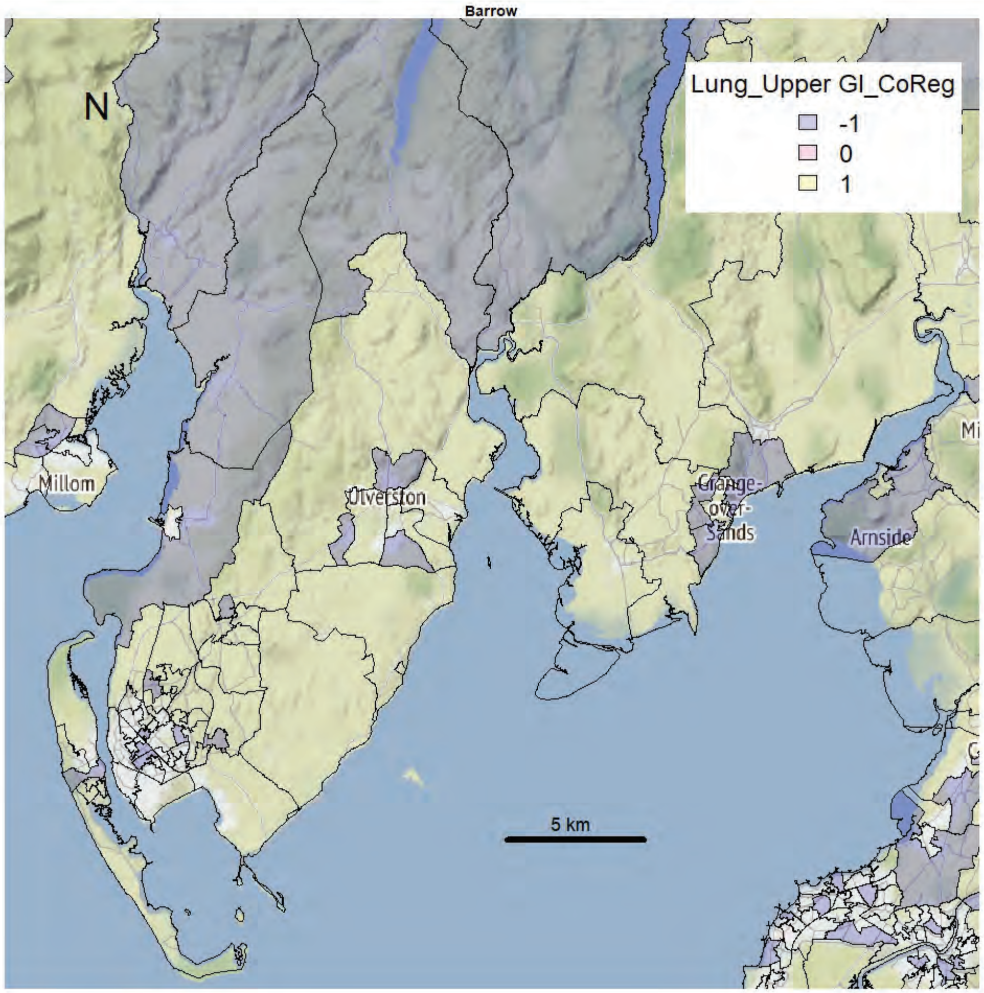
Barrow
N
Lung_Upper GI_CoReg
-1
0
1
Millom
Ulverston
Grange-over-Sands
Arnside
5 km

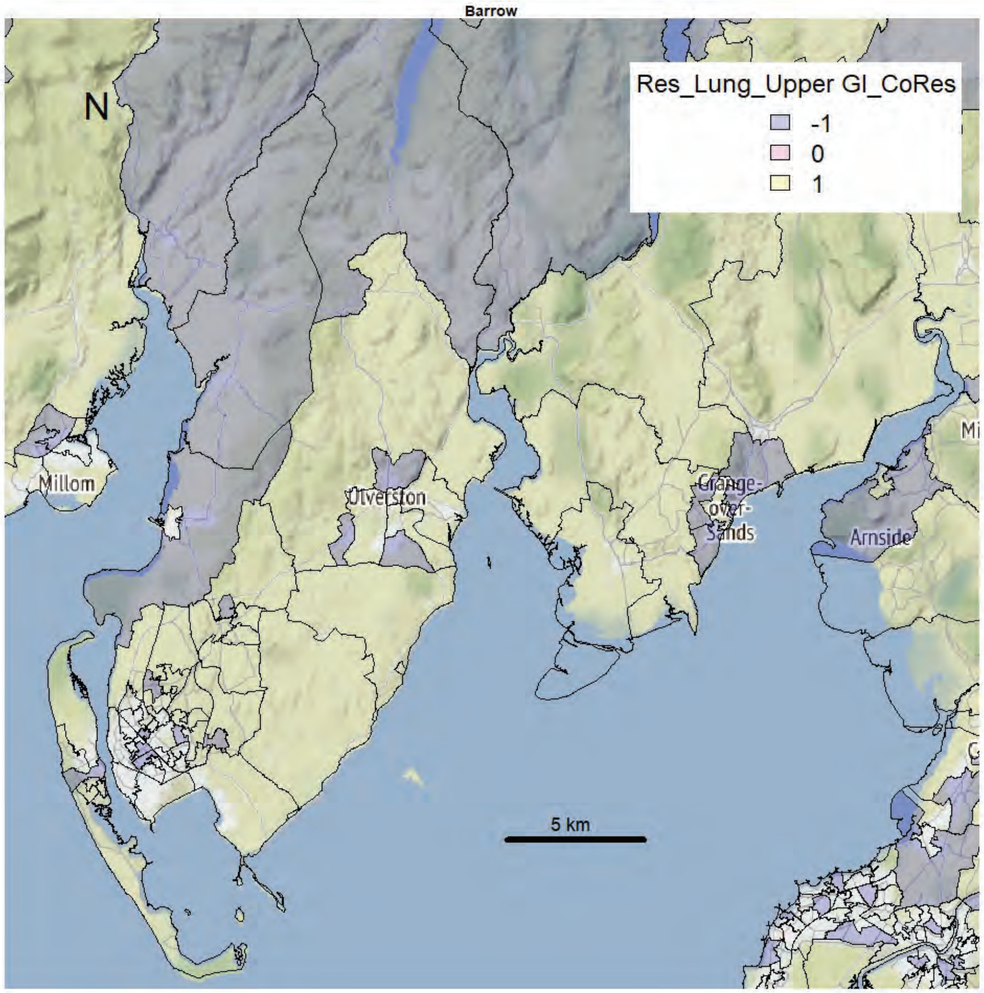
Barrow
N
Res_Lung_Upper GI_CoRes
-1
0
1
Millom
Ulverston
Grange-over-Sands
Arnside
5 km

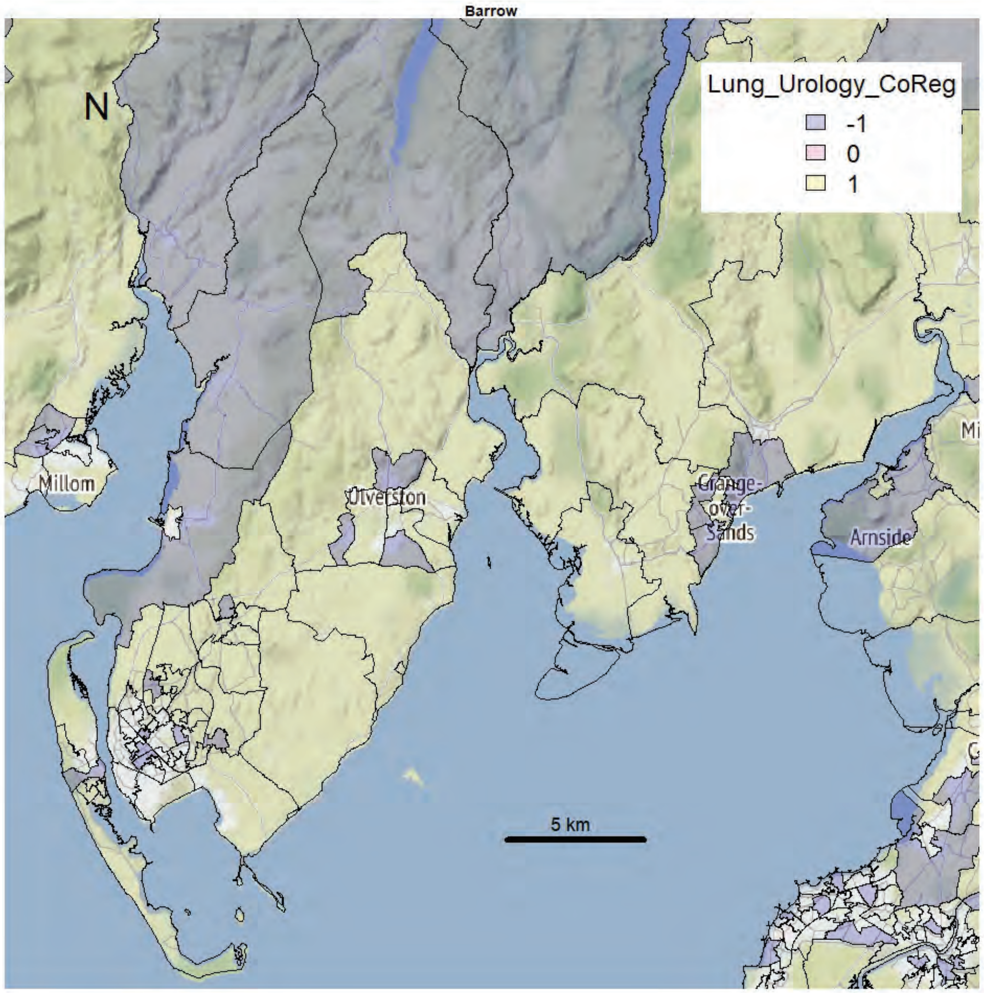
Barrow
N
Lung_Urology_CoReg
-1
0
1
Millom
Ulverston
Grange-over-Sands
Arnside
5 km

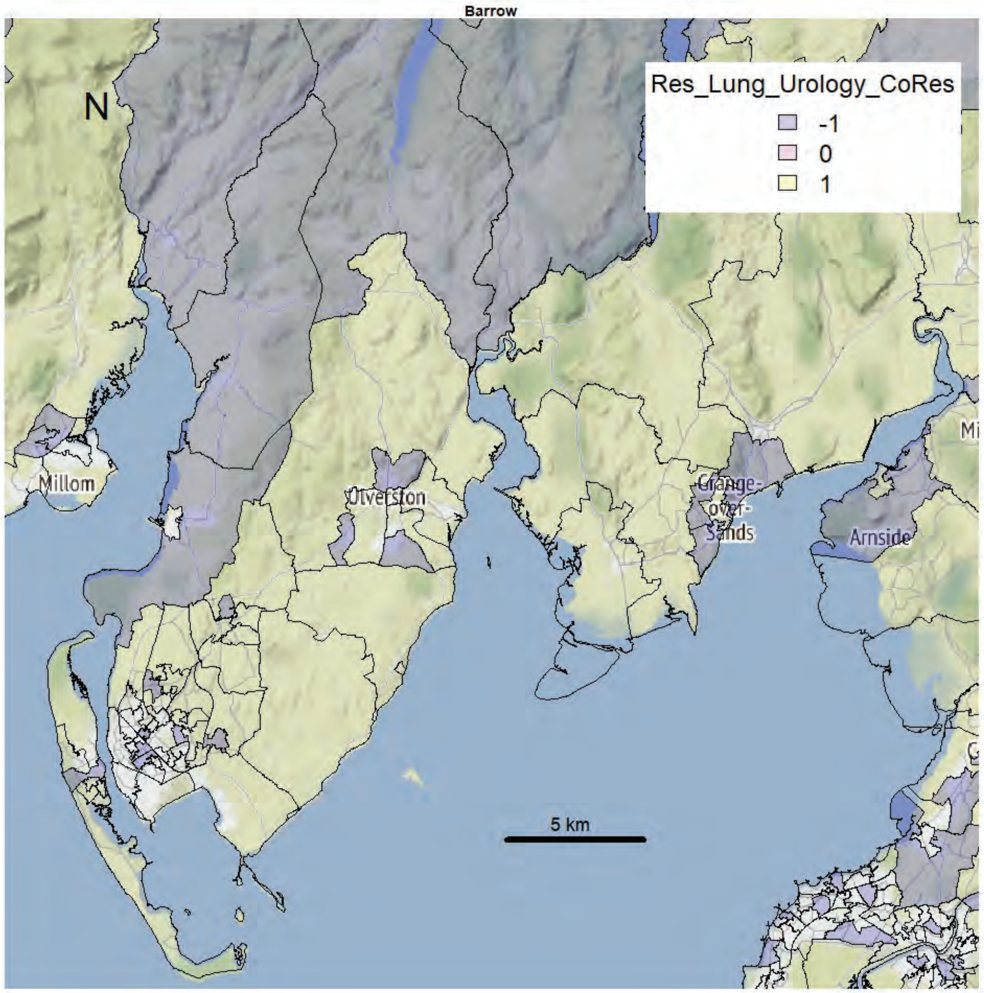
Barrow
N
Res_Lung_Urology_CoRes
-1
0
1
Millom
Ulverston
Grange-over-Sands
Arnside
5 km

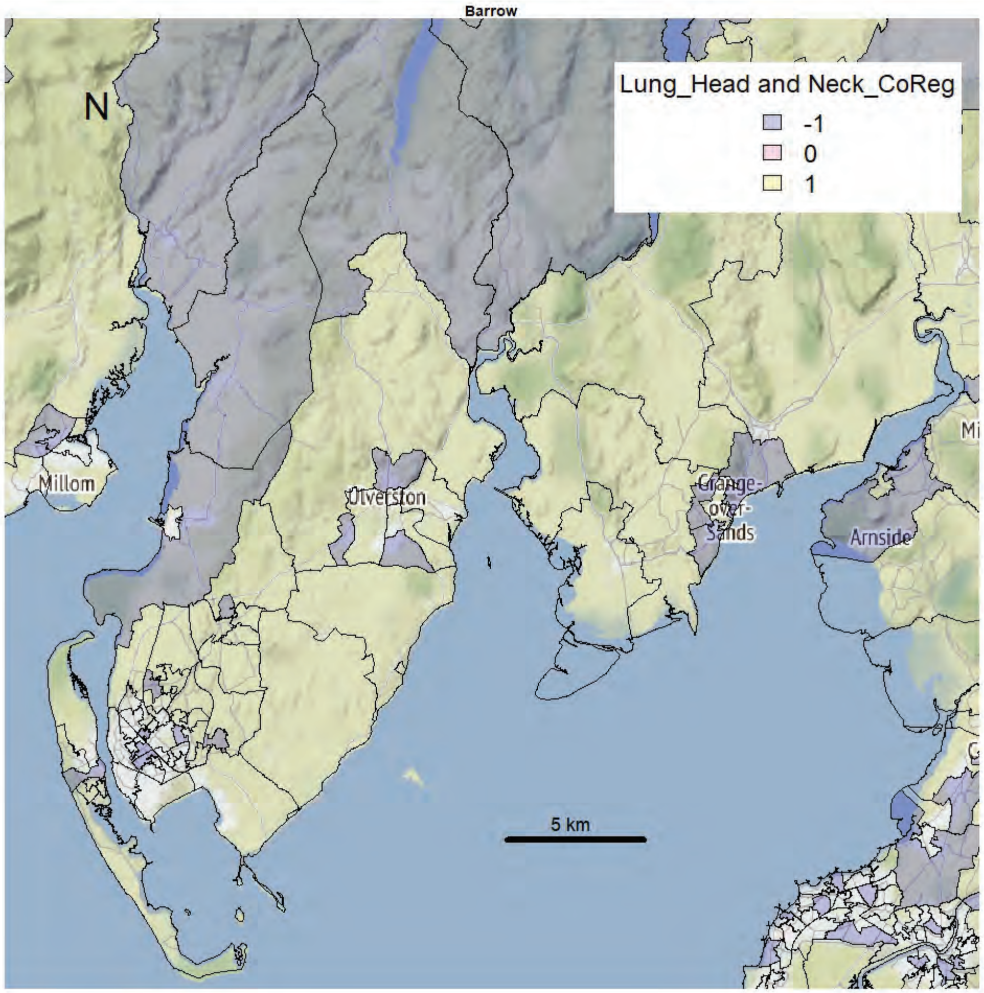

Barrow
N
Lung_Head and Neck_CoReg
-1
0
1
Millom
Ulverston
Grange-over-Sands
Arnside
5 km


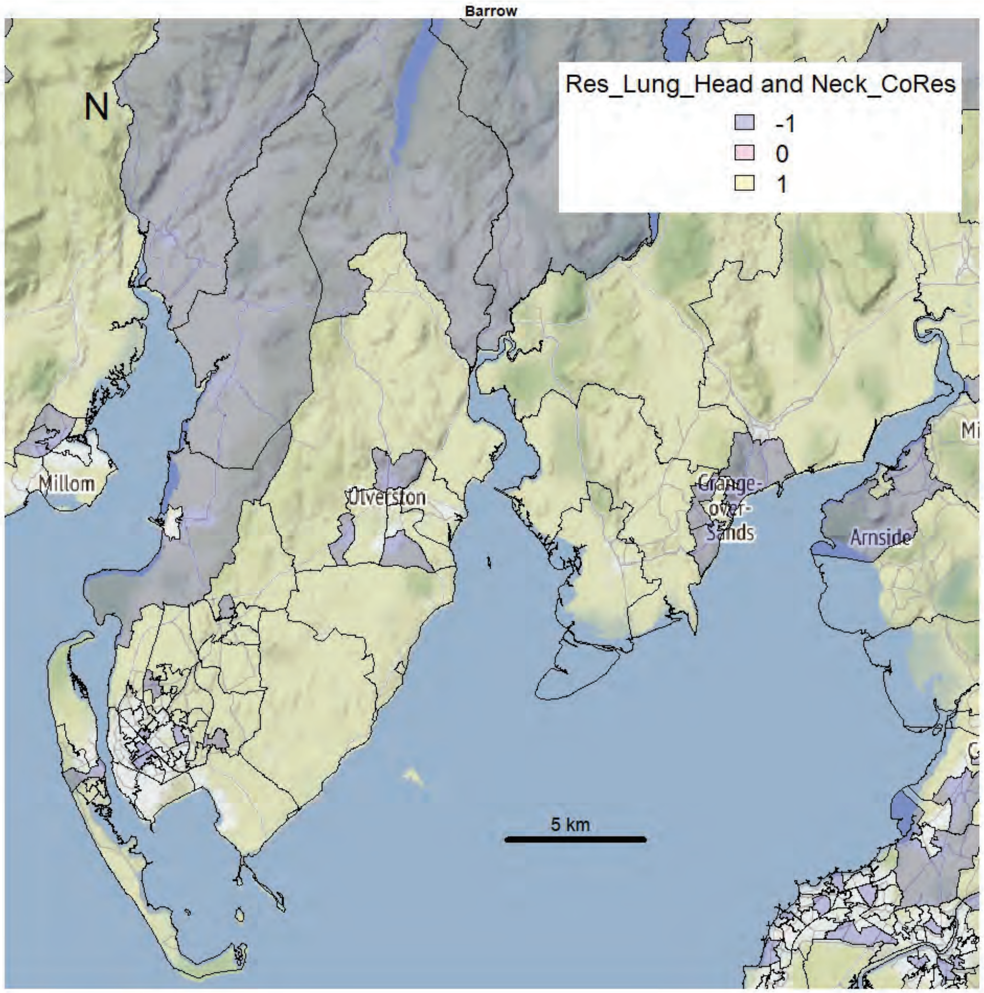

Barrow
N
Res_Lung_Head and Neck_CoRes
-1
0
1
Millom
Ulverston
Grange-over-Sands
Arnside
5 km

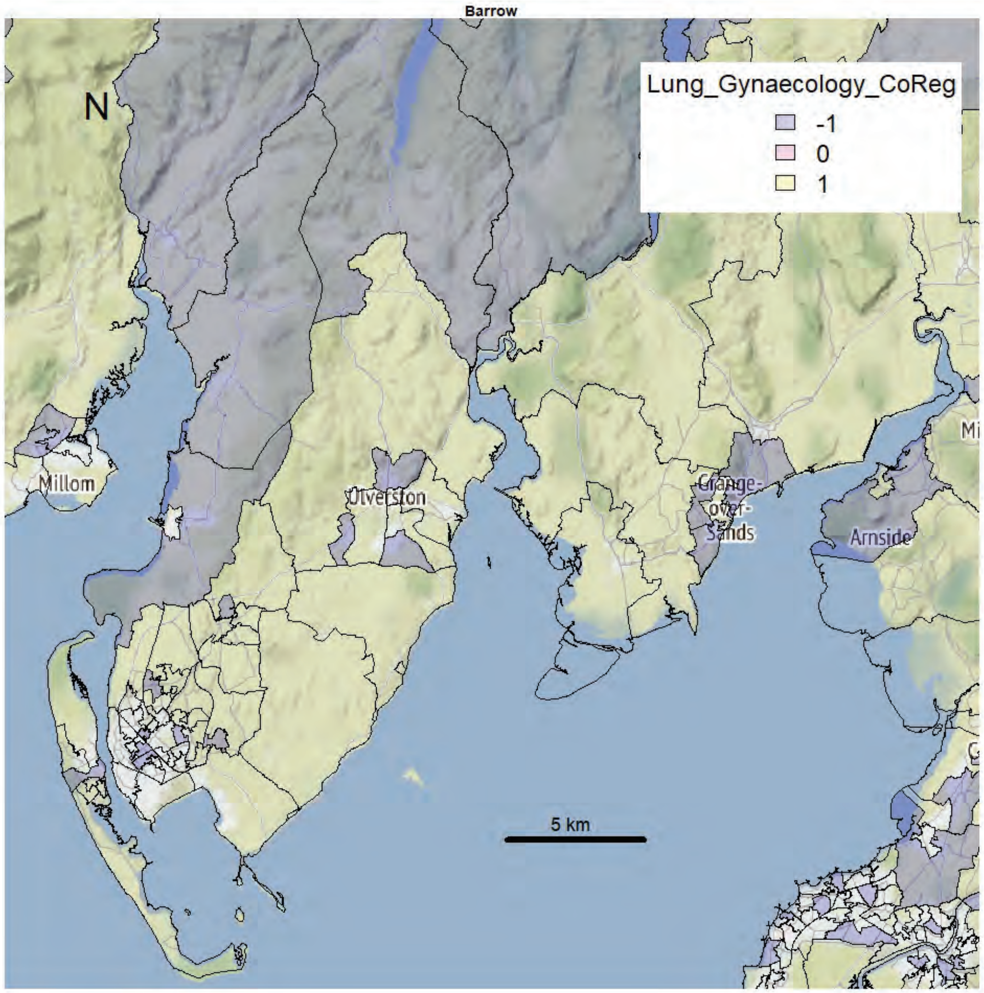
Barrow
N
Lung_Gynaecology_CoReg
-1
0
1
Millom
Ulverston
Grange-over-Sands
Arnside
5 km

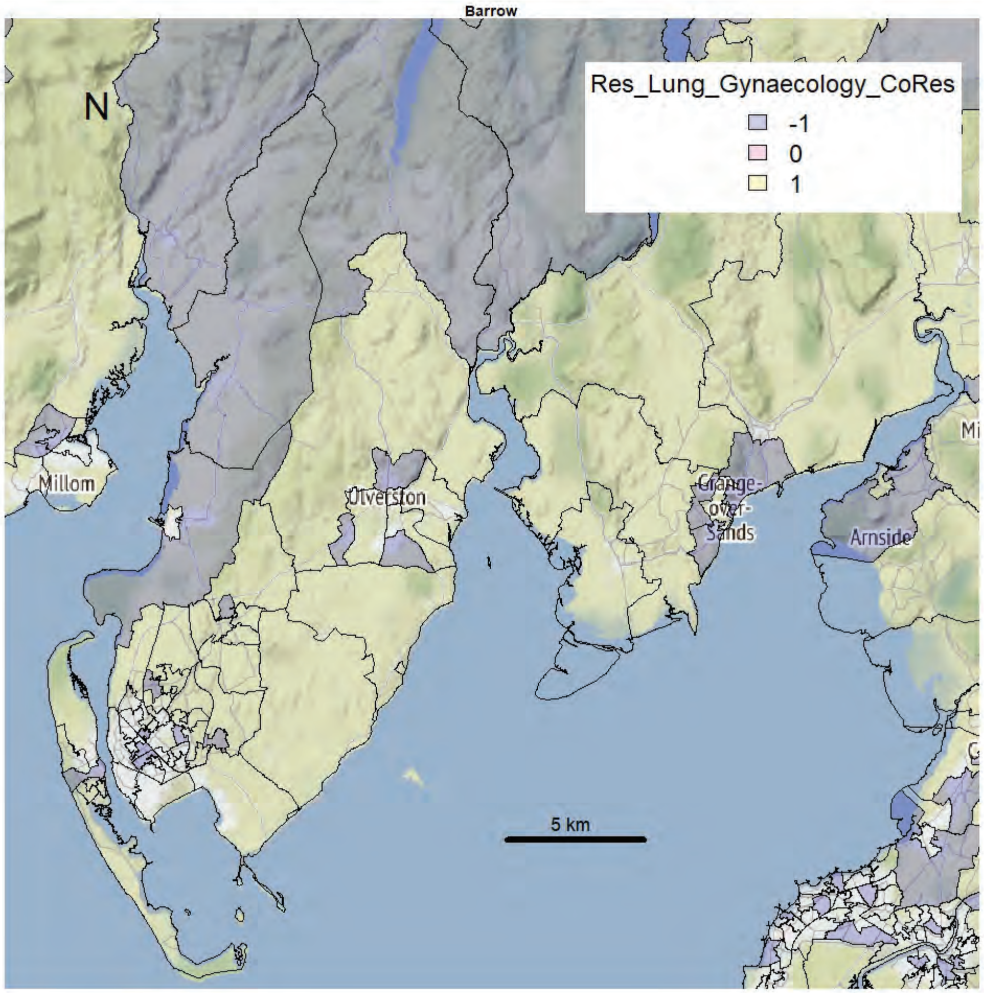
Barrow
N
Res_Lung_Gynaecology_CoRes
-1
0
1
Millom
Ulverston
Grange-over-Sands
Arnside
5 km

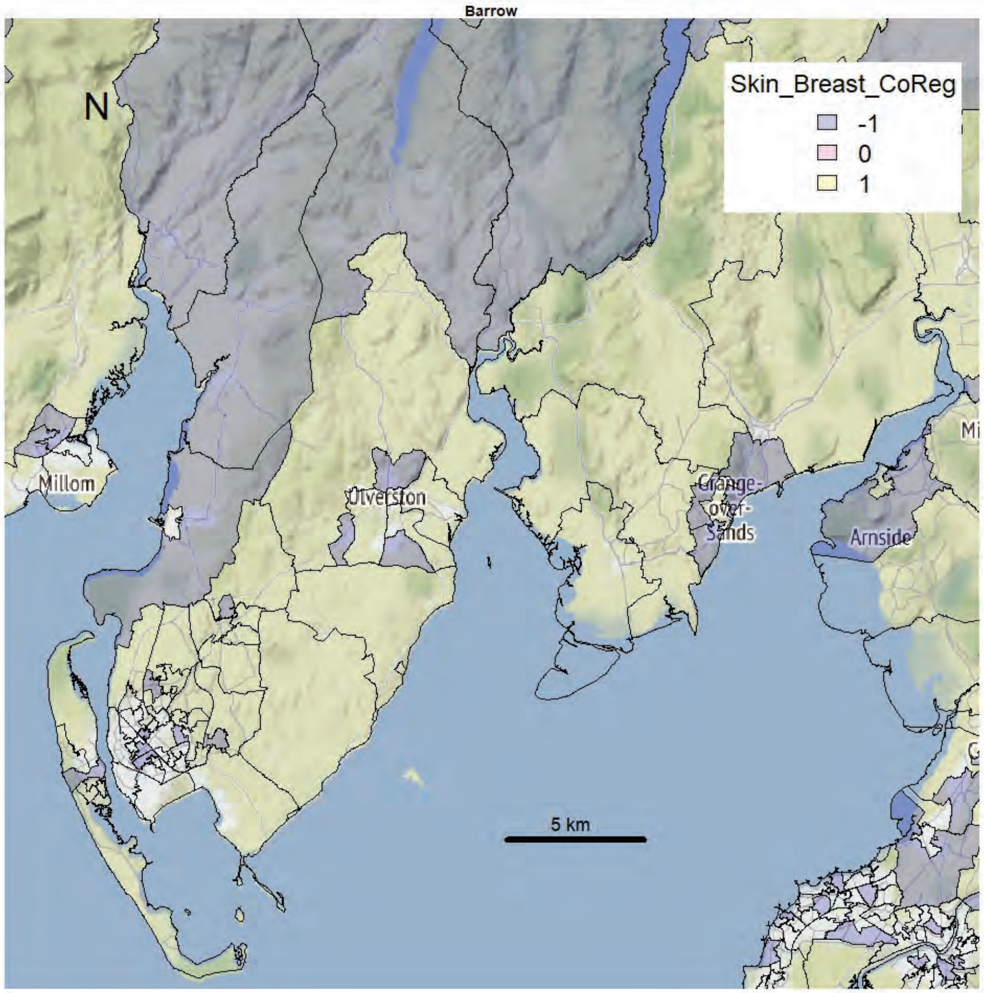
Barrow
N
Skin_Breast_CoReg
-1
0
1
Millom
Ulverston
Grange-over-Sands
Arnside
5 km

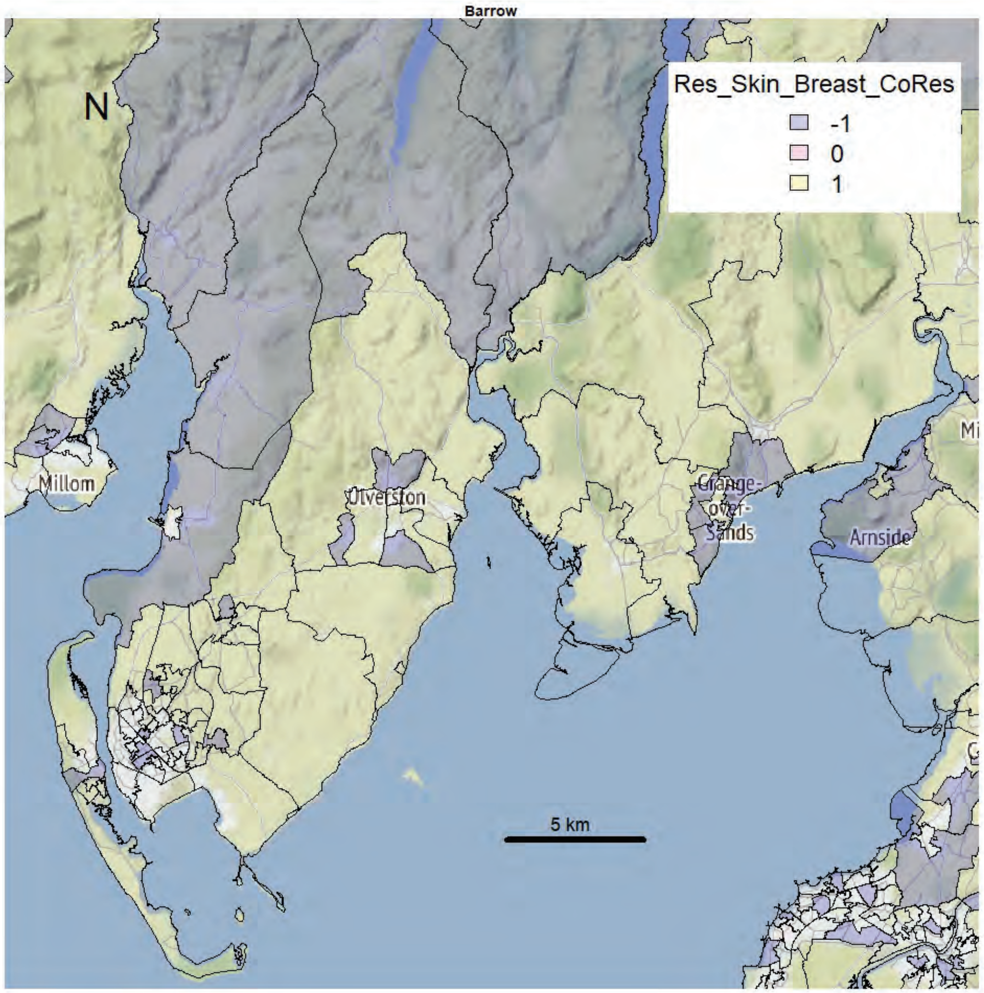
Barrow
N
Res_Skin_Breast_CoRes
-1
0
1
Millom
Ulverston
Grange-over-Sands
Arnside
5 km

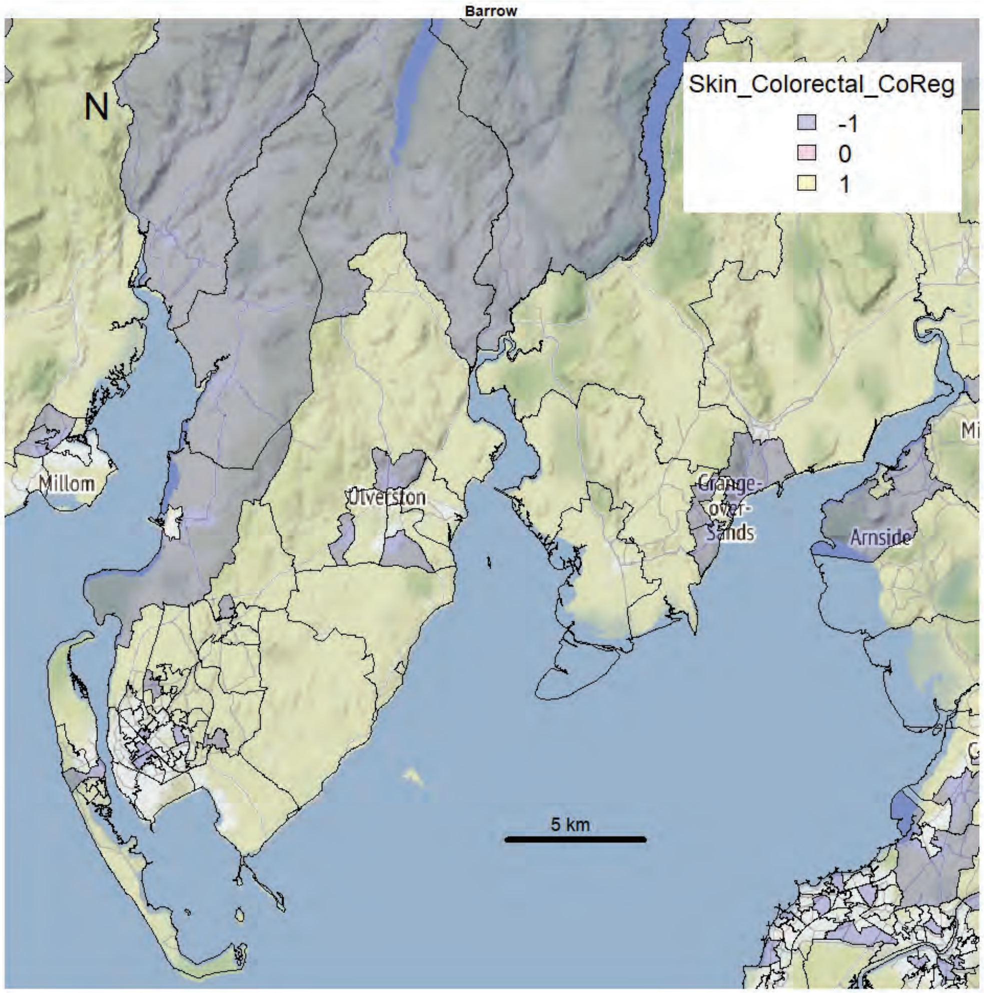
Barrow
N
Skin_Colorectal_CoReg
-1
0
1
Millom
Ulverston
Grange-over-Sands
Arnside
5 km

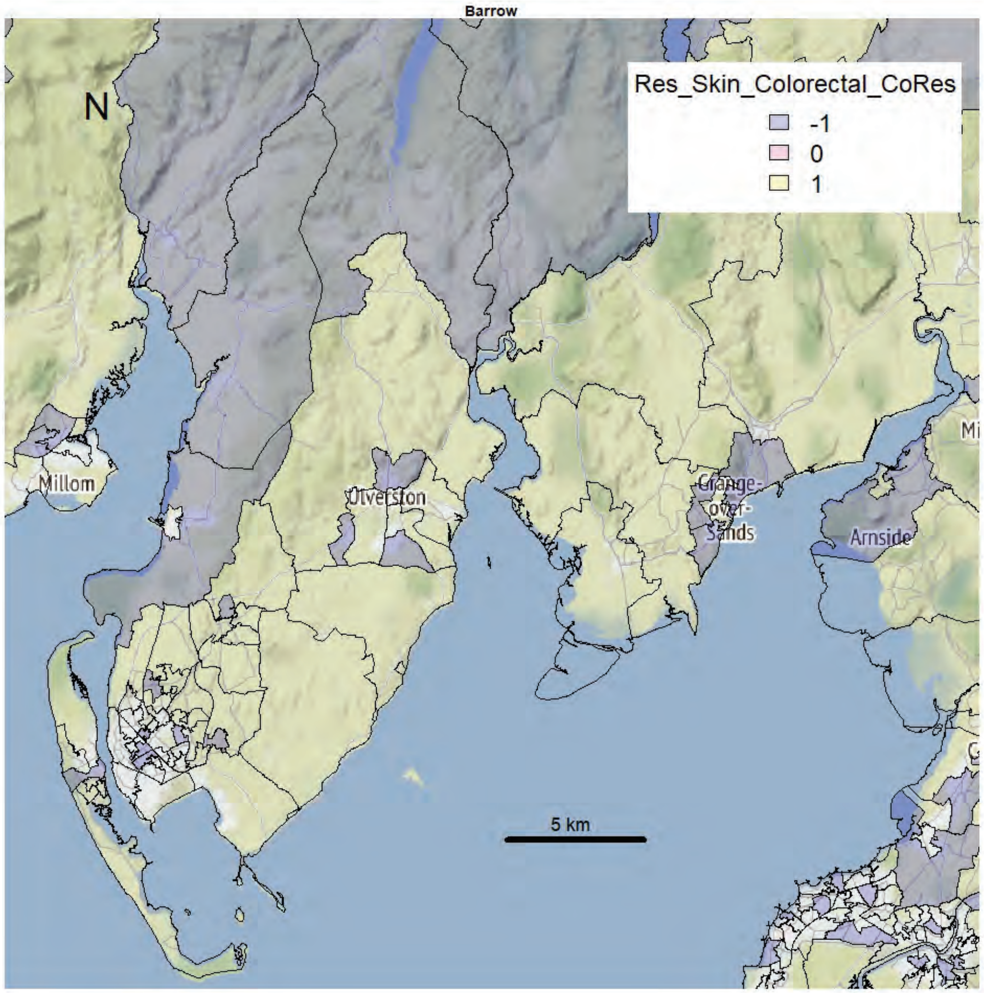
Barrow
N
Res_Skin_Colorectal_CoRes
-1
0
1
Millom
Ulverston
Grange-over-Sands
Arnside
5 km

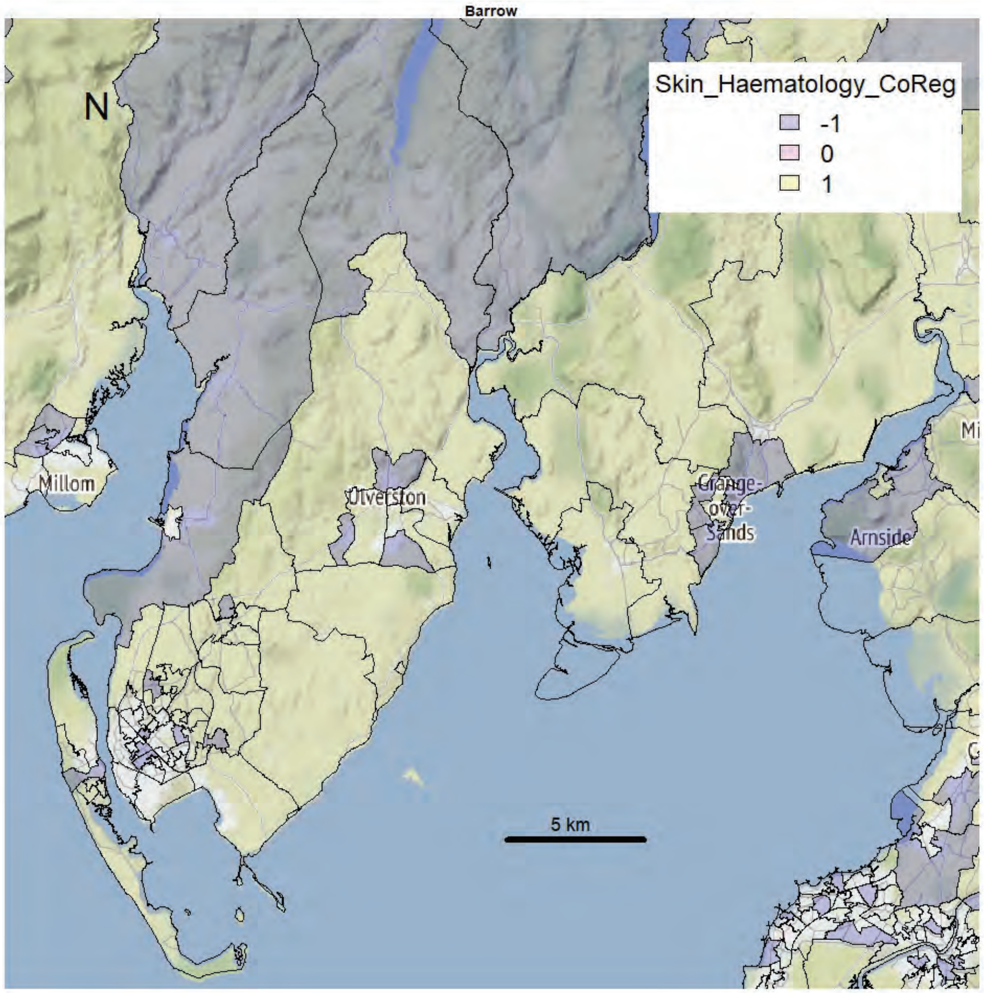
Barrow
N
Skin_Haematology_CoReg
-1
0
1
Millom
Ulverston
Grange-over-Sands
Arnside
5 km

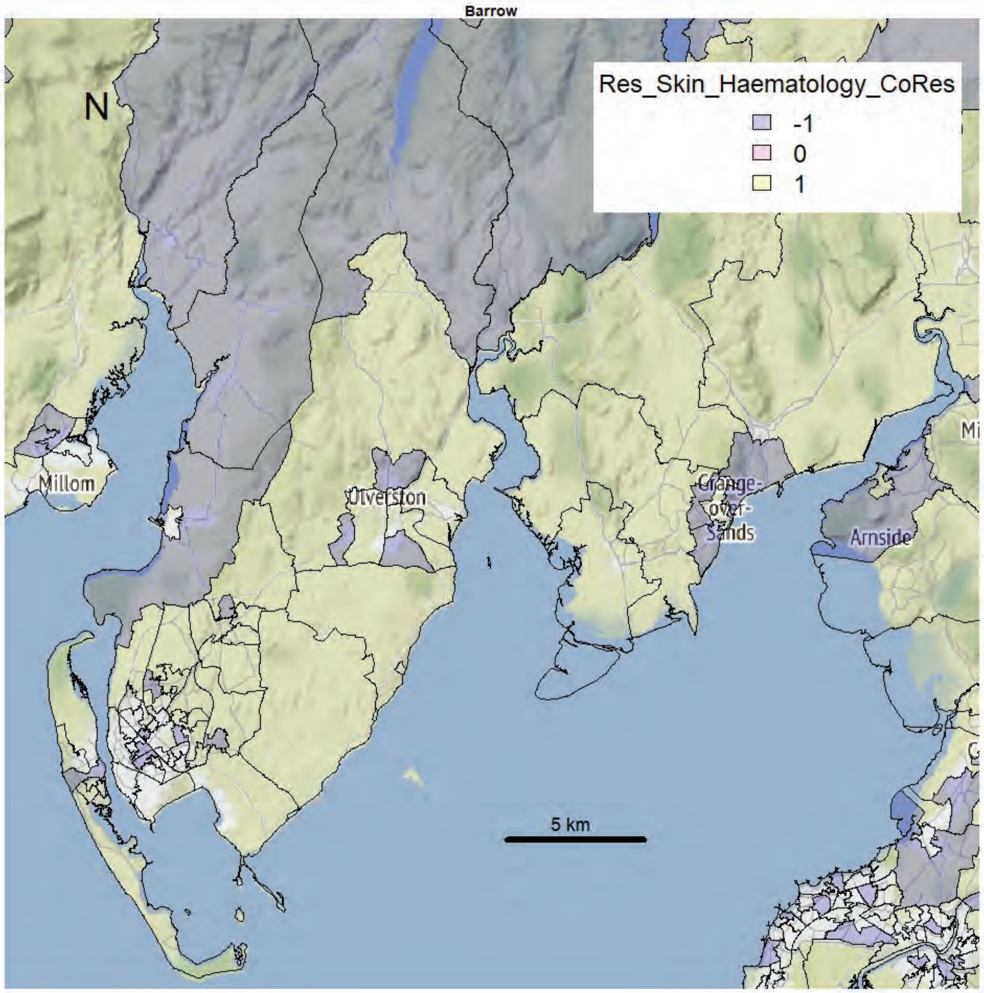
Barrow
N
Res_Skin_Haematology_CoRes
-1
0
1
Millom
Ulverston
Grange-over-Sands
Arnside
5 km

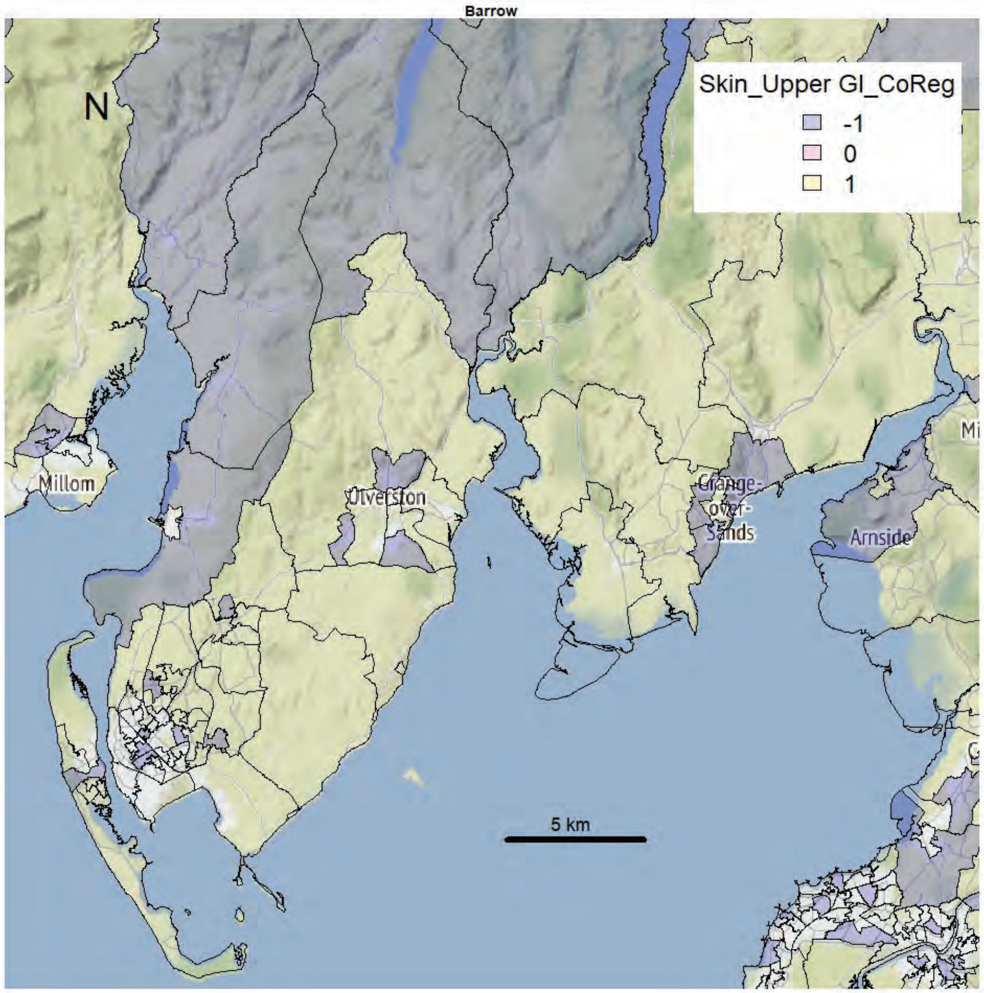
Barrow
N
Skin_Upper GI_CoReg
-1
0
1
Millom
Ulverston
Grange-over-Sands
Arnside
5 km

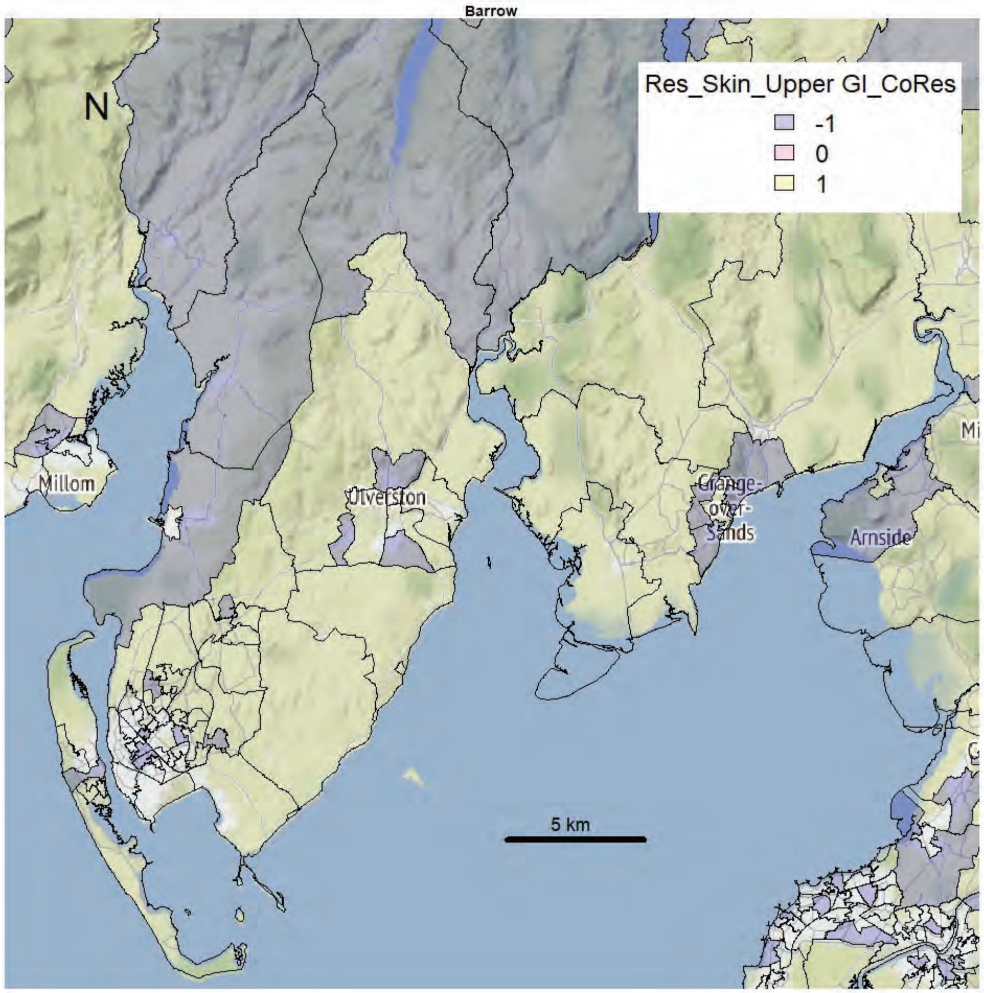
Barrow
N
Res_Skin_Upper GI_CoRes
-1
0
1
Millom
Ulverston
Grange-over-Sands
Arnside
5 km

Barrow

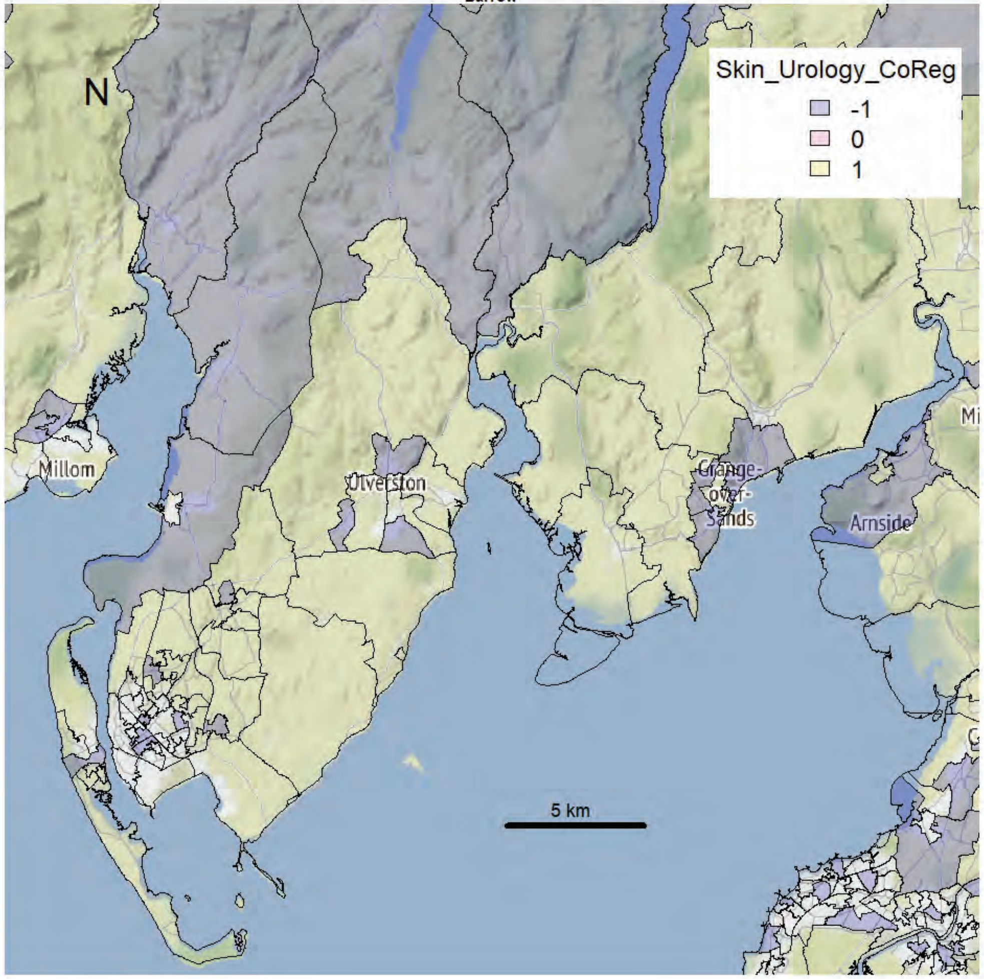


Barrow

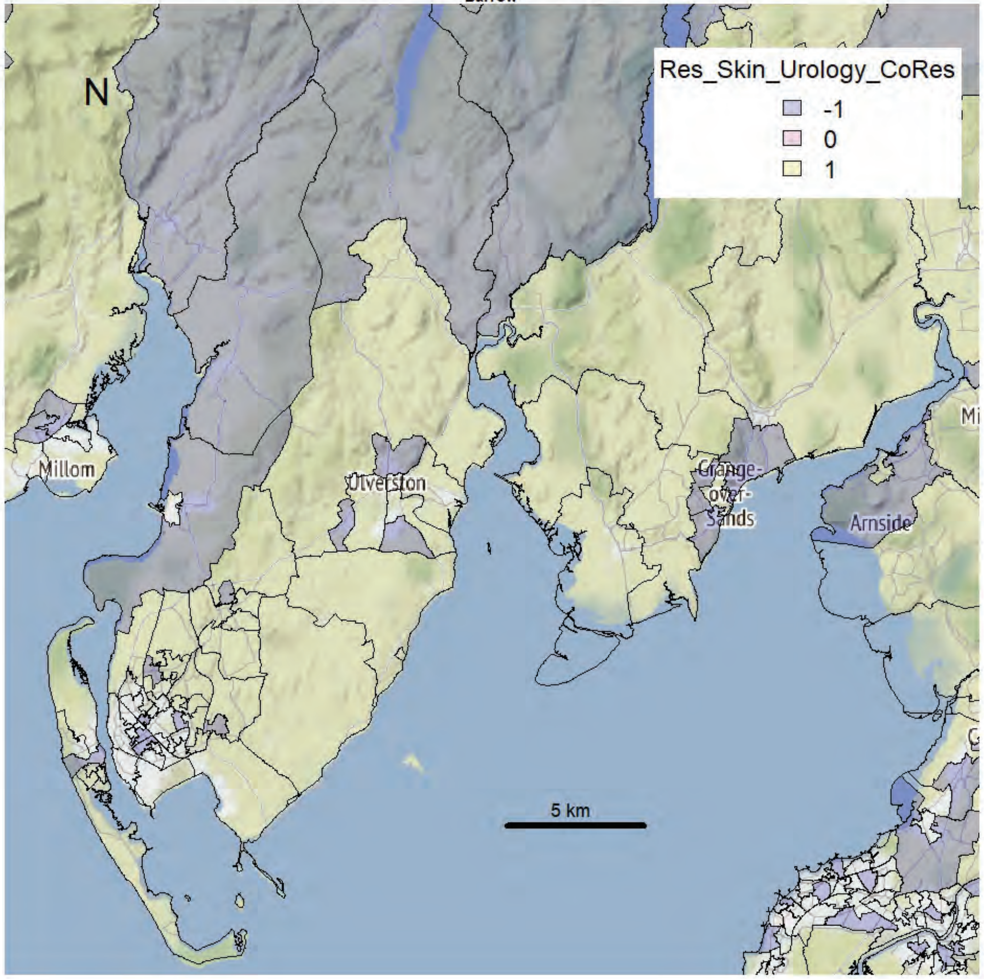

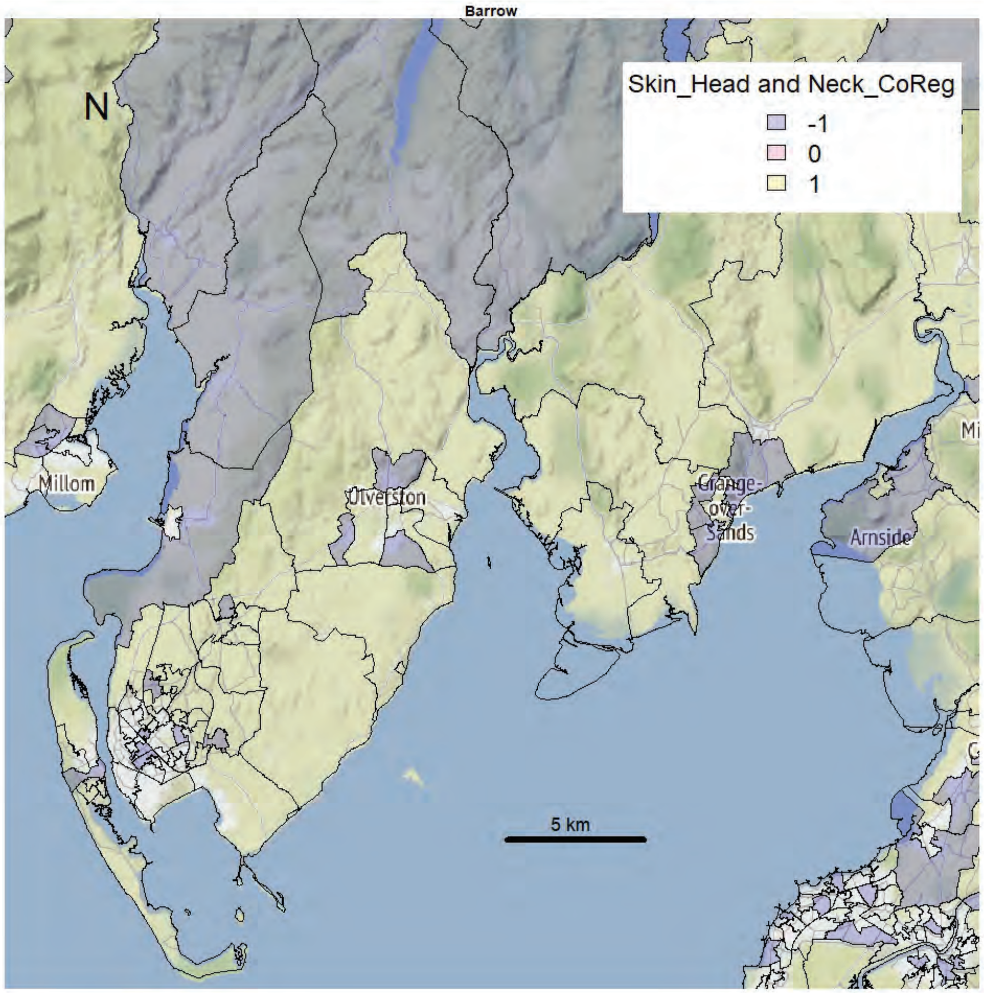
Barrow
N
Skin_Head and Neck_CoReg
-1
0
1
Millom
Ulverston
Grange-over-Sands
Arnside
5 km

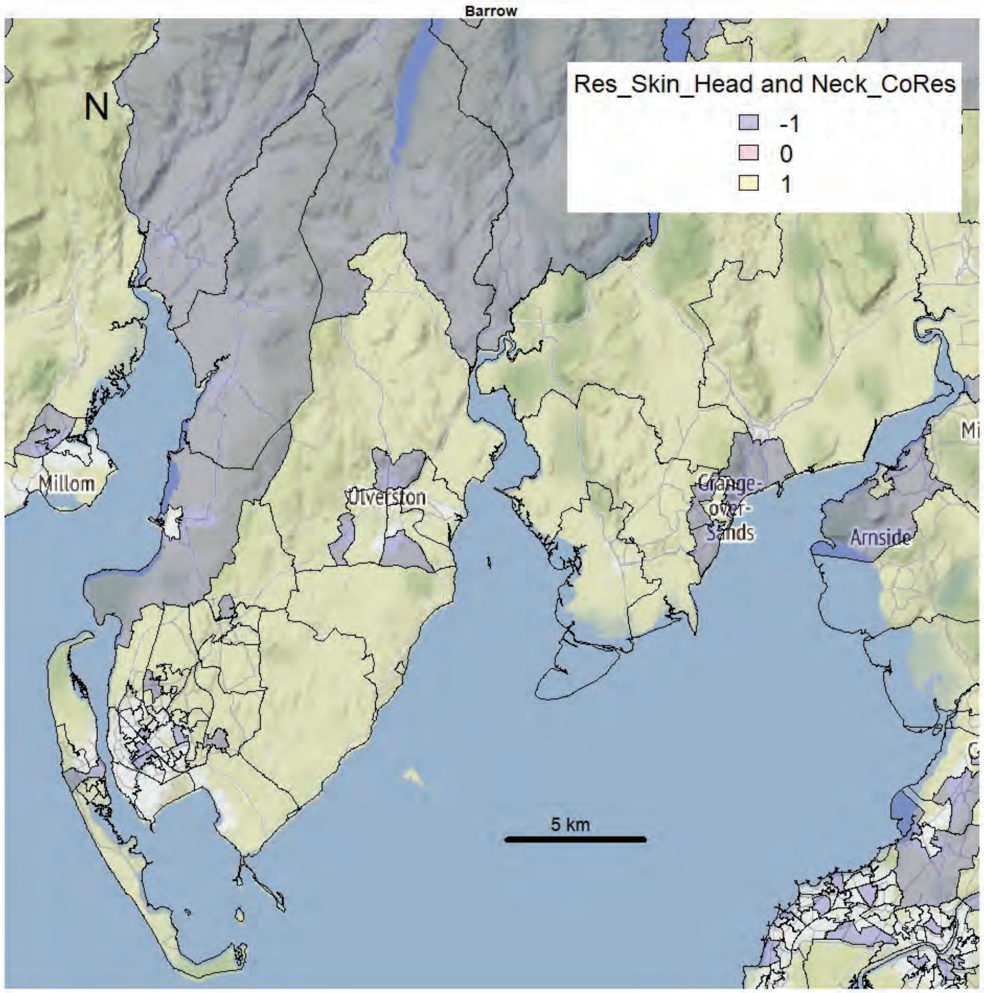
Barrow
N
Res_Skin_Head and Neck_CoRes
-1
0
1
Millom
Ulverston
Grange-over-Sands
Arnside
5 km

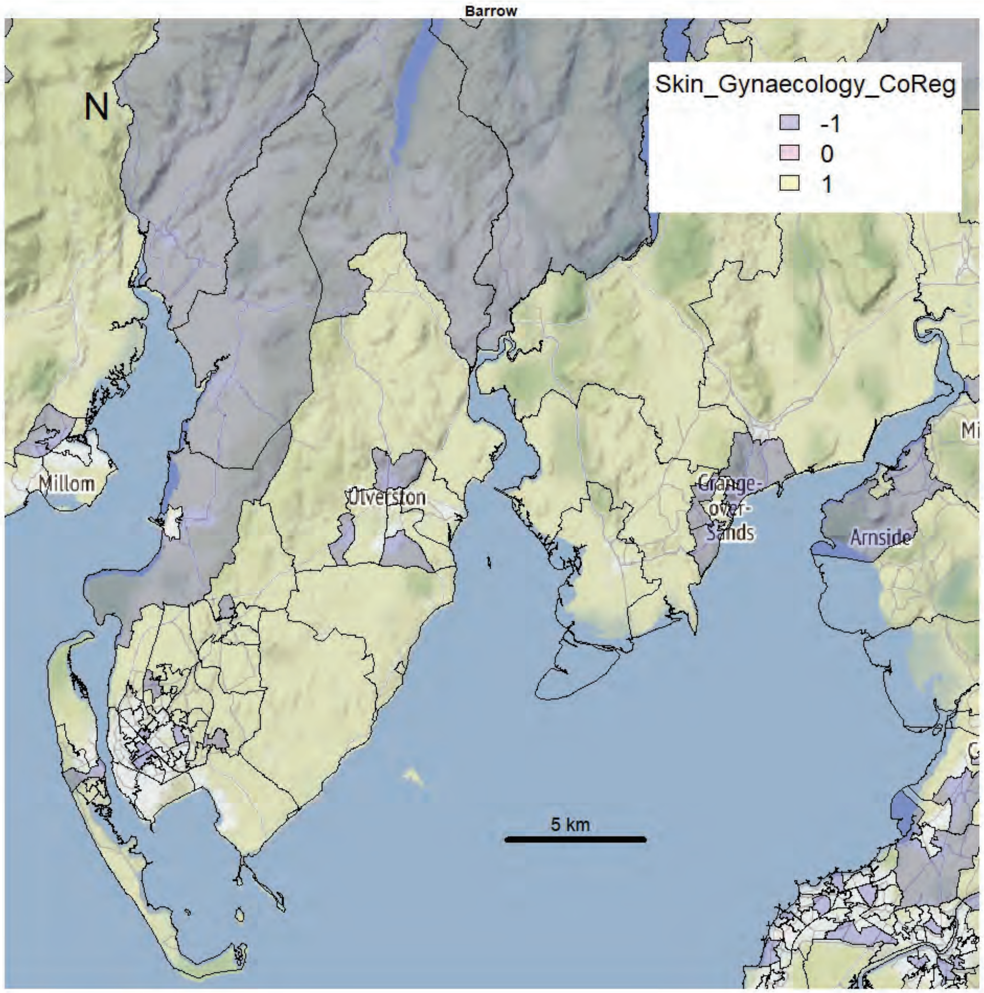
Barrow
N
Skin_Gynaecology_CoReg
-1
0
1
Millom
Ulverston
Grange-over-Sands
Arnside
5 km

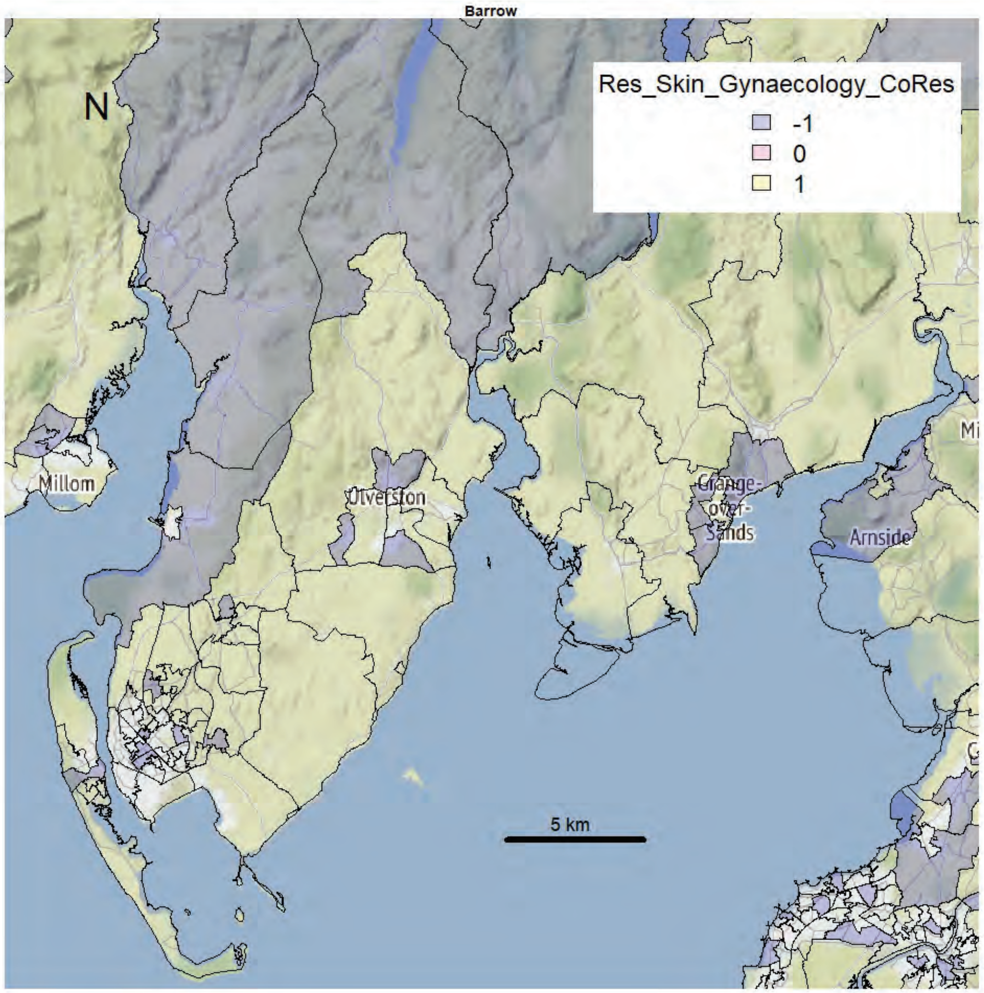
Barrow
N
Res_Skin_Gynaecology_CoRes
-1
0
1
Millom
Ulverston
Grange-over-Sands
Arnside
5 km

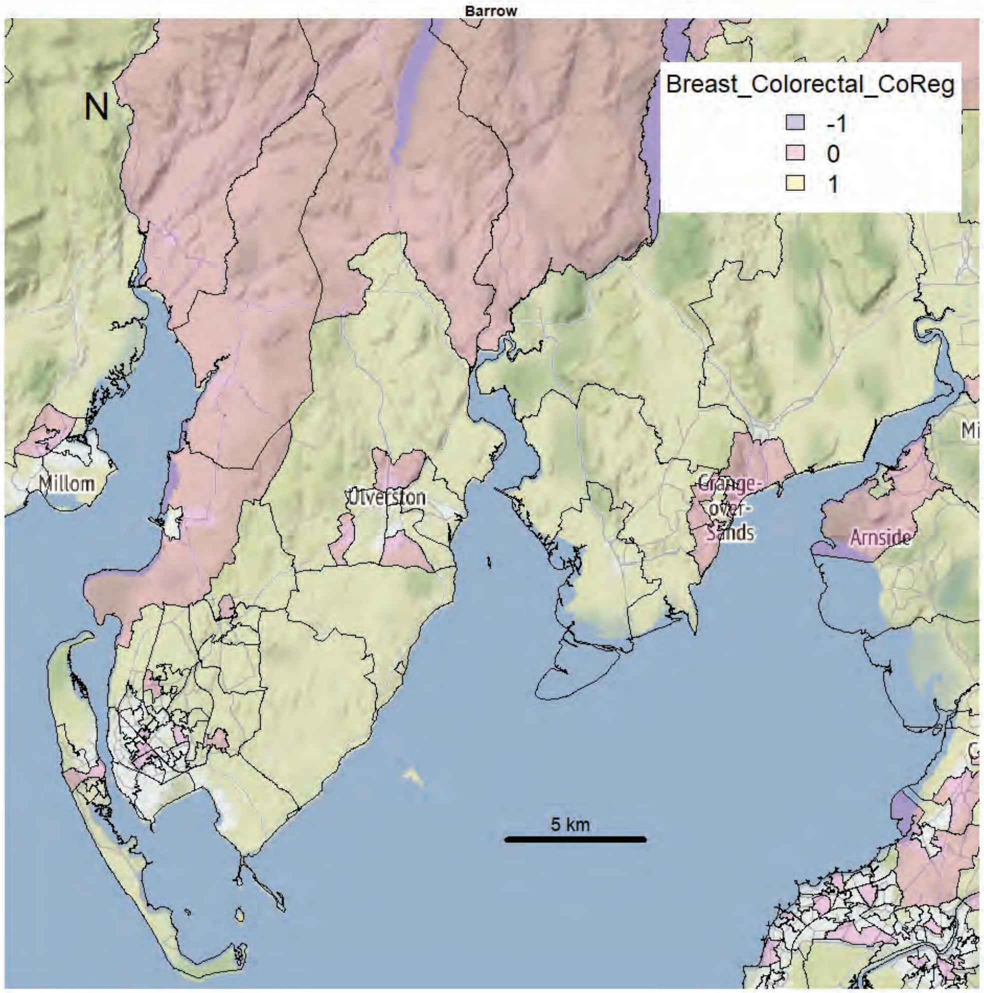
Barrow
N
Breast_Colorectal_CoReg
-1
0
1
Millom
Ulverston
Grange-over-Sands
Arnside
5 km

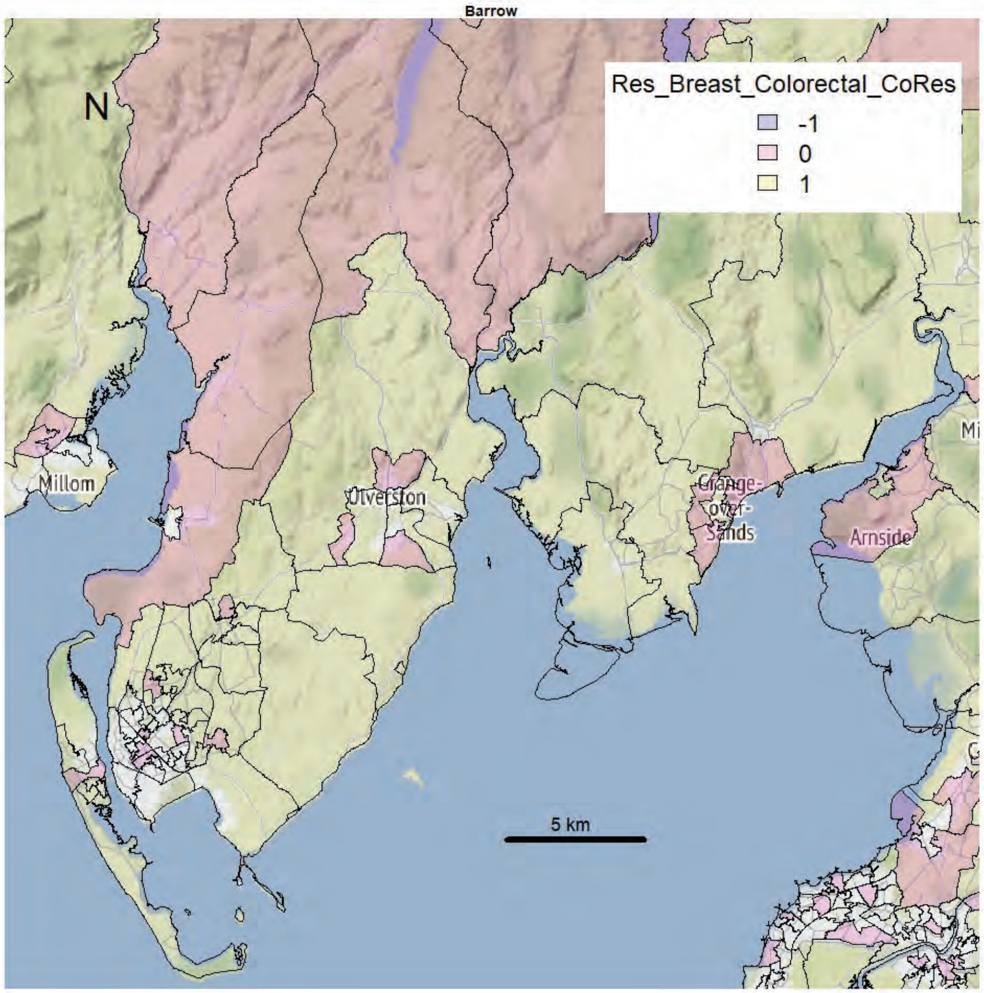
Barrow
N
Res_Breast_Colorectal_CoRes
-1
0
1
Millom
Ulverston
Grange-over-Sands
Arnside
5 km

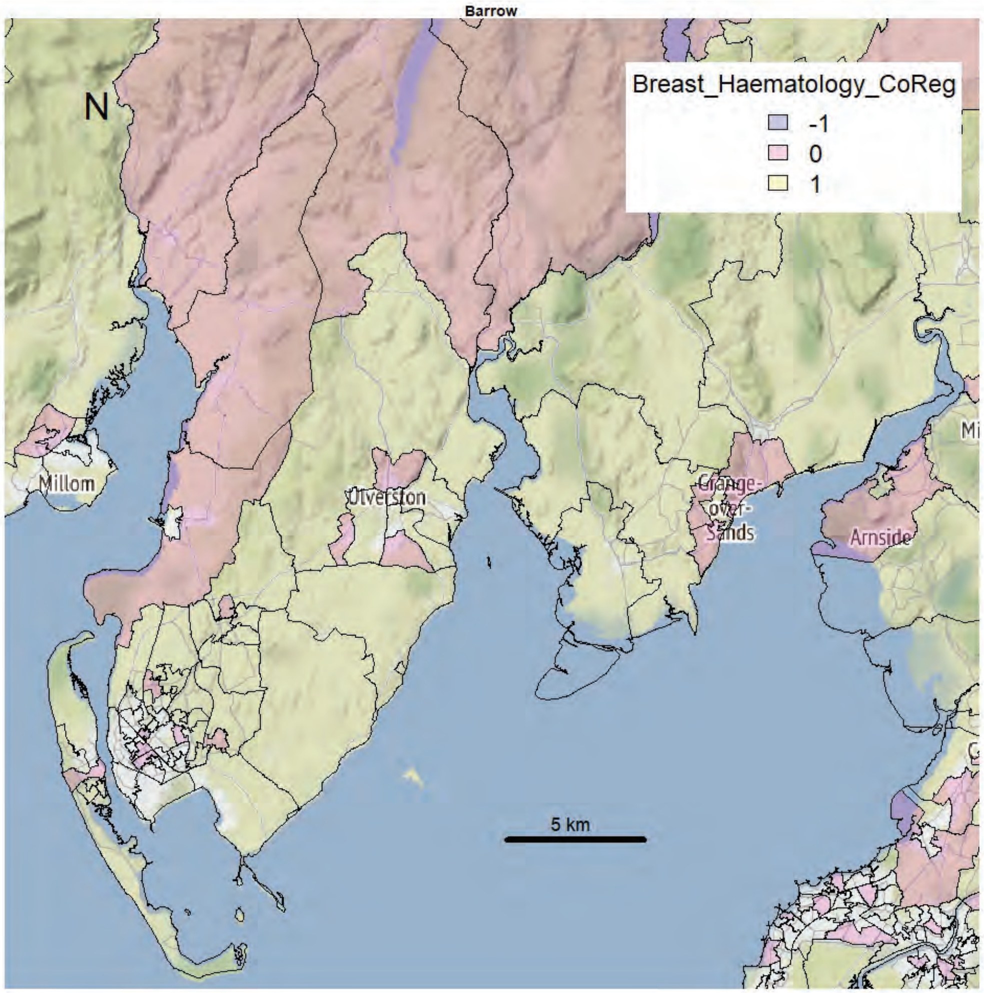
Barrow
N
Breast_Haematology_CoReg
-1
0
1
Millom
Ulverston
Grange-over-Sands
Arnside
5 km

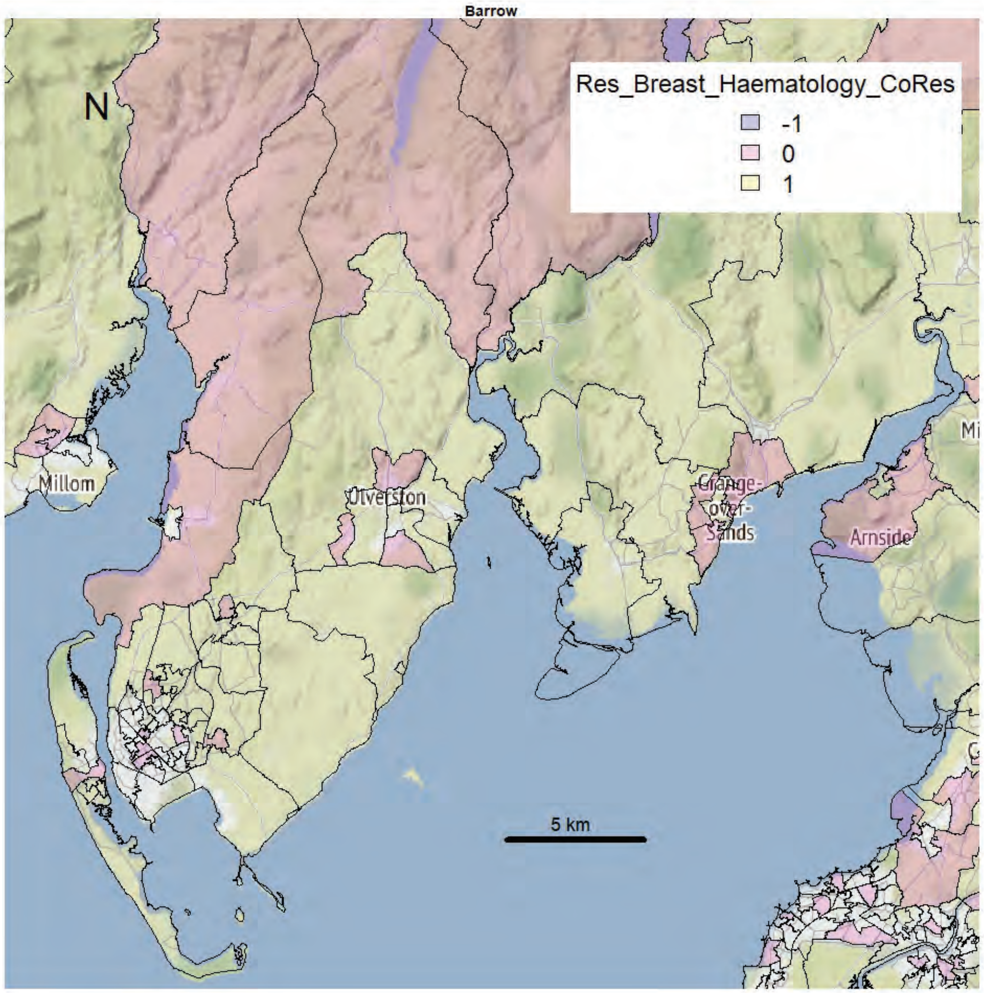
Barrow
N
Res_Breast_Haematology_CoRes
-1
0
1
Millom
Ulverston
Grange-over-Sands
Arnside
5 km

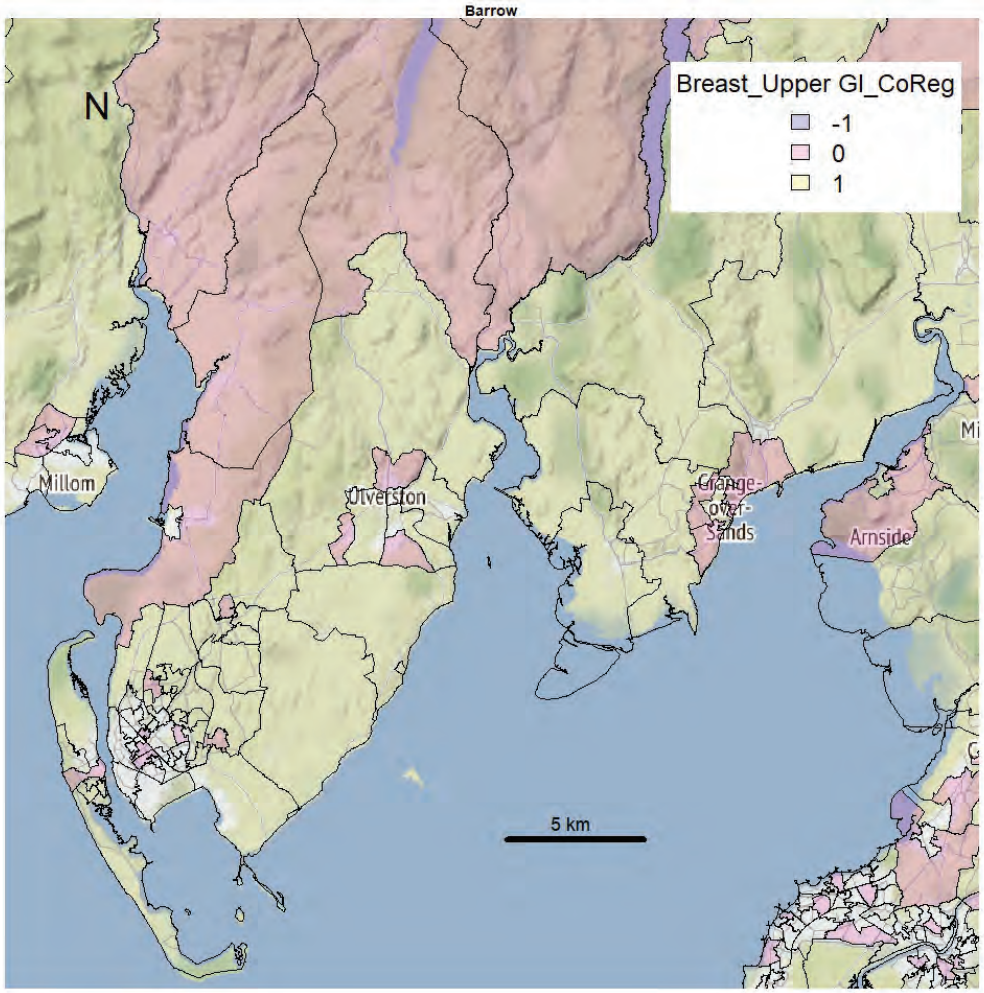
Barrow
N
Breast_Upper GI_CoReg
-1
0
1
Millom
Ulverston
Grange-over-Sands
Arnside
5 km

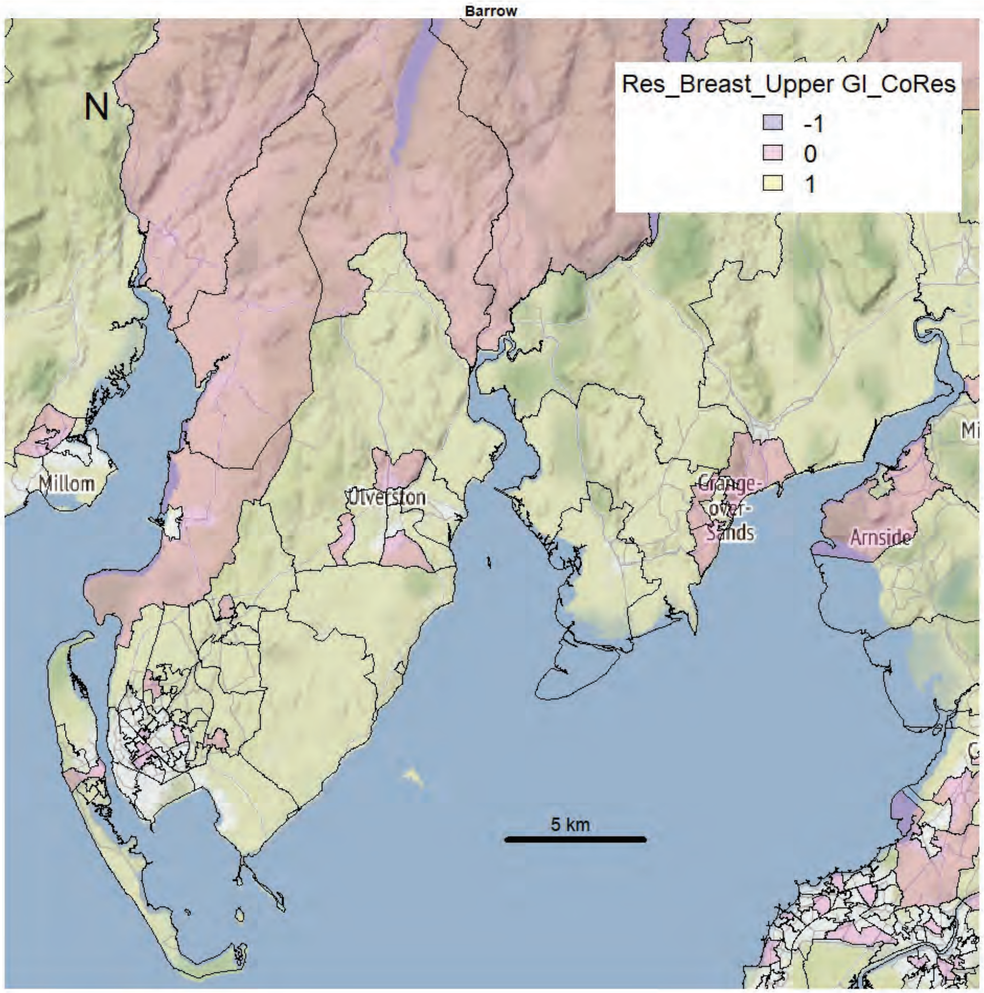
Barrow
N
Res_Breast_Upper GI_CoRes
-1
0
1
Millom
Ulverston
Grange-over-Sands
Arnside
5 km

Barrow

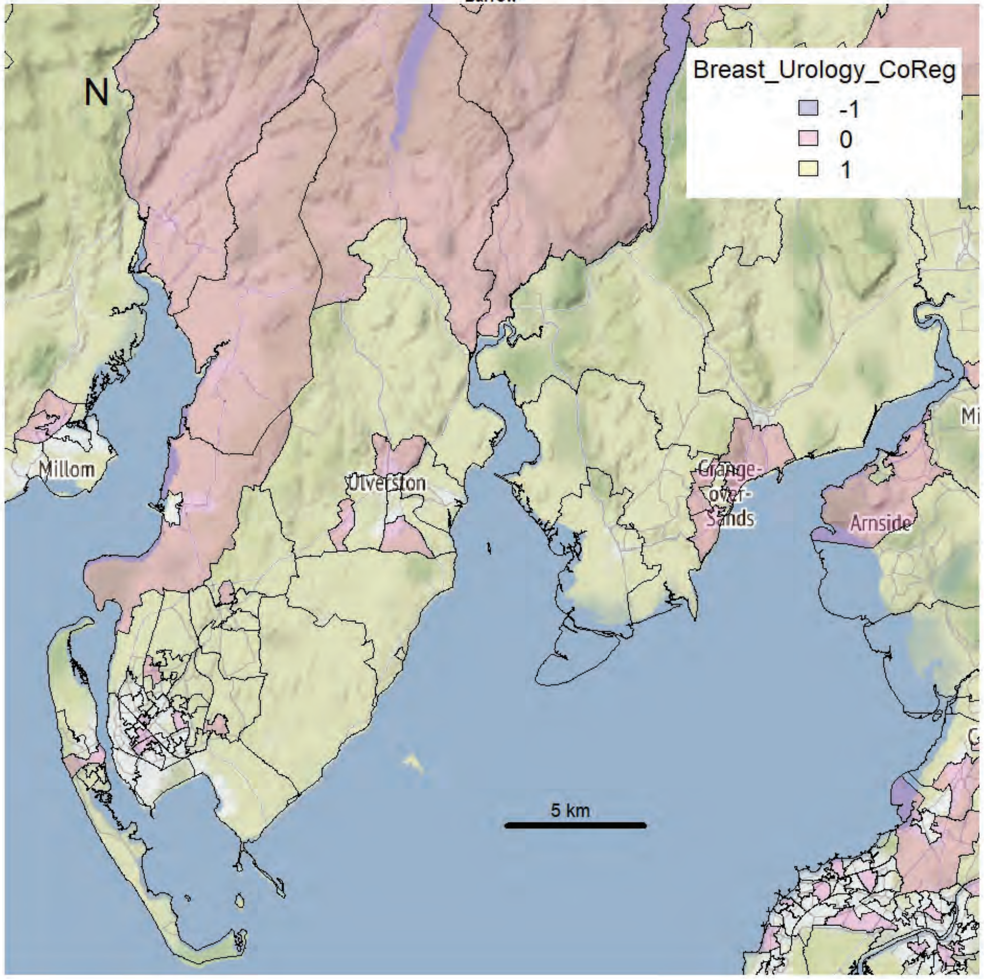


Barrow

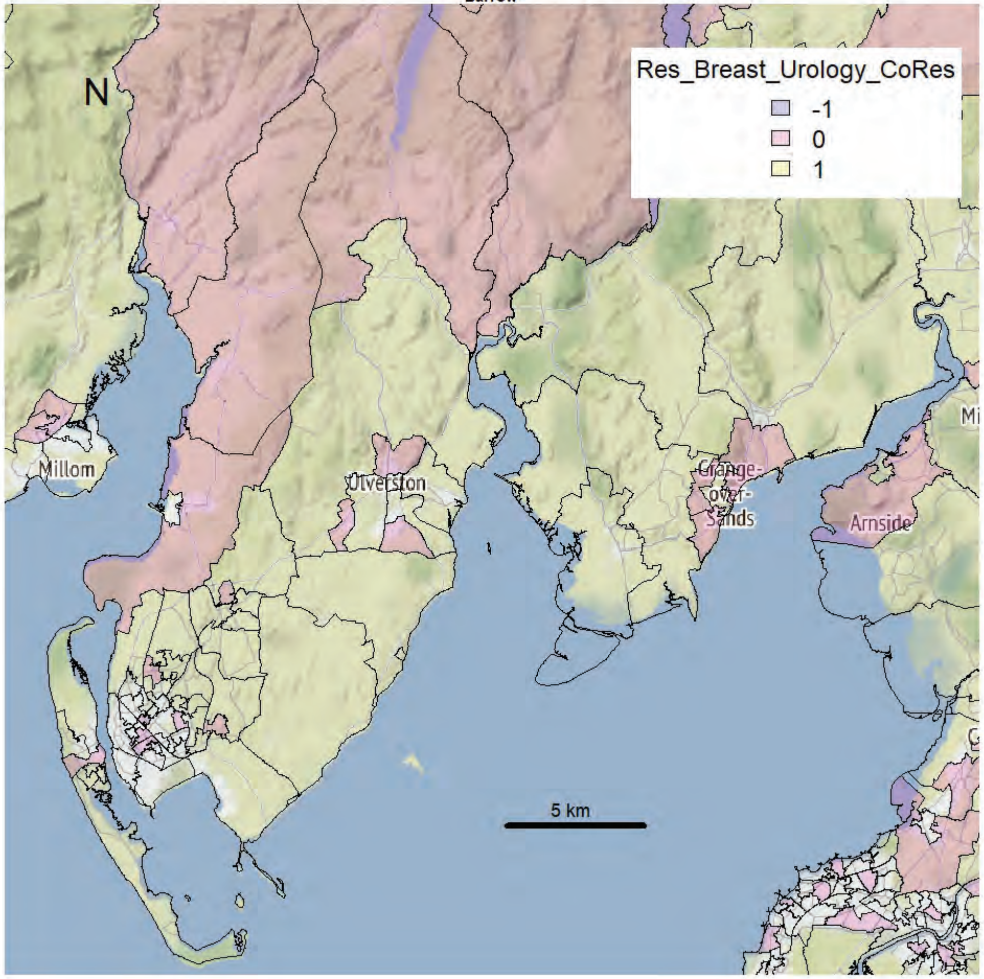

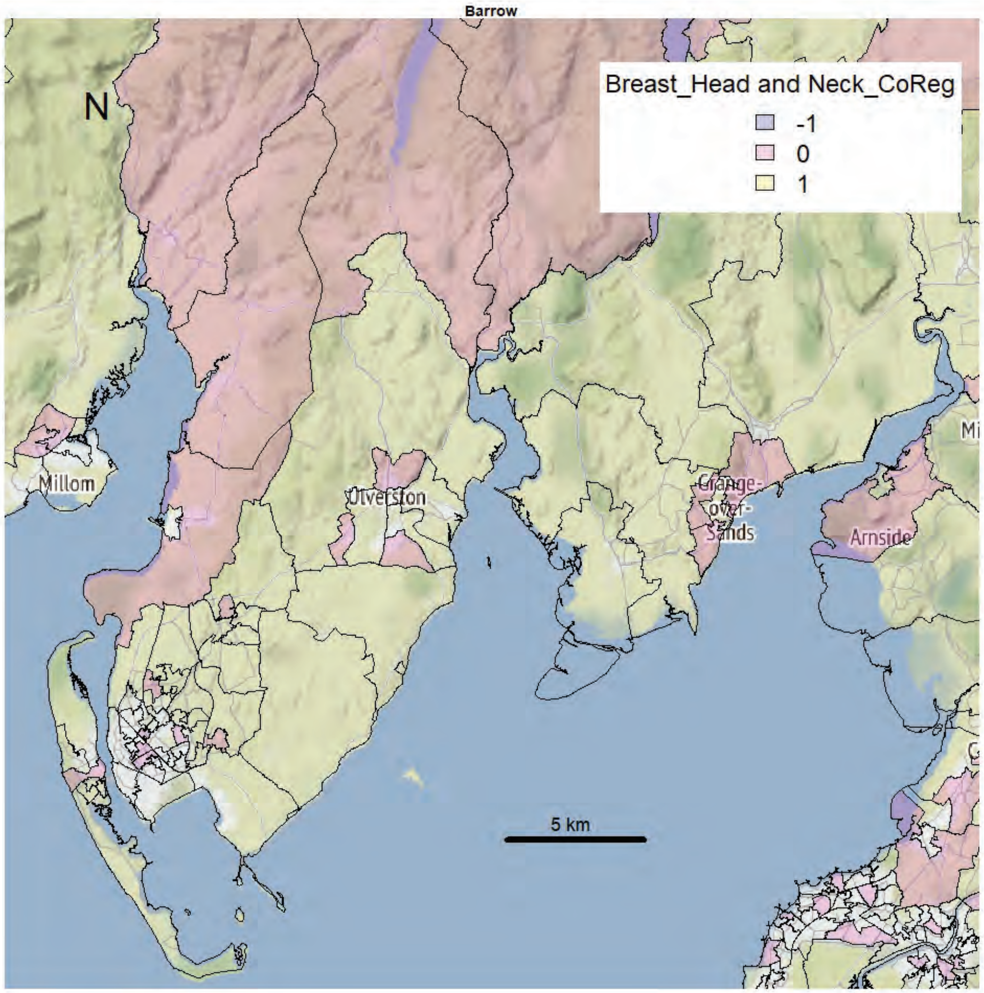
Barrow
Breast_Head and Neck_CoReg
-1
0
1
N
Millom
Ulverston
Grange-over-Sands
Arnside
5 km

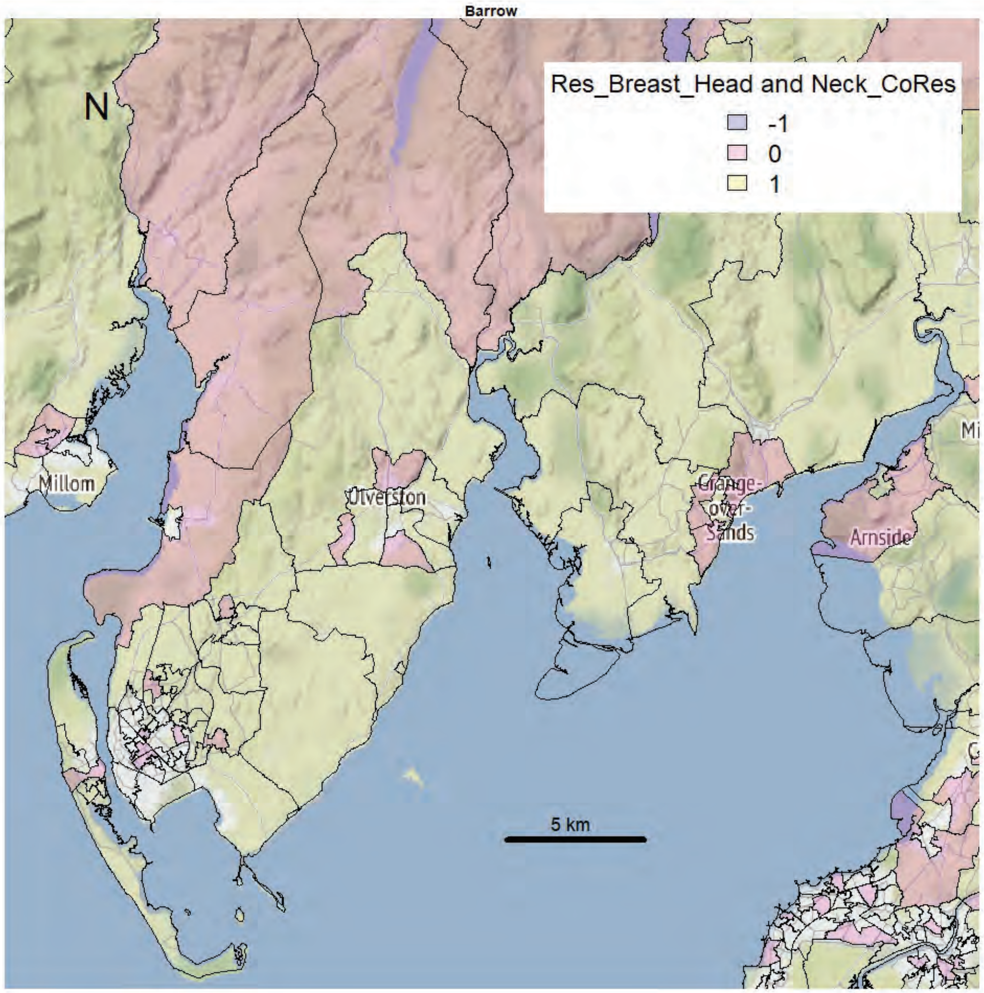
Barrow
Res_Breast_Head and Neck_CoRes
-1
0
1
N
Millom
Ulverston
Grange-over-Sands
Arnside
5 km

Barrow

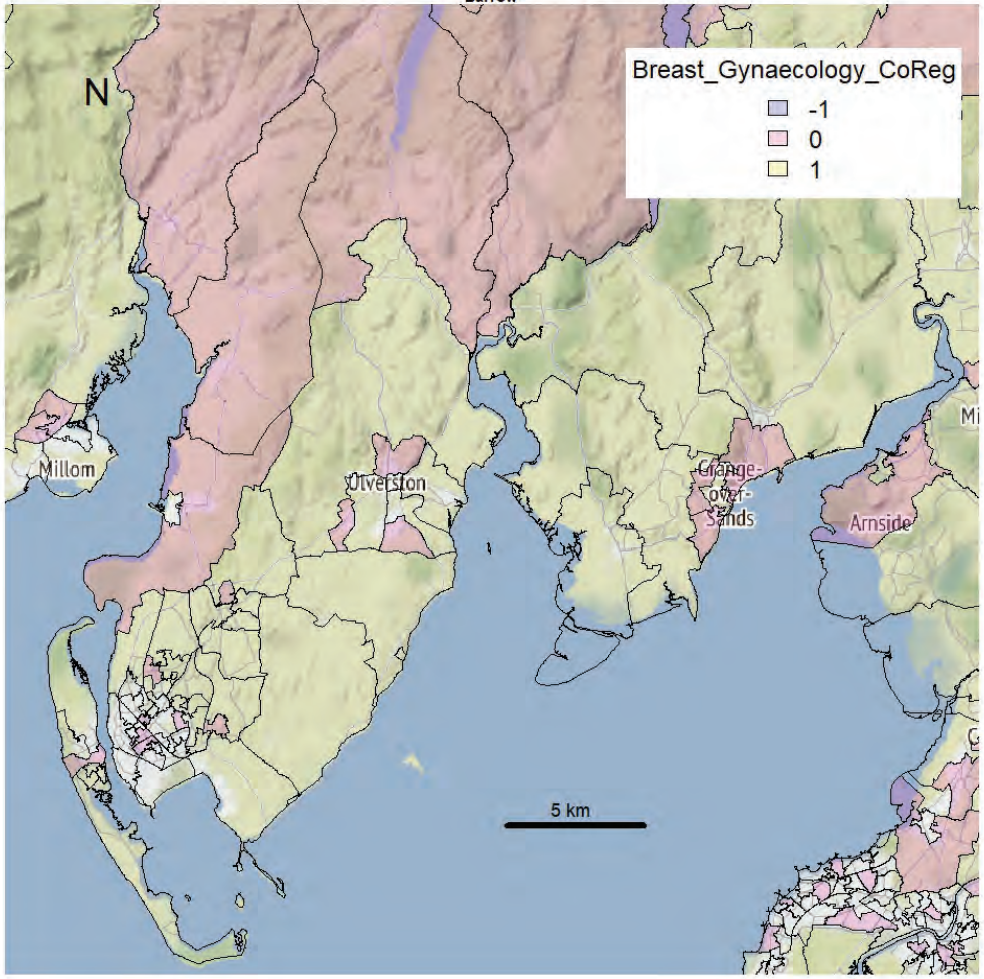


Barrow

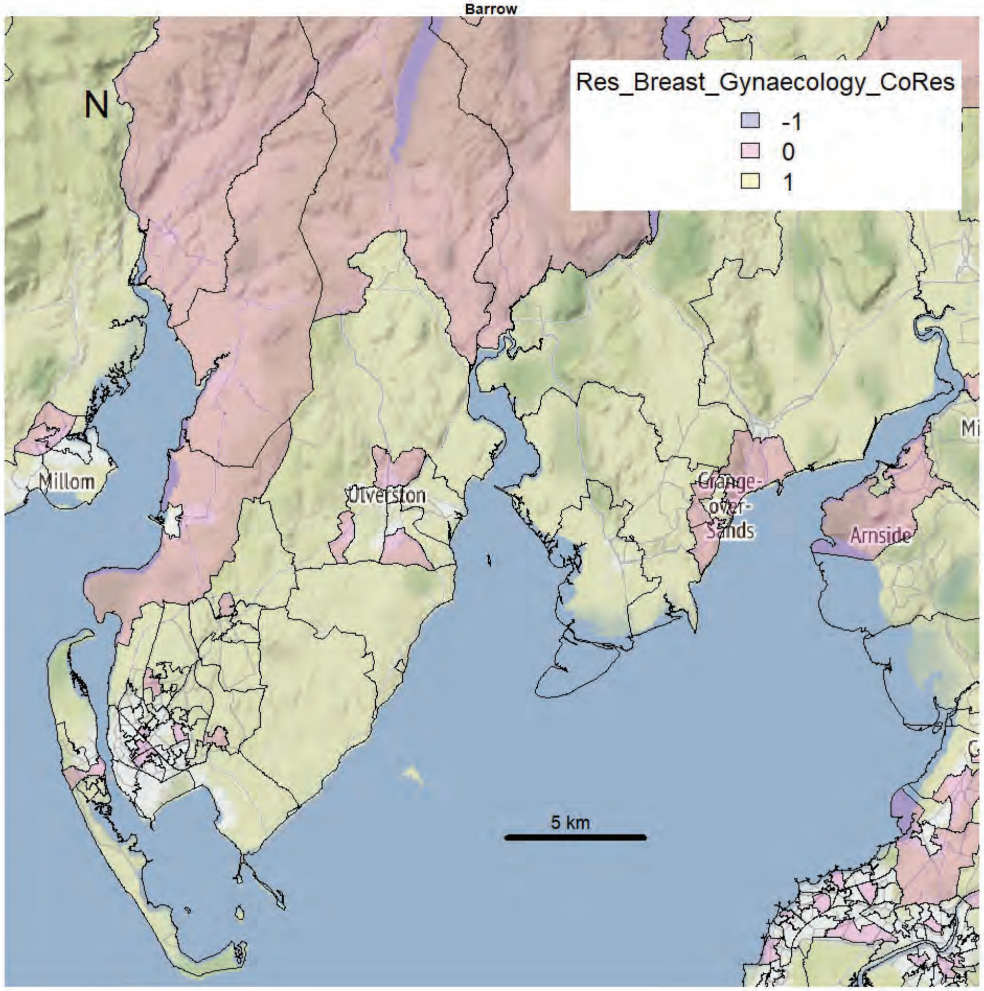

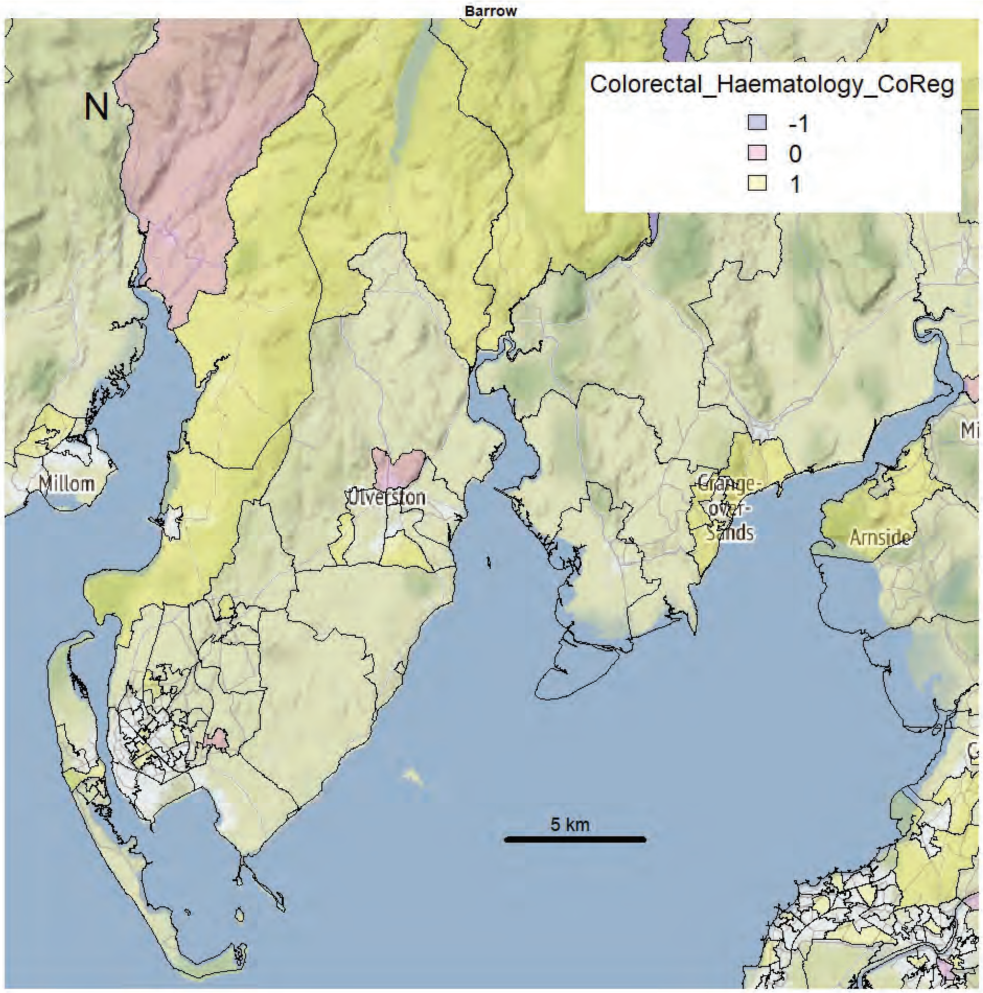
Barrow
N
Colorectal_Haematology_CoReg
-1
0
1
Millom
Ulverston
Grange-over-Sands
Arnside
5 km

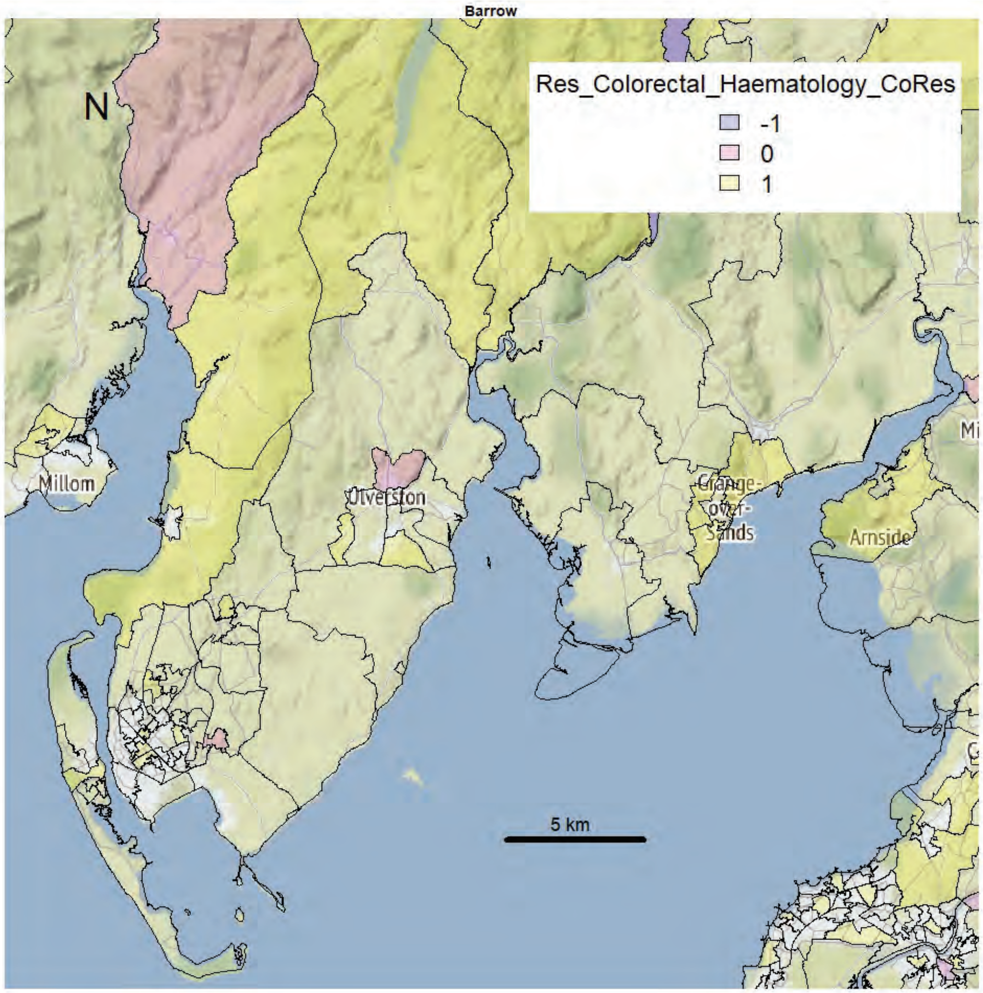
Barrow
N
Res_Colorectal_Haematology_CoRes
-1
0
1
Millom
Ulverston
Grange-over-Sands
Arnside
5 km

Barrow

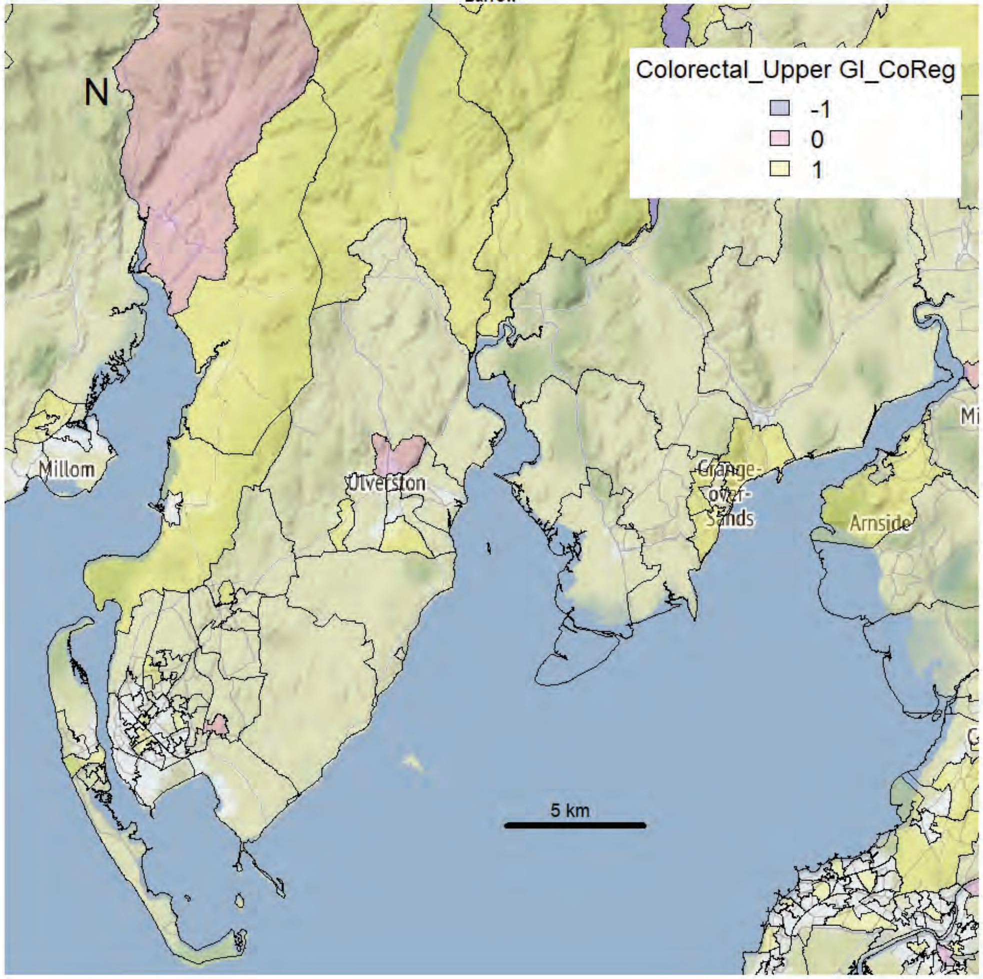


Barrow

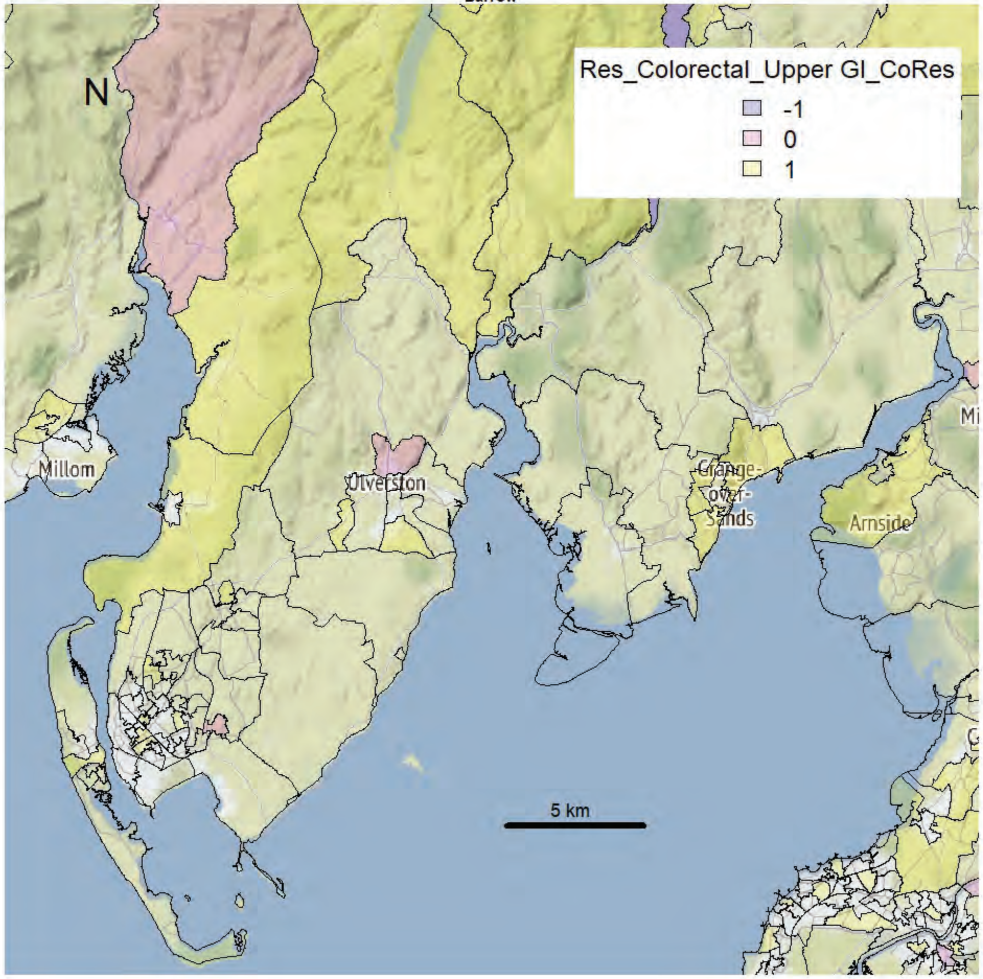

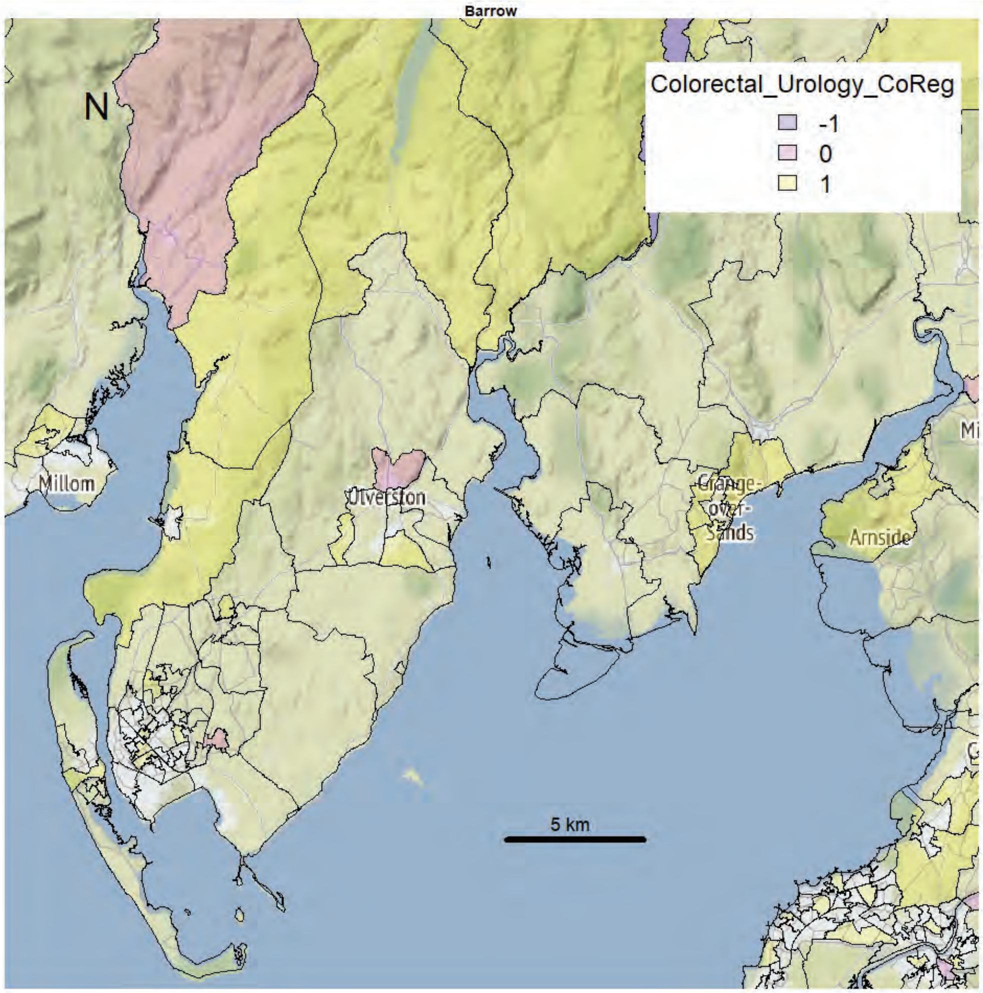
Barrow
N
Colorectal_Urology_CoReg
-1
0
1
Millom
Ulverston
Grange-over-Sands
Arnside
5 km

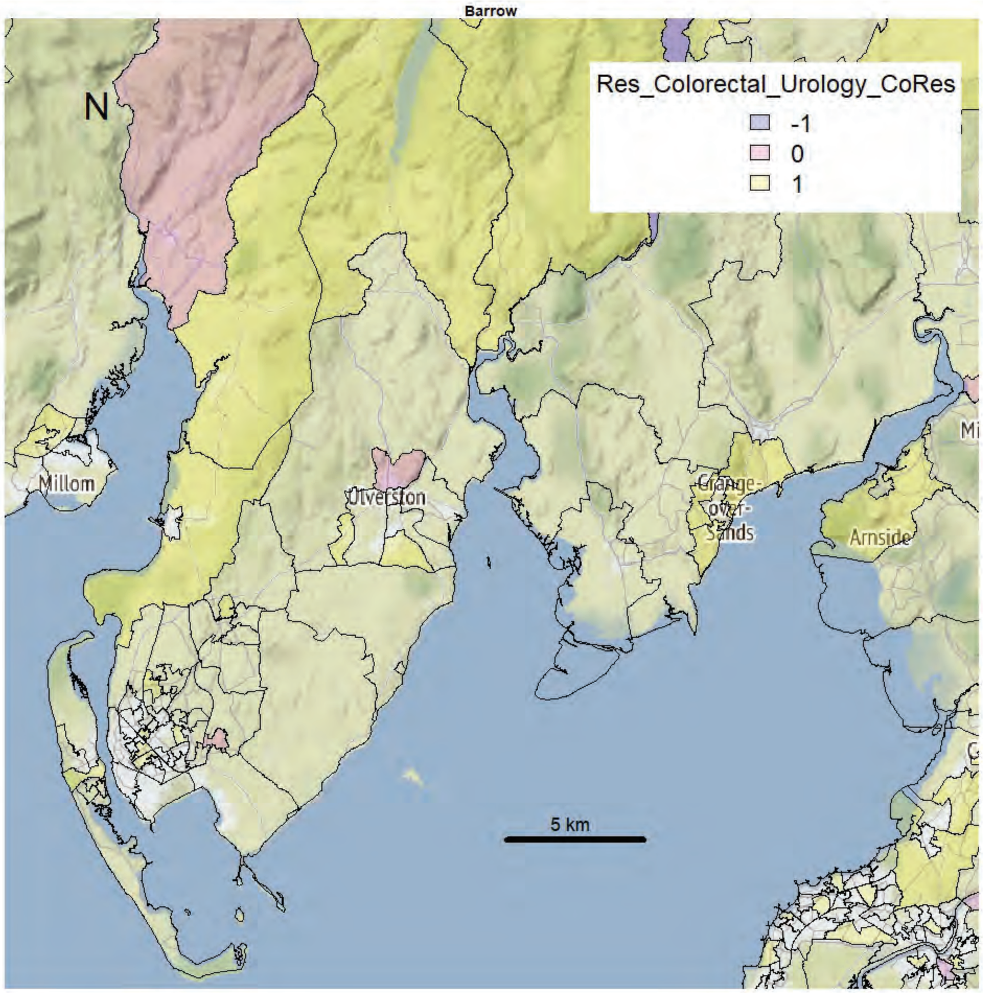
Barrow
N
Res_Colorectal_Urology_CoRes
-1
0
1
Millom
Ulverston
Grange-over-Sands
Arnside
5 km

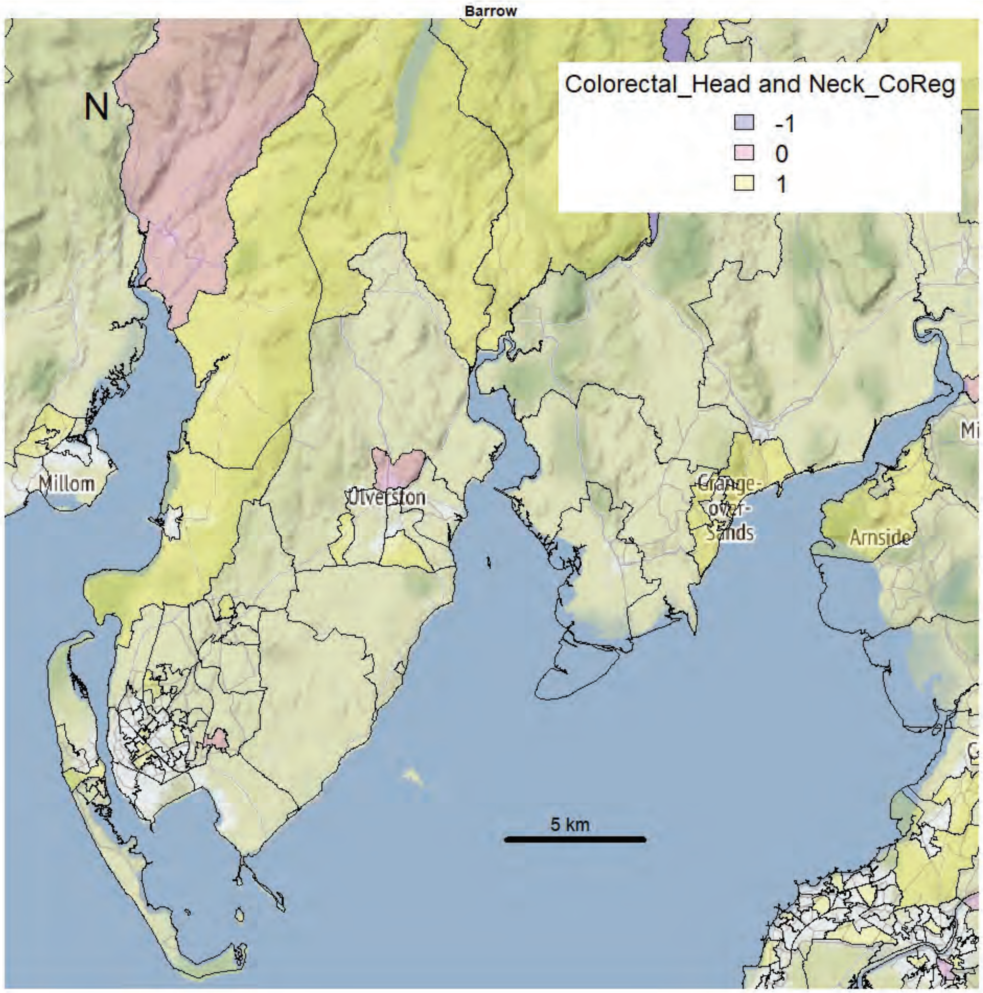
Barrow
N
Colorectal_Head and Neck_CoReg
-1
0
1
Millom
Ulverston
Grange-over-Sands
Arnside
5 km

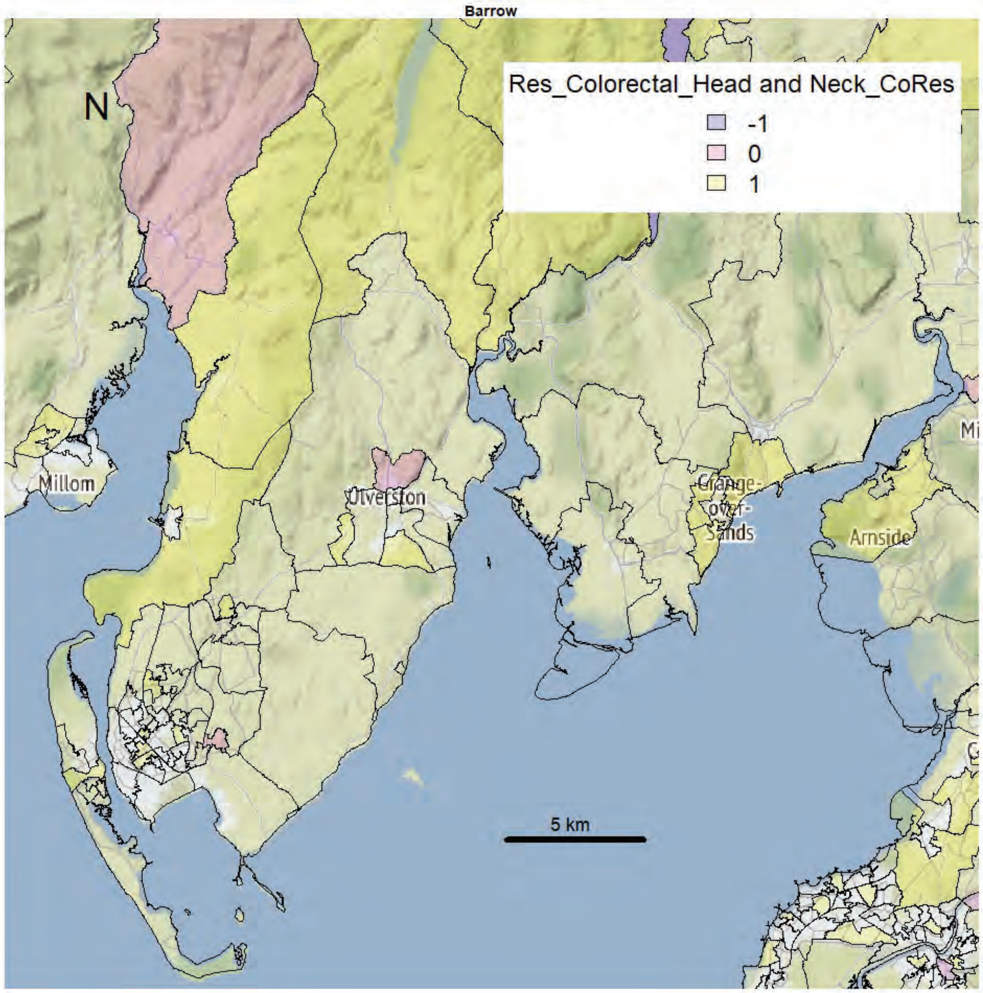
Barrow
N
Res_Colorectal_Head and Neck_CoRes
-1
0
1
Millom
Ulverston
Grange-over-Sands
Arnside
5 km

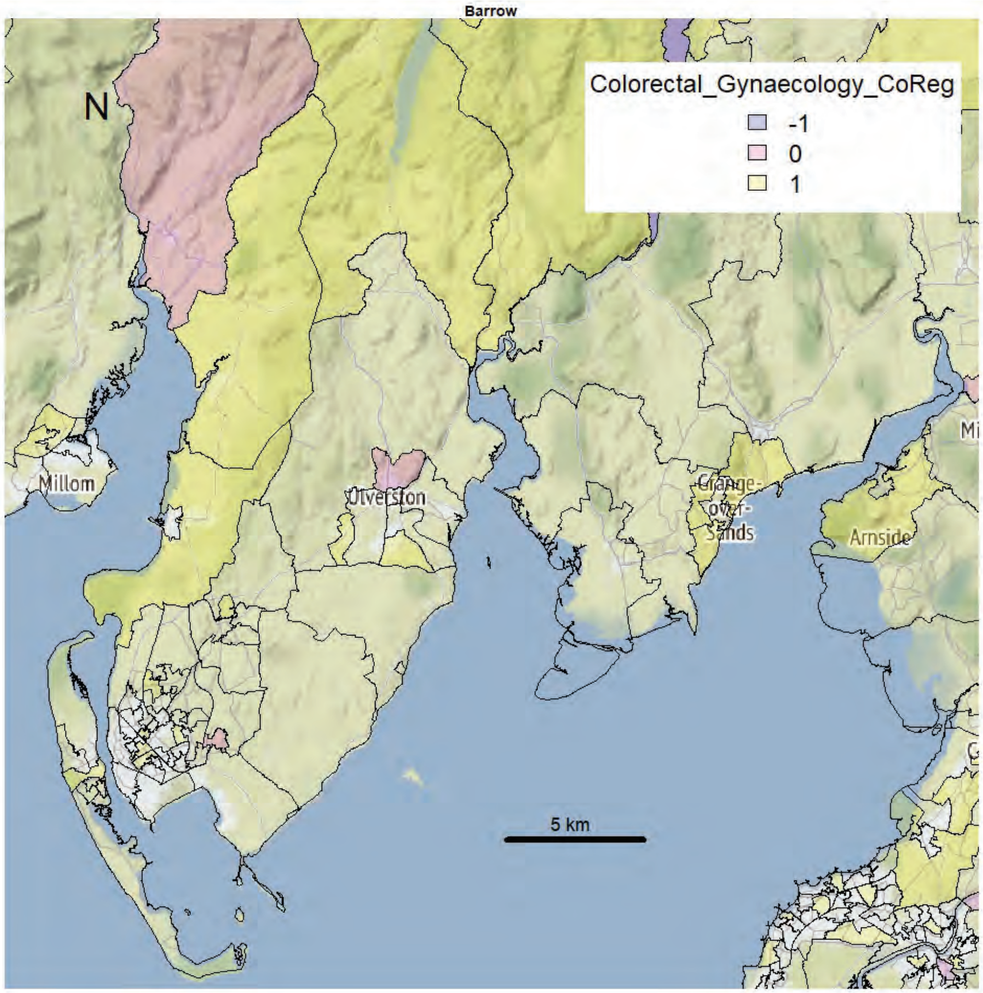
Barrow
N
Colorectal_Gynaecology_CoReg
-1
0
1
Millom
Ulverston
Grange-over-Sands
Arnside
5 km

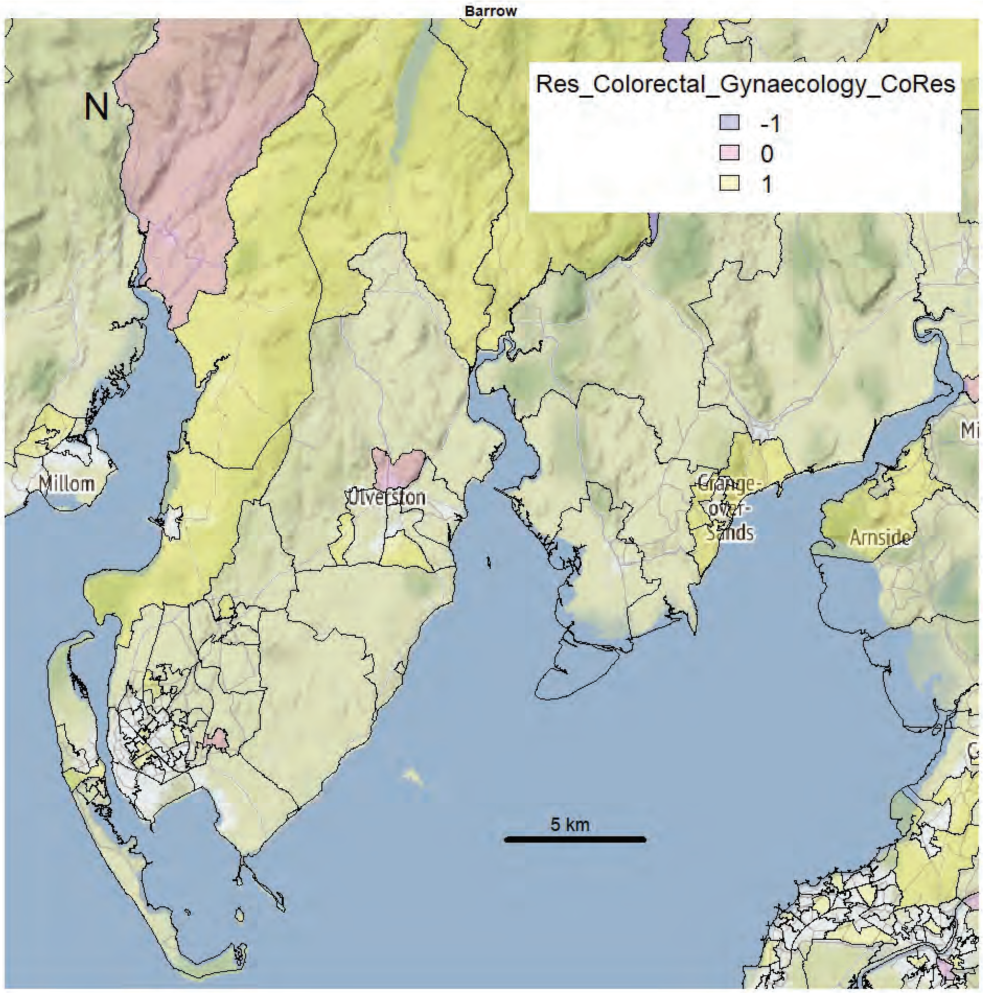
Barrow
N
Res_Colorectal_Gynaecology_CoRes
-1
0
1
Millom
Ulverston
Grange-over-Sands
Arnside
5 km

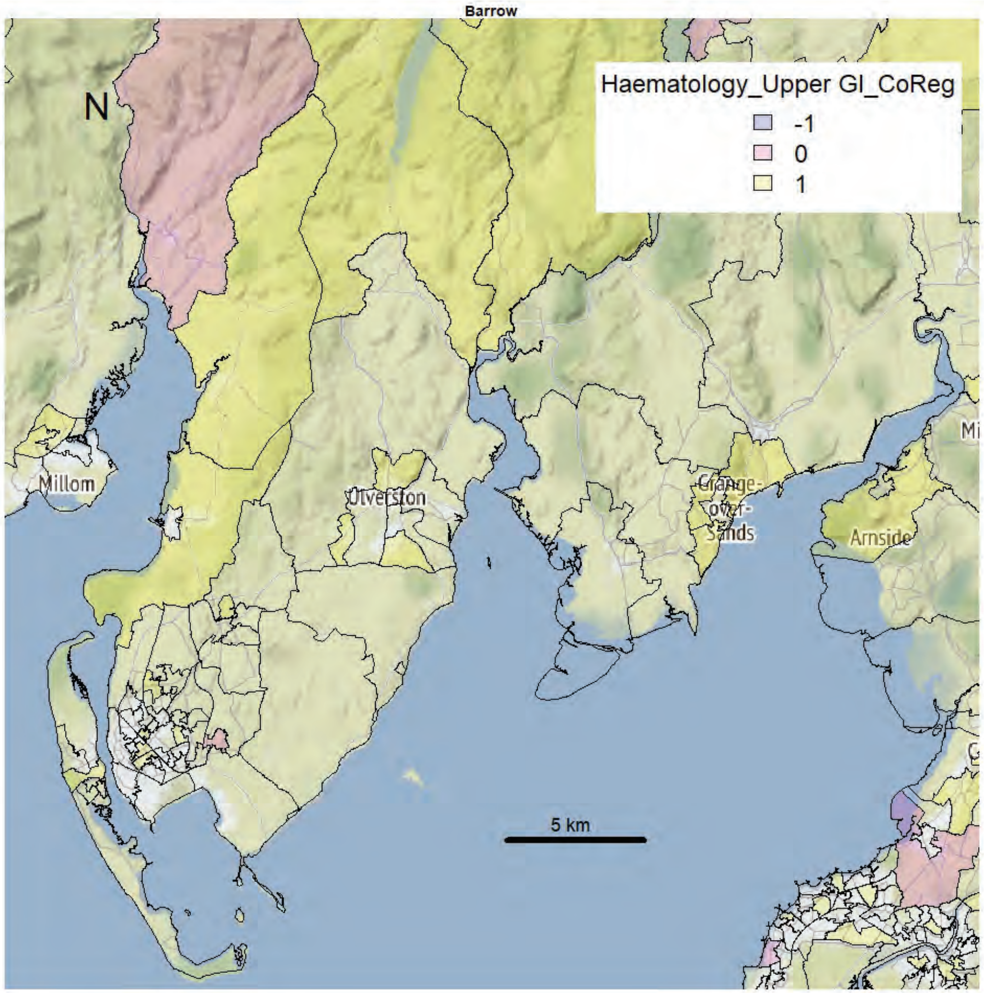
Barrow
N
Haematology_Upper GI_CoReg
-1
0
1
Millom
Ulverston
Grange-over-Sands
Arnside
5 km

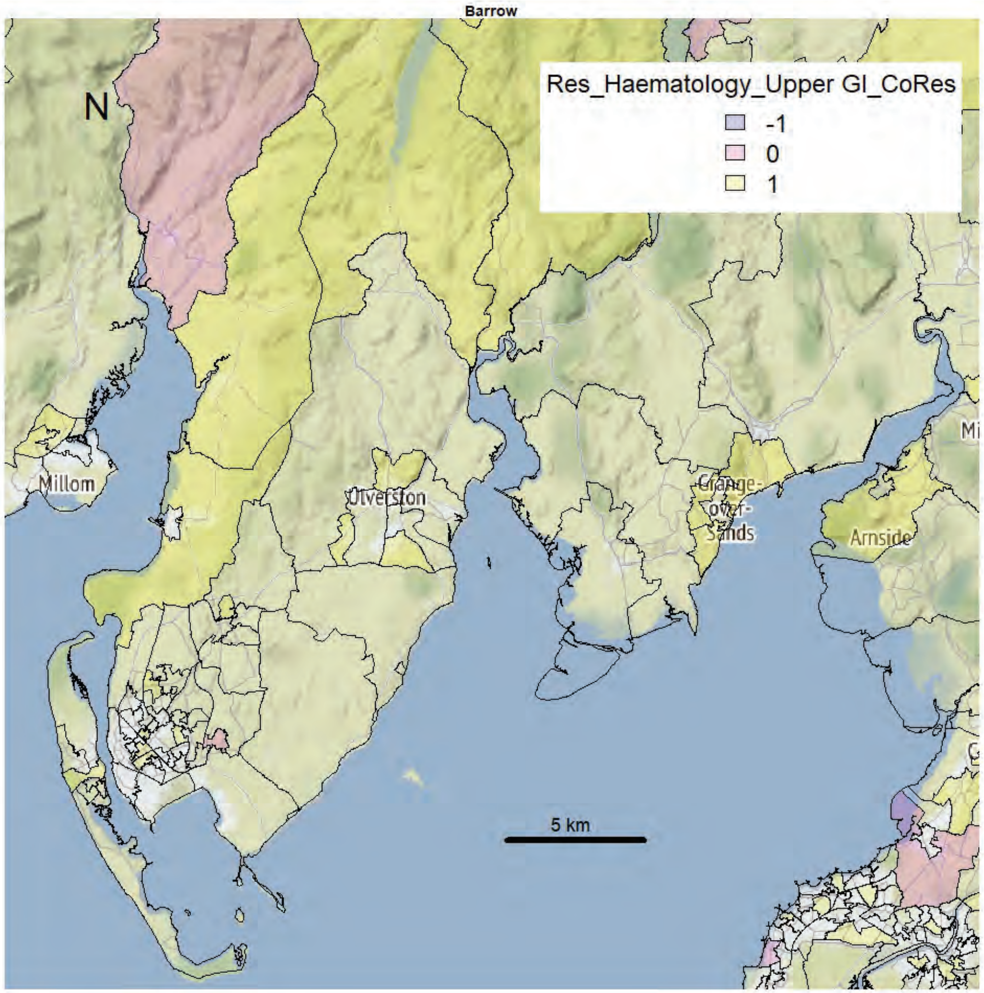
Barrow
N
Res_Haematology_Upper GI_CoRes
-1
0
1
Millom
Ulverston
Grange-over-Sands
Arnside
5 km

Barrow

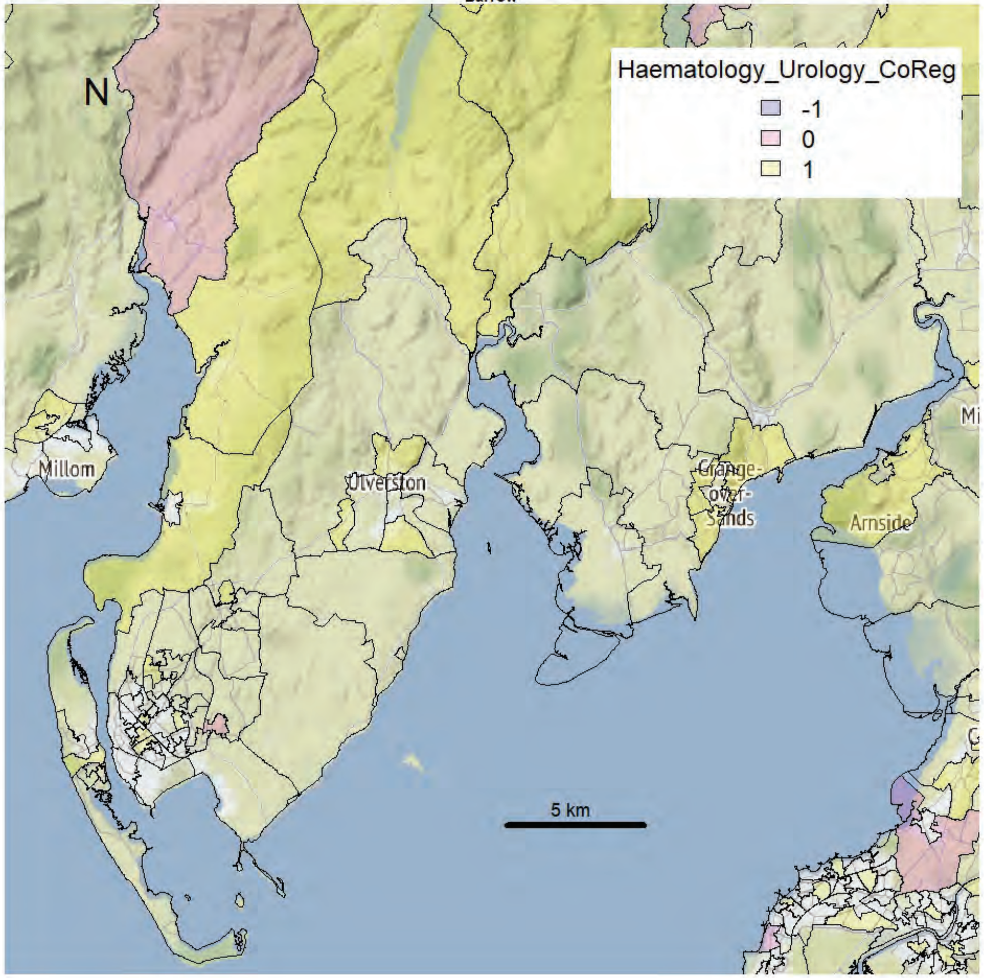


Barrow

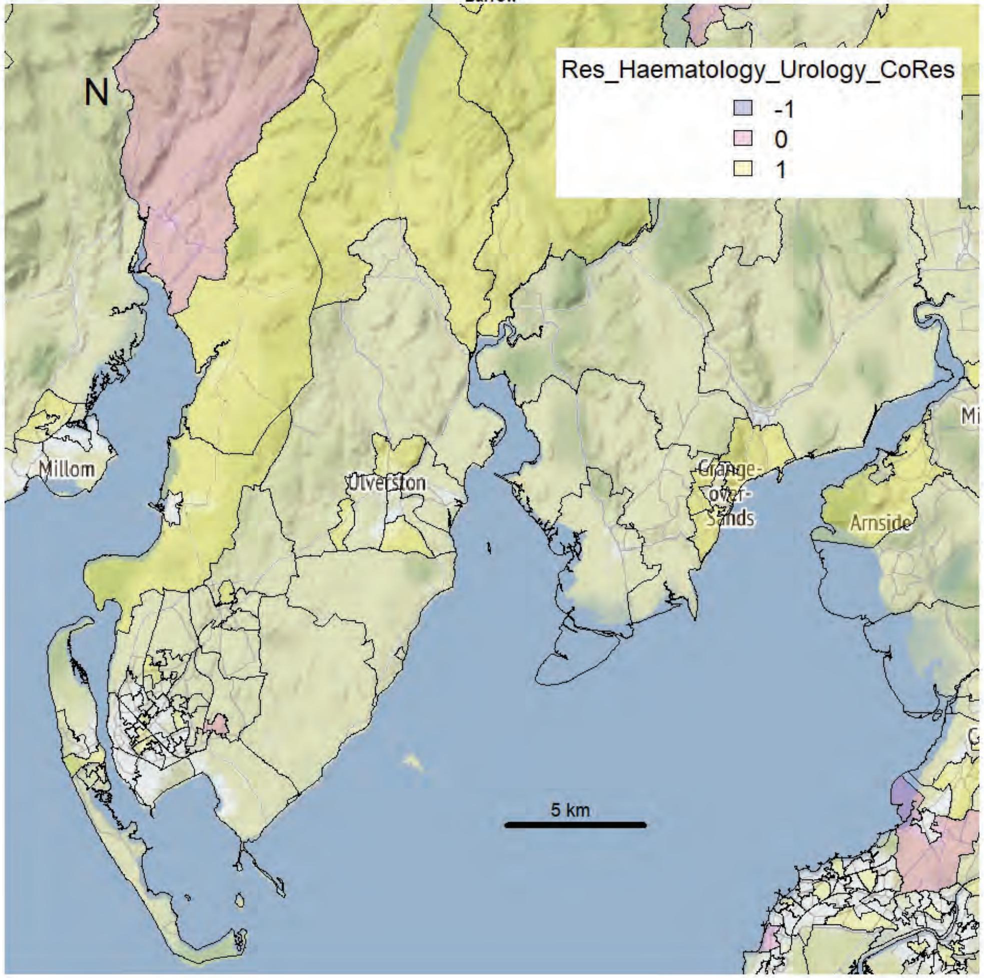

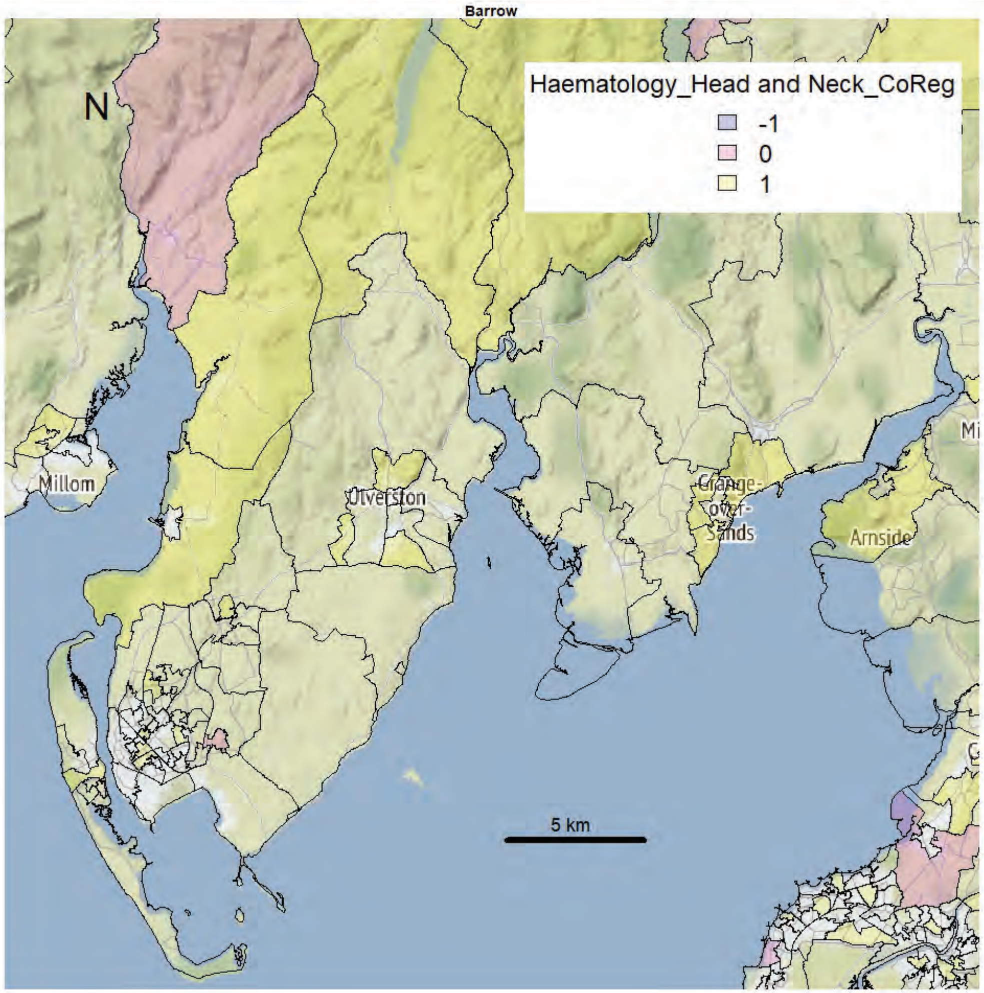
Barrow
N
Haematology_Head and Neck_CoReg
-1
0
1
Millom
Ulverston
Grange-over-Sands
Arnside
5 km

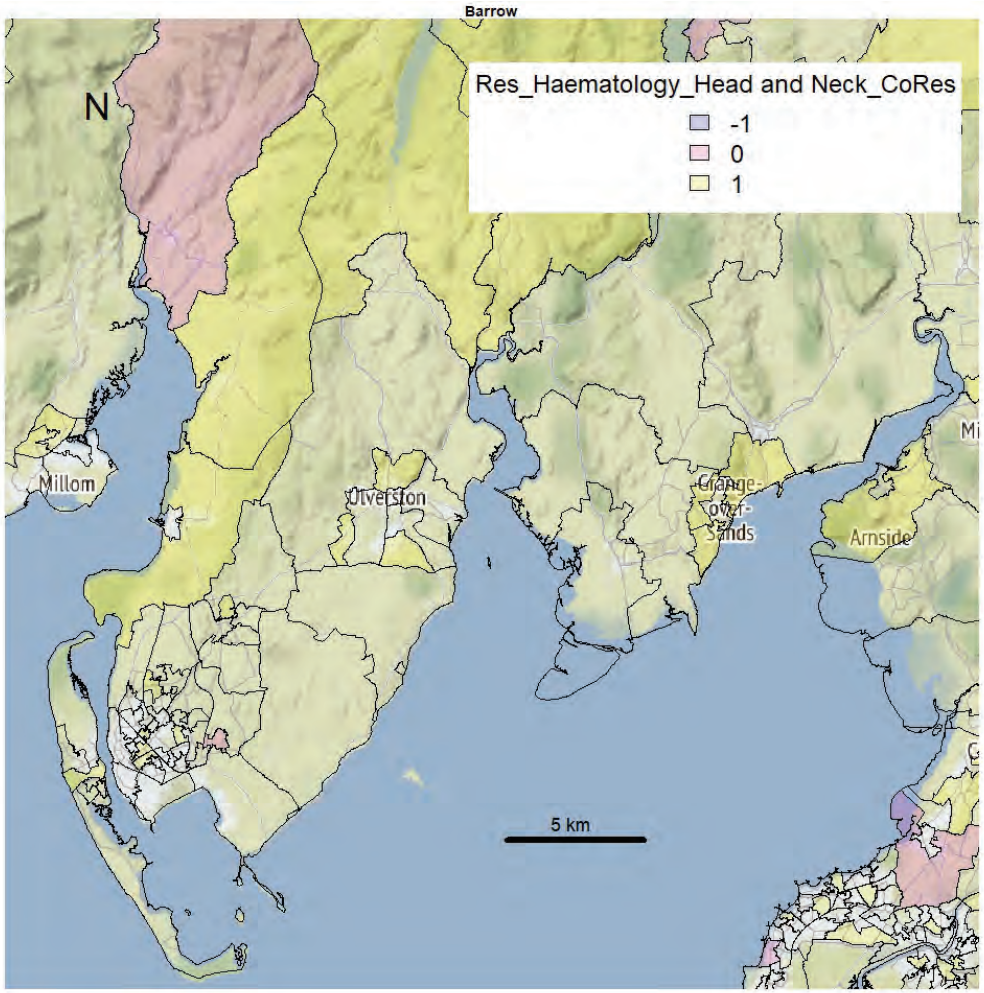
Barrow
N
Res_Haematology_Head and Neck_CoRes
-1
0
1
Millom
Ulverston
Grange-over-Sands
Arnside
5 km

Barrow

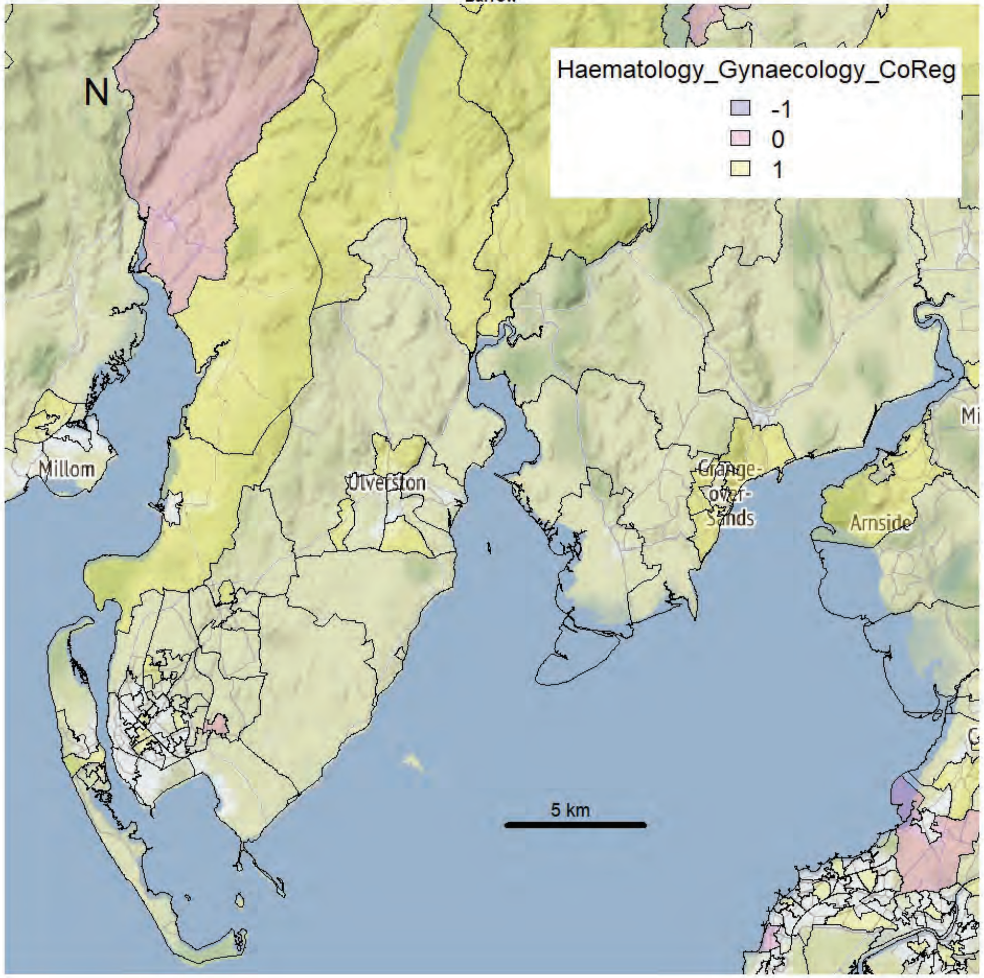


Barrow

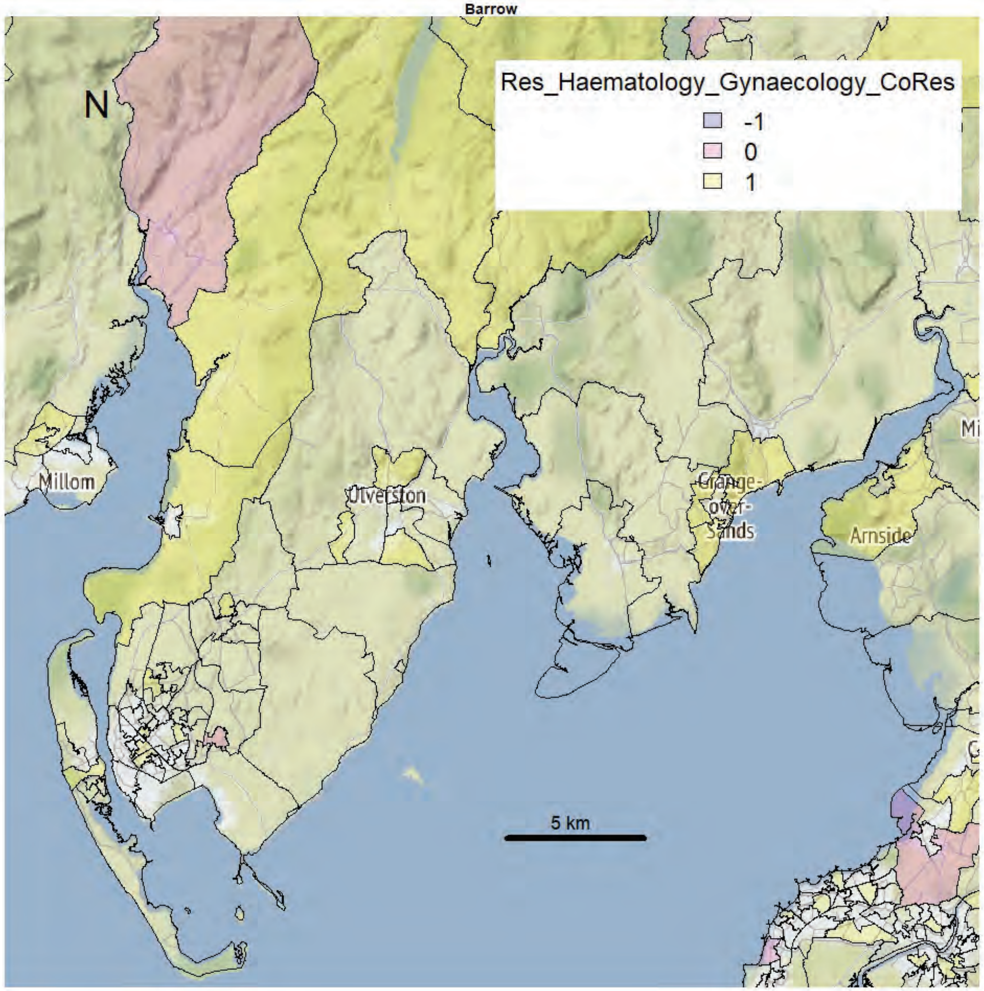

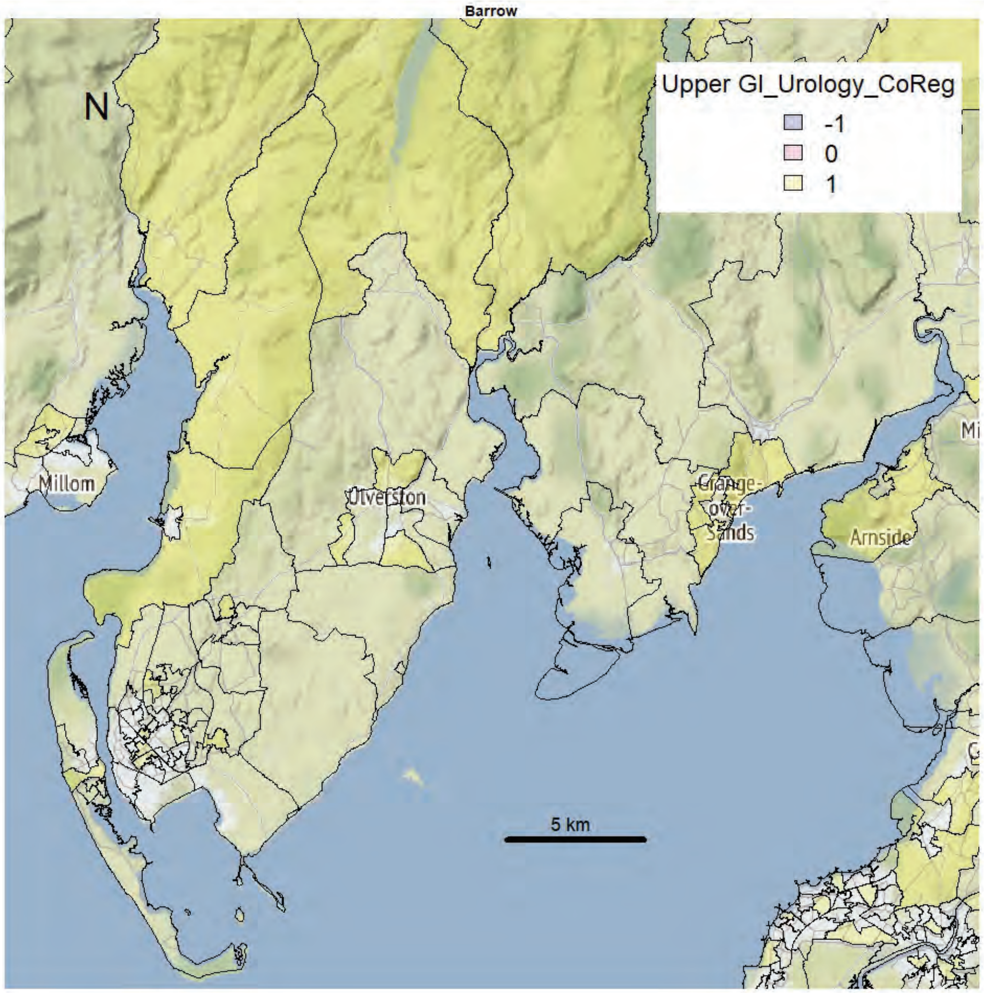
Barrow
N
Upper GI_Urology_CoReg
-1
0
1
Millom
Ulverston
Grange-over-Sands
Arnside
5 km

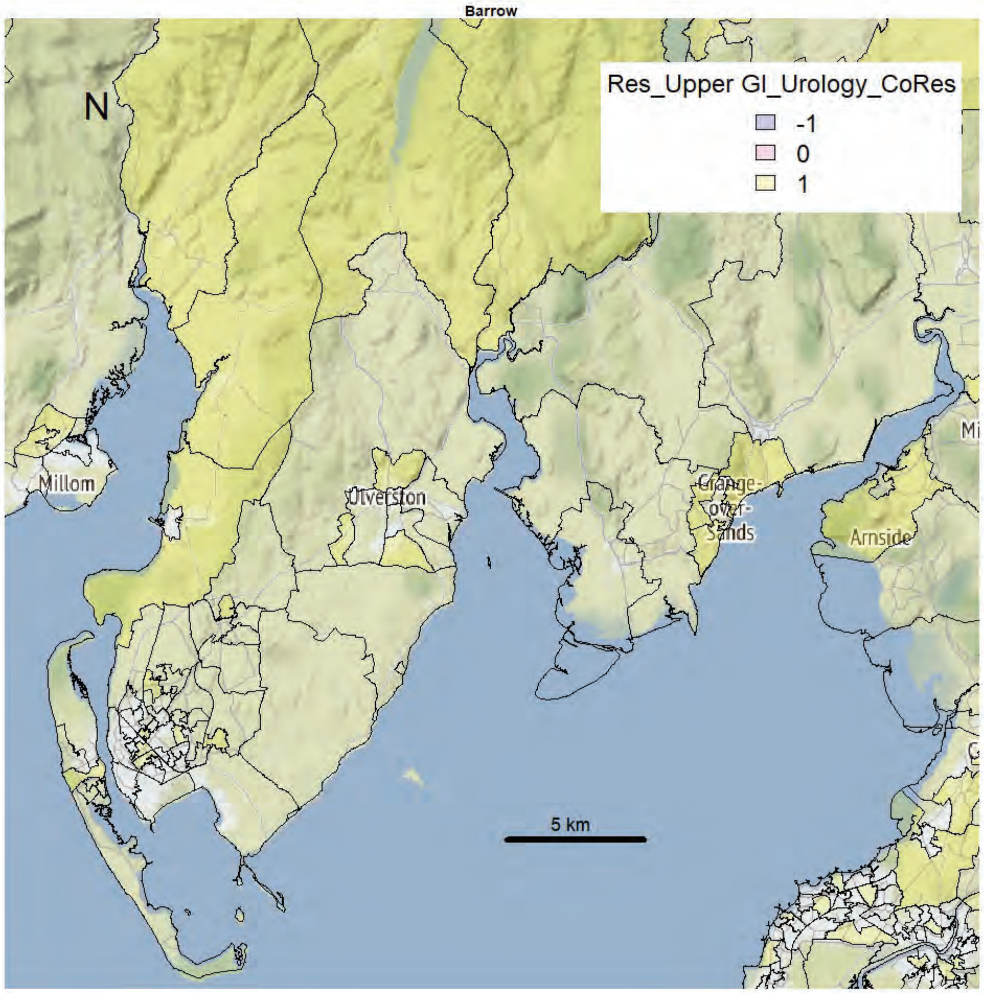
Barrow
N
Res_Upper GI_Urology_CoRes
-1
0
1
Millom
Ulverston
Grange-over-Sands
Arnside
5 km

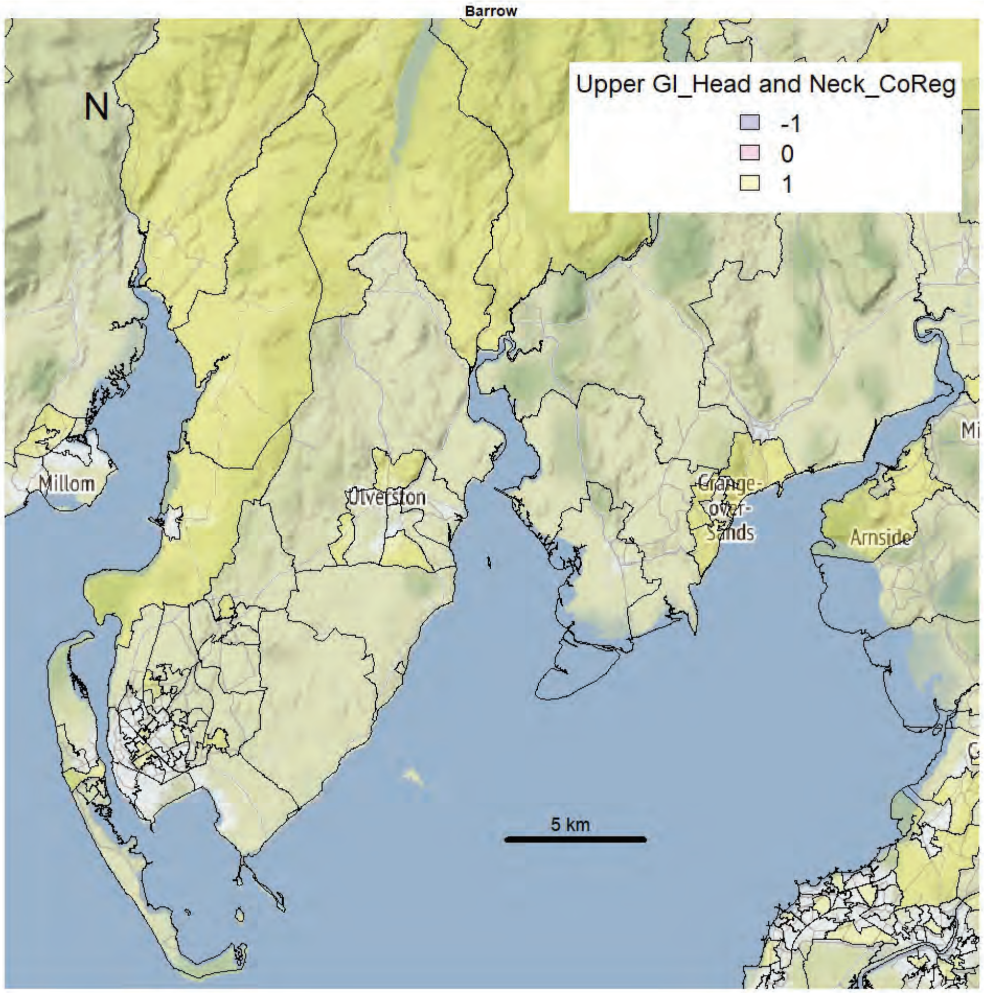
Barrow
Upper GI_Head and Neck_CoReg
-1
0
1
N
Millom
Ulverston
Grange-over-Sands
Arnside
5 km

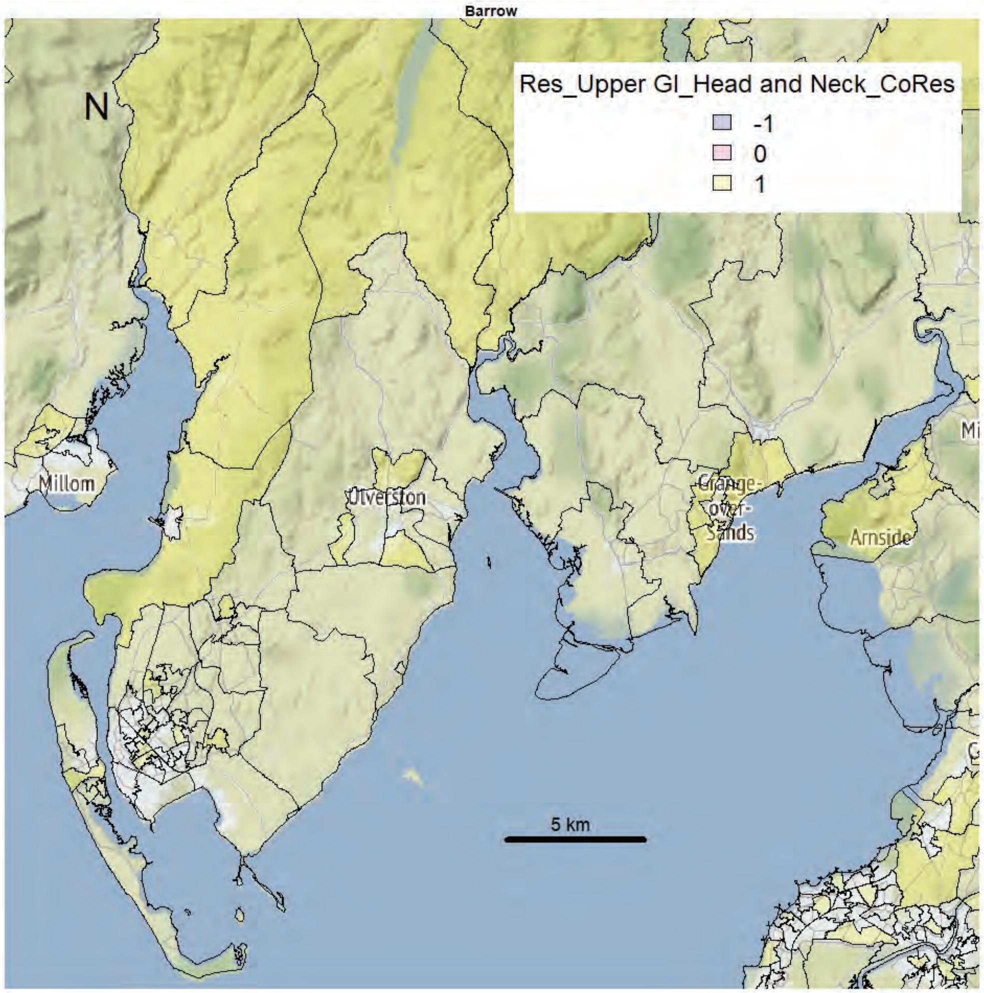
Barrow
Res_Upper GI_Head and Neck_CoRes
-1
0
1
N
Millom
Ulverston
Grange-over-Sands
Arnside
5 km

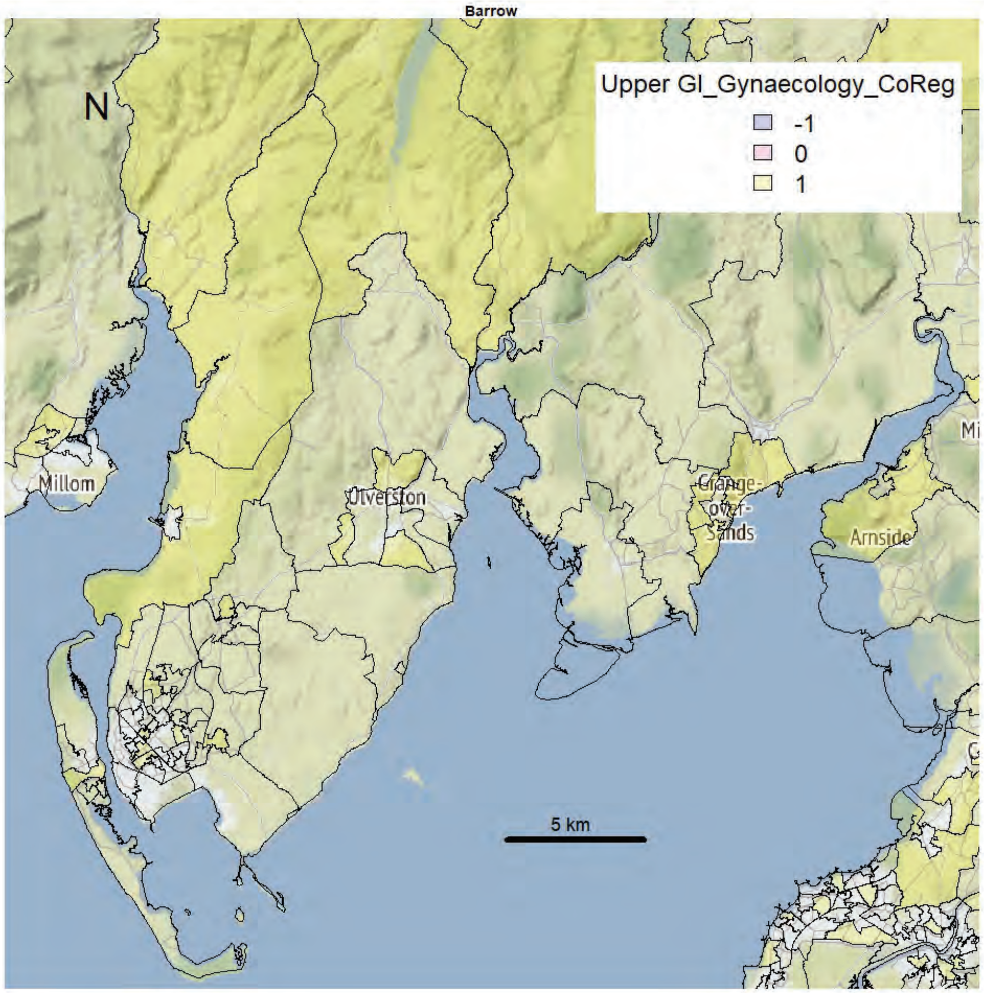
Barrow
N
Upper GI_Gynaecology_CoReg
-1
0
1
Millom
Ulverston
Grange-over-Sands
Arnside
5 km

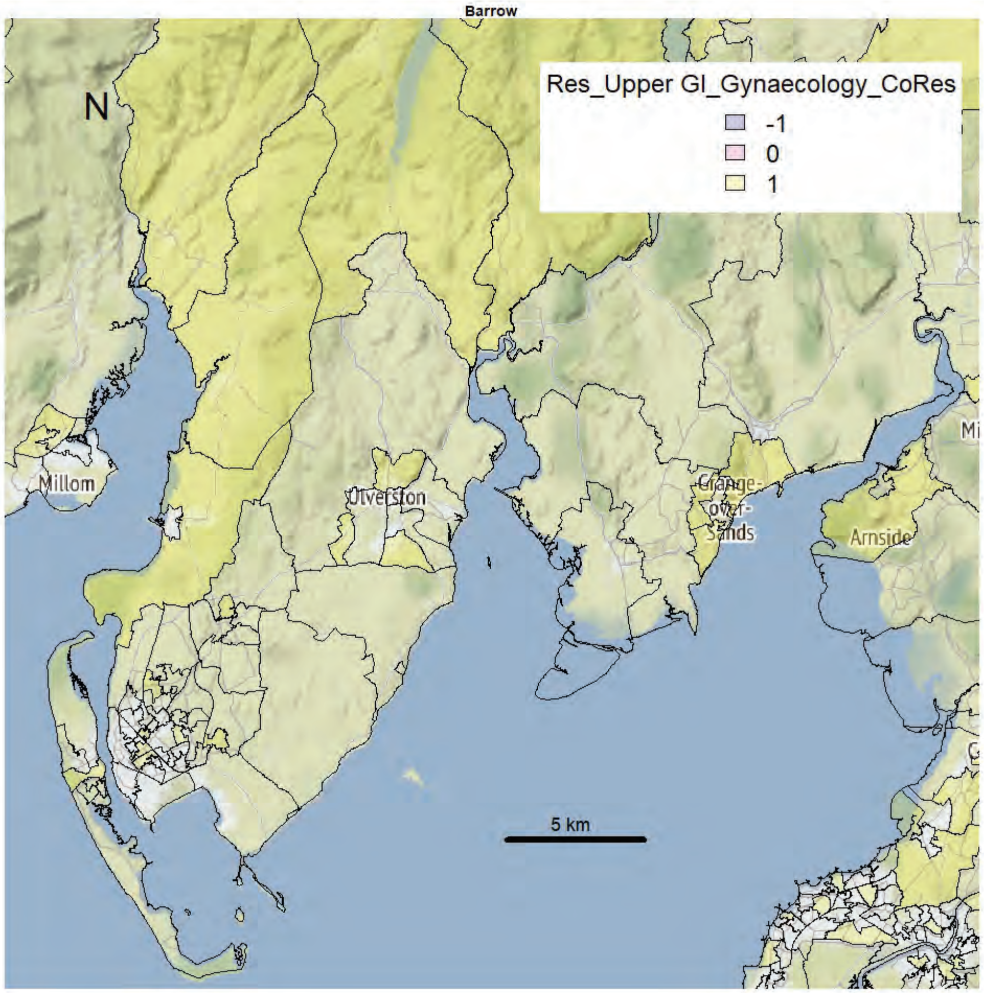
Barrow
N
Res_Upper GI_Gynaecology_CoRes
-1
0
1
Millom
Ulverston
Grange-over-Sands
Arnside
5 km

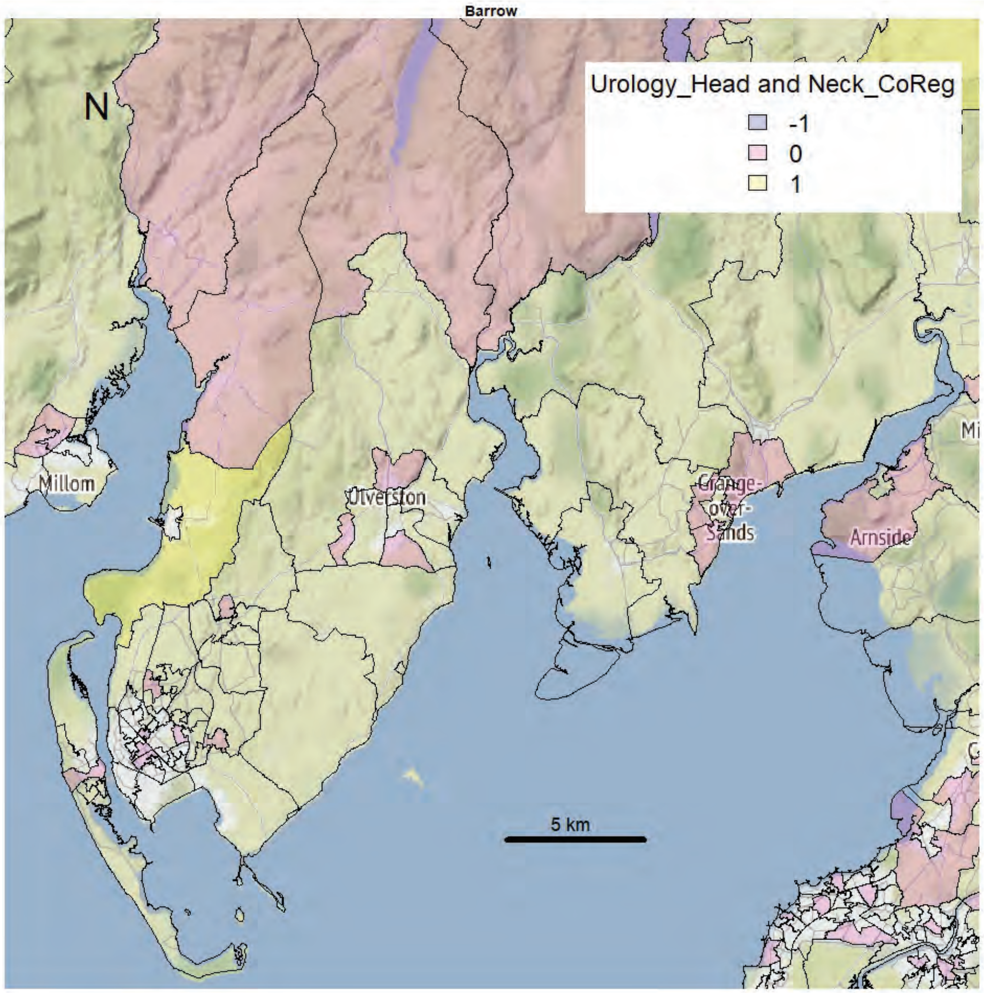
Barrow
N
Urology_Head and Neck_CoReg
-1
0
1
Millom
Ulverston
Grange-over-Sands
Arnside
5 km

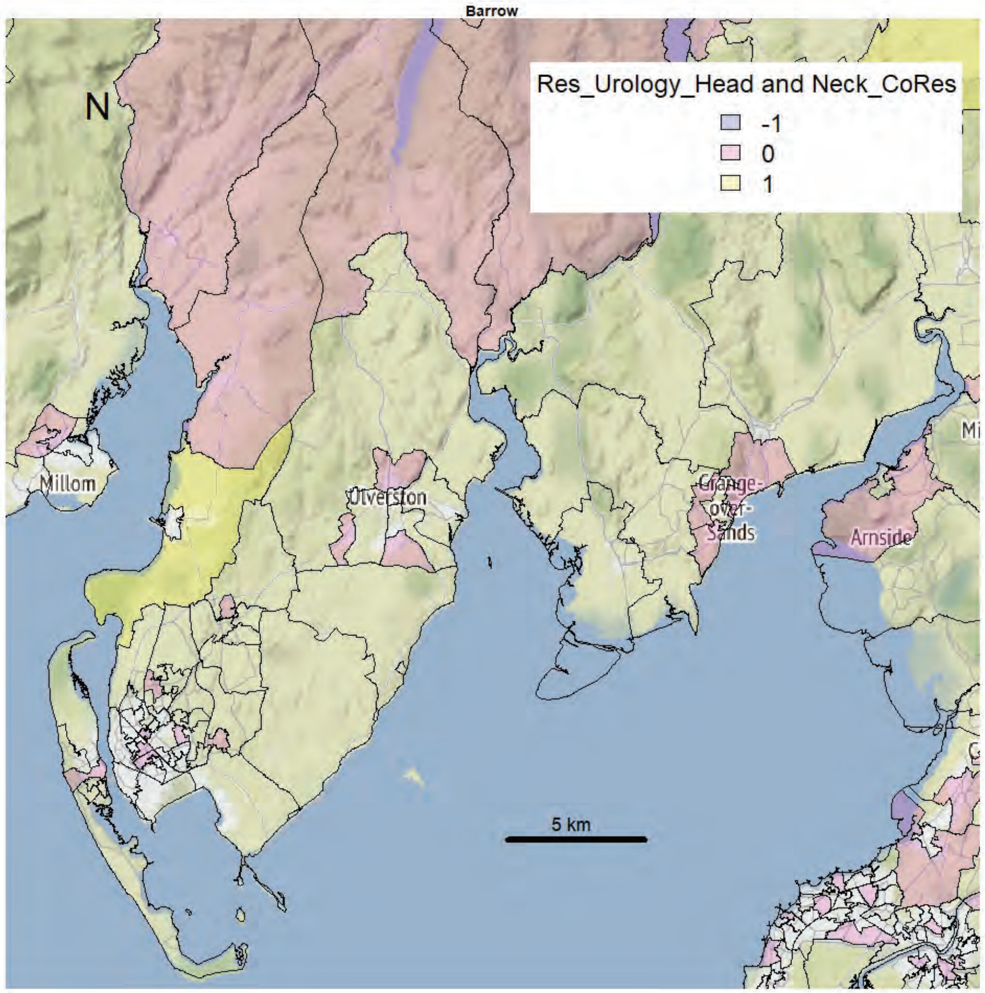
Barrow
N
Res_Urology_Head and Neck_CoRes
-1
0
1
Millom
Ulverston
Grange-over-Sands
Arnside
5 km

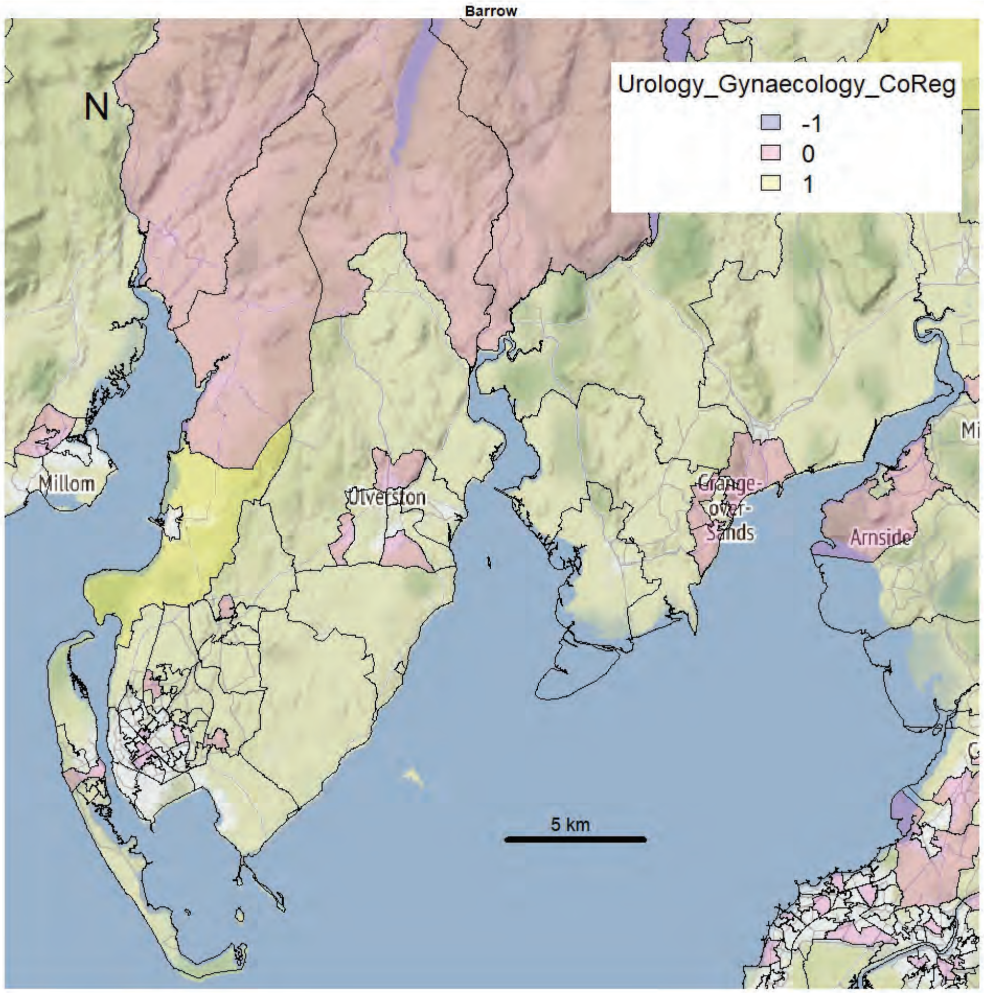
Barrow
N
Urology_Gynaecology_CoReg
-1
0
1
Millom
Ulverston
Grange-over-Sands
Arnside
5 km

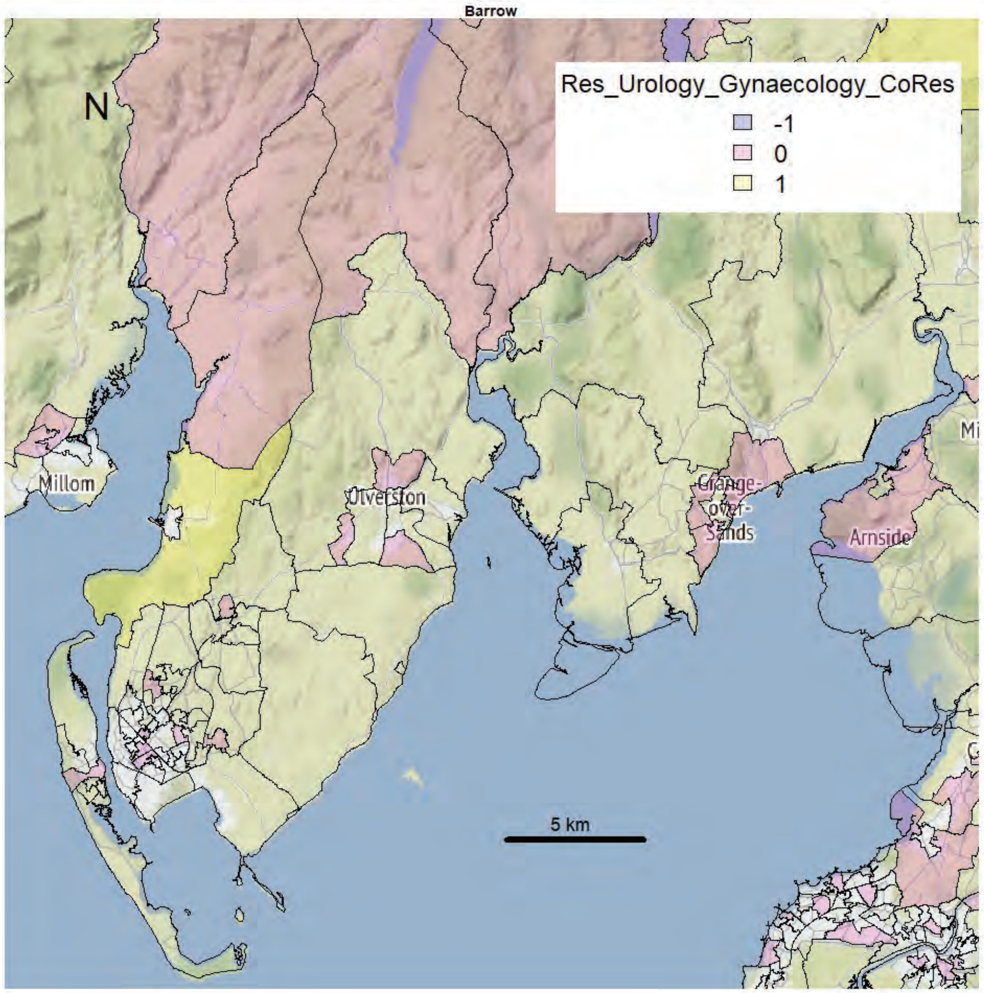
Barrow
N
Res_Urology_Gynaecology_CoRes
-1
0
1
Millom
Ulverston
Grange-over-Sands
Arnside
5 km

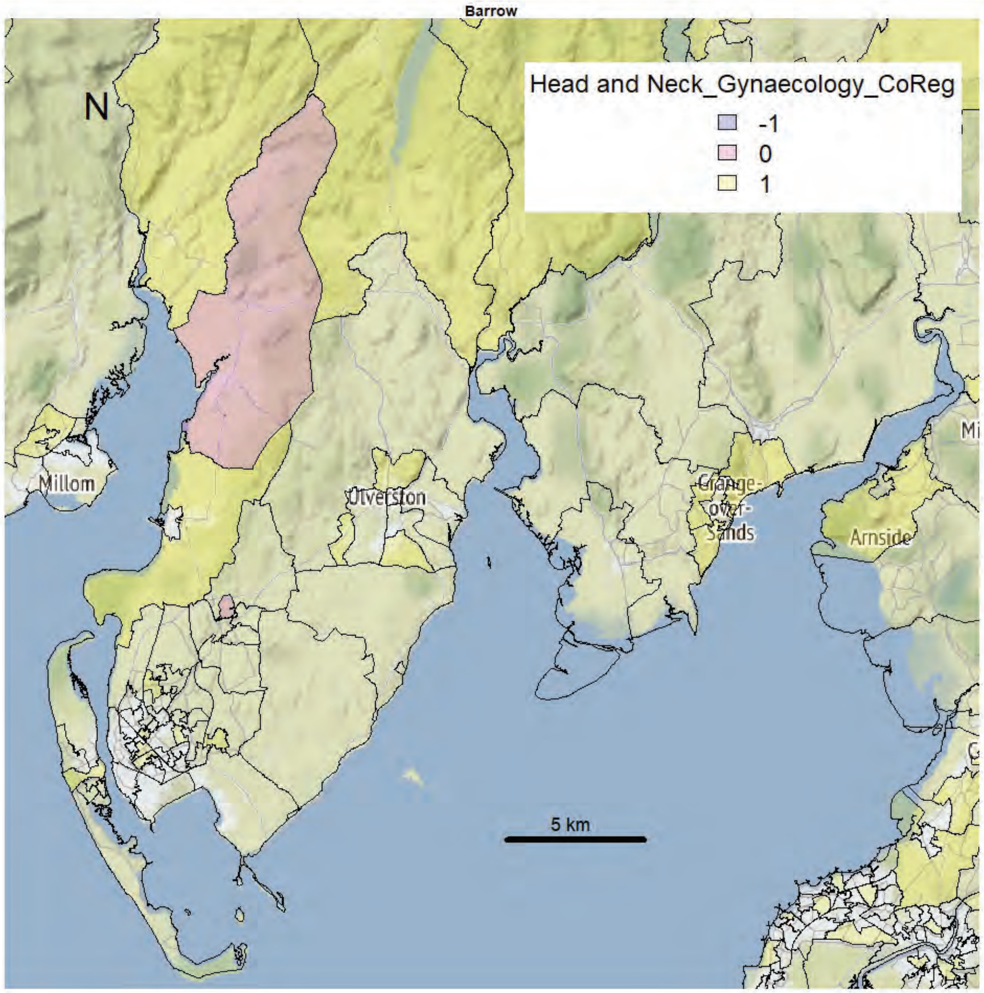
Barrow
N
Head and Neck_Gynaecology_CoReg
-1
0
1
Millom
Ulverston
Grange-over-Sands
Arnside
5 km

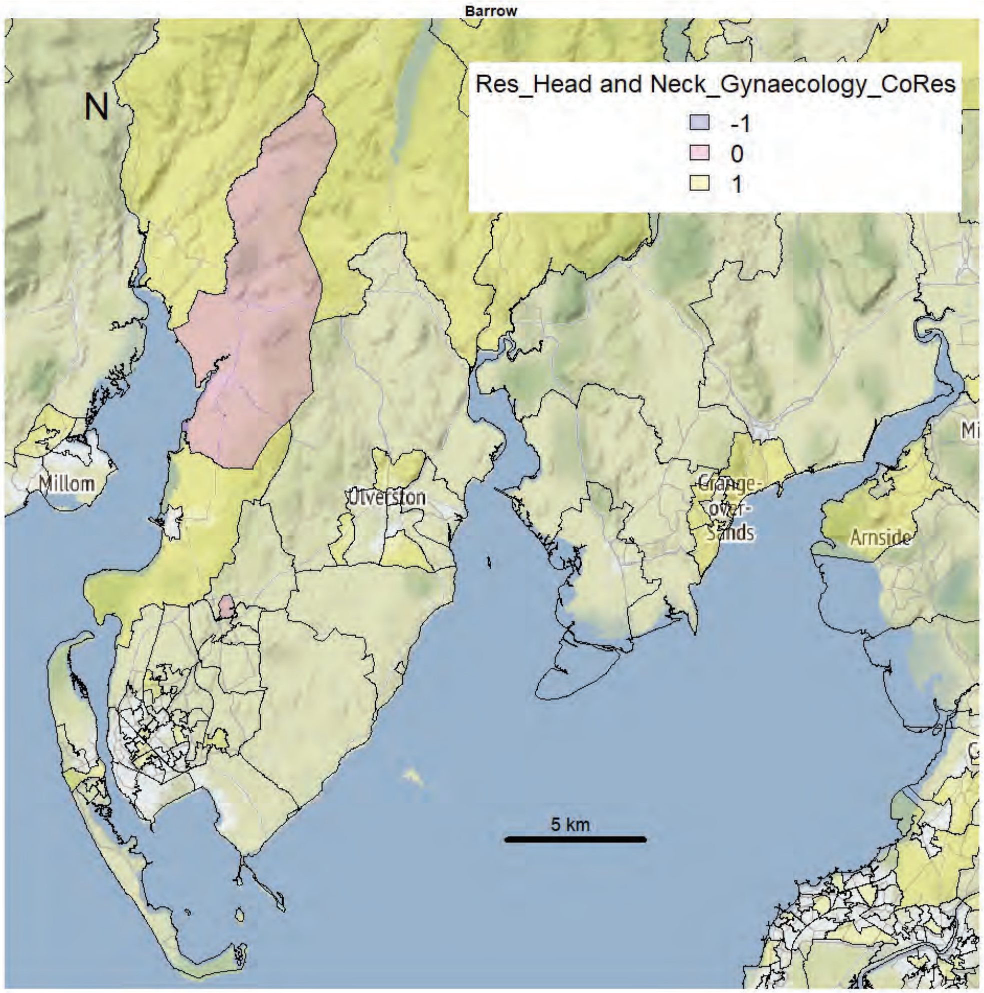
Barrow
N
Res_Head and Neck_Gynaecology_CoRes
-1
0
1
Millom
Ulverston
Grange-over-Sands
Arnside
5 km

Kendal area

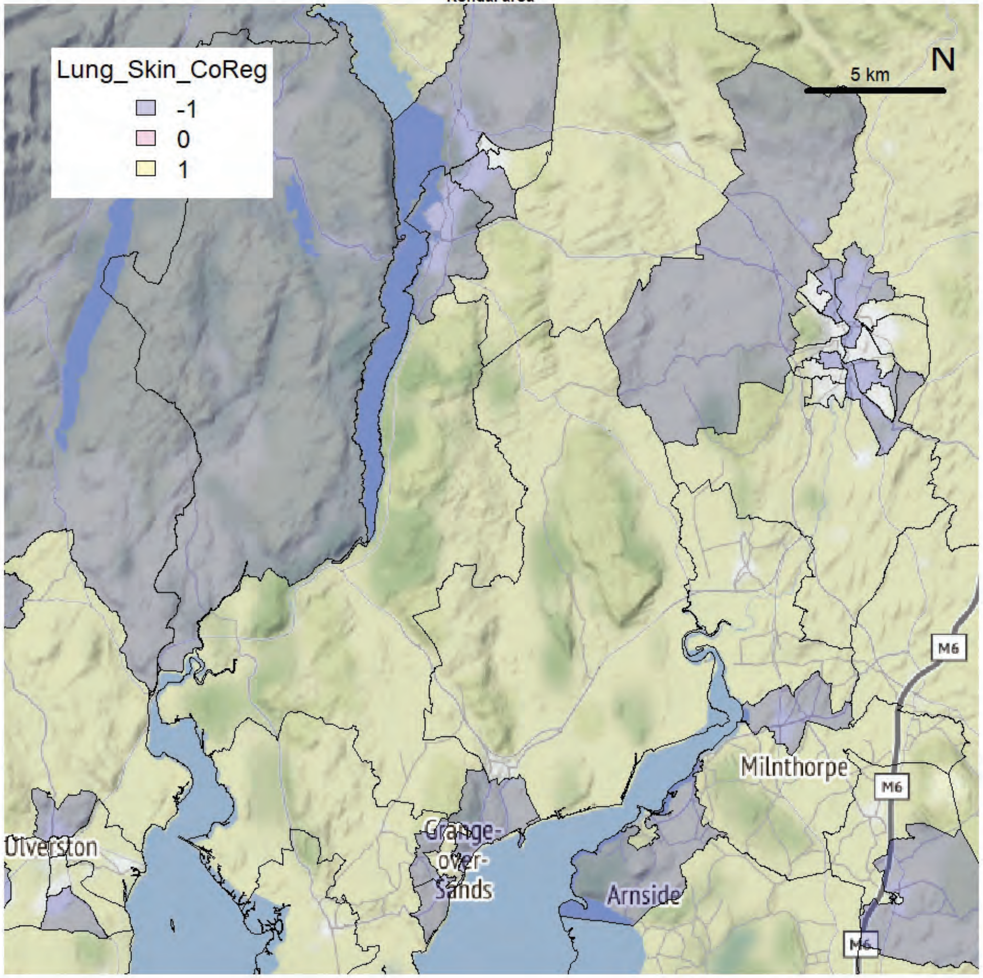


Kendal area

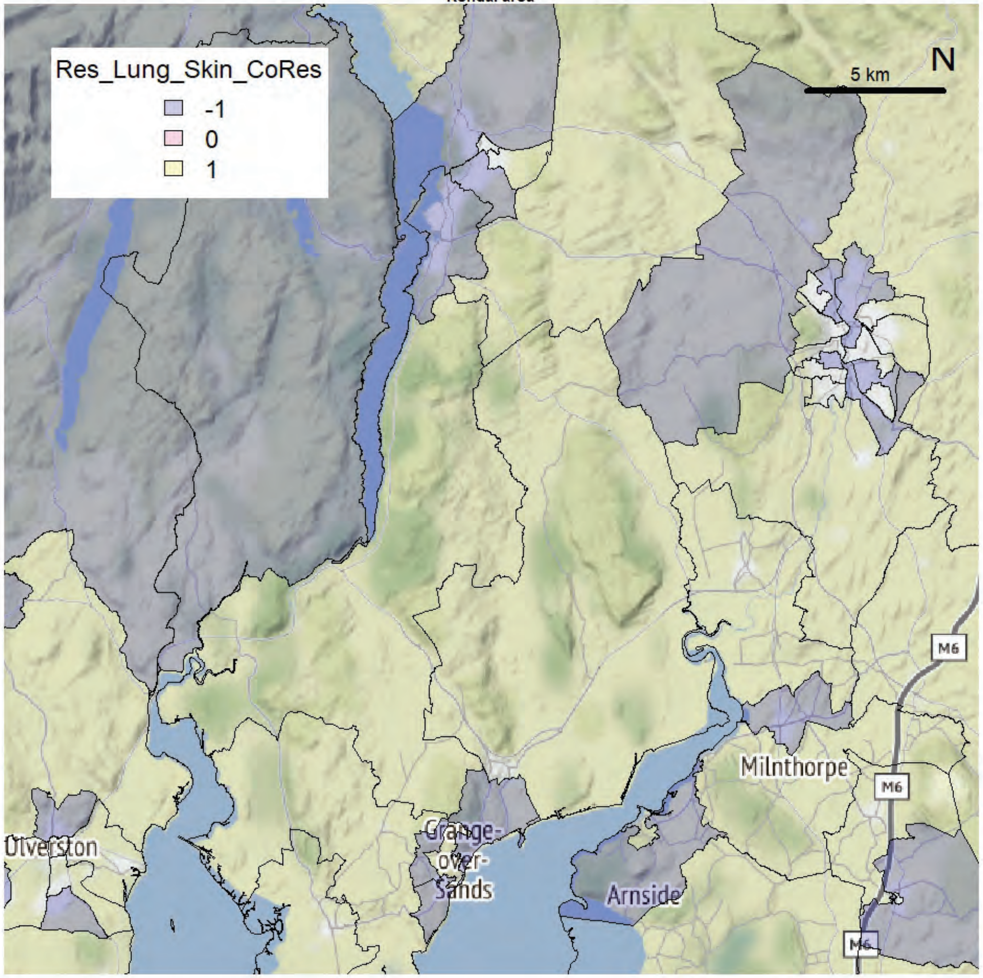

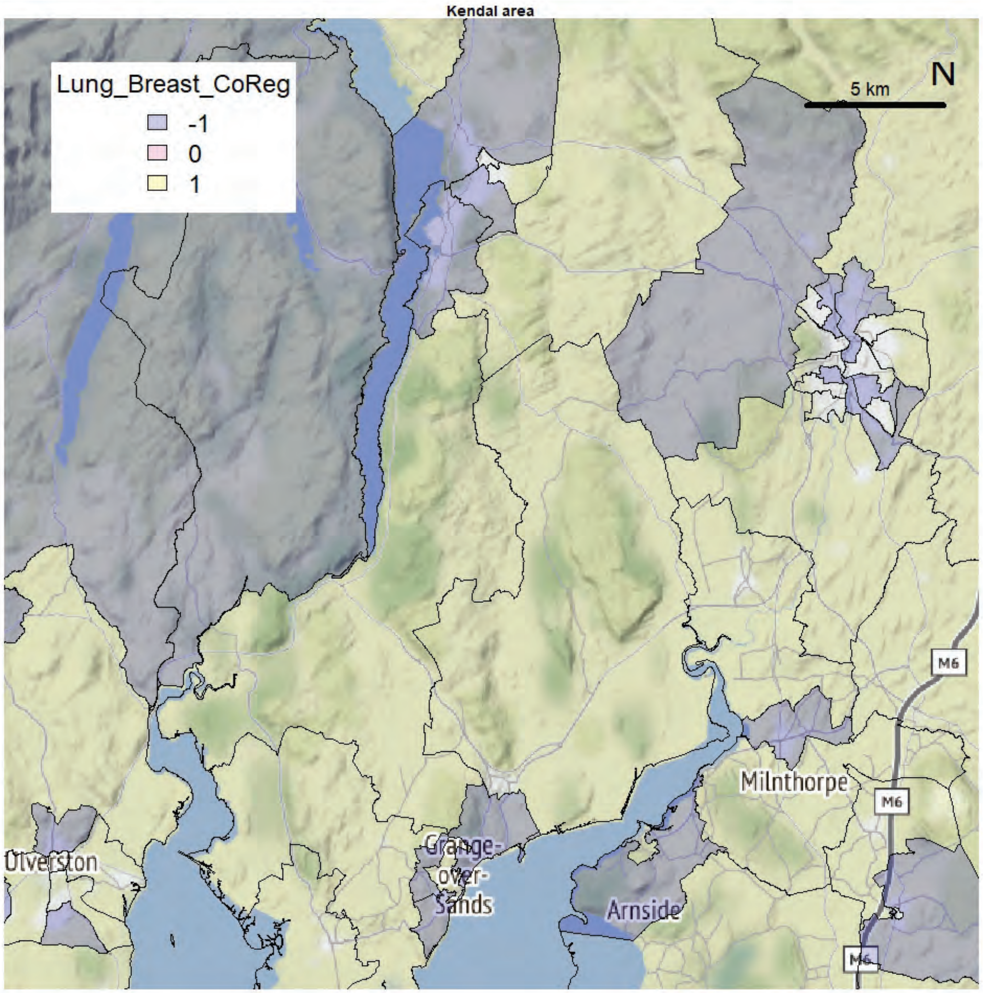
Kendal area
Lung_Breast_CoReg
-1
0
1
5 km
N
Ulverston
Grange-over-Sands
Arnside
Milnthorpe
M6

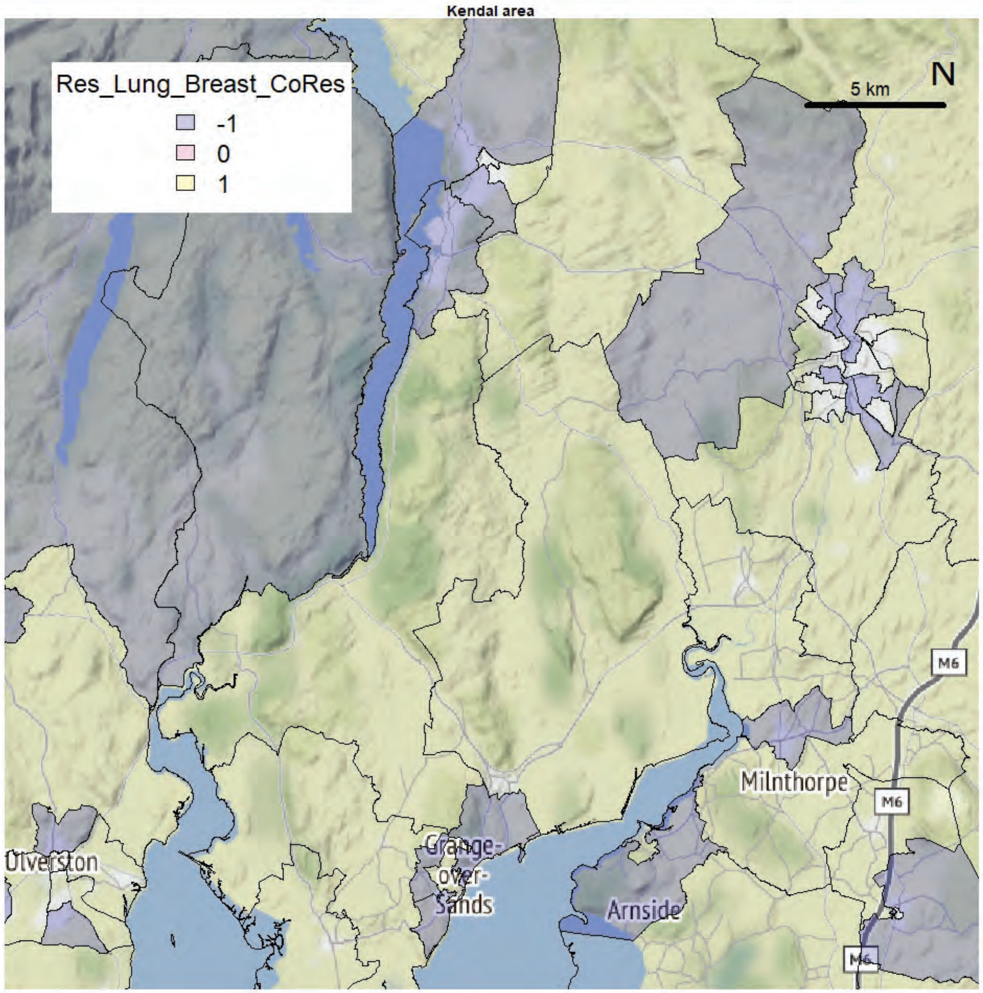
Kendal area
Res_Lung_Breast_CoRes
-1
0
1
5 km
N
Ulverston
Grange-over-Sands
Arnside
Milnthorpe
M6

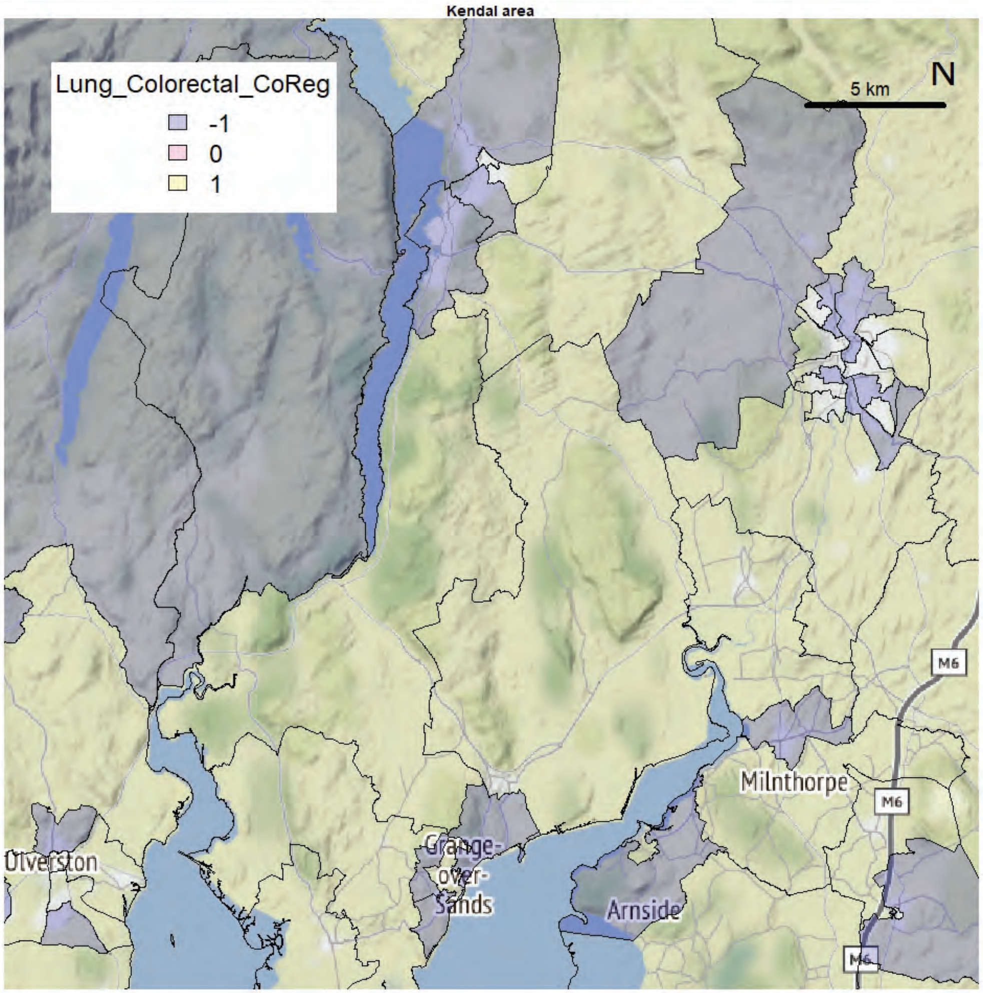
Kendal area
Lung_Colorectal_CoReg
-1
0
1
5 km
N
Ulverston
Grange-over-Sands
Arnside
Milnthorpe
M6

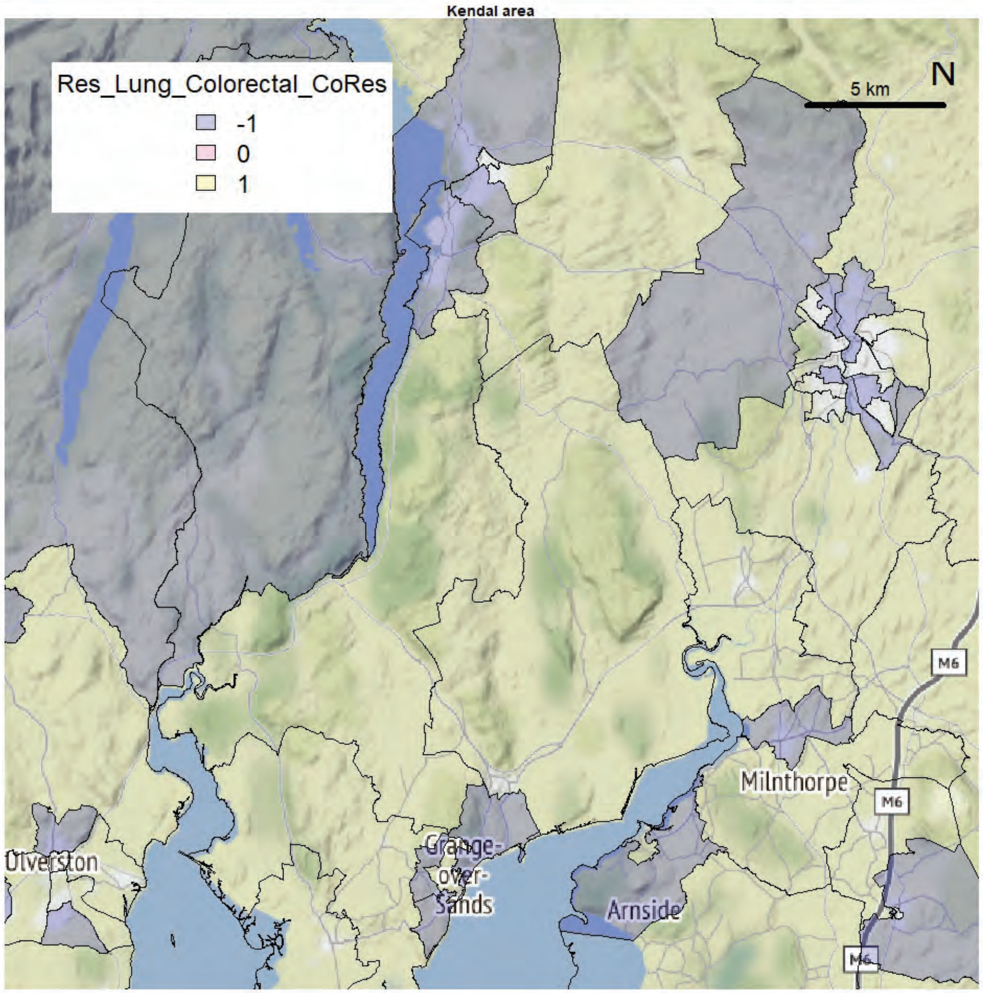
Kendal area
Res_Lung_Colorectal_CoRes
-1
0
1
5 km
N
Ulverston
Grange-over-Sands
Arnside
Milnthorpe
M6

Kendal area

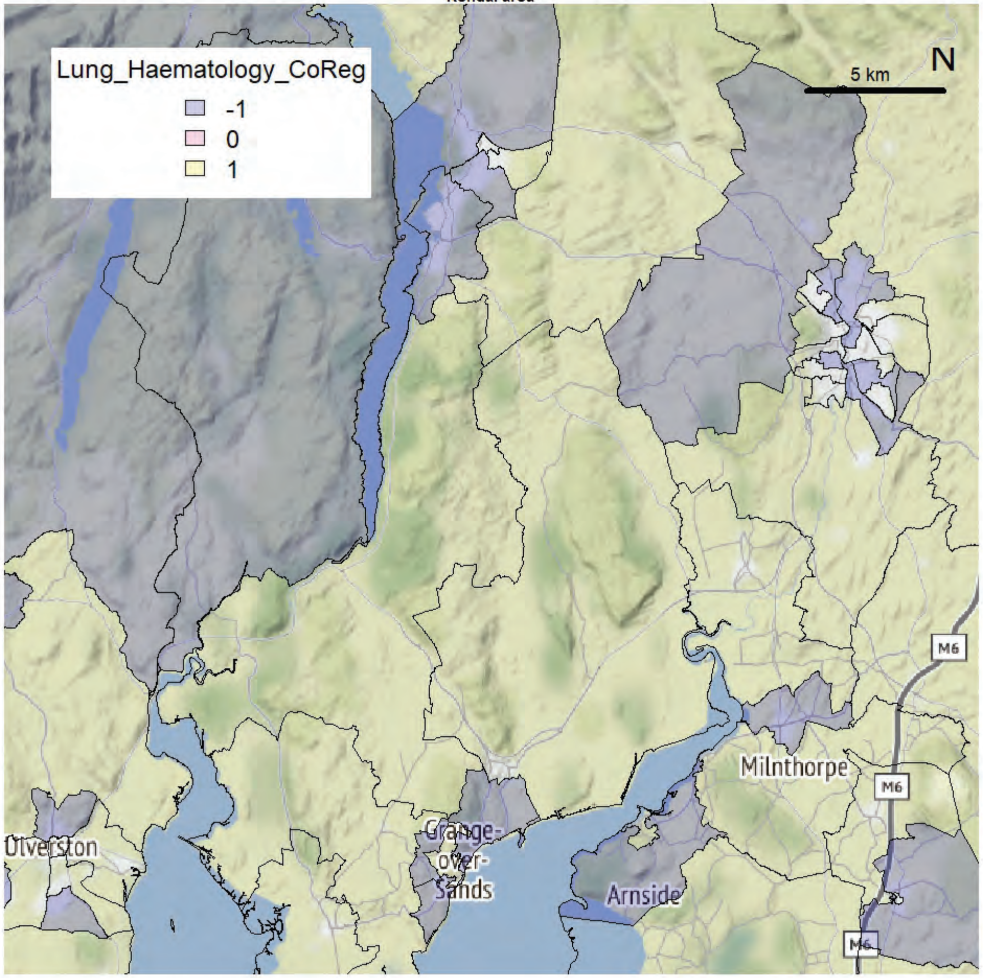


Kendal area

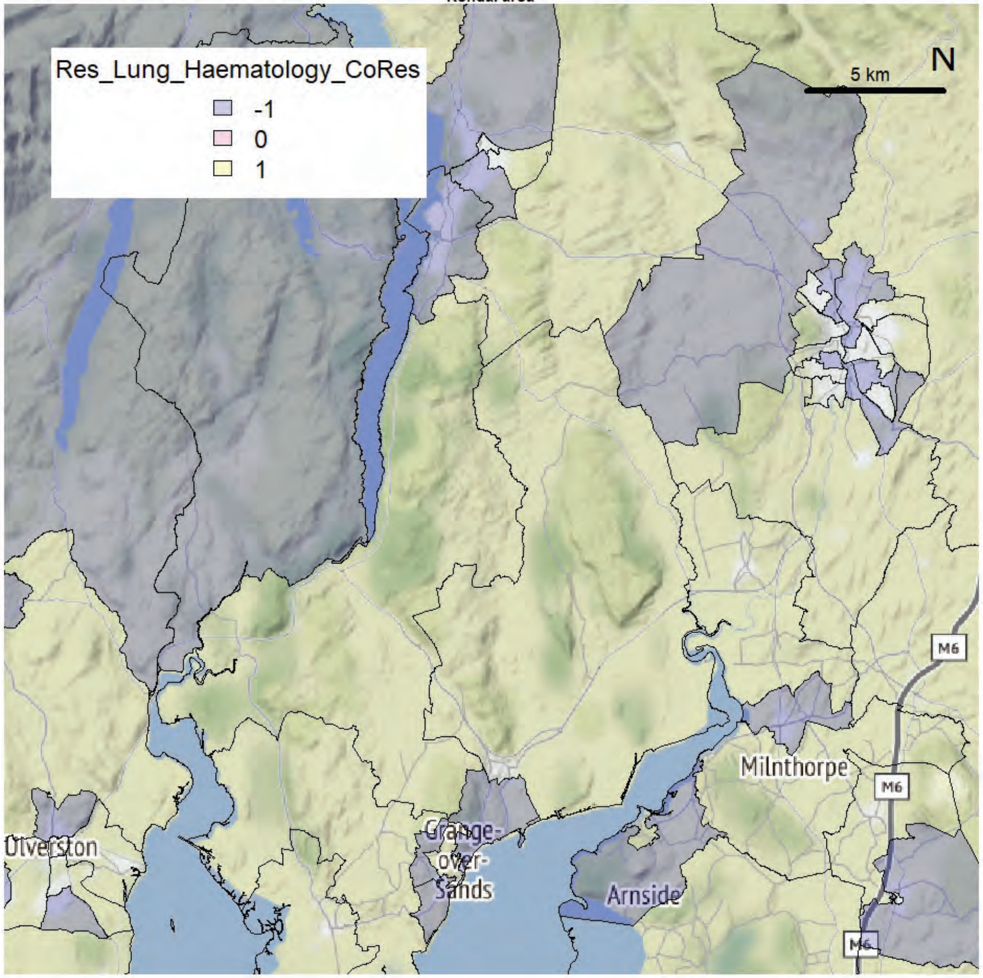

Kendal area

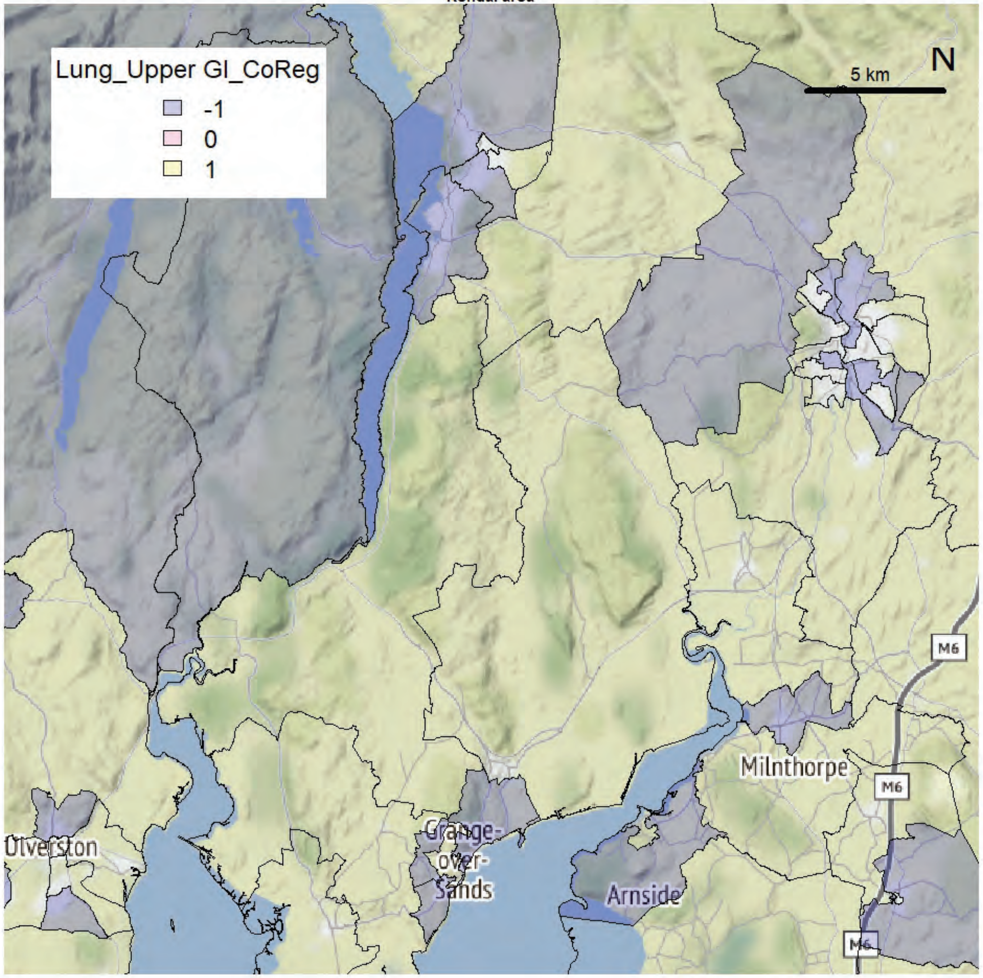


Kendal area

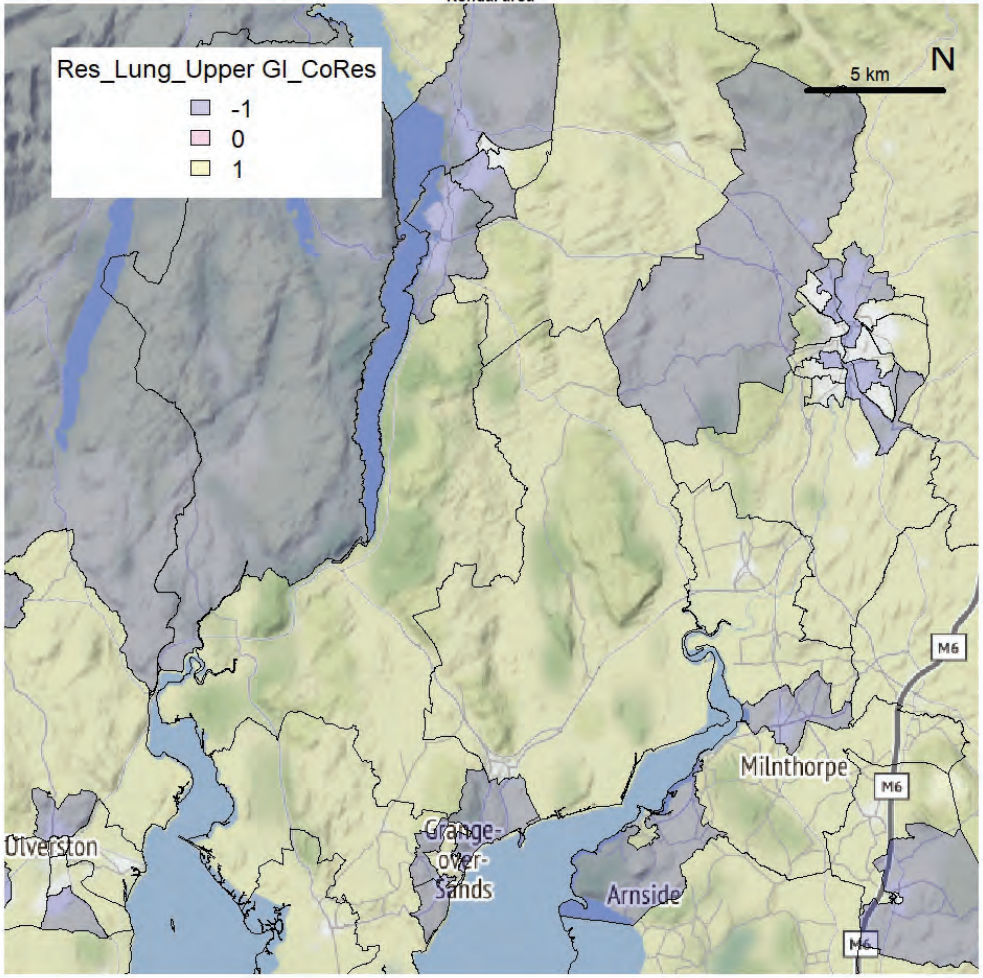

Kendal area

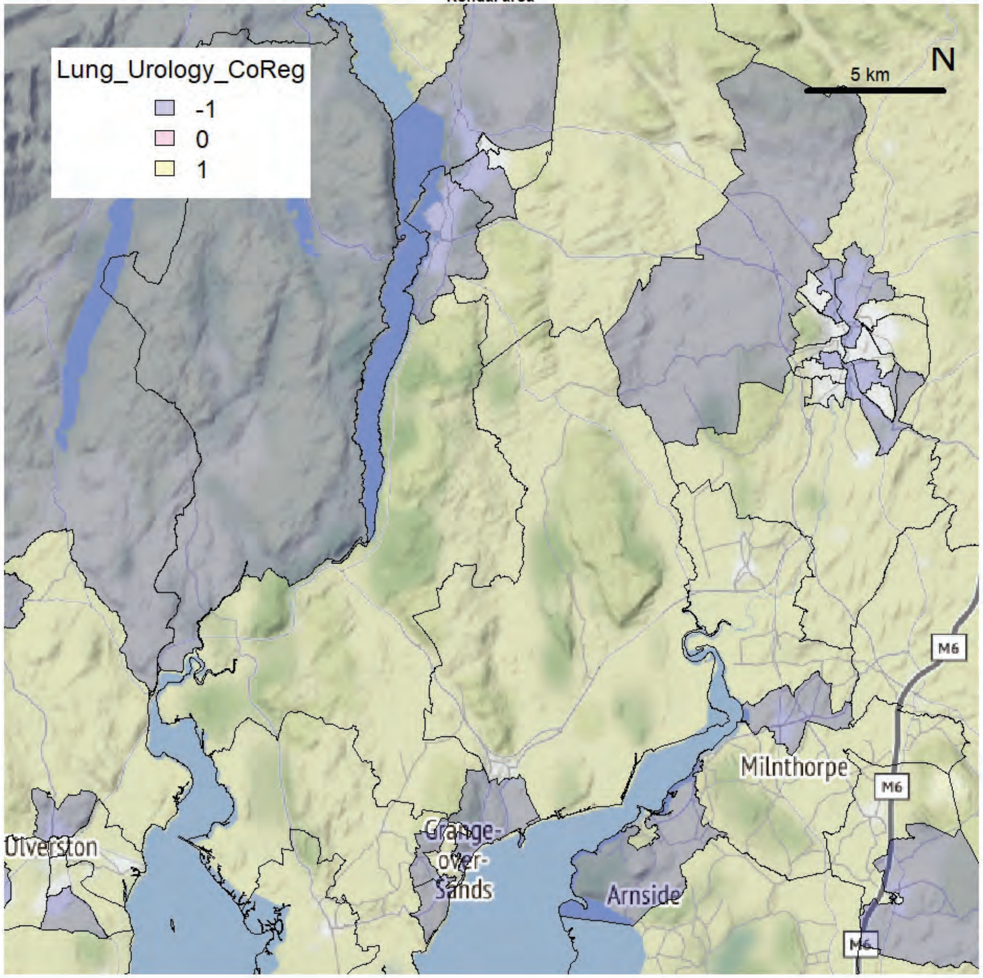


Kendal area

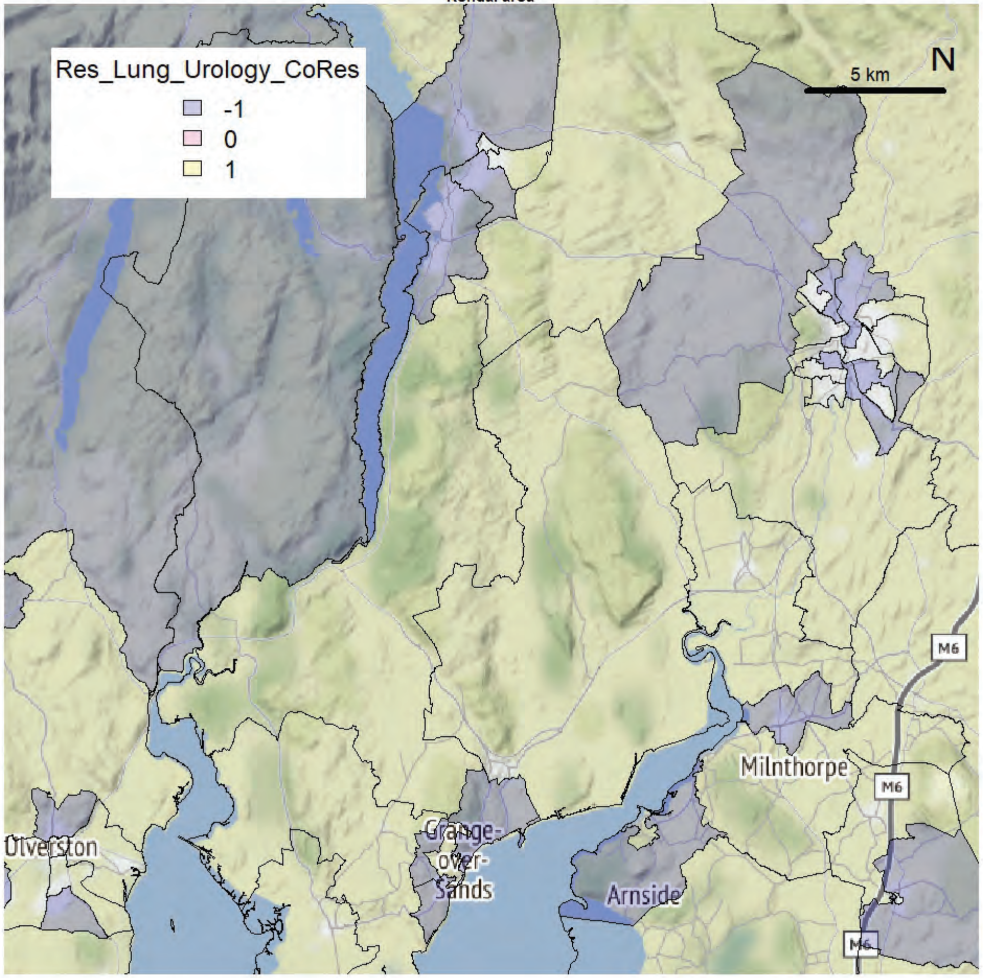

Kendal area

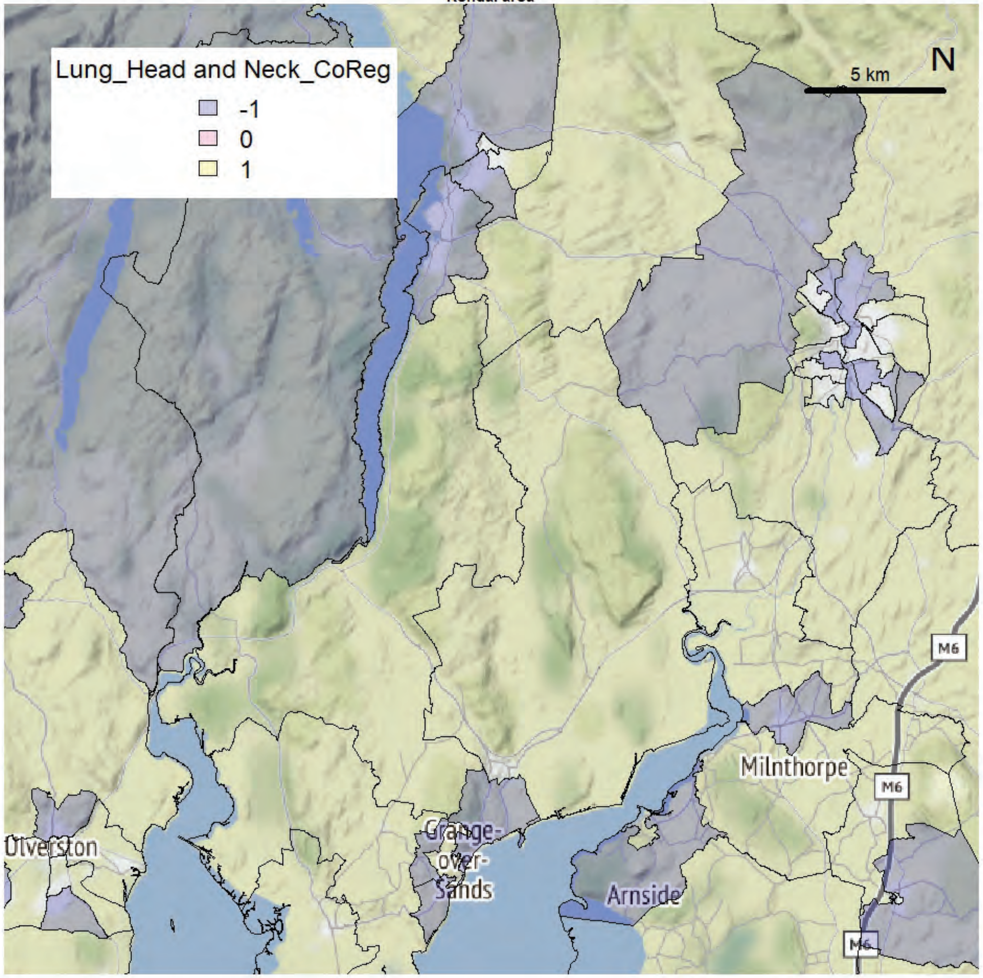


Kendal area

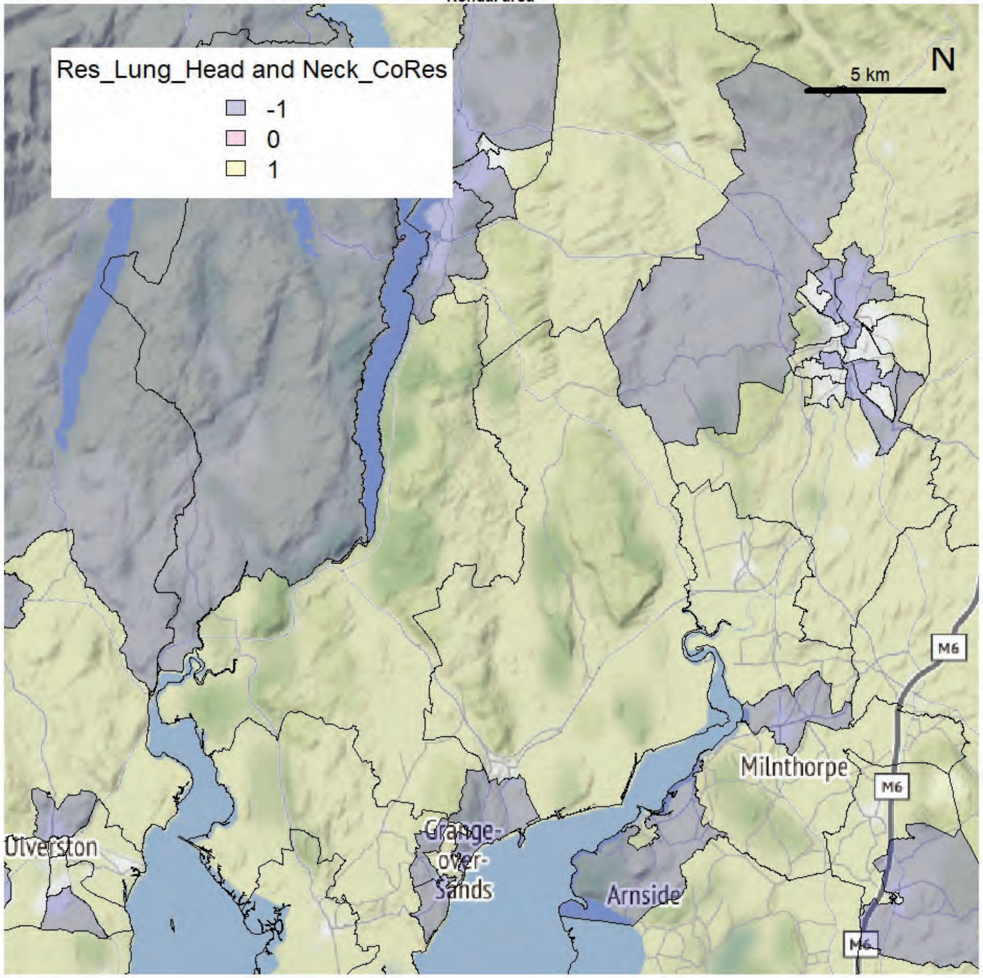

Kendal area

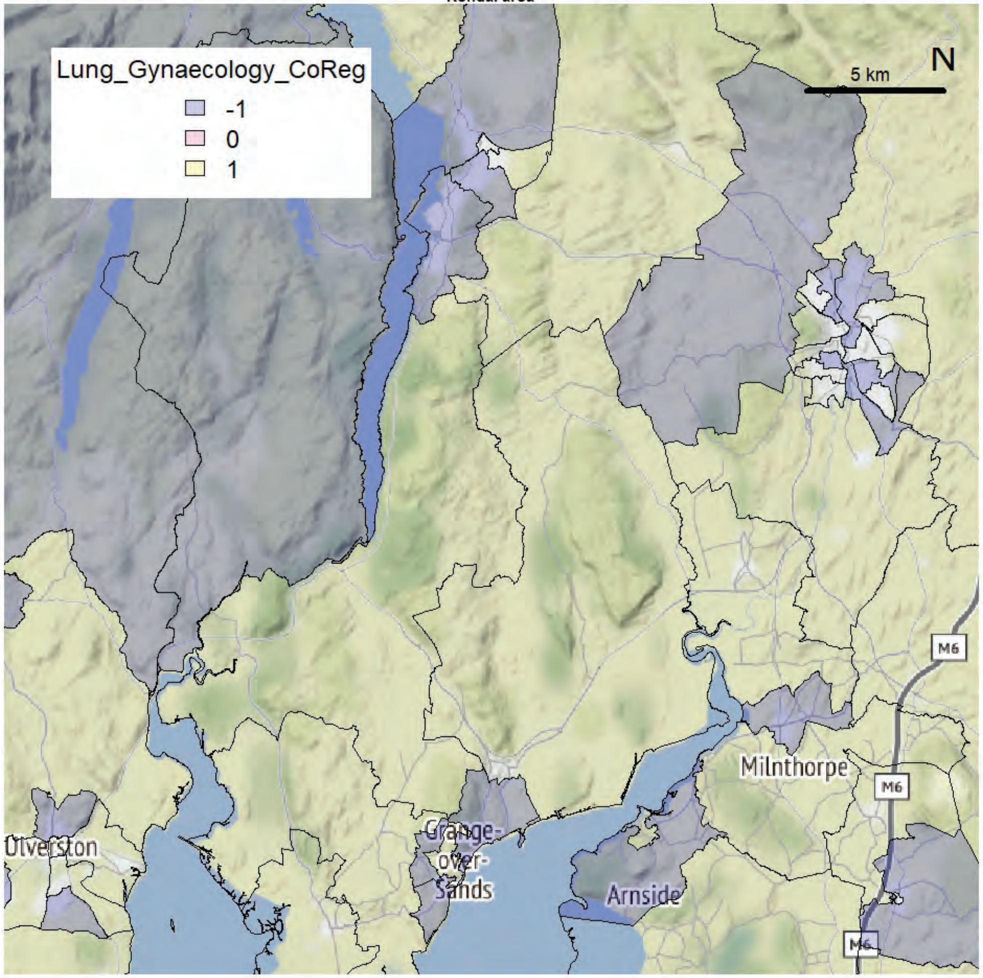


Kendal area

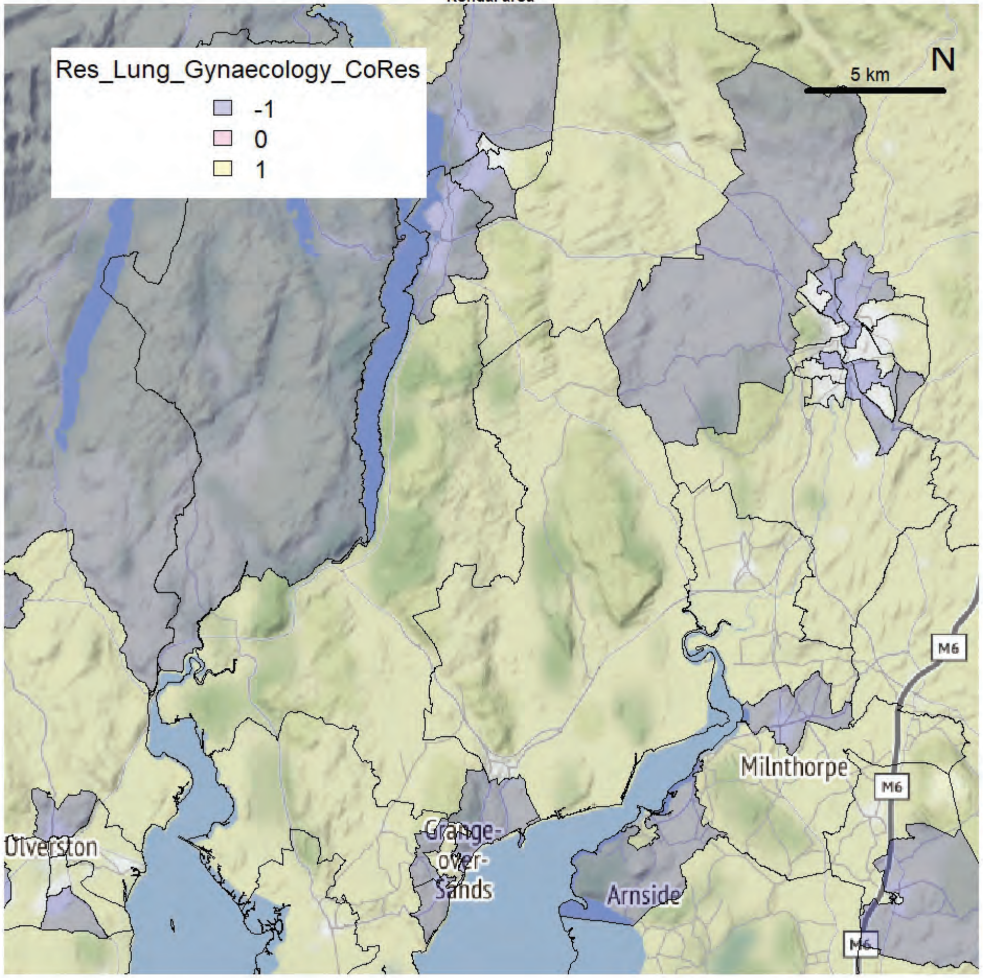

Kendal area

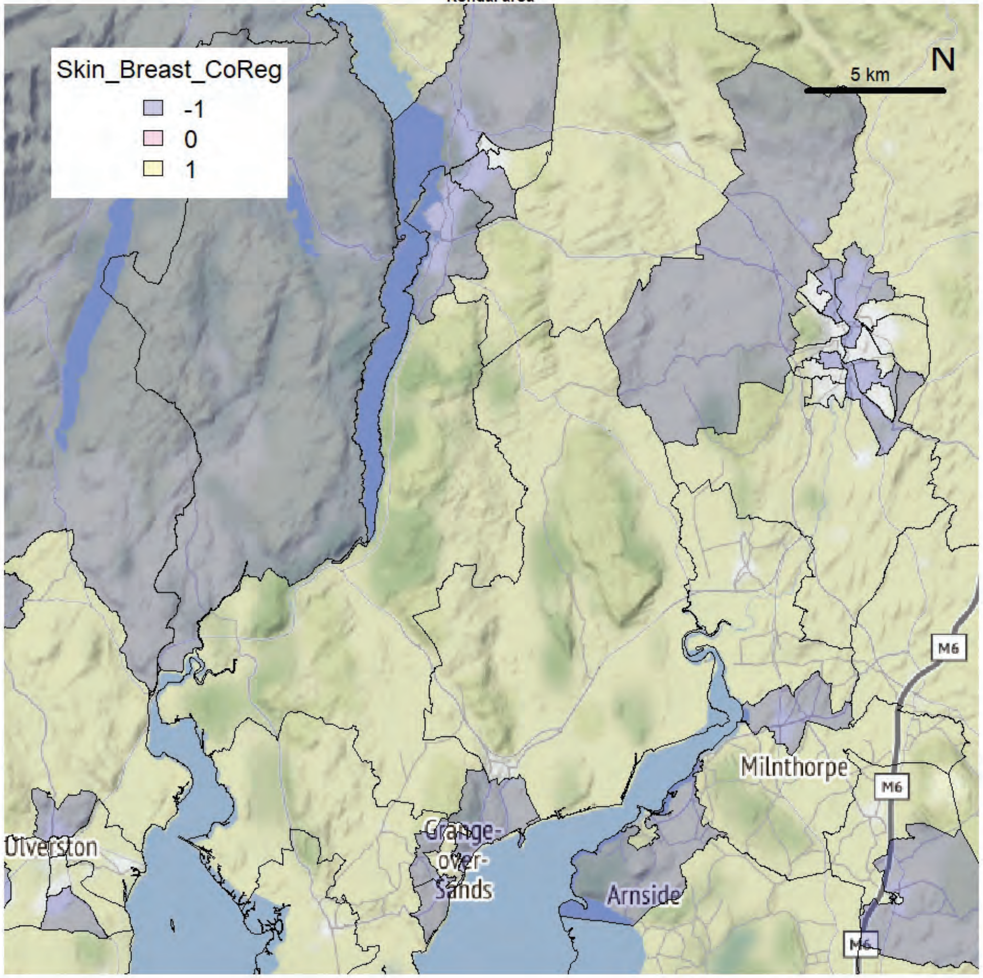


Kendal area

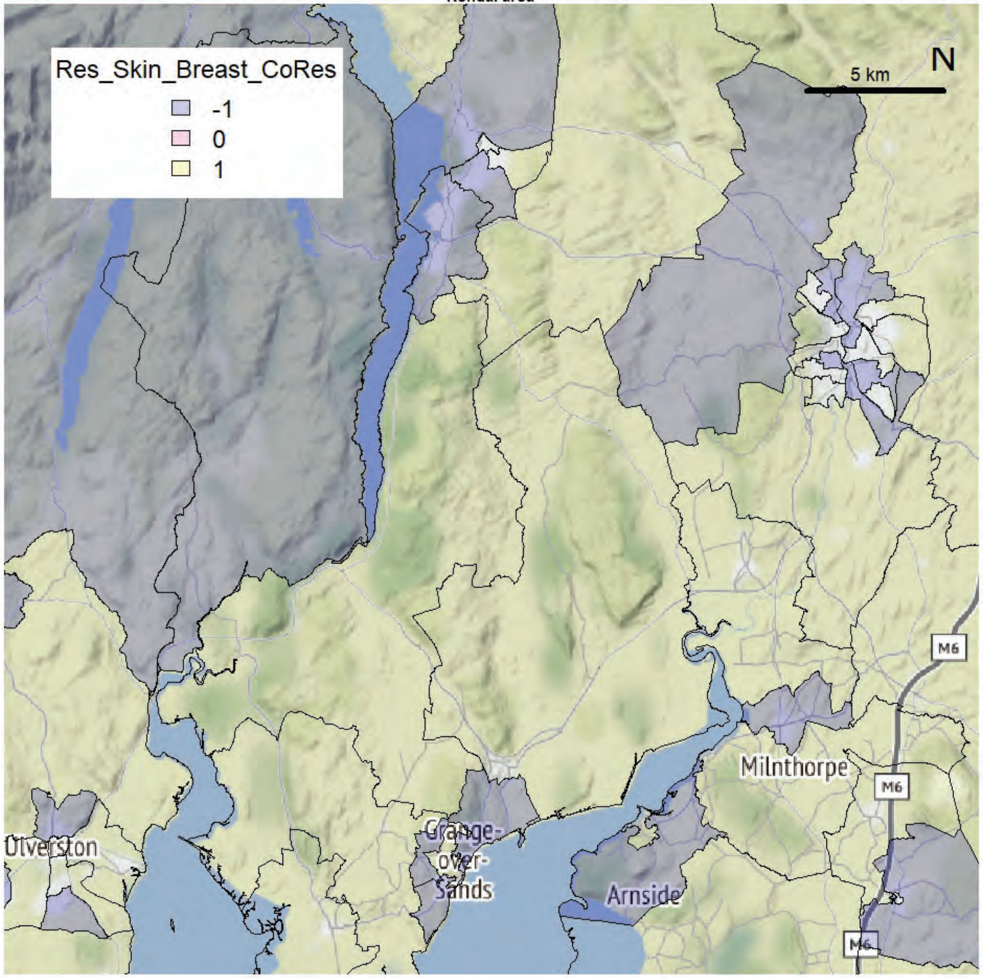

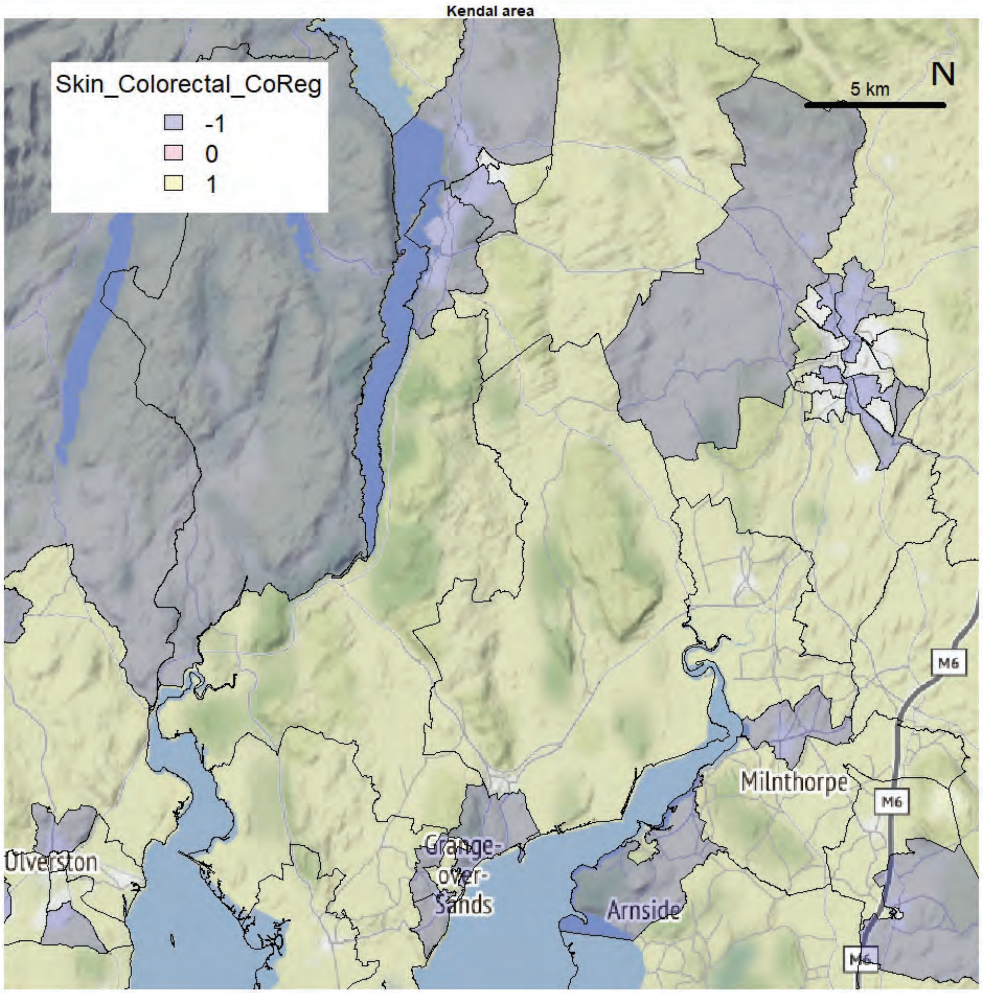
Kendal area
Skin_Colorectal_CoReg
-1
0
1
5 km
N
Ulverston
Grange-
over-
Sands
Arnside
Milnthorpe
M6

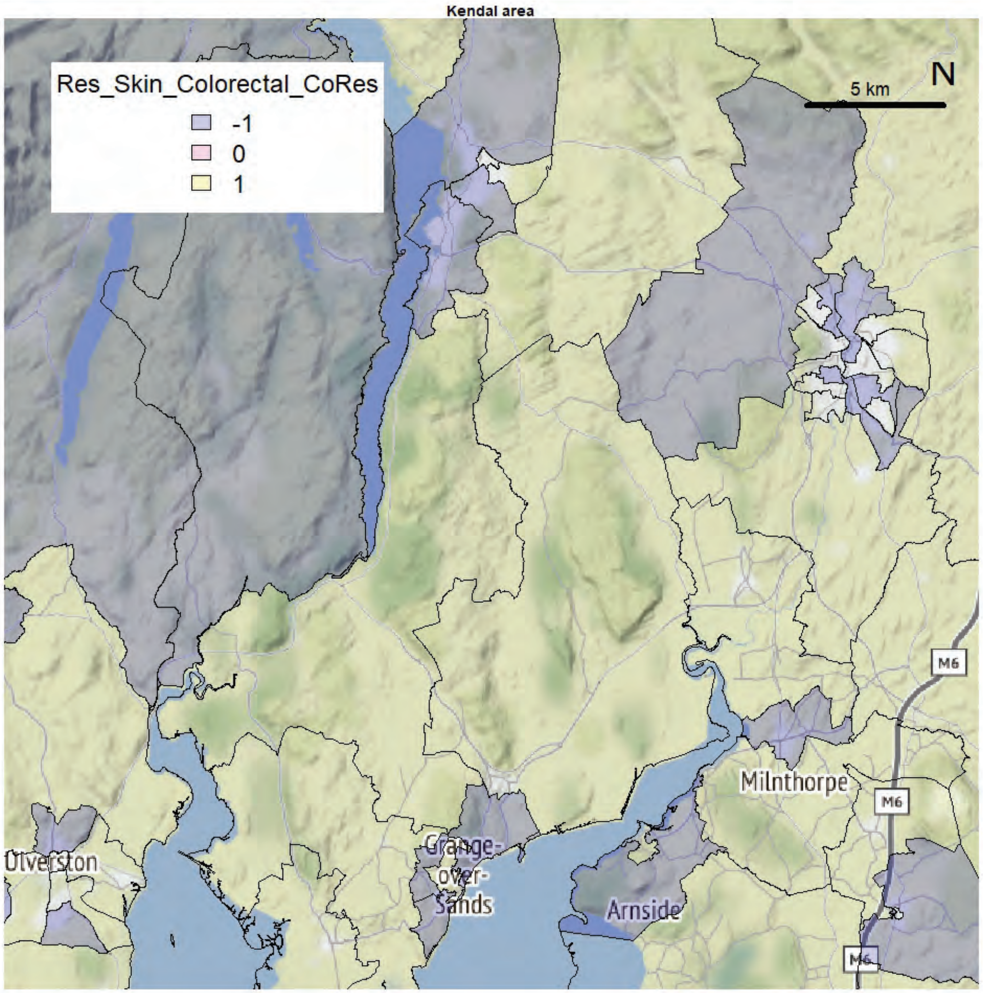
Kendal area
Res_Skin_Colorectal_CoRes
-1
0
1
5 km
N
Ulverston
Grange-
over-
Sands
Arnside
Milnthorpe
M6

Kendal area

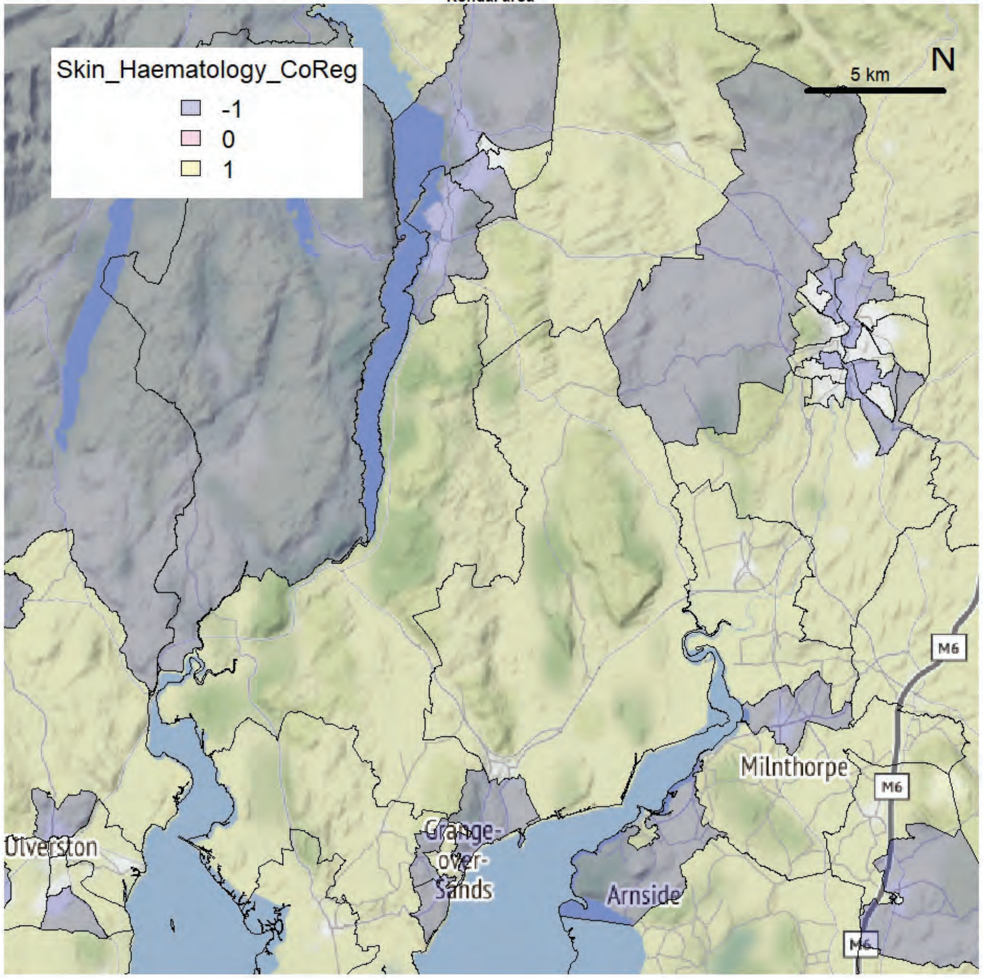


Kendal area

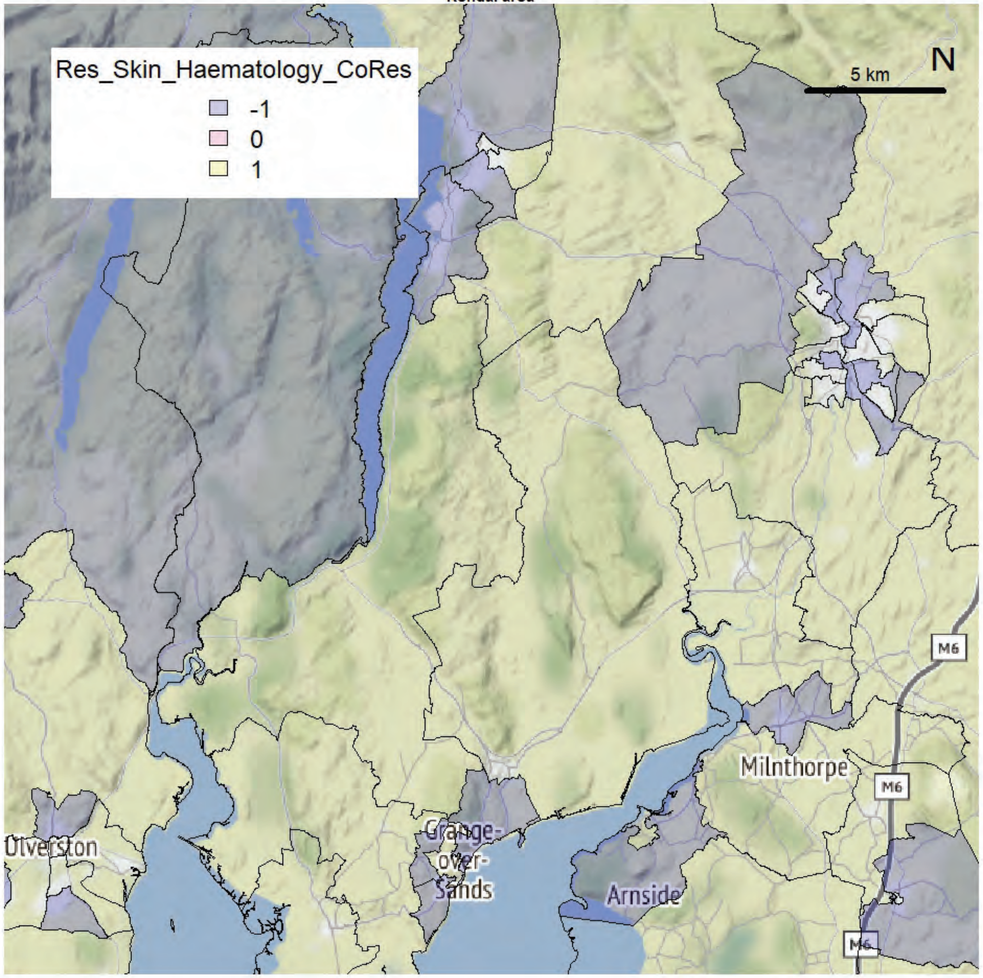

Kendal area

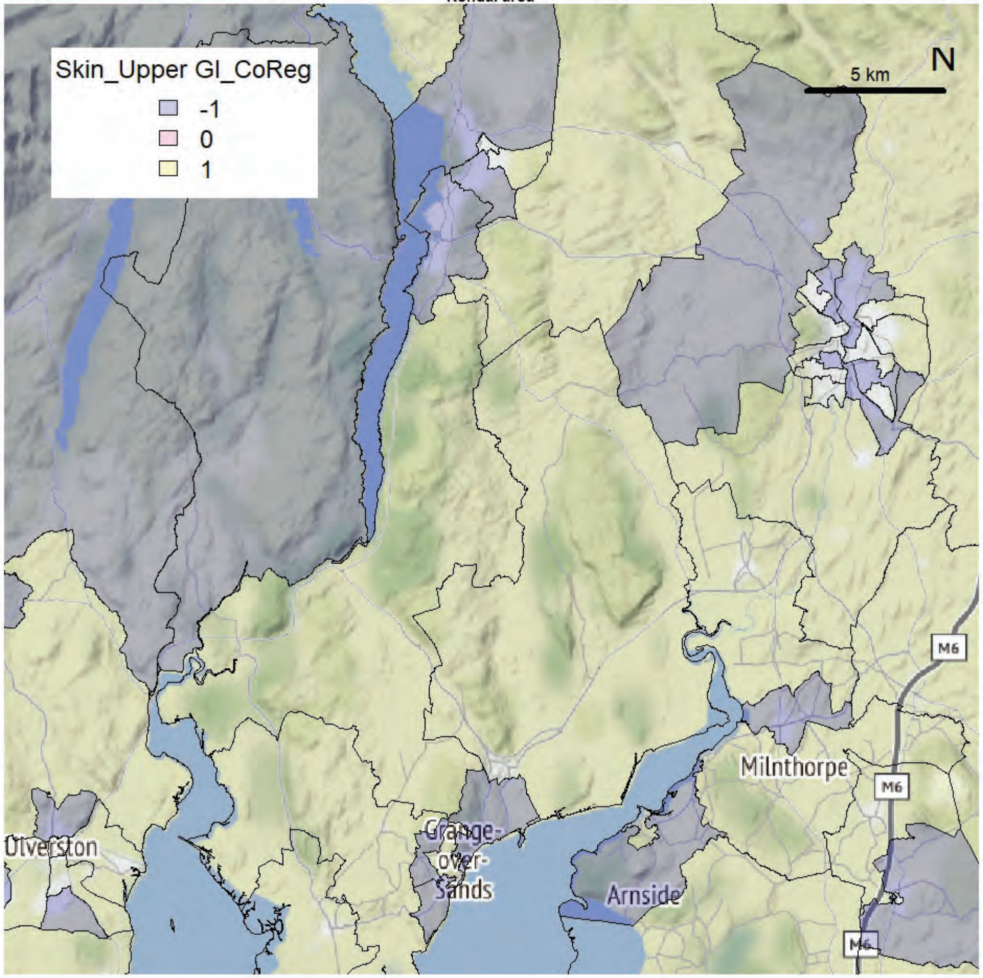


Kendal area

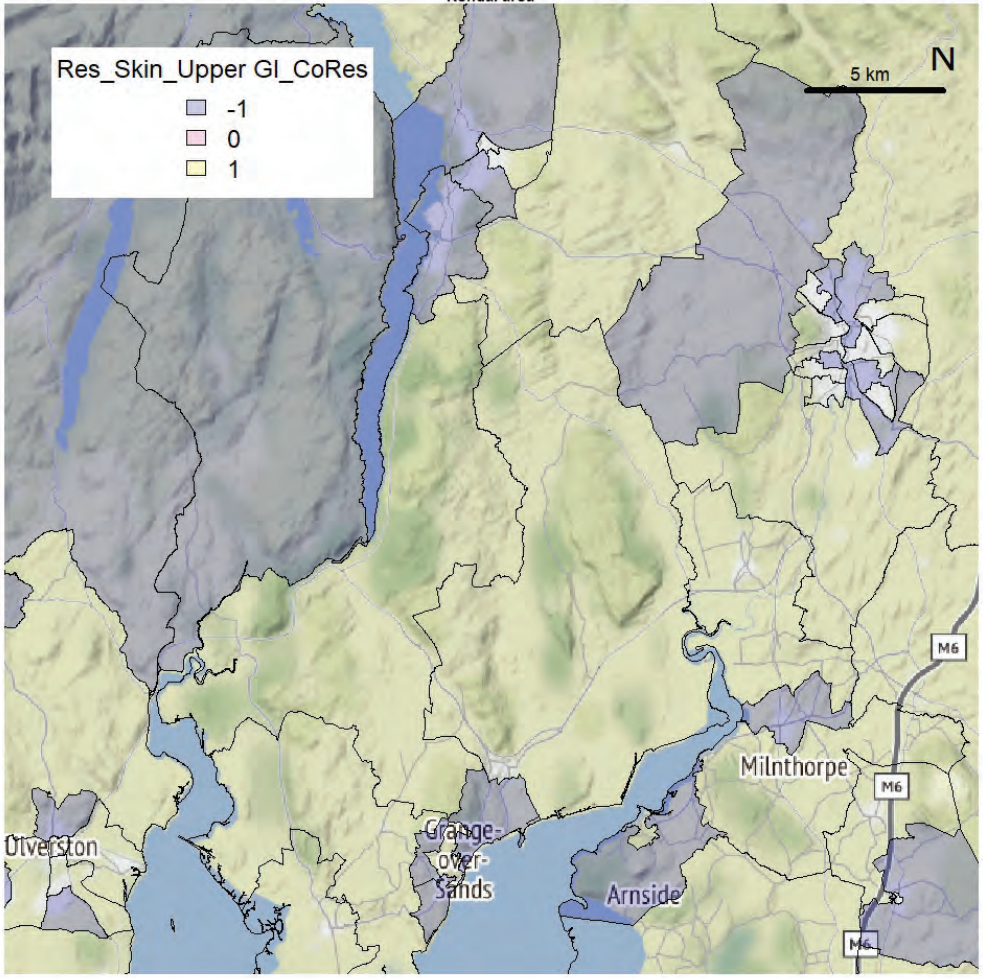

Kendal area

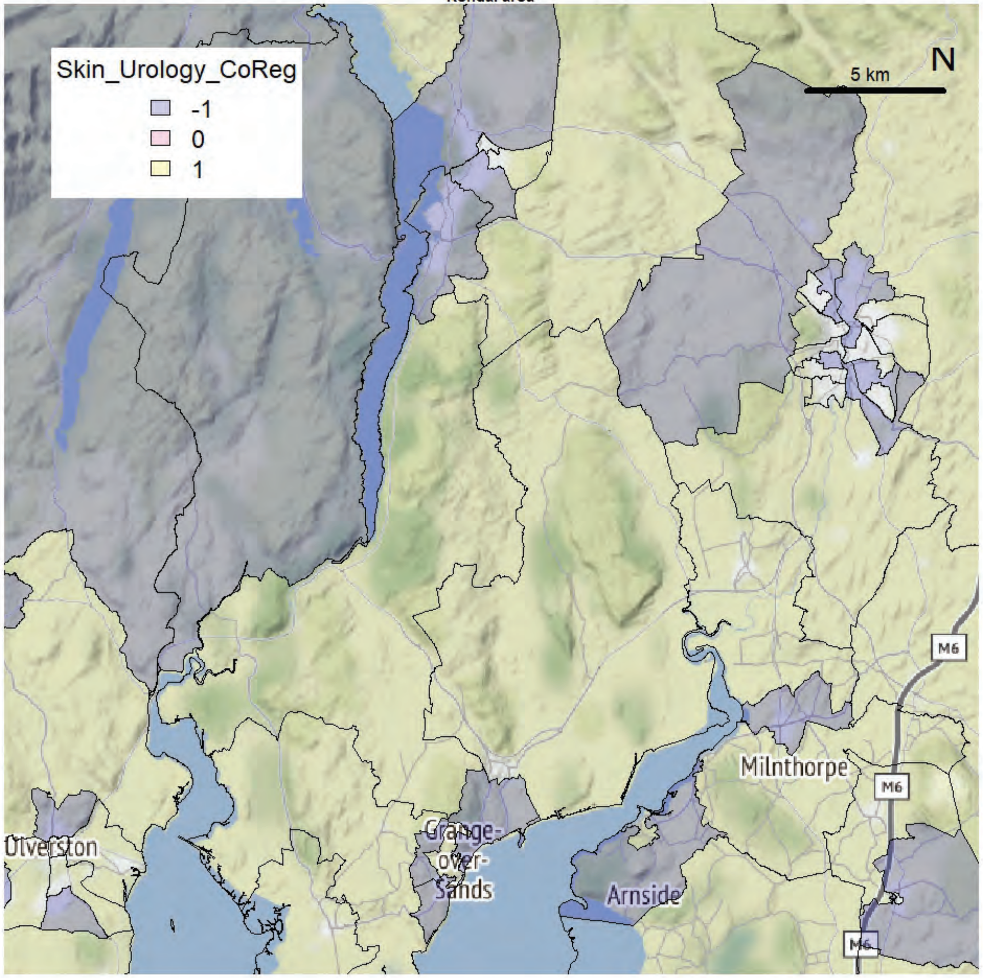


Kendal area

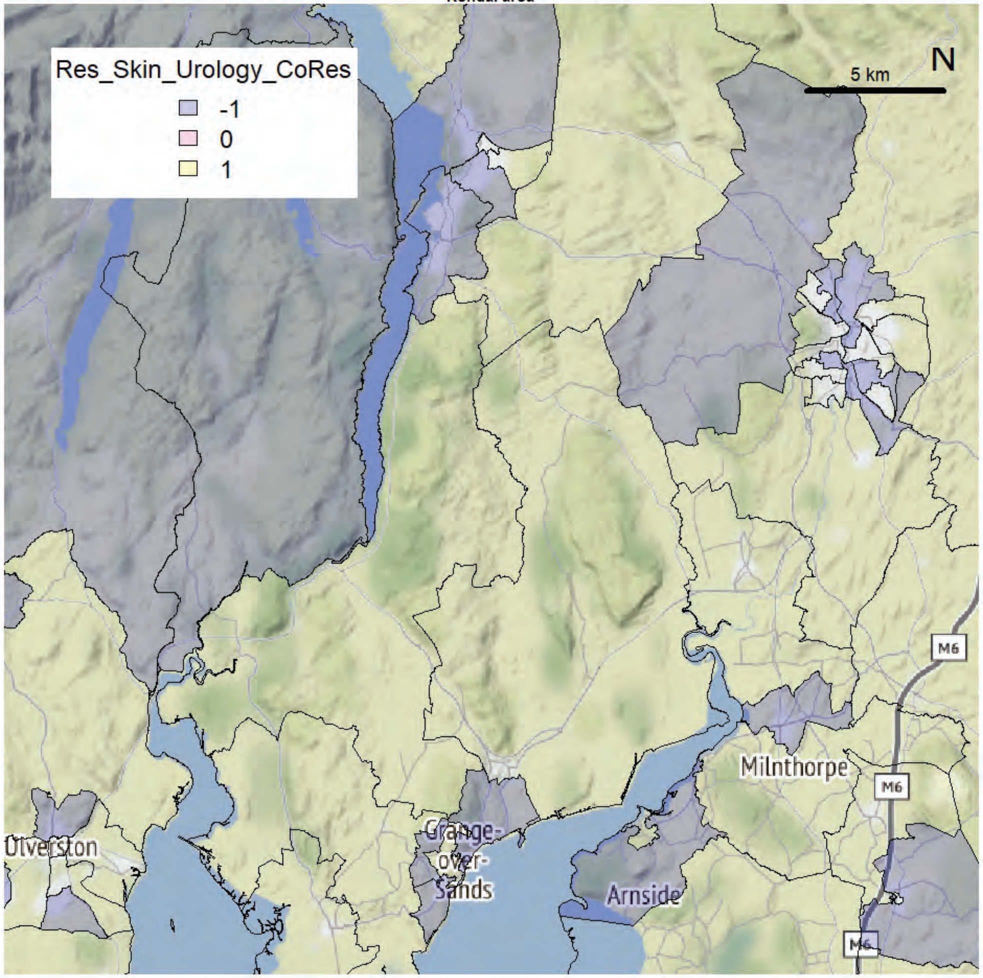

Kendal area

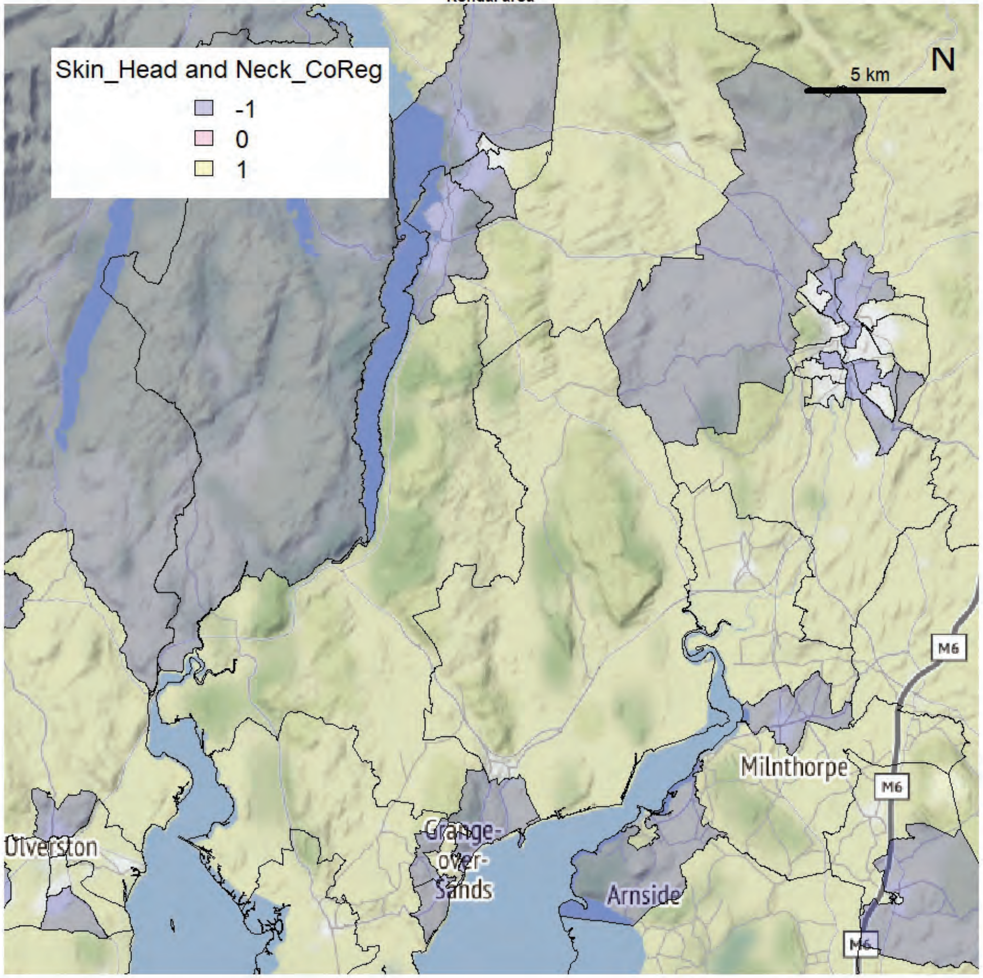


Kendal area

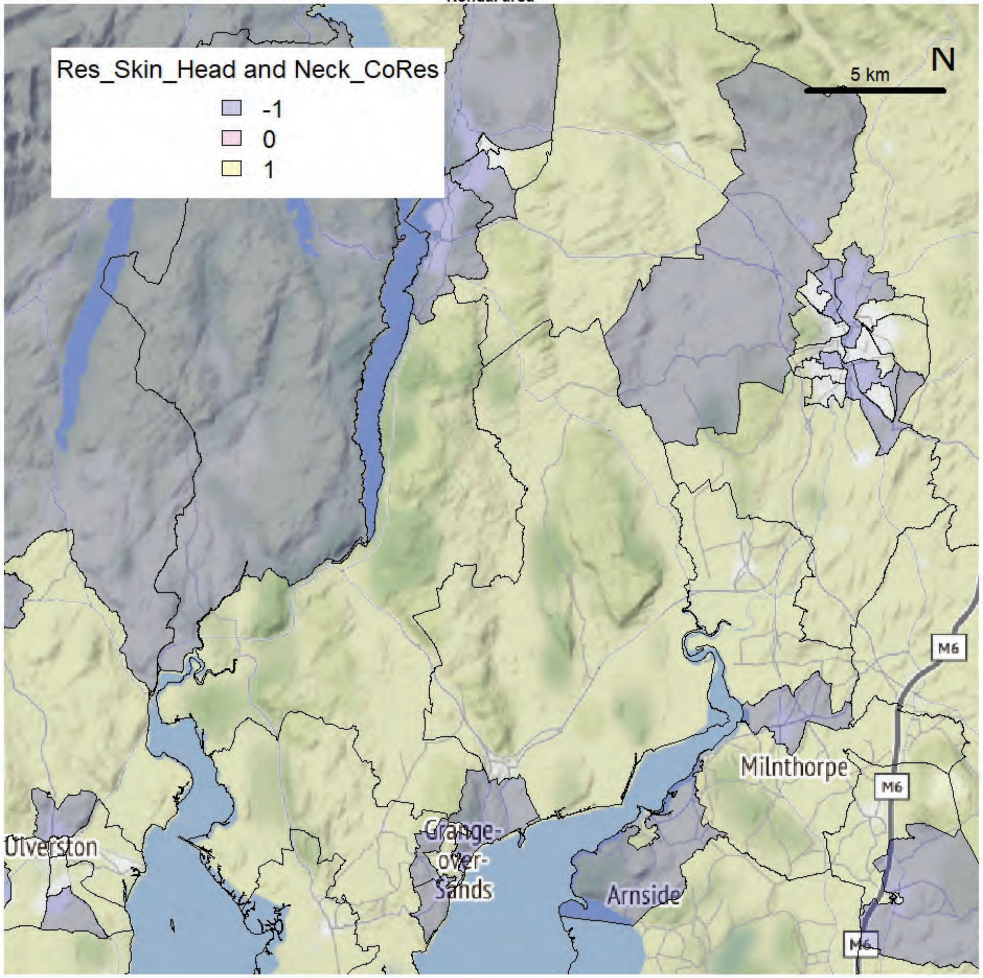

Kendal area

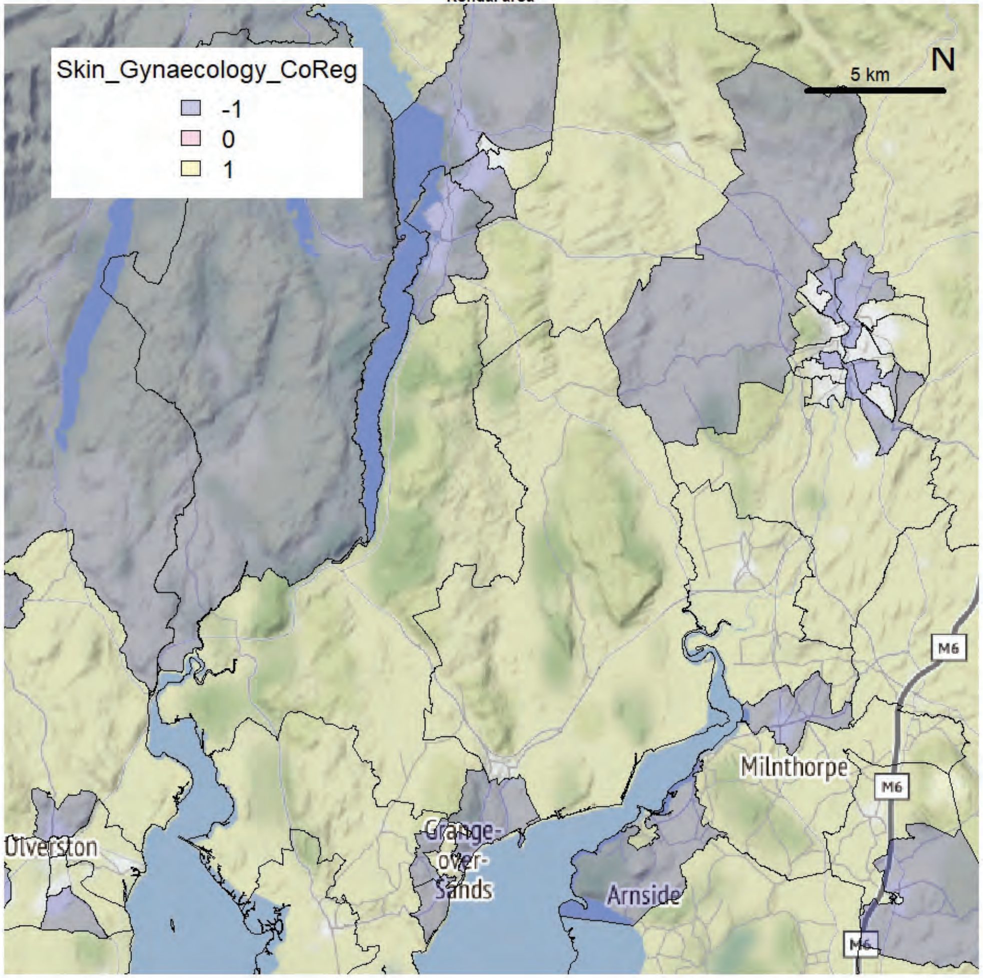


Kendal area

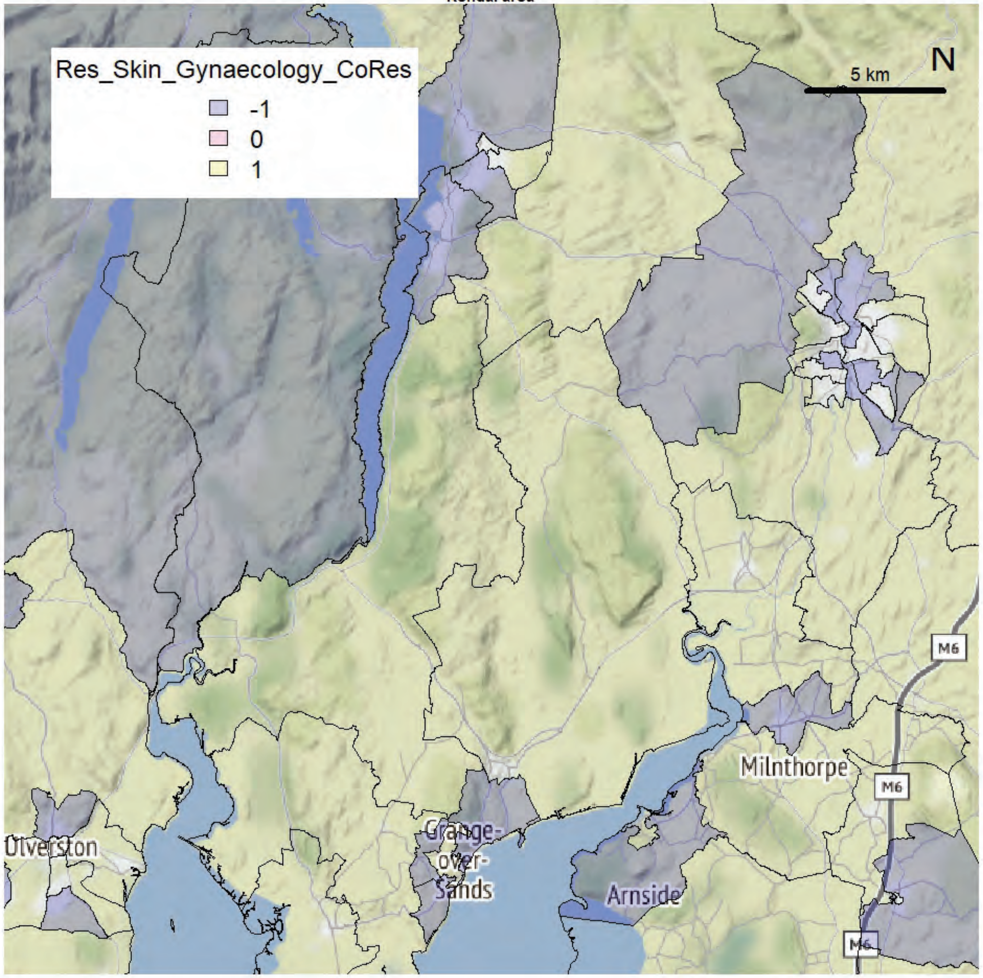

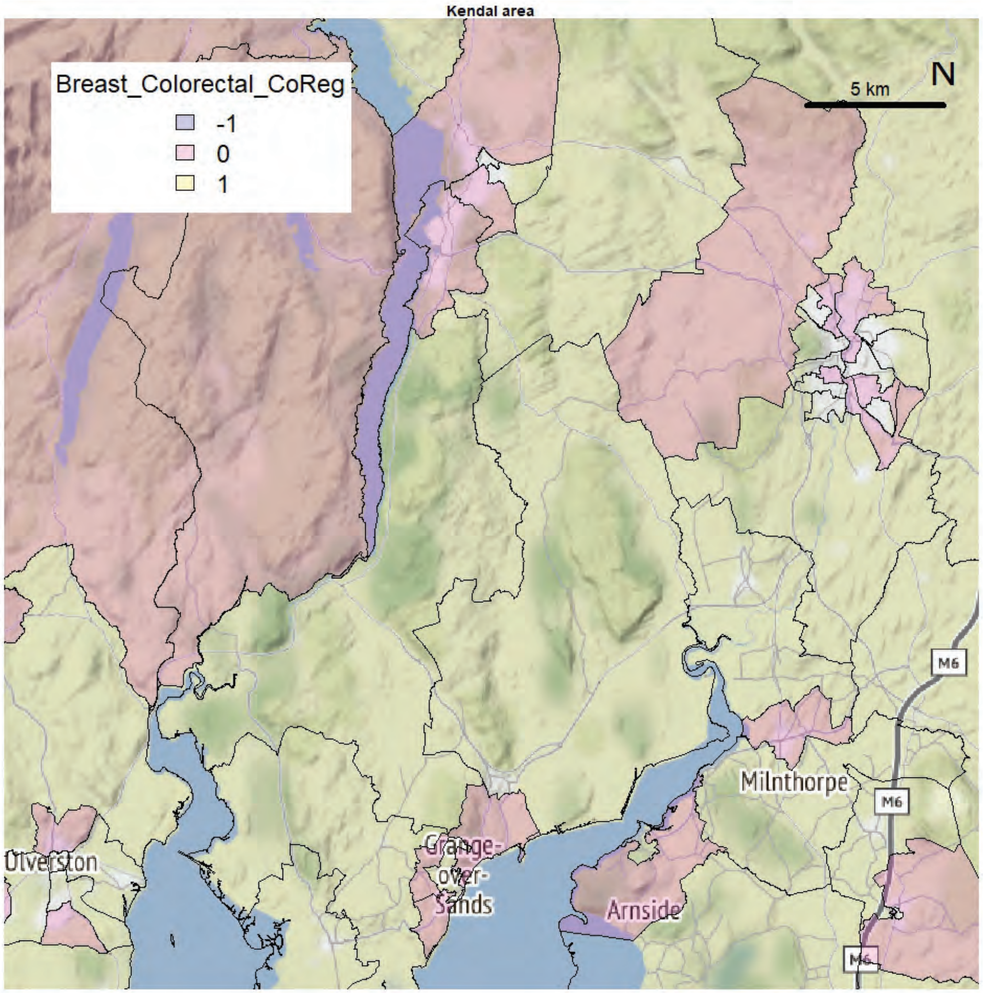
Kendal area
Breast_Colorectal_CoReg
-1
0
1
5 km
N
M6
M6
M6
Milnthorpe
Arnside
Grange-
over-
Sands
Ulverston

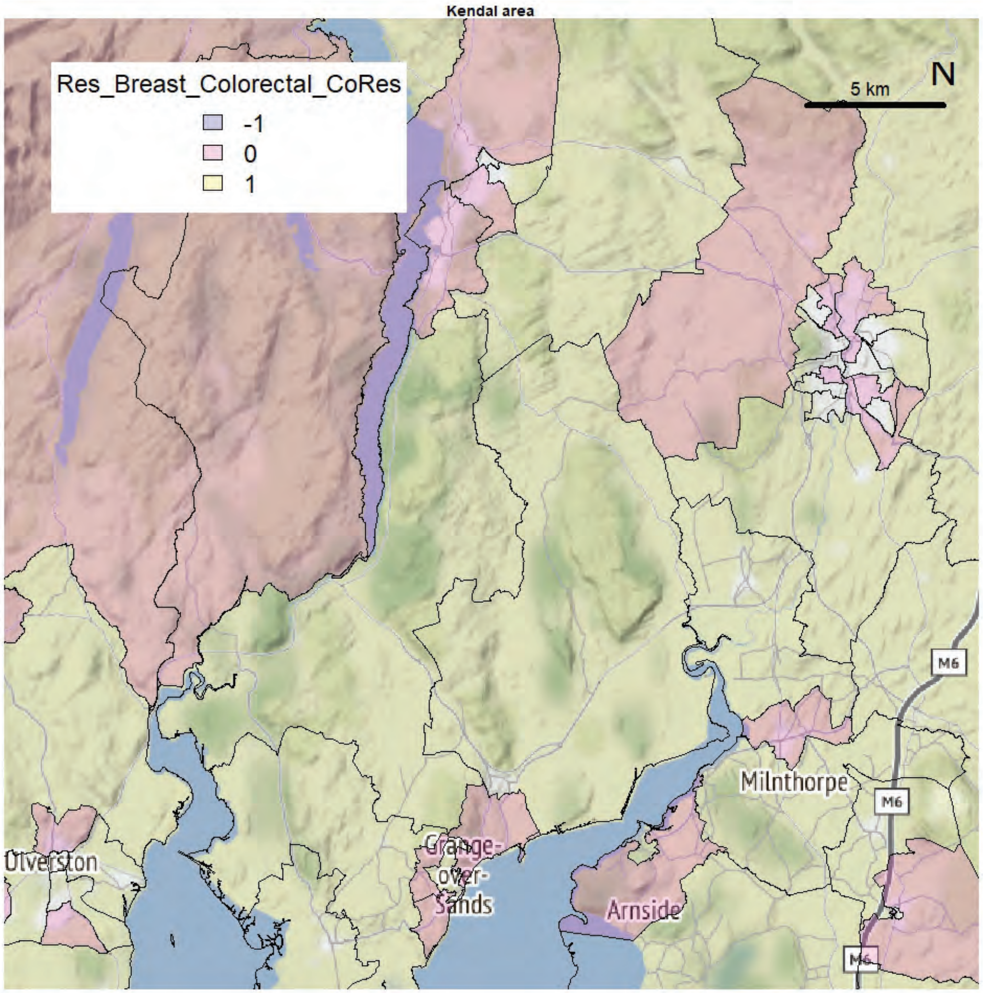
Kendal area
Res_Breast_Colorectal_CoRes
-1
0
1
5 km
N
M6
M6
M6
Milnthorpe
Arnside
Grange-
over-
Sands
Ulverston

Kendal area

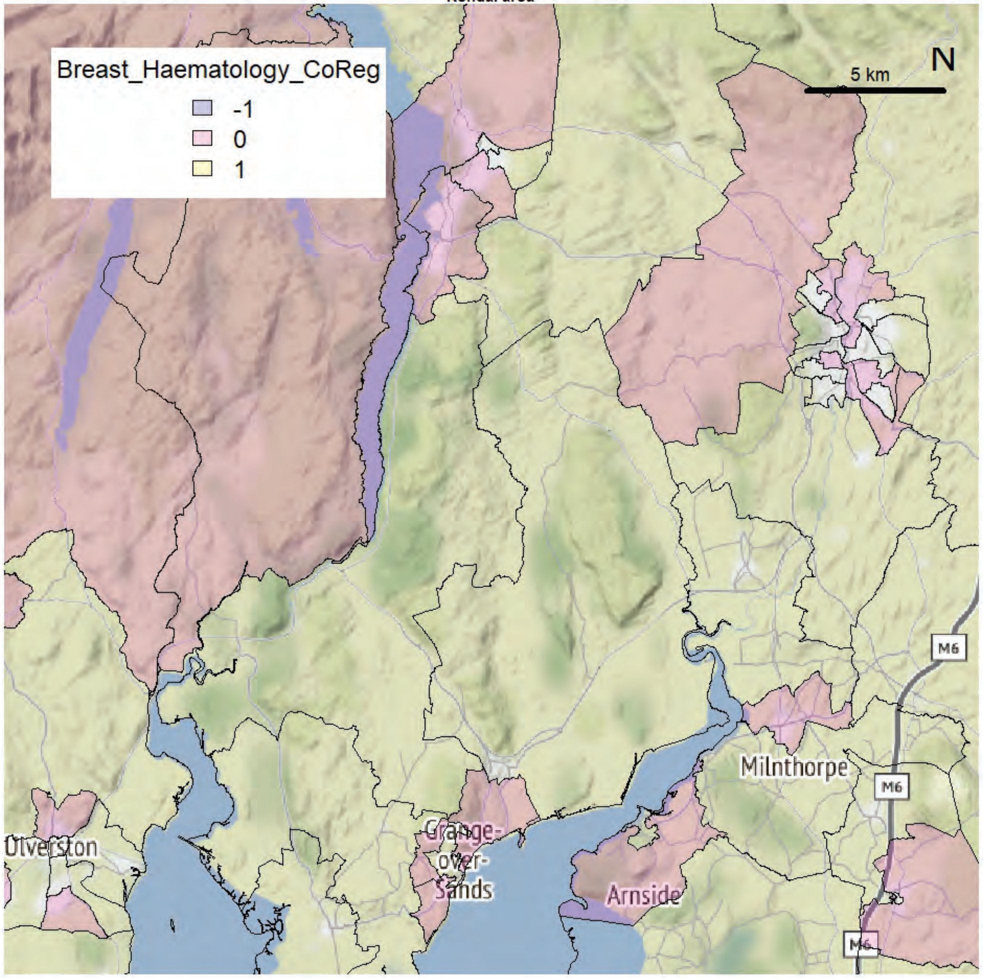


Kendal area

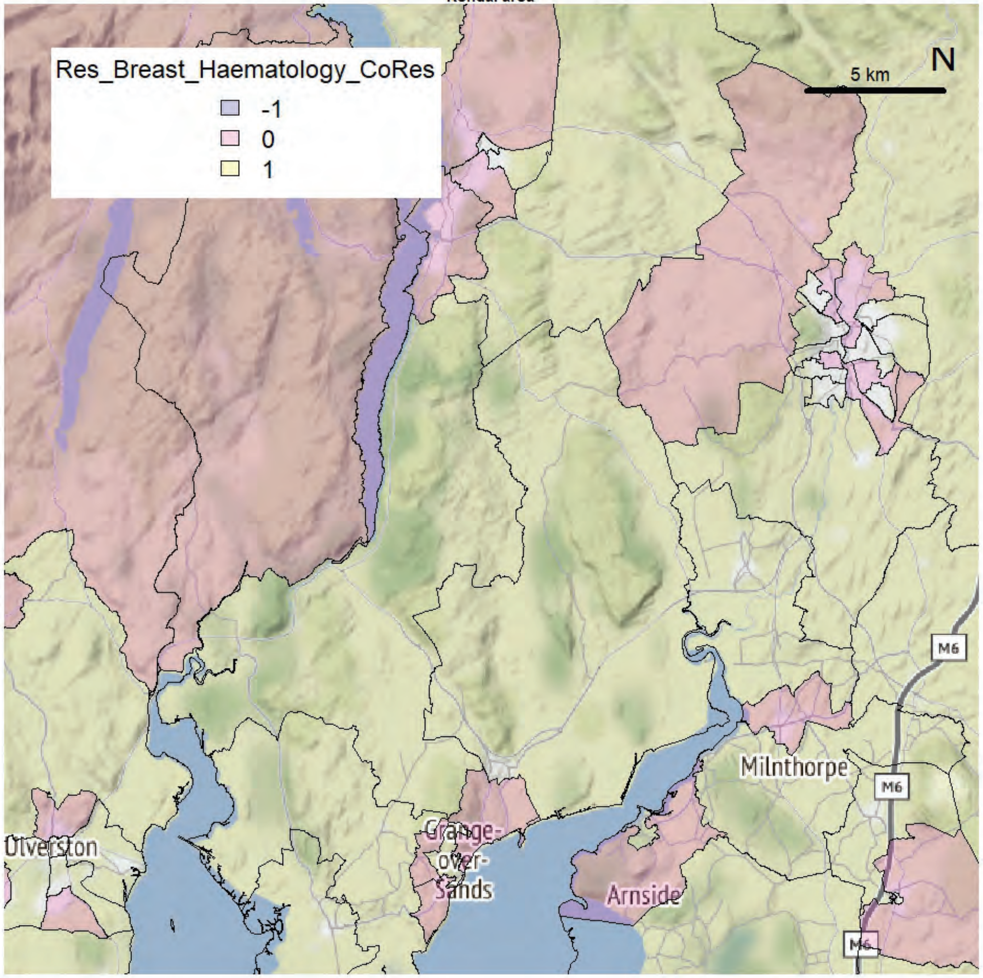

Kendal area

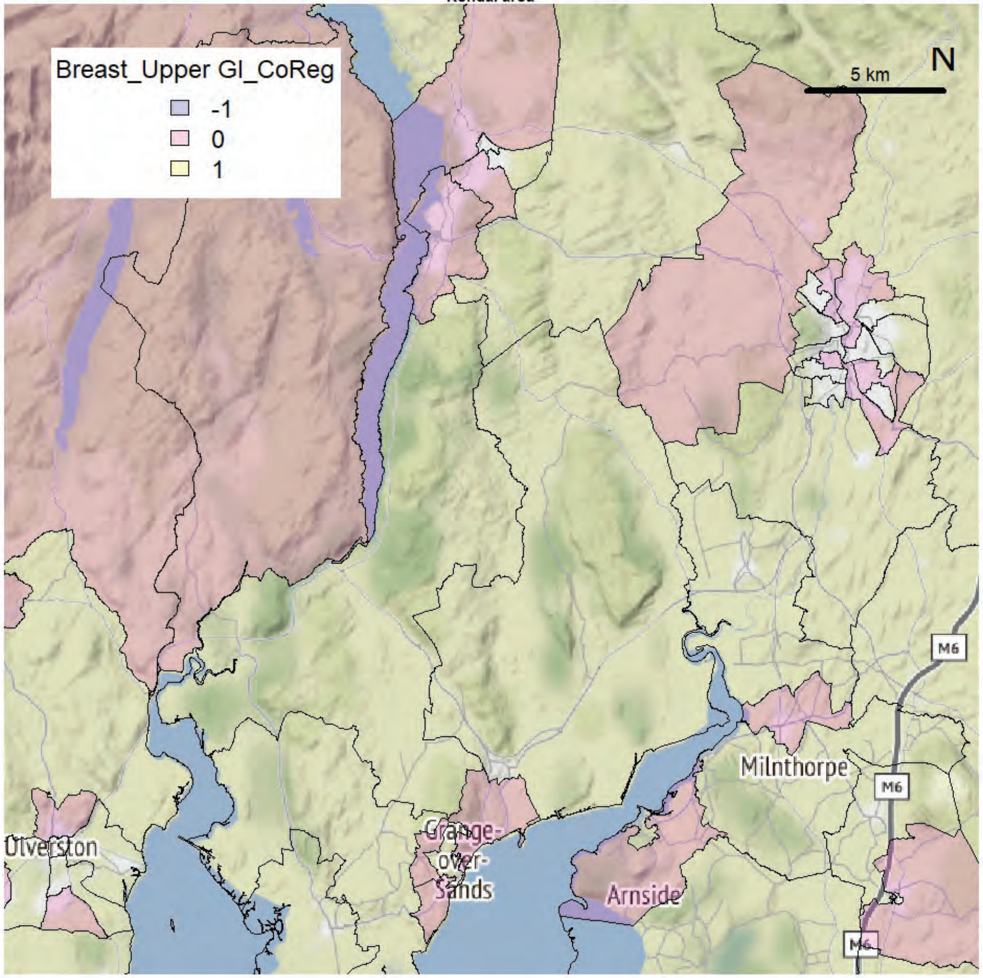


Kendal area

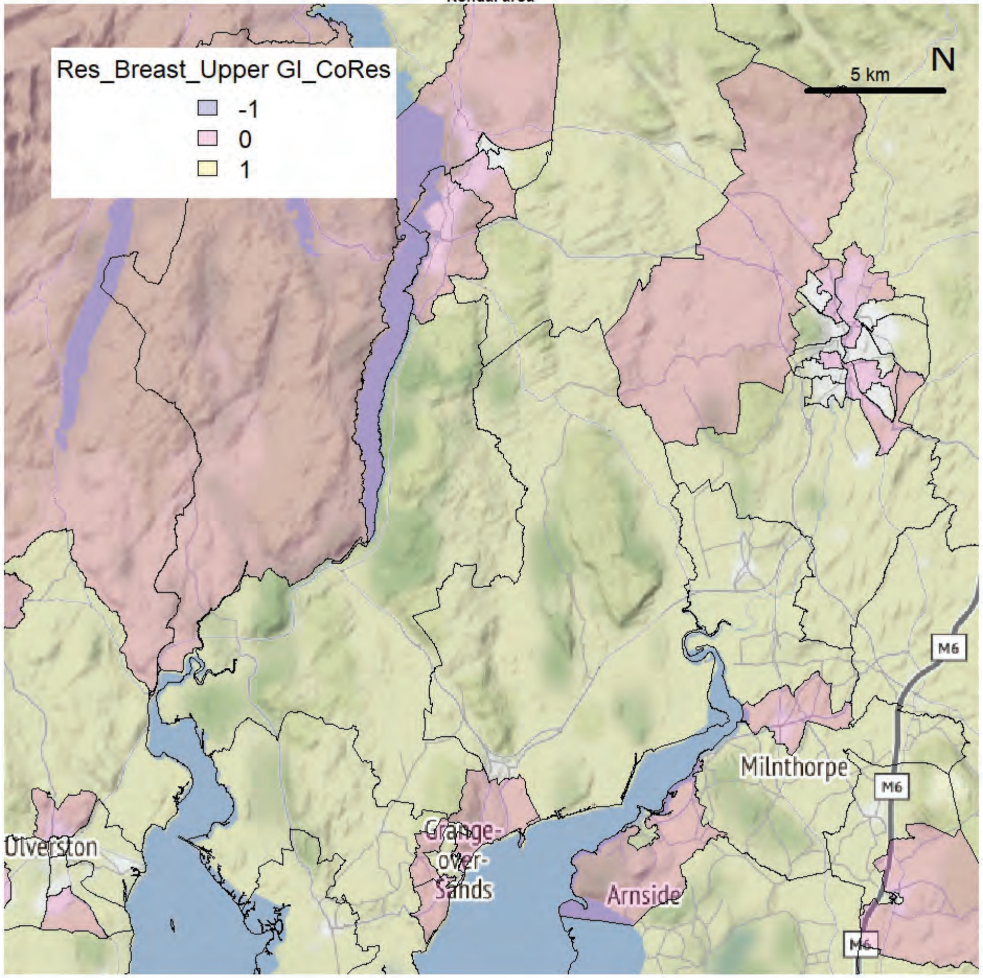

Kendal area

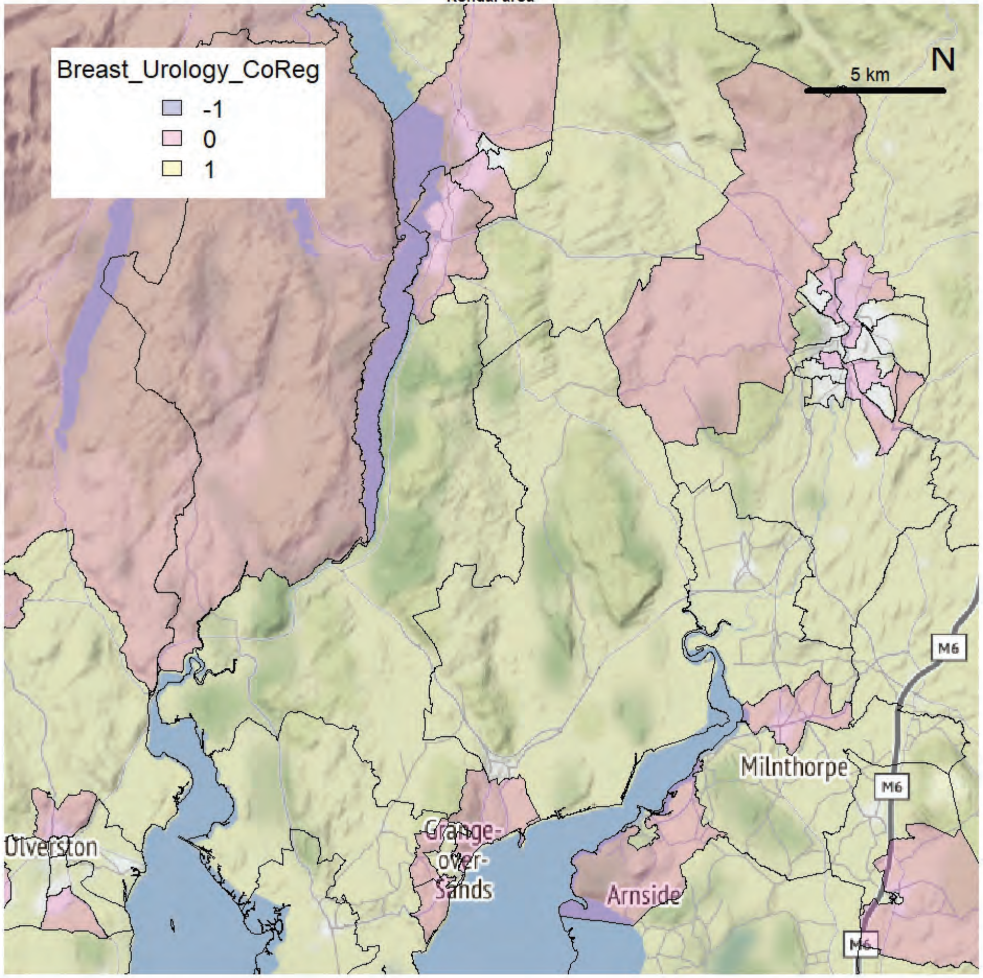


Kendal area

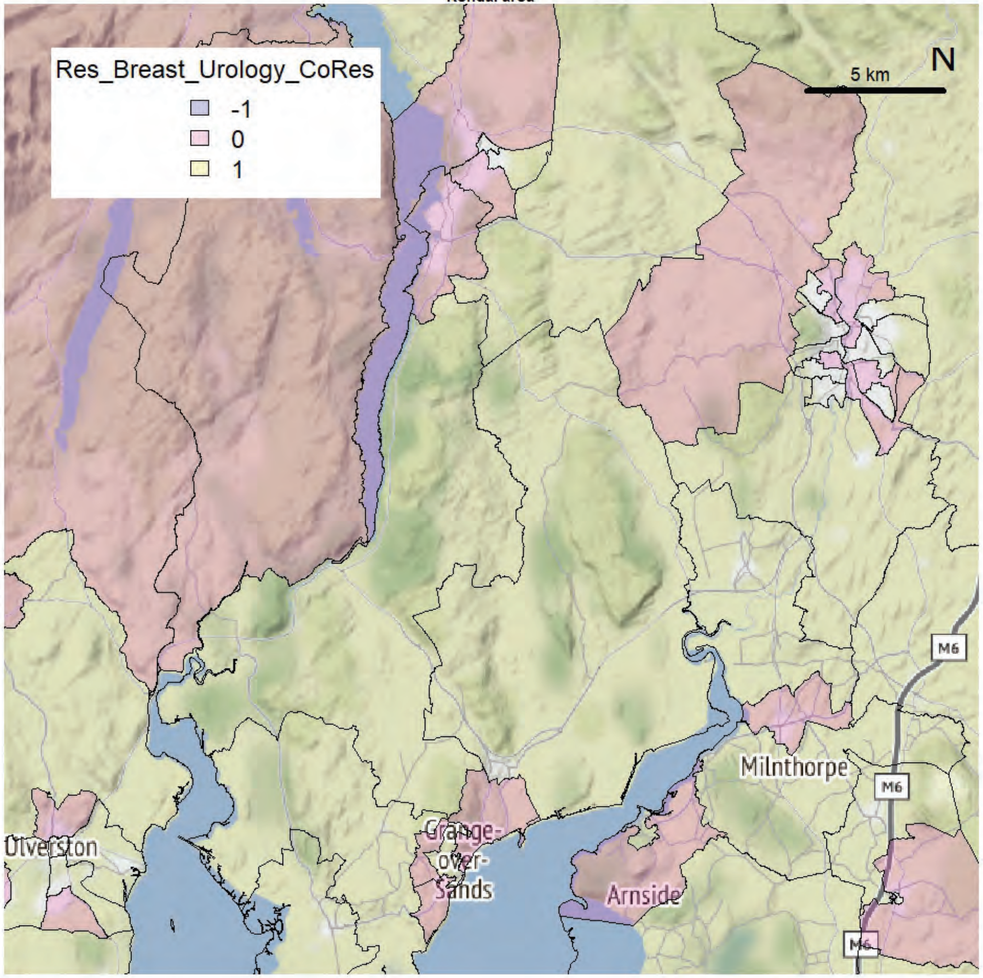

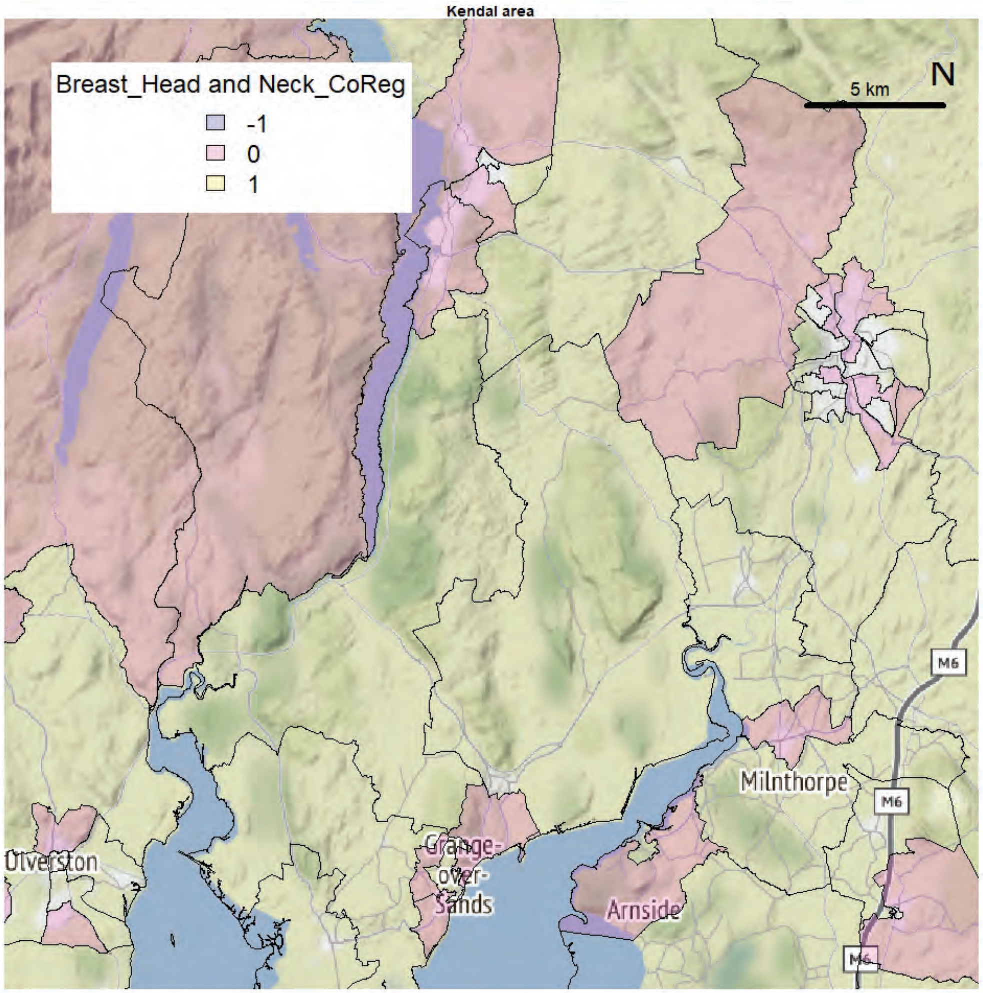
Kendal area
Breast_Head and Neck_CoReg
-1
0
1
5 km
N
M6
Milnthorpe
Ulverston
Grange-over-Sands
Arnside

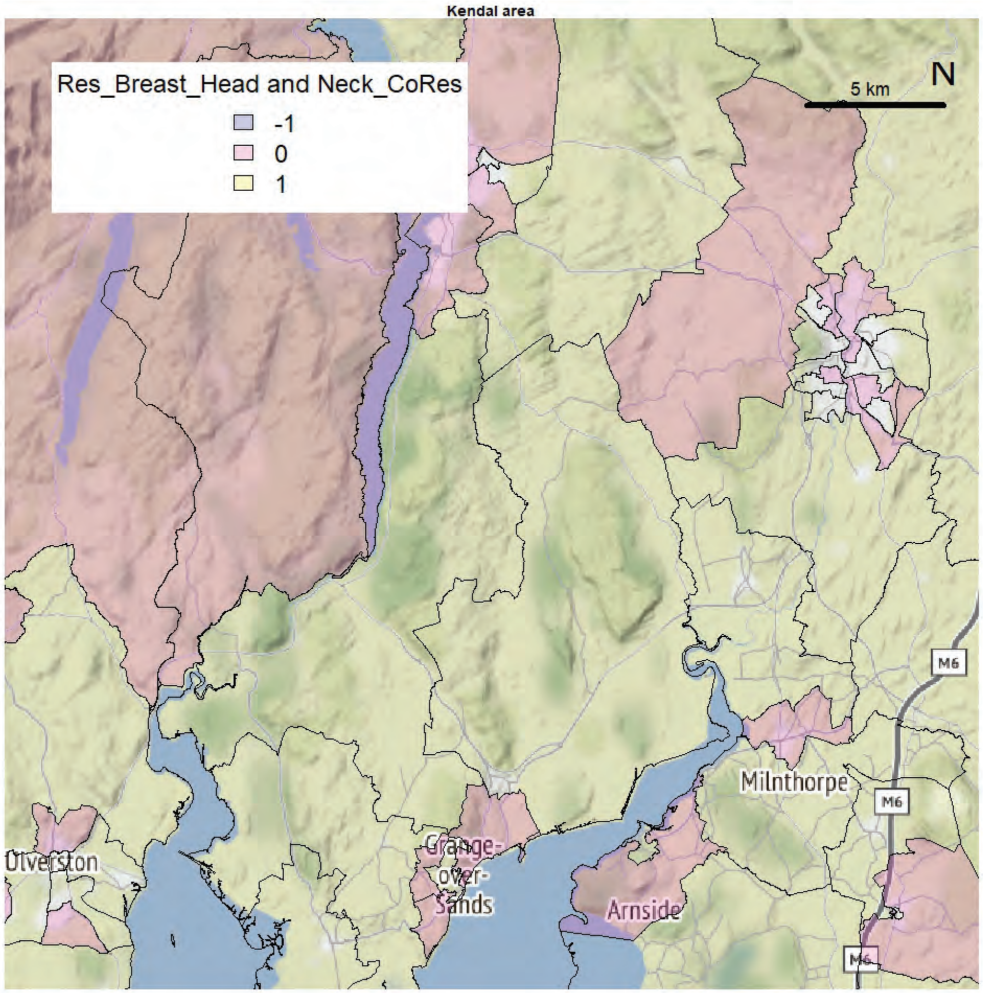
Kendal area
Res_Breast_Head and Neck_CoRes
-1
0
1
5 km
N
M6
Milnthorpe
Ulverston
Grange-over-Sands
Arnside

Kendal area

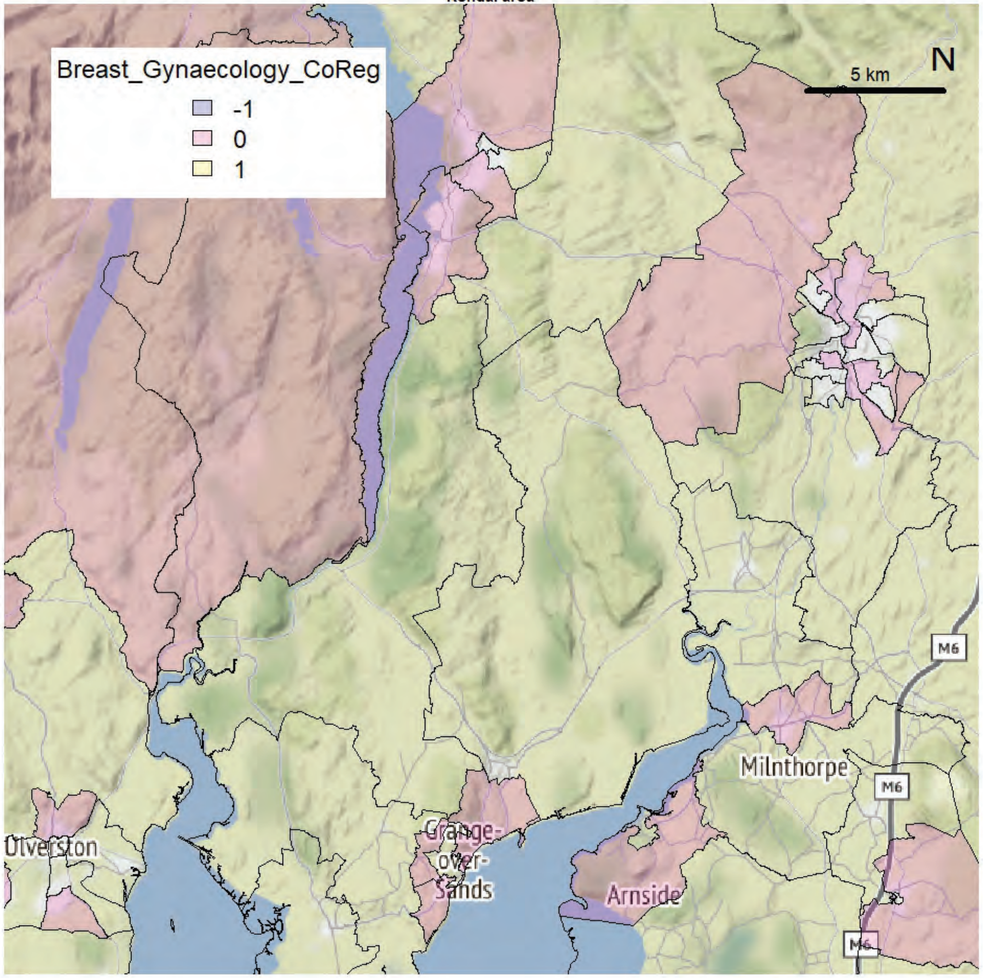


Kendal area

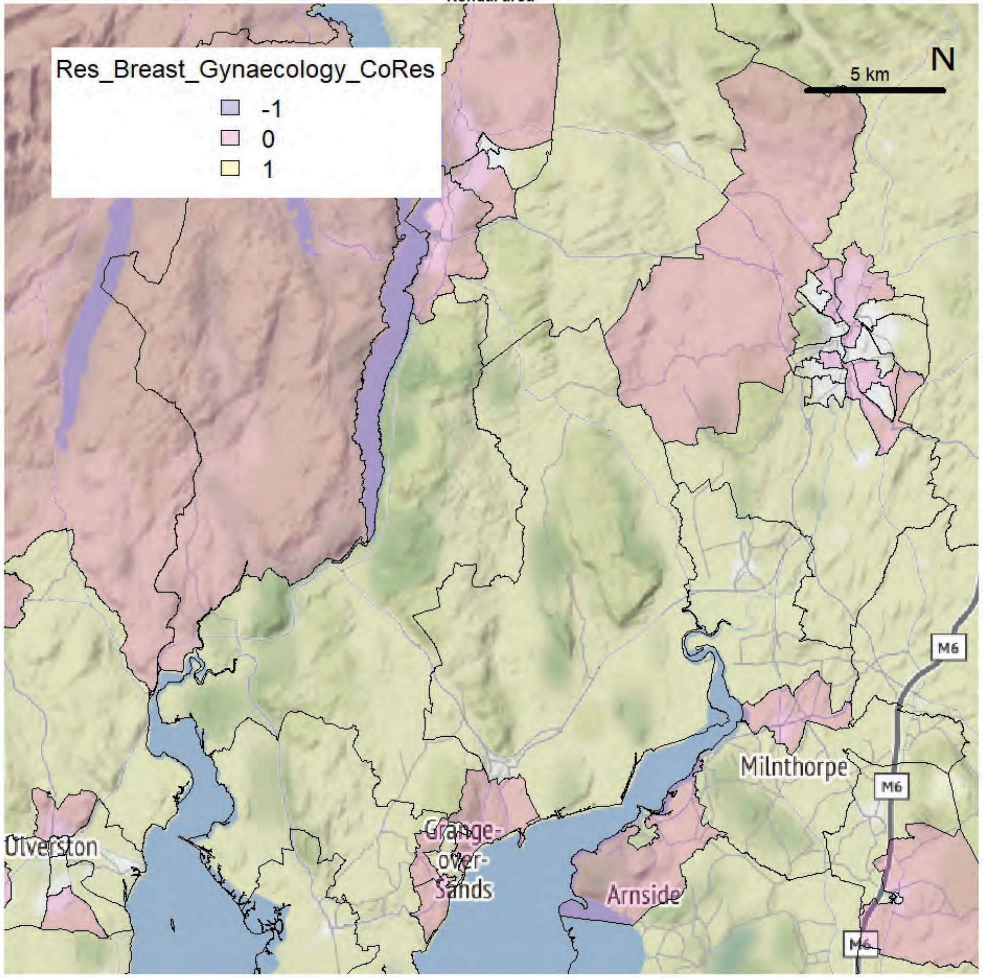

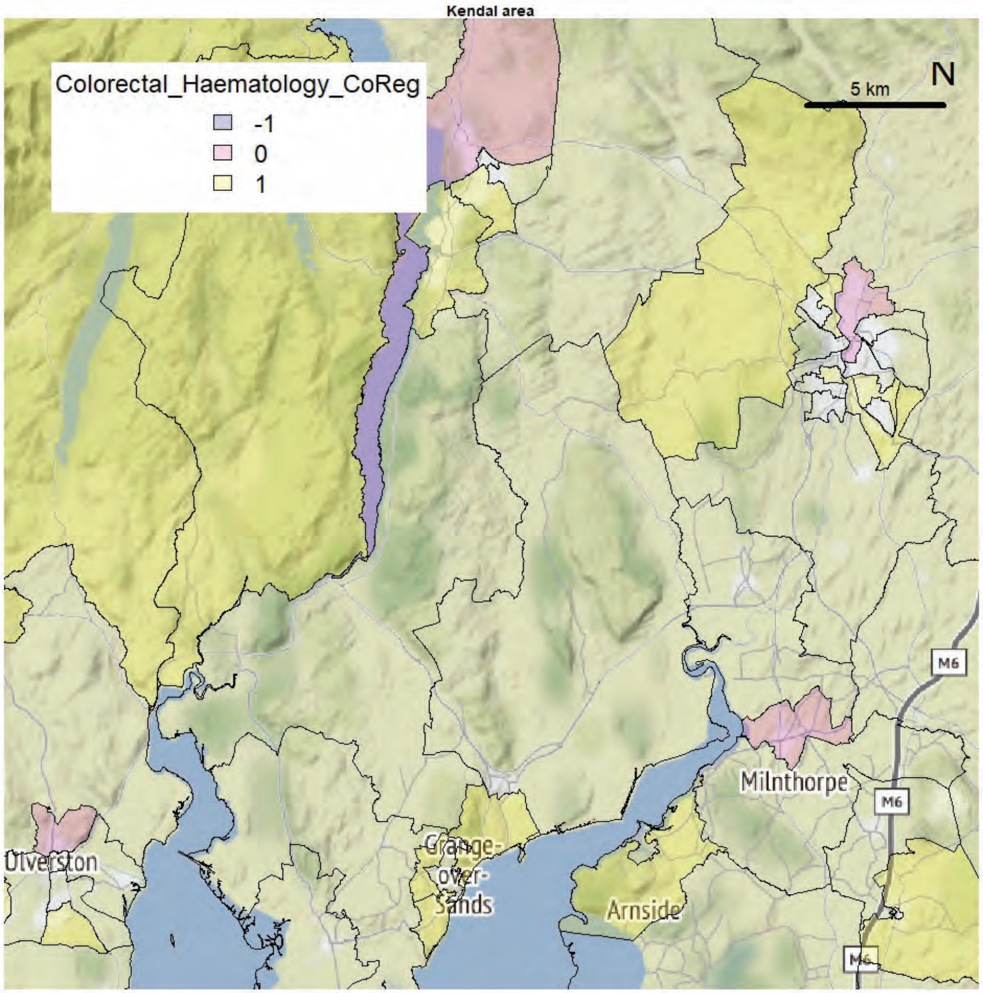
Kendal area
Colorectal_Haematology_CoReg
-1
0
1
5 km
N
Milnthorpe
M6
Ulverston
Grange-over-Sands
Arnside

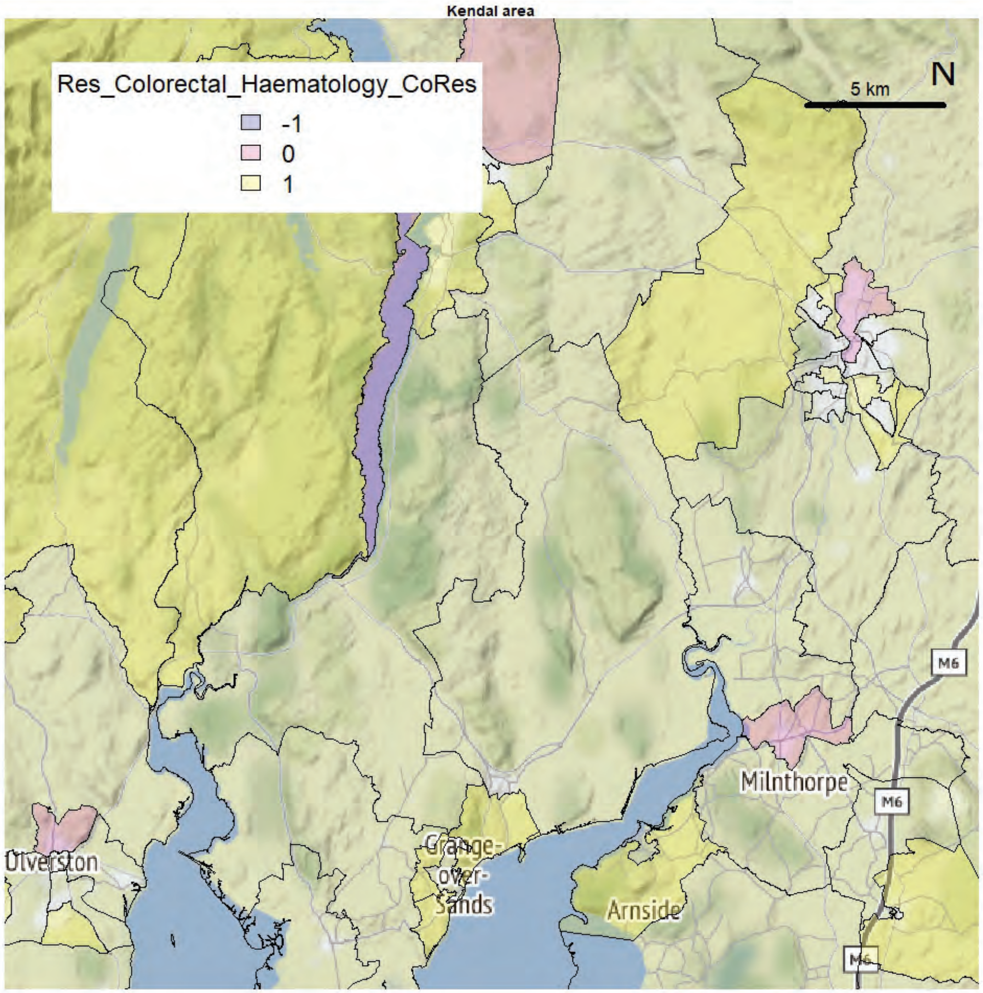
Kendal area
Res_Colorectal_Haematology_CoRes
-1
0
1
5 km
N
Milnthorpe
M6
Ulverston
Grange-over-Sands
Arnside

Kendal area

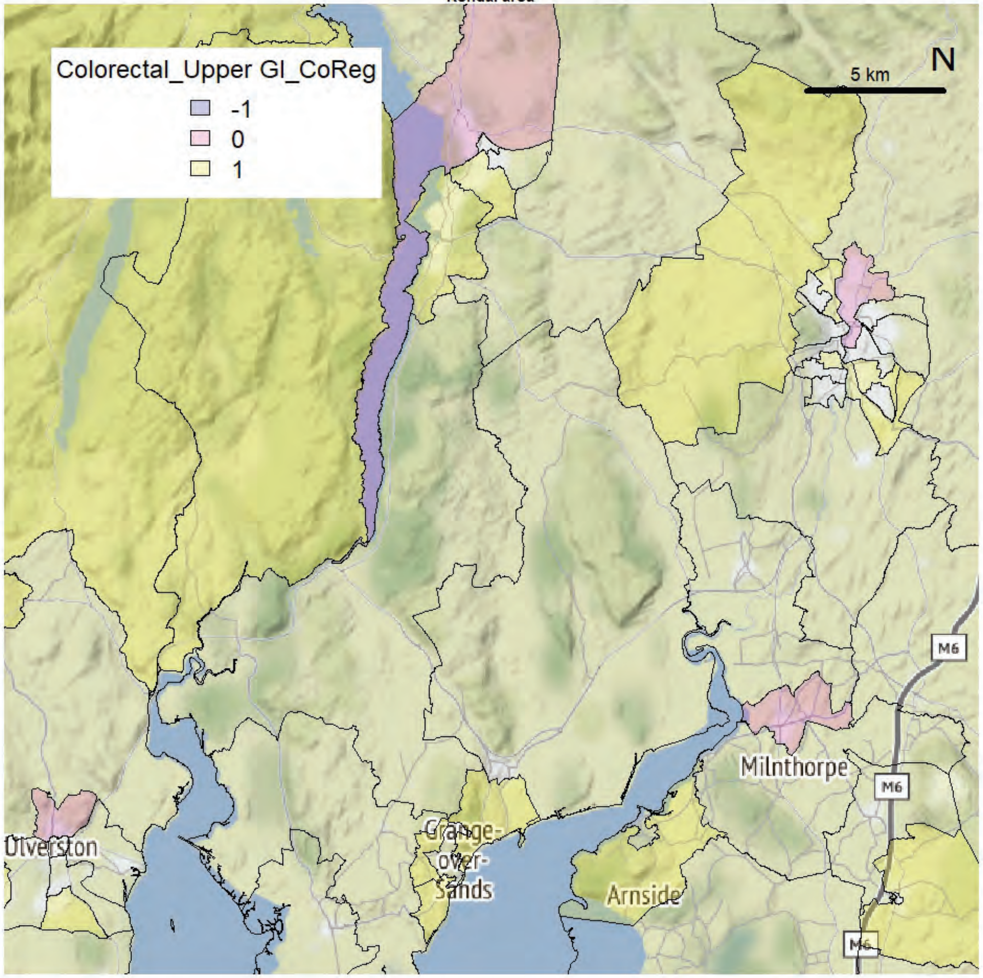


Kendal area

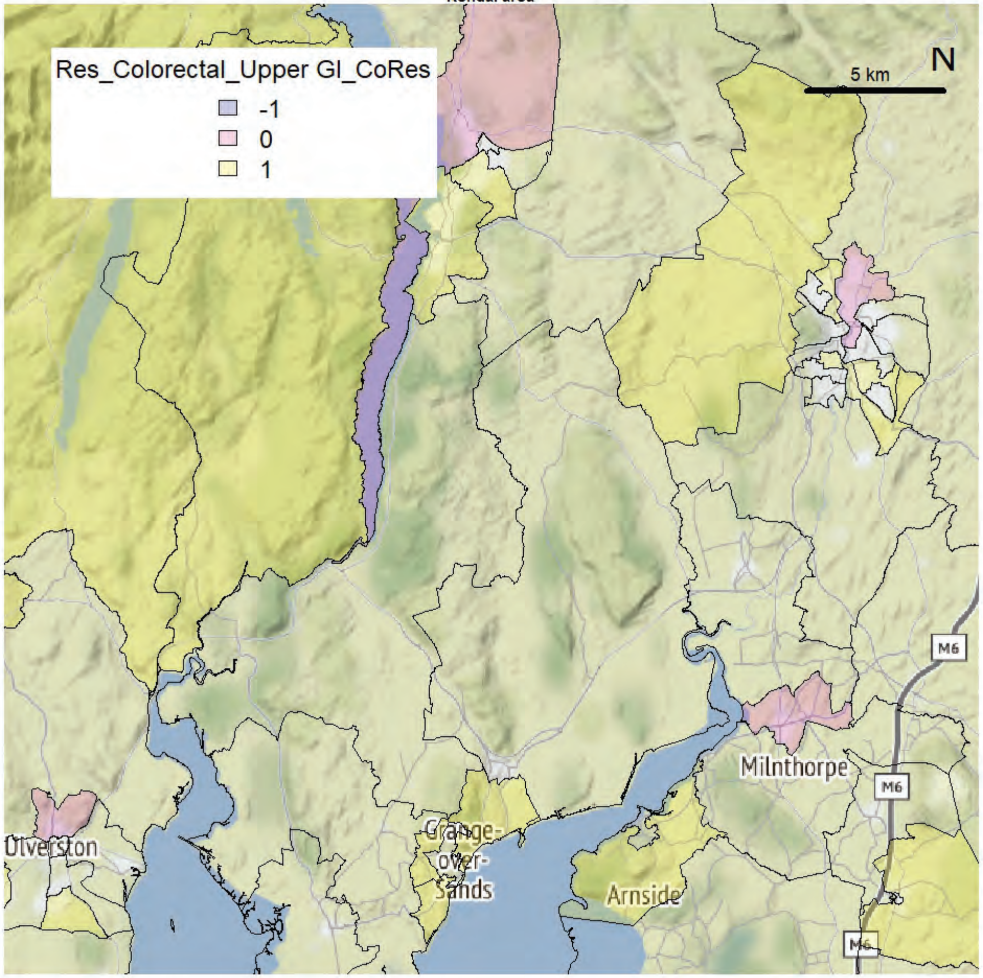

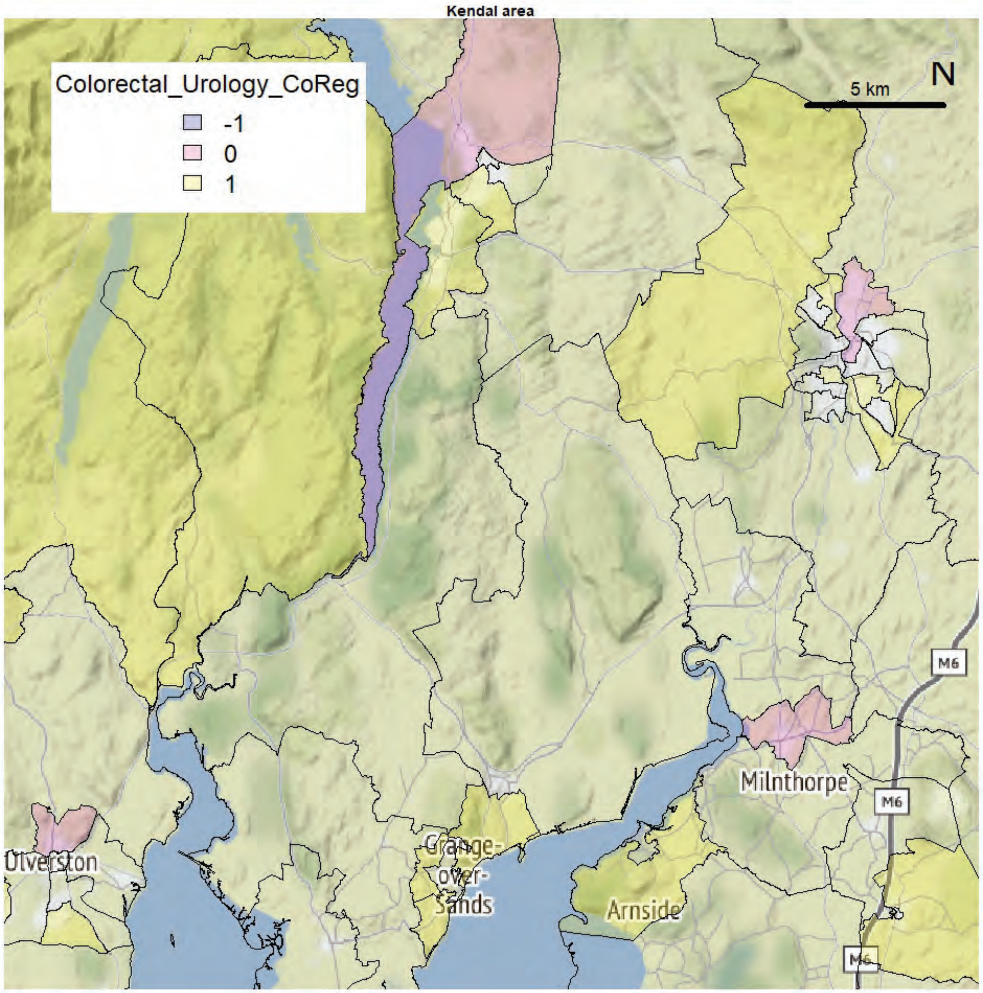
Kendal area
Colorectal_Urology_CoReg
-1
0
1
5 km
N
M6
Milnthorpe
Ulverston
Grange-over-Sands
Arnside

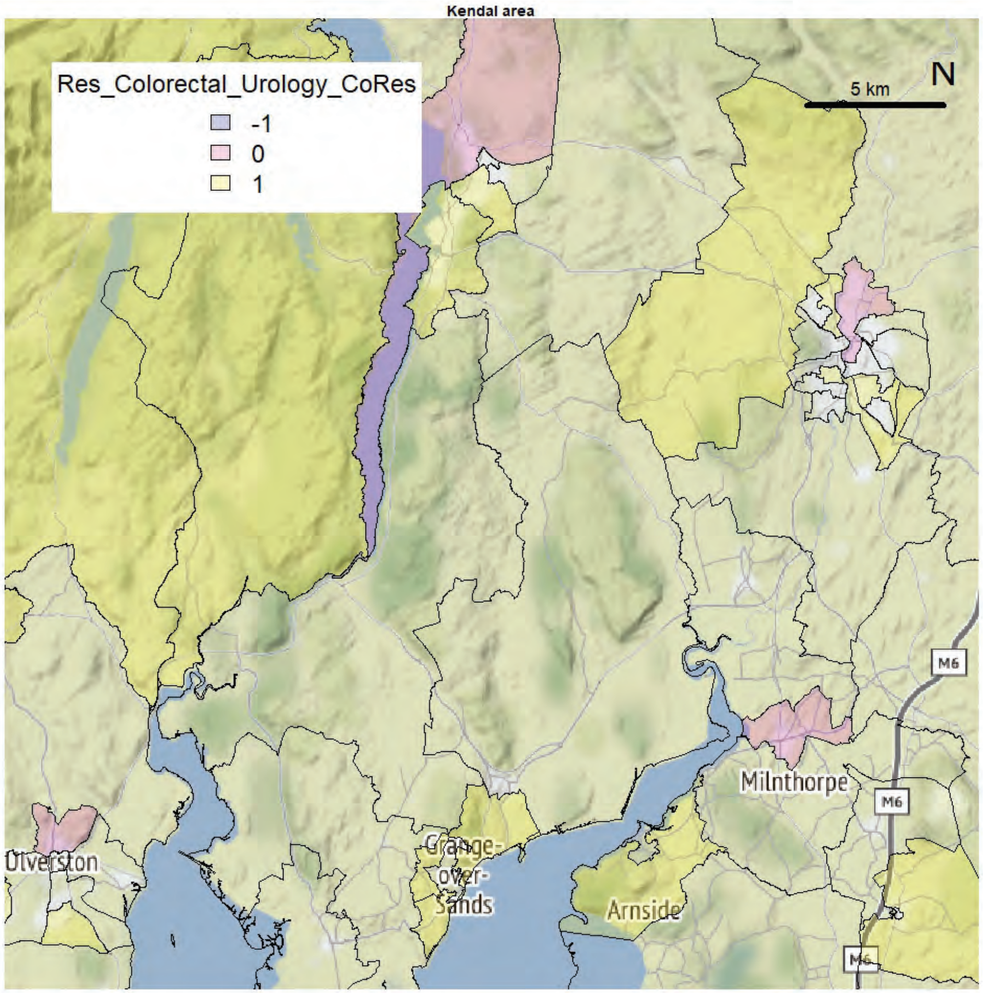
Kendal area
Res_Colorectal_Urology_CoRes
-1
0
1
5 km
N
M6
Milnthorpe
Ulverston
Grange-over-Sands
Arnside

Kendal area

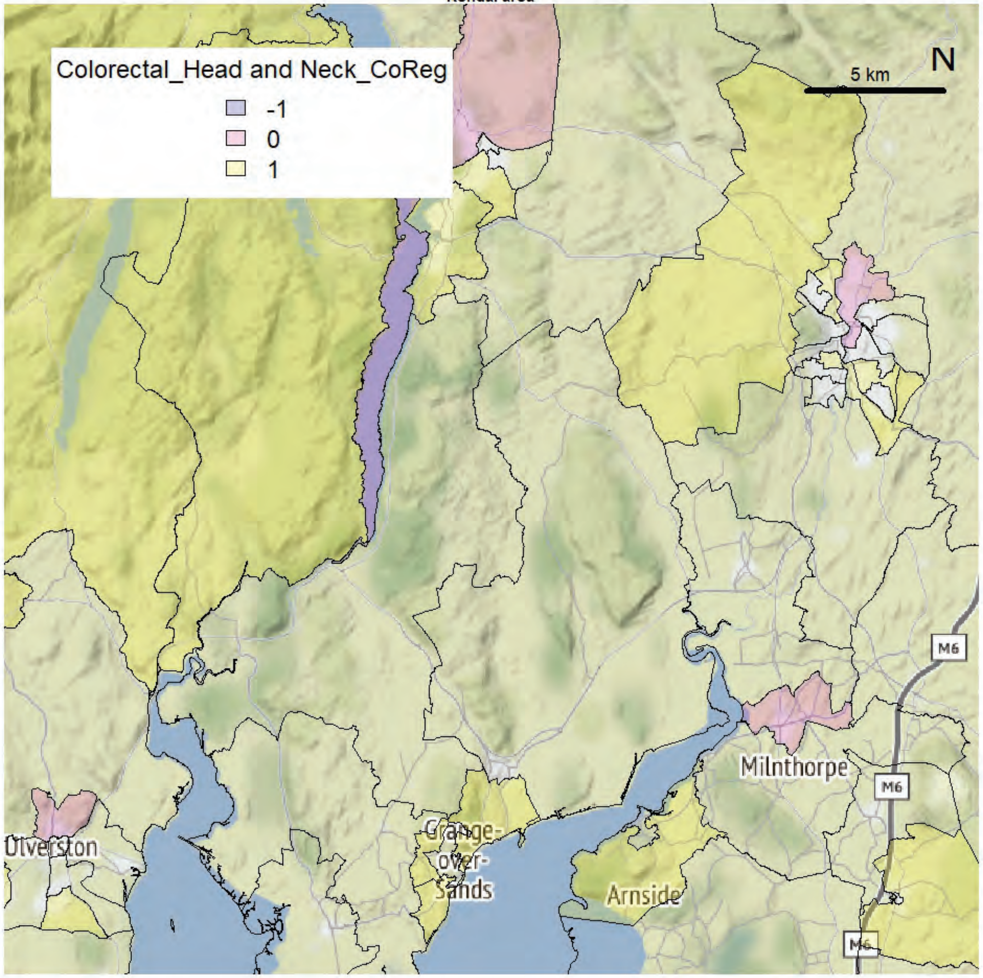


Kendal area

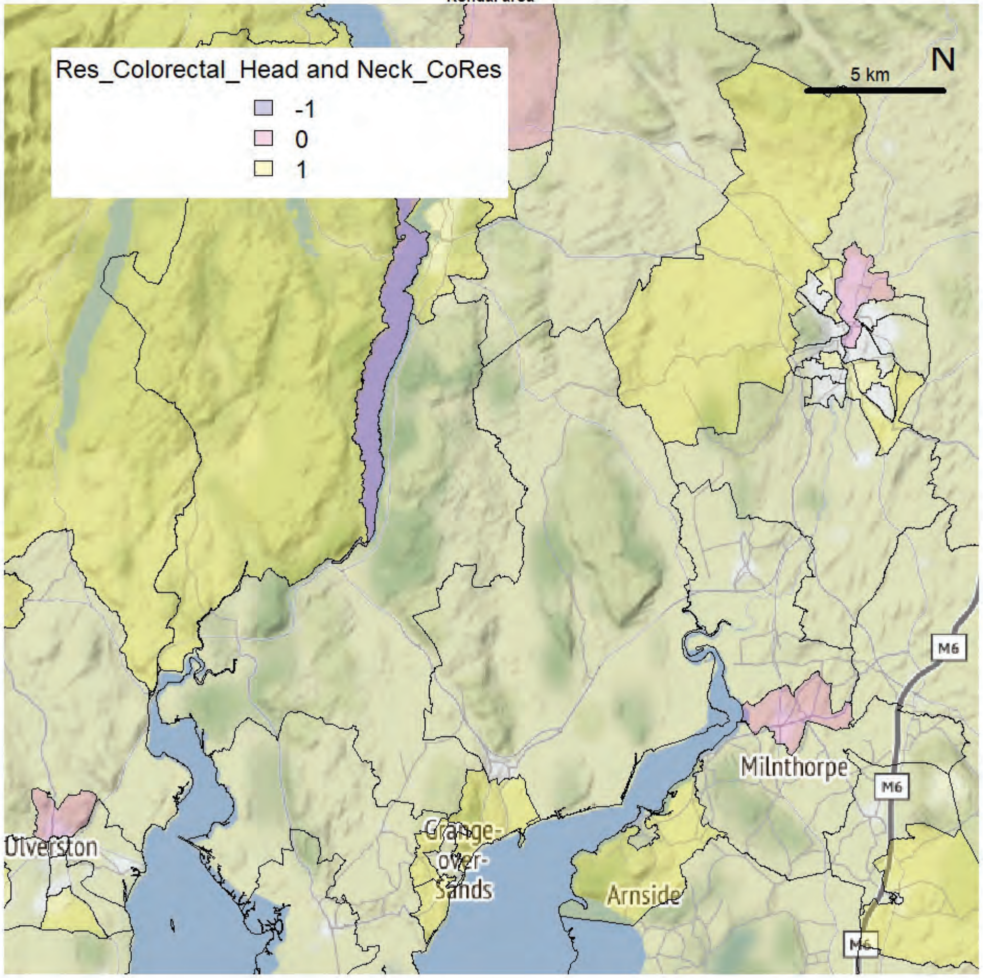

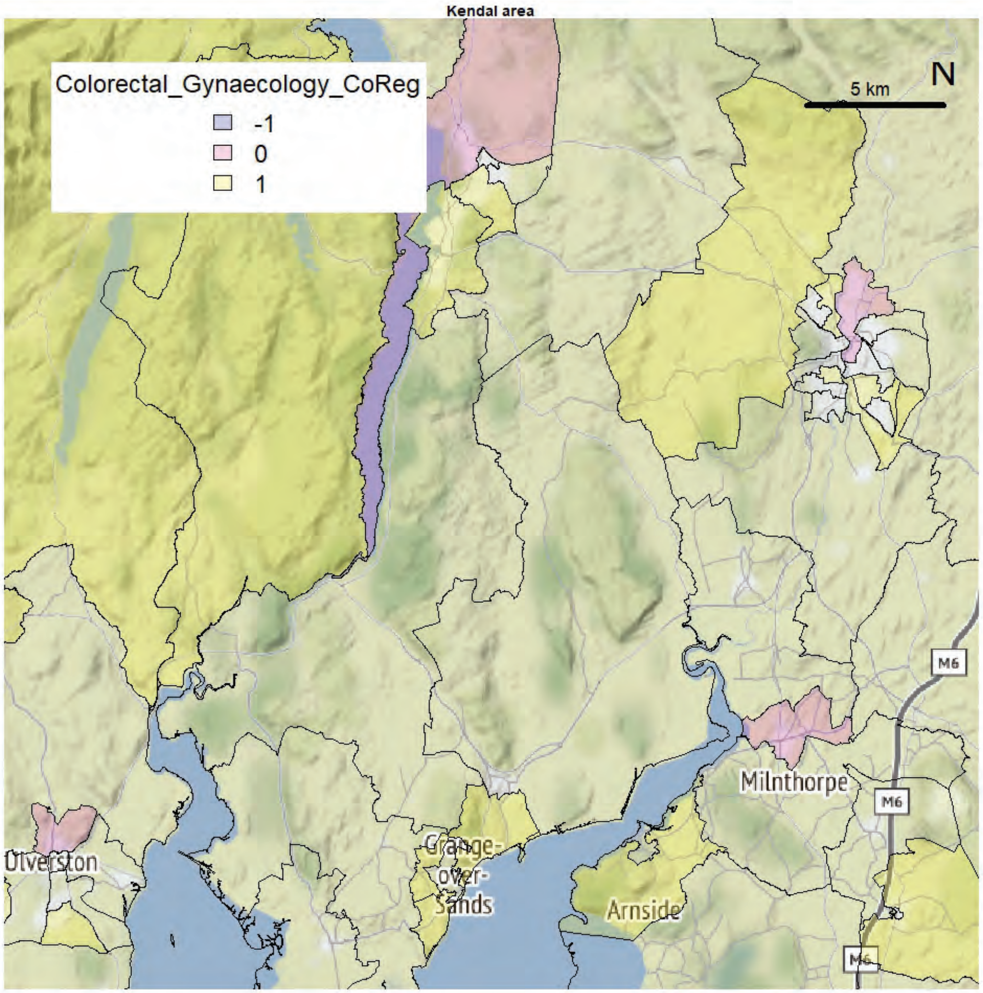
Kendal area
Colorectal_Gynaecology_CoReg
-1
0
1
5 km
N
M6
M6
M6
Milnthorpe
Ulverston
Grange-over-Sands
Arnside

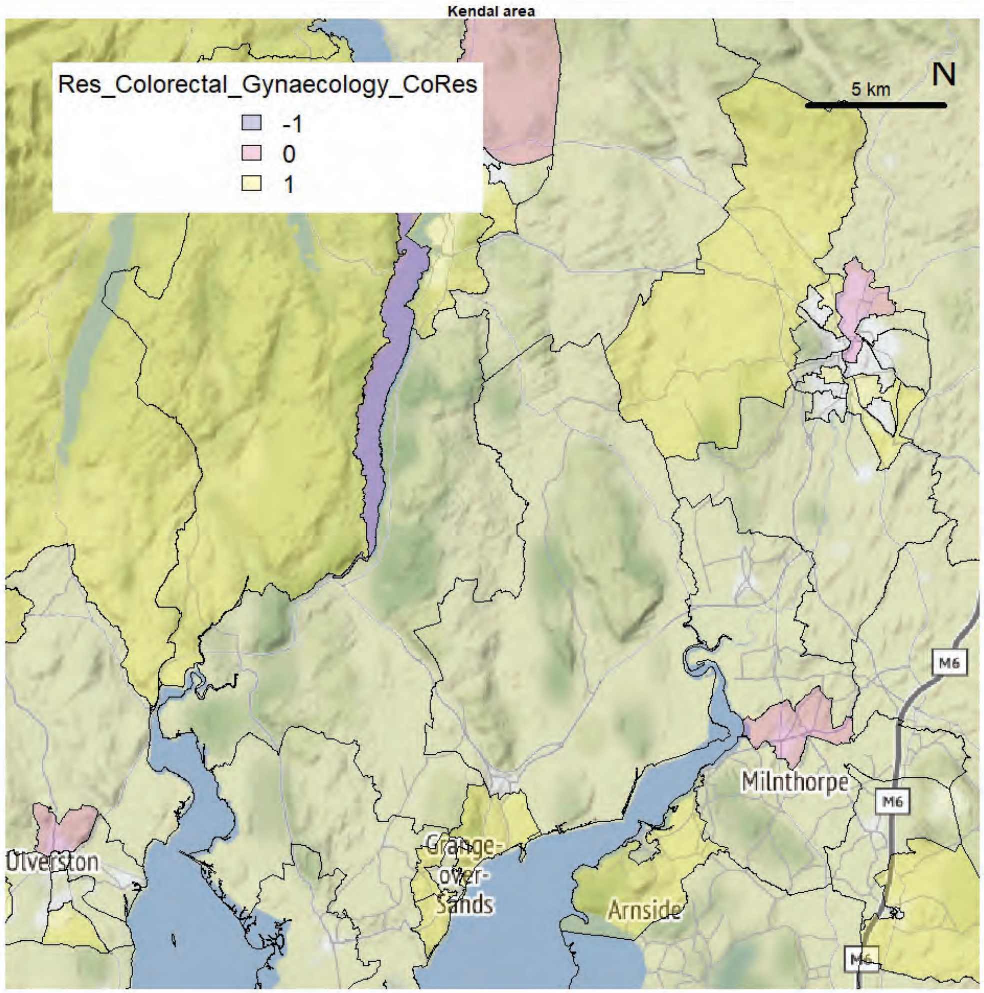
Kendal area
Res_Colorectal_Gynaecology_CoRes
-1
0
1
5 km
N
M6
M6
M6
Milnthorpe
Ulverston
Grange-over-Sands
Arnside

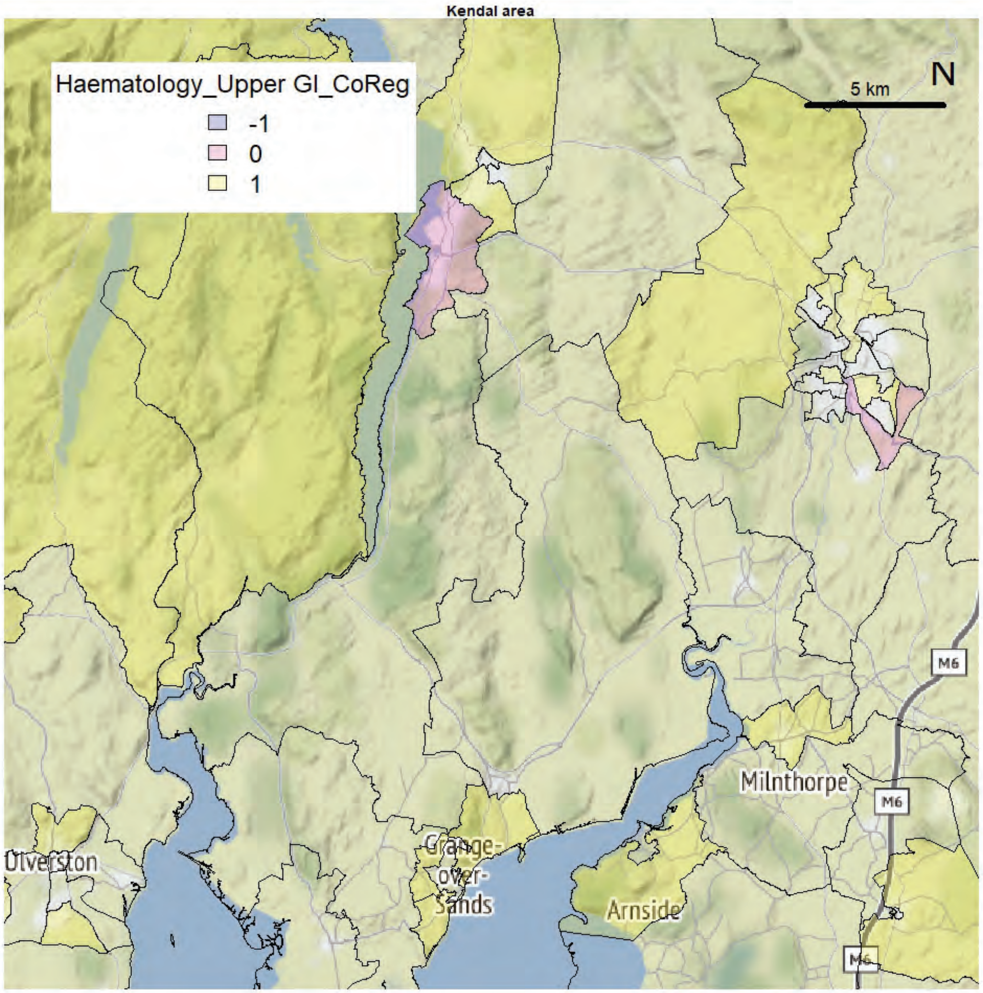
Kendal area
Haematology_Upper GI_CoReg
-1
0
1
5 km
N
Milnthorpe
M6
Ulverston
Grange-over-Sands
Arnside

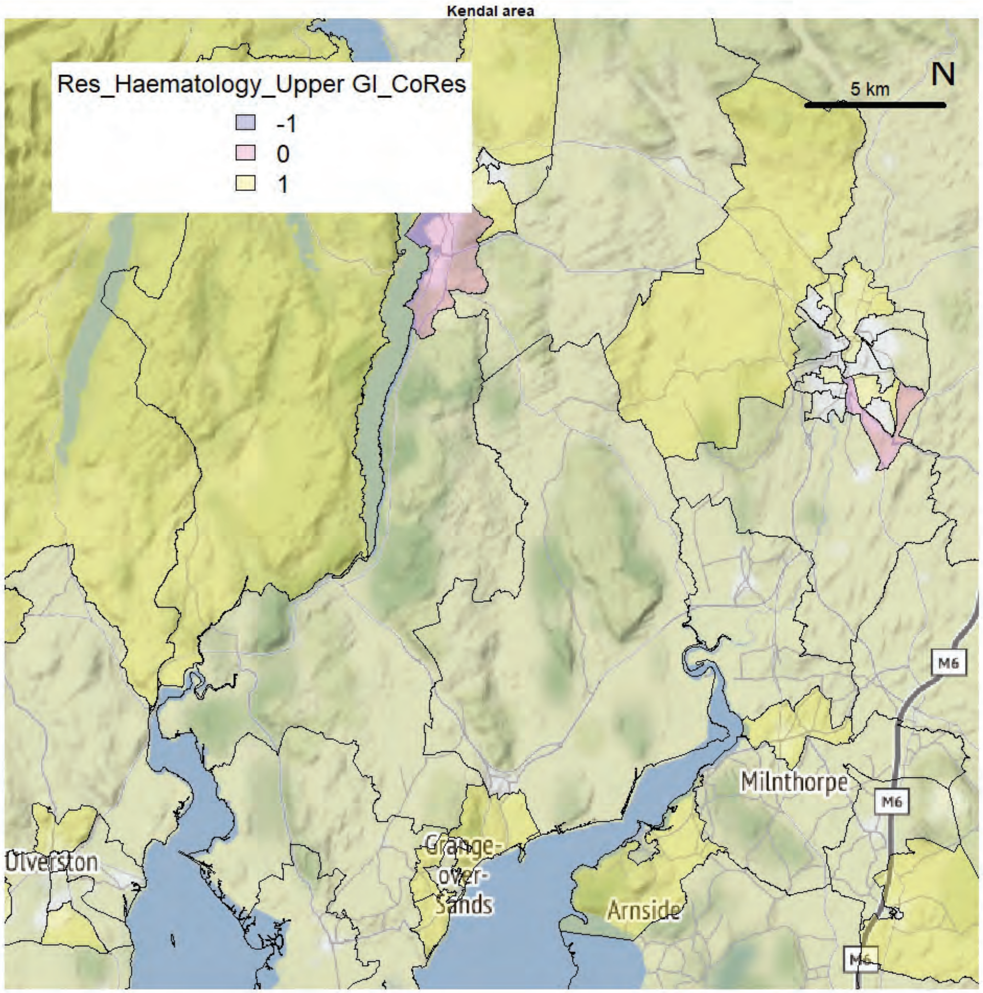
Kendal area
Res_Haematology_Upper GI_CoRes
-1
0
1
5 km
N
Milnthorpe
M6
Ulverston
Grange-over-Sands
Arnside

Kendal area

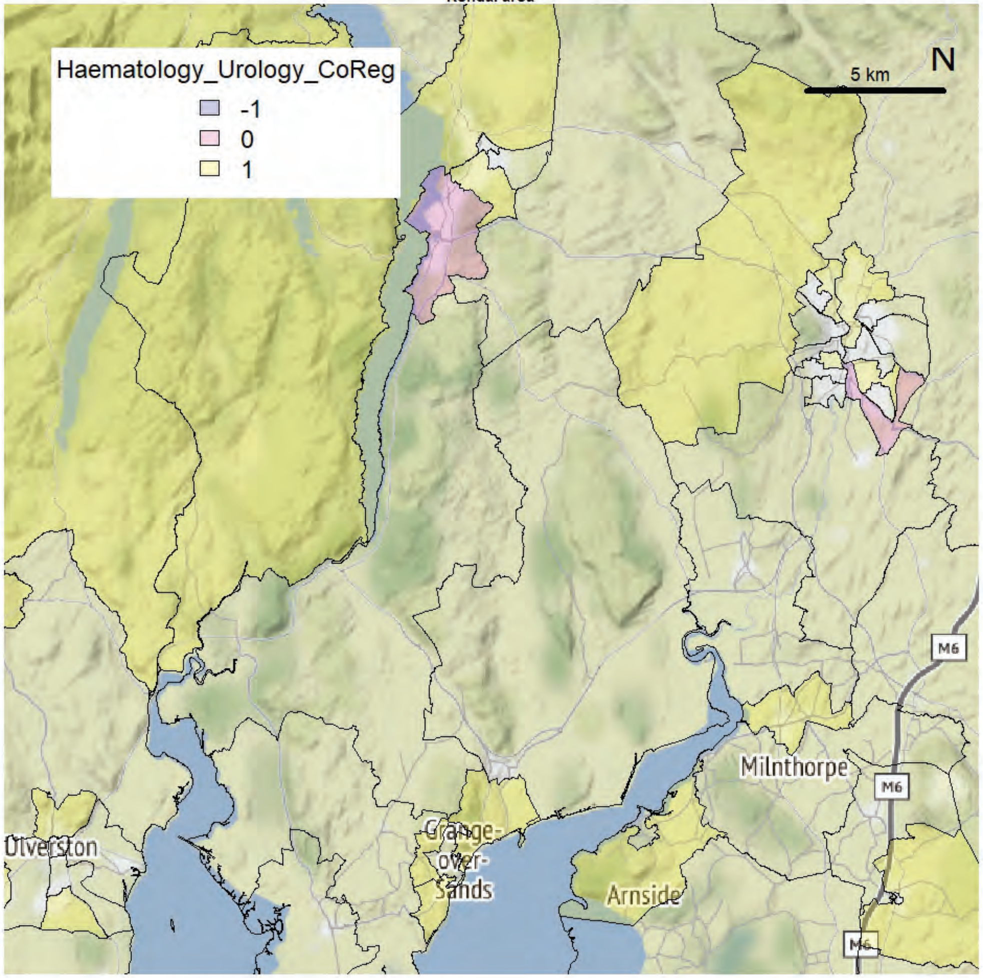


Kendal area

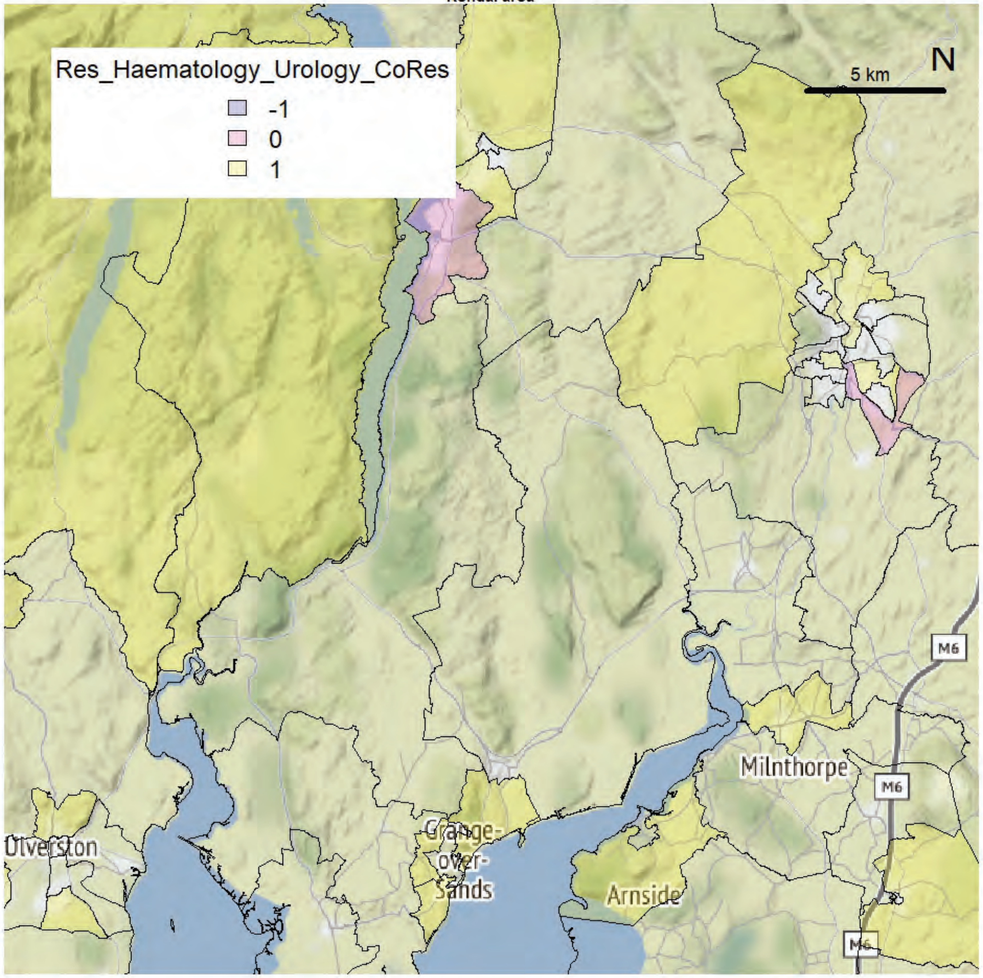

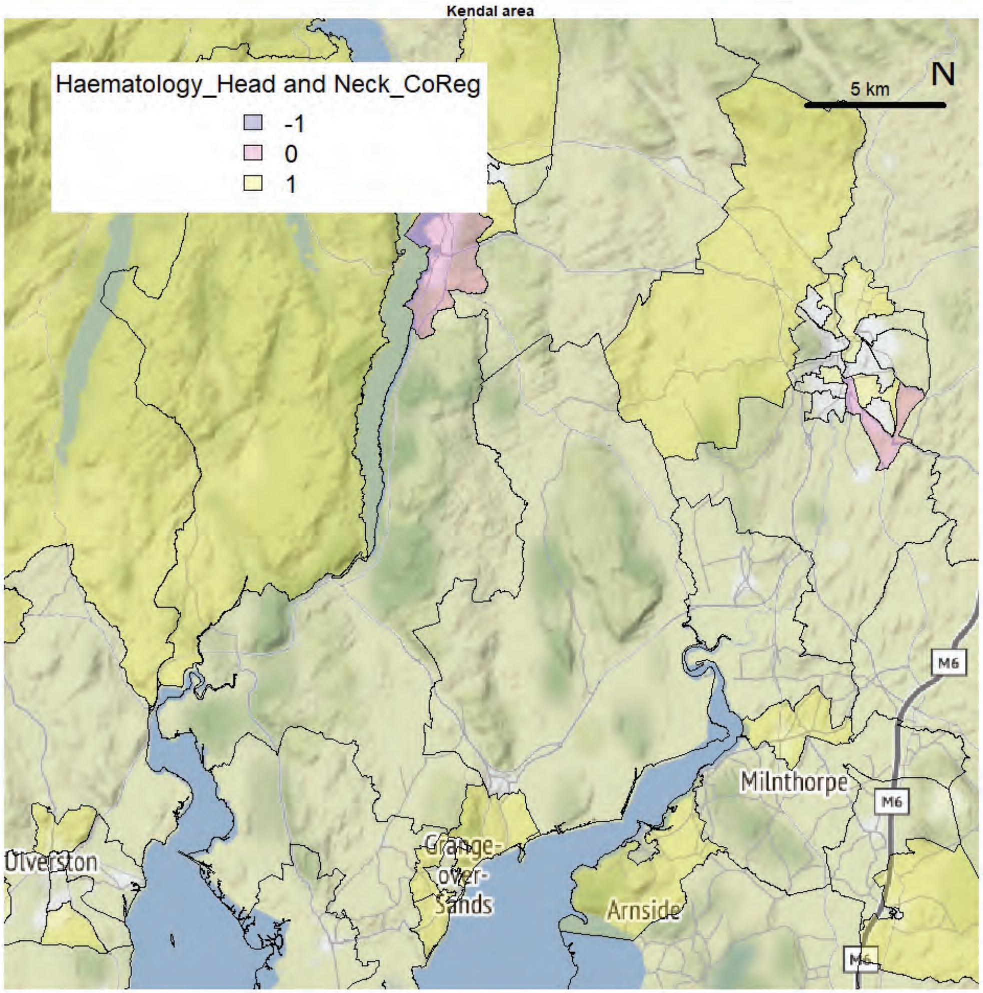
Kendal area
Haematology_Head and Neck_CoReg
-1
0
1
5 km
N
Ulverston
Grange-
over-
Sands
Arnside
Milnthorpe
M6

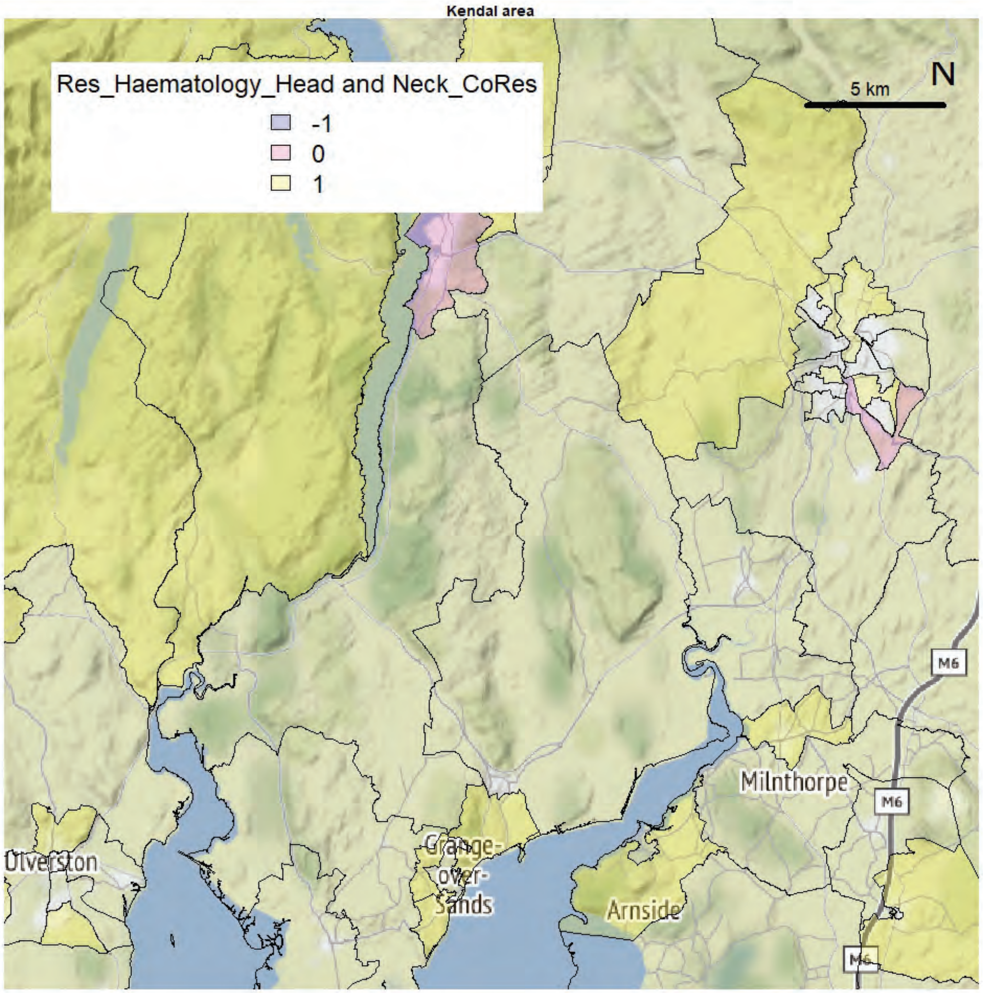
Kendal area
Res_Haematology_Head and Neck_CoRes
-1
0
1
5 km
N
Ulverston
Grange-
over-
Sands
Arnside
Milnthorpe
M6

Kendal area

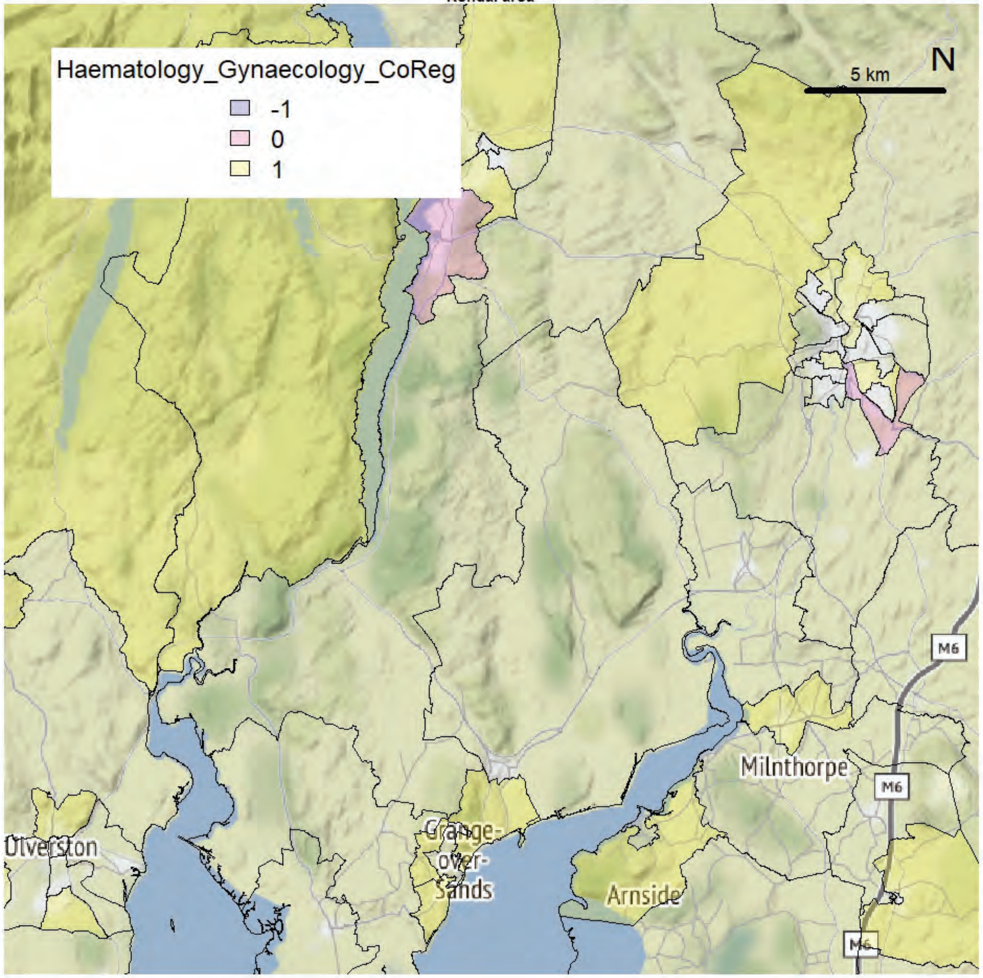


Kendal area

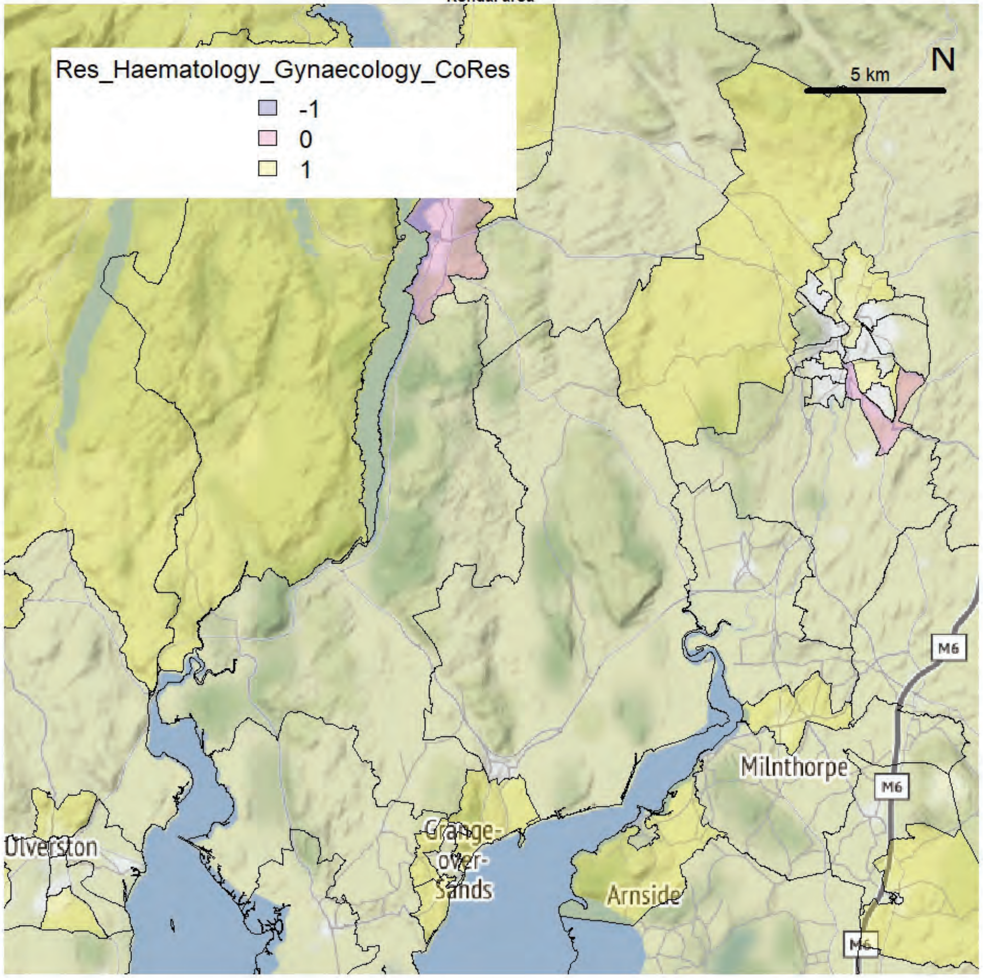

Kendal area

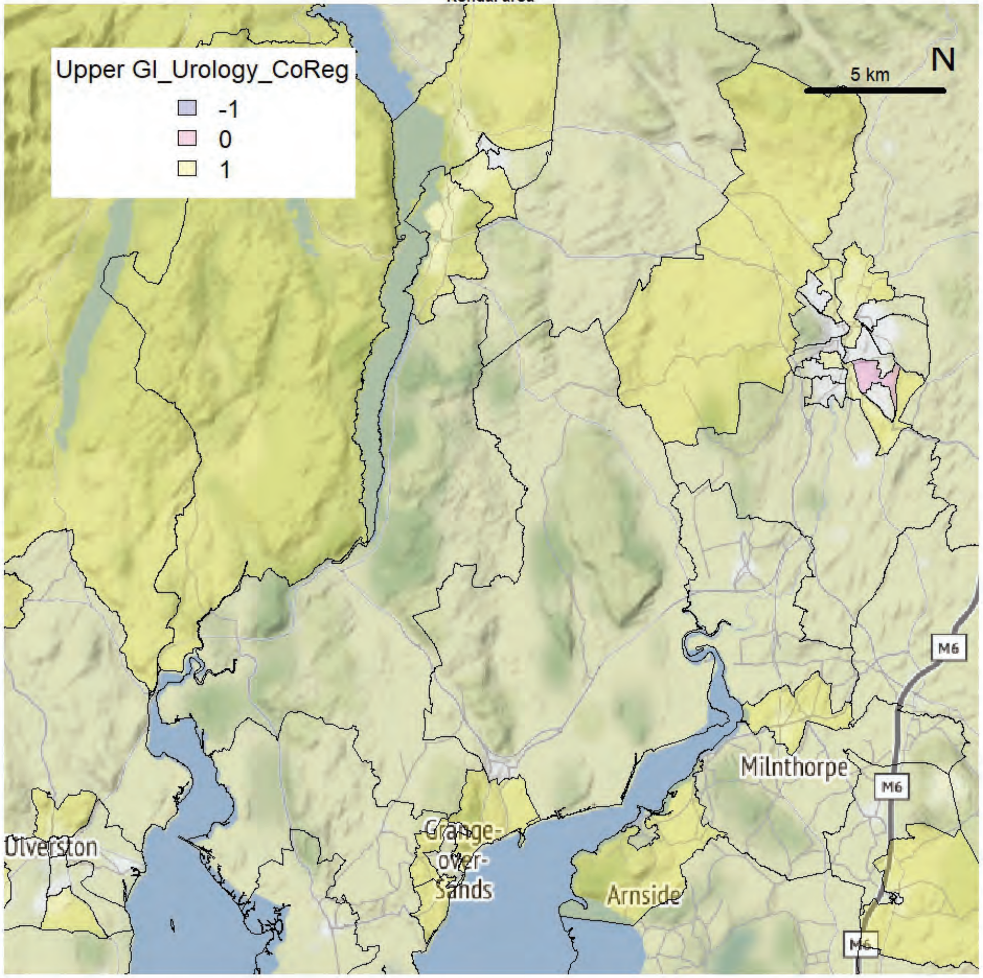


Kendal area

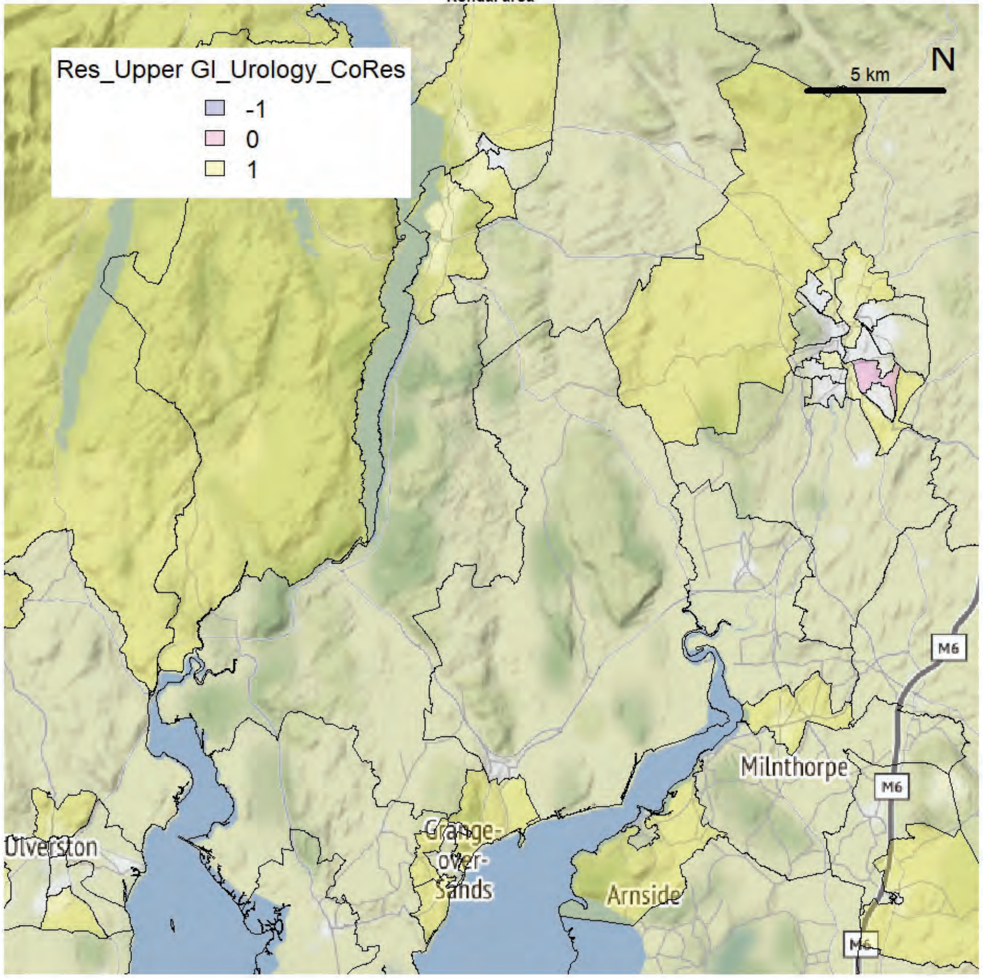

Kendal area

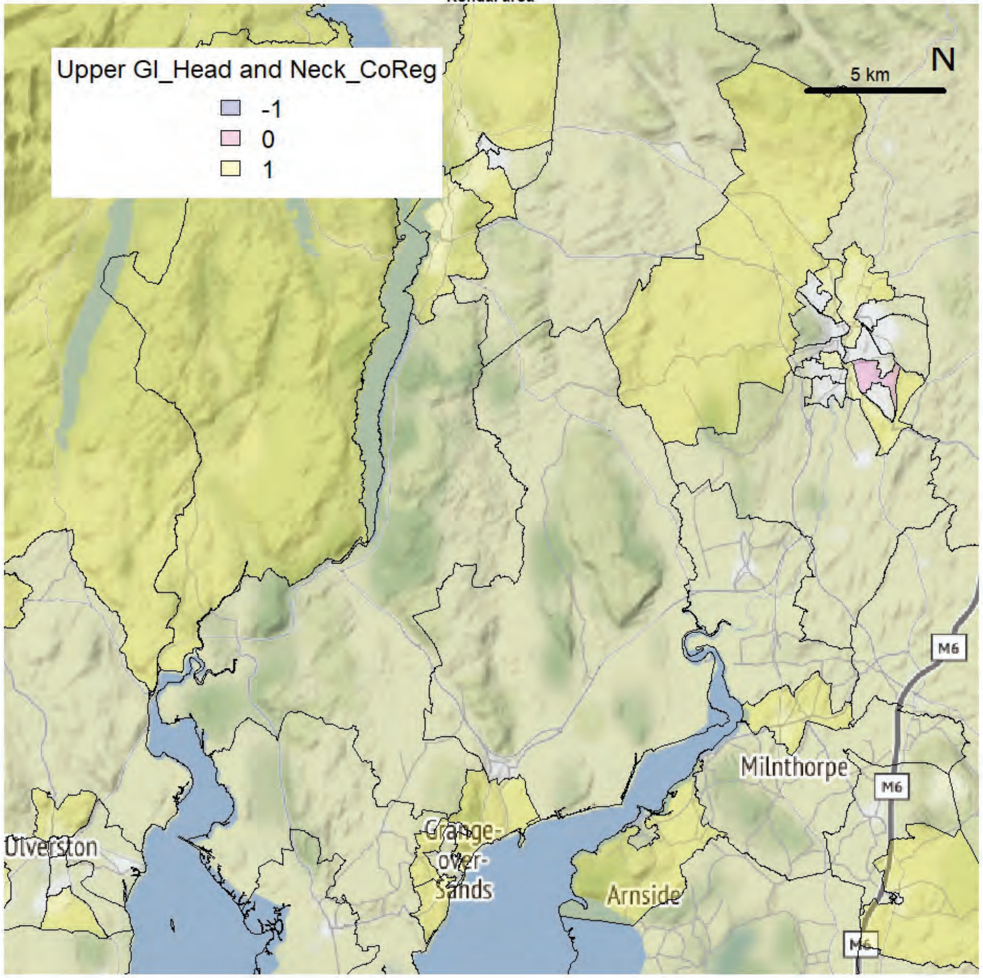


Kendal area

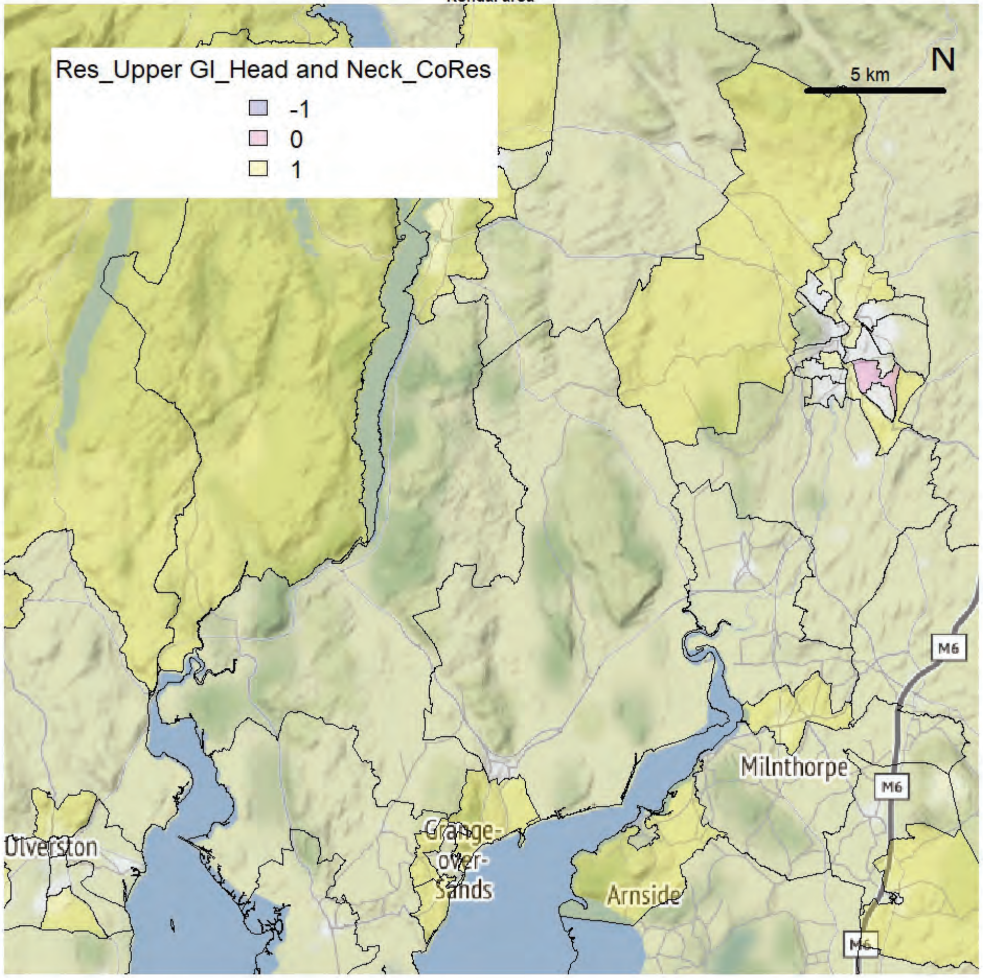

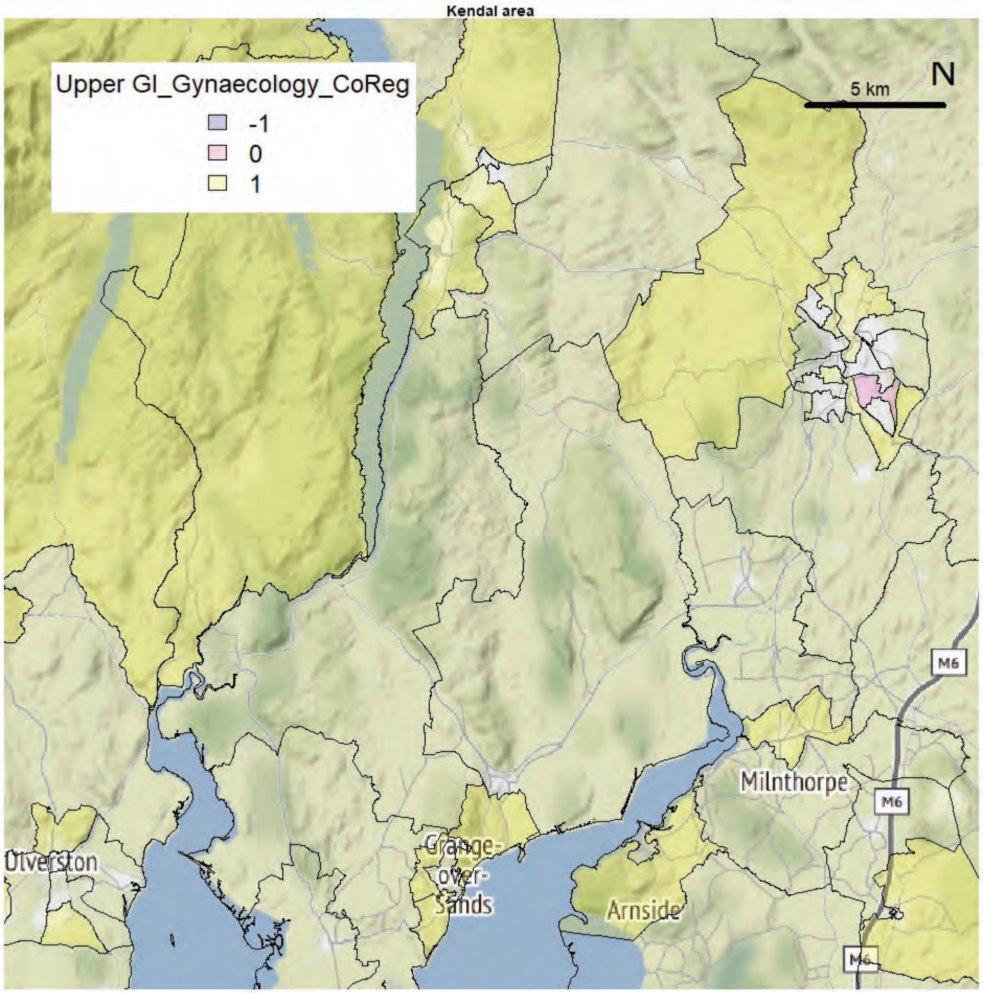
Kendal area
Upper GI_Gynaecology_CoReg
-1
0
1
5 km
N
M6
M6
M6
Milnthorpe
Ulverston
Grange-over-Sands
Arnside

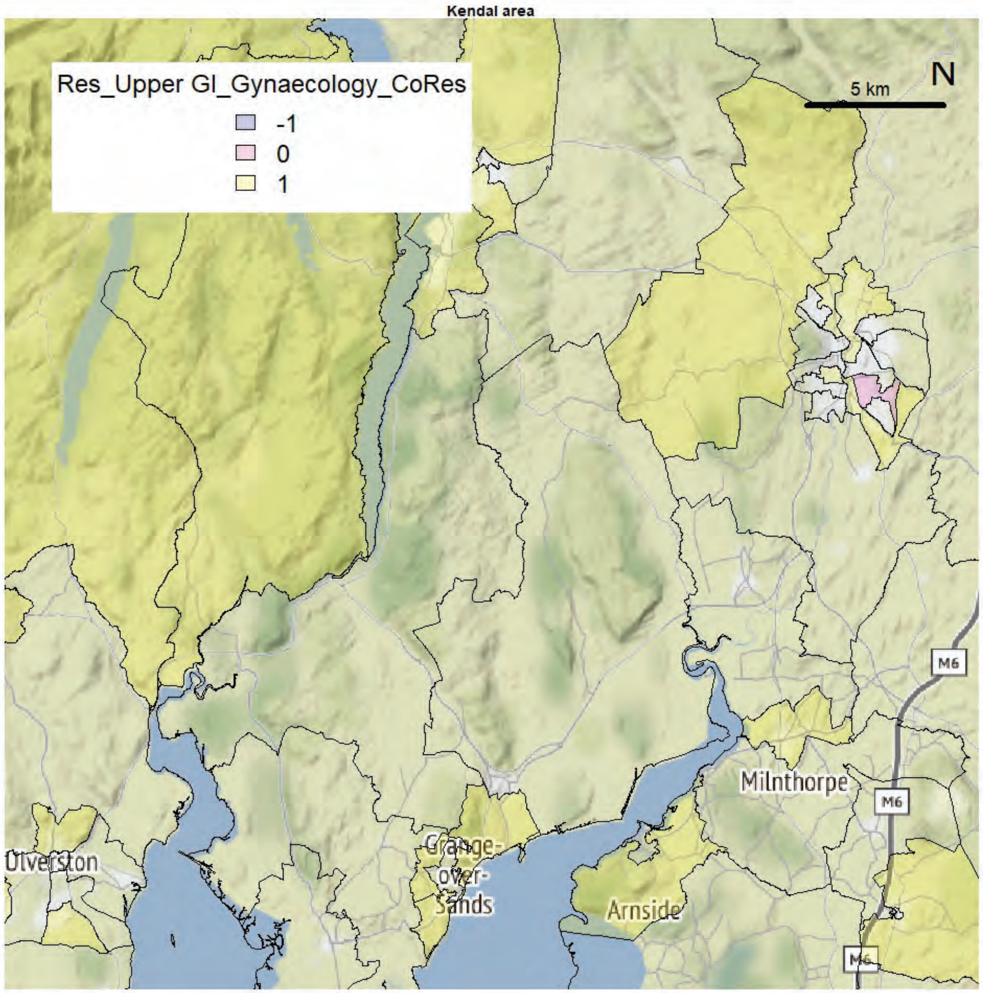
Kendal area
Res_Upper GI_Gynaecology_CoRes
-1
0
1
5 km
N
M6
M6
M6
Milnthorpe
Ulverston
Grange-over-Sands
Arnside

Kendal area

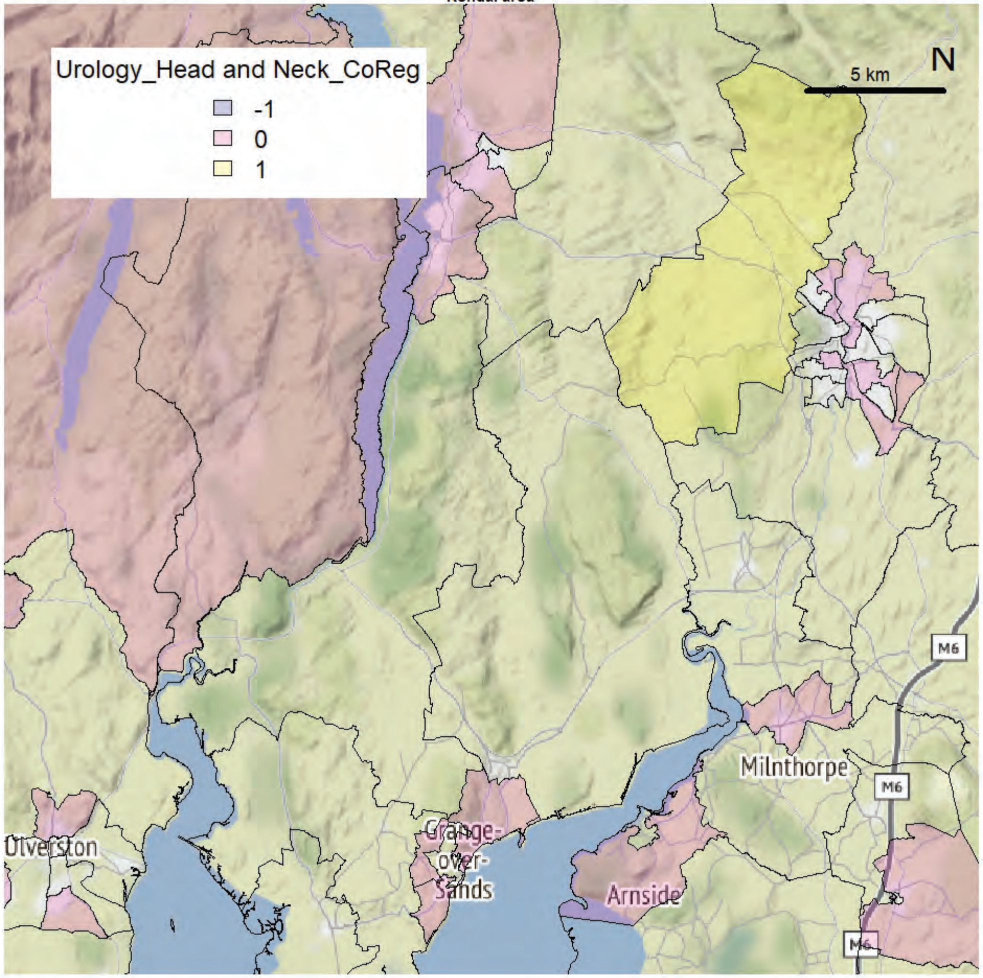


Kendal area

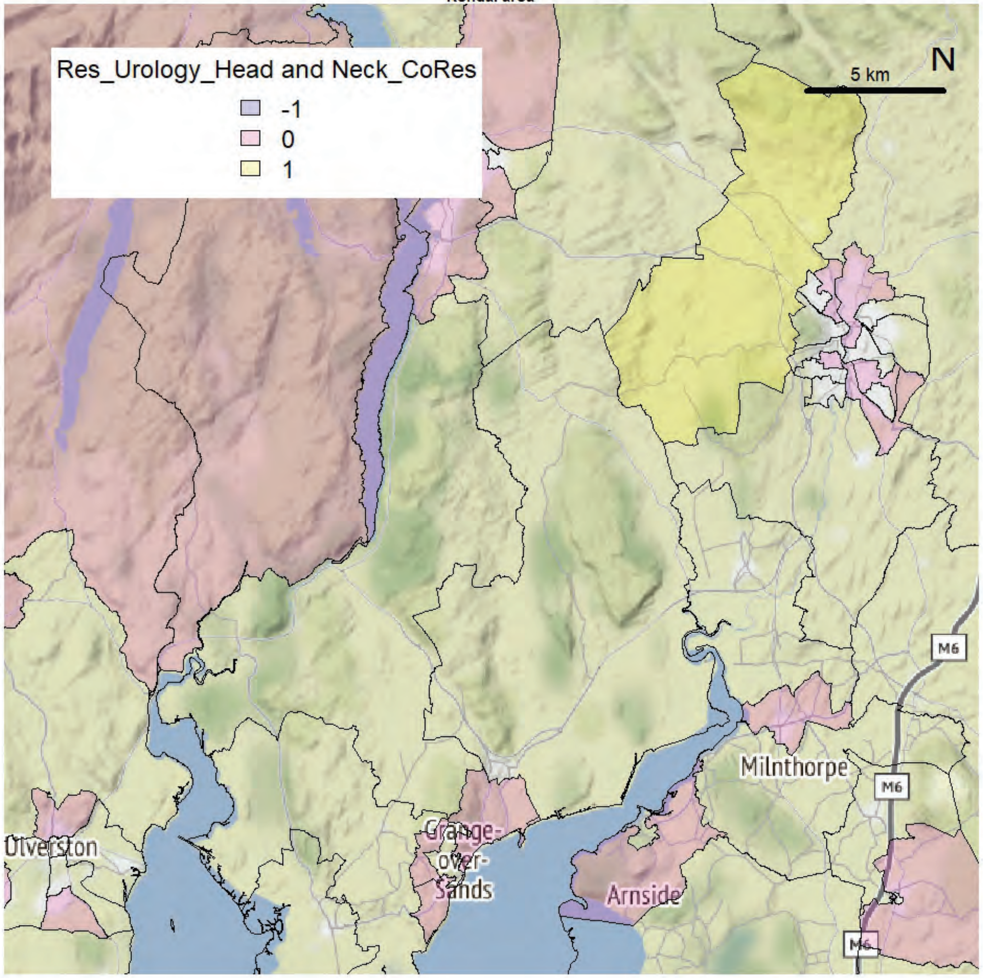

Kendal area

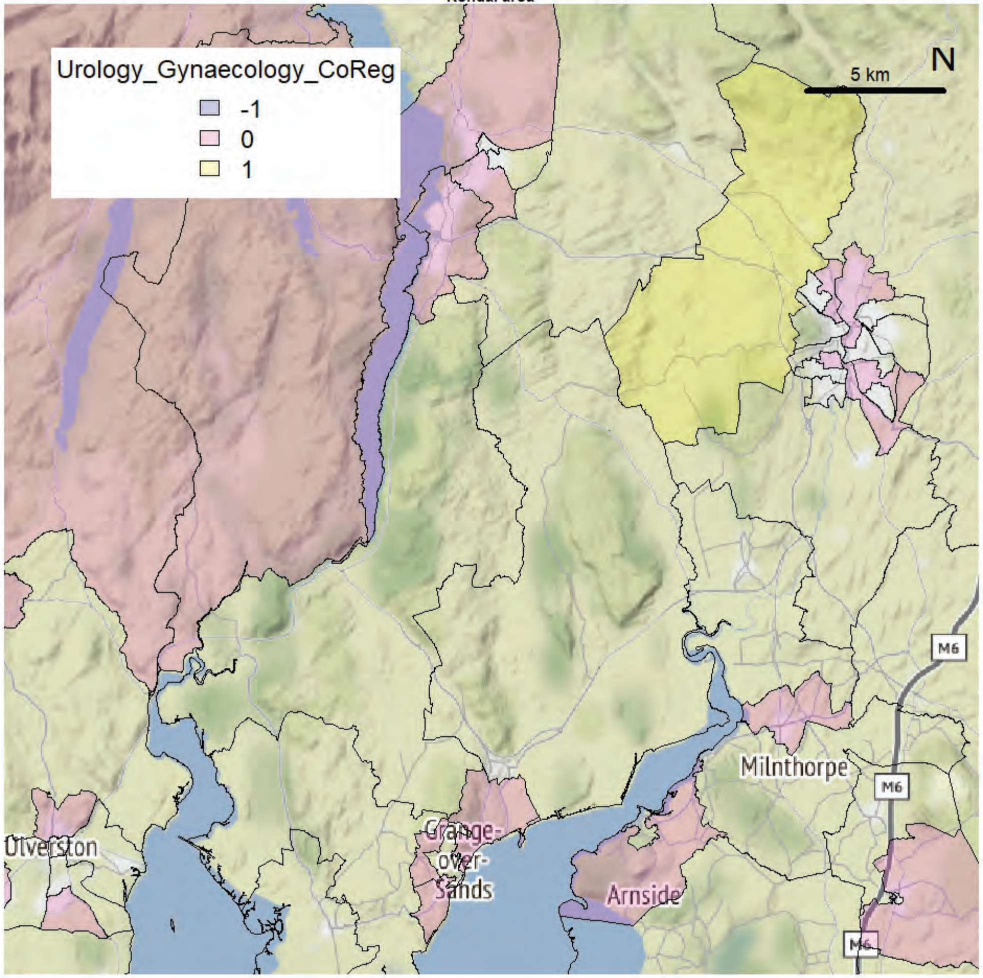


Kendal area

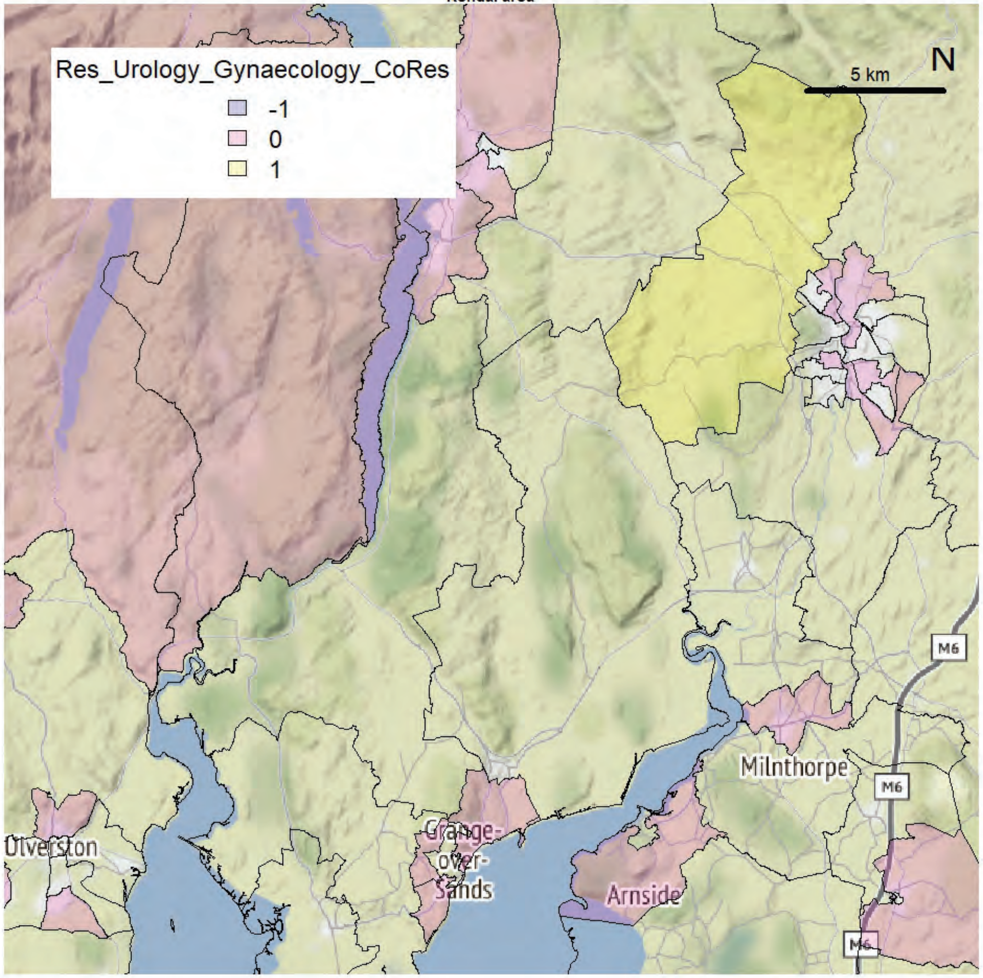

Kendal area

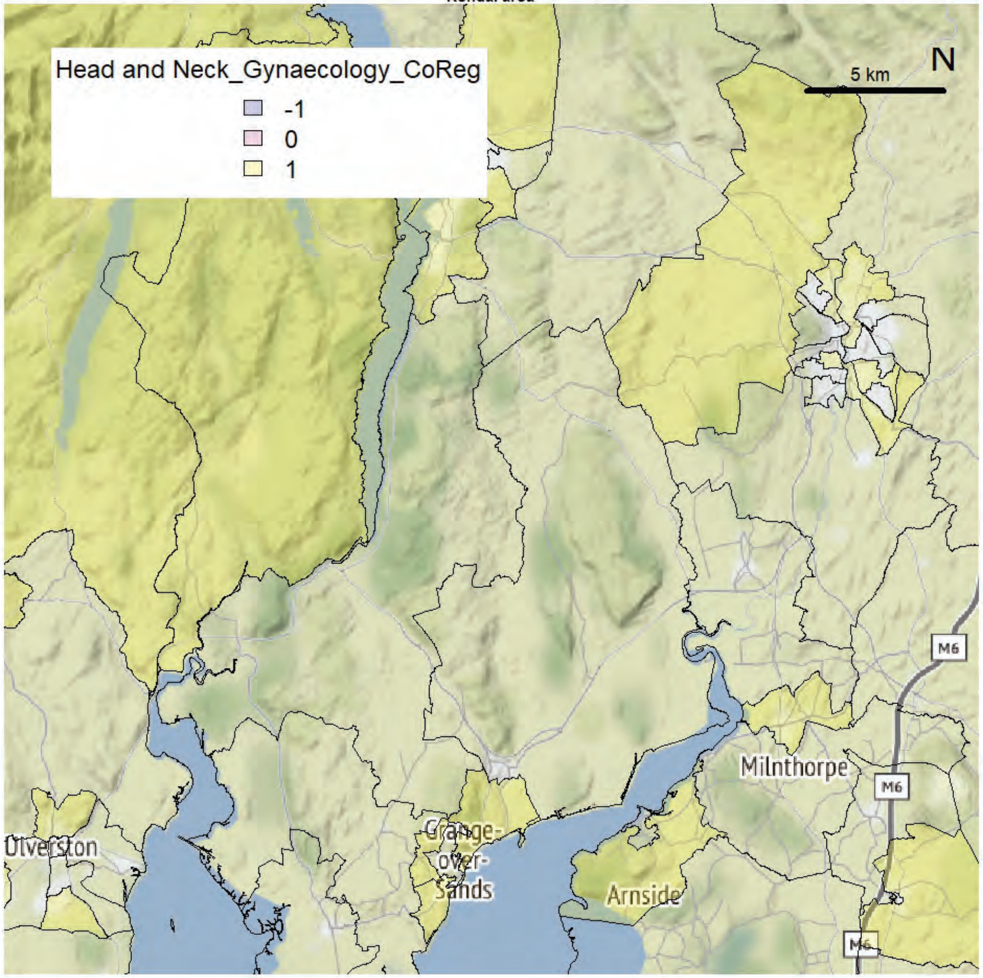


Kendal area

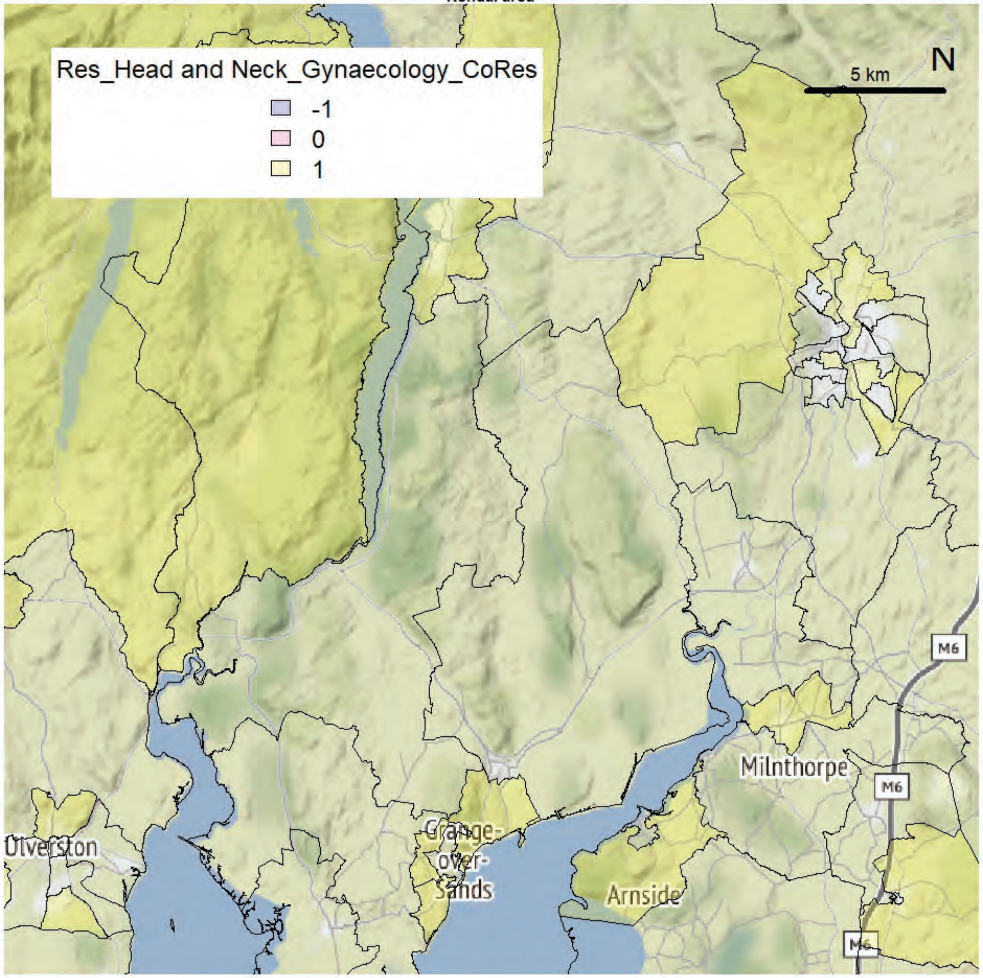
